# Functional analysis across model systems implicates ribosomal proteins in growth and proliferation defects associated with hypoplastic left heart syndrome

**DOI:** 10.1101/2022.07.01.22277112

**Authors:** Tanja Nielsen, Anaïs Kervadec, Jeanne L. Theis, Maria A. Missinato, James Marchant, Michaela Romero, Katya Marchetti, Aashna Lamba, Xin-Xin I. Zeng, Marie Berenguer, Stanley M. Walls, Analyne Schroeder, Katja Birker, Greg Duester, Paul Grossfeld, Timothy J. Nelson, Timothy M. Olson, Karen Ocorr, Rolf Bodmer, Georg Vogler, Alexandre R. Colas

## Abstract

Hypoplastic left heart syndrome (HLHS) is the most lethal congenital heart disease (CHD). The pathogenesis of HLHS is poorly understood, and due to the likely oligogenic complexity of the disease, definitive HLHS-causing genes have not yet been identified. Postulating impaired cardiomyocyte proliferation as a likely important contributing mechanism to HLHS pathogenesis, and we conducted a genome-wide siRNA screen to identify genes affecting proliferation of human iPSC-derived cardiomyocytes (hPSC-CMs). This yielded ribosomal protein (RP) genes as the most prominent class of effectors of CM proliferation. In parallel, whole genome sequencing and rare variant filtering of a cohort of 25 HLHS proband-parent trios with poor clinical outcome revealed enrichment of rare variants of RP genes. In addition, in a familial CHD case we identified a rare, predicted-damaging promoter variant affecting *RPS15A* that was shared between the HLHS proband and a distant relative with CHD. Functional testing with an integrated multi-model system approach reinforced the idea that RP genes are major regulators of cardiac growth and proliferation, thus potentially contributing to the hypoplastic phenotype observed in HLHS patients. Cardiac knockdown (KD) of RP genes with promoter or coding variants (*RPS15A, RPS17, RPL26L1, RPL39, RPS15*) reduced proliferation in generic hPSC-CMs and caused malformed hearts, heart-loss or even lethality in *Drosophila*. In zebrafish, diminished *rps15a* function caused reduced CM numbers, heart looping defects, or weakened contractility, while reduced *rps17* or *rpl39* function caused reduced ventricular size or systolic atrial dysfunction of the atrium, respectively. Importantly, genetic interactions between *RPS15A* and core cardiac transcription factors *TBX5* in CMs, *Drosocross, pannier* and *tinman* in flies, and *tbx5* and *nkx2-7* (*nkx2-5* paralog) in fish, support a specific role for RP genes in heart development. Furthermore, *RPS15A* KD-induced heart/CM proliferation defects were significantly attenuated by *p53* KD in both hPSC-CMs and zebrafish, and by Hippo activation (*YAP/yorkie* overexpression) in developing fly hearts. Based on these findings, we conclude that RP genes play novel critical roles in cardiogenesis and constitute an emerging class of gene candidates likely involved in HLHS.

## Introduction

Hypoplastic left heart syndrome (HLHS) accounts for 2-3% of all cases of congenital heart disease (CHD) [1, 2] and is characterized by underdevelopment of the left ventricle, mitral and aortic valves, and aortic arch [3]. HLHS has a recognized genetic component based on its familial association with left-sided obstructive congenital heart diseases [4–6]. However, segregation analyses in multiplex HLHS-CHD families [6] and genome-wide association studies (GWAS) of large cohorts [5] have lacked sufficient power to conclusively identify candidate HLHS-susceptibility genes with small to moderate effect sizes. Defects in a small number of genes involved in cardiogenesis, such as *NOTCH1* [7], *NKX2–5* [8], *MYH6* [9], and *GATA4* [8, 10] have been implicated as contributors to HLHS, as well as multiple other CHDs. However, the variety of phenotypic manifestations of HLHS, together with numerous studies linking it to diverse genetic loci [7–9, 11], suggests that HLHS is genetically heterogeneous and has a multigenic etiology [12, 13].

It has also been hypothesized that the underdevelopment of the left ventricular myocardium includes restricted blood flow across the mitral valve and its hemodynamic effect during ventricular growth and development (“no flow – no grow” [14]), as well as endocardial defects [15]. Therefore, HLHS manifestation may be due to a combination of cardiomyocyte autonomous and non-autonomous genetic as well as mechanical effects, all of which likely affect proliferation and differentiation in the developing heart.

In a digenic HLHS mouse model, loss of both Sin3A Associated Protein 130 (*Sap130)* and the protocadherin *Pcdha9* cause decreased CM proliferation resulting in left ventricular (LV) hypoplasia [16]. In addition, LVs from HLHS patients have cardiac damage associated with CM proliferation defects compared to healthy subjects [17]. These studies suggest that intrinsically defective cardiac differentiation and impaired CM proliferation are likely contributing to HLHS-associated heart defects. Further strengthening this hypothesis, we found that HLHS-patient-derived hPSC-CMs exhibited reduced proliferation compared to the parents [18]. There is a need to identify the potentially causative genes, which contribute to those processes described above and to elucidate their role HLHS pathogenesis.

To advance CHD gene discovery, we have developed a multi-model systems platform enabling to both rigorously prioritize candidate genes from whole-genome sequencing (WGS) based on rare, predicted-damaging variants and their mode of inheritance, and to functionally characterize gene function in complementary and genetically tractable model systems: human induced pluripotent stem cells (hPSC), the fruit fly *Drosophila melanogaster* and zebrafish *Danio rerio*. The genetic basis of cardiac development, originally uncovered in *Drosophila*, is fundamentally conserved across species [19–22] and ∼80% of human disease genes have fly orthologs [23]. In addition, the non-redundant fly genome together with the simple structure and function of the fly heart allow straightforward genotype-phenotype correlations. Zebrafish [24–26], which has a two-chambered heart and can be easily manipulated using morpholino injections and CRISPR technologies to affect heart development and function. In addition, phenotypes can be directly observed in the developing larva for several days post fertilization [27]. Finally, cardiomyocytes (CMs) differentiated from hPSCs (hPSC-CMs) enable to identify and quantify cellular phenotypes associated with human diseases, including HLHS [7, 28–30] and are amenable to large scale functional screens [31, 32], thereby enabling to serve both as a discovery and validation platform. This integrated approach allows to rapidly identify novel HLHS-associated gene candidates and to characterize their ability to regulate developmental processes associated with disease (e.g., hypoplasia) [18].

Defective CM proliferation is likely a hallmark of HLHS [16–18], thus to uncover novel regulators of human CM proliferation with potential links to HLHS, we performed a whole genome siRNA screen (18,055 genes) using an EdU assay in hPSC-CMs and identified ribosomal proteins (RPs) as the major class of genes controlling CM proliferation. In parallel, whole genome sequencing (WGS) of 25 poor-outcome HLHS proband-parent trios followed by unbiased variant filtering, revealed an enrichment for rare, predicted-damaging variants in RP genes. Moreover, analysis of a high-value family (75H) comprised of a HLHS proband, his phenotypically normal parents, and a fifth degree relative born with left-sided CHD led to the identification of a predicted-damaging variant in RP protein gene *RPS15A* that segregated with disease. Consistent with a central role for RPs in the regulation of heart development, heart-specific knock-down (KD) of HLHS-associated RPs in *Drosophila* caused severe phenotypes, including near complete heart-loss and lethality. Similarly, in zebrafish, *rps15a* CRISPR mutants and morphants had greatly reduced heart size and CM number, looping defects and diminished contractility. In this context, a multi-model system evaluation (hPSC-CMs, flies and zebrafish) of RP function show their important role on heart growth and differentiation and CM proliferation by modulating the p53, Myc and Hippo pathways. Moreover, we find evidence that RPs may regulate heart development and CM proliferation in a cardiac-specific manner by synergistically interacting with core cardiac transcription factors such as T-box, Nkx and Gata factors. In sum, here we show that RP genes are potent regulators of CM proliferation and cardiac growth and differentiation, consistent with a potentially critical role in the hypoplastic phenotype of HLHS. We suggest that RP genes are an emerging class of novel genetic effectors in HLHS.

## Results

### Whole genome siRNA screen identifies ribosomal proteins as central regulators of cardiomyocyte proliferation

Based on previous and our recent data [16–18], impaired CM proliferation is emerging as an important mechanism in HLHS pathogenesis. Thus, to comprehensively map the human genome for novel regulators of CM proliferation, we screened a library of 18,055 siRNAs for their ability to modulate proliferation in human iPSC-derived CMs (hPSC-CMs). To assess CM proliferation, we used a dual read-out and quantified both DNA synthesis using EdU incorporation [31], and total number of CMs, 3 days after treatment (**Figure 1A-C**). Briefly, siRNAs were transfected into 25-day old hPSC-CMs [29, 33]. After 2 days, EdU was added to the wells for 24 hours. On day 3, cells were fixed and co-stained for sarcomeric protein alpha-Actinin (ACTN2), EdU, and DAPI. Next, the number of EdU/alpha-Actinin double-positive cells, and the total number of CMs were determined using a commercially available high-throughput image analysis software (MetaXpress, Molecular Devices)[31]. Note that alpha-Actinin-, EdU+ cells which typically express fibroblast markers (i.e POSTN)[34], were excluded from this analysis, to focus on CM biology only. In total, we found 153 siRNAs that decreased proliferation (< 0.5-fold EdU incorporation) and number of CMs (<0.8-fold), and 162 siRNAs that increased EdU (>1.2-fold) and CM number (>1.1-fold; **Figure 1B, C and Supplemental Table 1 WGS data**). Consistent with previous studies [35, 36], KD of cell cycle checkpoint agonist, CHEK1 and MDM2, the negative regulator of TP53, both were among top hits that reduced CM proliferation. As expected, we identified TP53 and CDKN1A among the genes that significantly increased CM cell number upon KD (**Figure 1B, C**). Next, to gain insight into the molecular pathways regulating proliferation in hPSC-CMs, we performed Gene Ontology (GO) term analysis of hits decreasing EdU incorporation or CM numbers and found an enrichment for genes associated with “translation/ribosome” and p53 signaling as top hits (**Figure 1D, E**). Consistent with these observations, we had previously found that the p53 pathway is dysregulated in hPSCs differentiated from a HLHS family trio [18]. Remarkably, the most represented gene family are ribosomal proteins (RPs), KD of which caused consistently the strongest inhibition of cell proliferation (**Figure 1B**). Next, since, the primary screen was performed as a single data point, we next sought to validate these findings by re-testing all 80 RP genes for function [37] using a distinct set of siRNAs in biological quadruplicate conditions. Remarkably and consistent with the primary screen, KD of 59/80 RPs significantly reduced proliferation as compared to siControl (p<0.05; **Supplemental Figure 1A, B**). In this context, further functional testing in fibroblasts revealed that the antiproliferative effect of RP loss of function is cell type–independent (**Supplemental Figure 1C, D)**. Together, these findings establish RP function as essential for cell proliferation, including in hPSC-CMs, and suggest that RPs may represent a previously unrecognized class of regulators of ventricular growth during embryonic development.

**Figure 1:**
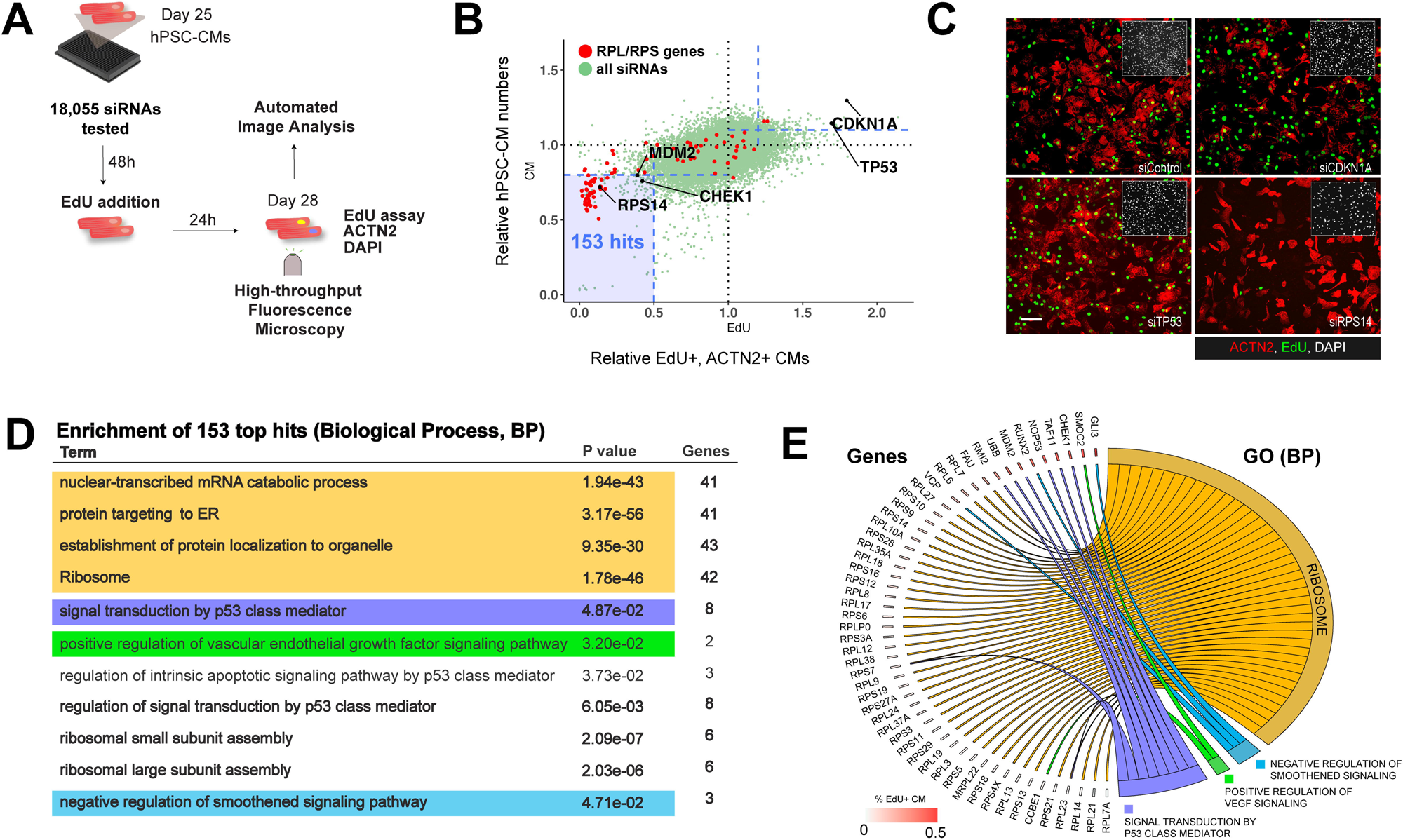
Whole Genome siRNA Screen identified Ribosomal proteins as agonists of cardiomyocyte proliferation. **(A)** High-throughput iPSC-derived CM proliferation screen overview. **(B)** Screen result showing normalized % EdU+ CMs (X-axis) and relative total number of CMs (Y-axis) upon knockdown of genome-wide siRNAs (18,055 siRNAs). siRNAs for RPL and RPS genes highlighted in red. **(C)** Representative immunofluorescence images of proliferation (EdU incorporation, green, CM marker ACTN2, red) of induced hPSC-CMs upon *TP53* and *RPS14* knockdown. Insets: nuclei (DAPI) **(D)** Gene ontology enrichment analysis for WGS hits (BP, biological process; FDR-corrected analysis using gprofiler2). (**E**) Overview of hits corresponding to top 4 non-redundant BP categories.

### Enrichment of RP gene variants in an HLHS patient cohort with poor clinical outcome

To identify candidate genes involved in HLHS pathogenesis, we performed whole genome sequencing (WGS), variant filtering, and enrichment analysis (see also [18], and Methods: WGS and bioinformatic strategies) in a cohort of 25 HLHS patient-parent trios with poor clinical outcome defined as either prenatal – restrictive atrial septal defect (n=2); postnatal – reduced right ventricular ejection fraction following stage II or III surgical palliation (n=19); protein-losing enteropathy (n=2); or cardiac transplantation/failed surgical palliation (n=2). The 25 probands were comprised of 18 males and 7 females, 24 of whom were of white ancestry. The HLHS phenotype with respect to valve morphology (mitral atresia, MA; mitral stenosis, MS; aortic atresia, AA; aortic stenosis, AS) was: MA/AA in 10, MS/AS in 9, MA/AA in 4, MA/AS in 1 and unknown in 1. CHD in the parents was excluded by echocardiography in all but two families, in which a bicuspid aortic valve was present in the mother. Genomic sequences were filtered for uncommon (MAF < 1%) *de novo* and recessive variants in coding and regulatory regions of genes expressed in the developing heart and predicted to alter protein structure or expression, yielding 292 unique HLHS candidate genes that primarily fit a recessive mode of inheritance (Figure **2A** **and Supplemental Table 2**). To determine whether certain gene networks were over-represented among these variants, we used two online bioinformatics tools (STRING and PANTHER [38, 39]). After applying Fisher’s exact test and false discovery rate corrections, RPs were the most enriched class of proteins when compared to the reference proteome, which includes data annotated by protein class (5.24-fold, p=0.0028), and cellular component (7.27-fold, p=0.011). In total, 14 variants found in 9 RP genes among 6 HLHS probands were identified, most fitting a recessive inheritance disease model (**Table 1**). These RP variants encompass mutations in upstream promoter regions that potentially affect transcription factor binding sites, as well as non-synonymous substitutions inside the protein coding regions.

**Figure 2:**
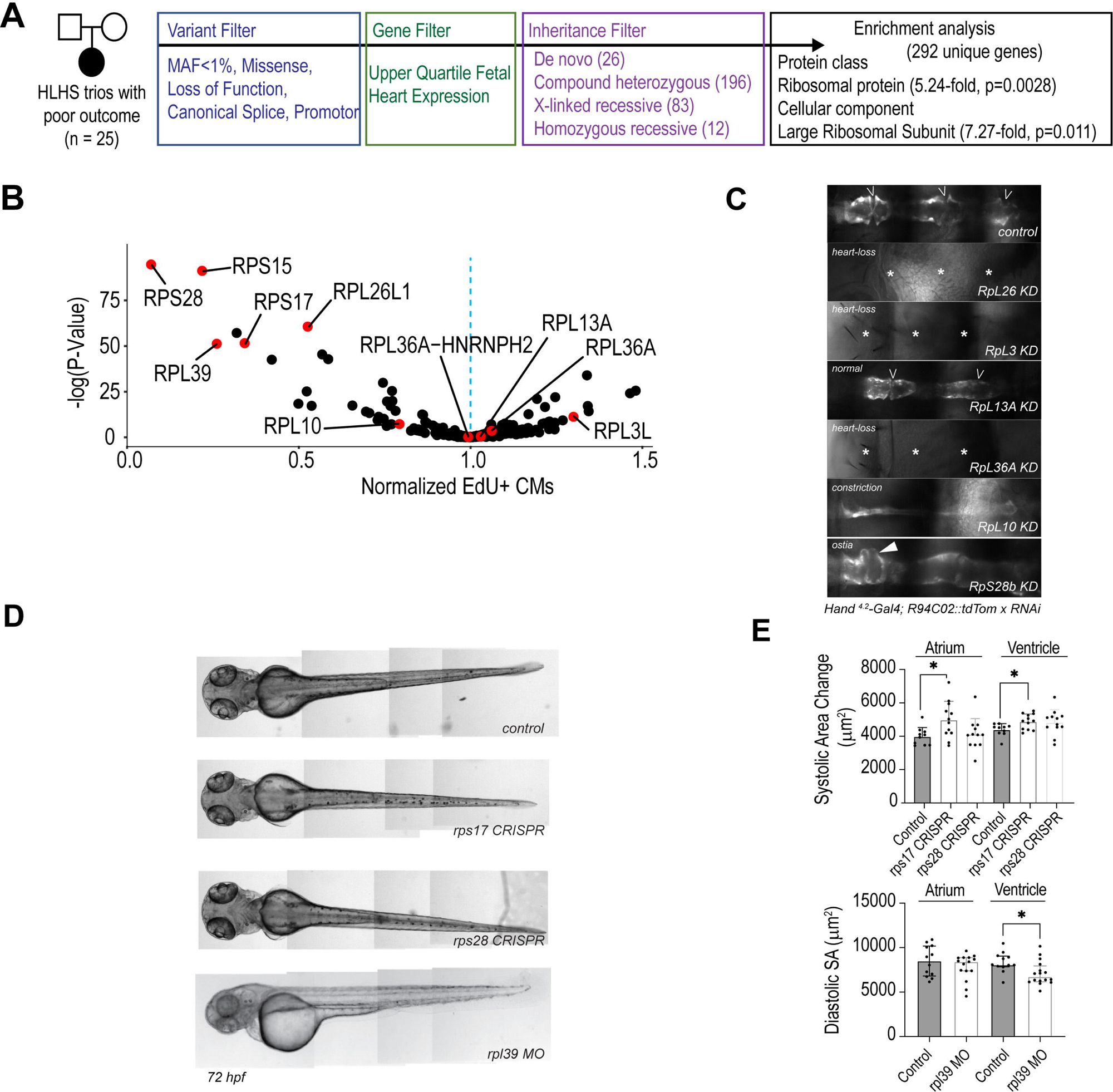
Ribosomal gene variants identified in HLHS. **(A)** Gene prioritization scheme of 25 poor-outcome proband-parent trios. **(B)** Testing 292 HLHS candidate genes from all poor-outcome families in CMs identified RPs as major regulators of hPSC-CM proliferation (Normalized fraction of ACTN2+/EdU+ cells). **(C)** *Drosophila* cardiac phenotypes induced by loss of RP genes affected in HLHS patients with poor outcome. Heart is visualized by RFP expression specifically in cardiomyocytes (R94C02::tdTom). Knockdown is achieved by sustained Gal4/UAS activity using Hand4.2-Gal4. (**D**) Wild-type zebrafish larva, *rps17* and *rps28* CRISPR mutants and *rpl39* morphants at 72 hpf. *rpl39* morphants, injected with 1 ng MO in lateral view, show mild edema. **(E)** Systolic surface area (SA) upon *rps17* and *rps28* CRISPR, and diastolic SA after *rpl39* MO in the atrium and ventricle of zebrafish hearts.

**Table 1:** Table 1 – Variants in ribosomal genes in HLHS probands. Abbrevations: MAF – minor allele frequency; TFBS – transcription factor binding site; n.t. – not tested; ASD – atrial septal defect; PLE - Protein-losing enteropathy; RV – right ventricle; RVEF – right ventricular ejection fraction; RVEDP - right ventricular end diastolic pressure

Next, all 292 prioritized candidate genes were systematically evaluated for effects on the proliferation of hPSC-CMs *in vitro* and for cardiac differentiation of fly hearts *in vivo.* Again, loss of RP function (*RPL39, RPL26L1, RPS15, RPS17, RSPS28*) caused the most severe phenotypes on proliferation in hPSC-CMs (**Figure 2B, Supplemental Table 3**), which was consistent with our genome-wide proliferation screen (see **Figure 1B**). In the fly, KD of RPs in the cardiac lineage and throughout embryonic development was achieved using a *Hand^4.2^*-Gal4 driver (see [40] and methods) and led to partial or complete heart loss (*RpL26, RpL36A, RpL3*) or caused lethality (*RpS17, RpL39, RpS15, RpL26, RpL36A*) (Figure **2C**). Also, KD of *RpL10* gave rise to severely constricted hearts, while KD of *RpS28b* caused inflow tract defects (**Figure 2C**). Finally, functional testing of RPs in zebrafish embryos revealed that KD of *rpl39* leads to reduced ventricle size, whereas *rps17* CRISPR mutants exhibit systolic dysfunction (**Figure 2D, E**). In sum, our patient centric approach (cohort of 25 probands) identifies RPs as the most enriched gene category affected by HLHS-associated variants (STRING, PANTHER analysis). In this context, systematic testing in multi-model systems (fly, zebrafish, hPSC-CMs) identifies RPs as most potent regulators of cardiac differentiation, proliferation and contractility among HLHS-associated genes. Collectively, these findings highlight that RPs regulate critical steps of cardiogenesis that are also found to be defective in HLHS probands [41, 42] and thus suggest a potential link between RP biology and HLHS phenotypic etiology.

### RPS15A variant associated with HLHS in familial CHD

To further establish a phenotypic link between RP function and HLHs associated phenotypes, we selected a rare familial CHD case (75H) (**Supplemental Figure 2A**), that is comprised of a young teenager with HLHS (MS/AS) and normal ventricular function following Fontan operation, his phenotypically normal parents, and a 5^th^ degree female relative born with left-sided CHD (bicuspid aortic valve and coarctation of the aorta). Based on these observations, we hypothesized the presence of a heterozygous driver variant exhibiting incomplete penetrance and variable expression. To investigate further, we filtered whole-genome sequencing (WGS) data for rare, predicted-damaging variants in coding and regulatory regions. This analysis identified six prioritized variants (**Supplemental Table 4**), including one located in the *RPS15A* gene locus, shared by the HLHS proband, the unaffected mother, and the proband’s fifth-degree relative with congenital heart disease (CHD).

We next investigated whether family members carrying these variants (the mother and proband) exhibit defects in cardiomyocyte (CM) proliferation compared to the father. Notably, analysis of EdU incorporation in CMs from the parent-proband trio revealed significantly reduced proliferation in both the proband and mother compared to the father. However, the proband displayed a markedly more severe phenotype than the mother (**Figure 3 A,B)**. Interestingly, this reduction in proliferation appeared to be specific to cardiomyocytes (CMs), as EdU incorporation in the non-CM cell population showed no significant differences among the family members (**Supplemental Fig. 2B**). Collectively, these findings suggest a phenotypic association between the presence of the variants and impaired CM proliferation.

**Figure 3:**
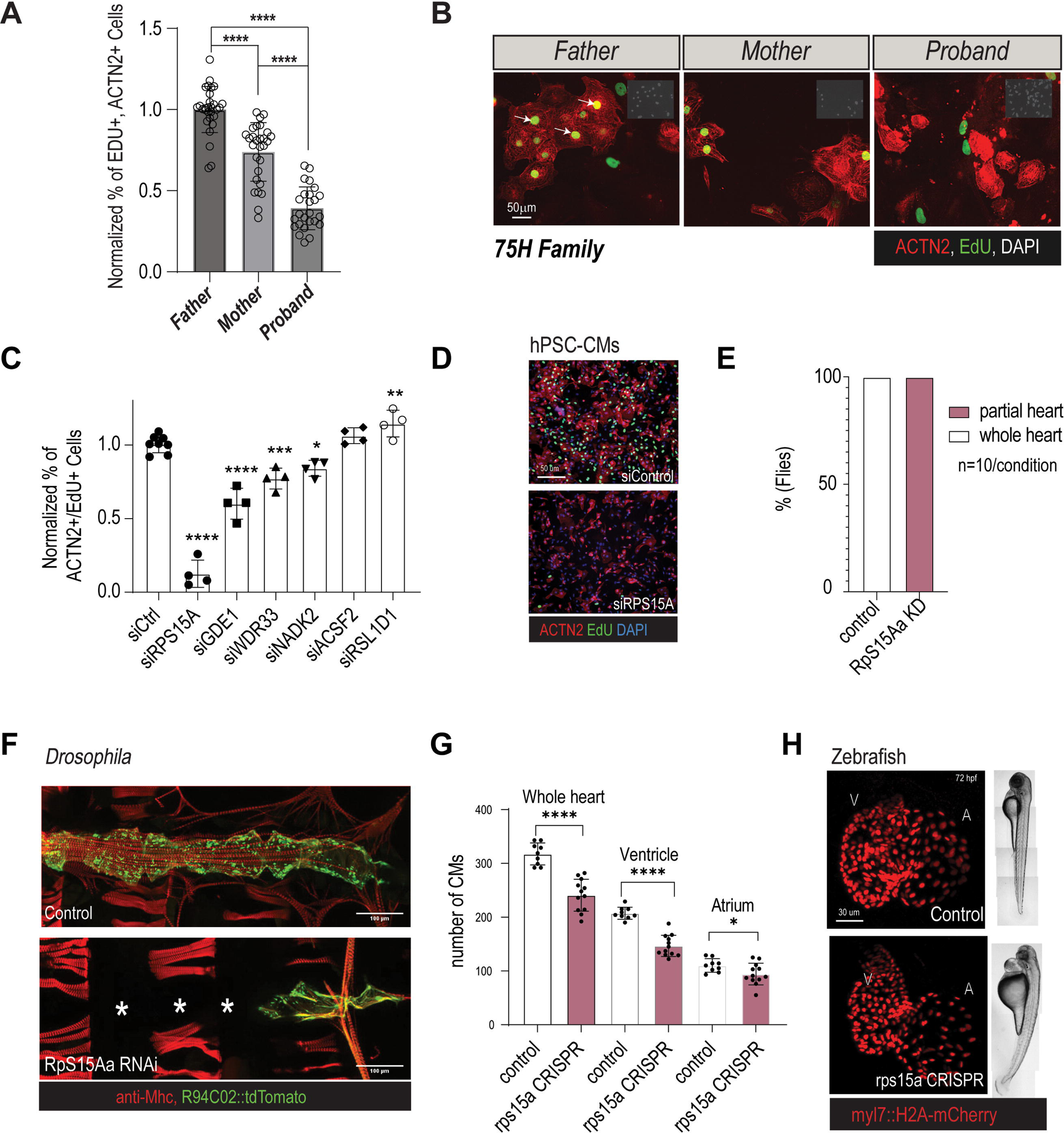
Characterization of RPS15A from 75H HLHS index family. **(A) (B) (C)** Prioritized candidate genes from the 75H family and relative hPSC-CM proliferation capacity upon KD. (**D**) Representative immunofluorescence images of proliferation (EdU incorporation, green; CM marker ACTN2, red) of induced hPSC-CMs upon siRPS15A knockdown. **(E)** Heart-specific KD of *RpS15Aa* in *Drosophila* adult hearts cause loss of heart tissue and is fully penetrant. **(F)** Representative immunofluorescence images of control and *RpS15Aa*-RNAi shows partial heart loss (Myosin heavy chain, Mhc, red; heart tissue-reporter, green). **(G)** CRISPR-mediated loss of *rps15a* in F_0_ larval zebrafish hearts causes decrease in CM number. **(H)** Representative immunofluorescence of hearts and whole-mount images of control and *rps15a*-CRISPR F_0_ larval hearts (CM nuclei reporter, red).

Finally, to determine the potential relative contribution of the six genes harboring rare and damaging variants in the regulation of CM proliferation, we performed functional KD experiments in hPSC-CMs *in vitro*. Remarkably, while KD of 5 of the other prioritized genes from the 75H family also resulted in moderate reduced CM proliferation, RPS15A KD produced the most pronounced effect, with over an 85% reduction in proliferation, emerging as the top candidate (**Figure 3C, D**). Together, these findings indicate that RPS15A plays a critical role in regulating cardiomyocyte (CM) proliferation in human pluripotent stem cells (hPSCs) and may influence heart developmental processes that are disrupted in HLHS.

### RPS15A KD in *Drosophila* and zebrafish causes severe cardiac proliferation and differentiation defects

To characterize the role of *RPS15A* during heart development, we first induced a heart-specific KD of *RpS15Aa* in flies (see methods), which caused a partial or complete loss of the heart (**Figure 3E,F**). Note that this phenotype is consistent with our results above and a previous study involving another RP gene (*RPL13*) [43].

Next, to evaluate the role of *RP* genes during heart development, we knocked-out *rps15a* in zebrafish (*Danio rerio*) using CRISPR [24–26]. We examined F_0_ larva at 72 hpf (hours post fertilization) using high speed digital imaging (SOHA [44], see also methods) and confocal microscopy to monitor heart size and cardiomyocyte numbers respectively. Remarkably, *rps15a* mutant hearts were smaller in size (**Supplemental Fig. 2C,D**), with reduced CM numbers (**Figure 3G,H**). We observed similar effects in response to injection of a *rps15a* morpholino (MO) compared to uninjected larva [45, 46] (**Supplemental Figure 3 C,D**). In this context, we also noted that the overall body plan and morphology of *rps15a* CRISPR mutants or morphants were largely unaffected, except for a mild pericardial edema, a possible indicator of heart dysfunction (**Figure 3H**) [47].

We quantified cardiomyocyte (CM) number using nuclear markers in Tg(*myl7*:*eGFP*); Tg(*myl7*:*H2A-mCherry*) embryos 72 hpf. CM numbers were predominantly reduced in the ventricle compared to the atrium (**Figure 3G,H)**. We also assessed total cardiac cell numbers and cardiac cell proliferation by phospho-histone H3 (PH3) immunostaining and DAPI to quantify total cardiac cells. We again found that the total cell numbers were significantly reduced in both rps15a morphants and crispants (**Supplemental Figure 4A-C**). Importantly, the proportion of proliferating (PH3+) cardiac cells was also decreased in rps15a morphants and crispants at both 24 and 48 hpf, but not at 72 hpf when proliferation normally declines (**Supplemental Fig. 4C**). Collectively, these data support the hypothesis that *RPS15A* is required for CM proliferation and heart morphogenesis (hPSC-CMs, fly and zebrafish), thus suggesting a potential role for RPs as genetic drivers of hypoplasticity observed in HLHS.

### Ribosomal proteins control CM cell cycle by regulating TP53 pathway activity

To investigate how RPs regulate cell cycle activity in hPSC-CMs, we performed RNA-seq upon KD of *RPS15A* and *RPL39*, which belong to the small and the large ribosomal subunits that harbor predicted-damaging variants in 75H and 151H probands, respectively. Comparison of differential gene expression between *siRPS15A* and *siRPL39* KD revealed that ∼53% of genes were commonly dysregulated (**Figure 4A**), indicating the existence of a conserved RP-dependent transcriptional network. Consistent with a central role for RPs in the regulation of proliferation, GO term analysis for differentially expressed genes revealed an enrichment for genes involved in DNA replication, mitosis and DNA damage response mediated by p53, and included the downregulation of positive regulators of cell cycle such as *CDK1*, *CCNA2, AURKA, PCNA* and the upregulation of cell cycle arrest mediators such as *CDKN1A, BTG2, GADD45A* (**Figure 4B** and **Supplemental Tables 5,6**). To confirm these findings, we performed immunostaining for canonical p53 downstream transcriptional target [48], *CDKN1A,* and observed that the percentage of CDKN1A+ CMs was increased by ∼60% in response to *RPS15A* KD (**Figure 4C, D**), indicating that RP KD induces cell cycle arrest *via* the activation of the p53 signaling pathway in hPSC-CMs.

**Figure 4:**
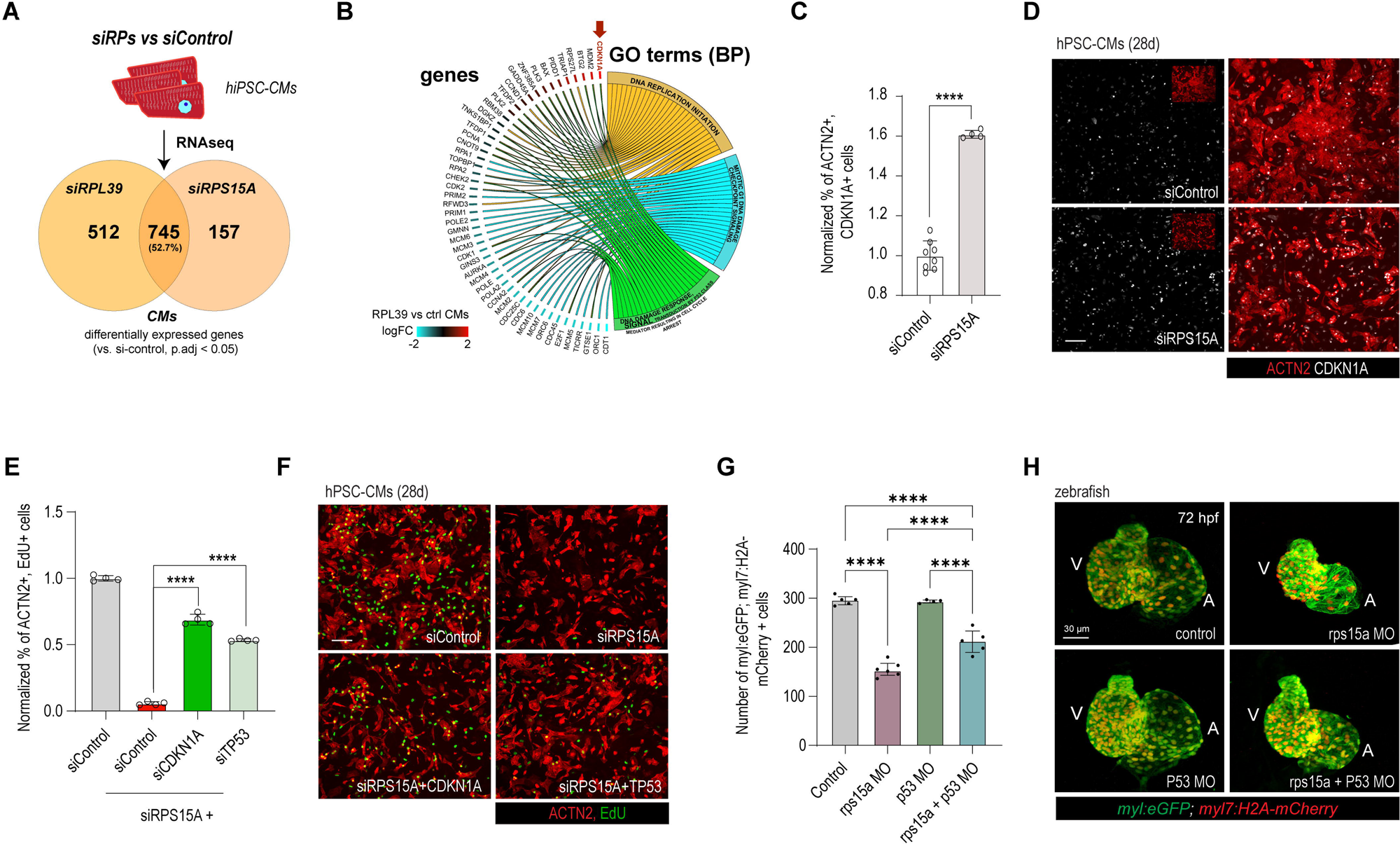
Loss of ribosomal gene function in CMs invokes TP53-stress response. **(A)** RNA sequencing of hPSC-CMs following siRNA-treatment for *RPL39* and *RPS15A* shows both, convergent and divergent transcriptomic response. **(B)** GO term analysis of differentially expressed genes following RP KD shows TP53-mediated response, including upregulation of *CDKN1A*. **(C, D)** CDKN1A is highly upregulated in CMs following *siRPS15A* treatment. **(E, F)** Reduced CM proliferation upon *RPS15A* KD is mediated by *CDKN1A*/*TP53* and can be rescued upon their co-KD. **(G, H)** Larval zebrafish CM number is reduced by morpholino-treatment for rps15a and can be attenuated by P53 morpholino co-injection. Control and morphant (MO) hearts of 72 hpf zebrafish larva stained with *Tg(myl7:EGFP)* and *Tg(myl7:H2A-mCherry)*. Note that smaller heart with aberrant looping by *rps15a* MO is partially reversed by *p53* co-KD. Student’s t-test, *p<0.05, **p<0.01, ***p<0.001, ****p<0.0001.

Next, to evaluate if the reduction in proliferation caused by RP KD is mediated by the p53 signaling pathway in CMs, we co-KD *TP53* or *CDKN1A* along with *RPS15A*, and observed a significant rescue of CM proliferation by EdU incorporation and CM number in comparison to RPS15A KD (**Figure 4E, F** and **Supplemental Figure 4D, E**).

Next, we tested if RP-mediated defects in zebrafish can also be rescued by inhibition of p53. To this aim, we used morpholinos to co-KD *p53* with *rps15a* [49] and examined larval hearts at 72 hpf. *p53* MO-mediated KD on its own had little effect on heart function but partially reversed the *rps15a* MO-mediated reduction in heart size and cardiomyocyte numbers (**Figure 4G, H**). Moreover, bradycardia, heart looping and reduced contractility phenotypes of *rps15a* morphants were also rescued by co-KD of *p53* (**Supplemental Figure 4F, G)**. Collectively, these observations support our hypothesis that RPs control cardiac development, including heart size and function, *via* a p53 pathway-mediated regulation of CM proliferation.

### Heart loss in *Drosophila* by RpS15Aa KD is partially rescued by Hippo pathway activation

The fly heart develops similarly to the vertebrate heart at early stages [20] and a link between RPs and cell cycle-regulating pathways, such as p53 and Hippo pathways [50], has been proposed to be conserved between mammals and flies [51]. To test whether p53 has a function in fly heart development similar to hPSC-CMs and zebrafish, we performed cardiac co-KD of *RpS15Aa* and *p53* and examined heart structure and function. Co-KD did not rescue the cardiac defects of *RpS15Aa* KD suggesting that *p53* has a different role in flies, as compared to vertebrates, as has been reported previously [52].

In vertebrates, TP53 is known to act through the negative regulation of the Hippo pathway [53]. In this context, YAP, a downstream effector of Hippo, has been shown to promote CM proliferation in the mouse heart [54]. We therefore tested if YAP/yorkie can substitute the function of p53 in the fly heart and found that overexpressing the Hippo pathway gene *yorkie* (*yki*), the fly ortholog of YAP, in the developing heart considerably restored *RpS15Aa* KD-induced heart loss in *Drosophila* (**Figure 5A’,B**). We also observe formation of adult ostial structures, which indicates that the larval heart underwent partial remodeling to an adult heart during metamorphosis. Further, we found that *yki*-mediated rescue depends on the transcriptional co-factor encoded by *scalloped* (*sd*, *Drosophila TEAD1/2/3/4*), since upon *sd* KD, *yki* OE can no longer rescue the *RpS15Aa* KD-induced heart loss (**Figure 5A”, B**). Moreover, KD of *sd* in an *RpS15Aa* KD background (without *yki* OE) worsens the *RpS15Aa* KD phenotype (**Figure 5A”, B**).

**Figure 5:**
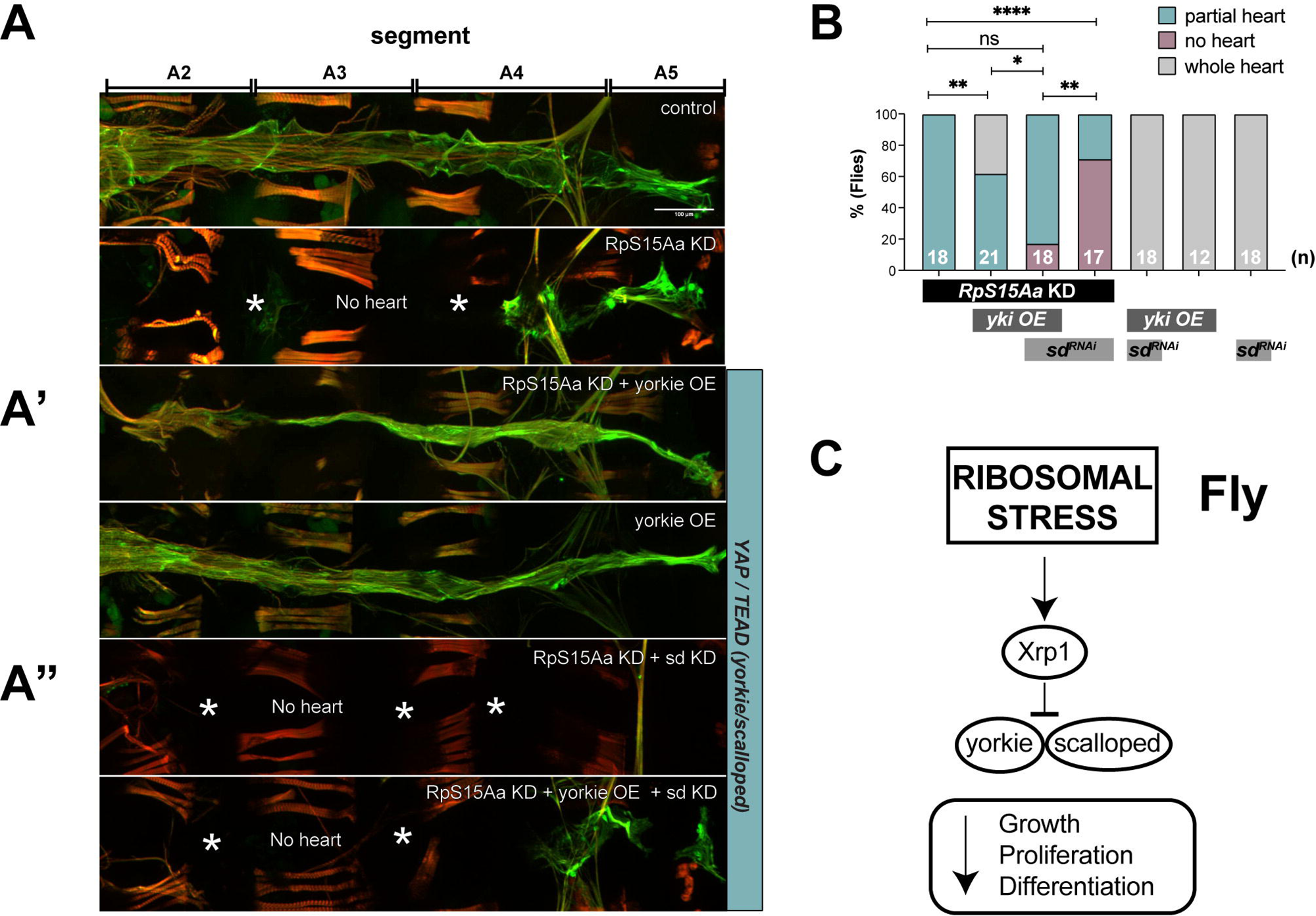
Rescue of *RpS15Aa* KD-mediated heart tube loss in Drosophila by YAP/yorkie overexpression depending on its co-factor TEAD/scalloped. (**A**) Representative images of RFP expressing fly hearts. RpS15Aa KD-mediated heart tube loss can be partially rescued by overexpression of *yorkie* (*RpS15Aa* RNAi + *yorkie* OE) . The rescue by *yki* OE depends on its co-factor *sd*. Flies were raised at 25 °C. **(B)** Quantification of events presented as a percentage of flies exhibiting whole heart tube versus partial heart loss (defined as 25-75% heart tube length compared to wildtype) or no heart tube. Statistics: Fisher’s exact test, *p<0.05. **(C)** Proposed signaling cascade underlying cardiac growth, proliferation and differentiation impairment following ribosomal stress (adapted from Baker et al., 2019).

### *RPS15A* genetically interacts with cardiac transcription factors *nkx2.7*/*tinman,* and TBX5/*tbx5a/Dorsocross*, *nkx2.7*/*tinman,* and *Gata4,5,6/pannier* in model systems

The profound importance of RPs for cell growth and proliferation in most cell types ([55] and **Supplemental Figure 1**), raises the question of how tissue specific phenotypes such as those observed in HLHS can arise from the defective function of ubiquitously expressed genes. To address this apparent paradox, we hypothesized that RPs might functionally interact with tissue-specific proteins such as cardiac transcription factors (cTFs), to control CM proliferation. Analysis of the impact of cTFs alone on hPSC-CM proliferation from our whole genome screen, revealed that KD of most cTFs did not cause major proliferation defects (except for e.g., *HES4* and *HOPX*; **Figure 6A**). Among the cTFs, TBX5 is specifically expressed in the left ventricle at stages of intense CM proliferation [56] and thus we next asked if *TBX5* could functionally interact with RPs. Remarkably, while hPSC-CM proliferation was not affected by increasing doses of siTBX5 (0-2nM), increasing si*TBX5* dosage in siRP backgrounds led to proportional decrease in EdU incorporation (Two-way ANOVA; **Figure 6B and Supplemental Figure 5A,B**), thereby indicating that *RPs* and *TBX5* genetically interact to regulate proliferation in hPSC-CMs.

**Figure 6:**
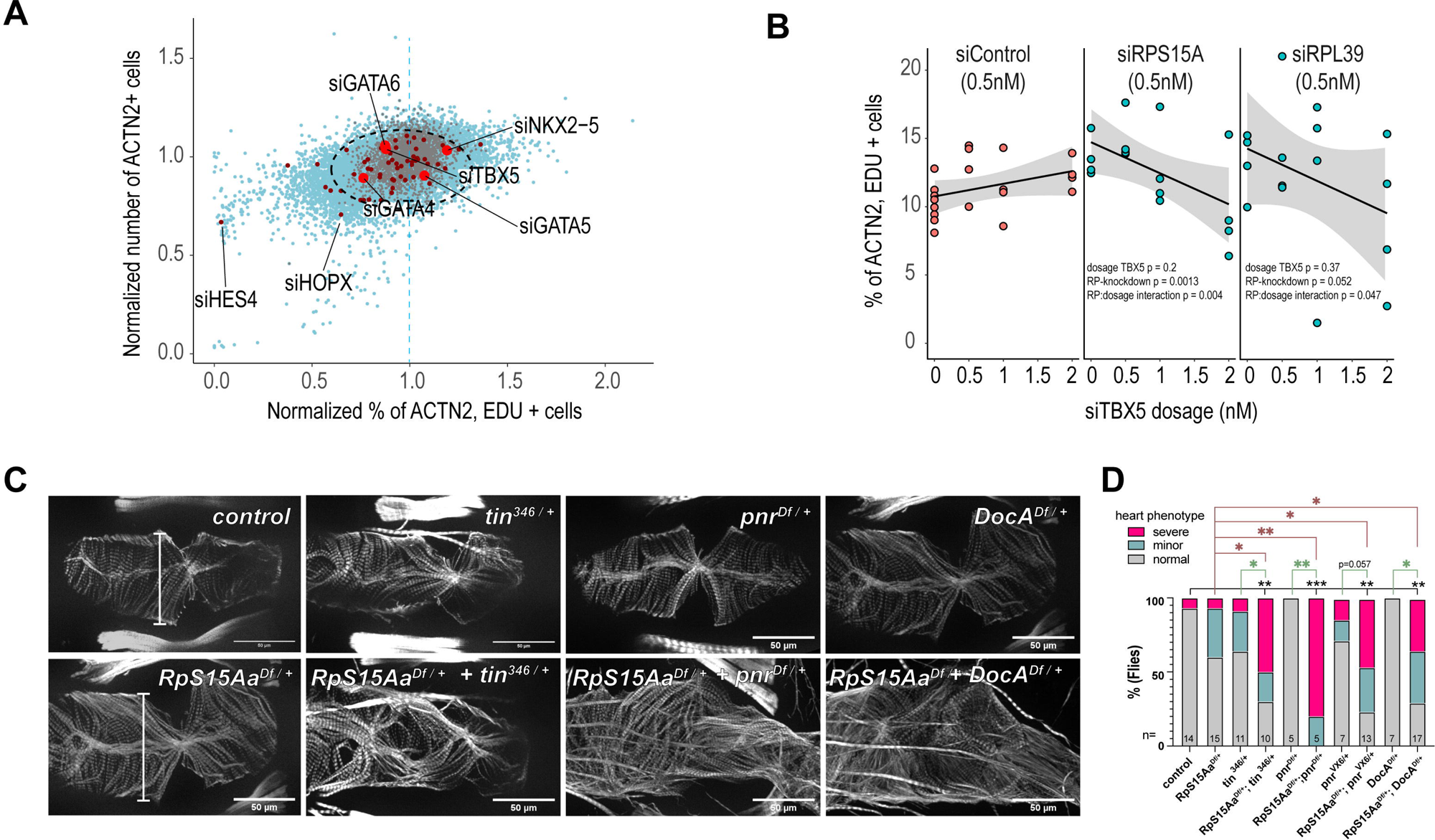
*RPS15A* genetically interacts with cardiac transcription factors. **(A)** The majority of cardiac transcription factors do not impact CM proliferation, except for e.g., HES4 and HOPX. **(B)** *TBX5* genetically interacts with *RPS15A* and *RPL39*. *siTBX5* does not impact CM proliferation at 0.5, 1, 1.5 or 2nM si-concentration, and neither do *siRPS15A* and *siRPL39* alone at 0.5nM. CM proliferation is reduced in siRP backgrounds with increased titration of *siTBX5*. Two-way ANOVA for si*TBX5* dosage, RP-knockdown and their interaction. **(C)** Representative fly heart segment (A4) from control flies, heterozygous mutants (*tin*^346/+^, *pnr*^VX6/+^, *Doc*^Df/-^, *Df*(*RpS15Aa*) ^+/-^) and transheterozygous mutants. *tin*^+/-^ = *tin^346^*/+, pnr^+/-^ = *Df(pnr)*/+ , Doc^+/-^ = *Df(DocA)*/+. Note the deformation and myofibrillar disorganization in the transheterozygous mutants. **(D)** Quantification of adult *Drosophila* heart defects and genetic interaction. - Statistics: Fisher’s exact test on absolute numbers testing *normal* vs. *severely deformed* hearts. *p<0.05, **p<0.005, ***p<0.001.

Conservation of genetic interactions between human and fly cardiac transcription factors and constitutive genes have been identified before [57]. We therefore specifically tested for genetic interaction between *RpS15Aa* and the cardiac TFs *tinman/NKX2-5, pannier/GATA4/5/6* and *Dorsocross/TBX5* in adult *Drosophila* hearts. A heterozygous deficiency covering *RpS15Aa* causes moderate dilation of hearts compared to controls (**Supplemental Figure 5C)**. However, when *RpS15Aa* was placed *in trans* to loss of function alleles of cardiac TFs *tinman, pannier* or *Dorsocross1/2/3*, these hearts exhibited significantly deformed hearts, which is only rarely observed in the single heterozygotes (Figure **5C****, D**). In addition, *RpS15Aa* heterozygous flies showed a prolonged heart period when combined with a *tinman* or *pannier* heterozygous mutations (**Supplemental Figure 5C**).

Next, we tested whether similar genetic interactions are conserved in zebrafish. Zebrafish expresses the NKX2-5 ortholog *nkx2.7* in the heart and functional studies have shown that both *nkx2.5* and *nkx2.7* play critical roles in cardiac development [58]. To determine whether *rps15a* and *nkx2.7 or tbx5a* genetically interact in zebrafish, we injected low doses of *rps15a* [45] MO, in combination with *nkx2.7* [59] or *tbx5a* MO. *nkx2.7* MO alone had little effect on heart size, cross-sectional area, or contractility, whereas double morphants exhibited cardiac dysfunction, with contraction virtually abolished in some animals (**Supplemental Figure 5D**). Furthermore, we observed a significant prolongation of the heart period in double morphants (*rps15a* + *nkx2.7* or *rps15a* + *tbx5a*) in comparison to each MO alone (Two-way ANOVA; **Supplemental Figure 5D, E**) consistent with the effects observed in *Drosophila*. Together, these results highlight the existence of evolutionarily conserved (fly, zebrafish and hPSC-CMs) genetic interactions between RPs and cardiac TFs genes in the regulation of cardiogenesis. These data also illustrate how the activity of ubiquitously expressed genes such as RPs, can be modulated by cell type-specific genes, like cardiac TFs, to achieve tissue-specific outcomes.

### RP-regulated HLHS-associated genes control proliferation in a CM-specific manner

Previous studies and this work show that reduced RP function can lead to tissue-specific developmental defects, that in turn can be rescued by loss of p53 [60, 61]. While these findings highlight a downstream role for p53 in the development of RP-dependent phenotypes, they also imply that RPs can regulate cell cycle and/or p53 activity in a tissue-specific manner. Thus, to delineate how RPs might control proliferation in a CM-specific manner, we compared differential gene expression profiles upon the two RP KDs in hPSC-CMs and undifferentiated hPSCs (**Figure 7A**). Consistent with our hypothesis, this analysis revealed that RPs regulate the expression of 493 genes in a CM-specific manner (**Supplemental Tables 5-8**), thereby suggesting the existence of a cell type-specific (cardiac) transcriptional response to RP loss of function.

**Figure 7:**
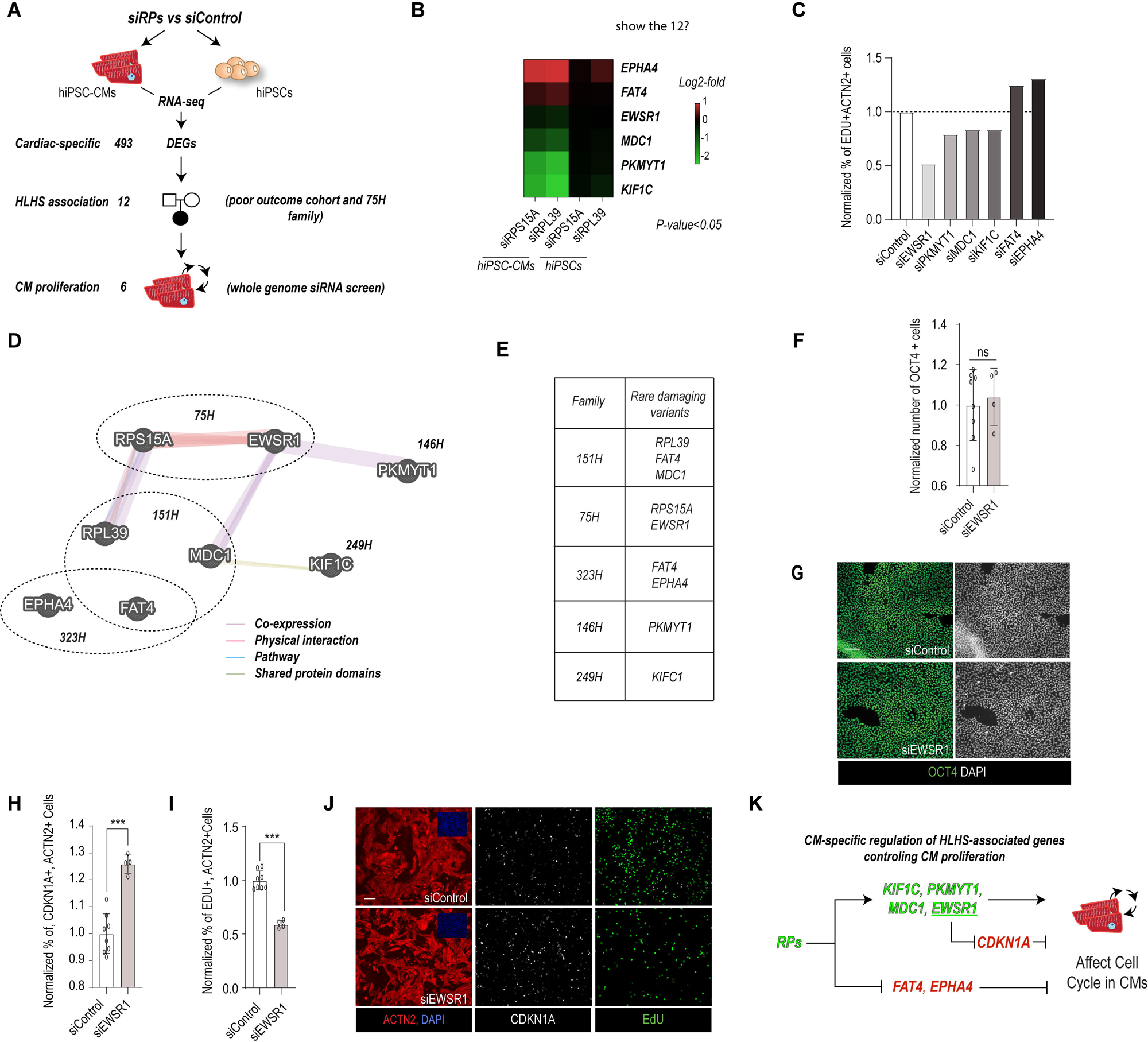
RP-dependent cardiac-specific regulation of cell proliferation. **(A)** Schematic illustrating approach to identify novel RP-dependent and cardiac-specific HLHS-associated gene network controlling CM proliferation. **(B)** Heatmap showing differential expression (hiPSC-CMs vs hiPSCs) of genes regulating CM proliferation. **(C)** Histogram showing effect of cardiac-specific and HLHS-associated genes on CM proliferation. **(D)** Visualization of RP-dependent and cardiac-specific HLHS-associated gene network (GeneMania). HLHS families. **(E)** Table of HLHS families harboring rare and damaging variant in RP-dependent and cardiac-specific genes. **(F)** Histogram showing lack of effect of siEWSR1 on OCT4+ hiPSCs. **(G)** Representative images of OCT4+ cells in siControl and siEWSR1. **(H)** Histogram showing that siEWSR1 increases the % of CDKN1A+ CMs as compared to siControl. **(I)** Histogram showing that siEWSR1 concomitantly decreases the % of EDU+ CMs as compared to siControl. **(J)** Representative images showing immunostaining for CDKN1A (white), EDU (green) and ACTN2 (Red) in siEWSR1 an siControl condition. **(K)** Pathway reconstruction of cardiac and RP-dependent regulation of CM proliferation by HLHS-associated genes. Chi-square test, *p<0.05, **p<0.005, ***p<0.001

Next, to evaluate whether HLHS-associated genes were part of this lineage-specific transcriptional response, we selected those differentially expressed genes harboring rare and predicted damaging variants in probands from the poor outcome cohort and 75H family (**Supplemental Tables 2, 4)**. Remarkably, this prioritization strategy identified 12 genes potentially associated with HLHS (**Supplemental Table 9**), 6 of which were found to also regulate cell cycle activity in hPSC-CMs (**Figure 7B, C**). Thus, collectively this approach led us to identify an RP-dependent and cardiac-specific HLHS-associated gene network that supports CM proliferation *via* the upregulation of *EWSR1, MDC1, PKMYT1* and *KIF1C,* and downregulation of *FAT4* and *EPHA4* (**Figure 7D, E**).

To further explore if RP-regulated HLHS-associated genes control proliferation in a cell type-specific manner, we asked if *EWSR1* KD, which elicited the strongest proliferation phenotype in hPSC-CMs, could regulate proliferation in non-cardiac cell types such hPSCs. In contrast to hPSC-CMs, siEWSR1 did not affect the number of OCT4+ cells (**Figure 7F, G**), which suggests that EWSR1 is able to regulate proliferation CM-specifically. EWSR1 is an RNA/DNA binding protein that regulates transcription and RNA splicing and plays a major role as oncogenic driver when fused to ETS transcription factors [62]. In a next step, we tested if EWSR1 regulates CM proliferation by modulating the p53 pathway. Consistent with this hypothesis, immunostaining for p53 downstream target, CDKN1A, revealed that EWSR1 KD both increased the percentage of CDKN1A-positive CMs and decreased EdU incorporation as compared to siControl (**Figure 7H-J**). Consistent with these observations, *RpS15Aa* deficiency fly line placed *in trans* to loss of function allele for *cabeza* (*caz*), the ortholog of *EWSR1,* caused prolongation of systolic interval lengths (**Supplemental Figure 5F)** as compared to controls, suggesting a genetic interaction between these two genes. Collectively, these results support the conclusion that RPs can modulate proliferation in a cell type-specific manner, by controlling the expression of downstream effectors (i.e. *EWSR1*), with a cell type-specific ability to modulate the p53 pathway activity (**Figure 7K and Figure 8**).

**Figure 8:**
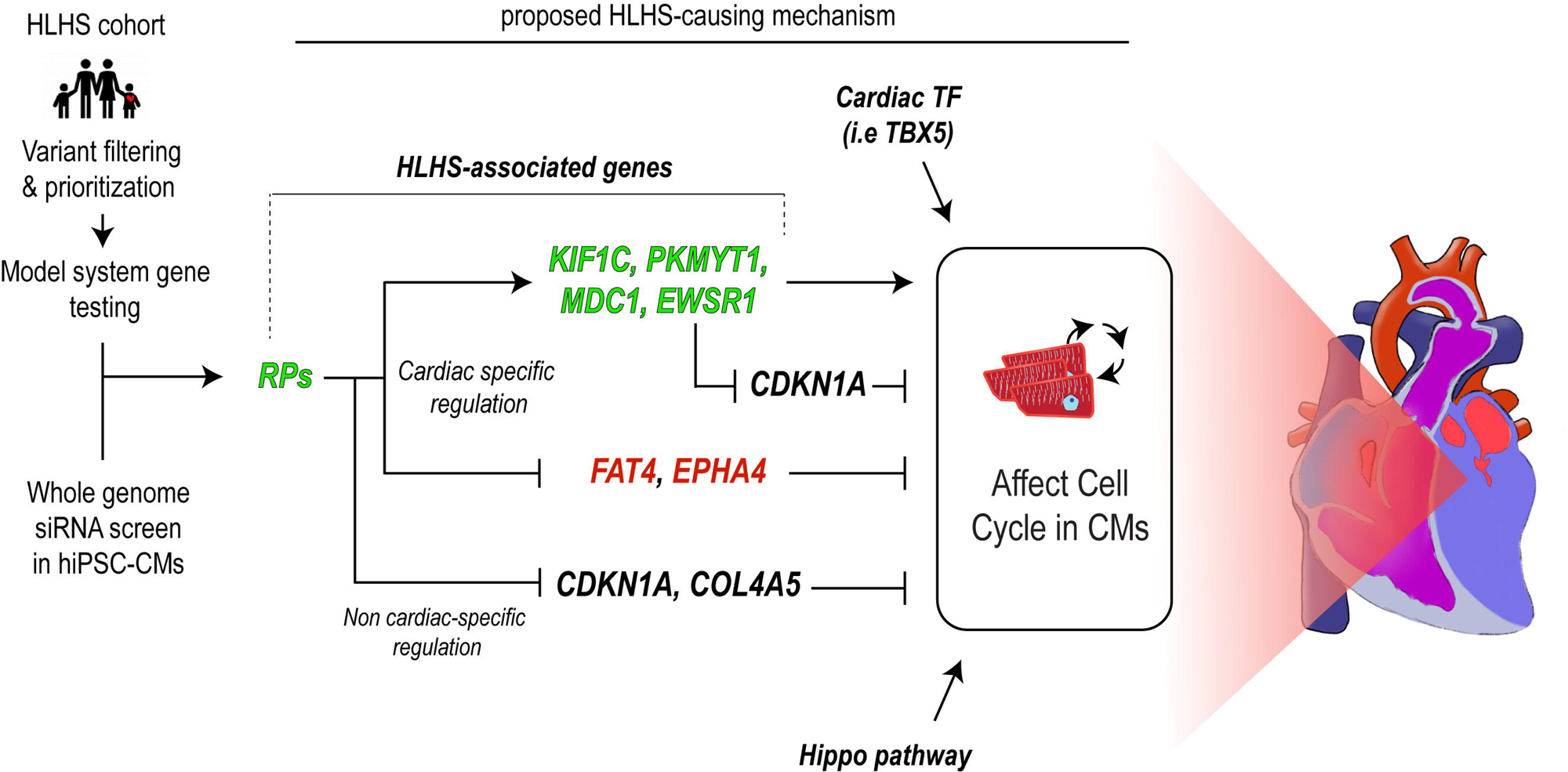
Model for RP-dependent involvement in HLHS-associated phenotypes. Schematic showing that combined prioritization and unbiased screening led to the identification of a novel HLHS-associated gene network regulating CM proliferation as potential disease-causing mechanism.

## Discussion

### Implication of ribosomal proteins as genetic modulators in HLHS pathogenesis

Recent work, including our own, strongly suggest that defective cardiac differentiation and impaired CM proliferation contribute to HLHS-associated heart defects [16–18]. In this study, we find that ribosomal proteins are critical for maintaining CM proliferation, as well as cardiac structure and function in fish and fly heart models. We show a potential role for RP function in cardiac development and HLHS pathogenesis as : (1) RPs are central regulators of cardiomyocyte proliferation, and (2) RP variants are enriched in a cohort of poor-outcome HLHS patients; (3) a rare predicted-damaging promoter variant in *RPS15A* was associated with HLHS in familial CHD (75H); (4) KD of RPs *in vivo* results in cardiac deficiencies in hPSC-CMs, *Drosophila* and zebrafish, and (5) induces TP53 signaling and blocks proliferation that can be alleviated by inhibition of TP53 in the vertebrate models. Interestingly, the transcriptomic response of a previously reported HLHS proband [18] paralleled these findings and raises the possibility of an underlying ribosomopathy condition in that HLHS family. While universally important for cellular growth, an increasing number of variants in RP genes have been linked to CHD in humans indicating tissue-specific differences in the penetrance of RP gene mutations. Most notably, ∼30% of patients with Diamond-Blackfan anemia [63], a ribosomopathy characterized by hypo-proliferative, proapoptotic erythropoiesis, also have CHD. In agreement with another study [46] *rps15a* KD in fish caused a noticeable reduction in circulating red blood cells.

In addition to RPS15A, *RpL13* was recently identified as a potential candidate gene involved in CHD from a screen for *de novo* copy number variations (CNVs) in 167 patients with CHD [43]. Furthermore, the Pediatric Cardiac Genomics Consortium (PCGC) exome dataset of 2871 patients with CHD identified several rare, predicted-damaging *de novo* variants in RP genes, 2 of which were found in patients with HLHS [64]. Our finding that RPs genetically interact with core cardiac transcription factors provides a potential mechanism for the cardiac specificity of RP phenotypes. In this context, we hypothesize that RP gene variants might play a role as genetic modifiers (or sensitizers) in the oligogenic pathogenesis of HLHS.

### Cardiac-specific and RP-dependent regulation of proliferation in the context of HLHS

Defective proliferation in left ventricular CMs is a phenotypic hallmark of HLHS [16, 17], thereby implying the existence of HLHS-associated mechanisms that regulate proliferation in a CM-specific specific-manner. In this study, we identified RPs as novel candidate genetic regulators of hypoplastic phenotypes observed in HLHS, as 1) rare, predicted damaging variants affecting RP genes are enriched in HLHS probands as compared to healthy controls, 2) KD of most RPs impairs proliferation in hiPSC-CMs (59/80 tested) and heart development in flies (6/6 tested), and 3) systemic loss of *rps15A* function causes heart-specific hypoplastic phenotypes in zebrafish. However, in this context, the evaluation of RPs function in multiple cell types revealed that they are generally required for proliferation (hPSCs, hPSC-CMs and dermal fibroblasts), thus, indicating that RP loss of function triggers a non-cell type specific cell cycle block.

Thus, an appealing mechanism by which RP function could be invoked in a spatially restricted manner would involve the regulation of RP expression by cell type-specific TFs. Consistent with this model, all probands harboring RP variants contain mutations affecting their promoter regions (**Table 1**). Also, several of these mutations are predicted to disrupt the binding site of previously identified HLHS-associated TF, ETS1 [15, 65]. Consistent with these observations, a recent study from the Bruneau lab [66] show that 38 RP genes are downregulated in TBX5-null hiPSC-CMs and ChIP analysis [67] reveals that a third of these dysregulated RPs (14/38 genes) are bound by TBX5 at their core promoter regions. Thus, we speculate that cell type specific TFs controlling RP expression represent a promising gene class enabling to modulate RP-dependent proliferation in a tissue-specific manner.

A second mechanism by which RPs might control proliferation and differentiation in a CM-specific manner, is based on the hypothesis that CMs may be molecularly more sensitive to RP loss-of-function than other cell types. Consistent with this model, RP deficiency predisposes flies, fish and hPSC-CM for interactions with core cardiogenic TFs, including TBX5/*Doc*, GATA4/*pnr* and NKX2-5/*tin*, the latter being the most cardiac restricted. (**Fig.** 6). Moreover, our comparative transcriptomics analysis (**Figure 4**) revealed that RPs regulate gene expression in a CM-specific manner. Among these RP regulated genes, *EWSR1, MDC1, PKMYT1, KIF1C, FAT4 and EPHA4*, were found to be mutated in HLHS families and regulate CM proliferation (**Figure 7**). Remarkably, EWSR1 KD did not affect proliferation in hPSCs, while it strongly reduced EdU incorporation in hPSC-CMs, thus suggesting that EWSR1 regulates proliferation in a CM-specific manner. Interestingly, in flies *RpS15Aa* genetically interacts with *cabeza* (*caz,* the fly ortholog of *EWSR1*). Consistent with our observations, a recent study has found that overexpression of a fusion Ewsr1-Fli1 protein in mice specifically induces dilated cardiomyopathy by promoting apoptosis in cardiac myocytes [68]. Similarly, Fat4, a negative regulator of the hippo pathway, which is upregulated upon RP KD, was found to specifically regulate heart size via the regulation of CM size and proliferation [69]. Thus, collectively we propose that RPs represent a central regulatory hub for cell proliferation and differentiation during embryonic development, which can be modulated in a cell type-specific manner by tissue-restricted modulators of RP expression and/or activity.

### An RP–MDM2–p53/YAP surveillance network in heart development

Defects in ribosome biogenesis caused, for example, by reduction in RP gene copy numbers induce a stress response program that activates TP53 (p53) and a cascade of cellular events resulting in apoptosis, inhibition of the cell cycle or DNA damage response (reviewed in e.g. [70]), commonly referred to as nucleolar stress response [71] with activation of TP53 being a key hallmark [72]. A key mechanism involves the release of RPs to the nucleoplasm, where several RPs bind and inhibit MDM2, which again results in p53 stabilization and pathway activation [73, 74]. A recent study showed that RP haploinsufficiency in the developing mouse limb bud also activates a common TP53 cascade but results in TP53-dependent tissue-specific changes of the translatome, which might confer the specificity often observed in ribosomopathy [60]. The TP53-MDM2 feedback loop is the central molecular node in response to a wide variety of stress signals, including in the human diseased adult heart [75, 76]. As mentioned above, the transcriptomic response of the HLHS family 5H [18] also showed activation of the TP53 pathway potentially due to an underlying ribosomopathy. Since low RP levels induce a proliferation blockade that can be overridden by p53-p21/CDKN1A KD, we hypothesize that RP levels may act as signaling components sensing cellular fitness and with a functional outcome either being p53 activation or non-activation. Such an RP–MDM2– p53 surveillance network was previously proposed to be important in response to nutrient availability changes and inhibition of oncogenic activity [77, 78]. Thus, the inhibition of the RP-MDM2-p53 axis might be a therapeutic avenue to consider for functional intervention, although further studies are needed, to identify the upstream mechanisms leading to the *p53* pathway activation observed in HLHS proband cells.

Heart-specific KD of the *RpS15Aa* in *Drosophila* causes constriction of larval hearts and atrophy in adult hearts, due to heart loss during metamorphosis, which could not be rescued by p53 reduction, unlike in hPSC-CMs and zebrafish. This might be due to a different, context-dependent role of *p53* in flies previously reported to cause a dwarfing, *Minute*-like phenotype [79]. We hypothesized that the loss of RPs in flies, in conjunction with reduced protein synthesis, might cause nucleolar stress, triggering a cell-intrinsic signaling cascade that prevents the heart to further differentiate and grow. To determine whether cardiac KD of *RpS15Aa* causes nucleolar stress in the *Drosophila* heart, we stained larval hearts for Fibrillarin, a marker for nucleoli and nucleolar integrity. We found that *RpS15Aa* KD causes expansion of nucleolar Fibrillarin staining in cardiomyocyte, which is a hallmark of nucleolar stress (**Suppl. Fig. 6A-C**). As a control, we also performed cardiac KD of *Nopp140*, which is known to cause nucleolar stress upon loss-of-function. We found a similar expansion of Fibrillarin staining in larval cardiomyocyte nuclei (**Suppl. Fig. 6C,D**). This suggests that *RpS15Aa* KD indeed causes nucleolar stress in the *Drosophila* heart, that likely contributes to the dramatic heart loss in adults.

The role of the Hippo pathway to regulate cell growth/organ size, proliferation, and survival, and its importance in cardiac biology is conserved between humans and fly [80–83]. The Hippo-YAP pathway is a cell-intrinsic pathway that regulates cardiomyocyte proliferation and thus heart size during development, as demonstrated by the ability of activated Yap to induce postnatal CM regeneration [84]. Interestingly, we find here that overexpression the *Drosophila* ortholog of YAP/TAZ, Yorkie, rescues *RpS15Aa* KD-induced heart loss, dependent on its downstream factor *scalloped* (the *Drosophila* ortholog of *TEAD1/2/3/4)* (Figure **5**). Of note, YAP1 is not only an important regulator of cardiomyocyte proliferation in the embryo but also promotes cardiomyocyte survival and growth in the postnatal heart [85, 86], which is in line with our findings. Interestingly, a proband among the PCGC HLHS patients was transheterozygous for mutations in *RPL15* and *TEAD4* [64], making this an interesting disease candidate pair.

### Rapid Gene Discovery and Prioritization Approach for Complex Genetic Diseases

Uncovering the genetic basis of polygenic and heterogeneous diseases, such as HLHS, remains a significant challenge. This difficulty arises from the lack of experimental approaches capable of rapidly determining the role and contribution of genetic variants, as well as their epistatic relationships, in generating disease-associated phenotypes. The current state-of-the-art approach involves using CRISPR-mediated editing or correction of gene variants [87], which is effective for analyzing single-family pedigrees. However, this method is impractical for prioritizing the large number of candidate genes identified through cohort-wide analyses. Consequently, progress in gene discovery for HLHS has been limited in recent years [12, 88]. In this study, we aimed to address these limitations by employing a HT and unbiased exploration of genes regulating a conserved HLHS-relevant phenotype [41, 89–91], such as reduced CM proliferation, in hiPSCs. Notably, results integration from a whole-genome functional screen in hiPSC-derived CMs (**Figure 1**, RPs are top hits from the screen), genomic data from a HLHS parent-proband cohort (**Figure 2**, RPs represent the most enriched gene class containing rare damaging variants), hiPSC-CM phenotyping from a high value HLHS family (75H), and functional validation in two independent *in vivo* model systems (**Figure 3**), identified RPs as a novel class of HLHS-associated genes. While our study highlights the potential of this approach for gene prioritization, additional research is needed to directly demonstrate the functional consequence of the identified genetic variants, to verify an association between RP encoding genes and HLHS in other patient cohorts with and without poor outcome, and determine if RP variants have a broader role in CHD susceptibility. In conclusion, we propose that the approach outlined in this study provides a novel framework for rapidly prioritizing candidate genes and systematically testing them, individually or in combination, using a CRISPR/Cas9 genome-editing strategy in mouse embryos [33]. Applied in the context of complex genetic diseases, this framework has the potential to yield deeper insights into their underlying mechanisms.

## Data Availability

All data produced in the present study are available upon reasonable request to the authors.

## Acknowledgements

This work was supported by a grant from the Wanek Foundation at Mayo Clinic in Rochester, M.N., to J.L.T., T.J.N., T.M.O., R.B. and A.R.C. We gratefully acknowledge the patients and families who participated in this study. We thank Marco Tamayo for excellent technical assistance. This work was supported by National Institutes of Health (R01 HL054732 to R.B.; R01 HL153645, R01 HL148827, R01 HL149992, R01 AG071464 to A.R.C.); by California Institute for Regenerative Medicine (DISC2-10110 to A.R.C.); by the American Heart Association (AHA Predoctoral Fellowship 18PRE33960593 to K.B.).

## MATERIALS AND METHODS

### Key resources table

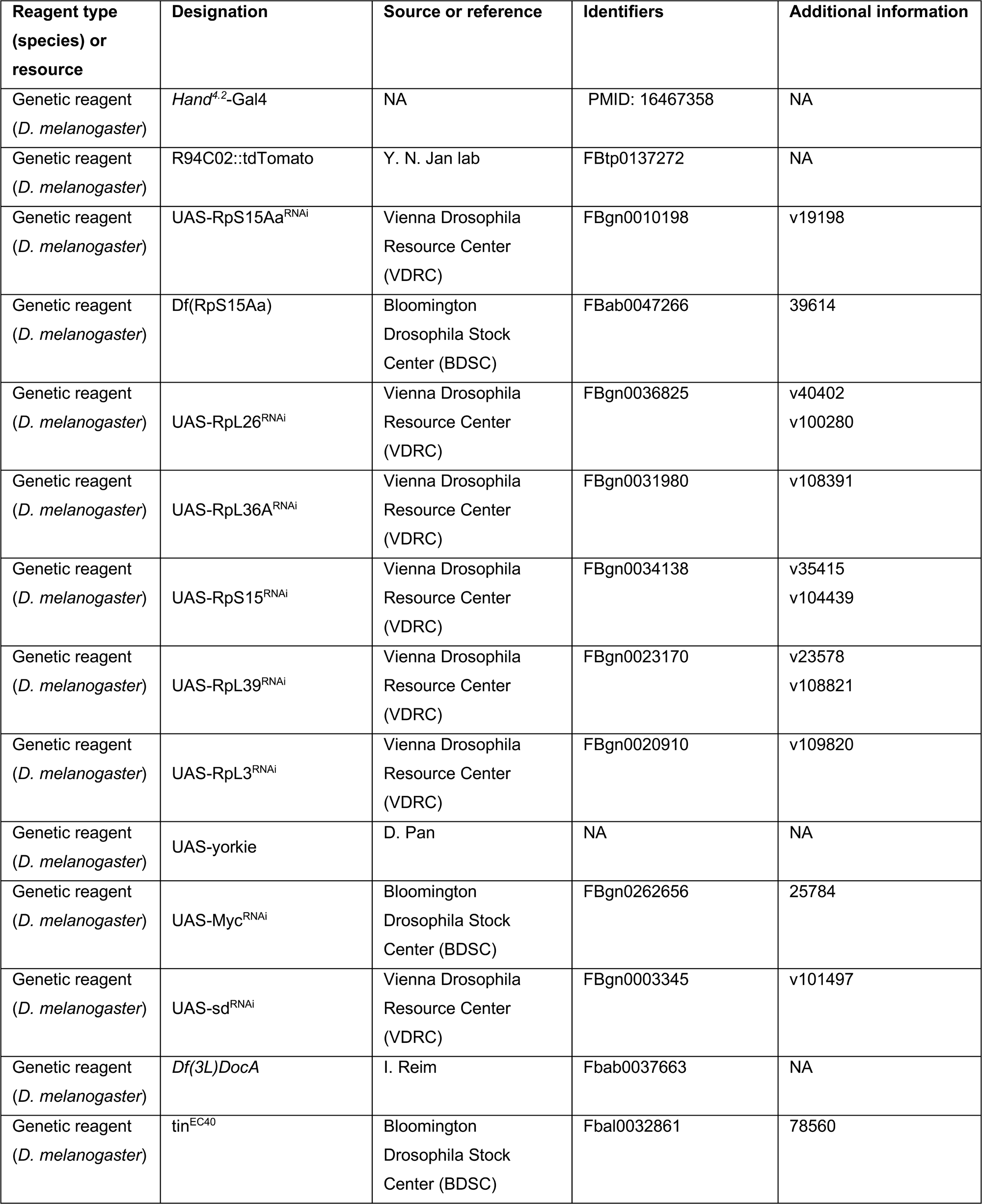
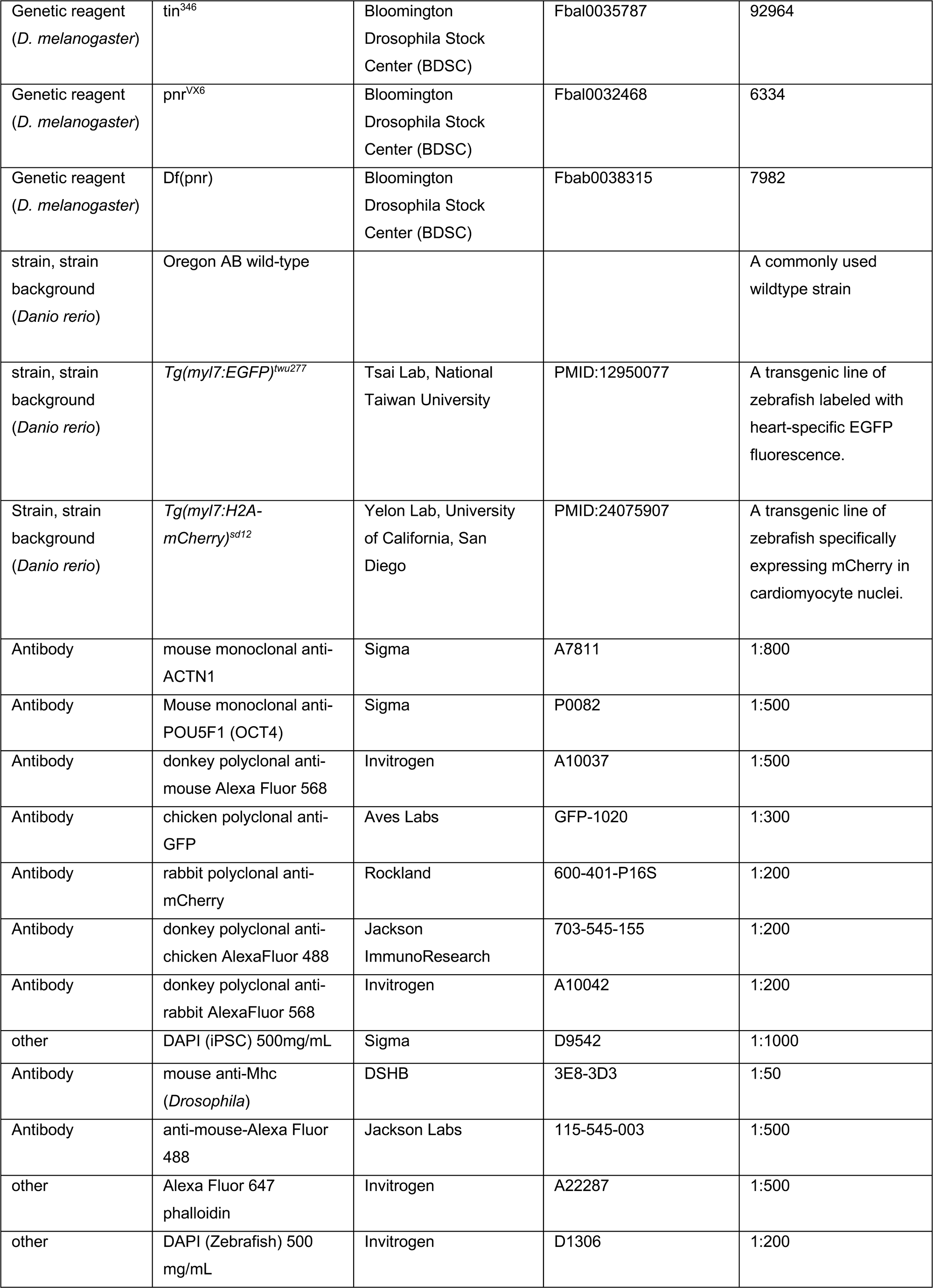
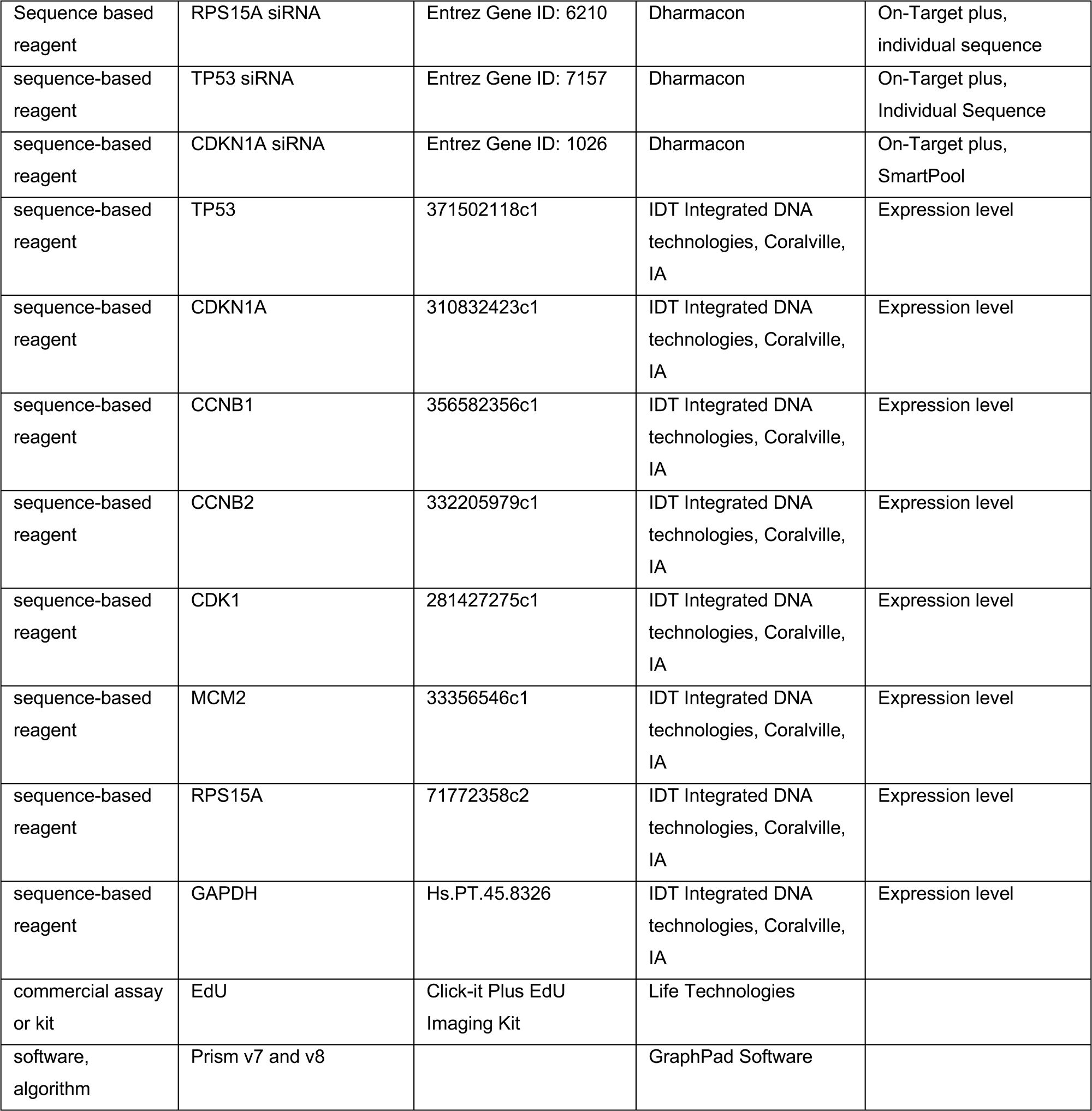

### Study subjects

Written informed consent was obtained for HLHS probands and family members under a research protocol approved by the Mayo Clinic Institutional Review Board. Cardiac anatomy was assessed by echocardiography. Candidate genes were identified and prioritized by WGS of genomic DNA and RNA sequencing of patient-specific iPSC and cardiomyocytes. Methods for genomic analyses, RNA Sequencing, iPSC technology, bioinformatics and statistics are described in the Online Appendix. Data are available at NCBI GEO database.

### WGS and bioinformatic strategies

Our methods for family phenotyping, WGS, variant filtering, and candidate gene prioritization in the rare HLHS-CHD family and 25 HLHS proband-parent trios have been previously described [7, 9, 18]. In brief, WGS of DNA isolated from blood or cheek swabs was performed on a HiSeq 4000 platform at the Mayo Clinic Medical Genome Facility. BAM files underwent primary and secondary analyses using an established workflow with standard data quality metrics, and reads were aligned to the human hg38 reference genome. Variant call format files with single nucleotide variants and small insertion-deletion (indel) calls were analyzed using Ingenuity Variant Analysis software and knowledge database (Qiagen). To retain only high-confidence data, variants were required to have a base call quality of at least 20 and to pass Variant Quality Score Recalibration. Functionally annotated coding and regulatory variants underwent primary filtering and prioritization by an iterative approach with variants required to have a minor allele frequency (MAF) <0.01 across all races in gnomAD v2.1.[93] Variants predicted to impact protein structure were retained and included missense, frameshift, stop-gain, stop-loss, canonical splice and non-canonical variants predicted to disrupt splicing based upon MaxEntScan[94]. Regulatory variants were defined as those that (a) impact a microRNA or microRNA binding site (b) reside in an enhancer binding site annotated in the VISTA database (http://enhancer.lbl.gov/) or (c) disrupt a predicted promoter or transcription factor binding site informed by the position-weighted matrices available in JASPAR (http://jaspar.cgb.ki.se/). Genes harboring a rare functional variant were further required to have upper quartile cardiac expression during mouse embryonic (e14.5) (Homsy et al., 2015) or human fetal heart development (ranked percentiles from RNASeq experiments available in ENCODE: ENCSR047LLJ (120 day male); ENCSR863BUL (91 day female); ENCSR000AEZ (28 week female and 19 week female) (Davis et al., 2018).

For the HLHS-CHD family, a secondary segregation filter was applied to model autosomal dominant inheritance with incomplete penetrance, which required sharing of prioritized rare variants between affected family members. For the poor-outcome HLHS proband-parent trios, a secondary Mendelian filter was applied to identify major-effect driver variants that arose *de novo* or fit recessive modes of inheritance (i.e., homozygosity, hemizygosity, compound heterozygosity).

To determine whether certain gene networks were over-represented, two online bioinformatics tools were used. First, STRING[39] was used to investigate experimental and predicted protein-protein and genetic interactions, and clustering of RP genes was demonstrated when the highest stringency filter was applied. In addition, PANTHER[38] was employed to identify potential enrichment of genes by ontology classification.

### Generation of hPSC-VCMs

Id1 overexpressing hPSCs [33, derived from, 95] were dissociated with 0.5 mM EDTA (ThermoFisher Scientific) in PBS without CaCl_2_ and MgCl_2_ (Corning) for 7 min at room temperature. hPSC were resuspended in mTeSR-1 media (StemCell Technologies) supplemented with 2 µM Thiazovivin (StemCell Technologies) and plated in a Matrigel-coated 12-well plate at a density of 3 x 10^5^ cells per well. After 24 hours after passage, cells were fed daily with mTeSR-1 media (without Thiazovivin) for 3-5 days until they reached ≥ 90% confluence to begin differentiation. hPSC-VCMs were differentiated as previously described [95]. At day 0, cells were treated with 6 µM CHIR99021 (Selleck Chemicals) in S12 media [96]. for 48 hours. At day 2, cells were treated with 2 µM Wnt-C59 (Selleck Chemicals), an inhibitor of WNT pathway, in S12 media. 48 hours later (at day 4), S12 media is fully changed. At day 5, cells were dissociated with TrypLE Express (Gibco) for 2 min and blocked with RPMI (Gibco) supplemented with 10% FBS (Omega Scientific). Cells were resuspended in S12 media supplemented with 4 mg/L Recombinant Human Insulin (Gibco) (S12+ media) and 2 µM Thiazovivin and plated onto a Matrigel-coated 12-well plate at a density of 9 x 10^5^ cells per well. S12+ media was changed at day 8 and replaced at day 10 with RPMI (Gibco) media supplemented with 213 µg/µL L-ascorbic acid (Sigma), 500 mg/L BSA-FV (Gibco), 0.5 mM L-carnitine (Sigma) and 8 g/L AlbuMAX Lipid-Rich BSA (Gibco; CM media). Typically, hPSC-ACMs start to beat around day 10. At day 15, cells were purified with lactate media (RPMI without glucose, 213 µg/µL L-ascorbic acid, 500 mg/L BSA-FV and 8 mM Sodium-DL-Lactate (Sigma), for 4 days. At day 19, media was replaced with CM media.

### siRNA transfection, proliferation assay and immunostaining in hPSC-VCMs

At day 25 of differentiation, hPSC-VCMs were dissociated with TrypLE Select 10X (Gibco), 10 min and neutralized with RPMI supplemented with 10% FBS. Cells were resuspended in RPMI with 2% KOSR (Gibco) and 2% B27 50X with vitamin A (Life Technologies) supplemented with 2 μM Thiazovivin and plated at a density of 5,000 cells/well in a Matrigel-coated 384-well plate. hPSC-VCMs were transfected with siRNA (Dharmacon: ON-TARGETplus, custom RNAi cherry-pick libraries 0.1 nmol). For the whole genome screening, siRNAs directed to 18,000 human genes were purchased from the Genomic Center Facility at SBP at a final concentration of 25 nM using lipofectamine RNAiMax (ThermoFisher) and opti-MEM (Gibco). 48 hours post-transfection, cells were labelled with 10 μM EdU (ThermoFisher). After 24h of EdU incubation, cells were fixed with 4% paraformaldehyde for 30 min and blocked in blocking buffer (10% Horse Serum, 10% Gelatin and 0.5% Triton X-100) for 20 min. EdU was detected according to protocol and cells were stained with cardiac specific marker ACTN2 (A7811, Sigma, dilution 1/800), secondary antibody Alexa Fluor 568 (Invitrogen, 1/500) and DAPI (1/1000) in Blocking Buffer. Cells were imaged with ImageXpress Micro XLS microscope (Molecular Devices) and custom algorithms were used to quantify % of EdU+ ACTN1+ hPSC-VCMs and the number of ACTN1+ cells. To quantify TP53 levels in hPSC-VCMs, cells were stained with Phospho-p53 (700439, ThermoFisher, 1/500), counterstained with ACTN1 (anti-mouse A7811, Sigma, or anti-rat ab50599, Abcam), and imaged and quantified with ImageXpress microscope. Whole genome screening was performed in one-replicate. All the other siRNAs experiments were performed in quadruplicates.

### RNA-seq and data analysis

hPSCs and hPSC-derived CMs were transfected with 25nM final concentrations of siRNA against RPS15A, RPL39 and with scrambled control siRNAs as above. 2 days after siRNA transfection RPs KD were verified and confirmed by proliferation assay (see above), and cells were pelleted and resuspended in 500µl TRIzol reagent followed by total RNA extraction. Library preparation and sequencing of the samples was done at La Jolla Institute of Immunology (La Jolla, CA). FASTQ files were processed using nf-core/rnaseq (version 21.03.0.edge [97]). Differential gene expression was determined using R/DESeq2 [98] and GO term enrichment was done using gprofiler2[99]. Analysis scripts can be downloaded at github.com/gvogler. Raw reads and counts tables are available at GEO accession number GSE207658.

### Proliferation assay in parent/proband iPSCs

hPSCs were dissociated as described previously and plated in a Matrigel-coated 384-well plate at a density of 3,000 cells per well. Cells were transfected with siRNA at a final concentration of 25nM using lipofectamine RNAiMax and opti-MEM. After 3 days, cells were labelled with 10 μM EdU for 1 hour. Cells were fixed with 4% paraformaldehyde for 30 min and blocked in blocking buffer for 20 min. EdU was detected according to the protocol and stained with stem cell specific marker OCT4 (P0082, Sigma, 1/500), secondary antibody Alexa Fluor 568 and DAPI in Blocking Buffer. Cells were imaged with ImageXpress microscope and % of EdU+ cells and number of OCT4+ cells were quantified.

### Quantitative Real Time PCR (RT-qPCR) in parent/proband iPSCs and hPSC-CMs

Total RNA was extracted using TRIzol and chloroform. 1ug of RNA was converted in cDNA using QuantiTect Reverse Transcription kit (Qiagen). qRT-PCR was performed using SYBR green (Biorad). Human primers sequences for qRT-PCR were obtained from Harvard Primer Bank: TP53 (Primer Bank ID: 371502118c1), CDKN1A (Primer Bank ID: 310832423c1), CCNB1 (Primer Bank ID: 356582356c1), CCNB2 (Primer Bank ID: 332205979c1), CDK1 (Primer Bank ID: 281427275c1), MCM2 (Primer Bank ID: 33356546c1), RPS15A (Primer Bank ID: 71772358c2). All values were normalized to *GAPDH (Primer Bank ID: 378404907c1)*. At least 3 independent biological replicates were performed for each experiment.

### Statistical analysis

To determine any statistical significance between experimental and control groups in hPSC-VCMs and iPSCs from parent/proband trio, we calculated two-sided p values with Student’s t-test using GraphPad Prism 8.1.2 software.

### *Drosophila* heart function studies

*Drosophila* orthologs were determined using the DIOPT database [100], and fly stocks were obtained from the Vienna Drosophila Resource Center (VDRC) stock center or Bloomington Drosophila Stock Center (BDSC) as indicated in the key source table. For *in vivo* functional heart analysis, we developed a high-throughput method based on genetically modified flies with cardiomyocyte-specific RFP fluorescence (Klassen et al.). The reporter line by itself or combined with the heart-specific *Hand^4.2^*-Gal4 driver (including all cardioblasts/cardiomyocytes, pericardial cells and wing hearts throughout development starting at post-mitotic, mid-embryonic stages through adulthood [101–103] was crossed to UAS-lines or mutant flies. For interaction studies a fly line harboring the fluorescent reporter, *Hand^4.2^*-Gal4 driver and a Deficiency for RpS15Aa was generated. Adult progeny flies were immobilized and exposed to fluorescence light to record 5 sec high frame rate movies of the beating heart. Movies were analyzed by fully automated quantification of contractility and rhythmicity parameters of the heart [104]. Semi-intact adult fly hearts were filmed and analyzed according to standard protocol [27, 105].

### Immunostainings of the fly heart

The immunostaining of fly adult hearts was performed as described previously [106]. In short, adult flies were dissected in a semi-intact fashion to expose the heart according to protocol [27, 107]. Myofibrils were relaxed using 10mM EGTA followed by fixation in 4% formaldehyde for 15 min. Hearts were washed with PBS + Triton (PBT; 0.03 % triton X-100) and stained using Alexa 647 phalloidin (Life Technologies) 2h at room temperature (RT). For Mhc labelling, hearts were stained using anti-Mhc antibody (1:50, incubation over night at 4°C) and after three washes with PBT, secondary antibody was applied for 2 h at RT. Hearts were washed with PBT three times and PBS one time and mounted using ProLong Gold mountant with DAPI (Life Technologies). Heart preparations were imaged with the Imager.Z1 with an Apotome (Carl Zeiss), Hamamatsu Orca Flash 4.0 and ZEN imaging software (Carl Zeiss).

### Zebrafish Husbandry

All zebrafish experiments were performed in accordance with protocols approved by IACUC. Zebrafish were maintained under standard laboratory conditions at 28.5°C. In addition to Oregon AB wild-type, the following transgenic lines were used: *Tg(myl7:EGFP)^twu277^* [108] and *Tg(myl7:H2A-mCherry)^sd12^* [109].

In zebrafish, gene expression was manipulated using standard microinjection of morpholino (MO) antisense oligonucleotides [110]. In addition, we performed targeted mutagenesis using CRISPR/Cas9 genome editing[111–113], to create insertion/deletion (INDEL) mutations in relevant ribosomal genes (F0). Subsequently, zebrafish were raised to 72 hours post fertilization (hpf), immobilized in low melt agarose and the hearts were filmed and analyzed according to standard protocol[27].

### MO sequence information

All MOs were purchased from Gene Tools, LLC, synthesized at 300nmol, except zebrafish p53 oligos at 100nmol.

Zebrafish *rpl13* 5’-UTR MO: TTGTTCACTCCGTCCTTAGCGGAAA

Zebrafish *rps15a* 5’-UTR MO: CGCACCATGATGCCAGTTCTGCAAT

Zebrafish *rpl39* 5’-UTR MO: GGATCGCAATCCGTTCACCACTATG

Zebrafish p53 MO: GCGCCATTGCTTTGCAAGAATTG

Zebrafish Control MO: 5’-CCT CTT ACC TCA GTT ACA ATT TAT A-3’.

### Zebrafish SOHA (Semi-automated Optical Heartbeat Analysis)

Larval zebrafish (72 hpf) were immobilized in a small amount of low melt agarose (1.5%) and submerged in conditioned water. Beating hearts were imaged with direct immersion optics and a digital high-speed camera (up to 200 frame/sec, Hamamatsu Orca Flash) to record 30 seconds movies; images were captured using HC Image (Hamamatsu Corp.). Cardiac function was analyzed from these high-speed movies using semi-automatic optical heartbeat analysis software[27, 114], which for zebrafish quantifies heart period (R-R interval), cardiac rhythmicity, as well as chamber size and fractional area change. All hearts were imaged at room temperature (20-21°C). Statistical analyses were performed using Prism software (Graphpad). Significance was determined using two-tailed, unpaired student t-test or one-way ANOVA and Dunnett’s multiple comparisons post hoc test as appropriate.

### Zebrafish Cardiomyocyte Cell Counts and Cardiac immunofluorescent imaging

To count cardiomyocytes, we used the expression of H2AmCherry in the nuclei (*Tg(myl7:H2A-mCherry*)[109] to qualify as an individual cell, performed the “Spot” function in Imaris to distinguish individual cells in reconstructions of confocal z-stacks.[115, 116] To compare data sets, we used Prism software (GraphPad) to perform Student’s t-test with two-tail distribution. Graphs display mean and standard deviation for each data set.

Whole-mount immunofluorescence was performed as previously described[18, 115–117] (see also key resources table). Confocal imaging was performed on an LSM 710 confocal microscope (Zeiss, Germany) with a 40x water objective. Exported z-stacks were processed with Imaris software (Bitplane), Zeiss Zen, and Adobe Creative Suite software (Photoshop and Illustrator 2020). All confocal images shown are projection views of partial reconstructions from multiple z-stack slices, except where noted that images are views of a single slice.

For proliferation assays, embryos at 24, 48, and 72 hpf were fixed in 4% paraformaldehyde overnight at 4 °C, washed in PBS with 0.1% Tween-20 (PBST). Samples were blocked in 5% normal goat serum in PBST prior to incubation with primary antibodies. Proliferating cells were labeled with anti-phospho-histone H3 (Sigma Aldrich-H0412 PH3; 1:500) and total nuclei were counterstained with DAPI. Embryos were mounted in ProLong Gold for imaging. Whole-mount hearts were imaged on a Zeiss Apotome microscope at 10× magnification. Z-stacks were acquired and reconstructed using Fiji/ImageJ. Regions corresponding to atrium, ventricle, atrioventricular canal, and outflow tract were manually segmented. For some experiments, cardiomyocyte nuclei were identified based on Tg(myl7:H2A-mCherry) expression, with DAPI used to quantify total cell counts. For proliferation experiments DAPI was used to identify cardiac cells and proliferating cells were identified by PH3 staining. Proliferative indices were calculated as PH3+/DAPI+ ratios from z-stacks analyzed in Fiji/ImageJ using the Cell Counter plugin. Cell counts were performed on at least 10 embryos per condition per timepoint.

### Zebrafish CRISPR/Cas9 experiments

Detailed steps were previously described[118] and we followed IDT manufacture instruction for Complexes preparation. <u>crRNA:tracrRNA Duplex Preparation</u>: Target-specific Alt-R® crRNA ( sequence information see below) and common Alt-R® tracrRNA were synthesized by IDT and each RNA was dissolved in duplex buffer (IDT) as 100 μM stock solution. Stock solutions were stored at −20°C. To prepare the crRNA:tracrRNA duplex, equal volumes of 100 μM Alt-R® crRNA and 100μM Alt-R® tracrRNA stock solutions were mixed together and annealed by heating followed by gradual cooling to room temperature by manufacture instruction: 95°C, 5 min on PCR machine; cool to 25°C; cool to 4°C rapidly on ice. The 50 μM crRNA:tracrRNA duplex stock solution was stored at −20°C. <u>Preparation of crRNA:tracrRNA:Cas9 RNP Complexes:</u> Cas9 protein (Alt-R® S.p. Cas9 nuclease, v.3, IDT) was adjusted to 25 μM stock solution in 20mM HEPES-NaOH (pH 7.5), 350 mM KCl, 20% glycerol, dispensed as 8 ul aliquots, and stored at −80°C. 25μM crRNA:tracrRNA duplex was produced by mixing equal volumes of 50 μM crRNA:tracrRNA duplex stock and duplex buffer (IDT). We used 5 μM RNP complex. To generate 5μM crRNA:tracrRNA:Cas9 RNP complexes: 1 μl 25 μM crRNA:tracrRNA duplex was mixed with 1 μl 25 μM Cas9 stock, 2 μl H2O, and 1 μl 0.25% phenol red solution. Prior to microinjection, the RNP complex solution was incubated at 37°C, 5 min and then placed at room temperature. Approximately one nanoliter of 5 μM RNP complex was injected into the cytoplasm of one-cell stage embryos to generate F0 larva.

### IDT crRNA sequence information

All crRNAs were purchased from Integrated DNA Technologies, Inc., synthesized at 2 nmol with standard desalting condition.

Dr.Cas9.RPS15A.1.AC: /AltR1/rUrU rGrUrU rGrUrC rArArU rCrUrC rArCrA rGrGrG rGrUrU rUrUrA rGrArG rCrUrA rUrGrC rU/AltR2/

Dr.Cas9.RPS15A.1.AB: /AltR1/rGrC rGrUrA rCrUrA rUrGrA rCrUrU rUrArG rArGrC rGrUrU rUrUrA rGrArG rCrUrA rUrGrC rU/AltR2/

Dr.Cas9.RPS17.1.AC: /AlTR1/rUrGrArCrUrUrCrCrArCrArUrUrArArCrArArGrCrGrUrUrUrUrArGrArGrCrUrArUrG rCrU/AlTR2/

Dr.Cas9.RPS28.1.AA: /AlTR1/rCrUrGrGrGrArArGrArArCrUrGrGrCrUrCrCrCrArGrUrUrUrUrArGrArGrCrUrArUr GrCrU/AlTR2/

---

**Supplemental Figure 1: (A, B)** Confirmatory siRNA screening for all RP genes confirms critical role for RPs in CM proliferation. (**C, D**) KD of RPs reduces proliferation in human fibroblasts.

**Supplemental Figure 2: Model organism phenotypes caused by loss of RP genes. (A)** Pedigree of the extended 75H family investigated for shared variants between HLHS proband (black square) and distant cousin with CHD (half-filled circle). (**B**) **(C)** Diastolic and **(D)** and Systolic Surface Area after rps15a CRISPR. Statistics: Mann-Whitney test, *p<0.05, **p<0.01, ***p<0.001, ****p<0.0001

**Supplemental Figure 3: *rps15a* zebrafish larval morphants show heart dysfunction and reduction in cardiomyocyte number. (A)** Lateral view (head to the right) of a wild-type control zebrafish larva (top) and a morphant (bottom) following injection of 2 ng/µl *rps15a* morpholino (MO). The morphant exhibits near normal body/tail but notable pericardial edema (arrowhead). **(B)** Control (left) and morphant hearts (right) stained with Tg(*myl7*:*EGFP*) and Tg(*myl7*:*H2A*-*mCherry*). Note the smaller heart with aberrant looping in morphant heart. **(C)** Total CM counts (mCherry+ cells) in control and *rps15a* morphants shown in (B). **(D)** Fractional area change (FAC) in control and *rps15a* morphants. **(E-H)** Injection of Control MO had little effect on zebrafish cardiac function. There was no significant difference between uninjected and injected controls with respect to (**E**) Systolic or (**F**) Diastolic Surface Areas, (**G**) Fractional Area Change or (**H**) R-R intervals. Student’s t-test, *p<0.05, **p<0.01, ***p<0.001, ****p<0.0001.

**Supplemental Figure 4: Proliferation and heart function depends on RPS15A and is regulated via TP53 in CMs and zebrafish. (A-C)** Cardiac proliferation (by PH3, **A**), total CM number (DAPI, **B**) and ratio of proliferating CMs (C) assessed for *rps15a* morphants (KD) and CRIPSR (KO) for three timepoints, 24/48/72hpf **(D, E)** The number of *siRPS15A-*treated CMs is strongly reduced, which can be attenuated by co-KD of *CDKN1A* or *TP53*. **(F,G)** Loss of *rps15a* causes strong reduction in zebrafish heart contractility (measured by Fractional Area Change, **C**) and heart period **(D)**, which can be rescued by co-KD of *p53* in both, zebrafish heart atrium and ventricle.

**Supplemental Figure 5: Genetic interaction between *RpS15Aa/RPS15A* and *cabeza/EWSR1* (A, B)** Dose-dependent inhibition of hPSC-CMs proliferation by RPS15A KD. **(C)** Diastolic Diameter (DD), and Heart Period (HP) of 3-week-old female control and heterozygous RpS15Aa deficient flies with or without additional heterozygous mutation in tinman, pannier or Dorsocross. Statistics: Kruskal-Wallis, *p<0.05, **p<0.005, ***p<0.001, ****p<0.0001. Note, the prolonged HP in transheterozygous mutants for Df(RpS15Aa) and tinman. **(D, E)** FAC, and HP of the atria and ventricles at 72 hpf in zebrafish embryos injected with 0.5 ng rps15a and/or 1 ng nkx2.7 MOs (**E**) or 0.5 ng rps15a and/or 0.5 ng tbx5 MOs (F). Note, that FAC and heart periods show synergistic genetic interaction between rps15a and nkx2.7 and tbx5a. Statistics: 2-Way ANOVA, *p<0.05, **p<0.01, ****p<0.0001.**(F)** Systolic intervals (SI) of 1-week-old adult flies upon heart-specific knockdown of *cabeza* (*caz^32990^* or *caz^34839^* RNAi) using Hand^4.2^-Gal4 driver with or without additional RpS15Aa heterozygosity (RpS15Aa^Df/+^) and age-matched controls. Automated analysis of SI depicted as median with interquartile range obtained from 5s high-frame-rate videos. Statistics: Kruskal-Wallis test, *p<0.05, **p<0.01, ***p<0.001.

**Suppl. Fig. 6: RpS15Aa KD induces nucleolar stress in 3^rd^ instar larval hearts. (A)** Nucleolar stress induction in 3^rd^ instar larval hearts upon *RpS15Aa* KD as indicated by enlarged nucleoli. Hearts from control and *RpS15Aa* KD. 3^rd^ instar larva stained for F-Actin (red), DAPI (blue), and nucleolus marker Fibrillarin (green) and cardiomyocyte (CM) nuclei in magnifications. Note, heart constriction upon *RpS15Aa* KD in the larval heart.

**(A)** Quantification of nucleolus area and area ratio(nucleolus/nucleus) indicates significantly enlarged nucleoli upon *RpS15Aa* KD. Statistics: Mann-Whitney test, ****p<0001. **(C)** Schematic representation of different nucleolar phenotypes upon RNAi-mediated loss of function. **(D)** As a positive control, KD of another nucleolar key player *Nopp140* specifically in the heart using Hand^4.2^-Gal4 combined with Fibrillarin^EGFP^ (kind gift of the Wieschaus lab) leads to loss of nucleolar maintenance. 3^rd^ instar larva stained for F-Actin (red) and DAPI (blue). Fibrillarin in (green). Cardiomyocyte nuclei in magnifications. PC= Pericardial Cell

**Supplemental Table 1: Whole genome siRNA screen results table.**

**Supplemental Table 2: 292 filtered variants from poor-outcome HLHS probands.**

MOI – mode of inheritance

**Supplemental Table 3: Subset from all 292 genes that cause CM proliferation defects and/or fly heart defects**

**Supplemental Table 4: Segregating variants of 75H proband.**

**Supplemental Tables 5-8: Differential gene expression lists for siRPL39 and siRPS15A in hPSCs and CMs.**

**Supplemental Table 9: 12 genes prioritized after RNAseq and WGS analysis.**

## REFERENCES

1. Gordon BM, Rodriguez S, Lee M, Chang R-K. Decreasing number of deaths of infants with hypoplastic left heart syndrome. J Pediatr. 2008;153(3):354–8. doi: 10.1016/j.jpeds.2008.03.009.

2. Reller MD, Strickland MJ, Riehle-Colarusso T, Mahle WT, Correa A. Prevalence of congenital heart defects in metropolitan Atlanta, 1998-2005. J Pediatr. 2008;153(6):807–13. doi: 10.1016/j.jpeds.2008.05.059.

3. Crucean A, Alqahtani A, Barron DJ, Brawn WJ, Richardson RV, O’Sullivan J, et al. Re-evaluation of hypoplastic left heart syndrome from a developmental and morphological perspective. Orphanet J Rare Dis. 2017;12(1):138. doi: 10.1186/s13023-017-0683-4.

4. Konno Y, Toki T, Tandai S, Xu G, Wang R, Terui K, et al. Mutations in the ribosomal protein genes in Japanese patients with Diamond-Blackfan anemia. Haematologica. 2010;95(8):1293–9. doi: 10.3324/haematol.2009.020826.

5. Agopian AJ, Goldmuntz E, Hakonarson H, Sewda A, Taylor D, Mitchell LE, et al. Genome-Wide Association Studies and Meta-Analyses for Congenital Heart Defects. Circ Cardiovasc Genet. 2017;10(3):e001449. doi: 10.1161/CIRCGENETICS.116.001449.

6. Martin LJ, Pilipenko V, Benson DW. Role of Segregation for Variant Discovery in Multiplex Families Ascertained by Probands With Left Sided Cardiovascular Malformations. Front Genet. 2018;9:729. doi: 10.3389/fgene.2018.00729.

7. Theis JL, Hrstka SCL, Evans JM, O’Byrne MM, de Andrade M, O’Leary PW, et al. Compound heterozygous NOTCH1 mutations underlie impaired cardiogenesis in a patient with hypoplastic left heart syndrome. Hum Genet. 2015;134(9):1003–11. doi: 10.1007/s00439-015-1582-1.

8. Elliott DA, Kirk EP, Yeoh T, Chandar S, McKenzie F, Taylor P, et al. Cardiac homeobox gene NKX2-5 mutations and congenital heart disease: associations with atrial septal defect and hypoplastic left heart syndrome. J Am Coll Cardiol. 2003;41(11):2072–6. doi: 10.1016/s0735-1097(03)00420-0.

9. Theis JL, Zimmermann MT, Evans JM, Eckloff BW, Wieben ED, Qureshi MY, et al. Recessive MYH6 Mutations in Hypoplastic Left Heart With Reduced Ejection Fraction. Circ Cardiovasc Genet. 2015;8(4):564–71. doi: 10.1161/CIRCGENETICS.115.001070.

10. Zanon A, Kalvakuri S, Rakovic A, Foco L, Guida M, Schwienbacher C, et al. SLP-2 interacts with Parkin in mitochondria and prevents mitochondrial dysfunction in Parkin-deficient human iPSC-derived neurons and Drosophila. Hum Mol Genet. 2017;26(13):2412–25. doi: 10.1093/hmg/ddx132. PubMed PMID: 28379402.

11. Dasgupta C, Martinez AM, Zuppan CW, Shah MM, Bailey LL, Fletcher WH. Identification of connexin43 (alpha1) gap junction gene mutations in patients with hypoplastic left heart syndrome by denaturing gradient gel electrophoresis (DGGE). Mutat Res. 2001;479(1-2):173–86. doi: 10.1016/s0027-5107(01)00160-9.

12. Yagi H, Liu X, Gabriel GC, Wu Y, Peterson K, Murray SA, et al. The Genetic Landscape of Hypoplastic Left Heart Syndrome. Pediatr Cardiol. 2018;39(6):1069–81. doi: 10.1007/s00246-018-1861-4. PubMed PMID: 29569026.

13. Yaich L, Ooi J, Park M, Borg JP, Landry C, Bodmer R, et al. Functional analysis of the Numb phosphotyrosine-binding domain using site-directed mutagenesis. The Journal of biological chemistry. 1998;273(17):10381–8. PubMed PMID: 9553095.

14. Grossfeld P, Ye M, Harvey R. Hypoplastic left heart syndrome: new genetic insights. J Am Coll Cardiol. 2009;53(12):1072–4. doi: 10.1016/j.jacc.2008.12.024. PubMed PMID: 19298922.

15. Miao Y, Tian L, Martin M, Paige SL, Galdos FX, Li J, et al. Intrinsic Endocardial Defects Contribute to Hypoplastic Left Heart Syndrome. Cell Stem Cell. 2020;27(4):574–89.e8. doi: 10.1016/j.stem.2020.07.015.

16. Liu X, Yagi H, Saeed S, Bais AS, Gabriel GC, Chen Z, et al. The complex genetics of hypoplastic left heart syndrome. Nat Genet. 2017;49(7):1152–9. doi: 10.1038/ng.3870.

17. Gaber N, Gagliardi M, Patel P, Kinnear C, Zhang C, Chitayat D, et al. Fetal reprogramming and senescence in hypoplastic left heart syndrome and in human pluripotent stem cells during cardiac differentiation. Am J Pathol. 2013;183(3):720–34. doi: 10.1016/j.ajpath.2013.05.022.

18. Theis JL, Vogler G, Missinato MA, Li X, Nielsen T, Zeng X-XI, et al. Patient-specific genomics and cross-species functional analysis implicate LRP2 in hypoplastic left heart syndrome. eLife. 2020;9:e59554. doi: 10.7554/eLife.59554.

19. Bodmer R. The gene tinman is required for specification of the heart and visceral muscles in Drosophila. Development. 1993;118(3):719–29.

20. Bodmer R. Heart development in Drosophila and its relationship to vertebrates. Trends in Cardiovascular Medicine. 1995;5(1):21–8. doi: 10.1016/1050-1738(94)00032-Q.

21. Cripps RM, Olson EN. Control of cardiac development by an evolutionarily conserved transcriptional network. Dev Biol. 2002;246(1):14–28. doi: 10.1006/dbio.2002.0666.

22. Bodmer R, Frasch M. Chapter 1.2 -Development and Aging of the Drosophila Heart. In: Rosenthal N, Harvey RP, editors. Heart Development and Regeneration. Boston: Academic Press; 2010. p. 47–86.

23. Bier E, Bodmer R. Drosophila, an emerging model for cardiac disease. Gene. 2004;342(1):1–11. doi: 10.1016/j.gene.2004.07.018.

24. Evans SM, Yelon D, Conlon FL, Kirby ML. Myocardial lineage development. Circ Res. 2010;107(12):1428–44. doi: 10.1161/CIRCRESAHA.110.227405.

25. Bakkers J. Zebrafish as a model to study cardiac development and human cardiac disease. Cardiovasc Res. 2011;91(2):279–88. doi: 10.1093/cvr/cvr098.

26. Liu J, Stainier DYR. Zebrafish in the study of early cardiac development. Circ Res. 2012;110(6):870–4. doi: 10.1161/CIRCRESAHA.111.246504.

27. Fink M, Callol-Massot C, Chu A, Ruiz-Lozano P, Izpisua Belmonte JC, Giles W, et al. A new method for detection and quantification of heartbeat parameters in Drosophila, zebrafish, and embryonic mouse hearts. Biotechniques. 2009;46(2):101–13. doi: 10.2144/000113078.

28. Huang G-Y, Xie L-J, Linask KL, Zhang C, Zhao X-Q, Yang Y, et al. Evaluating the role of connexin43 in congenital heart disease: Screening for mutations in patients with outflow tract anomalies and the analysis of knock-in mouse models. J Cardiovasc Dis Res. 2011;2(4):206–12. doi: 10.4103/0975-3583.89804.

29. Yu MS, Spiering S, Colas AR. Generation of First Heart Field-like Cardiac Progenitors and Ventricular-like Cardiomyocytes from Human Pluripotent Stem Cells. JoVE (Journal of Visualized Experiments). 2018;(136):e57688. doi: 10.3791/57688.

30. Paige SL, Galdos FX, Lee S, Chin ET, Ranjbarvaziri S, Feyen DAM, et al. Patient-Specific Induced Pluripotent Stem Cells Implicate Intrinsic Impaired Contractility in Hypoplastic Left Heart Syndrome. Circulation. 2020;142(16):1605–8. doi: 10.1161/CIRCULATIONAHA.119.045317.

31. Diez-Cunado M, Wei K, Bushway PJ, Maurya MR, Perera R, Subramaniam S, et al. miRNAs that Induce Human Cardiomyocyte Proliferation Converge on the Hippo Pathway. Cell reports. 2018;23(7):2168–74. Epub 2018/05/17. doi: 10.1016/j.celrep.2018.04.049. PubMed PMID: 29768213; PubMed Central PMCID: PMCPMC6261450.

32. Murphy SA, Miyamoto M, Kervadec A, Kannan S, Tampakakis E, Kambhampati S, et al. PGC1/PPAR drive cardiomyocyte maturation at single cell level via YAP1 and SF3B2. Nature communications. 2021;12(1):1648. Epub 2021/03/14. doi: 10.1038/s41467-021-21957-z. PubMed PMID: 33712605; PubMed Central PMCID: PMCPMC7955035.

33. Cunningham TJ, Yu MS, McKeithan WL, Spiering S, Carrette F, Huang C-T, et al. Id genes are essential for early heart formation. Genes Dev. 2017;31(13):1325–38. doi: 10.1101/gad.300400.117.

34. Cunningham TJ, Yu MS, McKeithan WL, Spiering S, Carrette F, Huang CT, et al. Id genes are essential for early heart formation. Genes & development. 2017. doi: 10.1101/gad.300400.117. PubMed PMID: 28794185.

35. Zhang Z, Li M, Wang H, Agrawal S, Zhang R. Antisense therapy targeting MDM2 oncogene in prostate cancer: Effects on proliferation, apoptosis, multiple gene expression, and chemotherapy. Proc Natl Acad Sci U S A. 2003;100(20):11636–41. Epub 2003/09/18. doi: 10.1073/pnas.1934692100. PubMed PMID: 13130078; PubMed Central PMCID: PMCPMC208810.

36. Yuan LL, Green A, David L, Dozier C, Recher C, Didier C, et al. Targeting CHK1 inhibits cell proliferation in FLT3-ITD positive acute myeloid leukemia. Leuk Res. 2014;38(11):1342–9. Epub 2014/10/05. doi: 10.1016/j.leukres.2014.08.020. PubMed PMID: 25281057.

37. Zhou X, Liao WJ, Liao JM, Liao P, Lu H. Ribosomal proteins: functions beyond the ribosome. J Mol Cell Biol. 2015;7(2):92–104. Epub 2015/03/05. doi: 10.1093/jmcb/mjv014. PubMed PMID: 25735597; PubMed Central PMCID: PMCPMC4481666.

38. Mi H, Muruganujan A, Ebert D, Huang X, Thomas PD. PANTHER version 14: more genomes, a new PANTHER GO-slim and improvements in enrichment analysis tools. Nucleic Acids Res. 2019;47(D1):D419–D26. doi: 10.1093/nar/gky1038.

39. Szklarczyk D, Gable AL, Lyon D, Junge A, Wyder S, Huerta-Cepas J, et al. STRING v11: protein-protein association networks with increased coverage, supporting functional discovery in genome-wide experimental datasets. Nucleic Acids Res. 2019;47(D1):D607–D13. doi: 10.1093/nar/gky1131.

40. Brand AH, Perrimon N. Targeted gene expression as a means of altering cell fates and generating dominant phenotypes. Development. 1993;118(2):401–15.

41. Gaber N, Gagliardi M, Patel P, Kinnear C, Zhang C, Chitayat D, et al. Fetal reprogramming and senescence in hypoplastic left heart syndrome and in human pluripotent stem cells during cardiac differentiation. Am J Pathol. 2013;183(3):720–34. doi: 10.1016/j.ajpath.2013.05.022. PubMed PMID: 23871585.

42. Liu X, Yagi H, Saeed S, Bais AS, Gabriel GC, Chen Z, et al. The complex genetics of hypoplastic left heart syndrome. Nature genetics. 2017;49(7):1152–9. doi: 10.1038/ng.3870. PubMed PMID: 28530678.

43. Schroeder AM, Allahyari M, Vogler G, Missinato MA, Nielsen T, Yu MS, et al. Model system identification of novel congenital heart disease gene candidates: focus on RPL13. Hum Mol Genet. 2019;28(23):3954–69. doi: 10.1093/hmg/ddz213.

44. Fink M, Callol-Massot C, Chu A, Ruiz-Lozano P, Izpisua Belmonte JC, Giles W, et al. A new method for detection and quantification of heartbeat parameters in Drosophila, zebrafish, and embryonic mouse hearts. Biotechniques. 2009;46(2):101–13. doi: 10.2144/000113078. PubMed PMID: 19317655; PubMed Central PMCID: PMC2855226.

45. Uechi T, Nakajima Y, Nakao A, Torihara H, Chakraborty A, Inoue K, et al. Ribosomal Protein Gene Knockdown Causes Developmental Defects in Zebrafish. PLOS ONE. 2006;1(1):e37. doi: 10.1371/journal.pone.0000037.

46. Ikeda F, Yoshida K, Toki T, Uechi T, Ishida S, Nakajima Y, et al. Exome sequencing identified RPS15A as a novel causative gene for Diamond-Blackfan anemia. Haematologica. 2017;102(3):e93–e6. doi: 10.3324/haematol.2016.153932.

47. Miura GI, Yelon D. A Guide to Analysis of Cardiac Phenotypes in the Zebrafish Embryo. Methods Cell Biol. 2011;101:161–80. doi: 10.1016/B978-0-12-387036-0.00007-4.

48. Fischer M. Census and evaluation of p53 target genes. Oncogene. 2017;36(28):3943–56. Epub 2017/03/14. doi: 10.1038/onc.2016.502. PubMed PMID: 28288132; PubMed Central PMCID: PMCPMC5511239.

49. Robu ME, Larson JD, Nasevicius A, Beiraghi S, Brenner C, Farber SA, et al. p53 activation by knockdown technologies. PLoS Genet. 2007;3(5):e78. doi: 10.1371/journal.pgen.0030078.

50. Furth N, Aylon Y, Oren M. p53 shades of Hippo. Cell Death Differ. 2018;25(1):81–92. doi: 10.1038/cdd.2017.163.

51. Baker NE, Kiparaki M, Khan C. A potential link between p53, cell competition and ribosomopathy in mammals and in Drosophila. Dev Biol. 2019;446(1):17–9. doi: 10.1016/j.ydbio.2018.11.018.

52. Ollmann M, Young LM, Di Como CJ, Karim F, Belvin M, Robertson S, et al. Drosophila p53 is a structural and functional homolog of the tumor suppressor p53. Cell. 2000;101(1):91–101. doi: 10.1016/S0092-8674(00)80626-1. PubMed PMID: 10778859.

53. Raj N, Bam R. Reciprocal Crosstalk Between YAP1/Hippo Pathway and the p53 Family Proteins: Mechanisms and Outcomes in Cancer. Front Cell Dev Biol. 2019;7:159. Epub 20190809. doi: 10.3389/fcell.2019.00159. PubMed PMID: 31448276; PubMed Central PMCID: PMCPMC6695833.

54. Monroe TO, Hill MC, Morikawa Y, Leach JP, Heallen T, Cao S, et al. YAP Partially Reprograms Chromatin Accessibility to Directly Induce Adult Cardiogenesis In Vivo. Developmental Cell. 2019;48(6):765–79.e7. doi: 10.1016/j.devcel.2019.01.017.

55. Kang J, Brajanovski N, Chan KT, Xuan J, Pearson RB, Sanij E. Ribosomal proteins and human diseases: molecular mechanisms and targeted therapy. Signal Transduct Target Ther. 2021;6(1):323. Epub 2021/09/01. doi: 10.1038/s41392-021-00728-8. PubMed PMID: 34462428; PubMed Central PMCID: PMCPMC8405630.

56. Bruneau BG, Logan M, Davis N, Levi T, Tabin CJ, Seidman JG, et al. Chamber-specific cardiac expression of Tbx5 and heart defects in Holt-Oram syndrome. Developmental biology. 1999;211(1):100–8. doi: 10.1006/dbio.1999.9298. PubMed PMID: 10373308.

57. Qian L, Wythe JD, Liu J, Cartry J, Vogler G, Mohapatra B, et al. Tinman/Nkx2-5 acts via miR-1 and upstream of Cdc42 to regulate heart function across species. J Cell Biol. 2011;193(7):1181–96. doi: 10.1083/jcb.201006114. PubMed PMID: 21690310; PubMed Central PMCID: PMCPMC3216339.

58. Lee KH, Xu Q, Breitbart RE. A new tinman-related gene, nkx2.7, anticipates the expression of nkx2.5 and nkx2.3 in zebrafish heart and pharyngeal endoderm. Dev Biol. 1996;180(2):722–31. doi: 10.1006/dbio.1996.0341.

59. Targoff KL, Schell T, Yelon D. Nkx Genes Regulate Heart Tube Extension and Exert Differential Effects on Ventricular and Atrial Cell Number. Dev Biol. 2008;322(2):314–21. doi: 10.1016/j.ydbio.2008.07.037.

60. Tiu GC, Kerr CH, Forester CM, Krishnarao PS, Rosenblatt HD, Raj N, et al. A p53-dependent translational program directs tissue-selective phenotypes in a model of ribosomopathies. Developmental Cell. 2021;0(0). doi: 10.1016/j.devcel.2021.06.013.

61. McGowan KA, Pang WW, Bhardwaj R, Perez MG, Pluvinage JV, Glader BE, et al. Reduced ribosomal protein gene dosage and p53 activation in low-risk myelodysplastic syndrome. Blood. 2011;118(13):3622–33. Epub 20110725. doi: 10.1182/blood-2010-11-318584. PubMed PMID: 21788341; PubMed Central PMCID: PMCPMC3186336.

62. Flucke U, van Noesel MM, Siozopoulou V, Creytens D, Tops BBJ, van Gorp JM, et al. EWSR1-The Most Common Rearranged Gene in Soft Tissue Lesions, Which Also Occurs in Different Bone Lesions: An Updated Review. Diagnostics (Basel). 2021;11(6). Epub 20210615. doi: 10.3390/diagnostics11061093. PubMed PMID: 34203801; PubMed Central PMCID: PMCPMC8232650.

63. Vlachos A, Osorio DS, Atsidaftos E, Kang J, Lababidi ML, Seiden HS, et al. The Increased Prevalence of Congenital Heart Disease (CHD) in Children with Diamond Blackfan Anemia (DBA) Suggests Unrecognized DBA as a Cause of CHD in the General Population: A Report of the Diamond Blackfan Anemia Registry. Circ Genom Precis Med. 2018;11(5):e002044. doi: 10.1161/CIRCGENETICS.117.002044.

64. Jin SC, Homsy J, Zaidi S, Lu Q, Morton S, DePalma SR, et al. Contribution of rare inherited and de novo variants in 2,871 congenital heart disease probands. Nat Genet. 2017;49(11):1593–601. doi: 10.1038/ng.3970.

65. Ye M, Coldren C, Liang X, Mattina T, Goldmuntz E, Benson DW, et al. Deletion of ETS-1, a gene in the Jacobsen syndrome critical region, causes ventricular septal defects and abnormal ventricular morphology in mice. Hum Mol Genet. 2010;19(4):648–56. Epub 20091126. doi: 10.1093/hmg/ddp532. PubMed PMID: 19942620; PubMed Central PMCID: PMCPMC2807373.

66. Kathiriya IS, Rao KS, Iacono G, Devine WP, Blair AP, Hota SK, et al. Modeling Human TBX5 Haploinsufficiency Predicts Regulatory Networks for Congenital Heart Disease. Developmental cell. 2021;56(3):292–309 e9. Epub 2020/12/16. doi: 10.1016/j.devcel.2020.11.020. PubMed PMID: 33321106; PubMed Central PMCID: PMCPMC7878434.

67. Ang YS, Rivas RN, Ribeiro AJS, Srivas R, Rivera J, Stone NR, et al. Disease Model of GATA4 Mutation Reveals Transcription Factor Cooperativity in Human Cardiogenesis. Cell. 2016;167(7):1734–49 e22. doi: 10.1016/j.cell.2016.11.033. PubMed PMID: 27984724; PubMed Central PMCID: PMCPMC5180611.

68. Tanaka M, Yamaguchi S, Yamazaki Y, Kinoshita H, Kuwahara K, Nakao K, et al. Somatic chromosomal translocation between Ewsr1 and Fli1 loci leads to dilated cardiomyopathy in a mouse model. Sci Rep. 2015;5:7826. Epub 20150116. doi: 10.1038/srep07826. PubMed PMID: 25591392; PubMed Central PMCID: PMCPMC5379005.

69. Ragni CV, Diguet N, Le Garrec JF, Novotova M, Resende TP, Pop S, et al. Amotl1 mediates sequestration of the Hippo effector Yap1 downstream of Fat4 to restrict heart growth. Nat Commun. 2017;8:14582. Epub 20170227. doi: 10.1038/ncomms14582. PubMed PMID: 28239148; PubMed Central PMCID: PMCPMC5333361.

70. Danilova N, Gazda HT. Ribosomopathies: how a common root can cause a tree of pathologies. Dis Model Mech. 2015;8(9):1013–26. doi: 10.1242/dmm.020529.

71. Boulon S, Westman BJ, Hutten S, Boisvert F-M, Lamond AI. The nucleolus under stress. Mol Cell. 2010;40(2):216–27. doi: 10.1016/j.molcel.2010.09.024.

72. Yang K, Yang J, Yi J. Nucleolar Stress: hallmarks, sensing mechanism and diseases. Cell Stress. 2018;2(6):125–40. doi: 10.15698/cst2018.06.139.

73. Dai M-S, Zeng SX, Jin Y, Sun X-X, David L, Lu H. Ribosomal protein L23 activates p53 by inhibiting MDM2 function in response to ribosomal perturbation but not to translation inhibition. Mol Cell Biol. 2004;24(17):7654–68. doi: 10.1128/MCB.24.17.7654-7668.2004.

74. Zhang Y, Lu H. Signaling to p53: ribosomal proteins find their way. Cancer Cell. 2009;16(5):369–77. doi: 10.1016/j.ccr.2009.09.024.

75. Men H, Cai H, Cheng Q, Zhou W, Wang X, Huang S, et al. The regulatory roles of p53 in cardiovascular health and disease. Cell Mol Life Sci. 2021;78(5):2001–18. Epub 20201111. doi: 10.1007/s00018-020-03694-6. PubMed PMID: 33179140.

76. Mak TW, Hauck L, Grothe D, Billia F. p53 regulates the cardiac transcriptome. Proc Natl Acad Sci U S A. 2017;114(9):2331–6. Epub 20170213. doi: 10.1073/pnas.1621436114. PubMed PMID: 28193895; PubMed Central PMCID: PMCPMC5338492.

77. Deisenroth C, Zhang Y. The Ribosomal Protein-Mdm2-p53 Pathway and Energy Metabolism. Genes Cancer. 2011;2(4):392–403. doi: 10.1177/1947601911409737.

78. Liu Y, Deisenroth C, Zhang Y. RP–MDM2–p53 Pathway: Linking Ribosomal Biogenesis and Tumor Surveillance. Trends Cancer. 2016;2(4):191–204. doi: 10.1016/j.trecan.2016.03.002.

79. Cui Z, DiMario PJ. RNAi knockdown of Nopp140 induces Minute-like phenotypes in Drosophila. Mol Biol Cell. 2007;18(6):2179–91. doi: 10.1091/mbc.e07-01-0074.

80. Barron DA, Kagey JD. The role of the Hippo pathway in human disease and tumorigenesis. Clin Transl Med. 2014;3:25. doi: 10.1186/2001-1326-3-25.

81. Del Re DP. The hippo signaling pathway: implications for heart regeneration and disease. Clin Transl Med. 2014;3(1):27. doi: 10.1186/s40169-014-0027-0.

82. Yu L, Daniels JP, Wu H, Wolf MJ. Cardiac hypertrophy induced by active Raf depends on Yorkie-mediated transcription. Sci Signal. 2015;8(362):ra13-ra. doi: 10.1126/scisignal.2005719.

83. Wang J, Liu S, Heallen T, Martin JF. The Hippo pathway in the heart: pivotal roles in development, disease, and regeneration. Nat Rev Cardiol. 2018;15(11):672–84. doi: 10.1038/s41569-018-0063-3.

84. Xiao Y, Leach J, Wang J, Martin JF. Hippo/Yap Signaling in Cardiac Development and Regeneration. Curr Treat Options Cardiovasc Med. 2016;18(6):38. doi: 10.1007/s11936-016-0461-y.

85. von Gise A, Lin Z, Schlegelmilch K, Honor LB, Pan GM, Buck JN, et al. YAP1, the nuclear target of Hippo signaling, stimulates heart growth through cardiomyocyte proliferation but not hypertrophy. Proceedings of the National Academy of Sciences of the United States of America. 2012;109(7):2394–9. doi: 10.1073/pnas.1116136109.

86. Del Re DP, Yang Y, Nakano N, Cho J, Zhai P, Yamamoto T, et al. Yes-associated protein isoform 1 (Yap1) promotes cardiomyocyte survival and growth to protect against myocardial ischemic injury. J Biol Chem. 2013;288(6):3977–88. doi: 10.1074/jbc.M112.436311.

87. Gifford CA, Ranade SS, Samarakoon R, Salunga HT, de Soysa TY, Huang Y, et al. Oligogenic inheritance of a human heart disease involving a genetic modifier. Science. 2019;364(6443):865-70. Epub 20190530. doi: 10.1126/science.aat5056. PubMed PMID: 31147515; PubMed Central PMCID: PMCPMC6557373.

88. Birla AK, Brimmer S, Short WD, Olutoye OO, 2nd, Shar JA, Lalwani S, et al. Current state of the art in hypoplastic left heart syndrome. Front Cardiovasc Med. 2022;9:878266. Epub 20221028. doi: 10.3389/fcvm.2022.878266. PubMed PMID: 36386362; PubMed Central PMCID: PMCPMC9651920.

89. Theis JL, Vogler G, Missinato MA, Li X, Nielsen T, Zeng XI, et al. Patient-specific genomics and cross-species functional analysis implicate LRP2 in hypoplastic left heart syndrome. eLife. 2020;9. Epub 2020/10/03. doi: 10.7554/eLife.59554. PubMed PMID: 33006316; PubMed Central PMCID: PMCPMC7581429.

90. Miao Y, Tian L, Martin M, Paige SL, Galdos FX, Li J, et al. Intrinsic Endocardial Defects Contribute to Hypoplastic Left Heart Syndrome. Cell stem cell. 2020;27(4):574–89 e8. Epub 20200817. doi: 10.1016/j.stem.2020.07.015. PubMed PMID: 32810435; PubMed Central PMCID: PMCPMC7541479.

91. Xu X, Jin K, Bais AS, Zhu W, Yagi H, Feinstein TN, et al. Uncompensated mitochondrial oxidative stress underlies heart failure in an iPSC-derived model of congenital heart disease. Cell stem cell. 2022;29(5):840–55 e7. Epub 20220407. doi: 10.1016/j.stem.2022.03.003. PubMed PMID: 35395180; PubMed Central PMCID: PMCPMC9302582.

92. Bray MA, Singh S, Han H, Davis CT, Borgeson B, Hartland C, et al. Cell Painting, a high-content image-based assay for morphological profiling using multiplexed fluorescent dyes. Nature protocols. 2016;11(9):1757–74. Epub 20160825. doi: 10.1038/nprot.2016.105. PubMed PMID: 27560178; PubMed Central PMCID: PMCPMC5223290.

93. Karczewski KJ, Francioli LC, Tiao G, Cummings BB, Alföldi J, Wang Q, et al. The mutational constraint spectrum quantified from variation in 141,456 humans. Nature. 2020;581(7809):434-43. doi: 10.1038/s41586-020-2308-7.

94. Grodecká L, Kramárek M, Lockerová P, Kováčová T, Ravčuková B, Richterová R, et al. No major effect of the CDH1 c.2440-6C>G mutation on splicing detected in last exon-specific splicing minigene assay. Genes, Chromosomes and Cancer. 2014;53(9):798–801. doi: 10.1002/gcc.22186.

95. Burridge PW, Holmström A, Wu JC. Chemically Defined Culture and Cardiomyocyte Differentiation of Human Pluripotent Stem Cells. Curr Protoc Hum Genet. 2015;87:21.3.1-.3.15. doi: 10.1002/0471142905.hg2103s87.

96. Pei F, Jiang J, Bai S, Cao H, Tian L, Zhao Y, et al. Chemical-defined and albumin-free generation of human atrial and ventricular myocytes from human pluripotent stem cells. Stem Cell Research. 2017;19:94–103. doi: 10.1016/j.scr.2017.01.006.

97. Ewels PA, Peltzer A, Fillinger S, Patel H, Alneberg J, Wilm A, et al. The nf-core framework for community-curated bioinformatics pipelines. Nat Biotechnol. 2020;38(3):276–8. doi: 10.1038/s41587-020-0439-x. PubMed PMID: 32055031.

98. Love MI, Huber W, Anders S. Moderated estimation of fold change and dispersion for RNA-seq data with DESeq2. Genome Biol. 2014;15(12):550. doi: 10.1186/s13059-014-0550-8. PubMed PMID: 25516281; PubMed Central PMCID: PMCPMC4302049.

99. Raudvere U, Kolberg L, Kuzmin I, Arak T, Adler P, Peterson H, et al. g:Profiler: a web server for functional enrichment analysis and conversions of gene lists (2019 update). Nucleic Acids Res. 2019;47(W1):W191–W8. doi: 10.1093/nar/gkz369. PubMed PMID: 31066453; PubMed Central PMCID: PMCPMC6602461.

100. Hu Y, Flockhart I, Vinayagam A, Bergwitz C, Berger B, Perrimon N, et al. An integrative approach to ortholog prediction for disease-focused and other functional studies. BMC Bioinformatics. 2011;12:357. doi: 10.1186/1471-2105-12-357. PubMed PMID: 21880147; PubMed Central PMCID: PMCPMC3179972.

101. Han Z, Yi P, Li X, Olson EN. Hand, an evolutionarily conserved bHLH transcription factor required for Drosophila cardiogenesis and hematopoiesis. Development. 2006;133(6):1175–82. doi: 10.1242/dev.02285.

102. Tögel M, Meyer H, Lehmacher C, Heinisch JJ, Pass G, Paululat A. The bHLH transcription factor hand is required for proper wing heart formation in Drosophila. Dev Biol. 2013;381(2):446–59. doi: 10.1016/j.ydbio.2013.05.027.

103. Hallier B, Hoffmann J, Roeder T, Tögel M, Meyer H, Paululat A. The bHLH Transcription Factor Hand Regulates the Expression of Genes Critical to Heart and Muscle Function in Drosophila melanogaster. PLOS ONE. 2015;10(8):e0134204. doi: 10.1371/journal.pone.0134204.

104. Vogler G. gvogler/FlyHearts-tdtK-Rscripts: First release of the R tdtK script. First ed: Zenodo; 2021.

105. Cammarato A, Ocorr S, Ocorr K. Enhanced assessment of contractile dynamics in Drosophila hearts. BioTechniques. 2015;58(2):77–80. doi: 10.2144/000114255. PubMed PMID: 25652030.

106. Alayari NN, Vogler G, Taghli-Lamallem O, Ocorr K, Bodmer R, Cammarato A. Fluorescent labeling of Drosophila heart structures. J Vis Exp. 2009;(32). doi: 10.3791/1423.

107. Ocorr K, Vogler G, Bodmer R. Methods to assess Drosophila heart development, function and aging. Methods. 2014;68(1):265–72. doi: 10.1016/j.ymeth.2014.03.031.

108. Huang C-J, Tu C-T, Hsiao C-D, Hsieh F-J, Tsai H-J. Germ-line transmission of a myocardium-specific GFP transgene reveals critical regulatory elements in the cardiac myosin light chain 2 promoter of zebrafish. Dev Dyn. 2003;228(1):30–40. doi: 10.1002/dvdy.10356.

109. Schumacher JA, Bloomekatz J, Garavito-Aguilar ZV, Yelon D. tal1 Regulates the formation of intercellular junctions and the maintenance of identity in the endocardium. Dev Biol. 2013;383(2):214–26. doi: 10.1016/j.ydbio.2013.09.019.

110. Westerfield M. The zebrafish book: A guide for the laboratory use of zebrafish (Brachydanio rerio). 1993 ed: M. W, editor: University of Oregon Press; 1993.

111. Talbot JC, Amacher SL. A streamlined CRISPR pipeline to reliably generate zebrafish frameshifting alleles. Zebrafish. 2014;11(6):583–5. doi: 10.1089/zeb.2014.1047.

112. Gagnon JA, Valen E, Thyme SB, Huang P, Akhmetova L, Ahkmetova L, et al. Efficient mutagenesis by Cas9 protein-mediated oligonucleotide insertion and large-scale assessment of single-guide RNAs. PLOS ONE. 2014;9(5):e98186. doi: 10.1371/journal.pone.0098186.

113. Irion U, Krauss J, Nüsslein-Volhard C. Precise and efficient genome editing in zebrafish using the CRISPR/Cas9 system. Development. 2014;141(24):4827–30. doi: 10.1242/dev.115584.

114. Ocorr K, Fink M, Cammarato A, Bernstein SI, Bodmer R. Semi-automated Optical Heartbeat Analysis of Small Hearts. Journal of Visualized Experiments : JoVE. 2009;(31). doi: 10.3791/1435.

115. Zeng X-XI, Yelon D. Cadm4 restricts the production of cardiac outflow tract progenitor cells. Cell Reports. 2014;7(4):951–60. doi: 10.1016/j.celrep.2014.04.013.

116. Pradhan A, Zeng X-XI, Sidhwani P, Marques SR, George V, Targoff KL, et al. FGF signaling enforces cardiac chamber identity in the developing ventricle. Development. 2017;144(7):1328–38. doi: 10.1242/dev.143719.

117. Alexander J, Stainier DY, Yelon D. Screening mosaic F1 females for mutations affecting zebrafish heart induction and patterning. Dev Genet. 1998;22(3):288–99. doi: 10.1002/(SICI)1520-6408(1998)22:3<288::AID-DVG10>3.0.CO;2-2.

118. Hoshijima K, Jurynec MJ, Klatt Shaw D, Jacobi AM, Behlke MA, Grunwald DJ. Highly Efficient CRISPR-Cas9-Based Methods for Generating Deletion Mutations and F0 Embryos that Lack Gene Function in Zebrafish. Developmental Cell. 2019;51(5):645–57.e4. doi: 10.1016/j.devcel.2019.10.004.

