## Supplemental Table 1 for "Functional analysis across model systems implicates ribosomal proteins in growth and proliferation defects associated with hypoplastic left heart syndrome"

| Gene Name | Normalized % EdU+CM | Normalized Tot Cell num |
|---|---|---|
| MEX3B | 2.140494219 | 1.182998053 |
| KRTAP5-9 | 1.801725824 | 0.922128488 |
| CDKN1A | 1.796442731 | 1.296560675 |
| HECTD3 | 1.776662765 | 1.181700195 |
| ETV7 | 1.755501908 | 0.979883193 |
| CDKN1B | 1.734760818 | 1.028552888 |
| PCGF6 | 1.728359282 | 1.121998702 |
| SH3RF3 | 1.723701433 | 1.134328358 |
| USP28 | 1.709444121 | 1.068786502 |
| KCTD14 | 1.708193631 | 1.003893576 |
| TP53 | 1.693106582 | 1.145360156 |
| KRT33B | 1.666043482 | 1.032806804 |
| SLCO6A1 | 1.661387196 | 1.073977936 |
| OR14A16 | 1.654052877 | 1.06399352 |
| INPP5A | 1.65252897 | 1.057105775 |
| TBC1D10C | 1.642099204 | 1.057918186 |
| ALDH9A1 | 1.640854713 | 1.142764439 |
| ITGAL | 1.632958845 | 0.870214147 |
| SLC3A2 | 1.625710861 | 1.081116158 |
| TTYH1 | 1.617509163 | 1.070084361 |
| TCTN1 | 1.614449487 | 1.176994735 |
| PPAP2B | 1.613975438 | 1.019467878 |
| SEC22B | 1.607937804 | 0.931551235 |
| PIK3C2B | 1.603911753 | 0.979234263 |
| UBE2J2 | 1.594528469 | 1.172615185 |
| PPAP2A | 1.587261304 | 1.046722907 |
| TRIP12 | 1.582951107 | 1.093445814 |
| TNK1 | 1.578321084 | 1.066190785 |
| GRIN3B | 1.560959774 | 1.074626866 |
| SCN9A | 1.55746647 | 1.00843608 |
| OXGR1 | 1.55581095 | 0.872160934 |
| BRDT | 1.555171298 | 1.006489293 |
| TRIM71 | 1.553605346 | 1.196625568 |
| ISOC2 | 1.547082465 | 1.042829332 |
| RNASE7 | 1.545594226 | 0.985074627 |
| HS3ST1 | 1.542452442 | 0.903958469 |
| RAB13 | 1.537616117 | 1.011031798 |
| DUSP14 | 1.536935715 | 1.209604153 |
| GRIN3A | 1.536622311 | 0.914341337 |
| SEPHS1 | 1.534994616 | 1.169370539 |

| | | |
|---|---|---|
| CRABP2 | 1.530037441 | 1.036988968 |
| PGK1 | 1.528476757 | 0.996106424 |
| PCSK2 | 1.526845522 | 1.098637249 |
| UBE2D4 | 1.526453746 | 1.014925373 |
| NUDT12 | 1.523912263 | 1.002595717 |
| CASP2 | 1.522184875 | 1.109020117 |
| DUSP28 | 1.520822663 | 1.117861482 |
| KCNT2 | 1.520235626 | 0.9733939 |
| MTRR | 1.52011708 | 1.082414017 |
| FAM45B | 1.516305187 | 1.074929121 |
| KLF16 | 1.512580636 | 1.045425049 |
| TNFRSF10D | 1.512473533 | 1.052563271 |
| CLOCK | 1.511847959 | 1.031797534 |
| RNF208 | 1.511442435 | 1.060350422 |
| POLA1 | 1.51061097 | 1.053861129 |
| AK2 | 1.508437555 | 1.083711875 |
| ARSH | 1.507098485 | 0.988968202 |
| PPIP5K1 | 1.50603668 | 1.049967554 |
| SLC16A11 | 1.503370355 | 1.088294856 |
| SRPX2 | 1.502648736 | 1.073714054 |
| ZAK | 1.50112577 | 1.007138222 |
| PMP2 | 1.500548167 | 1.051842851 |
| SPINK1 | 1.497744838 | 1.012978585 |
| IFNAR1 | 1.497328282 | 0.895522388 |
| TLE4 | 1.496785937 | 0.8137573 |
| GAL3ST2 | 1.490483682 | 0.924724205 |
| PRCP | 1.48982666 | 0.907203115 |
| P2RY10 | 1.487478966 | 0.887086308 |
| CSNK2B | 1.48607692 | 1.179104478 |
| SLC17A6 | 1.485085336 | 1.079169371 |
| ADA | 1.484108983 | 1.072680078 |
| PPM1K | 1.48172342 | 1.128487995 |
| PAK1 | 1.479605573 | 1.15898767 |
| FFAR4 | 1.478568465 | 0.986372485 |
| DDR1 | 1.478003975 | 1.00843608 |
| KHDC1 | 1.476914981 | 1.001620089 |
| ZNF526 | 1.470930405 | 0.880095923 |
| PTPN1 | 1.470057643 | 1.072031149 |
| GON4L | 1.469749379 | 0.988254354 |
| CCDC144NL | 1.467614069 | 1.137302552 |
| AGL | 1.466955457 | 1.107073329 |
| ARHGAP35 | 1.466903758 | 1.08890331 |
| PRKAA2 | 1.46562536 | 0.903958469 |
| NPBWR1 | 1.465074371 | 0.842310188 |

| | | |
|---|---|---|
| MCCD1 | 1.464991516 | 0.894289186 |
| CDK17 | 1.463635114 | 1.069435432 |
| UCHL3 | 1.462690619 | 1.139519792 |
| DYRK1A | 1.461814371 | 0.972096042 |
| B3GAT3 | 1.458732025 | 0.842310188 |
| KLK13 | 1.453057567 | 1.066190785 |
| PDE7A | 1.453035093 | 1.154445165 |
| MET | 1.452208074 | 1.1330305 |
| PON2 | 1.451210645 | 1.012329656 |
| GPR108 | 1.450778284 | 0.791693705 |
| CHD8 | 1.448047046 | 1.066190785 |
| FAM221A | 1.446555298 | 0.962737951 |
| ASB9 | 1.445035415 | 0.975989617 |
| ZNF398 | 1.444974167 | 0.927968851 |
| GALNT7 | 1.444834629 | 0.99091499 |
| FCGR3B | 1.444267093 | 0.998702141 |
| SLC26A7 | 1.44252184 | 1.073977936 |
| CAMK2N2 | 1.442413403 | 1.041312272 |
| HSF5 | 1.439478892 | 0.997569866 |
| CRHR2 | 1.438936664 | 0.931862427 |
| SLC41A1 | 1.435609406 | 0.952628164 |
| GAPDHS | 1.434614694 | 1.038935756 |
| KNCN | 1.433982855 | 1.099230458 |
| RASD1 | 1.432914143 | 1.155743024 |
| PPP2R2A | 1.432222958 | 0.90266061 |
| CYBB | 1.429688719 | 1.03504218 |
| ILK | 1.428439629 | 0.994808566 |
| CPOX | 1.426487663 | 1.112264763 |
| PVRL4 | 1.425602075 | 1.027541515 |
| CBLB | 1.424185165 | 0.802725503 |
| UFC1 | 1.423941817 | 1.046722907 |
| LPPR4 | 1.423317313 | 0.885788449 |
| RNPEPL1 | 1.420985755 | 0.909798832 |
| NUDT5 | 1.42047922 | 0.992861778 |
| KAL1 | 1.420162193 | 0.925373134 |
| CHRNA4 | 1.418797027 | 0.971447112 |
| SETD1A | 1.417322536 | 1.081765088 |
| EAPP | 1.415957826 | 1.082624544 |
| PHF10 | 1.413906863 | 0.978585334 |
| DMKN | 1.413272973 | 0.992709599 |
| HAO1 | 1.412824395 | 1.070084361 |
| ASIC3 | 1.412305429 | 1.01752109 |
| SCRN1 | 1.411823509 | 1.074626866 |
| NME6 | 1.411255314 | 1.181051265 |

| | | |
|---|---|---|
| CCT8L2 | 1.410096038 | 1.0532122 |
| SAT1 | 1.410009352 | 1.102530824 |
| PTPN14 | 1.409350766 | 1.121998702 |
| MYOM2 | 1.408051553 | 0.989064399 |
| COX15 | 1.407840356 | 1.151883354 |
| ERLEC1 | 1.407787174 | 1.06399352 |
| ZDHHC19 | 1.406419509 | 1.030832477 |
| LMNB1 | 1.406072936 | 1.230900993 |
| MST1R | 1.40551862 | 1.037637897 |
| NRD1 | 1.404592648 | 0.985074627 |
| KCTD15 | 1.40450094 | 1.152498378 |
| NT5C | 1.40440454 | 0.935756003 |
| NR1H4 | 1.401968842 | 1.101232966 |
| MIPOL1 | 1.401914874 | 0.844809866 |
| SMC3 | 1.4007098 | 1.066423653 |
| MFSD4 | 1.400615103 | 1.075334143 |
| CRISP3 | 1.400605744 | 1.108256252 |
| ATRX | 1.399973492 | 0.892926671 |
| GBE1 | 1.398643972 | 1.004542505 |
| SFTPC | 1.397865873 | 0.993833505 |
| COX4I2 | 1.397741133 | 1.005191434 |
| TRIM11 | 1.396209674 | 1.10577547 |
| RSPH3 | 1.395854852 | 0.836245289 |
| EREG | 1.395516512 | 0.872809864 |
| CRAT | 1.394904686 | 0.896171317 |
| PTPRG | 1.394518179 | 1.030499676 |
| SLC10A7 | 1.393990734 | 0.892926671 |
| TXNRD3 | 1.393832426 | 0.981181051 |
| HACL1 | 1.392104626 | 0.932511356 |
| BCL2L15 | 1.391789765 | 0.964713943 |
| RHOG | 1.391452956 | 1.009733939 |
| MTMR6 | 1.391086041 | 1.029201817 |
| TP53RK | 1.389551783 | 0.865022713 |
| SERTAD2 | 1.388992808 | 1.103828683 |
| PPP1R3A | 1.388336774 | 1.038935756 |
| CRNN | 1.387908092 | 0.900308325 |
| GK2 | 1.386691051 | 1.155743024 |
| STS | 1.386604529 | 1.040233615 |
| ZNF93 | 1.386319149 | 0.950992305 |
| ARHGEF1 | 1.386204451 | 1.038026721 |
| ZNF326 | 1.385556422 | 1.008100446 |
| MMP27 | 1.385426566 | 0.998702141 |
| DOT1L | 1.383078378 | 0.888384166 |
| PPAPDC2 | 1.382949789 | 1.06229721 |

| | | |
|---|---|---|
| WTAP | 1.382815973 | 1.075368277 |
| ELP3 | 1.382527554 | 1.020116807 |
| DCLK1 | 1.381950468 | 0.989617132 |
| PTPN2 | 1.381924783 | 1.082414017 |
| PARG | 1.381116286 | 0.98247891 |
| SGPP1 | 1.38004024 | 1.11486048 |
| FAM122A | 1.380007509 | 0.910490077 |
| FZD5 | 1.379237341 | 0.99156392 |
| SET | 1.379072036 | 1.014925373 |
| AR | 1.3789921 | 1.0266061 |
| PTPRD | 1.378738545 | 1.065541856 |
| KCNK15 | 1.377520161 | 1.140168722 |
| SLC26A5 | 1.376499929 | 1.001297859 |
| GUCY2F | 1.375994135 | 1.129785853 |
| AK3 | 1.37586283 | 1.168721609 |
| RFWD2 | 1.375693162 | 0.978585334 |
| HIF1A | 1.375540699 | 1.021414666 |
| DLEC1 | 1.374687254 | 1.014276444 |
| SPAST | 1.374508283 | 1.08825438 |
| OLFML1 | 1.374433787 | 1.149858242 |
| ADPRM | 1.373744516 | 0.90562981 |
| DUSP23 | 1.373048775 | 0.894873459 |
| C11orf16 | 1.372061288 | 1.081534772 |
| RYR2 | 1.372053734 | 0.9733939 |
| C9orf50 | 1.371318795 | 1.075739166 |
| CELSR1 | 1.371219635 | 1.067488644 |
| SLC24A2 | 1.370380514 | 1.064243997 |
| LPAR3 | 1.369526987 | 1.057105775 |
| VDAC3 | 1.368917631 | 1.135626217 |
| PIM3 | 1.368759488 | 1.081116158 |
| GTPBP1 | 1.368315272 | 0.952628164 |
| PPP1R7 | 1.368025412 | 1.105126541 |
| NFKBIE | 1.366856586 | 1.027903958 |
| GUCY2C | 1.365551105 | 0.958468527 |
| PHF21A | 1.365311462 | 1.056456846 |
| COX10 | 1.365261409 | 1.048669695 |
| PGC | 1.364916886 | 0.998053212 |
| SNTN | 1.364789691 | 1.034021871 |
| MUL1 | 1.364556228 | 1.068786502 |
| KMT2A | 1.363003283 | 0.983776768 |
| ITPR1 | 1.362143911 | 0.988319273 |
| MYOCD | 1.36200215 | 1.062946139 |
| CEP41 | 1.361863024 | 0.961064244 |
| GBA | 1.361737563 | 0.949383517 |

| | | |
|---|---|---|
| PPP1R8 | 1.361131171 | 1.059701493 |
| C12orf65 | 1.360782227 | 1.099230458 |
| ALG2 | 1.360170541 | 0.974042829 |
| PWWP2B | 1.359808098 | 0.980153908 |
| DEDD | 1.358984213 | 1.0222679 |
| FOXP2 | 1.358768562 | 0.75617659 |
| CAMTA2 | 1.358117123 | 1.060637205 |
| ACOXL | 1.358071052 | 1.018818949 |
| OR52D1 | 1.357513373 | 1.049674546 |
| ATAD5 | 1.35726654 | 0.966788173 |
| GNAT1 | 1.357054476 | 1.018631025 |
| PYCRL | 1.357038142 | 0.948734588 |
| ASUN | 1.356636665 | 1.011340624 |
| MYOM1 | 1.356217156 | 0.909900651 |
| CAMSAP2 | 1.35597541 | 1.075739166 |
| ALX3 | 1.355854828 | 1.099286178 |
| PPP2R3A | 1.355523065 | 1.131083712 |
| THTPA | 1.355321456 | 1.064243997 |
| PRMT7 | 1.355114824 | 1.103828683 |
| KCNK17 | 1.354300812 | 1.0532122 |
| SLC27A3 | 1.353935462 | 1.081116158 |
| ORAOV1 | 1.353807456 | 1.047961631 |
| ATP5E | 1.353476675 | 1.006489293 |
| PLCH1 | 1.353035257 | 0.983127839 |
| DPYD | 1.353011384 | 0.888384166 |
| TMEM44 | 1.352871472 | 0.994329688 |
| TYR | 1.352705388 | 1.058403634 |
| CALML5 | 1.352683401 | 1.009720535 |
| SCN10A | 1.352489454 | 1.046722907 |
| MMP21 | 1.352453068 | 0.908500973 |
| PPID | 1.352453068 | 0.999351071 |
| PTPN6 | 1.352342837 | 1.038935756 |
| SLCO4C1 | 1.352342837 | 1.038935756 |
| CP | 1.351983907 | 0.975989617 |
| CBWD1 | 1.351866575 | 1.07290401 |
| SUMO4 | 1.351701828 | 0.900308325 |
| B3GALNT1 | 1.351168743 | 0.980532122 |
| IGSF22 | 1.350371736 | 0.993114621 |
| EXOC8 | 1.349967332 | 1.032545392 |
| EDEM2 | 1.34972998 | 1.036988968 |
| FAM193B | 1.349647491 | 0.917780062 |
| MLX | 1.348949309 | 1.001946788 |
| SGTB | 1.348757475 | 1.076053443 |
| SEC14L2 | 1.348656979 | 1.065541856 |

| | | |
|---|---|---|
| DHX40 | 1.348622074 | 1.025957171 |
| PPP1R16B | 1.348132498 | 1.042180402 |
| RTN4IP1 | 1.346726252 | 1.019467878 |
| SCN4A | 1.345714253 | 1.115509409 |
| PPARD | 1.34503311 | 0.889682025 |
| CFC1B | 1.344687627 | 0.992709599 |
| CILP | 1.344502537 | 0.969500324 |
| IZUMO2 | 1.344416575 | 1.061968408 |
| VIPR2 | 1.343324903 | 0.898767034 |
| CHKB | 1.343006642 | 1.042180402 |
| BAZ2B | 1.342747818 | 1.002595717 |
| POMT2 | 1.340272276 | 0.980532122 |
| PCDHB1 | 1.339516447 | 0.94210346 |
| PNCK | 1.33883837 | 1.045425049 |
| CLCC1 | 1.338714226 | 1.041531473 |
| RAB14 | 1.338690764 | 0.853990915 |
| CDA | 1.338137964 | 0.866320571 |
| CD1B | 1.336867291 | 1.155933576 |
| CHEK2 | 1.336613255 | 1.055158988 |
| SLC25A36 | 1.33542813 | 0.948085659 |
| TRPM7 | 1.335359308 | 1.116158339 |
| RHCG | 1.335316882 | 1.055498458 |
| PPP2R5E | 1.334971319 | 0.984425698 |
| MED12L | 1.334147442 | 1.020656136 |
| TXNL4B | 1.33409186 | 1.149253731 |
| CBX8 | 1.333941126 | 0.981181051 |
| DUSP22 | 1.333941126 | 1.121349773 |
| MMP8 | 1.333844725 | 1.025308241 |
| MS4A13 | 1.33369865 | 1.116135663 |
| TRAF6 | 1.333304109 | 1.125892278 |
| DNAL1 | 1.332962319 | 1.119076549 |
| ASB10 | 1.332435187 | 0.853990915 |
| PAIP1 | 1.332399047 | 1.010382868 |
| FZD2 | 1.33212878 | 0.914341337 |
| FGF20 | 1.33161994 | 0.99091499 |
| KCNQ5 | 1.330042956 | 0.996106424 |
| ARID1A | 1.329517075 | 0.900064893 |
| ATP5O | 1.32931349 | 1.020765737 |
| SMYD4 | 1.329102489 | 0.98218568 |
| ALPK2 | 1.329008977 | 0.992861778 |
| WSCD1 | 1.328958014 | 1.069663832 |
| VN1R1 | 1.32816327 | 0.933160286 |
| AMBRA1 | 1.327835225 | 1.046248715 |
| BRE | 1.327833968 | 1.034831916 |

| | | |
|---|---|---|
| G6PC | 1.32749143 | 1.138870863 |
| LIF | 1.32740993 | 0.897469176 |
| GRAMD2 | 1.327379683 | 1.094552929 |
| GATSL3 | 1.327266409 | 1.003425831 |
| LIPA | 1.326937889 | 1.006489293 |
| TPSD1 | 1.326349689 | 0.954574951 |
| MAPK10 | 1.326346396 | 1.071382219 |
| NFE4 | 1.326099279 | 1.07571086 |
| EXD2 | 1.326068381 | 0.99108951 |
| SLC30A1 | 1.325176748 | 0.987670344 |
| TEX9 | 1.324909543 | 1.153908465 |
| BCKDHB | 1.324376319 | 1.00843608 |
| DUSP19 | 1.324041099 | 0.96430889 |
| SLC6A14 | 1.323911522 | 1.109669046 |
| FOLR3 | 1.323784049 | 0.918122645 |
| VIL1 | 1.323548983 | 1.025008565 |
| CLDN5 | 1.323376399 | 1.015758822 |
| RAPGEF6 | 1.323371235 | 0.896171317 |
| ENPP7 | 1.323143858 | 1.037637897 |
| HDHD3 | 1.322956244 | 0.997404283 |
| ZNF354A | 1.322699313 | 1.082414017 |
| HNRNPA1 | 1.322693764 | 1.1113395 |
| ANKRD13C | 1.322439505 | 0.864317537 |
| KCTD21 | 1.320134003 | 0.995544755 |
| TMPRSS11E | 1.319884435 | 1.024010383 |
| TPRX1 | 1.319714358 | 1.038072094 |
| ZFP14 | 1.319117798 | 0.983799109 |
| AQP3 | 1.318716515 | 0.968202466 |
| TCERG1 | 1.317856237 | 0.936404932 |
| ADAMTS1 | 1.317386748 | 0.993510707 |
| IRF7 | 1.317359087 | 1.139519792 |
| RPAP1 | 1.316973564 | 0.979445015 |
| TRIAP1 | 1.316564982 | 1.042122317 |
| GRXCR2 | 1.316323326 | 0.906817403 |
| HEMK1 | 1.316247971 | 0.925373134 |
| OBSL1 | 1.315989103 | 0.995544755 |
| IMPA2 | 1.314954837 | 1.060350422 |
| UQCRH | 1.314706633 | 1.040233615 |
| G6PC2 | 1.314611385 | 1.085009734 |
| MAN2B2 | 1.314251056 | 0.878001298 |
| MBD1 | 1.314196147 | 1.077222583 |
| DPP7 | 1.313788482 | 0.947436729 |
| RERE | 1.313788482 | 0.947436729 |
| SIK2 | 1.313781238 | 1.00843608 |

| | | |
|---|---|---|
| KCNG4 | 1.312551411 | 1.01752109 |
| EFNA1 | 1.31166858 | 1.088729017 |
| AMER1 | 1.311598722 | 1.069663832 |
| CCNT1 | 1.311530207 | 0.71891454 |
| CASR | 1.310766149 | 0.900713822 |
| FZD9 | 1.310590307 | 0.941596366 |
| ABCB11 | 1.310017337 | 1.105126541 |
| RWDD1 | 1.309934718 | 1.009592326 |
| SLC7A7 | 1.309807248 | 1.015574302 |
| ZNF774 | 1.309024256 | 1.089104901 |
| EPHA4 | 1.30883781 | 1 |
| NT5C1A | 1.308548197 | 1.065541856 |
| ACP1 | 1.30848061 | 1.1064244 |
| CCDC144B | 1.30815106 | 1.044193217 |
| FBP1 | 1.308111966 | 1.155743024 |
| PLCXD2 | 1.307626918 | 1.119402985 |
| POU6F1 | 1.307456179 | 1.033744322 |
| ABCB7 | 1.307421357 | 0.96430889 |
| PRCD | 1.306867213 | 1.063035286 |
| CCDC24 | 1.306050725 | 0.982584042 |
| UBR1 | 1.30597749 | 1.145360156 |
| IFNK | 1.305721091 | 1.088699878 |
| DYM | 1.305673157 | 1.090319968 |
| SLC6A3 | 1.305519114 | 0.990266061 |
| TPH1 | 1.304975534 | 0.87605451 |
| SMC1B | 1.304927868 | 0.994808566 |
| COX20 | 1.304694301 | 1.006509078 |
| NLGN3 | 1.30389216 | 1.110317975 |
| DPYS | 1.303761513 | 1.003893576 |
| PTPRO | 1.303761513 | 0.860480208 |
| FCGR1A | 1.303577932 | 1.0532122 |
| MPO | 1.303365291 | 1.036988968 |
| TSG101 | 1.303313592 | 0.971447112 |
| VPREB3 | 1.302787084 | 0.997259335 |
| AKR7L | 1.302779068 | 0.861129137 |
| MRPL4 | 1.302708698 | 1.006851662 |
| SNCG | 1.302391622 | 1.047619048 |
| FPR3 | 1.302288339 | 1.033744322 |
| DUSP8 | 1.30215053 | 1.095392602 |
| GLUL | 1.302039339 | 0.976103686 |
| GPRIN3 | 1.30199767 | 1.068043742 |
| MAP4K2 | 1.301981849 | 0.78780013 |
| ABCG4 | 1.301948838 | 1.066839714 |
| F2RL2 | 1.301643254 | 0.935756003 |

| | | |
|---|---|---|
| SMURF2 | 1.301574679 | 0.972744971 |
| BACH1 | 1.301313302 | 1.145360156 |
| NLGN4X | 1.301232345 | 1.041717294 |
| ATP8B2 | 1.300194941 | 0.792991564 |
| STPG2 | 1.30012945 | 0.915724563 |
| OR5L1 | 1.299894565 | 0.872901679 |
| ACSL6 | 1.299840458 | 1.011031798 |
| PXK | 1.299312354 | 1.064892927 |
| PPP2R2C | 1.299201382 | 0.879948086 |
| HDHD1 | 1.298710571 | 0.995457495 |
| KCNK18 | 1.298269566 | 1.073977936 |
| INPP5B | 1.298173988 | 1.012329656 |
| PSKH1 | 1.298173988 | 1.012329656 |
| B3GNT9 | 1.298153408 | 1 |
| HBE1 | 1.298110764 | 0.975340688 |
| RASA4 | 1.297963565 | 0.948967193 |
| PRLH | 1.29777585 | 1.041312272 |
| AKR7A2 | 1.297532865 | 0.987444309 |
| TGM7 | 1.297357507 | 1.13238157 |
| XPC | 1.296428818 | 0.988968202 |
| FAP | 1.296020578 | 1.030499676 |
| COPS4 | 1.295953975 | 1.010935601 |
| RILP | 1.295420604 | 1.030489894 |
| EIF4E3 | 1.29534528 | 1.125892278 |
| CHST11 | 1.295040437 | 0.878650227 |
| PRICKLE3 | 1.294913742 | 1.027255029 |
| ARNT2 | 1.294912248 | 0.987849332 |
| CHUK | 1.294682331 | 1.03569111 |
| MED21 | 1.294164271 | 0.891628812 |
| AIFM3 | 1.294011727 | 1.006075334 |
| ZNF225 | 1.293853481 | 0.941760877 |
| KRBA2 | 1.293802873 | 1.168894289 |
| HMOX1 | 1.293295432 | 1.073977936 |
| COPS2 | 1.292949345 | 0.979234263 |
| TGM2 | 1.292896905 | 1.012329656 |
| PKLR | 1.292835903 | 0.842959117 |
| FGL2 | 1.292764282 | 1.033095393 |
| KLF13 | 1.292446515 | 0.950681376 |
| ALG3 | 1.291912648 | 0.922128488 |
| UPRT | 1.291824458 | 0.985419198 |
| ADAMTS19 | 1.291774016 | 0.975989617 |
| RNF2 | 1.291640406 | 1.062946139 |
| FGD5 | 1.291477582 | 1.007194245 |
| LCP2 | 1.29117727 | 1.114422748 |

| | | |
|---|---|---|
| NME2 | 1.291148442 | 1.067488644 |
| CCDC115 | 1.291076453 | 1.115431349 |
| EPPK1 | 1.290814567 | 1.006509078 |
| GAL3ST3 | 1.290801614 | 0.96430889 |
| IAPP | 1.290650745 | 0.992709599 |
| IL5 | 1.290498171 | 1.113562622 |
| BAG6 | 1.290347606 | 1.105515588 |
| CBS | 1.29026355 | 0.968851395 |
| NARF | 1.290100714 | 1.010382868 |
| NAALAD2 | 1.2900924 | 0.836469825 |
| SLC22A4 | 1.290029632 | 0.967598218 |
| COMT | 1.289993118 | 1.085009734 |
| AKR1B15 | 1.289986384 | 0.938841636 |
| CFI | 1.289580596 | 1.068786502 |
| OR8G1 | 1.289548553 | 1.028756582 |
| MTMR14 | 1.289399979 | 1.023361454 |
| CCR10 | 1.289344081 | 1.151849448 |
| ZNF280B | 1.289318609 | 1.108950992 |
| ITGA5 | 1.288730691 | 0.961713173 |
| CD74 | 1.288679321 | 0.98247891 |
| PRR14 | 1.287788824 | 0.938841636 |
| TNFSF12 | 1.287626221 | 1.087358684 |
| STK32A | 1.287543261 | 1.074626866 |
| CYP4F11 | 1.287496995 | 1.116158339 |
| HIST1H4H | 1.287235924 | 1.12024666 |
| LILRB1 | 1.286459311 | 1.050616483 |
| OR4A16 | 1.286408244 | 1.039287161 |
| SLC25A53 | 1.286402907 | 0.92428914 |
| TAOK3 | 1.286144013 | 1.067488644 |
| GALNT2 | 1.285496952 | 1.038935756 |
| RAB5C | 1.285463529 | 0.997404283 |
| FUT3 | 1.285259899 | 0.921425678 |
| GPR155 | 1.285124193 | 0.993510707 |
| RIT2 | 1.285124193 | 0.993510707 |
| SRC | 1.285099084 | 0.972744971 |
| LEKR1 | 1.284858134 | 1.090319968 |
| PPFIA3 | 1.28481334 | 1.010382868 |
| ADCY8 | 1.283793026 | 0.998702141 |
| PKD2L2 | 1.283165147 | 1.011680727 |
| TMEM61 | 1.28266628 | 1.069663832 |
| SFI1 | 1.282526943 | 0.976704351 |
| BDKRB1 | 1.282498493 | 1.053861129 |
| CDK20 | 1.282446959 | 1.083062946 |
| PRICKLE2 | 1.282422098 | 1.033095393 |

| | | |
|---|---|---|
| TRAF3IP2 | 1.282207207 | 1.066828676 |
| TPST2 | 1.282160639 | 1.129136924 |
| P2RX5 | 1.281956395 | 1.037637897 |
| MIEF1 | 1.281893772 | 0.922235723 |
| NOMO1 | 1.281783385 | 1.059133252 |
| ITCH | 1.281469947 | 1.071382219 |
| OAS2 | 1.281397091 | 1.021414666 |
| NSUN5 | 1.281238043 | 1.08825438 |
| GLT8D2 | 1.280940856 | 0.996755354 |
| AADAC | 1.280846188 | 0.913325233 |
| DEFB108B | 1.280343867 | 1.131227217 |
| TPSG1 | 1.280326066 | 0.922128488 |
| BSPH1 | 1.280307815 | 0.933538883 |
| SOGA1 | 1.280190373 | 1.018499486 |
| GTF3C4 | 1.280157386 | 1.114211551 |
| IL6R | 1.279881273 | 1.014276444 |
| TRIM28 | 1.279881273 | 1.014276444 |
| C1QTNF3 | 1.2794429 | 0.968139774 |
| ING1 | 1.27903318 | 1.148604802 |
| TBC1D23 | 1.278800685 | 1.071283921 |
| ABCC10 | 1.278757561 | 1.040233615 |
| CS | 1.27857324 | 0.885788449 |
| UNC5B | 1.277853076 | 0.869565217 |
| UBE2Q1 | 1.277607094 | 1.036988968 |
| ATP6V0E1 | 1.277257385 | 1.07073329 |
| GRIK3 | 1.277154152 | 1.003893576 |
| WARS2 | 1.276982343 | 0.824140169 |
| MTMR1 | 1.27685005 | 1.112913692 |
| DCAF17 | 1.276436551 | 1.055488052 |
| ZFP41 | 1.276348323 | 0.906844876 |
| SUCLG1 | 1.276347049 | 0.725799919 |
| THNSL2 | 1.276329109 | 0.753406879 |
| EPM2A | 1.276059665 | 1.075924724 |
| ADAM15 | 1.275840191 | 1.101232966 |
| TUSC1 | 1.275472712 | 1.048197651 |
| FAM173B | 1.275092817 | 0.956834532 |
| SCN11A | 1.27478135 | 1.043478261 |
| SRXN1 | 1.274606824 | 0.913692408 |
| GLRX3 | 1.274490891 | 1.045767517 |
| NPPB | 1.274278931 | 1.110966905 |
| KIFC2 | 1.274189775 | 0.909798832 |
| PEX14 | 1.274086355 | 0.985611511 |
| CSE1L | 1.27404999 | 0.872160934 |
| ATPAF1 | 1.273993787 | 1.002740665 |

| | | |
|---|---|---|
| HSPA1B | 1.273812076 | 1.069435432 |
| HBD | 1.273504435 | 1.082219522 |
| ANAPC5 | 1.273453147 | 1.065541856 |
| TULP2 | 1.273039637 | 0.881245944 |
| CDKN3 | 1.272916975 | 0.898118105 |
| NARS | 1.27269816 | 1.129136924 |
| BZRAP1 | 1.272392727 | 0.950325454 |
| ABCB1 | 1.272092179 | 1.104477612 |
| MGA | 1.272056304 | 0.916409729 |
| RUNDC3B | 1.272017745 | 0.964028777 |
| PAGE1 | 1.271943169 | 0.99108951 |
| ASTL | 1.271832696 | 1.003893576 |
| STOX1 | 1.271409825 | 0.985014176 |
| MED9 | 1.271167707 | 1.103685703 |
| TMEM154 | 1.270974644 | 1.019441069 |
| BMP15 | 1.27083675 | 0.950032446 |
| TMX1 | 1.270381338 | 0.84944841 |
| IFT27 | 1.269879578 | 0.975989617 |
| TSPAN7 | 1.269512304 | 1.033616849 |
| TRIM33 | 1.269500561 | 0.824789098 |
| RIPPLY3 | 1.268714784 | 0.969218307 |
| SLC33A1 | 1.268699392 | 0.939000649 |
| FKTN | 1.268668686 | 1.096265844 |
| ETAA1 | 1.268632618 | 1.05872823 |
| FAM83F | 1.268306448 | 0.893074119 |
| P2RX4 | 1.267956095 | 1.137573005 |
| ALG10B | 1.267938561 | 0.863724854 |
| ATG5 | 1.267859448 | 0.914341337 |
| DGAT1 | 1.267859448 | 0.914341337 |
| RAB26 | 1.267859448 | 0.914341337 |
| PQBP1 | 1.26747496 | 0.88420692 |
| ZNF142 | 1.26742308 | 0.996916752 |
| KERA | 1.267412795 | 0.939362795 |
| NT5C1B | 1.267381644 | 0.944192083 |
| PHC1 | 1.267254846 | 1.185680027 |
| SEMA4B | 1.267099981 | 1.046982584 |
| HERC4 | 1.266986575 | 0.864373783 |
| SNX29 | 1.266645391 | 0.902389631 |
| GPR39 | 1.266511158 | 0.776119403 |
| KPNA3 | 1.266511158 | 0.852044127 |
| MINPP1 | 1.266511158 | 1.062946139 |
| TESC | 1.266468526 | 0.953017416 |
| CTXN3 | 1.266274495 | 0.998784933 |
| KCTD6 | 1.266140868 | 1.109669046 |

| | | |
|---|---|---|
| RABGEF1 | 1.266136378 | 1.029804728 |
| MBD2 | 1.266078303 | 0.949383517 |
| PARP10 | 1.265829406 | 1.049674546 |
| GIYD1 | 1.265690622 | 1.167523124 |
| NGLY1 | 1.265679035 | 0.987670344 |
| ANXA3 | 1.265575061 | 0.878001298 |
| ZC3H13 | 1.265196397 | 0.869177805 |
| PTPN11 | 1.265127469 | 0.890979883 |
| TRMT1L | 1.264875972 | 1.005191434 |
| FAM105A | 1.264875678 | 1.016200891 |
| RING1 | 1.264773891 | 0.946138871 |
| TREML4 | 1.264485986 | 1.086674767 |
| PRIM2 | 1.264405906 | 0.975989617 |
| METTL22 | 1.26404897 | 1.036452005 |
| WDTC1 | 1.264031566 | 1.1113395 |
| GPR112 | 1.263821411 | 1.069435432 |
| RENBP | 1.263497469 | 1.090850097 |
| SLC37A1 | 1.263401316 | 1.057105775 |
| TRMU | 1.263225226 | 0.875405581 |
| HMGCS2 | 1.26259496 | 1.049318624 |
| HYOU1 | 1.262408004 | 0.901362751 |
| RPL22 | 1.262396932 | 1.158273381 |
| NDUFV3 | 1.262386375 | 1.047619048 |
| CLASRP | 1.262238439 | 1.042822885 |
| MTRF1L | 1.261801438 | 0.969623329 |
| MYL5 | 1.261786204 | 1.055498458 |
| MPPED2 | 1.261737893 | 1.033095393 |
| RNASEH2C | 1.26152794 | 0.904414743 |
| CYP19A1 | 1.261485325 | 1.062946139 |
| HDLBP | 1.261365792 | 1.118105127 |
| AQP4 | 1.26124237 | 1.177804779 |
| HAAO | 1.260968418 | 0.889682025 |
| PPIF | 1.260955169 | 1.027541515 |
| MYL6B | 1.260935353 | 1.025921426 |
| PPP1R1C | 1.260907169 | 0.953277093 |
| FZR1 | 1.260867597 | 1.02672148 |
| FAM45A | 1.260783501 | 1.094775213 |
| LHFPL2 | 1.260622248 | 1.046722907 |
| CHST14 | 1.260592941 | 0.90266061 |
| BCHE | 1.26029205 | 1.123296561 |
| SYNJ1 | 1.260161732 | 0.970798183 |
| MAP3K13 | 1.259787079 | 1.038935756 |
| MOK | 1.259787079 | 1.038935756 |
| PSMB6 | 1.259528337 | 0.941596366 |

| | | |
|---|---|---|
| TBRG1 | 1.259432869 | 1.035286057 |
| TSGA10 | 1.25941982 | 0.94655704 |
| ABTB1 | 1.259381884 | 0.979883193 |
| APOH | 1.259381884 | 0.979883193 |
| CCR6 | 1.259127588 | 0.946138871 |
| G6PC3 | 1.258993483 | 0.92926671 |
| SCN3A | 1.258748654 | 1.005840363 |
| STAG2 | 1.258599943 | 1.023638232 |
| DRICH1 | 1.258298384 | 1.0854597 |
| MEOX2 | 1.258282046 | 1.048669695 |
| ZNF804A | 1.258019436 | 0.965978129 |
| C2orf71 | 1.258004408 | 1.031591738 |
| POP7 | 1.257715991 | 1.121349773 |
| C12orf54 | 1.257681346 | 1.010530579 |
| MAPK12 | 1.257554226 | 0.917585983 |
| HLX | 1.257503844 | 1.049318624 |
| DUSP5 | 1.257402752 | 0.947436729 |
| BAHD1 | 1.257281242 | 1.024010383 |
| CRISP1 | 1.257199468 | 0.985268928 |
| TAC3 | 1.257126557 | 1.007138222 |
| CHDH | 1.256966685 | 0.990266061 |
| SPSB1 | 1.256843201 | 1.020116807 |
| RASSF10 | 1.256719594 | 1.054813292 |
| SLC9C2 | 1.256633325 | 0.877012676 |
| CCNYL1 | 1.256563107 | 1.065613609 |
| DARS2 | 1.256549506 | 1.095180235 |
| IFNA6 | 1.256270712 | 1.036656389 |
| NEU3 | 1.256105584 | 0.829331603 |
| CUL3 | 1.25606574 | 0.944192083 |
| RPRM | 1.25606574 | 0.944192083 |
| USP49 | 1.255979795 | 0.897469176 |
| DCLK2 | 1.255868247 | 1.003893576 |
| TTLL10 | 1.255816935 | 1.04981774 |
| PSAT1 | 1.255592958 | 0.978585334 |
| MCM3AP | 1.255567145 | 0.938335046 |
| OXSM | 1.25555332 | 1.005670312 |
| IMPA1 | 1.25513096 | 1.119402985 |
| IL17A | 1.255078405 | 0.979748886 |
| RBM47 | 1.254890627 | 1.022953066 |
| CASP3 | 1.254777217 | 1.085658663 |
| GLYATL1 | 1.254701315 | 1.00908501 |
| GRK1 | 1.254601704 | 1.000648929 |
| TICRR | 1.254567526 | 0.871607938 |
| SLC35F2 | 1.254510389 | 0.99109284 |

| | | |
|---|---|---|
| EDNRA | 1.254504233 | 0.924075276 |
| TRNT1 | 1.254449159 | 1.022063595 |
| ZMPSTE24 | 1.254449159 | 0.953926022 |
| RFC5 | 1.254125059 | 0.745240988 |
| C7orf60 | 1.254086956 | 1.089104901 |
| TMEM74 | 1.254006827 | 1.00526529 |
| TOR1A | 1.253999509 | 1.018170019 |
| B3GALT4 | 1.253830664 | 1.069435432 |
| POLR3B | 1.253758385 | 1.031148605 |
| HTRA3 | 1.253653176 | 1.15055159 |
| PMM1 | 1.253523352 | 1.04412719 |
| KCNMB3 | 1.253344957 | 0.967553537 |
| KDM7A | 1.25307963 | 1.040233615 |
| VSNL1 | 1.252939958 | 0.868448099 |
| PRKAR2B | 1.252873245 | 0.903958469 |
| USP53 | 1.252744739 | 0.895522388 |
| PFKM | 1.252644222 | 1.155743024 |
| SUV420H2 | 1.252198195 | 1.06229721 |
| TOMM20L | 1.252122755 | 0.971908188 |
| NME4 | 1.252009181 | 1.105126541 |
| ADAMTSL5 | 1.251672901 | 1.134062373 |
| PPP1R13B | 1.251496719 | 0.930564568 |
| TYW3 | 1.251454979 | 0.865532604 |
| OPRM1 | 1.25139579 | 1.033095393 |
| SLC19A2 | 1.251359403 | 0.922128488 |
| PRSS50 | 1.25122711 | 0.994808566 |
| F13A1 | 1.251144869 | 1.016223232 |
| TBX1 | 1.251066003 | 1.037637897 |
| C1RL | 1.2508476 | 1.101881895 |
| NGEF | 1.250760839 | 0.891058582 |
| SPATA19 | 1.250743956 | 0.951802349 |
| ODF1 | 1.250567397 | 1.034426893 |
| AFP | 1.250546791 | 1.003893576 |
| ACAT1 | 1.250537818 | 1.157689812 |
| PHF1 | 1.25047945 | 0.922777417 |
| GAPT | 1.250151063 | 1.038072094 |
| CABYR | 1.250031853 | 0.956149366 |
| CACNA1D | 1.249549033 | 0.944841012 |
| KANK3 | 1.249401784 | 1.048602673 |
| IQCH | 1.248977656 | 1.093560146 |
| MAGT1 | 1.248820557 | 1.128392062 |
| CAPN6 | 1.248777088 | 1.112264763 |
| BAGE5 | 1.248622362 | 0.907502569 |
| LDLRAP1 | 1.248589033 | 1.097205346 |

| | | |
|---|---|---|
| SLC2A14 | 1.248429602 | 1.022712524 |
| STRBP | 1.248428573 | 1.076738609 |
| SERINC5 | 1.248420269 | 1.069258809 |
| STK25 | 1.248299943 | 0.992861778 |
| NSMAF | 1.248266274 | 1.068174032 |
| TIPARP | 1.24822223 | 1.078520441 |
| C20orf144 | 1.248154157 | 1.119076549 |
| SLC18A3 | 1.248066804 | 0.868916288 |
| ATR | 1.247915658 | 0.817650876 |
| LMTK3 | 1.247846836 | 0.924724205 |
| QRFPR | 1.24776476 | 1.096041531 |
| PMPCB | 1.247651812 | 0.980532122 |
| PCDHB16 | 1.247612411 | 1.043850634 |
| FAT4 | 1.24747594 | 1.089914945 |
| TSR2 | 1.247369882 | 1.164439044 |
| CHRNA2 | 1.247321614 | 1.134977287 |
| NR1I2 | 1.247321614 | 1.113562622 |
| RAD21 | 1.247321614 | 0.899415964 |
| CHST1 | 1.247166105 | 1.040882544 |
| GAP43 | 1.247066929 | 1.031860226 |
| OR2W3 | 1.247025606 | 1.03562864 |
| OR52B6 | 1.246859184 | 1.101747174 |
| MAGI3 | 1.246608942 | 0.908500973 |
| C17orf103 | 1.24653667 | 0.965056526 |
| FSTL3 | 1.246489367 | 1.060979788 |
| ADARB1 | 1.246407826 | 1.062946139 |
| CABLES2 | 1.246211529 | 0.981500514 |
| CABLES1 | 1.246167134 | 1 |
| RAB42 | 1.246133309 | 1.036452005 |
| DOPEY1 | 1.246044572 | 1.145808019 |
| PPIB | 1.24594591 | 1.059052563 |
| TIMM50 | 1.245917509 | 1.037637897 |
| TRIM59 | 1.245824813 | 0.9733939 |
| TADA3 | 1.245612913 | 1.042180402 |
| BUB3 | 1.245402659 | 1.012329656 |
| INPP5J | 1.245390146 | 1.089552239 |
| RCC2 | 1.245360479 | 1.009592326 |
| MGAT4B | 1.245272671 | 0.948085659 |
| DSG2 | 1.245043357 | 1.119561494 |
| TMC5 | 1.244769098 | 0.966426859 |
| SLC25A41 | 1.244750895 | 1.112191171 |
| BTNL3 | 1.244429955 | 0.9554642 |
| C2orf66 | 1.244354787 | 1.029119561 |
| ZC3H12A | 1.244340215 | 1.008100446 |

| | | |
|---|---|---|
| NAGPA | 1.244291707 | 0.961713173 |
| SLC5A6 | 1.244233752 | 0.996106424 |
| SYF2 | 1.243973904 | 1.117861482 |
| PPP6C | 1.243924299 | 1.000648929 |
| SLC22A10 | 1.243862091 | 0.922235723 |
| SLC30A5 | 1.24355335 | 1.073977936 |
| DIABLO | 1.243338157 | 0.975340688 |
| DHX16 | 1.243198208 | 0.863724854 |
| C6orf120 | 1.242910498 | 1.000342583 |
| GOLGA6B | 1.242891324 | 1.010935601 |
| ATP1B1 | 1.242890648 | 1.096041531 |
| CHAF1A | 1.242885924 | 0.909680032 |
| ECHDC2 | 1.242623175 | 1.119076549 |
| RASL12 | 1.242518218 | 1.044776119 |
| TMEM110 | 1.242492972 | 1.014580802 |
| SLC17A8 | 1.242424205 | 1.057754705 |
| SLMO2 | 1.242319164 | 0.810449575 |
| ASB12 | 1.242188596 | 0.946138871 |
| SEPT5 | 1.242069721 | 0.924724205 |
| PRDX1 | 1.241945166 | 1.053861129 |
| PINX1 | 1.241773726 | 1.025008565 |
| ZNF99 | 1.24157971 | 0.962658445 |
| GALP | 1.24153875 | 0.945186708 |
| TLDC1 | 1.241317594 | 1.032887975 |
| OMD | 1.240836814 | 0.936964714 |
| AGPAT4 | 1.240638038 | 0.968851395 |
| NXT1 | 1.240569463 | 0.887086308 |
| C17orf67 | 1.240552304 | 1.064398542 |
| PTGS2 | 1.240264373 | 0.986372485 |
| PDK4 | 1.240207323 | 1.046722907 |
| TPM4 | 1.240207323 | 1.046722907 |
| VPS36 | 1.240080394 | 1.064803564 |
| HSPB6 | 1.239961115 | 0.89756766 |
| SPINK7 | 1.239951668 | 1.029971648 |
| PROCA1 | 1.239938053 | 1.002740665 |
| ALPP | 1.239839256 | 1.016872161 |
| PSTPIP2 | 1.239813875 | 0.918590522 |
| MGAT4A | 1.239782394 | 1.040097205 |
| BAHCC1 | 1.239685607 | 1.021871203 |
| SCYL2 | 1.239640067 | 1.00526529 |
| PSORS1C1 | 1.23961563 | 1.085645769 |
| GJA9 | 1.239360813 | 1.012150668 |
| C5orf38 | 1.239280175 | 0.859052248 |
| ZBTB1 | 1.239116952 | 0.995889003 |

| | | |
|---|---|---|
| LRP2 | 1.239011084 | 1.090850097 |
| OR52M1 | 1.238875226 | 1.027406646 |
| KCNF1 | 1.238844709 | 1.069435432 |
| LOC653978_pseudogene | 1.238844709 | 1.069435432 |
| TPST1 | 1.238796714 | 1.082414017 |
| ACN9 | 1.238690815 | 0.996231586 |
| MDK | 1.238575061 | 1.092750101 |
| LIG1 | 1.238537643 | 1.013627515 |
| DLGAP4 | 1.238425857 | 1.014580802 |
| ENC1 | 1.2383157 | 1.078520441 |
| FIGNL1 | 1.238284388 | 0.989037342 |
| RPL3L | 1.238274692 | 1.158273381 |
| GLO1 | 1.238133271 | 1.057105775 |
| NT5M | 1.238133271 | 1.057105775 |
| NDUFA4L2 | 1.238118865 | 1.143413368 |
| PTH1R | 1.238046338 | 1.169370539 |
| MDGA1 | 1.237976193 | 0.936622131 |
| C11orf24 | 1.237692917 | 0.928057554 |
| C9orf66 | 1.237687706 | 1.002055498 |
| CREB1 | 1.237470158 | 1.018818949 |
| SNX32 | 1.2373289 | 1.050017129 |
| TTC12 | 1.237203256 | 1.043850634 |
| EYS | 1.237138101 | 0.992463172 |
| BTRC | 1.237128763 | 0.937053861 |
| RRH | 1.237128763 | 0.937053861 |
| IL4I1 | 1.237097728 | 0.950032446 |
| GLT1D1 | 1.236927236 | 1.027903958 |
| ATP8B1 | 1.236780865 | 0.829331603 |
| SLC27A6 | 1.236615066 | 0.907203115 |
| COBLL1 | 1.236569291 | 1.019441069 |
| DHCR7 | 1.236454207 | 0.998053212 |
| FKBP7 | 1.236433626 | 1.011031798 |
| PDP2 | 1.23631327 | 0.91174562 |
| AQR | 1.236291616 | 0.997974889 |
| SLC12A8 | 1.236162223 | 0.949297705 |
| ODF4 | 1.236148494 | 0.979445015 |
| BCKDK | 1.236084493 | 1.080467229 |
| PIAS2 | 1.23583316 | 1.085009734 |
| PRODH2 | 1.23560175 | 1.063595068 |
| WASF3 | 1.235251772 | 1.215142172 |
| DYNLRB1 | 1.23513967 | 0.916937054 |
| MYCBP2 | 1.23513967 | 0.916937054 |
| CACNA1B | 1.235135636 | 0.929915639 |
| ZNF681 | 1.235128092 | 0.87363305 |

| | | |
|---|---|---|
| C15orf41 | 1.234984135 | 0.883758607 |
| CD164 | 1.234810955 | 0.623501199 |
| EIF1B | 1.234563983 | 0.939000649 |
| SPOCD1 | 1.234554862 | 1.046172539 |
| HOXD9 | 1.234465767 | 0.977723775 |
| NDST3 | 1.23446266 | 0.832320778 |
| CLEC2A | 1.234459987 | 1.010962658 |
| DUX4 | 1.23445532 | 1.007194245 |
| ZNF80 | 1.234433777 | 1.042932361 |
| C6orf58 | 1.234407848 | 0.989469421 |
| USP14 | 1.234394644 | 1.17715769 |
| MATN2 | 1.234223878 | 1.04008222 |
| SPATA21 | 1.234216466 | 1.07654921 |
| AMMECR1L | 1.234015915 | 1.044957473 |
| HIBADH | 1.233644844 | 0.887735237 |
| CDK3 | 1.233558076 | 1.03504218 |
| APOBEC3C | 1.233471719 | 0.970149254 |
| C3orf18 | 1.2333893 | 0.861482382 |
| LMOD1 | 1.233210257 | 1.069886948 |
| THYN1 | 1.233146926 | 0.920210612 |
| ALDOA | 1.233135758 | 0.935756003 |
| GGT1 | 1.233122092 | 1.083062946 |
| KLHDC9 | 1.233105308 | 0.829890644 |
| CYP2C18 | 1.233086858 | 0.909798832 |
| CCKBR | 1.232963949 | 0.992212849 |
| DNAL4 | 1.232951518 | 0.844905905 |
| ROCK2 | 1.232835607 | 1.06164828 |
| AKAP10 | 1.232810663 | 1.048669695 |
| C2CD3 | 1.232687566 | 0.792052073 |
| CHORDC1 | 1.232456804 | 1.039054471 |
| ACKR4 | 1.232447535 | 1.001297859 |
| TTBK1 | 1.23231623 | 0.949383517 |
| RGS10 | 1.232300046 | 1.07654921 |
| TSPAN15 | 1.232267088 | 1.063183475 |
| HUS1 | 1.232241727 | 1.127190136 |
| SREK1IP1 | 1.232153898 | 1.110166059 |
| NAALADL1 | 1.232095192 | 0.776119403 |
| KCNIP1 | 1.232090006 | 0.871512005 |
| TRIM41 | 1.232048268 | 0.953926022 |
| RXFP1 | 1.231863699 | 0.984425698 |
| ITPKB | 1.231746882 | 1.205710578 |
| GUSBP2 | 1.231732938 | 0.956491949 |
| HIC1 | 1.231730088 | 1.027903958 |
| ATP6V1F | 1.231683987 | 1.09733939 |

| | | |
|---|---|---|
| KRTAP4-12 | 1.231660747 | 1.134062373 |
| TTC6 | 1.23161835 | 1.067488866 |
| BTK | 1.231485177 | 1.032446463 |
| SLC25A18 | 1.231398408 | 1.006489293 |
| CAPN11 | 1.231227176 | 0.88513952 |
| PARP15 | 1.231207175 | 1.012150668 |
| MAP2K2 | 1.231001528 | 0.833225178 |
| ST14 | 1.230861825 | 1.072031149 |
| DBH | 1.230812925 | 1.059052563 |
| TRPC5 | 1.230799754 | 0.989617132 |
| TFF2 | 1.230773417 | 0.94210346 |
| DQX1 | 1.230765487 | 0.997601918 |
| XYLB | 1.230680714 | 1.089552239 |
| ANKRD37 | 1.230597778 | 1.130012151 |
| KRT25 | 1.230519075 | 0.995949777 |
| FAM114A1 | 1.230467422 | 1.054678007 |
| SERPINA13P | 1.230355572 | 0.973963686 |
| AQP1 | 1.230180271 | 0.972744971 |
| PTS | 1.230005828 | 1.103179753 |
| LMBR1L | 1.229945248 | 1.17368962 |
| MEMO1 | 1.229900458 | 1.147428109 |
| HRNR | 1.229809818 | 1.003425831 |
| CD36 | 1.22972453 | 0.93835172 |
| C5orf20 | 1.229678053 | 0.812069664 |
| IGFBP5 | 1.229632748 | 1.113394998 |
| MARCH9 | 1.229502751 | 0.899415964 |
| RBBP6 | 1.229209845 | 0.947436729 |
| SCNN1G | 1.22919692 | 1.134328358 |
| NR4A1 | 1.2291877 | 0.99091499 |
| OLR1 | 1.229142825 | 0.953422438 |
| RRAD | 1.229008071 | 1.051914341 |
| UGT8 | 1.228983178 | 1.054272985 |
| EPO | 1.228837428 | 1.025693731 |
| RNASEK | 1.228708658 | 1.006851662 |
| BAI3 | 1.228627573 | 1.030499676 |
| SENP2 | 1.228627573 | 1.030499676 |
| HCN1 | 1.228620411 | 0.943543154 |
| PTPRJ | 1.228230775 | 1.00908501 |
| TGFBR3 | 1.228168045 | 1.039584685 |
| PPP1R3C | 1.228160389 | 0.878650227 |
| SMARCA5 | 1.228108772 | 1.070084361 |
| KHK | 1.228100622 | 1.187540558 |
| DNAJC11 | 1.228082525 | 0.887290168 |
| MAF | 1.22793046 | 0.926670993 |

| | | |
|---|---|---|
| ATP1A2 | 1.22791663 | 0.809214796 |
| GRIN2A | 1.227814055 | 1.253082414 |
| N4BP2 | 1.22775577 | 1.092147956 |
| ARG2 | 1.227576058 | 0.918234912 |
| SLC7A3 | 1.227144355 | 0.970798183 |
| ZNF304 | 1.227118462 | 0.978533819 |
| MTHFS | 1.227112249 | 1.001297859 |
| CLIC3 | 1.227104511 | 0.896820247 |
| ULK4 | 1.227010663 | 0.988319273 |
| ELL3 | 1.226975884 | 1.017129154 |
| KLRK1 | 1.226885853 | 1.038711888 |
| EMR2 | 1.226836467 | 1.066839714 |
| ADAM29 | 1.226737761 | 1.053861129 |
| LYRM4 | 1.226666266 | 1.046172539 |
| KRBOX4 | 1.226610792 | 0.978938842 |
| PPM1F | 1.226426579 | 1.014925373 |
| AKAP6 | 1.226205541 | 0.988968202 |
| TCF7L2 | 1.226205541 | 0.988968202 |
| HIST1H4L | 1.225987319 | 0.986296677 |
| PPP1R14C | 1.225652254 | 0.981500514 |
| PLEKHA4 | 1.225562677 | 1.028434395 |
| ZNF500 | 1.225227817 | 0.929526124 |
| NAPEPLD | 1.225076396 | 1.120700844 |
| CCR1 | 1.225014571 | 0.950681376 |
| PTPRA | 1.22473105 | 1.003244646 |
| MRGPRX1 | 1.224685031 | 1.051265412 |
| RCE1 | 1.224551009 | 0.959766385 |
| C7orf31 | 1.224485487 | 0.854402193 |
| STXBP4 | 1.224267049 | 1.059952038 |
| LSR | 1.224062585 | 1.064892927 |
| FAM179A | 1.223887377 | 0.873228028 |
| WDR61 | 1.223819474 | 0.788983394 |
| OPTC | 1.223794208 | 1.006480356 |
| GGA3 | 1.22376008 | 0.967797191 |
| SLC27A4 | 1.22374989 | 0.974483597 |
| HSD17B6 | 1.223702566 | 0.869987849 |
| ASB16 | 1.223591367 | 1.091499027 |
| DHX38 | 1.223578607 | 0.995457495 |
| PNKD | 1.223487475 | 0.916937054 |
| GALNT3 | 1.223472011 | 1 |
| EFCAB2 | 1.223350977 | 1.049007695 |
| GIMAP1 | 1.223349296 | 0.946132037 |
| C20orf24 | 1.223301638 | 1.079821857 |
| FRYL | 1.223117468 | 1.044147428 |

| | | |
|---|---|---|
| PPP3R1 | 1.223052479 | 0.860480208 |
| GEMIN5 | 1.222691589 | 0.953408702 |
| OR2F2 | 1.222639891 | 0.992805755 |
| MRPL11 | 1.222589008 | 0.967454608 |
| LRRC26 | 1.222564257 | 0.992304577 |
| FAM171B | 1.222465223 | 1.128811237 |
| OR11G2 | 1.222450814 | 1.042480301 |
| SLC2A1 | 1.222347134 | 1.153796236 |
| MLLT1 | 1.222340713 | 0.939649578 |
| NHLRC1 | 1.222340713 | 0.939649578 |
| PTPLB | 1.222311817 | 1.022712524 |
| CFC1 | 1.222235684 | 1.036313806 |
| ATP6AP2 | 1.222079638 | 1.055498458 |
| PAGE2 | 1.221929492 | 1.040097205 |
| MIEF2 | 1.221901526 | 1.001027749 |
| AIDA | 1.221869382 | 1.068853787 |
| FAM111B | 1.221760472 | 1.02673147 |
| C11orf65 | 1.221669901 | 1.014985824 |
| DBX1 | 1.221612949 | 0.954232483 |
| KCNA6 | 1.221605319 | 0.988319273 |
| SMG7 | 1.22147103 | 0.981368975 |
| NUAK1 | 1.221278661 | 1.062946139 |
| TRIM22 | 1.22124392 | 0.844256976 |
| CEP164 | 1.221183422 | 0.956257594 |
| MTNR1B | 1.221162914 | 1.014925373 |
| PAGE2B | 1.22113997 | 0.985014176 |
| MIS18A | 1.221024785 | 1.037341555 |
| DENND2A | 1.220944003 | 1.069663832 |
| CDO1 | 1.220880277 | 0.833130822 |
| PLXNA4 | 1.220834939 | 1.124594419 |
| CHRNA6 | 1.220654733 | 1.129136924 |
| KIF2C | 1.220565165 | 1.028552888 |
| WIBG | 1.220539436 | 1.100445525 |
| C3orf33 | 1.220224167 | 0.987444309 |
| UPK3A | 1.220198746 | 0.918465228 |
| CCDC134 | 1.219602392 | 0.947347104 |
| GADD45G | 1.219391401 | 0.933160286 |
| MCL1 | 1.219391401 | 0.933160286 |
| NAP1L4 | 1.219184326 | 1.025516403 |
| DUSP7 | 1.218838271 | 1.034393251 |
| FMN2 | 1.218837102 | 1.036656389 |
| GNAI3 | 1.218735367 | 0.91174562 |
| SDK2 | 1.218713497 | 1.029119561 |
| ZNF121 | 1.218586902 | 0.881466256 |

| | | |
|---|---|---|
| SLC50A1 | 1.218585218 | 0.995544755 |
| DAGLB | 1.218582246 | 0.968851395 |
| SLC1A3 | 1.218426161 | 1.00843608 |
| MRPS21 | 1.218285367 | 1.052415211 |
| AP3M2 | 1.217867975 | 1.011990408 |
| FAM151B | 1.217775589 | 0.967454608 |
| SPRN | 1.217759776 | 0.903734156 |
| VAMP2 | 1.217739794 | 1.091127098 |
| OASL | 1.217661296 | 0.916937054 |
| FLYWCH2 | 1.217657467 | 1.023638232 |
| USP42 | 1.217418442 | 0.996106424 |
| RMND5A | 1.217320373 | 0.991494532 |
| WARS | 1.217230663 | 1.000648929 |
| CPLX2 | 1.217058972 | 0.975676602 |
| N6AMT2 | 1.217038267 | 0.940866748 |
| SEC16A | 1.217038109 | 1.0798183 |
| SOX9 | 1.216648556 | 1.071382219 |
| TCF19 | 1.216425377 | 1.058403634 |
| ZBTB7C | 1.216180689 | 1.124746861 |
| TMEM159 | 1.215865013 | 0.948562171 |
| ASIC5 | 1.215703485 | 1.116158339 |
| NECAB2 | 1.215700002 | 0.986981843 |
| REM1 | 1.215683054 | 1.079384366 |
| NCAN | 1.215399827 | 0.967598218 |
| PROM2 | 1.215271782 | 1.050616483 |
| CRIPT | 1.215257755 | 1.106925881 |
| YWHAZ | 1.215247661 | 1.226476314 |
| EPB41L4B | 1.215233358 | 1.067831449 |
| STH | 1.215182611 | 1.030832477 |
| APOBEC2 | 1.215104832 | 1.051032807 |
| GRIA1 | 1.214923637 | 1.099286178 |
| NYNRIN | 1.214901465 | 0.996574169 |
| IDS | 1.214435543 | 1.033744322 |
| PP2D1 | 1.214324571 | 0.937053861 |
| PLA2G4B | 1.214267131 | 1.032887975 |
| MS4A10 | 1.214226715 | 1.057211374 |
| PIR | 1.214220761 | 1.139519792 |
| ABLIM1 | 1.214077963 | 0.90647482 |
| LRRC16A | 1.21406061 | 0.944844125 |
| FAM75A2 | 1.214015477 | 0.936411503 |
| STARD10 | 1.213955265 | 1.089414183 |
| THG1L | 1.213941817 | 1.091130012 |
| SEMA3G | 1.213919403 | 1.021466181 |
| TFDP3 | 1.213849412 | 1.193195626 |

| | | |
|---|---|---|
| ZNF566 | 1.213832805 | 1.011340624 |
| VCAM1 | 1.213750537 | 1.10490077 |
| TMEM209 | 1.213592698 | 0.969510106 |
| KRT79 | 1.213541045 | 1.018156903 |
| NKPD1 | 1.213423829 | 0.981157931 |
| GPR137B | 1.213346077 | 1.043478261 |
| METTL16 | 1.213267522 | 0.967598218 |
| MRPS18C | 1.213122837 | 0.999314834 |
| NRBP1 | 1.213081408 | 1.092147956 |
| ARRDC4 | 1.212990442 | 1.092840014 |
| SLC9A7 | 1.212920983 | 1.064748201 |
| HAUS2 | 1.212861427 | 0.867557716 |
| FERD3L | 1.212486456 | 1.048197651 |
| MTFMT | 1.212472298 | 1.0266061 |
| PPP1R2 | 1.212468676 | 0.942894225 |
| CAMK4 | 1.212464478 | 0.859182349 |
| GIPR | 1.212418706 | 0.881245944 |
| KCNAB3 | 1.212375157 | 0.903309539 |
| SRSF8 | 1.21232681 | 0.918995545 |
| ALDH1A1 | 1.21212119 | 1.057754705 |
| MYO9A | 1.211982505 | 1.006075334 |
| ISG15 | 1.211874919 | 0.995949777 |
| KRT16 | 1.211646514 | 1.037684138 |
| ATP4A | 1.211602553 | 1.137573005 |
| KCNU1 | 1.211549373 | 1.031797534 |
| SPNS1 | 1.211334262 | 1.124594419 |
| TRIT1 | 1.211320843 | 1.146658014 |
| PALD1 | 1.211266263 | 0.996755354 |
| SHC4 | 1.211096859 | 1.057918186 |
| KLHDC8B | 1.210752045 | 0.978128797 |
| BTN2A3P | 1.210688352 | 0.935936965 |
| CHMP4B | 1.210651082 | 1.066828676 |
| METTL17 | 1.210647872 | 0.976508708 |
| PATE1 | 1.210632712 | 1.001370332 |
| YIPF4 | 1.210543445 | 0.974888619 |
| CYLC2 | 1.210500589 | 0.968139774 |
| ABO | 1.21016733 | 1.006489293 |
| LPA | 1.210082455 | 0.852044127 |
| C6orf203 | 1.210080857 | 1.024706359 |
| CACNG2 | 1.210000214 | 0.988968202 |
| RAD18 | 1.209749869 | 1.064243997 |
| KCNMB2 | 1.209731099 | 1.042180402 |
| MRGPRX2 | 1.209488164 | 1.095392602 |
| PHF2 | 1.209464619 | 1.073329007 |

| | | |
|---|---|---|
| MCHR2 | 1.209461162 | 0.936404932 |
| SHC3 | 1.209384607 | 1.102065614 |
| ZBTB25 | 1.209371108 | 1.125961928 |
| ZNF510 | 1.209330877 | 1.058924289 |
| SMOX | 1.209299808 | 0.940947437 |
| UBFD1 | 1.209110804 | 0.969218307 |
| TYMP | 1.208878873 | 1.0854597 |
| SEMA3C | 1.208789116 | 0.892669097 |
| CHD1L | 1.208690205 | 0.888384166 |
| IL31RA | 1.208676292 | 1.122647631 |
| UBE2R2 | 1.208623523 | 0.937053861 |
| MAOA | 1.208596191 | 1.078520441 |
| GPT2 | 1.208524159 | 0.892926671 |
| RNH1 | 1.208442265 | 1.022681247 |
| BHMT2 | 1.208310777 | 0.946138871 |
| TMEM86B | 1.208196529 | 0.947752126 |
| OAS3 | 1.208156585 | 0.950681376 |
| PRUNE | 1.208122309 | 0.89145403 |
| C22orf42 | 1.207901178 | 1.029971648 |
| CCDC67 | 1.20776797 | 0.925880923 |
| FOPNL | 1.207755591 | 1.019846092 |
| ZNF544 | 1.207489223 | 1.079821857 |
| PPME1 | 1.207475608 | 1.04412719 |
| RAB2A | 1.20746968 | 1.14146658 |
| C10orf67 | 1.207426062 | 1.122302158 |
| TTK | 1.207407362 | 1.022063595 |
| ZNF610 | 1.207216641 | 0.896197328 |
| FRAT1 | 1.207123017 | 1.252433485 |
| DUSP15 | 1.20705436 | 1.00908501 |
| PRKD1 | 1.206994675 | 1.03569111 |
| LPPR1 | 1.206938205 | 1.061563386 |
| FAM71F1 | 1.206837263 | 1.021240151 |
| PIP4K2C | 1.206832663 | 0.99156392 |
| UNC13A | 1.206737945 | 1.090784515 |
| OR6M1 | 1.206647054 | 1.029119561 |
| HSPB3 | 1.206370076 | 0.932853717 |
| SLFN5 | 1.206330492 | 1.058403634 |
| ZDHHC23 | 1.206183401 | 0.998629668 |
| AGER | 1.206072738 | 1.067488644 |
| GPRIN1 | 1.206047516 | 1.009315512 |
| ADCYAP1R1 | 1.20593888 | 0.970149254 |
| DEFB132 | 1.205830256 | 1.089914945 |
| ZNF182 | 1.205732553 | 1.019846092 |
| OR52K1 | 1.205663065 | 0.957862282 |

| | | |
|---|---|---|
| AHDC1 | 1.205484882 | 1.038882139 |
| R3HDM2 | 1.205439494 | 1.045767517 |
| ANKRD44 | 1.205405466 | 0.961927906 |
| NUMBL | 1.205194066 | 0.980130182 |
| RAB29 | 1.205122069 | 0.997404283 |
| BTBD1 | 1.205059496 | 0.971223022 |
| OR6N1 | 1.205058363 | 1.03562864 |
| UBOX5 | 1.204870818 | 0.926670993 |
| FSHB | 1.204605584 | 1.020554985 |
| CDC42EP1 | 1.204530506 | 1 |
| CTDNEP1 | 1.204472148 | 0.984583762 |
| ASCL1 | 1.20406578 | 0.980532122 |
| HM13 | 1.20406578 | 0.980532122 |
| TKTL2 | 1.204027582 | 0.927319922 |
| NR2F2 | 1.203984856 | 0.874107722 |
| WWC2 | 1.203829428 | 1.066018631 |
| CPA6 | 1.203783082 | 0.963011032 |
| PRR22 | 1.203778277 | 0.901336074 |
| MARCH10 | 1.203652764 | 0.967553537 |
| KRT20 | 1.203623375 | 0.989064399 |
| ZNF385A | 1.203590831 | 0.887290168 |
| LRCH1 | 1.203430979 | 0.922233642 |
| SDHAF2 | 1.203360371 | 0.992709599 |
| CRH | 1.203352696 | 1.22972096 |
| DVL2 | 1.203286577 | 1.166152792 |
| ARFGAP3 | 1.203269385 | 0.981181051 |
| TFE3 | 1.203144006 | 0.985723556 |
| DHDDS | 1.203004139 | 0.905905256 |
| PPP2R2B | 1.202989568 | 1.048020766 |
| SLC5A2 | 1.202911078 | 1.095990279 |
| FCGR1B | 1.202883231 | 1.04008222 |
| HDAC1 | 1.202792898 | 0.941596366 |
| PPM1G | 1.202743339 | 1.11486048 |
| PEX5L | 1.202671659 | 1.041312272 |
| ZNF772 | 1.202499685 | 0.900769542 |
| CDIPT | 1.202352386 | 0.986372485 |
| LILRA1 | 1.202237569 | 1.083247688 |
| EFNB3 | 1.202231783 | 0.99091499 |
| RINT1 | 1.202228021 | 1.034831916 |
| GJA10 | 1.202166078 | 1.120696638 |
| PPM1D | 1.202090104 | 0.93770279 |
| PLK2 | 1.201919119 | 0.9733939 |
| PCDH1 | 1.201914874 | 1.077081192 |
| CD58 | 1.201831248 | 1.086674767 |

| | | |
|---|---|---|
| ZC3HC1 | 1.201750406 | 0.999189955 |
| FAM98A | 1.201736805 | 1.042122317 |
| COPZ2 | 1.201665693 | 0.947752126 |
| IRF3 | 1.201595013 | 0.95587281 |
| CA5A | 1.201558005 | 0.98217902 |
| BIN2 | 1.201521379 | 0.953017416 |
| TIMM13 | 1.201500473 | 0.965056526 |
| TPSAB1 | 1.201500392 | 0.959902795 |
| ECSIT | 1.201488064 | 1.13163224 |
| OR8D1 | 1.201348539 | 1.100850547 |
| FCGR3A | 1.201323099 | 1.147306944 |
| ARHGEF2 | 1.201258722 | 0.93835172 |
| OTP | 1.201247192 | 0.900650908 |
| N4BP3 | 1.201177431 | 1.126771972 |
| GPR34 | 1.201053573 | 1.098637249 |
| CDKN2B | 1.200857232 | 1.076573653 |
| RFX1 | 1.200787586 | 0.925373134 |
| LYN | 1.200759102 | 1.049967554 |
| FCRLB | 1.200540577 | 0.996759822 |
| MFAP3 | 1.200455815 | 0.968482357 |
| USF1 | 1.200432362 | 0.970149254 |
| MFSD9 | 1.200409202 | 1.002025111 |
| PCK2 | 1.200332421 | 1.036988968 |
| FGFBP3 | 1.199846619 | 1.064398542 |
| RNF213 | 1.199789664 | 1.028552888 |
| TMEM242 | 1.1997589 | 0.832320778 |
| PRNT | 1.199724617 | 0.964763062 |
| SCFD2 | 1.199701908 | 0.967797191 |
| USP40 | 1.199685278 | 1.033095393 |
| SEPP1 | 1.1995566 | 0.987324426 |
| WDR64 | 1.19955244 | 1.011340624 |
| MYOT | 1.199551472 | 0.885783718 |
| SAMHD1 | 1.199429012 | 0.870392872 |
| SLC2A4 | 1.199411224 | 1.077871512 |
| CASD1 | 1.199373932 | 0.948734588 |
| USP3 | 1.19933417 | 0.917585983 |
| ANAPC16 | 1.199205315 | 1.097636177 |
| TPRG1 | 1.199195349 | 1.006480356 |
| LOC150786 | 1.199111388 | 1.052415211 |
| NCK1 | 1.198993352 | 0.931213498 |
| ABHD13 | 1.198984915 | 0.927501013 |
| CNTN1 | 1.19872259 | 0.975989617 |
| OR4D9 | 1.198638311 | 0.871531346 |
| PLB1 | 1.198617546 | 0.980532122 |

| | | |
|---|---|---|
| COX5A | 1.198536787 | 1.051914341 |
| ABCA4 | 1.198407046 | 1.092147956 |
| TLK1 | 1.198300272 | 0.891628812 |
| KPTN | 1.198230207 | 0.95991778 |
| MUM1 | 1.198142352 | 1.002740665 |
| EMR4P | 1.198007037 | 1.007787151 |
| LRTM1 | 1.197918788 | 0.900769542 |
| OR10R2 | 1.197736242 | 1.032887975 |
| RNF148 | 1.197724421 | 0.985723556 |
| LOH12CR1 | 1.197547893 | 1.045767517 |
| FGF23 | 1.197529974 | 0.959117456 |
| LENG9 | 1.197435069 | 1.013703323 |
| LAGE3 | 1.197427317 | 1.145808019 |
| PRKCA | 1.197331987 | 1.003893576 |
| ABHD4 | 1.197324577 | 0.932511356 |
| GABRA5 | 1.19707201 | 1.08890331 |
| SLC29A3 | 1.197059908 | 1.0532122 |
| WBSCR17 | 1.197033071 | 0.981829981 |
| C3orf14 | 1.196982151 | 0.829080599 |
| LCN12 | 1.19666344 | 0.888384166 |
| EMCN | 1.196653796 | 1.007879411 |
| SUMO3 | 1.196630325 | 1.032545392 |
| PIGB | 1.196588443 | 1.040233615 |
| FBXW7 | 1.196558477 | 0.892926671 |
| PCDH11X | 1.1965446 | 0.936622131 |
| PTGER3 | 1.196466193 | 1.00908501 |
| MDH1 | 1.196430959 | 0.9733939 |
| ACTG2 | 1.196340509 | 0.862292426 |
| C7orf62 | 1.196233459 | 1.032887975 |
| APCDD1 | 1.196095899 | 0.961630695 |
| LAMC2 | 1.19601984 | 1.085054678 |
| FAM78B | 1.195986729 | 0.978128797 |
| PKDREJ | 1.195929199 | 1.036340039 |
| OAZ1 | 1.195811559 | 1.081116158 |
| OR4C46 | 1.195740354 | 0.917780062 |
| OR8D2 | 1.195694365 | 0.881466256 |
| NEK4 | 1.195660496 | 0.933809215 |
| GGT1 | 1.195580366 | 0.963343611 |
| MBD4 | 1.195565495 | 0.93835172 |
| GNPTG | 1.195554247 | 1.078574322 |
| MPL | 1.195502682 | 1.059052563 |
| USP46 | 1.195412127 | 0.8669695 |
| AGO2 | 1.195404882 | 1.103828683 |
| PALM2 | 1.195396137 | 1.10243234 |

| | | |
|---|---|---|
| BCL2L14 | 1.195378374 | 0.947436729 |
| KLF17 | 1.195149476 | 1.092750101 |
| RNF175 | 1.195014588 | 0.925373134 |
| SIKE1 | 1.194969046 | 1.004860267 |
| TGS1 | 1.194929301 | 1.010382868 |
| PUS1 | 1.194758918 | 0.949777238 |
| ALPI | 1.194750577 | 0.979234263 |
| NDUFB10 | 1.194750577 | 0.979234263 |
| PORCN | 1.19464932 | 0.943543154 |
| MYO5B | 1.194560246 | 0.948085659 |
| CCDC107 | 1.19454828 | 0.960307817 |
| KCNC2 | 1.194455037 | 1.077871512 |
| RGS11 | 1.194421709 | 0.949982871 |
| MURC | 1.194239655 | 1.085303186 |
| NEK2 | 1.194177196 | 0.926022064 |
| FAM175A | 1.194089768 | 1.078169299 |
| MSTO1 | 1.194041069 | 0.986634265 |
| TMEM126A | 1.19397179 | 1.050627785 |
| MFN2 | 1.193713634 | 0.993510707 |
| TMPRSS13 | 1.193686796 | 1.16807268 |
| AGBL1 | 1.193525962 | 1.173754557 |
| CCDC43 | 1.193521275 | 1.076954232 |
| MSH3 | 1.193443284 | 1.096690461 |
| CACNA2D1 | 1.193090282 | 0.985074627 |
| POLD3 | 1.193090282 | 1.029850746 |
| CLINT1 | 1.193083292 | 1.013365735 |
| CEND1 | 1.193045596 | 1.006480356 |
| TSEN15 | 1.193041941 | 1.074683111 |
| SYNGAP1 | 1.193030212 | 1.025516403 |
| FBXO48 | 1.192941195 | 1.038935756 |
| APPL2 | 1.192902862 | 1.151883354 |
| IFLTD1 | 1.192766648 | 0.958282706 |
| NHP2L1 | 1.192665464 | 1.062007537 |
| OR5W2 | 1.192604885 | 0.991778006 |
| SLC26A9 | 1.192579714 | 1.06164828 |
| PLA2G12A | 1.192399014 | 1.120051914 |
| IER5 | 1.192194153 | 0.998629668 |
| PJA2 | 1.192083058 | 0.999351071 |
| EMID1 | 1.192003853 | 0.923261391 |
| FMR1 | 1.191995974 | 0.993519644 |
| ACAA1 | 1.191955069 | 1.005670312 |
| RBL2 | 1.191951258 | 0.815704088 |
| AKAP4 | 1.191796737 | 1.01752109 |
| SYT5 | 1.191755375 | 1.084960603 |

| | | |
|---|---|---|
| RCBTB2 | 1.191710709 | 1.040502228 |
| LIPG | 1.191709792 | 0.952207371 |
| GZMH | 1.191708569 | 1.120700844 |
| CDC42SE2 | 1.191588542 | 1.031148605 |
| GPBP1L1 | 1.191460898 | 0.987039287 |
| PLEKHH2 | 1.191342819 | 1.054678007 |
| NMNAT2 | 1.191309549 | 1 |
| GPR174 | 1.1912986 | 0.950681376 |
| IARS | 1.191134365 | 0.910447761 |
| KRT26 | 1.191127444 | 0.926690968 |
| C5 | 1.19087655 | 1.047961631 |
| GRPEL1 | 1.190750816 | 1.016786571 |
| ACADM | 1.190678047 | 0.834523037 |
| MTSS1 | 1.19052164 | 1.067831449 |
| ANKRD55 | 1.19049385 | 0.764277035 |
| AVPR1B | 1.190417329 | 0.95587281 |
| LSM14B | 1.190361609 | 1.017415958 |
| HEPN1 | 1.190283357 | 0.984583762 |
| ECE2 | 1.190149203 | 0.974042829 |
| NKX2-5 | 1.190090671 | 1.032446463 |
| PAPSS1 | 1.190038313 | 1.090850097 |
| PFN3 | 1.190004162 | 1.031591738 |
| PRKACB | 1.189847735 | 1.050616483 |
| MYH8 | 1.18980575 | 0.942894225 |
| PIPOX | 1.189752899 | 1.113562622 |
| IFIT1B | 1.189748659 | 0.939362795 |
| ZNF444 | 1.189625009 | 1.016443988 |
| CDC25B | 1.189605375 | 0.844256976 |
| STEAP3 | 1.189306707 | 0.920830629 |
| LY96 | 1.189298327 | 0.988009592 |
| FAM153B | 1.189296106 | 0.960602946 |
| PASK | 1.189181328 | 0.871512005 |
| VASN | 1.189169558 | 1.094775213 |
| SLC5A3 | 1.189166451 | 0.978533819 |
| KCNA2 | 1.189113329 | 1.051265412 |
| GPR22 | 1.18904105 | 0.822193381 |
| UTY | 1.188941513 | 0.980472765 |
| COX18 | 1.188908948 | 1.127987039 |
| BBS4 | 1.188857328 | 0.933538883 |
| AVEN | 1.188764197 | 1.020116807 |
| C10orf90 | 1.188748268 | 1.018499486 |
| SYNPO2L | 1.188742197 | 1.039397054 |
| KCTD4 | 1.188507513 | 0.979883193 |
| B3GNT8 | 1.188450051 | 0.984425698 |

| | | |
|---|---|---|
| RSL1D1 | 1.188333782 | 1.017415958 |
| CD40 | 1.188296601 | 0.872160934 |
| CDK5RAP2 | 1.188230062 | 1.04898938 |
| RASA2 | 1.188228931 | 0.939649578 |
| LILRA6 | 1.188177549 | 0.967193196 |
| F11 | 1.18811903 | 1.074626866 |
| GALR2 | 1.188115819 | 1.011680727 |
| C9orf3 | 1.187985666 | 0.894873459 |
| ATP7A | 1.187978915 | 1.151200519 |
| ACSM3 | 1.187941376 | 0.962362103 |
| NEIL3 | 1.187924676 | 1.068859198 |
| OSMR | 1.187885479 | 0.966904607 |
| MMGT1 | 1.187518026 | 1.139732685 |
| CRIP2 | 1.187391797 | 0.96435804 |
| RHOBTB3 | 1.187121272 | 1.102530824 |
| C1orf27 | 1.186955545 | 0.959497772 |
| TPGS2 | 1.186917849 | 1.060348319 |
| BAIAP2L1 | 1.186845871 | 1.095180235 |
| TMEM18 | 1.186833711 | 0.961288112 |
| PPAP2C | 1.186822274 | 1.06229721 |
| ADAM19 | 1.186730895 | 1.071382219 |
| CC2D2A | 1.186605532 | 0.994176088 |
| MAGEA11 | 1.186569239 | 1.002055498 |
| RCAN2 | 1.186514732 | 0.894826996 |
| CLPTM1L | 1.186469932 | 1.031186715 |
| UBE2L3 | 1.186423006 | 1.103179753 |
| ARHGEF15 | 1.186414824 | 0.976019185 |
| MMP23B | 1.186335034 | 1.071599863 |
| DNTTIP1 | 1.186245271 | 0.972744971 |
| TRIM10 | 1.186091985 | 1.139519792 |
| GRIN2B | 1.186055022 | 1.067488644 |
| BAG2 | 1.185704078 | 1.027255029 |
| BEND5 | 1.185582985 | 1.145597807 |
| CD248 | 1.185509812 | 1.066828676 |
| HAT1 | 1.185454474 | 1.054510058 |
| OGDHL | 1.185256157 | 1.077222583 |
| KREMEN1 | 1.185108458 | 1.081877355 |
| C6orf10 | 1.185067588 | 1.008564577 |
| C8orf22 | 1.185045387 | 0.949640288 |
| SLC6A9 | 1.185042281 | 0.933160286 |
| FAM49A | 1.184879325 | 1.070878898 |
| DNLZ | 1.184627179 | 0.972250771 |
| KCNE3 | 1.184430126 | 1.001297859 |
| TMEM174 | 1.184400233 | 1.128797084 |

| | | |
|---|---|---|
| VRK3 | 1.184380979 | 0.915639195 |
| LOC389834 | 1.184312459 | 1.045906132 |
| SORBS2 | 1.184290508 | 1.052247874 |
| ZNF408 | 1.184277978 | 1.061664954 |
| OTUD6A | 1.184253525 | 1.074929121 |
| FAM65C | 1.184211509 | 1.063035286 |
| CCT2 | 1.184193772 | 0.935601458 |
| WWC1 | 1.184176758 | 1.102875658 |
| EID2 | 1.184172042 | 1.033095393 |
| KCNN4 | 1.184101244 | 1.042180402 |
| FSIP1 | 1.183953108 | 0.962315862 |
| APLNR | 1.183949687 | 0.965606749 |
| TLN2 | 1.183849893 | 0.979445015 |
| CDC37 | 1.183848143 | 1.070068854 |
| B4GALNT1 | 1.183700824 | 1.096690461 |
| TRIM74 | 1.183576105 | 1.11486048 |
| TAS2R13 | 1.183509346 | 0.932511134 |
| TCEAL6 | 1.183478369 | 1.059943297 |
| GAD2 | 1.183256066 | 1.028756582 |
| DTWD2 | 1.183216791 | 1.034021871 |
| GPR20 | 1.183177989 | 1.070084361 |
| AMIGO3 | 1.183176592 | 1.023295649 |
| ILKAP | 1.183147859 | 1.074626866 |
| ACADSB | 1.183117893 | 1.079169371 |
| CD8A | 1.183100653 | 1.032545392 |
| ZMYND8 | 1.18300083 | 1.09733939 |
| CALCRL | 1.182959504 | 0.993510707 |
| MMP15 | 1.182959504 | 0.993510707 |
| CCT6A | 1.182887553 | 1.115509409 |
| PCDH11Y | 1.182669034 | 0.893799246 |
| WBP1 | 1.182507279 | 0.912298732 |
| SUV420H1 | 1.182390238 | 0.962362103 |
| DCAF11 | 1.182314804 | 0.829736211 |
| FUCA2 | 1.182224933 | 1.111615834 |
| RFNG | 1.182077081 | 1.012329656 |
| BROX | 1.18207079 | 1.021061158 |
| TCEB3B | 1.181970719 | 1.030499676 |
| RPGRIP1L | 1.181874823 | 1.052652896 |
| SPOPL | 1.181844671 | 0.932511134 |
| C11orf74 | 1.181843342 | 1.057918186 |
| KIR3DX1 | 1.181636117 | 0.937626569 |
| PRR5 | 1.181636117 | 0.937626569 |
| ATP12A | 1.181535229 | 1.112264763 |
| ITGA2B | 1.18151613 | 0.976638546 |

| | | |
|---|---|---|
| OFD1 | 1.18151613 | 0.976638546 |
| USP15 | 1.181440805 | 0.990266061 |
| MMP3 | 1.181347615 | 0.863724854 |
| HVCN1 | 1.18131979 | 1.157689812 |
| GABRA4 | 1.181249897 | 1.0266061 |
| HDAC8 | 1.181181651 | 1.040233615 |
| GSTO1 | 1.18105051 | 1.067488644 |
| CLRN1 | 1.181043785 | 1.009720535 |
| CACNA2D3 | 1.181008278 | 1.076573653 |
| VAPB | 1.180998198 | 1.021240151 |
| LCE1C | 1.180985466 | 0.941075711 |
| C2 | 1.180983993 | 0.927319922 |
| ACO1 | 1.180959955 | 0.931862427 |
| HES7 | 1.180912537 | 0.940947437 |
| DCC | 1.18083549 | 0.929526124 |
| TSC22D3 | 1.180829174 | 0.990684488 |
| GIT1 | 1.180786692 | 1.066118534 |
| LCE1B | 1.180761319 | 1.099006509 |
| FAM120B | 1.180702944 | 0.922640745 |
| NIPAL1 | 1.180656741 | 0.899959498 |
| KIAA1755 | 1.180650119 | 0.994329688 |
| TACR1 | 1.180543399 | 1.018170019 |
| SYNE3 | 1.18048561 | 1.032202809 |
| FN1 | 1.180325737 | 0.891628812 |
| PARS2 | 1.180321198 | 1.095990279 |
| XRCC4 | 1.180321198 | 0.929931146 |
| ARID5B | 1.180293232 | 0.901984609 |
| CCDC62 | 1.18021891 | 0.984204131 |
| NDUFB5 | 1.180118282 | 0.937053861 |
| EVC | 1.18002164 | 0.960260363 |
| EPB41L4A | 1.179970169 | 1.147023086 |
| BLNK | 1.179952679 | 1.060294621 |
| LACC1 | 1.179936855 | 1.047387606 |
| FAM124B | 1.179924221 | 1.01417578 |
| C4orf3 | 1.179914797 | 1.052652896 |
| MAST3 | 1.17987691 | 0.996106424 |
| SPINT2 | 1.179872364 | 0.953017416 |
| CSF3R | 1.179808253 | 1.014276444 |
| RAPSN | 1.179791459 | 1.018818949 |
| PPAPDC3 | 1.179650851 | 0.846852693 |
| C1QC | 1.179603095 | 1.016200891 |
| TGIF2LY | 1.17947081 | 1.114211551 |
| NUP210L | 1.179465913 | 0.970838396 |
| ABCA7 | 1.179450064 | 0.896820247 |

| | | |
|---|---|---|
| TSPYL1 | 1.179425502 | 0.947584789 |
| HIST1H3J | 1.179379784 | 0.956834532 |
| MAFF | 1.17934963 | 0.924075276 |
| P2RY1 | 1.179333412 | 0.928617781 |
| PPBP | 1.179317442 | 0.933160286 |
| SLA2 | 1.179290841 | 1.000342583 |
| KIAA0226L | 1.179267009 | 0.892428914 |
| POLR2L | 1.179122747 | 0.992212849 |
| CLDN18 | 1.179077479 | 0.996574169 |
| OTUD5 | 1.179048163 | 0.963686194 |
| TMEM9B | 1.178990335 | 1.05872823 |
| LOC100132832 | 1.17877395 | 1.028434395 |
| RNF115 | 1.178751222 | 1.128487995 |
| FRMD1 | 1.178694476 | 0.962658445 |
| CD244 | 1.178687088 | 0.908464966 |
| ADAMTS9 | 1.178417155 | 0.965606749 |
| TIGIT | 1.178379868 | 1.062778453 |
| PRRT1 | 1.178331475 | 1.024301337 |
| ZFP90 | 1.178292444 | 1.0222679 |
| GMPR2 | 1.178264794 | 0.99108951 |
| FNDC3A | 1.178222157 | 1.006885379 |
| KRT15 | 1.178106177 | 1.094895512 |
| LIFR | 1.178098564 | 1.115509409 |
| PGBD2 | 1.178049494 | 0.890715999 |
| ERICH3 | 1.178015187 | 1.091130012 |
| MXRA5 | 1.177991602 | 0.972053463 |
| ITSN1 | 1.177987103 | 0.994861254 |
| ZNF410 | 1.17770323 | 1.028351559 |
| ABI3 | 1.17759658 | 0.891058582 |
| SLC38A4 | 1.177534814 | 1.025308241 |
| GOLGA2P5 | 1.177413889 | 0.848116646 |
| GCOM1 | 1.177251237 | 1.114621304 |
| PRR25 | 1.177169038 | 0.952038369 |
| AQP11 | 1.177024727 | 1.060294621 |
| SMEK2 | 1.176887066 | 1.062373431 |
| RNF26 | 1.176817285 | 0.989617132 |
| PPP1R16A | 1.176813745 | 0.994159637 |
| EPHB4 | 1.176799915 | 1.012329656 |
| NR0B2 | 1.176793247 | 1.021414666 |
| LRIT3 | 1.176709463 | 1.112024666 |
| LRP5 | 1.176604526 | 1.052757794 |
| PRPF4 | 1.176484682 | 1.091127098 |
| ARL14 | 1.176465995 | 1.031186715 |
| GSTT2B | 1.176418802 | 0.932853717 |

| | | |
|---|---|---|
| TOX | 1.176292977 | 1.078108941 |
| TTC7A | 1.176267888 | 0.875253139 |
| NFX1 | 1.176147669 | 1.049007695 |
| DHRS4L2 | 1.176086592 | 0.948955122 |
| BMP7 | 1.176046081 | 1.212848799 |
| SHPK | 1.176046081 | 0.949383517 |
| SPR | 1.176046081 | 0.985723556 |
| SUB1 | 1.176046081 | 0.963011032 |
| TRIM65 | 1.176046081 | 0.85853342 |
| TSN | 1.176009611 | 1.00445358 |
| GHDC | 1.175990366 | 1.063183475 |
| FAM3B | 1.175868925 | 1.12470024 |
| KRT27 | 1.175831535 | 1.098420413 |
| GNRH1 | 1.175564037 | 0.92428914 |
| PODN | 1.175433364 | 0.854402193 |
| STYXL1 | 1.175352343 | 1.099935107 |
| FEM1B | 1.175337608 | 1.077222583 |
| SMARCAL1 | 1.1753097 | 1.036340039 |
| MCM9 | 1.175247607 | 1.076144188 |
| MUS81 | 1.175242771 | 0.950032446 |
| GYPA | 1.175227588 | 1.007695423 |
| HSPH1 | 1.175215028 | 0.918234912 |
| ITGB8 | 1.175185145 | 0.886437378 |
| CYP3A7 | 1.175122585 | 1.07125728 |
| FAM83A | 1.175081339 | 1.032401782 |
| KIAA2018 | 1.174991323 | 0.96437136 |
| MIA3 | 1.174807593 | 1.005481329 |
| TAC1 | 1.174751409 | 0.988352175 |
| LMLN | 1.174513799 | 0.996106424 |
| ALPPL2 | 1.174499722 | 0.987021415 |
| ALLC | 1.174425137 | 0.941596366 |
| ATF6 | 1.174401345 | 0.927968851 |
| OLFM4 | 1.174340108 | 1.055893074 |
| LRRC43 | 1.174229352 | 0.991435423 |
| TCHP | 1.174042414 | 1.057918186 |
| CAPS | 1.173990208 | 1.063035286 |
| RAB22A | 1.17394972 | 1.092147956 |
| SCMH1 | 1.173940994 | 1.087605451 |
| EID1 | 1.173938649 | 0.989469421 |
| GALR1 | 1.173616229 | 0.942245295 |
| MFSD2B | 1.173592843 | 0.940390545 |
| ANXA6 | 1.173533114 | 1.02673147 |
| MFSD7 | 1.173522671 | 1.021466181 |
| ANP32C | 1.173485957 | 0.904761905 |

| | | |
|---|---|---|
| ATP11A | 1.173342504 | 0.846852693 |
| PGPEP1 | 1.173264215 | 1.09733939 |
| ASB11 | 1.173168967 | 1.060999351 |
| IFNG | 1.173079975 | 1.029201817 |
| ARHGAP15 | 1.173031728 | 0.934224049 |
| UBA6 | 1.173026712 | 1.011031798 |
| RPS6KA6 | 1.172899523 | 0.970149254 |
| ANK3 | 1.172854657 | 0.956521739 |
| NDUFA13 | 1.172798964 | 0.983394087 |
| OR11H1 | 1.17276857 | 1.105173004 |
| SNX25 | 1.172683432 | 1.110166059 |
| MBLAC1 | 1.172650675 | 1.069544365 |
| RPP25L | 1.172650675 | 1.069544365 |
| CDH22 | 1.172627839 | 1.002055498 |
| ZNF135 | 1.172608582 | 0.992805755 |
| EMILIN1 | 1.172605229 | 0.963686194 |
| ITPA | 1.172585048 | 1.102530824 |
| RAB8B | 1.172432653 | 1.071688943 |
| SERPINF2 | 1.17206233 | 0.917780062 |
| BRI3 | 1.171972617 | 0.929770469 |
| LIMS2 | 1.171948512 | 0.900650908 |
| NBR1 | 1.171936547 | 1.056298096 |
| FAM84A | 1.171799263 | 1.059943297 |
| FZD8 | 1.171717114 | 1.057754705 |
| CRYBA1 | 1.171696591 | 1.035971223 |
| UBP1 | 1.171648019 | 1.057211374 |
| MYO1F | 1.171593738 | 1.038711888 |
| ERMARD | 1.171490477 | 0.951353203 |
| TMEM184A | 1.171490477 | 0.951353203 |
| C2orf15 | 1.171487356 | 1.056703119 |
| FKSG2_pseudogene | 1.171462838 | 1.032202809 |
| CYP11B1 | 1.171419022 | 0.989617132 |
| SPATA31E1 | 1.171410959 | 0.963343611 |
| TOX4 | 1.171373964 | 0.989874443 |
| ZNF287 | 1.171258764 | 1.088043851 |
| ZNHIT6 | 1.171222977 | 1.028756582 |
| RAB3IL1 | 1.17118799 | 0.918465228 |
| BCAT1 | 1.171183345 | 0.982989064 |
| PROX1 | 1.17106709 | 0.766385464 |
| ANKRD20A1 | 1.171038573 | 1.021871203 |
| ABCF3 | 1.171037454 | 0.761842959 |
| ZNF683 | 1.17096522 | 0.970838396 |
| PAX1 | 1.170910511 | 1.040233615 |
| THAP11 | 1.170903945 | 0.955806783 |

| | | |
|---|---|---|
| EMC1 | 1.170756734 | 0.949297705 |
| SARS2 | 1.170700422 | 0.999351071 |
| JPH4 | 1.170622799 | 0.932168551 |
| TCEAL2 | 1.17061363 | 0.837181045 |
| HSPD1 | 1.17060147 | 0.981181051 |
| C11orf72_pseudogene | 1.170540277 | 1.046982584 |
| ORAI1 | 1.170498648 | 0.963011032 |
| PTP4A3 | 1.170449254 | 1.090850097 |
| KATNA1 | 1.170419042 | 0.949383517 |
| OR2T10 | 1.170401325 | 1.071599863 |
| SCNN1A | 1.170378374 | 1.077222583 |
| OR2L3 | 1.170376496 | 0.958890031 |
| DCAF10 | 1.170368455 | 0.920210612 |
| RUSC1-AS1 | 1.1703546 | 1.040097205 |
| LRRC4 | 1.170308123 | 1.022610483 |
| STX1B | 1.170305765 | 1.063595068 |
| ACYP1 | 1.170231262 | 1.049967554 |
| CHDC2 | 1.170096231 | 1.107330903 |
| C5AR1 | 1.17007633 | 0.894873459 |
| PPP4R1 | 1.170049657 | 1.018170019 |
| KCNMB4 | 1.17002883 | 1.14146658 |
| SLC7A14 | 1.17001275 | 1.093182597 |
| HDHD2 | 1.169905868 | 0.816524909 |
| TPT1 | 1.169612984 | 1.052415211 |
| ALDH4A1 | 1.16958873 | 0.945489942 |
| MAB21L3 | 1.169514138 | 1.02673147 |
| FAM151A | 1.169507617 | 1.130822195 |
| PLCXD1 | 1.169480145 | 1.046073978 |
| LOC553158 | 1.169388825 | 0.876873228 |
| ZCCHC13 | 1.169367837 | 0.950992305 |
| PPP6R3 | 1.169249656 | 1.034021871 |
| GPR111 | 1.169228483 | 0.895522388 |
| IRX6 | 1.169043261 | 1.058239123 |
| HDGFRP2 | 1.169024042 | 1.071283921 |
| TNFSF8 | 1.168841497 | 0.913669065 |
| CACNA1C | 1.168755042 | 1.046722907 |
| MAP4K4 | 1.168716186 | 0.937053861 |
| DIDO1 | 1.168714581 | 0.836776023 |
| C5orf15 | 1.168706339 | 0.960260363 |
| POLR3D | 1.168658889 | 1.033095393 |
| PDX1 | 1.168651568 | 1.013365735 |
| KCNJ9 | 1.168593359 | 1.024010383 |
| PECR | 1.168560101 | 1.019467878 |
| CLN5 | 1.168455187 | 1.032202809 |

| | | |
|---|---|---|
| TNFAIP6 | 1.168419846 | 0.989722508 |
| LRRC70 | 1.168370504 | 0.99109284 |
| PCDH8 | 1.168321298 | 0.992463172 |
| SPAG17 | 1.168131723 | 1.027946537 |
| ZNF782 | 1.167925161 | 0.708383961 |
| PSTK | 1.167889414 | 1.029201817 |
| PLEKHO2 | 1.167875596 | 0.952207371 |
| CATSPER1 | 1.167737116 | 1.194029851 |
| LRBA | 1.167688277 | 0.943816375 |
| MBNL3 | 1.167688277 | 0.943816375 |
| LMF1 | 1.167683144 | 0.848116646 |
| DLX2 | 1.167629437 | 0.997404283 |
| XRCC1 | 1.167621698 | 0.905905256 |
| NRSN1 | 1.167552177 | 0.993114621 |
| SLC22A11 | 1.167545656 | 1.03199676 |
| WDR63 | 1.167539645 | 1.070878898 |
| METTL23 | 1.167458422 | 1.017129154 |
| SYN3 | 1.167419591 | 0.995889003 |
| BNIP2 | 1.167405106 | 0.883192732 |
| DYSF | 1.167264633 | 0.944158959 |
| TXNIP | 1.167263142 | 1.111786148 |
| GTF3C2 | 1.167192547 | 1.034393251 |
| PABPC4 | 1.167142057 | 0.906844876 |
| TPK1 | 1.166952987 | 1.007138222 |
| FRMD8 | 1.166896311 | 1.113394998 |
| TBCD | 1.166881993 | 0.897910243 |
| ITGA4 | 1.166870252 | 0.998053212 |
| EFHC2 | 1.166831322 | 1.020251114 |
| MB21D1 | 1.166810177 | 1.013703323 |
| C4orf29 | 1.166756695 | 1.174159579 |
| FRMD5 | 1.166687263 | 0.931956258 |
| KCNQ3 | 1.166682637 | 0.815055159 |
| INCA1 | 1.166682585 | 0.983213429 |
| C12orf76 | 1.166634195 | 0.927372388 |
| DMRTC1B | 1.166526539 | 0.872901679 |
| EBI3 | 1.166498892 | 0.879299156 |
| PSMD10 | 1.166456199 | 0.978128797 |
| HIATL1 | 1.166284989 | 0.942891859 |
| SLIT3 | 1.166202893 | 1.00890716 |
| TRIM47 | 1.165994415 | 1.062946139 |
| DUSP10 | 1.165951278 | 0.907203115 |
| ELP4 | 1.165803715 | 0.950668037 |
| HYDIN | 1.165732307 | 0.988009592 |
| STX1A | 1.165679106 | 0.884973674 |

| | | |
|---|---|---|
| ZNF154 | 1.165415049 | 0.855087359 |
| CHGB | 1.165383695 | 0.95101062 |
| LGI1 | 1.165315414 | 1.048304214 |
| ETV6 | 1.165285695 | 1.205710578 |
| SIPA1L1 | 1.165246815 | 0.931140802 |
| NANOS2 | 1.165243466 | 0.986229243 |
| GTPBP8 | 1.165230833 | 1.000405022 |
| LYZ | 1.16520403 | 1.196625568 |
| KCTD2 | 1.165079146 | 1.182998053 |
| GZMM | 1.164979749 | 0.931551235 |
| MAGEF1 | 1.164943428 | 0.988254354 |
| CHRM1 | 1.164792084 | 0.949383517 |
| PTAFR | 1.164779654 | 0.880597015 |
| PLA2G4A | 1.164752405 | 1.013627515 |
| AMPD1 | 1.164479465 | 0.889428919 |
| L3MBTL3 | 1.164473454 | 0.931956258 |
| EVA1A | 1.164462859 | 1.017010936 |
| CRELD1 | 1.164346919 | 1.034831916 |
| PHLPP2 | 1.164277811 | 0.972744971 |
| C6orf1 | 1.164221402 | 0.939246659 |
| UBE2W | 1.164219115 | 1.032446463 |
| CRIP3 | 1.164132308 | 1.098825435 |
| ZDHHC4 | 1.164080546 | 1.04008222 |
| SPEM1 | 1.164077394 | 1.093560146 |
| SEZ6L | 1.164058648 | 1.022681247 |
| DNAH9 | 1.164014167 | 1.017471737 |
| NECAB3 | 1.164008051 | 0.930798219 |
| HTR4 | 1.163938557 | 0.945489942 |
| CAPN15 | 1.163880108 | 0.940947437 |
| CBX7 | 1.16386282 | 1.064892927 |
| TADA1 | 1.163857944 | 0.96435804 |
| C20orf96 | 1.163796228 | 0.949640288 |
| CCT6B | 1.163787576 | 0.996106424 |
| FBXL18 | 1.163674794 | 0.987021415 |
| ZNF22 | 1.163670811 | 0.928400137 |
| TRAPPC3 | 1.163586626 | 0.993833505 |
| SMIM8 | 1.163539459 | 0.907159986 |
| RBM12B | 1.163461222 | 0.998379911 |
| FAM213B | 1.163324244 | 0.922235723 |
| SULT1A3 | 1.163263013 | 0.895522388 |
| TRRAP | 1.163201929 | 0.950681376 |
| CACNA1I | 1.163147513 | 1.005840363 |
| IQSEC2 | 1.163103011 | 0.995139733 |
| ANXA13 | 1.163091211 | 1.03562864 |

| | | |
|---|---|---|
| ABCD2 | 1.163089064 | 1.001297859 |
| ECHDC1 | 1.163036839 | 0.989874443 |
| ASMTL | 1.162887619 | 0.927968851 |
| AVPR2 | 1.162792453 | 0.863724854 |
| CEP76 | 1.162787691 | 0.982989064 |
| ARMC3 | 1.162786163 | 1.057513163 |
| LECT1 | 1.162768335 | 0.930455635 |
| HSP90AA1 | 1.162757384 | 0.918883842 |
| HSP90B1 | 1.162601052 | 0.96495782 |
| CISD2 | 1.162543637 | 1.013365735 |
| ASB6 | 1.162519716 | 1.015574302 |
| FAM50A | 1.162508488 | 0.961927906 |
| FOXG1 | 1.162503746 | 1.07073329 |
| MYT1L | 1.162503746 | 1.07073329 |
| SPTSSB | 1.162443193 | 0.966769442 |
| NR1D2 | 1.16243443 | 1.121349773 |
| BCO2 | 1.162420682 | 0.896171317 |
| SYT8 | 1.162400602 | 1.033573141 |
| TAS2R20 | 1.162355362 | 1.095180235 |
| TRIM58 | 1.162264267 | 1.162881246 |
| DNAJC9 | 1.162204276 | 0.948269955 |
| GLIPR1L2 | 1.162187616 | 1.010125557 |
| G2E3 | 1.162150085 | 1.043478261 |
| C15orf54 | 1.162132369 | 1.029804728 |
| IGSF8 | 1.162127838 | 1.043165468 |
| LY6G5B | 1.162101513 | 1.00445358 |
| STRA6 | 1.16201391 | 0.985419198 |
| CD28 | 1.162008867 | 1.008505468 |
| ZWILCH | 1.161955707 | 0.929770469 |
| C15orf26 | 1.161859918 | 1.090319968 |
| C7orf55 | 1.161831876 | 0.840356286 |
| USP43 | 1.16168463 | 0.956521739 |
| FAM60A | 1.161681933 | 0.97326853 |
| CHST10 | 1.161677797 | 1.06229721 |
| ACTBL2 | 1.161661607 | 1.097610369 |
| PLCL1 | 1.161616137 | 0.740428293 |
| CA5B | 1.161579503 | 0.896820247 |
| FOXQ1 | 1.16150909 | 1.011647825 |
| SCN3B | 1.161451161 | 0.993510707 |
| FAM168A | 1.161447968 | 0.946214457 |
| PAFAH1B1 | 1.161428505 | 1.09072499 |
| SERTAD4 | 1.161265959 | 1.074683111 |
| SMAD9 | 1.161263382 | 0.92926671 |
| INPP5E | 1.161234404 | 1.030499676 |

| | | |
|---|---|---|
| TOP1 | 1.161234404 | 0.772874757 |
| MYL12B | 1.161229666 | 1.076053443 |
| STAC | 1.161202433 | 0.918590522 |
| PROSC | 1.161153174 | 0.904414743 |
| CNOT10 | 1.161151054 | 0.942446043 |
| TEX33 | 1.161094592 | 1.087484812 |
| FXR1 | 1.161021151 | 1.042822885 |
| LBX1 | 1.160968582 | 0.961713173 |
| NT5C3A | 1.160968582 | 1.113562622 |
| TSHR | 1.160968582 | 1.012329656 |
| IL4R | 1.160915589 | 1.030781693 |
| HMG20A | 1.160903876 | 1.058403634 |
| SEC61A1 | 1.160901987 | 0.992120589 |
| PDE3A | 1.160900666 | 1.007787151 |
| PLSCR5 | 1.160830111 | 1.089509923 |
| WNT16 | 1.160824372 | 0.953408702 |
| SLC35A5 | 1.160797514 | 1.201168073 |
| TST | 1.160751907 | 0.948085659 |
| PEBP4 | 1.160712541 | 1.038072094 |
| LASP1 | 1.1606792 | 0.972250771 |
| OGFOD2 | 1.160636182 | 1.000810045 |
| FOLR4 | 1.160608878 | 0.977723775 |
| HIST1H1T | 1.160580198 | 0.954637505 |
| FPGT | 1.160453103 | 0.999189955 |
| ASRGL1 | 1.16039717 | 0.985014176 |
| ATXN7L1 | 1.16021888 | 0.942486837 |
| NDST4 | 1.160192521 | 1.059052563 |
| CUBN | 1.160177456 | 0.865671642 |
| KRTAP10-7 | 1.160140483 | 1.051032807 |
| FOXE1 | 1.160128309 | 0.815055159 |
| ESRRA | 1.160085366 | 0.995949777 |
| FBN1 | 1.16006308 | 0.994176088 |
| CDY | 1.159982185 | 0.90266061 |
| NSUN2 | 1.159937935 | 1.013018157 |
| ABCB10 | 1.159928164 | 0.853786958 |
| MOB3C | 1.159925656 | 1.027749229 |
| SPIDR | 1.159807589 | 1.038882139 |
| WDR54 | 1.159697507 | 1.001620089 |
| UCMA | 1.159663122 | 1.019441069 |
| PGBD1 | 1.159582073 | 0.867420349 |
| F2RL1 | 1.159559455 | 0.972096042 |
| NOX3 | 1.159559455 | 0.972096042 |
| TUBB2A | 1.159555916 | 1.018170019 |
| LRP6 | 1.159482072 | 0.921479559 |

| | | |
|---|---|---|
| PDZD11 | 1.159466748 | 1.02673147 |
| LCE6A | 1.159453575 | 0.97327852 |
| ATP6V1D | 1.159403947 | 0.963011032 |
| CDH6 | 1.159387786 | 0.94655704 |
| MGME1 | 1.159349227 | 0.947927372 |
| ZNF226 | 1.159349227 | 0.947927372 |
| BAIAP2 | 1.159245475 | 0.953926022 |
| RGS21 | 1.159198075 | 0.938677629 |
| CHRNB3 | 1.159044771 | 1.032446463 |
| RBMY1A1 | 1.159019963 | 1.151764303 |
| GADD45GIP1 | 1.158941441 | 1.021925317 |
| ARHGAP31 | 1.158874292 | 1.039397054 |
| CDH3 | 1.158795635 | 1.131894484 |
| TMIGD2 | 1.158791791 | 0.795868773 |
| OR5B12 | 1.158674885 | 1.06235012 |
| ZBTB49 | 1.158553047 | 1.0222679 |
| SH2B3 | 1.158518548 | 1.072093965 |
| CCDC64 | 1.15845258 | 1.040097205 |
| SRGAP2 | 1.158442854 | 1.071942446 |
| TEX2 | 1.158440456 | 1.057918186 |
| ATP8A2 | 1.158385773 | 1.166774822 |
| C5orf24 | 1.158354928 | 1.052652896 |
| C17orf74 | 1.158316264 | 0.801539085 |
| ZNF493 | 1.158315217 | 0.940733128 |
| FSCN1 | 1.158300537 | 1.153819801 |
| CRY2 | 1.15829176 | 1.074626866 |
| FAM71A | 1.158273118 | 1.038477116 |
| GLA | 1.158262371 | 0.987021415 |
| OGN | 1.158197166 | 1.006480356 |
| MCAM | 1.158181222 | 1.042122317 |
| SERPINA7 | 1.158177084 | 1.153147307 |
| TP53INP1 | 1.157895741 | 1.064405618 |
| AMPD2 | 1.157836348 | 1.005840363 |
| FRMD7 | 1.157819292 | 1.065613609 |
| KIF2A | 1.157741923 | 1.167423751 |
| ZC3H6 | 1.157731267 | 1.095990279 |
| METTL12 | 1.157653561 | 0.934566632 |
| SYNDIG1 | 1.15759629 | 1.092840014 |
| NOX4 | 1.157511613 | 0.853786958 |
| ZNF766 | 1.157452421 | 1.00891049 |
| CCDC102B | 1.157449975 | 1 |
| IPO4 | 1.157436845 | 1.052757794 |
| PALM | 1.157373729 | 0.929427886 |
| RAI1 | 1.15737314 | 0.992463172 |

| | | |
|---|---|---|
| TCEB3 | 1.15735591 | 1.06164828 |
| H2AFZ | 1.157305402 | 0.995203837 |
| HHLA3 | 1.157174746 | 1.010935601 |
| GUCY2EP | 1.157172082 | 1.172615185 |
| GTF2H2C_2 | 1.157091903 | 1.051730045 |
| OPRK1 | 1.157030486 | 1.043478261 |
| UQCR11 | 1.157003896 | 1.001946788 |
| GALNT18 | 1.156947339 | 1.038935756 |
| CYB5B | 1.156883164 | 0.995139733 |
| GRWD1 | 1.156746418 | 1.050702295 |
| ZNF112 | 1.156743472 | 1.018499486 |
| L1TD1 | 1.156555924 | 0.877683273 |
| ARHGAP8 | 1.156513167 | 0.93770279 |
| DSCAML1 | 1.156492231 | 0.947584789 |
| ATP9B | 1.156471429 | 0.974691759 |
| IFI27L1 | 1.15627846 | 0.939705379 |
| SAAL1 | 1.156253849 | 0.888618874 |
| ZNF107 | 1.156161138 | 1.015390846 |
| GYS1 | 1.156146828 | 1.15055159 |
| FAM110C | 1.156129799 | 0.929085303 |
| C11orf54 | 1.156104034 | 0.979748886 |
| CCDC141 | 1.155965869 | 1.019184652 |
| PBDC1 | 1.155945797 | 1.070229531 |
| OR51S1 | 1.155934651 | 1.020554985 |
| C16orf58 | 1.155909595 | 0.978128797 |
| ANKEF1 | 1.155841492 | 1.095392602 |
| CTSF | 1.155827497 | 0.905905256 |
| ERO1L | 1.155827497 | 0.905905256 |
| SNAI1 | 1.155770478 | 1.147833131 |
| XKRY2 | 1.155714038 | 0.996574169 |
| TPP2 | 1.15568162 | 1.049318624 |
| C9orf96 | 1.155672564 | 1.086307592 |
| ZNF560 | 1.155618979 | 0.865022268 |
| EPYC | 1.155568958 | 1.04090725 |
| HIST1H2AB | 1.155424963 | 1.026378897 |
| TACR3 | 1.155302933 | 0.846203764 |
| ATP4B | 1.155292313 | 0.772225827 |
| C3orf52 | 1.155284763 | 1.073309032 |
| MANBAL | 1.155264285 | 1.064398542 |
| INIP | 1.155230494 | 1.087016101 |
| P2RX6 | 1.155148248 | 1.059052563 |
| AGPS | 1.155081566 | 1.128487995 |
| PALB2 | 1.155076622 | 0.953827461 |
| APOBEC3D | 1.155031349 | 0.980532122 |

| | | |
|---|---|---|
| PZP | 1.154980567 | 1.011990408 |
| KLC2 | 1.154978573 | 1.081877355 |
| TINAG | 1.154783391 | 1.040882544 |
| INPP1 | 1.154650933 | 1.10577547 |
| TNPO3 | 1.15464369 | 0.992120589 |
| FUT10 | 1.154269933 | 1.029566626 |
| TECPR1 | 1.154250221 | 0.956149366 |
| MAS1L | 1.154222523 | 0.944192083 |
| GRM8 | 1.154159381 | 1.046073978 |
| CYP2S1 | 1.15396765 | 1.036988968 |
| YWHAB | 1.153950946 | 0.992304577 |
| CCDC30 | 1.153828117 | 1.030489894 |
| PABPC3 | 1.153744294 | 1.034600891 |
| ECEL1 | 1.153711213 | 0.99091499 |
| SLC45A2 | 1.153639854 | 1.123126772 |
| KLHL11 | 1.15361634 | 1.003768414 |
| IVD | 1.153573835 | 1.105305792 |
| PRUNE2 | 1.153568232 | 0.946132037 |
| C17orf75 | 1.15347766 | 1.01417578 |
| RNF138 | 1.153473299 | 1.014276444 |
| KREMEN2 | 1.153462021 | 0.912394549 |
| DLG1 | 1.153443828 | 1.046722907 |
| CYP2U1 | 1.153429833 | 0.944841012 |
| AHSA2 | 1.153282726 | 1.076738609 |
| ASPN | 1.15328042 | 0.948157149 |
| TRPS1 | 1.153203526 | 1.102530824 |
| CPA5 | 1.153165822 | 1.000648929 |
| MBD5 | 1.15314711 | 1.084244633 |
| DYRK1B | 1.153126801 | 1.065541856 |
| OXCT2 | 1.153001981 | 0.971648441 |
| CHST3 | 1.152993831 | 0.95018226 |
| OLFML3 | 1.152886674 | 0.943816375 |
| TFAP2D | 1.152758075 | 0.917585983 |
| OR52A4_pseudogene | 1.152747529 | 1.008564577 |
| FAM75A3 | 1.152688444 | 1.097205346 |
| IFITM2 | 1.152660937 | 0.998784933 |
| PTPDC1 | 1.152421867 | 1.033744322 |
| SACM1L | 1.152421867 | 1.033744322 |
| PCGF5 | 1.152418492 | 1.001297859 |
| CCNA2 | 1.152411083 | 0.936404932 |
| EXOSC10 | 1.152310813 | 0.996755354 |
| STOML2 | 1.152302996 | 1.011647825 |
| DOCK1 | 1.152295792 | 1.031860226 |
| PFDN6 | 1.15219193 | 0.931146213 |

| | | |
|---|---|---|
| OCLN | 1.152123281 | 0.797534069 |
| KDM4D | 1.152093213 | 1.022610483 |
| INTS12 | 1.152092409 | 0.987670344 |
| ZNF670 | 1.152078639 | 0.879303362 |
| URI1 | 1.151966029 | 0.96882494 |
| MCF2L2 | 1.151847663 | 0.946138871 |
| DIRC2 | 1.15178991 | 1.059266872 |
| CSNK1G3 | 1.151730928 | 0.941596366 |
| BRS3 | 1.151698904 | 1.034393251 |
| SMYD5 | 1.151698904 | 1.034393251 |
| STMN4 | 1.151682028 | 0.921891059 |
| LLGL2 | 1.151653936 | 0.923261391 |
| SPTBN4 | 1.151618867 | 0.98663926 |
| ICAM5 | 1.151611617 | 0.88420692 |
| CYP7A1 | 1.15152866 | 0.96495782 |
| PDE4D | 1.151358498 | 0.971243418 |
| C10orf111 | 1.151348368 | 0.980130182 |
| OR4D1 | 1.151309402 | 1.044878383 |
| OSGEP | 1.151302518 | 1.048669695 |
| COX8C | 1.15127949 | 1.046577562 |
| WDR20 | 1.15125239 | 0.940866748 |
| ATP10A | 1.151194922 | 1.04412719 |
| FAM110B | 1.151190396 | 1.075334143 |
| VPS37A | 1.151146179 | 1.053867963 |
| AMOT | 1.15099586 | 0.892771497 |
| PAPSS2 | 1.150976847 | 1.03504218 |
| RC3H1 | 1.150964581 | 1.095392602 |
| LPIN2 | 1.150940789 | 0.980558931 |
| ST6GALNAC5 | 1.150860113 | 1.090850097 |
| FDX1 | 1.150845581 | 0.912515188 |
| NGRN | 1.150807988 | 1.097205346 |
| CACHD1 | 1.150795762 | 1.122316727 |
| COX6B2 | 1.150635206 | 0.961064244 |
| AASDHPPT | 1.150612887 | 0.948562171 |
| FAM172A | 1.15058706 | 1.023896314 |
| EPN1 | 1.150572215 | 1.001027749 |
| RPUSD2 | 1.150460525 | 1.006075334 |
| JAK1 | 1.150443969 | 0.924075276 |
| MC4R | 1.150392764 | 0.951979234 |
| CCR9 | 1.150368232 | 1.040233615 |
| PGLYRP1 | 1.15034444 | 0.979883193 |
| GTF3C3 | 1.150269773 | 0.947436729 |
| FAM133B | 1.150262062 | 1.079789388 |
| HEATR3 | 1.150262062 | 0.966788173 |

| | | |
|---|---|---|
| NFATC4 | 1.150121823 | 0.948967193 |
| SIX4 | 1.150067752 | 1.086956522 |
| TBCEL | 1.150065382 | 1.015390846 |
| FBXL21 | 1.149958756 | 1.082414017 |
| NUDT2 | 1.149858816 | 0.961713173 |
| FIGLA | 1.149835692 | 1.092750101 |
| NEK1 | 1.149705118 | 0.985074627 |
| CYP4F2 | 1.149677293 | 1.012978585 |
| ELK4 | 1.149677293 | 1.012978585 |
| CDC20 | 1.149672412 | 0.893799246 |
| CAMKV | 1.149651032 | 1.040882544 |
| BLCAP | 1.14958365 | 1.105858171 |
| TCEAL3 | 1.149581603 | 1.173754557 |
| GOLGA8G | 1.149559087 | 1.136087485 |
| COX7A2L | 1.149535368 | 1.036340039 |
| CPB1 | 1.149506225 | 0.9201817 |
| ST3GAL1 | 1.149381588 | 1.087605451 |
| C14orf180 | 1.149336849 | 1.122302158 |
| TAGLN3 | 1.14933057 | 1.071688943 |
| ZNF699 | 1.149284723 | 1.021466181 |
| OR2T12 | 1.149273551 | 1.056183625 |
| TMBIM6 | 1.14927047 | 0.985954094 |
| DPF2 | 1.149269711 | 1.083062946 |
| OR10X1 | 1.14925212 | 1.057553957 |
| NUP107 | 1.149238029 | 0.96437136 |
| PCDHA6 | 1.149230779 | 1.058924289 |
| DNAJC17 | 1.149210351 | 1.012555691 |
| B4GALT6 | 1.149095021 | 0.934458144 |
| CXorf36 | 1.149008271 | 0.849331713 |
| WDR53 | 1.148950097 | 1.014580802 |
| OR6S1 | 1.148885475 | 0.913326482 |
| KBTBD6 | 1.148774405 | 0.984204131 |
| GPATCH2 | 1.1487552 | 0.971648441 |
| WDR77 | 1.148743331 | 1.095585257 |
| RNASE10 | 1.148735436 | 0.95909275 |
| DCLK3 | 1.148696165 | 1.004542505 |
| KRT72 | 1.148651028 | 1.074119077 |
| TRIM46 | 1.148587169 | 1.139519792 |
| B3GNT3 | 1.148578525 | 1.055807917 |
| WDR24 | 1.148560578 | 0.916288125 |
| OR7G2 | 1.14851167 | 0.959575197 |
| MARK3 | 1.148459815 | 1.051265412 |
| EFCAB3 | 1.148399485 | 0.964763062 |
| OR7E24 | 1.148358796 | 1.043508051 |

| | | |
|---|---|---|
| DEFA5 | 1.148346336 | 1.069201781 |
| HMX1 | 1.148231779 | 1.070084361 |
| APOD | 1.148219349 | 1.042180402 |
| RPRD2 | 1.148202568 | 0.978759849 |
| TM4SF4 | 1.148163275 | 1.078169299 |
| GIMAP4 | 1.148113252 | 1.053057918 |
| ELP2 | 1.148095474 | 0.982989064 |
| OR1G1 | 1.148031253 | 0.989722508 |
| ASCC2 | 1.147976121 | 1.019036047 |
| SLC30A7 | 1.14793344 | 0.977287476 |
| ATXN2 | 1.147918406 | 0.993924666 |
| RHOH | 1.147835064 | 0.865671642 |
| CCSAP | 1.147829617 | 0.985014176 |
| SPATS2 | 1.14782301 | 1.029119561 |
| SRD5A2 | 1.147692727 | 1.238157041 |
| GPR115 | 1.147599455 | 1.019467878 |
| PRRT3 | 1.147507624 | 0.990684488 |
| SWAP70 | 1.147331174 | 1.010620075 |
| ACVRL1 | 1.147302432 | 1.24789098 |
| SPTAN1 | 1.147295108 | 0.90647482 |
| GPRC5D | 1.147214017 | 1.005840363 |
| RIN3 | 1.147209926 | 1.045906132 |
| L3MBTL1 | 1.147156799 | 1.028756582 |
| OR8B4 | 1.146938567 | 0.983898595 |
| MYOC | 1.14693467 | 0.956834532 |
| PARL | 1.146932917 | 1.059538275 |
| GSPT2 | 1.146818043 | 0.992212849 |
| TEX101 | 1.146748106 | 1.041717294 |
| ZAR1 | 1.146694846 | 0.97327852 |
| DCLRE1C | 1.146563499 | 1.007695423 |
| XIRP1 | 1.146549221 | 1.039739637 |
| LPCAT4 | 1.14651098 | 1.011340624 |
| DLST | 1.146491215 | 0.917780478 |
| EXOC1 | 1.146479652 | 0.79181855 |
| TMEM25 | 1.146467573 | 0.932853717 |
| PTPN18 | 1.146397865 | 1.081116158 |
| TPM2 | 1.14638852 | 1.09963548 |
| CRYGC | 1.146387375 | 1.023638232 |
| TMEM9 | 1.146278632 | 1.031860226 |
| KCNH1 | 1.146272734 | 1.025308241 |
| EIF3E | 1.146259132 | 0.980963953 |
| AFAP1L1 | 1.146153991 | 0.988254354 |
| FUZ | 1.146153991 | 1.004455245 |
| RAB35 | 1.146133278 | 0.969500324 |

| | | |
|---|---|---|
| RHNO1 | 1.146101115 | 0.975698663 |
| CCDC70 | 1.146099281 | 0.943296881 |
| DKC1 | 1.145992505 | 0.96495782 |
| HLA-DQA2 | 1.14594384 | 0.969852689 |
| RRP9 | 1.145874533 | 1.107330903 |
| PPA2 | 1.145791637 | 0.983776768 |
| FHL1 | 1.145766897 | 0.932766302 |
| ZNF776 | 1.145619119 | 1.060753341 |
| RNFT2 | 1.145582289 | 1.077222583 |
| DCAF4 | 1.145572509 | 1.064398542 |
| OR2L13 | 1.145572509 | 1.064398542 |
| SLC7A9 | 1.145415666 | 0.9467878 |
| NFATC3 | 1.145412127 | 1.021414666 |
| WDR25 | 1.14538836 | 1.010530579 |
| TCTE3 | 1.145343397 | 0.985268928 |
| CYTH4 | 1.145170541 | 0.998972251 |
| SMC6 | 1.14516672 | 1.14945322 |
| ANLN | 1.145129264 | 0.877012676 |
| FAT2 | 1.145107651 | 1.0668037 |
| UGP2 | 1.145097569 | 0.863075925 |
| OR2T33 | 1.145058675 | 1.036046983 |
| CXorf56 | 1.14503453 | 0.967598218 |
| MESDC1 | 1.145022605 | 1.137375814 |
| BOK | 1.145013543 | 0.980130182 |
| REPS2 | 1.145002306 | 1.012675574 |
| A26B3 | 1.144979289 | 0.982870846 |
| TBC1D1 | 1.144954912 | 0.953066119 |
| IL2 | 1.144937378 | 0.954436451 |
| OR4K14 | 1.144909574 | 0.99473471 |
| RPUSD3 | 1.144895259 | 0.942486837 |
| ANTXR1 | 1.144850428 | 0.961288112 |
| SOHLH2 | 1.144723082 | 0.97326853 |
| S100A5 | 1.144574493 | 0.918122645 |
| RNF6 | 1.144572347 | 0.994159637 |
| MSANTD1 | 1.144539242 | 0.920863309 |
| TBC1D14 | 1.144456643 | 1.02672148 |
| SLITRK5 | 1.144444455 | 0.895169579 |
| CANT1 | 1.144403585 | 1.012978585 |
| ALG13 | 1.144398083 | 0.998784933 |
| GNB3 | 1.144395792 | 1.098663926 |
| GALNT15 | 1.144358389 | 1.059701493 |
| MAGEA6 | 1.144322073 | 1.005138746 |
| RBPJ | 1.144303974 | 0.961713173 |
| GPATCH3 | 1.144258405 | 0.897934386 |

| | | |
|---|---|---|
| NRGN | 1.144172668 | 0.933809215 |
| CMTR2 | 1.143962814 | 1.002740665 |
| VKORC1L1 | 1.143934273 | 0.999189955 |
| VPS8 | 1.143859593 | 1.232965607 |
| CCL21 | 1.14370985 | 1.096608428 |
| PGLYRP2 | 1.143677823 | 0.990266061 |
| SOX14 | 1.143643336 | 0.960945529 |
| IL1R2 | 1.143601277 | 0.90927501 |
| LGALS12 | 1.143521903 | 1.032446463 |
| GPR153 | 1.143378167 | 1.027903958 |
| UBE2C | 1.143378167 | 0.957819598 |
| DHX36 | 1.143373422 | 0.927501013 |
| THAP7 | 1.143339897 | 1.061664954 |
| ZNF319 | 1.143326214 | 0.989722508 |
| WDR45 | 1.143318865 | 0.89145403 |
| VTI1A | 1.143312168 | 1.027749229 |
| TMX3 | 1.143242416 | 1.093445814 |
| TMEM161B | 1.14321189 | 0.859052248 |
| SOGA3 | 1.143195994 | 1.002055498 |
| KLK1 | 1.143195656 | 0.813108371 |
| ZNF157 | 1.143156937 | 0.856800274 |
| ST3GAL6 | 1.143122059 | 1.15898767 |
| KCNN1 | 1.143111357 | 1.112264763 |
| RNF113A | 1.143111357 | 1.112264763 |
| C10orf32 | 1.143091329 | 1.049674546 |
| PTP4A1 | 1.143065338 | 0.948734588 |
| COBL | 1.142984353 | 1.060637205 |
| ZNF26 | 1.14298168 | 0.757793765 |
| SMDT1 | 1.142969058 | 1.149858242 |
| GPR83 | 1.142949098 | 1.037637897 |
| TRIM77 | 1.142894765 | 0.920830629 |
| FBN2 | 1.142850327 | 1.036313806 |
| SYS1 | 1.142824898 | 1.038072094 |
| ARL10 | 1.142820772 | 0.974888619 |
| DLG4 | 1.14280355 | 1.033095393 |
| ZMAT2 | 1.14277299 | 0.915350344 |
| ARHGAP17 | 1.142764148 | 1.006509078 |
| ALS2CR11 | 1.142756587 | 1.002025111 |
| COLQ | 1.142705382 | 1.051387461 |
| AK9 | 1.142679465 | 1.054128126 |
| FBXO5 | 1.142656768 | 1.028552888 |
| RNF183 | 1.142640057 | 1.005191434 |
| MOGAT2 | 1.142444786 | 0.953926022 |
| CTBS | 1.142282741 | 1.025921426 |

| | | |
|---|---|---|
| HLA-DQA1 | 1.142259888 | 1.06164828 |
| RECK | 1.142179428 | 0.98663926 |
| CAPS2 | 1.142094721 | 0.749696233 |
| KBTBD7 | 1.142081242 | 0.9554642 |
| OR10H2 | 1.142021673 | 1.098420413 |
| ANKRD20A5P | 1.142006572 | 0.879410757 |
| PER2 | 1.141908476 | 0.931140802 |
| PPP2R5D | 1.141854528 | 1.071382219 |
| ITM2B | 1.141850435 | 0.894484412 |
| MAN1B1 | 1.141792292 | 0.935756003 |
| SLC13A3 | 1.141785047 | 1.024659312 |
| CAMSAP1 | 1.141753699 | 0.904076739 |
| SRGAP3 | 1.141730448 | 1.107735925 |
| NUDT14 | 1.141625258 | 0.931213498 |
| PREPL | 1.14159167 | 0.996755354 |
| FHL5 | 1.141537487 | 1.106115836 |
| C17orf96 | 1.141479576 | 1.021240151 |
| CATSPER3 | 1.14147864 | 1.015574302 |
| SC5D | 1.14147864 | 1.015574302 |
| ATP1B3 | 1.141456495 | 0.772225827 |
| OR2Z1 | 1.141450623 | 1.113737581 |
| OR6A2 | 1.141419858 | 1.072970195 |
| PTX3 | 1.141286421 | 1.180541281 |
| ZNF346 | 1.141235448 | 1.096265844 |
| RCBTB1 | 1.141217913 | 1.052757794 |
| L3MBTL2 | 1.141166553 | 1.006489293 |
| HCG27 | 1.141166215 | 0.967112025 |
| MEI1 | 1.141144381 | 0.978128797 |
| DNAJC6 | 1.141130511 | 1.017471737 |
| DBI | 1.141044131 | 1.039692183 |
| IQCA1 | 1.141013884 | 0.984926345 |
| SPANXN4 | 1.140995228 | 1.019846092 |
| MKRN1 | 1.140962062 | 1.04412719 |
| BBOX1 | 1.140951854 | 0.978585334 |
| PDE6G | 1.140879696 | 1.007290401 |
| LHFPL1 | 1.140803133 | 0.963953017 |
| CHMP1B | 1.140770022 | 0.967598218 |
| CDCA7 | 1.140705688 | 0.974306269 |
| SNX24 | 1.140648915 | 0.883179171 |
| TRHR | 1.140623055 | 0.969500324 |
| MAP7D1 | 1.14059978 | 1.040097205 |
| KCNA10 | 1.140525337 | 0.988319273 |
| BAIAP3 | 1.140366591 | 1.068516615 |
| ABHD8 | 1.140307593 | 0.918234912 |

| | | |
|---|---|---|
| TIGD4 | 1.140271441 | 1.030489894 |
| SAMM50 | 1.140206878 | 1.006075334 |
| ZNF236 | 1.140186712 | 0.937992463 |
| DKK1 | 1.14015871 | 0.993833505 |
| GPR173 | 1.140009335 | 1.101232966 |
| SPAM1 | 1.139971055 | 0.740428293 |
| SHROOM2 | 1.139860083 | 1.054510058 |
| MLXIP | 1.139821618 | 0.879708384 |
| CEP63 | 1.139786469 | 0.972053463 |
| LSM14A | 1.139647046 | 1.109761037 |
| EMP2 | 1.139605027 | 1.017471737 |
| OR1S1 | 1.139573129 | 0.73826653 |
| KCNV1 | 1.139457934 | 1.022063595 |
| POLQ | 1.139404754 | 1.083062946 |
| HS3ST4 | 1.139344493 | 0.956521739 |
| ACP2 | 1.139294688 | 1.01752109 |
| ARSJ | 1.139230508 | 0.983394087 |
| PTX4 | 1.139221935 | 0.957862282 |
| UBE2N | 1.139063524 | 1.031797534 |
| PPEF1 | 1.139029524 | 0.989617132 |
| MRPS22 | 1.138838117 | 1.018156903 |
| USP4 | 1.138765102 | 0.900713822 |
| EPHA6 | 1.138659235 | 1.163530175 |
| CXCR4 | 1.138633879 | 0.93835172 |
| TUBB2B | 1.138543376 | 1.016605913 |
| KRTAP5-1 | 1.138515002 | 0.98582422 |
| TM7SF3 | 1.138462727 | 1.017129154 |
| FBP2 | 1.13845886 | 1.055807917 |
| KIAA1211L | 1.138396872 | 1.004050223 |
| TNKS1BP1 | 1.138396872 | 1.004050223 |
| IL16 | 1.138277954 | 0.9109284 |
| C11orf85 | 1.138269663 | 0.912298732 |
| ENPEP | 1.138268117 | 0.92926671 |
| PPP1R15B | 1.138268117 | 0.92926671 |
| ACTN3 | 1.138233279 | 1.059609455 |
| CYB561D1 | 1.138233279 | 1.059609455 |
| CDS2 | 1.138153605 | 1.107722258 |
| ZNF100 | 1.138125306 | 0.864679685 |
| SAMD14 | 1.138005397 | 1.107735925 |
| RAP1GDS1 | 1.137977532 | 0.921020656 |
| TMEM67 | 1.137879422 | 1.05346294 |
| DDX23 | 1.137800686 | 0.957819598 |
| FCER2 | 1.137759606 | 1.056456846 |
| DGCR14 | 1.137750186 | 1.037262049 |

| | | |
|---|---|---|
| KCNK10 | 1.137719927 | 1.075275795 |
| PLAT | 1.137537745 | 0.99091499 |
| RADIL | 1.137530391 | 0.891058582 |
| MTMR10 | 1.137499102 | 1.001215067 |
| BAI2 | 1.137360339 | 0.986372485 |
| C9orf91 | 1.137359891 | 0.923946557 |
| FAM9B | 1.137355196 | 1.11381126 |
| CASP9 | 1.137325434 | 1.005191434 |
| RABL2B | 1.137248708 | 0.944192083 |
| DSG3 | 1.137182051 | 0.960945529 |
| TRMT61B | 1.137136062 | 1.059266872 |
| CABP4 | 1.137125686 | 0.97327852 |
| RAB11FIP4 | 1.137040692 | 1.092345079 |
| ITGBL1 | 1.137027319 | 0.905104488 |
| HORMAD2 | 1.137020876 | 1.00526529 |
| CSF2 | 1.136992436 | 0.859831278 |
| STC1 | 1.136919754 | 1.021240151 |
| CHPT1 | 1.136918428 | 1.033744322 |
| IL10RA | 1.136909784 | 1.170019468 |
| METTL15 | 1.136835708 | 0.9963548 |
| RAB44 | 1.136826507 | 1.0445358 |
| TRHDE | 1.136793296 | 0.99156392 |
| LOC155060 | 1.136788385 | 0.958282706 |
| IREB2 | 1.136757239 | 0.874107722 |
| SCGB1D4 | 1.136748855 | 1.014580802 |
| TSPAN10 | 1.136745748 | 1.063588497 |
| SHMT1 | 1.136741186 | 0.737832576 |
| PIGH | 1.136705046 | 0.91174562 |
| THRB | 1.136705046 | 0.91174562 |
| WRNIP1 | 1.136634412 | 0.968202466 |
| MOCOS | 1.136591687 | 1.005840363 |
| DCP2 | 1.136589175 | 1.108941418 |
| MAP4K1 | 1.136448609 | 0.963659961 |
| SESN2 | 1.136388001 | 0.850976362 |
| TELO2 | 1.136234461 | 1.029566626 |
| ZNF571 | 1.136166663 | 1.019184652 |
| TMEM243 | 1.13610332 | 1.038369305 |
| PLA2G12B | 1.136091472 | 0.953422438 |
| FAM20C | 1.136037576 | 0.941418294 |
| VDAC2 | 1.136001511 | 1.086307592 |
| SBSN | 1.135674833 | 0.999189955 |
| P2RY12 | 1.135663739 | 1.077222583 |
| FAM92B | 1.135632145 | 1.137302552 |
| TNNC2 | 1.135603465 | 0.948612539 |

| | | |
|---|---|---|
| TRMT112 | 1.135566719 | 0.960945529 |
| DHRS7B | 1.13552084 | 1.042932361 |
| FBXW8 | 1.135492754 | 1.091499027 |
| NFIB | 1.135287851 | 0.898767034 |
| PLEC | 1.135223227 | 1.093867763 |
| TRAF3IP1 | 1.135172389 | 0.957177115 |
| GSTZ1 | 1.135169306 | 1.138870863 |
| LHX2 | 1.135131339 | 0.973621103 |
| ZBTB46 | 1.135110415 | 0.737950587 |
| TRAP1 | 1.135021206 | 0.781310837 |
| MINA | 1.134903024 | 1.076396026 |
| KRT14 | 1.134863514 | 0.913669065 |
| KRT77 | 1.13477747 | 1.074524099 |
| NDFIP2 | 1.134749017 | 0.965741692 |
| COMMD10 | 1.134726452 | 0.976704351 |
| RBMY1E | 1.134663019 | 0.807125728 |
| MBD6 | 1.134572808 | 0.846865365 |
| RRAGB | 1.13454824 | 0.99108951 |
| ZBTB2 | 1.134464019 | 0.900308325 |
| NIPBL | 1.134365063 | 0.955121617 |
| TTC8 | 1.134330776 | 1.182260024 |
| NRN1 | 1.134302309 | 0.993490922 |
| CUEDC1 | 1.134258314 | 1.0222679 |
| PAM | 1.134257659 | 0.894873459 |
| PIGS | 1.134246216 | 1.030489894 |
| L2HGDH | 1.13423115 | 1.022063595 |
| LY6G6C | 1.13397608 | 1.046172539 |
| C1orf229 | 1.133956642 | 0.956834532 |
| ZNF423 | 1.133942525 | 1.069435432 |
| C18orf54 | 1.133942357 | 1.064398542 |
| TMC1 | 1.133932674 | 0.976019185 |
| MYLK | 1.1339147 | 1.050616483 |
| OR1D2 | 1.133905629 | 0.96435804 |
| POLD1 | 1.133885969 | 1.031797534 |
| EPS8L3 | 1.13384722 | 1.051387461 |
| PPAT | 1.133688558 | 0.918883842 |
| RRS1 | 1.133590994 | 0.911956149 |
| SAP30L | 1.133577129 | 0.927029805 |
| CLCN5 | 1.133518725 | 0.933160286 |
| SLC6A12 | 1.133511398 | 1.022712524 |
| RBMY1B | 1.133460049 | 1.077766358 |
| BATF2 | 1.133458222 | 0.78817335 |
| CASP5 | 1.133401909 | 1.145360156 |
| ACSS2 | 1.13336979 | 0.839206156 |

| | | |
|---|---|---|
| KIF11 | 1.133284465 | 0.537059538 |
| KIAA1467 | 1.133270762 | 0.904819765 |
| HPX | 1.13311556 | 1.032887975 |
| LYPD2 | 1.133108311 | 1.05344296 |
| HIST2H4B | 1.133098686 | 1.046982584 |
| GABRG1 | 1.133028902 | 1.046722907 |
| C1R | 1.133020011 | 1.117456197 |
| SLC7A8 | 1.132983871 | 1.027903958 |
| PDPK1 | 1.132888459 | 0.848799481 |
| SLC38A1 | 1.132837994 | 0.971447112 |
| MYL3 | 1.132834043 | 0.900713822 |
| AEBP2 | 1.132739898 | 1.0222679 |
| OR6B2 | 1.132739898 | 0.912641316 |
| PPP1R15A | 1.132739898 | 1.019527235 |
| SLX4 | 1.132739898 | 0.998972251 |
| STRA13 | 1.132739898 | 1.011305242 |
| TUSC5 | 1.132739898 | 1.026378897 |
| BCLAF1 | 1.132694221 | 0.9963548 |
| TKTL1 | 1.132618521 | 1.089552239 |
| PNMT | 1.132519239 | 0.92926671 |
| ARHGEF10L | 1.132400907 | 1.155123532 |
| OR2L3 | 1.132354722 | 1.007536828 |
| UBL4A | 1.132346854 | 0.995457495 |
| FAM193A | 1.13234566 | 0.984241178 |
| DDIT4L | 1.132330254 | 0.947242206 |
| SYMPK | 1.131999087 | 1.047619048 |
| RBM22 | 1.131930216 | 0.958547448 |
| LOC642219_pseudogene | 1.131803603 | 0.948734588 |
| DKK4 | 1.131783716 | 1.04090725 |
| ZNF354B | 1.131764202 | 0.795477903 |
| CLDN7 | 1.13166008 | 1.078108941 |
| MOCS2 | 1.131609238 | 0.996106424 |
| VSTM2L | 1.131606751 | 1.027406646 |
| CCDC89 | 1.131559681 | 1.03199676 |
| EFCAB6 | 1.131557907 | 0.984926345 |
| B9D2 | 1.131552968 | 0.980815348 |
| C22orf29 | 1.131520406 | 0.948157149 |
| CREB3L2 | 1.131504578 | 0.942446043 |
| SLC6A1 | 1.131504358 | 1.06229721 |
| CSMD1 | 1.131452245 | 0.904076739 |
| PAK2 | 1.131411827 | 1.128487995 |
| NIT2 | 1.131362104 | 0.802725503 |
| NDUFC1 | 1.131346628 | 0.835560123 |
| DNAJC19 | 1.131336512 | 1.030376671 |

| | | |
|---|---|---|
| ADAM12 | 1.13132654 | 0.972744971 |
| PRODH | 1.131280521 | 1.005840363 |
| VPS45 | 1.131257198 | 0.921020656 |
| GBGT1 | 1.131204043 | 0.902011681 |
| SLC8A2 | 1.131200174 | 0.987021415 |
| OR4M1 | 1.131126333 | 0.961973279 |
| TNS4 | 1.131079876 | 0.992304577 |
| CCNJL | 1.13105899 | 0.970433374 |
| IL5RA | 1.131030012 | 0.949383517 |
| P2RY6 | 1.131017334 | 0.864373783 |
| DAGLA | 1.13097555 | 1.114621304 |
| ZSCAN21 | 1.130965464 | 0.882948562 |
| CELA2B | 1.130958061 | 1.015574302 |
| ZFP1 | 1.13094813 | 1.082905104 |
| SLIT2 | 1.130922893 | 1.067831449 |
| MAP9 | 1.130922522 | 1.059943297 |
| TDO2 | 1.130907443 | 1.045362495 |
| KCNK2 | 1.130813502 | 1.062946139 |
| GGT7 | 1.130783536 | 0.977936405 |
| VAT1 | 1.13077942 | 0.859831278 |
| OR4F6 | 1.130745097 | 0.910490077 |
| MROH9 | 1.13051444 | 0.92345079 |
| OARD1 | 1.130410759 | 0.999657417 |
| POLDIP2 | 1.130397891 | 0.994176088 |
| PML | 1.130296018 | 0.850746269 |
| PTPRU | 1.130269345 | 0.916937054 |
| SEC23A | 1.13017465 | 1.058924289 |
| CAV1 | 1.130151623 | 0.931213498 |
| SAR1B | 1.130094183 | 0.960307817 |
| GDPD4 | 1.130085269 | 1.075334143 |
| MRPL48 | 1.130074652 | 1.019184652 |
| CD48 | 1.13005245 | 1.155190134 |
| TSPAN17 | 1.130020976 | 1.110966905 |
| ZNF781 | 1.129960618 | 1.009720535 |
| RARRES3 | 1.129926716 | 0.8935756 |
| COCH | 1.129894818 | 0.954779034 |
| HIPK4 | 1.129829081 | 1.073329007 |
| CTSG | 1.129819119 | 0.974042829 |
| HSD17B8 | 1.129727988 | 1.054510058 |
| OR4X1 | 1.12968431 | 0.889003083 |
| BRCC3 | 1.12964377 | 1.068448765 |
| CLP1 | 1.129581012 | 0.946132037 |
| STAP1 | 1.129460798 | 1.065090785 |
| OR51B4 | 1.129422375 | 1.052757794 |

| | | |
|---|---|---|
| YKT6 | 1.129329219 | 1.023980815 |
| KCNH6 | 1.129234707 | 1.059701493 |
| RBPMS | 1.12921599 | 0.99109284 |
| P2RX1 | 1.129188606 | 0.993510707 |
| NOTUM | 1.129105587 | 1.107330903 |
| GLS | 1.129004284 | 0.908500973 |
| PAICS | 1.129004284 | 1.022063595 |
| STXBP5 | 1.128991572 | 1.035286057 |
| CORO6 | 1.128909722 | 0.963142973 |
| SLC25A21 | 1.128790533 | 1.080849606 |
| APOL2 | 1.128620803 | 0.94210346 |
| ERI2 | 1.128580139 | 1.071283921 |
| CDC34 | 1.128578673 | 0.932511356 |
| NMT2 | 1.128578673 | 0.932511356 |
| SEPT7 | 1.128537017 | 0.979883193 |
| LAMC1 | 1.128517543 | 1.010962658 |
| SGCB | 1.128511834 | 1.009592326 |
| FAM71D | 1.128510112 | 0.917437479 |
| DHFRL1 | 1.128488771 | 1.004110997 |
| UNC119 | 1.128355062 | 1.062007537 |
| TMEM2 | 1.128350984 | 0.972593354 |
| MCRS1 | 1.128304604 | 1.115431349 |
| HIST1H2AM | 1.128280301 | 1.044193217 |
| GPM6A | 1.128187885 | 1.084360805 |
| TBC1D5 | 1.128153888 | 1.015416238 |
| C1D | 1.128150489 | 0.930113052 |
| NAP1L1 | 1.128110662 | 1.089756766 |
| GRM2 | 1.128074773 | 1.018170019 |
| ENTPD7 | 1.128012619 | 0.985074627 |
| SPC24 | 1.127836267 | 0.949640288 |
| KIAA0247 | 1.127800654 | 0.942788626 |
| GJB4 | 1.127798084 | 0.885788449 |
| TMEM184B | 1.127778996 | 0.938677629 |
| TDRD9 | 1.127735042 | 0.912920211 |
| CORO2B | 1.127638357 | 1.050222762 |
| HMCES | 1.127571116 | 0.982584042 |
| RASD2 | 1.127527936 | 0.893456663 |
| STAMBPL1 | 1.127517444 | 0.739130435 |
| BRPF1 | 1.127513163 | 1.085009734 |
| SERPINI2 | 1.127482622 | 0.942894225 |
| ZC3H7B | 1.127444632 | 1.055893074 |
| POLR1B | 1.127290397 | 1.033095393 |
| PTTG1 | 1.127274261 | 0.829331603 |
| MEGF6 | 1.127264711 | 1.015795869 |

| | | |
|---|---|---|
| RUFY2 | 1.127247832 | 1.130524152 |
| PELI2 | 1.127242893 | 1.058924289 |
| ARMC7 | 1.12722671 | 1.103685703 |
| CCNL1 | 1.127223501 | 0.95909275 |
| RSAD1 | 1.127178877 | 1.092750101 |
| XAGE1D | 1.127155954 | 1.134872418 |
| DYRK4 | 1.127013626 | 1.042829332 |
| LDHC | 1.126980285 | 0.995457495 |
| FNDC7 | 1.126864882 | 0.981368975 |
| SEMA6B | 1.126815419 | 1.016200891 |
| PGLS | 1.126756226 | 0.917375456 |
| KAZN | 1.126682459 | 1.025008565 |
| REEP5 | 1.126672328 | 1.074929121 |
| LY75 | 1.126638962 | 1.025516403 |
| ZNF740 | 1.126619859 | 1.021871203 |
| PDILT | 1.126506721 | 1.084244633 |
| FAM26E | 1.126502086 | 1 |
| AP2A1 | 1.126473491 | 1.052757794 |
| ZNF276 | 1.126470401 | 0.869987849 |
| A1CF | 1.126390891 | 1.039054471 |
| HIST1H2BB | 1.126357181 | 1.033573141 |
| CIITA | 1.126354052 | 0.967553537 |
| KRTAP15-1 | 1.126313004 | 0.966084275 |
| NDUFB11 | 1.126227731 | 1.013018157 |
| TNP1 | 1.126182784 | 0.946899623 |
| LONP2 | 1.126172777 | 0.948734588 |
| CBFB | 1.126144211 | 1.024659312 |
| DIRAS2 | 1.12609273 | 0.930741191 |
| SOX30 | 1.126079343 | 0.873929428 |
| IGBP1 | 1.126037108 | 0.915639195 |
| TM6SF2 | 1.126018582 | 0.981500514 |
| PPP5C | 1.126018009 | 0.99156392 |
| ZFYVE20 | 1.125997423 | 0.920863309 |
| ISY1 | 1.125982649 | 1.027946537 |
| COG1 | 1.125961809 | 0.97327852 |
| NARS2 | 1.125954275 | 0.947347104 |
| FLOT1 | 1.12587477 | 1.017471737 |
| UBE2Z | 1.125843319 | 0.820895522 |
| RANBP3 | 1.125779891 | 0.892086331 |
| FOXO3 | 1.125742144 | 1.001297859 |
| SMIM14 | 1.125701561 | 0.945727015 |
| KCNK16 | 1.125697854 | 1.015574302 |
| OR1N1 | 1.125683609 | 0.934909215 |
| IFIT3 | 1.125682407 | 1.015390846 |

| | | |
|---|---|---|
| NDUFC2 | 1.12565816 | 1.119886594 |
| HNF1A | 1.125652607 | 0.938436614 |
| FDCSP | 1.125635626 | 0.983213429 |
| UGT2B28 | 1.125571982 | 1.058403634 |
| PCDP1 | 1.125446249 | 1.013770757 |
| KIAA0355 | 1.125422511 | 1.079789388 |
| FLJ46838_pseudogene | 1.125400333 | 0.951695786 |
| FYTTD1 | 1.125399486 | 1.041312272 |
| KRT75 | 1.125329341 | 1.030376671 |
| GARS | 1.125314722 | 1.158338741 |
| ILDR2 | 1.125305681 | 0.991778006 |
| SMG6 | 1.125292409 | 0.922235723 |
| NLGN1 | 1.125243347 | 1.006489293 |
| C2orf40 | 1.125239839 | 1.050627785 |
| CRX | 1.125179053 | 1.125243348 |
| ANKRD6 | 1.12512689 | 0.968482357 |
| FAM160B1 | 1.125116061 | 0.967112025 |
| PDZD7 | 1.125046725 | 1.021466181 |
| UPF3B | 1.124975969 | 0.913325233 |
| PISD | 1.124971384 | 1.010530579 |
| WDR35 | 1.124895448 | 0.989379925 |
| SIRT1 | 1.12488236 | 1.059052563 |
| IL9 | 1.124878903 | 0.954574951 |
| RTP2 | 1.124838766 | 1.080507023 |
| FES | 1.124826298 | 0.759896171 |
| C3orf35 | 1.124801855 | 1.082219522 |
| GPC3 | 1.124718288 | 0.919150394 |
| TEN1 | 1.124696312 | 1.109626584 |
| ESX1 | 1.124438856 | 1 |
| CACFD1 | 1.124381039 | 1.082624544 |
| PUS7 | 1.12436433 | 1.020251114 |
| TFCP2L1 | 1.124299432 | 0.833130822 |
| S100A6 | 1.124237852 | 0.883841661 |
| FKBP9 | 1.124172158 | 1.073977936 |
| ULK3 | 1.124114696 | 0.955223881 |
| SLC35G1 | 1.124056903 | 0.983213429 |
| ADORA2A | 1.124008664 | 1.0266061 |
| PRMT1 | 1.124008664 | 1.0266061 |
| EPAS1 | 1.123951861 | 1.069435432 |
| TTC31 | 1.123889211 | 1.017010936 |
| PIGL | 1.123844078 | 0.92709599 |
| NCOA6 | 1.1238115 | 0.993510707 |
| CIZ1 | 1.123794212 | 1.007787151 |
| FBXL13 | 1.123757002 | 0.83192732 |

| | | |
|---|---|---|
| TRIM54 | 1.123729588 | 1.064892927 |
| tcag7.23_pseudogene | 1.123710055 | 1.022681247 |
| CCDC102A | 1.123680356 | 1.019036047 |
| CHM | 1.123680356 | 1.046577562 |
| STIM2 | 1.123592485 | 1.03564196 |
| SNX10 | 1.123587904 | 1.10243234 |
| GABRR1 | 1.123557615 | 1.003244646 |
| CCNI2 | 1.123530587 | 1.095580678 |
| AXDND1 | 1.123420714 | 0.934791414 |
| COA5 | 1.123350255 | 0.909215485 |
| CCDC74A | 1.123329582 | 0.951397327 |
| TMEM182 | 1.123274414 | 0.971648441 |
| DNAJA3 | 1.123264484 | 0.900993491 |
| OR2T2 | 1.123156539 | 0.984609154 |
| NKD2 | 1.123120943 | 0.847207948 |
| TSPAN33 | 1.123090571 | 1.028756582 |
| VGF | 1.123071084 | 1.00843608 |
| YIPF2 | 1.123030732 | 0.959232614 |
| PSRC1 | 1.123029952 | 1.07290401 |
| TMEM116 | 1.122984004 | 0.914945322 |
| CPSF2 | 1.122951213 | 0.99109284 |
| KRT82 | 1.122929753 | 1.010530579 |
| CLCNKB | 1.12282148 | 1.046722907 |
| R3HDM4 | 1.122788507 | 1.130866735 |
| PPP1R14A | 1.122701467 | 0.966426859 |
| GRAMD1B | 1.122625284 | 0.977723775 |
| FPR1 | 1.122589494 | 1.027903958 |
| MMP10 | 1.122589494 | 0.985074627 |
| ZMAT1 | 1.122483313 | 0.963142973 |
| ARHGDIA | 1.122445414 | 1.007290401 |
| COL4A5 | 1.122294949 | 1.077423775 |
| BIN1 | 1.122273926 | 0.964028777 |
| RBM18 | 1.122235322 | 0.886605002 |
| QARS | 1.122177235 | 0.918465228 |
| CBLN2 | 1.122175513 | 0.991778006 |
| TM9SF3 | 1.122021133 | 0.987444309 |
| STARD7-AS1 | 1.122003915 | 1.031591738 |
| THBS3 | 1.121988073 | 1.075739166 |
| ZFYVE9 | 1.1219859 | 0.804672291 |
| CPNE9 | 1.121979362 | 0.891859052 |
| MRM1 | 1.121973504 | 0.959902795 |
| ZXDA | 1.121911992 | 1.110996917 |
| AGO1 | 1.12181689 | 0.942894225 |
| CXCR3 | 1.121746668 | 1.152498378 |

| | | |
|---|---|---|
| AP1M1 | 1.121735013 | 1.022610483 |
| CROCC | 1.121620969 | 1.116820829 |
| ANKRD46 | 1.121618604 | 0.971243418 |
| GPRIN2 | 1.121618604 | 0.882948562 |
| NAGS | 1.121568908 | 1.042137718 |
| TUBG2 | 1.121493921 | 1.000685166 |
| NUDCD1 | 1.121467311 | 1.000810045 |
| BHLHE22 | 1.121467052 | 1.067146283 |
| ZDHHC8 | 1.121454864 | 1.100376841 |
| EHMT2 | 1.121446518 | 1.06229721 |
| KBTBD11 | 1.121432028 | 0.995203837 |
| ZNF595 | 1.121369365 | 0.989722508 |
| TRIM5 | 1.121364195 | 0.99091499 |
| LRTM2 | 1.121330988 | 0.952380952 |
| MCOLN1 | 1.121308873 | 0.948085659 |
| NXNL1 | 1.121297069 | 0.942081814 |
| NMU | 1.121290662 | 1.050702295 |
| RASSF9 | 1.121237831 | 0.9109284 |
| CBLL1 | 1.121230419 | 1.072031149 |
| KLLN | 1.121189168 | 1.007879411 |
| TTI1 | 1.121185136 | 1.041109969 |
| BLOC1S1 | 1.12113238 | 1.012150668 |
| CCDC86 | 1.121085005 | 1.028756582 |
| NCF4 | 1.121078205 | 0.998287085 |
| ZNF669 | 1.121069825 | 1.069258809 |
| TTC9B | 1.121062643 | 1.089509923 |
| TACSTD2 | 1.120959132 | 1.087016101 |
| NARFL | 1.120884046 | 1.030781693 |
| LDOC1 | 1.120726557 | 1.001370332 |
| CUL4B | 1.120711454 | 1.034393251 |
| MSANTD4 | 1.120689494 | 1.030489894 |
| NEMF | 1.120638793 | 1.058239123 |
| POLR3E | 1.120508981 | 0.983556012 |
| PROKR2 | 1.120496343 | 1.04412719 |
| ZNF124 | 1.120477491 | 1.012675574 |
| F3 | 1.120472093 | 0.974078574 |
| RNF169 | 1.120445056 | 0.905905256 |
| SLC1A2 | 1.120406858 | 1.001297859 |
| BPIFB1 | 1.12038805 | 1.068174032 |
| C9orf131 | 1.120365119 | 0.906034832 |
| CASC1 | 1.120331186 | 0.875642343 |
| CSPP1 | 1.120327016 | 1.040502228 |
| LILRB2 | 1.120315645 | 1.093182597 |
| TBC1D19 | 1.12010124 | 0.982528263 |

| | | |
|---|---|---|
| ARHGEF37 | 1.120083614 | 0.981157931 |
| GNAZ | 1.120037675 | 0.957877683 |
| CYB5D2 | 1.119918475 | 1.024301337 |
| CBX1 | 1.119897029 | 1.057553957 |
| PADI6 | 1.119881883 | 0.896820247 |
| TMEM200C | 1.119841978 | 1.022953066 |
| KDM5D | 1.119761773 | 1.030499676 |
| ITIH1 | 1.119710852 | 0.982870846 |
| MATN1 | 1.119710852 | 0.982870846 |
| PTPN9 | 1.119697643 | 0.839714471 |
| GADL1 | 1.119693829 | 0.875253139 |
| SLC26A4 | 1.119681179 | 1.001946788 |
| MRRF | 1.119653037 | 1.067488866 |
| FGFR1OP | 1.119637768 | 0.977389517 |
| C7orf50 | 1.119505101 | 0.996916752 |
| SLC7A5 | 1.119351721 | 1.049967554 |
| KIAA1377 | 1.119328258 | 1.070572114 |
| KTN1 | 1.119313357 | 0.993519644 |
| PYHIN1 | 1.119313357 | 0.903199676 |
| LTBP1 | 1.119254902 | 1.035971223 |
| TMEM213 | 1.119211731 | 1.093155124 |
| OR2B6 | 1.119169357 | 0.972250771 |
| CDKN2AIP | 1.119127171 | 1.192790603 |
| PLBD2 | 1.119103749 | 0.825282631 |
| ECHS1 | 1.119072069 | 0.897469176 |
| NR2E1 | 1.119072069 | 0.897469176 |
| FITM1 | 1.119011997 | 1.045906132 |
| EIF2D | 1.118968001 | 0.901678657 |
| IFNLR1 | 1.118952904 | 1.001215067 |
| NFU1 | 1.118929574 | 0.96435804 |
| AKAP9 | 1.118904047 | 0.988319273 |
| AP3B1 | 1.118846388 | 1.010530579 |
| KDELC1 | 1.118777701 | 0.861596437 |
| HDAC6 | 1.118720384 | 0.878650227 |
| HIST4H4 | 1.118710779 | 0.85748544 |
| C11orf48 | 1.118686283 | 1.032806804 |
| PLA2G15 | 1.118677988 | 0.931213498 |
| C3orf67 | 1.118642543 | 1.018499486 |
| ENPP5 | 1.118617069 | 0.916937054 |
| RABL6 | 1.118545127 | 1.09352518 |
| ICAM2 | 1.118509514 | 1.090784515 |
| EML2 | 1.11847748 | 1.033915725 |
| SLC4A7 | 1.118467323 | 1.060350422 |
| THUMPD3 | 1.11845854 | 1.032545392 |

| | | |
|---|---|---|
| RBM5 | 1.118445582 | 1.113052415 |
| PUM2 | 1.11843951 | 1.03117506 |
| TMPRSS15 | 1.118422456 | 0.874107722 |
| DPH3 | 1.118408405 | 0.961117861 |
| TRMT13 | 1.11840489 | 0.977723775 |
| GCHFR | 1.118401405 | 1.028434395 |
| RFC4 | 1.118394906 | 1.027541515 |
| GALNT14 | 1.118388793 | 1.060753341 |
| GTF2H5 | 1.118324605 | 1.022953066 |
| TRPV3 | 1.118293868 | 1.01752109 |
| CUL7 | 1.118247273 | 0.950681376 |
| TSPAN6 | 1.118207842 | 1.014731072 |
| RHOF | 1.118199041 | 0.863507493 |
| SLC2A5 | 1.11814265 | 0.893074119 |
| KRT6B | 1.118117359 | 1.141144227 |
| ANKMY2 | 1.118113462 | 0.955121617 |
| ZNF18 | 1.118106983 | 1.034258308 |
| TSPYL6 | 1.118105158 | 1.072093965 |
| ZNF821 | 1.118100573 | 1.022276225 |
| DDX46 | 1.118098185 | 1.027255029 |
| C1orf204 | 1.118048035 | 1.056526208 |
| SCN4B | 1.118028704 | 1.118105127 |
| OR5K1 | 1.117941742 | 0.978128797 |
| FAM107B | 1.117937361 | 0.961522884 |
| TOR3A | 1.117923098 | 0.911705144 |
| RNF14 | 1.11791197 | 1.142115509 |
| RGPD7_pseudogene | 1.117866299 | 0.991435423 |
| ATAD3C | 1.117823306 | 0.937626569 |
| GLB1L | 1.117769744 | 1.010962658 |
| MYO1C | 1.117732001 | 1.085988352 |
| SASS6 | 1.117664525 | 0.847306602 |
| DROSHA | 1.117630039 | 0.972863507 |
| WDR86 | 1.117620921 | 1.13163224 |
| SLC9A9 | 1.117584855 | 1.065208586 |
| COA4 | 1.117462344 | 1.024706359 |
| ARFGEF2 | 1.117443515 | 0.964028777 |
| SUSD1 | 1.117426584 | 0.912110166 |
| TTC33 | 1.117409672 | 1.083839611 |
| TRAPPC12 | 1.117354074 | 1.134977732 |
| KLK2 | 1.117335543 | 0.83192732 |
| MYRF | 1.117307495 | 1.005823912 |
| MARS | 1.117243752 | 1.090201168 |
| TRIP13 | 1.117243752 | 0.999351071 |
| USP2 | 1.117166286 | 0.985074627 |

| | | |
|---|---|---|
| WDR19 | 1.117164955 | 0.97326853 |
| MST4 | 1.116975296 | 0.994808566 |
| CCDC47 | 1.116960685 | 0.853381936 |
| CDIP1 | 1.116934196 | 1.034426893 |
| KRTAP21-2 | 1.116918331 | 0.907502569 |
| HAX1 | 1.11691056 | 1.093560146 |
| MCCC2 | 1.116890642 | 1.076954232 |
| ATF4 | 1.116809415 | 0.966255678 |
| IFT81 | 1.116781325 | 0.993924666 |
| NUP54 | 1.116752861 | 0.995203837 |
| CHD9 | 1.116688562 | 1.032806804 |
| CC2D2B | 1.116642033 | 0.988352175 |
| COX19 | 1.116553011 | 1.00891049 |
| ZNF513 | 1.116395707 | 1.116241393 |
| RABEP2 | 1.116391063 | 0.996916752 |
| GRTP1 | 1.116370855 | 0.948269955 |
| BZW2 | 1.11635779 | 0.818299805 |
| CARKD | 1.116299583 | 0.944158959 |
| CSTF3 | 1.116299583 | 0.944158959 |
| KIFC3 | 1.116247148 | 0.842310188 |
| GPN2 | 1.116199842 | 0.85581207 |
| ZBTB14 | 1.116176716 | 0.970433374 |
| SRM | 1.116151159 | 0.942894225 |
| ZSCAN29 | 1.116116199 | 1.038882139 |
| PRRG4 | 1.116104992 | 1.081409478 |
| ATN1 | 1.116090816 | 1.158338741 |
| ANKRD30A | 1.116014903 | 1.020897568 |
| GMIP | 1.115973011 | 0.99091499 |
| RHOC | 1.115973011 | 0.99091499 |
| CD24 | 1.115969865 | 1.018156903 |
| GKN2 | 1.115889973 | 1.024301337 |
| DOCK6 | 1.115738223 | 1.042932361 |
| PSMD6 | 1.115696875 | 0.933538883 |
| ZNF385C | 1.115694519 | 1.001712915 |
| HIST1H4B | 1.115607343 | 0.973963686 |
| MRPL54 | 1.115603175 | 0.905789654 |
| CELF5 | 1.115596197 | 0.928057554 |
| GOPC | 1.115529124 | 1.134977287 |
| ABHD3 | 1.115519904 | 0.996106424 |
| MICALL2 | 1.115478326 | 0.887808829 |
| SULF2 | 1.115463451 | 1.011745646 |
| HFE2 | 1.11536172 | 0.982528263 |
| KRT23 | 1.115313148 | 0.935251799 |
| TRPC4 | 1.115311379 | 0.967553537 |

| | | |
|---|---|---|
| PLVAP | 1.115288681 | 0.978417266 |
| USP44 | 1.11527277 | 0.92926671 |
| S100P | 1.115258366 | 0.958282706 |
| B3GALT5 | 1.115242639 | 0.99156392 |
| SOX11 | 1.115152752 | 1.19150394 |
| SLC52A3 | 1.115138555 | 1.019441069 |
| MRGPRG | 1.115111087 | 0.876703439 |
| RXFP4 | 1.115111087 | 0.876703439 |
| JKAMP | 1.115105313 | 1.034258308 |
| EEA1 | 1.11509668 | 1.101881895 |
| DLGAP3 | 1.115071512 | 0.988352175 |
| SLC16A9 | 1.115062088 | 1.141486811 |
| NTN1 | 1.114994622 | 1.049674546 |
| FBXW11 | 1.114911042 | 0.948734588 |
| ATP1A1 | 1.114891696 | 1.135626217 |
| PRMT5 | 1.114844273 | 0.855002025 |
| CCDC110 | 1.114787118 | 0.958687728 |
| KIAA0319L | 1.114787013 | 0.907845152 |
| LRFN4 | 1.114747186 | 0.970537855 |
| ANKHD1 | 1.114746552 | 0.927372388 |
| SEMA4D | 1.114732714 | 0.929121102 |
| MED24 | 1.114687205 | 1.044776119 |
| MTERFD2 | 1.114612801 | 0.869987849 |
| OR2T1 | 1.114591865 | 1.026378897 |
| MSANTD3 | 1.114567821 | 1.025516403 |
| GAB1 | 1.114508223 | 0.979102432 |
| UBQLN3 | 1.114437087 | 0.954093868 |
| CNGB3 | 1.114419243 | 1.040233615 |
| ZNF133 | 1.114416199 | 0.931825968 |
| SEMA4D | 1.114376133 | 1.079384366 |
| C22orf23 | 1.114363187 | 1.055841041 |
| STX17 | 1.114327846 | 0.969510106 |
| NSUN4 | 1.114253214 | 1.020251114 |
| GCC2 | 1.114251227 | 0.944501542 |
| C1orf53 | 1.11418959 | 1.016605913 |
| UBASH3A | 1.114188655 | 1.011680727 |
| BET1L | 1.114109512 | 0.935196436 |
| PIGZ | 1.113995278 | 1.045425049 |
| FEM1C | 1.113854723 | 1.047961631 |
| BCL7A | 1.113841674 | 1.006166495 |
| MADCAM1 | 1.113789976 | 1.003425831 |
| DLX1 | 1.113772594 | 0.829890644 |
| ENDOV | 1.113755404 | 0.91983556 |
| SENP1 | 1.113696362 | 1.064892927 |

| | | |
|---|---|---|
| SH2D4B | 1.113689887 | 1.059266872 |
| NUDT15 | 1.113686154 | 0.954574951 |
| HIP1 | 1.1136483 | 1.061563386 |
| HMGA1 | 1.113640053 | 0.868267359 |
| CSNK1G2-AS1 | 1.113621862 | 0.835560956 |
| ASB13 | 1.113579052 | 0.916288125 |
| TMEM82 | 1.113558645 | 1.106115836 |
| GPALPP1 | 1.113540909 | 1.010620075 |
| SEPT10 | 1.113492609 | 1.088729017 |
| CELSR3 | 1.113490472 | 0.853990915 |
| DCUN1D2 | 1.113436468 | 0.902011681 |
| MADD | 1.113408149 | 1.084360805 |
| SLC25A47 | 1.113347573 | 1.020554985 |
| AKAP5 | 1.113304174 | 1.046073978 |
| CACNA1H | 1.113304174 | 1.046073978 |
| GSPT1 | 1.113262173 | 0.856217092 |
| FRAS1 | 1.113237923 | 0.955121617 |
| ADAM11 | 1.113116148 | 1.127839066 |
| TMEM14B | 1.113068601 | 0.946899623 |
| SYVN1 | 1.113043704 | 0.763140818 |
| ZNF488 | 1.113008113 | 1.16032888 |
| FXYD1 | 1.112929521 | 0.955223881 |
| PRKCB | 1.112888031 | 1.051265412 |
| EIF4A2 | 1.112861687 | 1.123296561 |
| COPS5 | 1.112782904 | 0.940947437 |
| ERC2 | 1.112781247 | 0.965978129 |
| BAZ1A | 1.112698467 | 0.968139774 |
| ATXN7 | 1.112666479 | 1.05508303 |
| HTR3D | 1.112646089 | 0.984926345 |
| SULT1A4 | 1.112627889 | 0.950681376 |
| CLCN4 | 1.112610931 | 1.07073329 |
| NBPF10 | 1.11248666 | 1.021871203 |
| SCN5A | 1.112476085 | 0.888384166 |
| UCP3 | 1.112476085 | 0.8403634 |
| OSR2 | 1.112472237 | 1.033915725 |
| ZNF703 | 1.112463126 | 1.00891049 |
| UCN2 | 1.112439885 | 0.955806783 |
| MAGEA2 | 1.112423755 | 0.859540939 |
| OR13H1 | 1.11237251 | 0.990750257 |
| KRTAP9-4 | 1.112264663 | 1.068448765 |
| WBP1L | 1.11225108 | 1.060637205 |
| UGT2B15 | 1.112195116 | 1.027903958 |
| DENR | 1.112117825 | 1.053785543 |
| BCL10 | 1.111991366 | 0.8935756 |

| | | |
|---|---|---|
| IGSF11 | 1.111930037 | 1.107735925 |
| SNRNP27 | 1.111928385 | 1.044193217 |
| SYBU | 1.111917511 | 1.118191161 |
| CTSD | 1.111853228 | 0.927319922 |
| XKR6 | 1.111834626 | 1.091130012 |
| RHPN1 | 1.11180972 | 0.927029805 |
| TUBAL3 | 1.111808361 | 1.081116158 |
| SIX3 | 1.111793132 | 1.057105775 |
| EBF1 | 1.111717158 | 1.017010936 |
| SPOCK1 | 1.11171619 | 0.892669097 |
| ZFAND2A | 1.111683639 | 1.004050223 |
| ADAMTS6 | 1.111645361 | 1.042829332 |
| GSTA3 | 1.111605188 | 0.994808566 |
| SP6 | 1.111603788 | 0.991435423 |
| LY6H | 1.111595405 | 1.027749229 |
| CASQ2 | 1.111582447 | 0.953751285 |
| FSIP2 | 1.111578369 | 0.971908188 |
| ARHGAP10 | 1.111559293 | 0.916067146 |
| RP13-36C9.1 | 1.111551908 | 0.952380952 |
| FAM64A | 1.111541985 | 1.006851662 |
| LRRC75B | 1.111501025 | 0.877012676 |
| SLC25A20 | 1.111493393 | 1.028552888 |
| MREG | 1.111422061 | 0.891049008 |
| ANGPT4 | 1.111399033 | 0.927372388 |
| HHEX | 1.111307904 | 0.98582422 |
| PRAME | 1.111307508 | 0.9777321 |
| TRIM51EP | 1.11125021 | 0.942245295 |
| ZNRF3 | 1.11125021 | 0.942245295 |
| MTPN | 1.111219789 | 0.991778006 |
| CYS1 | 1.111077381 | 1.110654334 |
| VSIG1 | 1.111075478 | 1.003083248 |
| CNEP1R1 | 1.111059163 | 0.930336168 |
| PDE4A | 1.111046131 | 1.009733939 |
| OR14I1 | 1.110987287 | 0.948967193 |
| DNAJB1 | 1.110971945 | 0.962658445 |
| HOXA11 | 1.110838338 | 0.963953017 |
| PKD2 | 1.110830938 | 1.0532122 |
| ANP32E | 1.110826773 | 0.920863309 |
| ZNF431 | 1.110826184 | 0.973963686 |
| NCLN | 1.110679659 | 1.010530579 |
| TRPM1 | 1.110672383 | 1.120700844 |
| GPR52 | 1.110656824 | 0.793640493 |
| RPL39L | 1.110653056 | 0.983898595 |
| KIF18A | 1.110632868 | 1.096690461 |

| | | |
|---|---|---|
| C21orf59 | 1.110593978 | 1.177804779 |
| NDUFB4 | 1.110591624 | 1.072680078 |
| KCNA4 | 1.110475137 | 0.721609345 |
| UNC79 | 1.110462076 | 0.905789654 |
| NOD2 | 1.11043046 | 0.945727015 |
| NTNG1 | 1.110428015 | 1.09437019 |
| SPP1 | 1.110400813 | 0.955042527 |
| PLEKHA7 | 1.110390523 | 1.018226002 |
| MIB1 | 1.110378572 | 0.894873459 |
| RRP1B | 1.110315386 | 0.982989064 |
| MUC4 | 1.110283874 | 1.054128126 |
| RAB11B | 1.110261848 | 0.970028352 |
| ANKS1A | 1.110234646 | 0.979343864 |
| SFTPA1 | 1.110211455 | 1.020116807 |
| GCAT | 1.110156875 | 0.996106424 |
| CST3 | 1.110120986 | 1.080849606 |
| NCKAP1L | 1.1100859 | 1.066828676 |
| MAP4K3 | 1.110080891 | 1.145360156 |
| CCDC170 | 1.110069469 | 0.992805755 |
| NUDT17 | 1.11003816 | 0.991435423 |
| C7orf25 | 1.110031182 | 1.008221994 |
| C1orf168 | 1.109951928 | 1.12272175 |
| IFT122 | 1.109859624 | 1.128392062 |
| DCAF8 | 1.109847297 | 1.024301337 |
| RPS6KB2 | 1.109809883 | 1.083062946 |
| RTN1 | 1.1098011 | 1.065775951 |
| UPK1A | 1.109778989 | 1.014045906 |
| ANAPC11 | 1.109736286 | 1.001297859 |
| ZNF605_pseudogene | 1.109733181 | 0.860226105 |
| KY | 1.109692693 | 0.976103686 |
| FAM195A | 1.109627133 | 1.044957473 |
| RAB3GAP1 | 1.109622761 | 1.057553957 |
| SPINT1 | 1.109622761 | 1.007194245 |
| PIWIL2 | 1.109619881 | 1.011031798 |
| ESR2 | 1.109573944 | 1.044776119 |
| ZNF563 | 1.109531055 | 1.036656389 |
| ARID3C | 1.109449403 | 1.179019846 |
| DEFB115 | 1.109438668 | 1.032545392 |
| CHERP | 1.109432552 | 0.998972251 |
| FAM227B | 1.109332597 | 0.879708384 |
| GRIN2D | 1.109283673 | 1.040233615 |
| KCNRG | 1.109249509 | 1.073977936 |
| PHTF1 | 1.10923502 | 0.890979883 |
| ASIP | 1.109220186 | 1.022953066 |

| | | |
|---|---|---|
| KRT74 | 1.1091648 | 1.013770757 |
| ARL17 | 1.109127716 | 0.931956258 |
| GPR61 | 1.109109806 | 0.900713822 |
| KCTD10 | 1.109017522 | 1.127190136 |
| CTSO | 1.108920216 | 1.136924075 |
| RTBDN | 1.108770624 | 1.003768414 |
| BPNT1 | 1.108730378 | 0.963659961 |
| MS4A6A | 1.108689384 | 0.984241178 |
| CDH2 | 1.108638999 | 0.740664611 |
| APH1A | 1.108620606 | 1.098015391 |
| PLAC1 | 1.108514126 | 1.057211374 |
| GP2 | 1.108512404 | 0.912983899 |
| LIN7C | 1.108477743 | 1.023638232 |
| CALR | 1.10844661 | 0.824140169 |
| MXD1 | 1.108420564 | 0.787363305 |
| ZNHIT3 | 1.108420564 | 0.787363305 |
| MLEC | 1.108303618 | 0.984583762 |
| SIX5 | 1.108303618 | 0.984583762 |
| MRPL39 | 1.108302576 | 1.079821857 |
| CLHC1 | 1.10829229 | 1.047619048 |
| SAMD9 | 1.108238009 | 1.029462145 |
| CCL1 | 1.108235427 | 0.981843097 |
| SOCS4 | 1.108197253 | 1.046073978 |
| GLTSCR1L | 1.108172582 | 1.02672148 |
| RBP5 | 1.10815754 | 0.994518671 |
| ARFIP1 | 1.108087899 | 0.92874272 |
| CT47.7 | 1.108071013 | 0.929121102 |
| MID1IP1 | 1.10792349 | 1.018631025 |
| HTN3 | 1.107902447 | 1.03117506 |
| LUM | 1.107888537 | 0.968139774 |
| RYR1 | 1.107869524 | 0.940298507 |
| GRM7 | 1.107818566 | 1.085009734 |
| TP53INP2 | 1.107753016 | 0.931825968 |
| OR5H2 | 1.107683103 | 0.898252826 |
| CIART | 1.107659824 | 1.017010936 |
| ADAMTS5 | 1.107642641 | 0.926022064 |
| ATP8B4 | 1.107568946 | 1.03199676 |
| CEP44 | 1.107567926 | 0.955806783 |
| MST1 | 1.10751397 | 0.902011681 |
| NLRP9 | 1.10751397 | 0.902011681 |
| ECI2 | 1.107510266 | 1.046722907 |
| LRRC8E | 1.107489174 | 0.953827461 |
| DNAAF1 | 1.107482399 | 1.028351559 |
| FFAR1 | 1.107408597 | 0.91174562 |

| | | |
|---|---|---|
| KCNJ8 | 1.107408597 | 0.91174562 |
| HIST2H2BF | 1.107291169 | 0.862697448 |
| MIER2 | 1.10721211 | 0.980153908 |
| RAB1A | 1.10715644 | 1.085658663 |
| PPFIBP2 | 1.107122935 | 1.051914341 |
| GPATCH8 | 1.107102895 | 0.920615634 |
| ARPP19 | 1.107028675 | 0.734305387 |
| ITPR3 | 1.107008588 | 0.950681376 |
| DRAXIN | 1.106961587 | 1.034426893 |
| ZPR1 | 1.106902192 | 1.036313806 |
| SPOP | 1.106888191 | 1.065775951 |
| HAPLN2 | 1.106881803 | 1.005481329 |
| AHCY | 1.10686691 | 1.081116158 |
| SH2D6 | 1.106809479 | 0.918995545 |
| OSBPL2 | 1.106799294 | 1.077081192 |
| ITFG3 | 1.106778253 | 1.01782098 |
| OR1L8 | 1.106764813 | 1.030832477 |
| C17orf97 | 1.106759603 | 0.896197328 |
| OLA1 | 1.106709178 | 0.915350344 |
| DDX58 | 1.106691479 | 1.023361454 |
| MROH7 | 1.10668004 | 0.878088295 |
| DNTT | 1.106560338 | 0.878650227 |
| SLC6A5 | 1.106413419 | 0.913730255 |
| TROVE2 | 1.106382448 | 0.885783718 |
| MRPL14 | 1.106362056 | 1.029804728 |
| FHOD3 | 1.106330702 | 1.087358684 |
| MYEOV2 | 1.106324427 | 1.089104901 |
| FGL1 | 1.106318314 | 0.999594978 |
| DISP1 | 1.106281722 | 0.938677629 |
| USP5 | 1.106235161 | 0.994808566 |
| TMEM120B | 1.106220872 | 1.024323398 |
| PON1 | 1.106202891 | 1.081765088 |
| WDR76 | 1.106190454 | 0.96800324 |
| PRSS16 | 1.106138761 | 1.048020766 |
| DENND1C | 1.10612953 | 0.992304577 |
| METTL7A | 1.1060954 | 1.035236938 |
| LOC81691 | 1.106008088 | 0.970028352 |
| FAM65B | 1.105982271 | 1.000685166 |
| HLA-DRB4 | 1.105945616 | 0.999314834 |
| MTMR7 | 1.105914924 | 0.990266061 |
| SERINC2 | 1.10581202 | 1.07654921 |
| DAAM1 | 1.105789343 | 0.950325454 |
| GJB6 | 1.105755381 | 1.009733939 |
| CEACAM5 | 1.105749199 | 0.891401165 |

| | | |
|---|---|---|
| C5orf54 | 1.105723236 | 0.99109284 |
| WIF1 | 1.10571141 | 0.947584789 |
| SLC29A2 | 1.105655029 | 0.889033095 |
| HIST1H4G | 1.105625322 | 0.958890031 |
| DOC2B | 1.105614539 | 1.130181569 |
| GPR98 | 1.105591723 | 0.985723556 |
| LINC00173 | 1.105579696 | 0.971565605 |
| DAZ4 | 1.105547662 | 0.956149366 |
| MRPL1 | 1.105517848 | 1.026378897 |
| TMEM248 | 1.10548585 | 1.080599433 |
| EMC7 | 1.105480332 | 0.925316889 |
| KCNA5 | 1.105454243 | 1.048669695 |
| PARM1 | 1.105425598 | 0.966084275 |
| ZNF687 | 1.105363237 | 1.015390846 |
| APIP | 1.105249844 | 1.045362495 |
| PIK3AP1 | 1.105223813 | 0.95018226 |
| AGPAT1 | 1.105190644 | 0.99091499 |
| OR1M1 | 1.105151656 | 0.956491949 |
| SRSF4 | 1.105150025 | 0.998629668 |
| OR8U8 | 1.105062791 | 0.961522884 |
| XIRP2 | 1.104961537 | 1.005823912 |
| LOC100134257 | 1.104926286 | 1.018499486 |
| FRY | 1.104922502 | 1.015795869 |
| B2M | 1.104894445 | 0.933160286 |
| ARHGDIB | 1.104856993 | 0.954232483 |
| HAUS8 | 1.104828365 | 0.978533819 |
| GBP5 | 1.104800597 | 0.972250771 |
| KIF3A | 1.104770549 | 1.049318624 |
| SPDYE3 | 1.104752871 | 0.950587282 |
| PRR23B | 1.104745229 | 0.92874272 |
| IKZF3 | 1.104726739 | 0.94127177 |
| PGP | 1.10472453 | 1.005840363 |
| LPCAT3 | 1.104703408 | 1.066423653 |
| ARL11 | 1.104669063 | 0.940047962 |
| UBE3D | 1.104643237 | 1.049674546 |
| KRTAP6-2 | 1.10462543 | 0.855772525 |
| YIPF6 | 1.104609617 | 0.979445015 |
| EPSTI1 | 1.104590632 | 1.0890716 |
| CD163 | 1.104590179 | 1.025308241 |
| KIF14 | 1.104577583 | 1.078520441 |
| FAM78A | 1.104544424 | 0.818144998 |
| MIF4GD | 1.10452201 | 1.092750101 |
| DEFB110 | 1.104497305 | 1.083434589 |
| C5orf42 | 1.104475773 | 0.892428914 |

| | | |
|---|---|---|
| CCDC153 | 1.1044673 | 1.056868791 |
| MASTL | 1.104460684 | 1.044776119 |
| SPTLC2 | 1.104455177 | 1.006885379 |
| CYB561A3 | 1.104368525 | 0.916409729 |
| SIPA1L3 | 1.104232365 | 1.106115836 |
| TSC22D4 | 1.104228748 | 1.048197651 |
| SPINK8 | 1.104212243 | 1.014580802 |
| ATF5 | 1.104209516 | 1.030499676 |
| CELA3A | 1.104209516 | 1.030499676 |
| CATSPER4 | 1.104202601 | 0.977287476 |
| OSTM1 | 1.104195578 | 1.073997945 |
| HIST1H2AG | 1.104169933 | 0.96437136 |
| FAM184A | 1.104156023 | 0.950325454 |
| CYP27C1 | 1.10412958 | 1.262816353 |
| FIBCD1 | 1.104109399 | 1.057211374 |
| DLX3 | 1.104043223 | 0.890330954 |
| TEX29 | 1.103914549 | 0.979343864 |
| SOCS6 | 1.103879647 | 0.856586632 |
| GPR18 | 1.103867134 | 0.972744971 |
| ERGIC3 | 1.103847002 | 0.960712839 |
| AKT3 | 1.103832805 | 1.03569111 |
| CDKL2 | 1.103744966 | 0.992212849 |
| SAP30BP | 1.103741097 | 0.87605451 |
| WHAMM | 1.103638402 | 1.050627785 |
| SLC2A7 | 1.103616657 | 1.012675574 |
| CFTR | 1.103608474 | 1.074626866 |
| KRTAP4-1 | 1.103598262 | 1.025351148 |
| VAMP8 | 1.103598262 | 1.025351148 |
| LOC441956 | 1.103570668 | 0.957862282 |
| AFF4 | 1.103560413 | 0.95909275 |
| TUT1 | 1.103539629 | 1.046982584 |
| NFKB1 | 1.103528538 | 1.210253082 |
| COX4I1 | 1.103513853 | 1.086269745 |
| ARHGEF25 | 1.103441626 | 1.019869818 |
| ERGIC2 | 1.103417984 | 1.082624544 |
| TRPV5 | 1.103412545 | 0.987670344 |
| TIMM21 | 1.103383498 | 1.073309032 |
| PTGES3 | 1.103364139 | 0.871512005 |
| GPR151 | 1.103352943 | 1.060350422 |
| WWTR1 | 1.10329369 | 1.006075334 |
| ITGA8 | 1.103244125 | 0.972458485 |
| ZFP91 | 1.103241494 | 1.052415211 |
| ALCAM | 1.103216363 | 0.987444309 |
| AKAP3 | 1.10319373 | 1.0266061 |

| | | |
|---|---|---|
| PTRH2 | 1.103150557 | 1.022953066 |
| GRIK2 | 1.103141619 | 1.036340039 |
| GABBR2 | 1.103076913 | 0.983127839 |
| TEX19 | 1.103059485 | 1.032887975 |
| CLUAP1 | 1.103059365 | 1.013770757 |
| ZFAND3 | 1.102924985 | 0.882948562 |
| PTH | 1.102903847 | 1.131551901 |
| GPR161 | 1.102866083 | 0.886437378 |
| WDR47 | 1.102849695 | 1.006480356 |
| GGA1 | 1.10284354 | 0.934566632 |
| UGDH | 1.102738235 | 0.978585334 |
| HBG2 | 1.102736557 | 0.924260834 |
| ZNF548 | 1.102709559 | 1.072093965 |
| TAS2R42 | 1.102662337 | 1.023491292 |
| C3orf84 | 1.1026599 | 0.941760877 |
| PDIA3 | 1.102638789 | 0.998053212 |
| NAPRT1 | 1.102592193 | 0.989874443 |
| GRIA3 | 1.102590547 | 1.007787151 |
| RAG1 | 1.102584538 | 1.153147307 |
| CA8 | 1.10254762 | 0.8327258 |
| RAB24 | 1.102528502 | 1.040424803 |
| ZMYND15 | 1.102493161 | 1.026378897 |
| PIWIL4 | 1.102384372 | 0.971565605 |
| H1FNT | 1.102363471 | 1.051842851 |
| LYPD3 | 1.102358021 | 1.175374646 |
| TRAK1 | 1.102337033 | 1.0818145 |
| BLOC1S4 | 1.102269028 | 1.102470636 |
| RB1 | 1.102250792 | 0.993924666 |
| POU4F3 | 1.102240094 | 0.939651681 |
| DYDC1 | 1.102202857 | 1.053867963 |
| FABP9 | 1.102191803 | 1.068853787 |
| NINL | 1.102191803 | 1.068853787 |
| RNF113B | 1.102170123 | 1.022712524 |
| E2F1 | 1.102059562 | 0.857432159 |
| ANKS3 | 1.102050444 | 0.956662616 |
| MTA1 | 1.102016178 | 0.81440623 |
| PCDHGC4 | 1.101995797 | 0.971908188 |
| SLITRK3 | 1.101984832 | 0.895854745 |
| PATE3 | 1.101947769 | 0.945186708 |
| CCDC13 | 1.101860488 | 0.988659376 |
| OR2T27 | 1.101836624 | 1.017129154 |
| ANTXR2 | 1.101822894 | 1.078574322 |
| CRLF2 | 1.101723437 | 1.047371836 |
| CD44 | 1.101680306 | 1.036998972 |

| | | |
|---|---|---|
| CMC1 | 1.10165081 | 1.060979788 |
| SGK1 | 1.101583652 | 0.994159637 |
| DDX31 | 1.101545454 | 1.003893576 |
| KBTBD3 | 1.101522674 | 1.044193217 |
| EFHC1 | 1.101515244 | 1.031517643 |
| AOC2 | 1.101467739 | 0.953422438 |
| LAPTM4A | 1.101465141 | 0.998379911 |
| CCDC174 | 1.10140926 | 0.914135277 |
| SPATA17 | 1.10135399 | 1.009720535 |
| FAM149A | 1.10124248 | 0.997944502 |
| ABCB8 | 1.101224229 | 0.795587281 |
| FAM163A | 1.10118548 | 0.996759822 |
| SPTA1 | 1.101165317 | 1.179856115 |
| GOLPH3L | 1.101162463 | 0.958547448 |
| LRWD1 | 1.101161421 | 1.0445358 |
| GPR75 | 1.101101979 | 0.926670993 |
| LTB | 1.101078278 | 1.041795135 |
| PAPPA | 1.101066003 | 0.936404932 |
| MYO1D | 1.101062465 | 1.065775951 |
| MAPK8IP3 | 1.101059791 | 1.10243234 |
| DFNB31 | 1.101024767 | 0.99109284 |
| RPL23A | 1.101023136 | 1.027749229 |
| MEF2C | 1.100941778 | 0.863724854 |
| CCL4L1 | 1.100921325 | 0.975676602 |
| TFRC | 1.100904979 | 0.873458793 |
| SLC13A4 | 1.100728954 | 0.901579587 |
| MAMSTR | 1.100717749 | 1.102774923 |
| COLGALT2 | 1.100717391 | 0.886593763 |
| OR6C65 | 1.100706694 | 1.005481329 |
| ZNF367 | 1.100687664 | 1.13806098 |
| PRND | 1.100602889 | 1.074683111 |
| DBF4B | 1.100584488 | 1.01417578 |
| FAM98C | 1.100572109 | 0.999189955 |
| PFDN2 | 1.100399268 | 0.947927372 |
| TCL1A | 1.10030371 | 0.921205892 |
| ALOX5 | 1.100291327 | 0.997569866 |
| C18orf21 | 1.10022981 | 1.06235012 |
| SULT1B1 | 1.100223178 | 0.966255678 |
| TMBIM4 | 1.100219807 | 0.937626569 |
| MAG | 1.100211233 | 0.942446043 |
| LYRM9 | 1.100206104 | 0.97326853 |
| HSD17B13 | 1.100197411 | 0.975989617 |
| GRIA4 | 1.100147441 | 0.995457495 |
| AQP12A | 1.100128216 | 0.827055488 |

| | | |
|---|---|---|
| AOC3 | 1.100099446 | 1.014925373 |
| FOXC1 | 1.100099446 | 1.014925373 |
| LEMD2 | 1.100076301 | 0.962315862 |
| NDUFB9 | 1.100053181 | 1.034393251 |
| CHAMP1 | 1.10005038 | 1.025921426 |
| RORB | 1.100008644 | 0.903309539 |
| MCCC1 | 1.099983139 | 1.016605913 |
| ZNF248 | 1.099937156 | 1.112024666 |
| USP50 | 1.099927505 | 1.088043851 |
| FBXO4 | 1.099924345 | 1.092796885 |
| ZNF678 | 1.099902297 | 0.814094775 |
| LURAP1L | 1.099847896 | 1.050017129 |
| ADHFE1 | 1.09984268 | 0.971447112 |
| MRPL50 | 1.099804942 | 1.048646797 |
| PRDM2 | 1.09975797 | 1.010382868 |
| PNN | 1.0997329 | 1.034600891 |
| RBMY1F | 1.099517138 | 1.002025111 |
| ZMYND12 | 1.099508436 | 0.945871874 |
| LRRC39 | 1.099467981 | 1.120291616 |
| MRPS15 | 1.09945044 | 0.862624186 |
| IGFBPL1 | 1.099446815 | 1.002398082 |
| RCN1 | 1.099446815 | 1.002398082 |
| TULP3 | 1.099445773 | 1.04898938 |
| GINS1 | 1.099428247 | 0.844471446 |
| ZNF8 | 1.099401188 | 1.001027749 |
| PYGL | 1.099396766 | 0.942081814 |
| SHB | 1.099396406 | 1.015574302 |
| SPIN1 | 1.099323209 | 0.989064399 |
| MMP13 | 1.09928988 | 0.865022713 |
| SH3PXD2A | 1.099279095 | 0.99473471 |
| KANSL2 | 1.099224792 | 1.021061158 |
| HSD11B1 | 1.099219 | 0.903958469 |
| HIST1H4C | 1.099180123 | 0.982870846 |
| UPK3BL | 1.099094125 | 0.899623159 |
| IMPG1 | 1.099078675 | 0.991435423 |
| PDE6H | 1.09905213 | 1.011031798 |
| RNF217 | 1.098884685 | 1.147306944 |
| SYT13 | 1.098865131 | 0.939362795 |
| OR4C3 | 1.098860781 | 1.053785543 |
| CRHR1 | 1.098857271 | 0.879948086 |
| C1orf216 | 1.098841343 | 0.881466256 |
| TCEAL5 | 1.09882914 | 1.015795869 |
| NR4A3 | 1.098694436 | 1.016223232 |
| GLE1 | 1.098616245 | 0.932511134 |

| | | |
|---|---|---|
| GPS1 | 1.098609131 | 0.966426859 |
| GDF3 | 1.098606911 | 0.9777321 |
| IL15 | 1.098583486 | 1.113394998 |
| CFHR3 | 1.098515349 | 1.040502228 |
| MON2 | 1.098515349 | 1.040502228 |
| IGSF9 | 1.098468354 | 0.871873929 |
| TAAR5 | 1.098447713 | 0.88513952 |
| HEPACAM2 | 1.098414436 | 0.904419322 |
| HSPA1A | 1.098367265 | 1.063183475 |
| TLDC2 | 1.09836233 | 0.903048989 |
| GIMAP2 | 1.098360336 | 0.869133265 |
| FAIM3 | 1.098355193 | 0.912920211 |
| EGR2 | 1.098308669 | 1.070084361 |
| SAPCD2 | 1.09828254 | 1.069886948 |
| NUTM1 | 1.098226401 | 1.000685166 |
| CTR9 | 1.098111648 | 1.014580802 |
| TBC1D9B | 1.098087808 | 1.04090725 |
| ITFG1 | 1.098062185 | 0.999594978 |
| ATP6V0D2 | 1.098051985 | 0.870863076 |
| CDH1 | 1.098048788 | 1.029119561 |
| ATF1 | 1.098038731 | 0.900064893 |
| TRPV2 | 1.098038731 | 0.900064893 |
| TMSB4Y | 1.098028036 | 1.095580678 |
| RHOD | 1.097996664 | 1.007138222 |
| CWF19L1 | 1.097993329 | 1.005138746 |
| TSPAN9 | 1.097968136 | 0.948612539 |
| LETMD1 | 1.097958725 | 1.131227217 |
| C1orf68 | 1.097880191 | 0.968482357 |
| OR6Q1 | 1.097761841 | 1.024301337 |
| LGR6 | 1.097643004 | 0.963659961 |
| MMS19 | 1.097643004 | 0.87605451 |
| HHATL | 1.097618343 | 0.926690968 |
| C1orf43 | 1.097491655 | 1.095990279 |
| SLC10A5 | 1.097469414 | 1.045220966 |
| ODAM | 1.097468222 | 1.075334143 |
| MTG1 | 1.097418607 | 1.034021871 |
| LRG1 | 1.097394206 | 0.861077359 |
| TAX1BP3 | 1.097366334 | 0.987324426 |
| ZNF461 | 1.097233999 | 1.041717294 |
| RIC8B | 1.097206797 | 0.942081814 |
| KLHDC7A | 1.097205778 | 0.968408262 |
| TMEM151A | 1.097204148 | 1.021061158 |
| PHF13 | 1.097145524 | 0.715768981 |
| HDAC9 | 1.096995861 | 1.100584036 |

| | | |
|---|---|---|
| SAE1 | 1.096946549 | 1.022712524 |
| POU2AF1 | 1.096933048 | 1.003244646 |
| C14orf182 | 1.096905626 | 1.028776978 |
| WDR44 | 1.096898966 | 0.931140802 |
| SARS | 1.096873282 | 0.925373134 |
| SMPD3 | 1.096848255 | 0.896171317 |
| C17orf49 | 1.096836207 | 1.036452005 |
| GPBP1 | 1.096820059 | 0.899149453 |
| ADCY3 | 1.096740389 | 1.027136493 |
| SP5 | 1.096728964 | 1.088386434 |
| STAC3 | 1.096654793 | 0.935594382 |
| ZC2HC1A | 1.096642839 | 1.01782098 |
| SMTN | 1.096635038 | 0.978074683 |
| CLRN3 | 1.096628881 | 1.04981774 |
| SLC30A2 | 1.096614006 | 0.965168084 |
| CASP8 | 1.096593629 | 1.018170019 |
| LRRC42 | 1.0965324 | 1.040502228 |
| WNT2B | 1.096527019 | 1.071599863 |
| GRB10 | 1.096511524 | 0.964028777 |
| PILRB | 1.096454841 | 1.08016444 |
| CD163L1 | 1.096373059 | 0.952207371 |
| ZNF524 | 1.096353031 | 0.981157931 |
| TMEM244 | 1.096346642 | 1.023638232 |
| DDX19B | 1.09634559 | 0.823491239 |
| AFAP1L2 | 1.096344686 | 0.957877683 |
| AMHR2 | 1.09626409 | 1.033095393 |
| ELOVL4 | 1.09626409 | 1.033095393 |
| C1orf109 | 1.096181066 | 0.959902795 |
| CCNB3 | 1.096054072 | 0.822600243 |
| ZNF430 | 1.096051948 | 1.015416238 |
| RASEF | 1.095887656 | 1.042480301 |
| C4BPA | 1.095884801 | 0.873929428 |
| BEND4 | 1.095881232 | 0.958282706 |
| TMEM132E | 1.095870891 | 0.926285946 |
| SLFN12 | 1.095825718 | 0.979883193 |
| DEFB135 | 1.095813575 | 0.777663583 |
| SELO | 1.095806095 | 1.044957473 |
| TSPAN12 | 1.0958027 | 0.882494005 |
| CARHSP1 | 1.095785256 | 0.966084275 |
| MTM1 | 1.095769903 | 1.140817651 |
| C1orf110 | 1.09576519 | 0.879303362 |
| MMRN1 | 1.095732289 | 1.017129154 |
| MAPK6 | 1.095703715 | 1.101881895 |
| TMEM204 | 1.095694383 | 1.073309032 |

| | | |
|---|---|---|
| MYO18B | 1.095686428 | 1.092147956 |
| HARS2 | 1.095670391 | 0.971648441 |
| EIF4ENIF1 | 1.09563267 | 1.062946139 |
| C1orf177 | 1.095442943 | 0.990684488 |
| FAM167B | 1.095418237 | 0.958687728 |
| PITPNA | 1.095309058 | 1.068137573 |
| RNF43 | 1.095287571 | 1.058403634 |
| LRRN3 | 1.095266371 | 1.045906132 |
| SCGB3A2 | 1.095266371 | 1.045906132 |
| KCTD16 | 1.095242391 | 1.085054678 |
| WDFY1 | 1.095153323 | 0.980815348 |
| DMBT1 | 1.095080045 | 0.925070879 |
| KIAA1683 | 1.095017802 | 1.08016444 |
| POGK | 1.094981872 | 1.027749229 |
| MED18 | 1.094921061 | 1.057108141 |
| FTH1 | 1.094910613 | 0.912394549 |
| ADPRHL1 | 1.094862454 | 1.024659312 |
| NCOA5 | 1.094845567 | 1.036452005 |
| CYB5RL | 1.094835567 | 0.972593354 |
| SLC7A6 | 1.094799395 | 1.127190136 |
| OXA1L | 1.094790257 | 1.012332991 |
| TEX43 | 1.094782147 | 0.971223022 |
| RNF103 | 1.094780872 | 0.995457495 |
| FAM19A1 | 1.094760036 | 0.99109284 |
| CNST | 1.09472489 | 1.076144188 |
| PTPN20B | 1.094697643 | 1.08825438 |
| KRT9 | 1.094687178 | 1.009592326 |
| BDH2 | 1.09463376 | 0.9467878 |
| NGFRAP1 | 1.094601815 | 0.986981843 |
| NBPF3 | 1.094565855 | 1.034831916 |
| RNF150 | 1.094547403 | 0.80532122 |
| TRIM55 | 1.094537936 | 0.917585983 |
| DESI2 | 1.094514354 | 1.002835156 |
| TICAM1 | 1.094452869 | 0.921830701 |
| RQCD1 | 1.094403449 | 0.961630695 |
| OR9I1 | 1.094303632 | 1.029804728 |
| ZNF337 | 1.094303632 | 1.029804728 |
| XKRY | 1.094256873 | 0.927714971 |
| TMEM97 | 1.094255018 | 0.989874443 |
| RGPD1 | 1.094206268 | 0.952207371 |
| MRPS36 | 1.094206171 | 1.017129154 |
| LEFTY1 | 1.094171272 | 1.078574322 |
| C9orf153 | 1.09413755 | 1.035236938 |
| TIPRL | 1.094101026 | 0.991899554 |

| | | |
|---|---|---|
| FCRL2 | 1.094045314 | 1.08890331 |
| QDPR | 1.09401728 | 1.086269745 |
| EVI2B | 1.094013787 | 0.961973279 |
| NDFIP1 | 1.093961409 | 0.950668037 |
| MFSD1 | 1.093942243 | 1.020212402 |
| C8A | 1.093900678 | 0.950587282 |
| TP53AIP1 | 1.093898565 | 0.979102432 |
| ID4 | 1.093876963 | 1.040233615 |
| SLC8A3 | 1.093805177 | 1.020765737 |
| GRIP1 | 1.093737126 | 0.935251799 |
| RP6-166C19.1 | 1.093734816 | 0.974991435 |
| ANKRD45 | 1.093715612 | 0.963953017 |
| FANCA | 1.093698089 | 1.041312272 |
| SOWAHA | 1.093695389 | 0.903604698 |
| ASPRV1 | 1.093692065 | 0.99156392 |
| SMG1 | 1.093692065 | 0.99156392 |
| SHQ1 | 1.093654382 | 0.992304577 |
| CAND2 | 1.093629664 | 1.064892927 |
| ZBTB26 | 1.093619386 | 1.009315512 |
| DCTN5 | 1.093594069 | 0.948967193 |
| TWSG1 | 1.093581669 | 1.090099349 |
| PPP6R1 | 1.093563556 | 1.037667072 |
| PTGR2 | 1.093554997 | 1.045425049 |
| SLAIN1 | 1.093528 | 1.132037262 |
| TMEM102 | 1.093528 | 0.90562981 |
| GRAP | 1.093499478 | 1.186707777 |
| RNF13 | 1.093477366 | 1.025957171 |
| SAMD4B | 1.093407883 | 1.045362495 |
| ATG4C | 1.093400558 | 0.923426347 |
| TCF25 | 1.093350167 | 1.079384366 |
| RAD51 | 1.09330918 | 0.903958469 |
| CCDC146 | 1.093295177 | 0.973963686 |
| RAB6C | 1.093269912 | 0.977287476 |
| GPR97 | 1.093244885 | 1.133679429 |
| IL1RAPL1 | 1.093236159 | 1.050616483 |
| ADH1A | 1.093225951 | 0.967553537 |
| LONRF1 | 1.093225951 | 0.967553537 |
| CABP2 | 1.093165458 | 0.926690968 |
| TMEM261 | 1.093157707 | 1.127440904 |
| ALPL | 1.09315293 | 1.031148605 |
| AAGAB | 1.0931457 | 0.901678657 |
| LIN54 | 1.092961035 | 0.957067639 |
| OR9G1 | 1.092819516 | 0.9331963 |
| ZNF577 | 1.092804746 | 1.039739637 |

| | | |
|---|---|---|
| RACGAP1 | 1.092771216 | 0.883521754 |
| NMB | 1.092769908 | 1.110166059 |
| ZNF763 | 1.092687945 | 0.92709599 |
| RABGAP1L | 1.092651676 | 0.821385176 |
| CKM | 1.092644008 | 0.997404283 |
| SYT1 | 1.09263273 | 0.924075276 |
| PCDHB13 | 1.092582547 | 1.024323398 |
| HNRNPH2 | 1.092561388 | 1.052757794 |
| LETM2 | 1.092490252 | 1.079821857 |
| AFF2 | 1.092428048 | 1.036857027 |
| CPNE7 | 1.092412268 | 0.88513952 |
| GSDMA | 1.092392288 | 1.082219522 |
| NRDE2 | 1.09238681 | 0.952038369 |
| TMEM180 | 1.092316846 | 0.993519644 |
| TREML2 | 1.092284909 | 0.882494005 |
| MAGOH | 1.092254143 | 0.900993491 |
| SMIM7 | 1.092240894 | 1.106925881 |
| CYSLTR2 | 1.092220702 | 0.919532771 |
| NCR2 | 1.092195608 | 0.95018226 |
| MED7 | 1.092154761 | 0.9733939 |
| PRY | 1.092150934 | 1.023896314 |
| LMNB2 | 1.09214753 | 0.908187736 |
| IRS4 | 1.092077436 | 0.935251799 |
| MTX1 | 1.091988715 | 1.007787151 |
| PDIA6 | 1.09197612 | 0.817001947 |
| C9orf47 | 1.091940465 | 1.046248715 |
| FGF9 | 1.091933559 | 0.998053212 |
| SPC25 | 1.091909926 | 0.997944502 |
| NPS | 1.091884009 | 0.864337102 |
| LRRC37B | 1.09186847 | 1.00526529 |
| TNFRSF13B | 1.091820035 | 0.978585334 |
| ADAMTS18 | 1.091786776 | 0.851395198 |
| ACP5 | 1.091722976 | 1.022712524 |
| NCS1 | 1.09163902 | 0.944158959 |
| RICTOR | 1.091622165 | 1.066461117 |
| FAM120C | 1.091575259 | 0.85054678 |
| IQGAP2 | 1.091575259 | 1.009315512 |
| OAT | 1.091575259 | 1.026326448 |
| S100A16 | 1.091549398 | 0.979787598 |
| GABRR3 | 1.091544993 | 0.87605451 |
| TEX12 | 1.091476721 | 0.874614594 |
| CAMTA1 | 1.091433858 | 0.977046934 |
| ZNF106 | 1.091432227 | 1.023980815 |
| FAM96A | 1.091426972 | 0.994329688 |

| | | |
|---|---|---|
| PALMD | 1.091387688 | 1.004110997 |
| IFRD2 | 1.091386601 | 0.910243234 |
| TBX19 | 1.091364458 | 0.96430889 |
| ZNF160 | 1.091300493 | 0.954574951 |
| VHL | 1.091248905 | 1.056868791 |
| TMEM33 | 1.091240975 | 1.075368277 |
| MSL2 | 1.091224097 | 0.998702141 |
| MALSU1 | 1.091203776 | 1.10243234 |
| CD81 | 1.091182798 | 0.943131209 |
| EIF2S1 | 1.091168364 | 0.935107073 |
| LBH | 1.091167846 | 0.980130182 |
| DCAF5 | 1.091135449 | 0.988694758 |
| PRSS3 | 1.091116963 | 1.044193217 |
| CCDC80 | 1.090985983 | 1.000405022 |
| OR10A4 | 1.090965085 | 0.966084275 |
| ZNF583 | 1.090961732 | 0.984583762 |
| IMPG2 | 1.090960418 | 0.826652963 |
| CCDC155 | 1.09093438 | 1.149858242 |
| ZDHHC22 | 1.09093133 | 0.993148338 |
| ACTR5 | 1.090897257 | 1.038711888 |
| ZNF555 | 1.090891638 | 1.094210346 |
| ZNF667 | 1.090890572 | 1.076144188 |
| NDUFS8 | 1.090887066 | 0.896171317 |
| DNAH12 | 1.090844018 | 1.000342583 |
| MUC2 | 1.090756933 | 0.970537855 |
| ZNF320 | 1.090699801 | 1.010125557 |
| AGTR2 | 1.090681258 | 0.920830629 |
| NEFM | 1.090674585 | 0.981773998 |
| EFCAB11 | 1.090666676 | 0.959232614 |
| GLYR1 | 1.090646106 | 1.023295649 |
| S100A13 | 1.090630166 | 1.091535034 |
| MGAT5 | 1.090606837 | 0.91109669 |
| ELF1 | 1.090594508 | 1.051842851 |
| ZNF878 | 1.090565681 | 0.9109284 |
| DNAJC13 | 1.090536457 | 0.919492977 |
| MAN2B1 | 1.090351115 | 0.963142973 |
| TBATA | 1.090337865 | 0.960945529 |
| JMJD7 | 1.090266729 | 1.050702295 |
| TGM1 | 1.090258034 | 0.916288125 |
| TYMS | 1.090252088 | 0.779667882 |
| PANX1 | 1.090226504 | 0.960415315 |
| ZSCAN18 | 1.090217041 | 0.976508708 |
| KCNG3 | 1.090178674 | 0.906554186 |
| LEMD3 | 1.090118702 | 0.946899623 |

| | | |
|---|---|---|
| AMDHD2 | 1.090079886 | 1.083062946 |
| PPP1R42 | 1.09005699 | 1.063720452 |
| SRA1 | 1.090018432 | 0.935594382 |
| FBXO34 | 1.090011311 | 1.073329007 |
| EPB41L2 | 1.089970041 | 0.880095923 |
| MMD | 1.089874891 | 0.914354231 |
| RNF185 | 1.089830612 | 0.91174562 |
| NMI | 1.089807808 | 1 |
| UHRF1BP1L | 1.089803529 | 0.994176088 |
| SPATA33 | 1.089802948 | 1.081004455 |
| RBM17 | 1.089786583 | 0.948612539 |
| C21orf62 | 1.08970498 | 1.009934909 |
| FAM218A | 1.089665516 | 1 |
| LAMTOR2 | 1.089665516 | 1 |
| TSPAN3 | 1.089660305 | 0.918807811 |
| AS3MT | 1.08966012 | 0.892277742 |
| CARS | 1.089645631 | 1.068786502 |
| CARD17 | 1.0895879 | 1.007194245 |
| MINK1 | 1.089572117 | 0.926670993 |
| ZNF826P | 1.089566741 | 0.871873929 |
| CDC42EP4 | 1.089551879 | 1.024323398 |
| ENOX2 | 1.08953108 | 1.009315512 |
| CDX2 | 1.089418299 | 0.888213852 |
| SELPLG | 1.089358362 | 0.966084275 |
| WSCD2 | 1.089342958 | 0.924260834 |
| PLEKHM1 | 1.08933548 | 1.010277492 |
| ATG7 | 1.089321854 | 0.941596366 |
| MED1 | 1.089321854 | 0.941596366 |
| CAP2 | 1.089275309 | 0.874957177 |
| PMP22 | 1.089269691 | 1.08016444 |
| ZCCHC8 | 1.089245767 | 1.026036314 |
| SPPL2C | 1.089234839 | 0.931862427 |
| KLK6 | 1.089218127 | 0.887735237 |
| WNT6 | 1.089215954 | 1.043165468 |
| CLPS | 1.089201319 | 1.051730045 |
| BTBD6 | 1.089200458 | 1.087358684 |
| AKNAD1 | 1.089148571 | 1.032401782 |
| RSBN1 | 1.08913935 | 0.96800324 |
| CCDC103 | 1.089128042 | 1.14418793 |
| NDUFB8 | 1.089081384 | 0.9777321 |
| WDR66 | 1.089079292 | 1.062778453 |
| NDUFAF6 | 1.089065895 | 0.998379911 |
| TMEM121 | 1.089065895 | 0.880923451 |
| ZC4H2 | 1.089065895 | 1.057108141 |

| | | |
|---|---|---|
| CNTNAP3 | 1.088985962 | 0.957862282 |
| TTLL12 | 1.088967937 | 1.017415958 |
| PPP2R3B | 1.088962925 | 0.90266061 |
| HS2ST1 | 1.08893156 | 0.981181051 |
| TMEM144 | 1.088923619 | 1.000405022 |
| DUSP11 | 1.088904887 | 1.059701493 |
| APOC4 | 1.088887502 | 0.87727825 |
| MNT | 1.088852201 | 0.865937627 |
| MT1F | 1.088822395 | 0.936622131 |
| CTRB1 | 1.088806346 | 1.128487995 |
| TUB | 1.088791034 | 0.804672291 |
| BZW1 | 1.088771606 | 0.883192732 |
| RHOBTB1 | 1.088766061 | 0.996759822 |
| MANSC1 | 1.088727471 | 1.005138746 |
| STRIP2 | 1.088711159 | 0.969510106 |
| DMRTC2 | 1.088684789 | 1.039397054 |
| ENOX1 | 1.088664384 | 1.063183475 |
| GK5 | 1.088664339 | 0.951979234 |
| SKOR1 | 1.088640901 | 0.904009721 |
| CEP120 | 1.088614463 | 0.945727015 |
| RFK | 1.088613051 | 1.064892927 |
| METTL7B | 1.088571643 | 1.099286178 |
| GML | 1.088543423 | 0.948269955 |
| TRIM69 | 1.088408066 | 1.0798183 |
| CEP97 | 1.088372651 | 0.988352175 |
| BGN | 1.088362683 | 0.865707434 |
| MRPL19 | 1.088356567 | 1.005481329 |
| PLEKHA5 | 1.088312591 | 1.038882139 |
| OLIG1 | 1.088256387 | 0.977046934 |
| RGS19 | 1.088254204 | 0.834523037 |
| YY2 | 1.088228744 | 0.968813285 |
| RIOK3 | 1.08820868 | 1.094743673 |
| SPINK13 | 1.088193905 | 0.975676602 |
| ZNF224 | 1.088170344 | 0.922918808 |
| SV2B | 1.088139955 | 1.029566626 |
| GLT6D1 | 1.088113299 | 1 |
| RAB3IP | 1.088069485 | 1.068516615 |
| SGPP2 | 1.088062638 | 1.040882544 |
| AFAP1 | 1.08799391 | 0.823005265 |
| MOCS1 | 1.087972165 | 0.883841661 |
| ANP32B | 1.087920643 | 1.013018157 |
| EMC8 | 1.087912762 | 0.925475901 |
| RNASEH2B | 1.087817962 | 0.980558931 |
| ELOVL3 | 1.087805375 | 0.993148338 |

| | | |
|---|---|---|
| NDUFS4 | 1.087803649 | 0.977287476 |
| PNMAL2 | 1.087762422 | 1.121616992 |
| PCDH17 | 1.08763261 | 0.920520726 |
| XAGE2 | 1.087630146 | 0.971243418 |
| C8orf82 | 1.08761573 | 1.079384366 |
| TRIM72 | 1.087572897 | 1.060999351 |
| PKP1 | 1.087561746 | 0.944844125 |
| ASAH2B | 1.087552411 | 1.062373431 |
| IWS1 | 1.087486902 | 1.045362495 |
| CNTNAP2 | 1.087486129 | 1.016872161 |
| KRT2 | 1.087465354 | 1.075739166 |
| SPANXN5 | 1.087392968 | 0.986634265 |
| CLDN20 | 1.087366597 | 0.974991435 |
| PCDHA3 | 1.087363652 | 0.932168551 |
| PDXDC1 | 1.087236735 | 1.019036047 |
| SLC39A1 | 1.087227515 | 0.982989064 |
| AGXT | 1.08717824 | 0.918883842 |
| HSDL1 | 1.087076019 | 1.015390846 |
| SLC25A37 | 1.087074265 | 0.943543154 |
| DST | 1.087055502 | 0.993833505 |
| ZNF169 | 1.087038556 | 0.95101062 |
| DYNC2H1 | 1.086981117 | 0.962332928 |
| NAPG | 1.086980741 | 0.966769442 |
| TRAPPC6B | 1.086959898 | 0.923946557 |
| ADAT1 | 1.086919992 | 1.096041531 |
| CRP | 1.086884548 | 1.023980815 |
| C9orf72 | 1.086875305 | 1.006851662 |
| JAKMIP1 | 1.086825138 | 1.085864723 |
| PXYLP1 | 1.086792555 | 1.01752109 |
| LRRFIP2 | 1.086767604 | 0.936964714 |
| HPS6 | 1.08675528 | 0.91983556 |
| GPN1 | 1.086748937 | 0.911270983 |
| SLC39A5 | 1.086732263 | 0.978417266 |
| WDR16 | 1.086723818 | 0.820170109 |
| KCND1 | 1.086604447 | 1.032446463 |
| TMEM158 | 1.086567992 | 0.882948562 |
| DEFB103B | 1.086553562 | 0.991435423 |
| CPXM2 | 1.086499402 | 1.15055159 |
| TXNRD3NB | 1.086432902 | 0.871531346 |
| MORN4 | 1.086429006 | 0.930113052 |
| PACSIN3 | 1.086391994 | 0.995139733 |
| RNF216 | 1.086376905 | 0.88513952 |
| FGFR1OP2 | 1.086350936 | 0.95909275 |
| SZT2 | 1.086344008 | 1.01417578 |

| | | |
|---|---|---|
| PTPRC | 1.08631648 | 1.037637897 |
| PEX2 | 1.086283222 | 1.003244646 |
| ADRBK2 | 1.086276965 | 1.096690461 |
| CDH9 | 1.08626648 | 0.960260363 |
| UTF1 | 1.086256595 | 0.942081814 |
| LRRC32 | 1.086241007 | 0.911705144 |
| ARPC5L | 1.086188956 | 1.000342583 |
| MGAM | 1.086152779 | 1.059943297 |
| PI3 | 1.086136125 | 1.007536828 |
| ILVBL | 1.086131829 | 0.959117456 |
| CLLU1 | 1.086111874 | 1.078979344 |
| C1orf61 | 1.086066792 | 0.963142973 |
| CLCN2 | 1.086058397 | 0.983776768 |
| DCP1A | 1.086045448 | 0.932766302 |
| PRDM12 | 1.086003779 | 1.031591738 |
| C14orf169 | 1.085984383 | 0.987667009 |
| IL6 | 1.085982944 | 0.816929931 |
| CRYL1 | 1.085982298 | 1.029119561 |
| CDCA8 | 1.085970201 | 1.020554985 |
| CYTL1 | 1.085964504 | 1.050627785 |
| CHRNA7 | 1.08593837 | 0.821544452 |
| IL7R | 1.085858516 | 1.057754705 |
| TMEM70 | 1.085830124 | 1.077359255 |
| ENDOG | 1.085810522 | 1.023361454 |
| ZBTB40 | 1.08578983 | 0.942486837 |
| WIPF3 | 1.085772714 | 1.041312272 |
| ZNF609 | 1.085676437 | 1.024301337 |
| FETUB | 1.085638942 | 1.005138746 |
| ZNF43 | 1.085623492 | 0.996574169 |
| KLHL34 | 1.085610368 | 0.963547995 |
| DAP3 | 1.085581004 | 0.944841012 |
| KCNK5 | 1.085581004 | 1.062946139 |
| MCU | 1.085576747 | 1.007290401 |
| TGOLN2 | 1.085558155 | 1.028091812 |
| OR2T11 | 1.085557204 | 1.093867763 |
| ADAT3 | 1.085526439 | 1.010962658 |
| HIST1H1E | 1.085478909 | 1.018156903 |
| SUPT16H | 1.085475466 | 1.112264763 |
| SCGB1A1 | 1.0854577 | 1.108140948 |
| ANKRD26 | 1.08542953 | 0.935196436 |
| ETV2 | 1.085415747 | 0.893799246 |
| LARP7 | 1.085367077 | 0.97326853 |
| DBNL | 1.085297086 | 1.079789388 |
| ING3 | 1.08528975 | 1.05508303 |

| | | |
|---|---|---|
| ADNP | 1.085279864 | 0.923946557 |
| POLE | 1.085196308 | 0.915639195 |
| PLAC8 | 1.085164098 | 1.068516615 |
| MAPRE1 | 1.085145747 | 0.978417266 |
| S100A8 | 1.0851055 | 1.138922641 |
| PRSS2 | 1.085094185 | 1.009934909 |
| PPM1J | 1.08508122 | 0.822193381 |
| SERPINA3 | 1.085062533 | 0.905905256 |
| OSBP2 | 1.085041173 | 0.935594382 |
| C9orf135 | 1.085021754 | 1.034426893 |
| CLEC5A | 1.085008776 | 1 |
| PPP2R3C | 1.084985688 | 1.028756582 |
| ZNF662 | 1.084912233 | 1.017415958 |
| PEMT | 1.084879693 | 1.004542505 |
| TMEM255B | 1.084837147 | 1.006075334 |
| IPCEF1 | 1.084834108 | 1.00445358 |
| IKZF1 | 1.084826512 | 0.778066191 |
| ADAMTS15 | 1.084729618 | 1.103179753 |
| PPIA | 1.084696112 | 1.127839066 |
| GJB5 | 1.084691916 | 0.920615634 |
| BCL6 | 1.0846883 | 1.027136493 |
| FTO | 1.084674573 | 0.96882494 |
| EMC2 | 1.084643319 | 1.084244633 |
| NRXN3 | 1.084625004 | 0.951695786 |
| PYGO1 | 1.084619431 | 1.016101405 |
| TBPL1 | 1.08453283 | 0.941676792 |
| OR52I2 | 1.084520248 | 0.917437479 |
| B4GALT1 | 1.084497631 | 0.8669695 |
| FOXD4L4 | 1.084468646 | 1.036857027 |
| RITA1 | 1.084468646 | 1.036857027 |
| KDR | 1.084465689 | 1 |
| C12orf42 | 1.08442285 | 0.968408262 |
| OR10P1 | 1.084416172 | 0.947584789 |
| SUV39H2 | 1.084395632 | 0.940947437 |
| DCST1 | 1.084382326 | 1.059266872 |
| ARR3 | 1.084369277 | 1.075025694 |
| ARGLU1 | 1.084356545 | 1.090784515 |
| GRAMD1C | 1.084348325 | 0.938031592 |
| AP2M1 | 1.084265926 | 1.000685166 |
| ERCC6L | 1.084153739 | 0.966426859 |
| RBPJL | 1.084137687 | 1.010530579 |
| SCAF1 | 1.084122793 | 1.013703323 |
| SKA3 | 1.084104805 | 0.933538883 |
| APH1B | 1.08409186 | 0.946138871 |

| | | |
|---|---|---|
| APOL4 | 1.084054783 | 0.956491949 |
| RMDN1 | 1.084041308 | 1.056298096 |
| BAGE2 | 1.084005169 | 0.852004111 |
| AQP5 | 1.083975035 | 0.930741191 |
| PPM1E | 1.083848594 | 1.084360805 |
| FUT8 | 1.08384135 | 0.877352369 |
| SYTL3 | 1.083807186 | 0.926670993 |
| ZIC5 | 1.083744684 | 1.061322371 |
| CAMK1G | 1.083723628 | 1.074626866 |
| IL1F10 | 1.083703543 | 0.949640288 |
| CES1 | 1.083686418 | 0.86761843 |
| HIST1H2AA | 1.083681341 | 1.059952038 |
| WFDC12 | 1.083594573 | 0.971223022 |
| C15orf53 | 1.083592942 | 0.986981843 |
| TOB2 | 1.083528195 | 0.891058582 |
| FHDC1 | 1.083490361 | 1.008564577 |
| ACER3 | 1.083489524 | 1.020656136 |
| NUP62CL | 1.083469148 | 0.963547995 |
| ADCK4 | 1.083466862 | 1.055158988 |
| OVCH1 | 1.083449327 | 1.129136924 |
| RARB | 1.083398596 | 1.009315512 |
| PDZD3 | 1.083386371 | 1.079789388 |
| CALML6 | 1.083381582 | 0.971243418 |
| SLC2A12 | 1.083372566 | 0.952207371 |
| GUCA2A | 1.083358239 | 1.021582734 |
| GANAB | 1.083343122 | 0.89509923 |
| LEPREL2 | 1.083338108 | 1.020765737 |
| KTI12 | 1.083332527 | 0.876063183 |
| SFPQ | 1.083326341 | 1.028776978 |
| TSC22D2 | 1.083312966 | 0.927501013 |
| LINC00482 | 1.083308789 | 1.087484812 |
| C9orf24 | 1.083263949 | 0.894141829 |
| TAGLN | 1.083193266 | 0.908530319 |
| OR4P4 | 1.083111754 | 0.891401165 |
| SNX21 | 1.083041811 | 1.05346294 |
| ZNF462 | 1.08303355 | 0.960260363 |
| SSH1 | 1.082930196 | 1.040882544 |
| BTN3A2 | 1.082928296 | 1.020554985 |
| PHF20L1 | 1.082923611 | 1.016223232 |
| RBAK | 1.082916778 | 0.99156392 |
| DPYSL2 | 1.082909616 | 0.966904607 |
| C9 | 1.082830699 | 0.971908188 |
| SLC38A3 | 1.082781274 | 1.006489293 |
| MRPS5 | 1.082738403 | 1.032202809 |

| | | |
|---|---|---|
| TMEM200B | 1.082647076 | 1.07290401 |
| STX19 | 1.08261865 | 0.929085303 |
| DICER1 | 1.082577186 | 1.048197651 |
| C1orf21 | 1.082508366 | 0.996574169 |
| PCYOX1L | 1.08250317 | 1.055893074 |
| C5orf56 | 1.082495408 | 0.780061665 |
| GPCPD1 | 1.082478769 | 1.036857027 |
| ZNF468 | 1.082433127 | 1.128392062 |
| ST6GALNAC1 | 1.08240909 | 0.888384166 |
| OR4D6 | 1.082395908 | 0.924974306 |
| SETBP1 | 1.082378101 | 0.970880439 |
| ABCA1 | 1.082363645 | 0.895909275 |
| PCDH20 | 1.082359796 | 0.955121617 |
| ZBED4 | 1.082337006 | 1.023980815 |
| SP7 | 1.082318206 | 1.115509409 |
| GPAT2 | 1.082216935 | 1.059952038 |
| TMEM189 | 1.082185218 | 0.982528263 |
| VCL | 1.082182349 | 0.957067639 |
| CLEC4A | 1.082179802 | 1.035236938 |
| IQCB1 | 1.082121785 | 0.912298732 |
| C5orf30 | 1.08203182 | 1.096395302 |
| ZNF627 | 1.081998905 | 1.085988352 |
| GRM6 | 1.081995087 | 0.933160286 |
| RNF145 | 1.081988748 | 1.160285529 |
| ANKRD2 | 1.081967024 | 1.058323208 |
| LRRC37A3 | 1.081952557 | 0.980153908 |
| LTBP3 | 1.081949654 | 0.985611511 |
| HIST2H2BE | 1.081928993 | 1.015758822 |
| TSC2 | 1.081911529 | 1.199870214 |
| GALNT9 | 1.081844682 | 1.036988968 |
| SEC14L1 | 1.081792572 | 0.898767034 |
| SESN1 | 1.081781102 | 1.066118534 |
| SORL1 | 1.081763923 | 1.076573653 |
| PRRC1 | 1.081741207 | 0.944106926 |
| OR6C1 | 1.081669187 | 1.056183625 |
| VSTM1 | 1.081639377 | 0.860267315 |
| TRIB1 | 1.081619034 | 1.066839714 |
| NUP210 | 1.081596103 | 0.819458719 |
| MAB21L2 | 1.081580592 | 1.091130012 |
| MYO3B | 1.081412403 | 0.943543154 |
| KLC4 | 1.081371126 | 1.140947752 |
| RNF128 | 1.081321189 | 1.047371836 |
| PRDM14 | 1.081303325 | 0.741077223 |
| GMFB | 1.08130295 | 1.033573141 |

| | | |
|---|---|---|
| FOXA2 | 1.08128011 | 0.80532122 |
| ICA1 | 1.081198058 | 0.986296677 |
| LIMK1 | 1.081168068 | 1.037637897 |
| RPA1 | 1.081154783 | 1.032401782 |
| TNF | 1.081118592 | 1.012978585 |
| OR13G1 | 1.081109478 | 0.992120589 |
| TRMT10C | 1.081090946 | 1.096951011 |
| NOL3 | 1.081071101 | 0.976361768 |
| ATG16L2 | 1.080974608 | 1.028756582 |
| TMEM37 | 1.080961519 | 1.067488644 |
| SMCO3 | 1.080920174 | 1.010962658 |
| STAT2 | 1.080894014 | 0.914341337 |
| CENPT | 1.080878025 | 0.950587282 |
| RGN | 1.080876269 | 1.174717369 |
| TCOF1 | 1.080853099 | 1.018170019 |
| LURAP1 | 1.080822623 | 0.98663926 |
| SLC7A13 | 1.080819676 | 1.121998702 |
| CSTF2 | 1.080808033 | 0.993833505 |
| KCNC4 | 1.080766166 | 1.09733939 |
| FRAT2 | 1.080763584 | 0.903391572 |
| GPR149 | 1.080762626 | 0.865022713 |
| ASH1L | 1.08074609 | 0.952612394 |
| LENG8 | 1.080735597 | 1.006075334 |
| OR13C5 | 1.08070586 | 0.969510106 |
| POLL | 1.080691005 | 1.00843608 |
| OBP2B | 1.080615632 | 0.94127177 |
| USP30 | 1.080492359 | 0.934458144 |
| MYH7B | 1.080429548 | 1.04455245 |
| REC8 | 1.080426339 | 1.01782098 |
| OR6K2 | 1.080325477 | 0.838396112 |
| TMEM140 | 1.080304031 | 1.019846092 |
| ZNF3 | 1.080259243 | 1.013018157 |
| CACNG3 | 1.080236416 | 1.043478261 |
| OR7G1 | 1.08022875 | 0.879410757 |
| PNMA2 | 1.08022875 | 1.086330935 |
| RBM11 | 1.08017278 | 0.835820896 |
| APBA2 | 1.080156934 | 1.143884892 |
| ZNF668 | 1.080156458 | 0.955042527 |
| C2orf47 | 1.080145149 | 0.928311057 |
| RNF157 | 1.080089387 | 0.994159637 |
| CC2D1A | 1.080067924 | 0.90927501 |
| PEG3 | 1.080019238 | 1.030489894 |
| TBC1D3 | 1.079999393 | 0.985954094 |
| SCN1B | 1.079996032 | 1.00908501 |

| | | |
|---|---|---|
| EVA1C | 1.079970259 | 1.073655361 |
| INA | 1.079951229 | 0.963001028 |
| LOC440295 | 1.07991779 | 1.05789654 |
| PITPNM1 | 1.079888156 | 0.945727015 |
| GRIN1 | 1.079839206 | 0.9201817 |
| NLRP3 | 1.079839206 | 0.9201817 |
| FAM117A | 1.079817655 | 0.940056703 |
| SCFD1 | 1.079785849 | 0.974648852 |
| CPED1 | 1.079703385 | 0.899965742 |
| ZNF281 | 1.079649756 | 1.112191171 |
| GDPD3 | 1.079609105 | 0.997569866 |
| PAPLN | 1.079565871 | 0.868448099 |
| SLC16A2 | 1.079558072 | 0.96495782 |
| EAF1 | 1.079526939 | 0.856217092 |
| IDO1 | 1.079514408 | 1.087484812 |
| ZNF239 | 1.079501395 | 1.158958547 |
| HCN4 | 1.079495013 | 1.138221934 |
| SETDB1 | 1.079464537 | 1.095180235 |
| HIST1H2BD | 1.079463154 | 1.114422748 |
| TTR | 1.07946266 | 0.940298507 |
| RELL2 | 1.079449561 | 0.910243234 |
| ZBTB3 | 1.079424867 | 0.946214457 |
| DCUN1D1 | 1.07940421 | 1.073977936 |
| PHLDB1 | 1.079335335 | 1.104487838 |
| RUNDC3A | 1.079316034 | 1.002398082 |
| RNF20 | 1.079315054 | 1.049318624 |
| SRY | 1.079287805 | 0.970149254 |
| CASP7 | 1.079237423 | 1.064243997 |
| PRLHR | 1.079237423 | 1.064243997 |
| OR1E2 | 1.079213181 | 1.072970195 |
| AP1AR | 1.07905997 | 0.965573107 |
| OTOF | 1.079030674 | 0.960945529 |
| LNPEP | 1.079024536 | 0.841661259 |
| OPA3 | 1.079024536 | 0.841661259 |
| MRPS24 | 1.079014589 | 1.032887975 |
| BBS12 | 1.079007502 | 0.97326853 |
| TAB2 | 1.078947349 | 1.002740665 |
| ZNF215 | 1.078941233 | 1.045906132 |
| CDC123 | 1.07888876 | 1.091940057 |
| PACS2 | 1.078877401 | 1.009720535 |
| COG2 | 1.078875398 | 0.979787598 |
| GFOD1 | 1.078873223 | 1.008564577 |
| SELL | 1.078866155 | 1.116478246 |
| CTDSP1 | 1.078864746 | 1.005191434 |

| | | |
|---|---|---|
| TTC21B | 1.078848059 | 0.84528149 |
| THUMPD1 | 1.078799957 | 0.978417266 |
| PAX2 | 1.078752457 | 1.074626866 |
| POLM | 1.078681332 | 0.937221547 |
| JAM3 | 1.078663485 | 1.083590271 |
| C1orf111 | 1.078602935 | 0.931551235 |
| MRPL24 | 1.078548126 | 0.880781089 |
| CXorf23 | 1.078493755 | 0.965741692 |
| DPT | 1.078469107 | 0.893799246 |
| ZC3H12B | 1.078378289 | 1.09437019 |
| ZNF84 | 1.078377264 | 1.049331963 |
| CD53 | 1.078368338 | 1.027749229 |
| ELAVL1 | 1.078359095 | 1.006166495 |
| COL6A3 | 1.078352706 | 0.991778006 |
| DPP4 | 1.078318367 | 0.921479559 |
| TPRG1L | 1.078295715 | 0.998784933 |
| CNPPD1 | 1.078292218 | 1.019184652 |
| CCDC101 | 1.078245284 | 0.895504253 |
| ZGPAT | 1.07820662 | 1.01417578 |
| MRVI1 | 1.078193171 | 0.974648852 |
| TM9SF4 | 1.07816014 | 0.92428914 |
| TBKBP1 | 1.077954874 | 1.060348319 |
| DIAPH2 | 1.077949541 | 0.956149366 |
| AOAH | 1.077916465 | 1.011031798 |
| LINC00521 | 1.077898024 | 0.983556012 |
| SNX7 | 1.077856657 | 0.940390545 |
| TRMT10B | 1.077855705 | 1.095923261 |
| NAP1L5 | 1.077835724 | 0.996574169 |
| SEL1L3 | 1.077796304 | 1.03117506 |
| SORBS3 | 1.077796304 | 1.03117506 |
| ZNF275 | 1.077776176 | 1.014580802 |
| ZNF623 | 1.077773423 | 1.08016444 |
| FOXD1 | 1.077755935 | 1.110317975 |
| RAB27A | 1.077733131 | 0.82284231 |
| LHFPL3 | 1.077696261 | 1.071599863 |
| TMEM126B | 1.077667775 | 0.911300122 |
| NPFF | 1.077648958 | 1.091812264 |
| MGC50722 | 1.077621012 | 0.98217902 |
| HGC6.3 | 1.077590282 | 1.161014046 |
| OSGIN2 | 1.077580719 | 0.989874443 |
| IQCD | 1.077532784 | 0.829080599 |
| SGCE | 1.077502743 | 1.025693731 |
| NUFIP2 | 1.07749249 | 0.899959498 |
| HSPA6 | 1.077473566 | 1.006489293 |

| | | |
|---|---|---|
| TFR2 | 1.077451174 | 1.075924724 |
| KLC3 | 1.077419644 | 1.021414666 |
| ELMSAN1 | 1.07740492 | 0.953751285 |
| ARHGAP21 | 1.0773771 | 0.883179171 |
| URGCP | 1.0773771 | 0.883179171 |
| GSTA5 | 1.077329747 | 0.981829981 |
| TRUB2 | 1.077315298 | 0.896197328 |
| CNGA2 | 1.077276649 | 0.996755354 |
| SEMA3E | 1.07724765 | 1 |
| GIMAP7 | 1.077229588 | 1.017010936 |
| MCTS1 | 1.077205966 | 1.020212402 |
| LAMTOR1 | 1.077176916 | 0.982584042 |
| ZBED2 | 1.077147743 | 0.984241178 |
| METTL21C | 1.077095184 | 0.997259335 |
| SLC25A28 | 1.077057441 | 0.975676602 |
| BPHL | 1.077026716 | 1.001946788 |
| VTI1B | 1.076983269 | 1.05789654 |
| TSPAN31 | 1.076972697 | 0.978938842 |
| SERPINH1 | 1.076902348 | 0.994329688 |
| WSB2 | 1.076883802 | 0.823491239 |
| ZNF846 | 1.076879918 | 0.923946557 |
| ZNF528 | 1.076858033 | 1.027749229 |
| PSMD12 | 1.076831694 | 1.059538275 |
| OR52E6 | 1.076796956 | 0.922576225 |
| CCDC172 | 1.076742856 | 0.970195272 |
| FEZF2 | 1.076717894 | 1.011745646 |
| ZNF382 | 1.076713722 | 0.921205892 |
| OR10J5 | 1.076710188 | 0.990407674 |
| OR6C76 | 1.076672251 | 1.048197651 |
| PPP1CC | 1.076670832 | 1.205710578 |
| ARMC4 | 1.0766637 | 0.934224049 |
| VPS28 | 1.0766637 | 0.96882494 |
| RGS7 | 1.076634148 | 1.006075334 |
| PPP1CB | 1.076611559 | 1.066839714 |
| KIAA0391 | 1.076599967 | 1.092345079 |
| RGS13 | 1.076590163 | 0.836245289 |
| RNF170 | 1.076572538 | 1.081765088 |
| B9D1 | 1.076570728 | 0.942891859 |
| TBX10 | 1.076554731 | 0.987667009 |
| MSH2 | 1.076534505 | 0.843608047 |
| P2RX3 | 1.076534505 | 1.012329656 |
| ZNF747 | 1.076456469 | 1.023491292 |
| SLC35B3 | 1.076442868 | 1.081004455 |
| LOC653111 | 1.076395708 | 0.888384166 |

| | | |
|---|---|---|
| OR52R1 | 1.0763256 | 1.045563549 |
| PCDHGA1 | 1.0763256 | 1.045563549 |
| GATC | 1.076307386 | 0.948955122 |
| DDX60 | 1.076261368 | 0.962332928 |
| PNMA5 | 1.076233939 | 1.036998972 |
| NUP160 | 1.076177302 | 1.042822885 |
| TCEAL7 | 1.076170491 | 0.956662616 |
| OR51E1 | 1.076105593 | 0.9145403 |
| LIMD2 | 1.076070852 | 0.893479141 |
| ARPC3 | 1.076019655 | 1.037637897 |
| HEBP2 | 1.076016997 | 0.903048989 |
| PKD1L2 | 1.076008953 | 0.93835172 |
| ARHGAP11B | 1.075997831 | 0.923261391 |
| PCDHGB5 | 1.075969557 | 1.018499486 |
| B4GALT4 | 1.075871574 | 0.882138518 |
| SLC24A4 | 1.075719917 | 0.958468527 |
| OMP | 1.075706852 | 0.979787598 |
| LRRC3 | 1.075683146 | 1.066423653 |
| SRSF12 | 1.075647814 | 0.937992463 |
| GNB2L1 | 1.075610097 | 0.889033095 |
| PRIMA1 | 1.075523258 | 1.037684138 |
| MED28 | 1.075516215 | 1.018631025 |
| ZBTB21 | 1.075498202 | 0.962658445 |
| GOLIM4 | 1.075391135 | 0.954093868 |
| PPP1R36 | 1.075381031 | 0.994518671 |
| TMEM8B | 1.075338938 | 0.939705379 |
| RASSF2 | 1.075276366 | 0.985954094 |
| TMC4 | 1.075262909 | 0.877697842 |
| FSCN2 | 1.075195307 | 1.004796163 |
| CCL20 | 1.075156167 | 1.013627515 |
| LRAT | 1.075097965 | 1.058403634 |
| PRR32 | 1.075002152 | 0.934224049 |
| ZSCAN16 | 1.074987609 | 1.101660591 |
| CD160 | 1.074955545 | 0.988968202 |
| SLC16A3 | 1.074921052 | 1.018818949 |
| CRYAA | 1.074902879 | 0.986296677 |
| CARD18 | 1.074888699 | 1.048669695 |
| PRMT9 | 1.074881447 | 0.905447071 |
| NRSN2 | 1.074849204 | 1.074929121 |
| GATA5 | 1.074808269 | 0.904607398 |
| ZBBX | 1.074801385 | 0.92428914 |
| FNDC5 | 1.074703872 | 1.048197651 |
| CTNNA3 | 1.074622594 | 1.061664954 |
| TPD52L1 | 1.074608095 | 0.934566632 |

| | | |
|---|---|---|
| NUCB2 | 1.074599259 | 0.967797191 |
| SUFU | 1.074534709 | 1.232965607 |
| DACT3 | 1.074503117 | 0.992709599 |
| TSC1 | 1.074494594 | 0.892771497 |
| NBPF7 | 1.074410762 | 1.038882139 |
| SLC35D1 | 1.074410762 | 1.038882139 |
| ZNF764 | 1.074405006 | 0.987039287 |
| DLL1 | 1.074372532 | 1.178455548 |
| LMO3 | 1.074370854 | 0.817745803 |
| ZFP36L2 | 1.074318929 | 0.989722508 |
| MBTPS1 | 1.074315071 | 0.870214147 |
| SPACA1 | 1.074309493 | 1.152288376 |
| PHAX | 1.074281412 | 0.916067146 |
| TAS2R1 | 1.074275885 | 1.062007537 |
| SLC35A3 | 1.074250612 | 0.989617132 |
| OR5AK2 | 1.074245618 | 0.908872902 |
| TKT | 1.074236864 | 1.019467878 |
| FKBP14 | 1.074217682 | 1.064243997 |
| MRI1 | 1.074217682 | 1.064243997 |
| ZNF407 | 1.074157309 | 0.907502569 |
| CTBP1-AS2 | 1.074061924 | 1.127582017 |
| ACKR3 | 1.073992528 | 1.099286178 |
| FGF6 | 1.07398693 | 1.129136924 |
| ZDHHC13 | 1.073964373 | 0.987444309 |
| C1QL3 | 1.073936102 | 1.077764277 |
| MED25 | 1.073929225 | 1.010530579 |
| RIC8A | 1.073897234 | 0.958687728 |
| SLC18A2 | 1.073853018 | 1.064398542 |
| LSG1 | 1.073792094 | 0.953017416 |
| PTP4A2 | 1.073781205 | 1.029850746 |
| SLFNL1 | 1.073728877 | 0.999189955 |
| CD274 | 1.073687086 | 0.939705379 |
| ESCO1 | 1.073675543 | 0.955042527 |
| SGSM3 | 1.073628501 | 1.12915382 |
| DTX3L | 1.073539421 | 0.960415315 |
| SEC24B | 1.07349366 | 0.989037342 |
| C12orf49 | 1.073443816 | 0.95909275 |
| SLC6A16 | 1.07338062 | 1.1596366 |
| GSE1 | 1.073372004 | 0.980472765 |
| MPND | 1.073354314 | 1.051437829 |
| NLRP8 | 1.073347114 | 1.070084361 |
| WFDC9 | 1.073337614 | 0.947242206 |
| RNF135 | 1.073321429 | 1.010382868 |
| GLRB | 1.073314514 | 0.995457495 |

| | | |
|---|---|---|
| LATS2 | 1.073284796 | 0.935756003 |
| SLC22A3 | 1.07327681 | 0.920830629 |
| DOCK10 | 1.073244774 | 0.997944502 |
| STX12 | 1.073189522 | 0.993924666 |
| GYG1 | 1.073179175 | 0.771576898 |
| MYBL1 | 1.073084915 | 1.000648929 |
| TMPRSS11A | 1.073084915 | 1.000648929 |
| CDK2 | 1.073052232 | 0.95587281 |
| MCM7 | 1.073052232 | 0.95587281 |
| PTPLAD1 | 1.073027736 | 1.080599433 |
| UTP3 | 1.072988186 | 0.915724563 |
| JADE3 | 1.072884128 | 1.03569111 |
| CELF3 | 1.07287298 | 0.969218307 |
| RBMX2 | 1.072851804 | 0.907159986 |
| PSG7 | 1.072833499 | 0.99109284 |
| EBPL | 1.072829301 | 0.975989617 |
| SULT6B1 | 1.072817505 | 0.939020212 |
| ZBED6CL | 1.072769114 | 1.041795135 |
| STBD1 | 1.072700651 | 0.969510106 |
| PRX | 1.072671257 | 0.880923451 |
| AMY1B | 1.072663769 | 0.891401165 |
| FBXO7 | 1.072615343 | 0.996106424 |
| HTR2A | 1.072579286 | 0.966255678 |
| HYLS1 | 1.072523683 | 1.098420413 |
| HCRT | 1.072487832 | 1.030489894 |
| TRIM61 | 1.072485931 | 1.075924724 |
| MEGF10 | 1.072474692 | 0.94655704 |
| IFI44 | 1.072462776 | 0.991435423 |
| NR2C1 | 1.072424758 | 0.931146213 |
| SLCO2A1 | 1.072424758 | 0.87727825 |
| APOBEC3G | 1.072389366 | 1.001297859 |
| CYP4F22 | 1.072389366 | 1.001297859 |
| PHKG2 | 1.072389366 | 1.001297859 |
| ILF2 | 1.072318853 | 1.025516403 |
| PLA1A | 1.07230589 | 0.787151201 |
| HNRNPUL2 | 1.072264139 | 0.904761905 |
| OR5K4 | 1.072264139 | 0.904761905 |
| MAF1 | 1.072257433 | 1.039397054 |
| GNAT2 | 1.072090615 | 0.961713173 |
| IGSF10 | 1.072089833 | 1.068516615 |
| SNX5 | 1.072065375 | 0.906439854 |
| DOCK7 | 1.072057436 | 0.997601918 |
| MAP1LC3C | 1.072026399 | 0.894826996 |
| ZNF785 | 1.071966092 | 0.946942082 |

| | | |
|---|---|---|
| TRAPPC5 | 1.071924271 | 0.989037342 |
| ANG | 1.071921112 | 0.862292426 |
| CCS | 1.071888676 | 0.981829981 |
| HERC5 | 1.071888676 | 0.981829981 |
| DUSP26 | 1.071887491 | 1.010530579 |
| JOSD2 | 1.071809349 | 1.05872823 |
| ARHGAP22 | 1.071790381 | 1.012332991 |
| RTN4 | 1.071755312 | 0.986296677 |
| GRIPAP1 | 1.071675703 | 1.016786571 |
| C19orf33 | 1.071670447 | 0.984926345 |
| OSBPL8 | 1.071664738 | 0.953066119 |
| ABHD14A | 1.071663357 | 0.877352369 |
| IRX3 | 1.071655495 | 1.086330935 |
| HAPLN4 | 1.071625001 | 1.009592326 |
| CCDC85B | 1.071607371 | 1.195625759 |
| VSIG10 | 1.071581685 | 1.034258308 |
| PRSS22 | 1.071526948 | 0.927319922 |
| SPZ1 | 1.071513489 | 1.072498987 |
| SCN7A | 1.071508672 | 1.022063595 |
| SYDE1 | 1.071486592 | 1.041717294 |
| ENKUR | 1.07148137 | 1.070572114 |
| MMP11 | 1.071475002 | 1.007138222 |
| NEXN | 1.07146823 | 0.988009592 |
| RAET1G | 1.071446164 | 0.974991435 |
| PLAU | 1.071329701 | 0.947436729 |
| GREM1 | 1.071300222 | 0.896882494 |
| KHDRBS1 | 1.071263596 | 1.107073329 |
| SPACA7 | 1.071245238 | 0.920615634 |
| MIER3 | 1.071220583 | 0.991899554 |
| MBTD1 | 1.07121726 | 0.965056526 |
| PICALM | 1.071129813 | 0.963686194 |
| HAUS6 | 1.071105527 | 0.931825968 |
| CASP4 | 1.071081579 | 1.032446463 |
| RXFP2 | 1.071056553 | 0.93770279 |
| SLC26A6 | 1.070980019 | 1.087016101 |
| SLC35F5 | 1.070954102 | 0.979787598 |
| POLR3C | 1.070938527 | 0.949777238 |
| XPO7 | 1.070872273 | 1.034943474 |
| KRTAP3-2 | 1.070837158 | 0.965399109 |
| SPEF2 | 1.070753064 | 0.982870846 |
| HHIPL1 | 1.070720758 | 1.088729017 |
| DAZ3 | 1.070600189 | 1.036656389 |
| MEA1 | 1.070600182 | 1.011745646 |
| ARL4C | 1.070569288 | 0.880095923 |

| | | |
|---|---|---|
| ZNF23 | 1.070553572 | 1.04455245 |
| PPM1A | 1.070548865 | 1.077871512 |
| IPO9 | 1.070520172 | 0.910585817 |
| ZNF263 | 1.070510431 | 0.954093868 |
| ITIH2 | 1.070431275 | 0.971565605 |
| DEFB114 | 1.070412773 | 0.863507493 |
| GALNT12 | 1.070408834 | 1.033095393 |
| OR8B3 | 1.070383473 | 0.939705379 |
| CPN1 | 1.070370059 | 0.953277093 |
| TAS2R45 | 1.070355364 | 0.944511948 |
| LYAR | 1.070349083 | 0.83967112 |
| THAP5 | 1.070346908 | 0.883179171 |
| FABP7 | 1.07033835 | 0.8327258 |
| METTL25 | 1.07033835 | 0.8327258 |
| C14orf80 | 1.070291767 | 0.894826996 |
| PRG2 | 1.070263824 | 0.753746456 |
| TM2D3 | 1.070254298 | 1.050627785 |
| SARM1 | 1.070241462 | 0.99473471 |
| SDSL | 1.070201955 | 1.038286827 |
| ARL5B | 1.070187363 | 1.116241393 |
| DUSP21 | 1.070155607 | 1.153147307 |
| HNRNPF | 1.070103369 | 1.028434395 |
| GOLM1 | 1.070057289 | 1.132922234 |
| CYFIP1 | 1.07003928 | 1.046982584 |
| LRRC75A | 1.070003724 | 1.031591738 |
| METTL2A | 1.070003724 | 1.031591738 |
| NSD1 | 1.069924773 | 0.963659961 |
| THAP1 | 1.069922493 | 0.957519699 |
| HDAC2 | 1.069903473 | 0.904414743 |
| EIF3L | 1.069897165 | 0.988009592 |
| HEXB | 1.069885752 | 1.013627515 |
| STRN3 | 1.069829427 | 1.104145255 |
| CATSPER2 | 1.069827549 | 0.998702141 |
| SPIN2B | 1.069783866 | 1.027946537 |
| XRCC6BP1 | 1.069767536 | 0.983776768 |
| CST6 | 1.069764408 | 0.948955122 |
| LRRC23 | 1.069762471 | 1.101255569 |
| MZT2A | 1.069748505 | 1.05344296 |
| SSBP4 | 1.069743838 | 0.979445015 |
| SPOCK3 | 1.069726575 | 1.034600891 |
| CYP3A43 | 1.069706276 | 1.040424803 |
| RNF24 | 1.069677556 | 1.018818949 |
| PRELID1 | 1.069665022 | 0.786958283 |
| PPFIA1 | 1.069628903 | 1.118754056 |

| | | |
|---|---|---|
| GPR113 | 1.069533491 | 1.038935756 |
| ODF3L1 | 1.069514902 | 1.082219522 |
| HORMAD1 | 1.069503334 | 0.914354231 |
| LYSMD3 | 1.069502269 | 0.854597003 |
| HMHA1 | 1.069498339 | 1.138870863 |
| MACF1 | 1.069398624 | 0.943473792 |
| UGT2B7 | 1.069394858 | 1.059052563 |
| KRT1 | 1.06935055 | 0.985611511 |
| PITRM1 | 1.069332128 | 1.04412719 |
| PTPN4 | 1.069290637 | 0.879299156 |
| C22orf31 | 1.069268046 | 0.997569866 |
| TNPO2 | 1.069268046 | 0.997569866 |
| ZNF658B | 1.069235495 | 1.055488052 |
| TRAPPC11 | 1.069221781 | 0.87975334 |
| SIGIRR | 1.069176063 | 0.952380952 |
| PCDHB4 | 1.069136825 | 1.025008565 |
| CNGA4 | 1.069132824 | 0.949383517 |
| RNASE4 | 1.069132824 | 1.099286178 |
| KRTAP17-1 | 1.069123368 | 0.847893114 |
| ZNF195 | 1.069120242 | 1.000342583 |
| TMUB2 | 1.069114818 | 1.017010936 |
| SURF6 | 1.069084447 | 0.95101062 |
| MAK | 1.069068858 | 1.084360805 |
| BMPR1B | 1.069062273 | 0.984425698 |
| RAB19 | 1.069025952 | 0.907502569 |
| PHF8 | 1.068989663 | 0.969500324 |
| LCNL1 | 1.068982727 | 1.016443988 |
| SPIN3 | 1.068952913 | 0.930798219 |
| AGTPBP1 | 1.068935412 | 1.054510058 |
| ZNF581 | 1.06892899 | 1.003425831 |
| ARHGEF11 | 1.068923553 | 0.972935937 |
| C18orf25 | 1.06890262 | 1.045563549 |
| SEPT3 | 1.068865849 | 1.039584685 |
| ANXA1 | 1.068833568 | 0.971565605 |
| FRRS1L | 1.068808104 | 0.946899623 |
| ZNF608 | 1.068801262 | 0.983213429 |
| SEH1L | 1.068798317 | 0.916409729 |
| RXRB | 1.06879431 | 1.024659312 |
| SLC35A4 | 1.068784267 | 0.924665857 |
| DDIT4 | 1.068691613 | 1.060294621 |
| MRTO4 | 1.068675051 | 0.984609154 |
| NEUROD6 | 1.068638782 | 0.90202124 |
| C16orf87 | 1.068637604 | 0.974648852 |
| SYP | 1.068609603 | 1.131551901 |

| | | |
|---|---|---|
| RALGPS1 | 1.068607156 | 0.949982871 |
| ARL5A | 1.068590149 | 0.894873459 |
| CDH5 | 1.068578565 | 0.997944502 |
| CEACAM6 | 1.068370187 | 0.928400137 |
| FOXR2 | 1.068288086 | 0.963343611 |
| C4orf47 | 1.068249301 | 0.980815348 |
| NPRL3 | 1.068246854 | 0.896539911 |
| FAM107A | 1.06821119 | 0.901984609 |
| CYC1 | 1.068188576 | 0.955223881 |
| U2SURP | 1.068171323 | 0.973621103 |
| INSL6 | 1.068133404 | 1.012150668 |
| GUCY1A2 | 1.068117861 | 1.025308241 |
| PDXP | 1.068067895 | 1.021871203 |
| ELOVL2 | 1.068001457 | 1.060979788 |
| CCDC28B | 1.067940697 | 1.041312272 |
| CD300LG | 1.067923751 | 1.005823912 |
| DONSON | 1.067869289 | 0.9331963 |
| GIPC1 | 1.067869289 | 1.088729017 |
| LHX6 | 1.067867751 | 0.714864318 |
| CPNE2 | 1.067847586 | 0.944844125 |
| RMND1 | 1.067839072 | 1.018226002 |
| UBXN11 | 1.067805553 | 1.128392062 |
| ZNF622 | 1.067804127 | 1.010530579 |
| TRPV1 | 1.067790873 | 1.085658663 |
| GSTA1 | 1.067767658 | 0.965606749 |
| GABRA2 | 1.067737939 | 0.845554835 |
| ARIH2 | 1.067726084 | 1.03569111 |
| SPAG8 | 1.067702142 | 1.145597807 |
| QSOX2 | 1.067680495 | 1.029971648 |
| DENND6B | 1.067662615 | 0.912110166 |
| LGALS9B | 1.067658871 | 0.94210346 |
| SPATA13 | 1.067639932 | 0.953751285 |
| CCNK | 1.067607354 | 1.096265844 |
| TNFRSF10B | 1.067595985 | 0.989469421 |
| KIRREL | 1.067588868 | 1.048304214 |
| ARRB1 | 1.067560096 | 1.006166495 |
| ZFR2 | 1.067551805 | 1.011990408 |
| UCP1 | 1.067546373 | 1.005840363 |
| UBR7 | 1.067520315 | 0.969852689 |
| CEP192 | 1.067513156 | 0.844809866 |
| COQ9 | 1.067511272 | 1.016605913 |
| KRTAP9-2 | 1.067511272 | 1.016605913 |
| ADAMTSL3 | 1.06750355 | 0.981500514 |
| WDR96 | 1.067503478 | 1.059133252 |

| | | |
|---|---|---|
| FANCE | 1.067466396 | 0.873929428 |
| DIRAS1 | 1.067393906 | 1.036046983 |
| RGS16 | 1.067382573 | 1.074683111 |
| MAP1B | 1.067318005 | 1.020656136 |
| IQCE | 1.067258354 | 0.987849332 |
| ALOX12 | 1.067255854 | 0.961064244 |
| RAB2B | 1.067255854 | 0.961064244 |
| MDM1 | 1.067229155 | 0.977389517 |
| DIEXF | 1.067217754 | 0.980153908 |
| MYH14 | 1.067209491 | 0.994861254 |
| CYB5A | 1.067135741 | 0.862292426 |
| NECAB1 | 1.067092003 | 0.928057554 |
| OR5B3 | 1.067023042 | 0.997944502 |
| SCEL | 1.06700872 | 0.685918235 |
| SUDS3 | 1.067003296 | 1.061563386 |
| CCL2 | 1.067002245 | 1.021240151 |
| CASS4 | 1.066977415 | 1.050359712 |
| TSNARE1 | 1.066946294 | 0.951397327 |
| TMEM208 | 1.06693274 | 0.996574169 |
| ZNF565 | 1.066927892 | 1.002398082 |
| NKAIN2 | 1.066900343 | 1.037341555 |
| TMEM86A | 1.066860307 | 1.073309032 |
| KLRB1 | 1.066858251 | 1.095580678 |
| FAM135B | 1.066820931 | 0.988254354 |
| STAG3L3 | 1.066804332 | 0.918122645 |
| COL9A3 | 1.066801568 | 1.05344296 |
| ARMC10 | 1.066799485 | 1.02308627 |
| CXXC5 | 1.066775742 | 1.094210346 |
| C8orf34 | 1.066774116 | 0.903199676 |
| RNF149 | 1.066753929 | 0.99156392 |
| PCDHB5 | 1.06675064 | 0.852689277 |
| ZBTB12 | 1.066747559 | 0.999657417 |
| SPATA1 | 1.066694592 | 0.934224049 |
| OR4C15 | 1.066683763 | 0.951695786 |
| OR6C74 | 1.066670035 | 0.974991435 |
| CCND3 | 1.066646497 | 0.976638546 |
| CCL28 | 1.066641354 | 1.027406646 |
| DEFB113 | 1.066612718 | 1.085645769 |
| UBL4B | 1.066603212 | 1.04455245 |
| SERPINB11 | 1.066594504 | 1.126413155 |
| HECTD4 | 1.06653569 | 1.121998702 |
| SPECC1 | 1.066529553 | 0.847306602 |
| XK | 1.066515851 | 1.086956522 |
| HRH3 | 1.066512393 | 0.926670993 |

| | | |
|---|---|---|
| MARK2 | 1.066471972 | 1.016872161 |
| NRG3 | 1.066468679 | 1.013018157 |
| FCRL6 | 1.066398585 | 0.918995545 |
| FGFR4 | 1.066363059 | 1.001946788 |
| LRRC52 | 1.066350753 | 0.893884164 |
| RPIA | 1.06633441 | 0.966904607 |
| IGDCC4 | 1.066333993 | 0.998379911 |
| KRT85 | 1.066329103 | 1.033211827 |
| CUL4A | 1.066310701 | 1.057105775 |
| PCIF1 | 1.066302302 | 0.940390545 |
| OVOL1 | 1.066301079 | 0.946214457 |
| HMGN3 | 1.066297635 | 0.963686194 |
| CRLS1 | 1.066270445 | 0.896820247 |
| CUL5 | 1.066143338 | 0.881894873 |
| OMA1 | 1.066122922 | 1.152498378 |
| HSPB2 | 1.066108105 | 0.966769442 |
| DGAT2L6 | 1.066098038 | 1.046982584 |
| TUBG1 | 1.066090569 | 1.117456197 |
| CHI3L2 | 1.066038591 | 0.952207371 |
| NOMO3 | 1.066019978 | 1.035286057 |
| GALNT5 | 1.066019524 | 1.047371836 |
| OR6C75 | 1.066017894 | 1.011990408 |
| FAM160B2 | 1.066014677 | 0.977046934 |
| NR6A1 | 1.06598643 | 1.102530824 |
| HNRNPU | 1.065926368 | 1.004796163 |
| SLC25A13 | 1.065893075 | 0.942245295 |
| CCDC81 | 1.06581909 | 1.03564196 |
| TRIM73 | 1.065791735 | 0.872160934 |
| FAM47B | 1.065749557 | 1.089914945 |
| GLCCI1 | 1.065737201 | 0.984583762 |
| C11orf88 | 1.065692933 | 1.099691675 |
| LOC652859_pseudogene | 1.065672778 | 1.002595717 |
| TLX3 | 1.06566978 | 1.041452552 |
| GALNTL5 | 1.065656478 | 1.057754705 |
| FBXL5 | 1.065619268 | 0.967553537 |
| ASAP1 | 1.065595744 | 1.069201781 |
| AP5Z1 | 1.065587271 | 1.051730045 |
| ABCB5 | 1.065585326 | 0.873228028 |
| KIAA1143 | 1.065569374 | 1.016786571 |
| PLEK2 | 1.065563076 | 1.005138746 |
| TCN2 | 1.065533927 | 1.024301337 |
| MRPL46 | 1.065490984 | 0.892669097 |
| C4orf51 | 1.065475085 | 1.009592326 |
| EVX2 | 1.065469437 | 0.999189955 |

| | | |
|---|---|---|
| SLC35E2 | 1.065456236 | 0.980472765 |
| OR6C70 | 1.065434215 | 1.083932854 |
| DPYSL5 | 1.065427867 | 0.917585983 |
| FZD6 | 1.065359374 | 0.772225827 |
| C1QTNF2 | 1.065357642 | 0.97327852 |
| SNRNP25 | 1.06531043 | 1.030147311 |
| ACSBG2 | 1.065289811 | 0.90266061 |
| CDY1 | 1.065248407 | 0.968408262 |
| PNISR | 1.065241831 | 0.948612539 |
| ODC1 | 1.065170524 | 0.998053212 |
| CST9 | 1.065096115 | 0.992463172 |
| SDC2 | 1.065026293 | 0.928400137 |
| MDFI | 1.065005541 | 0.9109284 |
| APOPT1 | 1.064989502 | 0.979445015 |
| NXF3 | 1.064983486 | 1.003244646 |
| KDM4C | 1.064963222 | 1.116478246 |
| C14orf79 | 1.06494972 | 1.024665982 |
| HIGD1B | 1.064922237 | 0.984204131 |
| ACAA2 | 1.064909313 | 0.837767683 |
| API5 | 1.064872467 | 0.960602946 |
| CCL25 | 1.064865988 | 1.029119561 |
| CCDC109B | 1.064863147 | 0.976508708 |
| OR10A6 | 1.064857243 | 0.948955122 |
| KCTD1 | 1.064829876 | 1.142857143 |
| PCDHA10 | 1.064812251 | 0.845495033 |
| OSBPL6 | 1.064752442 | 1.010277492 |
| COMMD5 | 1.064720453 | 0.986981843 |
| SETD5 | 1.064660645 | 0.946214457 |
| IRF2BP1 | 1.064631225 | 0.863075925 |
| TAS2R7 | 1.064592454 | 1.144570058 |
| LNX1 | 1.064572528 | 0.958468527 |
| ANO9 | 1.064552921 | 0.938031592 |
| FOXD4L3 | 1.064492229 | 0.904076739 |
| NUDT16 | 1.064482471 | 1.011745646 |
| PLSCR4 | 1.064478351 | 0.998702141 |
| OR5D13 | 1.064474694 | 1.006166495 |
| LRRC55 | 1.064454403 | 0.940056703 |
| IQGAP1 | 1.064453435 | 0.967193196 |
| IGSF6 | 1.064426032 | 0.977046934 |
| MKRN3 | 1.064396192 | 0.717715769 |
| SIRT2 | 1.064381127 | 0.963659961 |
| ZNF134 | 1.06437746 | 0.896882494 |
| PCSK4 | 1.064321772 | 0.908500973 |
| TAMM41 | 1.064272795 | 0.895512162 |

| | | |
|---|---|---|
| STARD3 | 1.064259066 | 1.036998972 |
| ZNF322 | 1.064226035 | 1.11579308 |
| CRYM | 1.064191701 | 0.968851395 |
| AP3S2 | 1.064177288 | 0.882138518 |
| ZNF454 | 1.064015527 | 0.95991778 |
| MXRA8 | 1.063989746 | 0.948269955 |
| ATF2 | 1.063980557 | 0.977318753 |
| TTC32 | 1.063928089 | 1.060753341 |
| C1orf220 | 1.06390379 | 1.070473876 |
| POM121L12 | 1.063859798 | 0.974648852 |
| MAPKAPK5 | 1.063819024 | 0.979234263 |
| CLEC14A | 1.063804112 | 0.990750257 |
| C20orf26 | 1.063803614 | 1.030147311 |
| CITED2 | 1.063795068 | 0.883841661 |
| ACTL8 | 1.063705888 | 0.94653706 |
| PGM2L1 | 1.063677412 | 0.993114621 |
| USO1 | 1.063668243 | 1.103685703 |
| NCBP1 | 1.063641994 | 0.999657417 |
| LRRC49 | 1.06356351 | 1.004860267 |
| PMF1 | 1.063548339 | 0.891743748 |
| C2CD4B | 1.063504266 | 1.107735925 |
| TLR4 | 1.063479441 | 0.969500324 |
| SLC46A1 | 1.06347516 | 0.833874108 |
| ZNF165 | 1.063452057 | 0.996916752 |
| PTGDS | 1.063441865 | 0.942891859 |
| PCP2 | 1.063313862 | 0.849606029 |
| C4orf33 | 1.063304021 | 0.96435804 |
| SPATA2L | 1.063292763 | 0.927501013 |
| C11orf82 | 1.063287764 | 1.100719424 |
| SYNRG | 1.063284547 | 1.067146283 |
| PCDHB10 | 1.063205391 | 1.038026721 |
| CDH19 | 1.063165111 | 0.992805755 |
| HRASLS2 | 1.063139284 | 0.953408702 |
| SRRT | 1.063130222 | 1.103802672 |
| PEX11G | 1.06312107 | 0.947584789 |
| CDCA3 | 1.063098415 | 1.092154848 |
| LCA5L | 1.063096512 | 1.030832477 |
| TPMT | 1.063079844 | 1.020116807 |
| SMNDC1 | 1.063068964 | 0.991435423 |
| HLA-DQB1 | 1.063032672 | 0.864373783 |
| ROBO3 | 1.06300109 | 0.940390545 |
| H2AFY | 1.062986138 | 1.023638232 |
| IL12A | 1.062976985 | 0.962315862 |
| CDADC1 | 1.062949526 | 0.96495782 |

| | | |
|---|---|---|
| DRD5 | 1.062882103 | 1.065541856 |
| DNAH10 | 1.062875809 | 0.988694758 |
| SMARCD1 | 1.062873882 | 0.955042527 |
| OR8U9 | 1.062861627 | 0.871873929 |
| BRD1 | 1.062828698 | 0.927906035 |
| ZNF512B | 1.062821312 | 1.096800324 |
| TIMM44 | 1.062736845 | 0.947927372 |
| TMEM232 | 1.062726106 | 0.836930456 |
| PAMR1 | 1.062718033 | 1.07073329 |
| NLGN4Y | 1.062665944 | 1.034831916 |
| DYNLT3 | 1.062628146 | 0.875658161 |
| NPAT | 1.062598741 | 0.962658445 |
| TOE1 | 1.062530641 | 0.867762933 |
| COL1A2 | 1.062498743 | 0.961288112 |
| SV2A | 1.062444354 | 0.794248684 |
| ASB18 | 1.062418459 | 0.940298507 |
| CKB | 1.062418459 | 0.940298507 |
| NAA25 | 1.062388778 | 1.136087485 |
| LOC653566_pseudogene | 1.062388278 | 0.981843097 |
| PRSS53 | 1.062130002 | 0.980963953 |
| TYW5 | 1.062089189 | 0.999657417 |
| MAT1A | 1.062074348 | 1.071382219 |
| WDR37 | 1.062041841 | 0.988009592 |
| NPC1L1 | 1.062023232 | 1.117051438 |
| DSCAM | 1.062019458 | 0.960260363 |
| BEND6 | 1.06201606 | 1.004110997 |
| TRAM1 | 1.061993359 | 0.976361768 |
| OR6C4 | 1.061915155 | 0.849263446 |
| CADM1 | 1.061906732 | 1.053057918 |
| DOK3 | 1.061906732 | 1.053057918 |
| CTCFL | 1.061889157 | 1.082219522 |
| SLC37A3 | 1.061866811 | 0.935756003 |
| JPH1 | 1.061845242 | 0.985268928 |
| E2F8 | 1.061834753 | 1.03564196 |
| FAM198B | 1.061770161 | 0.998784933 |
| BTBD7 | 1.061762598 | 1.071599863 |
| SPATA4 | 1.061750772 | 1.005823912 |
| DRD2 | 1.061708339 | 0.981181051 |
| AP4E1 | 1.0616824 | 1.114080164 |
| FANCL | 1.06166124 | 0.944501542 |
| LINS | 1.061617799 | 0.944511948 |
| C8G | 1.061603434 | 0.983394087 |
| UGT2B4 | 1.061536365 | 0.986372485 |
| OR9A4 | 1.061454356 | 0.941760877 |

| | | |
|---|---|---|
| INMT | 1.061446793 | 0.890238963 |
| PRDM6 | 1.061444755 | 0.899959498 |
| ACLY | 1.06144068 | 0.919400567 |
| ZFP82 | 1.061376922 | 0.984241178 |
| COX7B2 | 1.061191017 | 1.095580678 |
| FARS2 | 1.061176531 | 0.976638546 |
| ARNT | 1.061161833 | 0.935196436 |
| SPRY2 | 1.06115409 | 0.921548476 |
| CCDC140 | 1.061080991 | 0.995544755 |
| FAM75A7 | 1.061077578 | 0.976103686 |
| NDRG3 | 1.06103855 | 1.098663926 |
| CDK4 | 1.06103123 | 1.001946788 |
| DUSP2 | 1.06103123 | 1.001946788 |
| KIAA0232 | 1.060989571 | 1.016786571 |
| TRIM32 | 1.060957057 | 1.213497729 |
| TMEM55B | 1.060836733 | 1.030781693 |
| SERTM1 | 1.060781373 | 0.95991778 |
| LIMD1 | 1.060715946 | 1.0668037 |
| KIF20B | 1.06070309 | 1.012329656 |
| SIRPA | 1.06070309 | 1.012329656 |
| SLC6A18 | 1.06069021 | 1.044878383 |
| IFNA16 | 1.060674578 | 0.974648852 |
| TRPM6 | 1.060636408 | 1.077871512 |
| NRG1 | 1.060605948 | 1.057754705 |
| STRN | 1.060531898 | 1.047961631 |
| NUPL1 | 1.060510965 | 0.897225077 |
| TDRKH | 1.060469734 | 1.020212402 |
| APLP2 | 1.060408021 | 0.813284731 |
| CNTN4 | 1.060403536 | 0.976361768 |
| SZT2 | 1.060401724 | 0.928057554 |
| ZNF540 | 1.060294839 | 0.926687222 |
| CXCR2 | 1.060265459 | 0.962362103 |
| LRRC2 | 1.060247535 | 1.043850634 |
| PSKH2 | 1.060223392 | 1.027903958 |
| OPRD1 | 1.060179514 | 0.922128488 |
| TDRD7 | 1.060100732 | 1.079136691 |
| PHF21B | 1.060023594 | 1.012978585 |
| NOP9 | 1.060010521 | 0.928400137 |
| ZNF586 | 1.059968156 | 1.055841041 |
| PPYR1 | 1.059955759 | 0.907203115 |
| AGAP7 | 1.059937663 | 0.991435423 |
| KRTAP19-3 | 1.059912063 | 0.943131209 |
| SLC17A7 | 1.059901726 | 0.953827461 |
| IFT80 | 1.059897447 | 1.01417578 |

| | | |
|---|---|---|
| CD200 | 1.05988782 | 0.944106926 |
| CENPV | 1.059734031 | 1.018226002 |
| ZNF765 | 1.059725575 | 1.068853787 |
| TUBB3 | 1.059704384 | 0.887808829 |
| ANO4 | 1.05965987 | 0.977046934 |
| VTA1 | 1.059629858 | 1.111786148 |
| RHBDF2 | 1.059487939 | 0.973673552 |
| KAT8 | 1.05945712 | 1.004860267 |
| MIS12 | 1.059427522 | 0.915724563 |
| REEP2 | 1.059375412 | 1.098420413 |
| GRM1 | 1.059326975 | 0.948085659 |
| KRR1 | 1.059288457 | 1.04981774 |
| LEO1 | 1.05926477 | 0.907654921 |
| NWD1 | 1.05925988 | 0.948562171 |
| PRPF39 | 1.05925988 | 0.948562171 |
| VAMP4 | 1.05924932 | 0.945186708 |
| ANXA5 | 1.059247553 | 0.971565605 |
| HMGN2P46 | 1.059218109 | 0.929121102 |
| AMICA1 | 1.059193046 | 1.001215067 |
| SMARCA1 | 1.059152243 | 0.981773998 |
| TOMM40L | 1.059152243 | 0.940866748 |
| DEFB130 | 1.059140169 | 0.891058582 |
| CNBD1 | 1.059131154 | 0.972053463 |
| SFXN3 | 1.059109993 | 1.069886948 |
| TMCO3 | 1.059087906 | 0.952612394 |
| FCRL4 | 1.059030293 | 1.00526529 |
| ARMC1 | 1.058999117 | 1.06763872 |
| MYH2 | 1.058998917 | 0.958468527 |
| BIVM | 1.058988925 | 1.026036314 |
| SAP130 | 1.058973792 | 0.85748544 |
| WNT10B | 1.058959882 | 0.983556012 |
| AP5B1 | 1.058948192 | 1.020212402 |
| ABCA3 | 1.058929436 | 0.93835172 |
| SLC9C1 | 1.058920916 | 0.914696814 |
| DDX42 | 1.058906962 | 0.983776768 |
| CYP4V2 | 1.058888837 | 1.087701268 |
| RPN2 | 1.058807786 | 0.948967193 |
| CLPTM1 | 1.058801298 | 1.049674546 |
| OR51L1 | 1.0587071 | 1.011647825 |
| OR2G2 | 1.05869908 | 0.911956149 |
| RPL10 | 1.058635465 | 0.989722508 |
| ACPP | 1.058629368 | 0.812459442 |
| ZP2 | 1.058622325 | 1.109969168 |
| PCDHGA5 | 1.058577016 | 0.994176088 |

| | | |
|---|---|---|
| TXN | 1.058576516 | 1.130434783 |
| EHBP1 | 1.058551506 | 0.986229243 |
| SRSF1 | 1.058535105 | 0.868105516 |
| ISCA1L_pseudogene | 1.058524411 | 0.915039397 |
| PPY | 1.058523794 | 1.007695423 |
| CTTN | 1.058523369 | 1.035286057 |
| FBXO24 | 1.058516338 | 1.019467878 |
| MXD4 | 1.058441424 | 1.181051265 |
| DHRS13 | 1.058438673 | 0.916160389 |
| SRPX | 1.058429669 | 0.986981843 |
| TOM1L1 | 1.058424912 | 1.033915725 |
| ATG2A | 1.058396321 | 0.970880439 |
| CLCNKA | 1.058345846 | 0.797534069 |
| RP6-166C19.10 | 1.058303754 | 1.006166495 |
| SNX27 | 1.058292879 | 1.016443988 |
| INO80D | 1.058227452 | 1.03117506 |
| TARS2 | 1.058215708 | 1.046982584 |
| ITGB1BP2 | 1.058189528 | 0.9109284 |
| C7orf33 | 1.058168187 | 0.931483385 |
| OR5AR1 | 1.058163838 | 1.045906132 |
| DUOXA1 | 1.058156465 | 0.943701904 |
| GAGE6 | 1.058137422 | 1.019527235 |
| DENND4C | 1.058067829 | 0.998379911 |
| OPCML | 1.058006446 | 0.87363305 |
| FRMD4A | 1.05798826 | 1.105710814 |
| TIMM17B | 1.057983324 | 1.090099349 |
| CERKL | 1.05795627 | 0.990684488 |
| ERICH4 | 1.057917444 | 0.969852689 |
| SOS2 | 1.057908654 | 1.05789654 |
| TCTEX1D1 | 1.057806455 | 1.047387606 |
| LRP10 | 1.057798712 | 0.920615634 |
| DYNC1LI2 | 1.057754683 | 1 |
| PABPC1L2B | 1.057744724 | 0.993490922 |
| ZWINT | 1.05768931 | 0.889345666 |
| EXOC5 | 1.057689191 | 0.795868773 |
| NUDC | 1.057683012 | 1.127098321 |
| TRIM24 | 1.057646264 | 0.863724854 |
| FAM217B | 1.057624518 | 0.904076739 |
| OR2J3 | 1.057602996 | 0.9554642 |
| TCEA1 | 1.057583039 | 0.889033095 |
| TWIST2 | 1.057566822 | 0.959766385 |
| ZNF101 | 1.05756566 | 0.908530319 |
| CORO1C | 1.057549971 | 0.905224787 |
| HCRTR2 | 1.057523355 | 0.914341337 |

| | | |
|---|---|---|
| HPGDS | 1.057479928 | 0.842851357 |
| SYK | 1.057475361 | 0.868916288 |
| TBCCD1 | 1.057472338 | 1.045767517 |
| IKZF5 | 1.057425779 | 1.100445525 |
| UNC45B | 1.05737092 | 1.055498458 |
| CEP19 | 1.05735487 | 1.102470636 |
| ZFP28 | 1.05731537 | 1.131894484 |
| CCDC60 | 1.057290736 | 0.932766302 |
| SLC38A8 | 1.057279485 | 0.930798219 |
| FSTL1 | 1.057278307 | 0.951353203 |
| FAM89A | 1.057248966 | 1.051842851 |
| DRG1 | 1.05720424 | 0.870392872 |
| TBCC | 1.057173279 | 1.021925317 |
| OR5D14 | 1.05717115 | 0.980815348 |
| BABAM1 | 1.057162775 | 0.903604698 |
| ZBTB44 | 1.057124315 | 0.936816525 |
| STAT6 | 1.057111245 | 0.975340688 |
| GJA5 | 1.057088112 | 1.071382219 |
| RSF1 | 1.05704975 | 1.096690461 |
| RECQL5 | 1.057028099 | 1.025957171 |
| OR6C68 | 1.057013563 | 0.983898595 |
| LSM11 | 1.057006767 | 0.953066119 |
| SLC52A1 | 1.05696584 | 0.962332928 |
| RNF182 | 1.056924042 | 1.005840363 |
| WDR91 | 1.056894172 | 0.941418294 |
| NEGR1 | 1.056868074 | 1.01781432 |
| CAPN13 | 1.056852256 | 0.960415315 |
| PTBP2 | 1.056845147 | 0.956149366 |
| ZBTB32 | 1.056809912 | 1.06399352 |
| CMYA5 | 1.056805888 | 0.966383151 |
| RPE | 1.056785206 | 0.92345079 |
| SLC8A1 | 1.0567808 | 1.011031798 |
| PARD3B | 1.056760653 | 0.956662616 |
| ZNF692 | 1.056749446 | 0.935196436 |
| TMEM63C | 1.056707166 | 1.077764277 |
| KRTAP6-3 | 1.056620457 | 0.943131209 |
| RSAD2 | 1.056589398 | 0.824140169 |
| TJP3 | 1.056560376 | 1.013703323 |
| DNASE1L3 | 1.056554822 | 0.84944841 |
| HCFC2 | 1.056554822 | 0.970798183 |
| PCDH18 | 1.056536975 | 0.984204131 |
| PNPLA5 | 1.056536975 | 0.984204131 |
| GH2 | 1.056536362 | 0.677286742 |
| BMPER | 1.056533327 | 1.04898938 |

| | | |
|---|---|---|
| LRRC59 | 1.056447116 | 1.019441069 |
| FXYD6 | 1.056424669 | 1.09733939 |
| C6orf47 | 1.056401385 | 1.006509078 |
| ZNF358 | 1.056387792 | 1.052072628 |
| RBL1 | 1.056316167 | 1.077222583 |
| PLXNA4 | 1.056298167 | 1.023491292 |
| PCMTD1 | 1.056295751 | 1.102530824 |
| SLC9A1 | 1.056246686 | 0.834523037 |
| ZC3HAV1 | 1.056229934 | 0.989037342 |
| ZCCHC12 | 1.056215084 | 1.091940057 |
| SAT2 | 1.056206424 | 0.970838396 |
| OR52E4 | 1.056203382 | 0.912641316 |
| CCNG2 | 1.056086485 | 1.036988968 |
| CUEDC2 | 1.056055356 | 1.062692703 |
| SLC25A3 | 1.056052737 | 1.041312272 |
| LAMTOR5 | 1.055986893 | 0.920178143 |
| SAMD9L | 1.055986159 | 1.109761037 |
| SFRP4 | 1.055930787 | 1.033616849 |
| MAGEB18 | 1.05590282 | 1.012150668 |
| ARHGAP20 | 1.055892422 | 0.893799246 |
| DHRS9 | 1.055818359 | 1.047371836 |
| ANKFN1 | 1.055807104 | 1.025921426 |
| PRAMEF7 | 1.055789705 | 0.937992463 |
| PRH2 | 1.055757536 | 0.988009592 |
| TNFAIP8 | 1.055744906 | 1.061158364 |
| PGAM5 | 1.055716736 | 0.838396112 |
| TYROBP | 1.055642404 | 0.916067146 |
| AGBL5 | 1.055612461 | 0.975293641 |
| FAM53A | 1.055608284 | 1.020251114 |
| GTF3C1 | 1.055600032 | 0.897124342 |
| CXADR | 1.055581515 | 0.880597015 |
| TTI2 | 1.05553814 | 0.887403807 |
| C4BPB | 1.055499226 | 1.050017129 |
| CTSS | 1.055426006 | 0.835171966 |
| CTSV | 1.055426006 | 0.809863725 |
| SLC47A2 | 1.055390362 | 1.026326448 |
| C15orf38 | 1.055316652 | 0.926690968 |
| SMIM13 | 1.055312142 | 1.082562521 |
| IKBKG | 1.05530071 | 1.093445814 |
| SRSF9 | 1.05525369 | 0.916970433 |
| KRTAP10-10 | 1.055198732 | 0.970880439 |
| SYCP1 | 1.055132354 | 0.940047962 |
| XPOT | 1.055131085 | 0.890030832 |
| DSCR3 | 1.055113686 | 1.029804728 |

| | | |
|---|---|---|
| RHBDD1 | 1.055090834 | 1.046172539 |
| SRCRB4D | 1.055035794 | 1.0532122 |
| POF1B | 1.054969738 | 1.032887975 |
| COL17A1 | 1.054947083 | 1.022610483 |
| ZNF75D | 1.054928597 | 1.127098321 |
| RUFY3 | 1.054881266 | 1.052247874 |
| HOOK3 | 1.054879753 | 0.927029805 |
| HSD17B3 | 1.05487174 | 0.835155934 |
| BCDIN3D | 1.054852892 | 1.007290401 |
| OTUD6B | 1.054753067 | 1.005138746 |
| BCAS1 | 1.054747947 | 0.940390545 |
| CAMP | 1.054725066 | 0.890373416 |
| PCNXL2 | 1.054712696 | 1.009592326 |
| INTS9 | 1.054674194 | 1.058323208 |
| ZP4 | 1.054645548 | 1.043508051 |
| RRM1 | 1.05463842 | 1.043478261 |
| RBM15 | 1.05463413 | 0.939020212 |
| PRDM1 | 1.054618745 | 1.018170019 |
| WWP1 | 1.054511642 | 1.048669695 |
| KEAP1 | 1.054488006 | 1.11579308 |
| SIAE | 1.054389235 | 0.939246659 |
| PFN1 | 1.054384383 | 0.965741692 |
| ABCC9 | 1.054360579 | 1.028552888 |
| GPR152 | 1.054292827 | 0.846203764 |
| UBE2S | 1.054244256 | 0.927319922 |
| ISOC1 | 1.054241375 | 1.271901363 |
| EPDR1 | 1.05419714 | 1.084244633 |
| TBC1D16 | 1.054176639 | 1.086673518 |
| GFRAL | 1.05417549 | 0.886593763 |
| OR51G1 | 1.054168347 | 0.948269955 |
| KRTAP19-4 | 1.054161324 | 0.963001028 |
| SLC23A3 | 1.054137794 | 1.051032807 |
| CCDC167 | 1.05412195 | 0.967454608 |
| CDH24 | 1.054104053 | 1.011647825 |
| IL22RA2 | 1.054094852 | 0.888618874 |
| SMOC1 | 1.054083573 | 0.873244262 |
| BSDC1 | 1.054071317 | 1.041312272 |
| NCAM2 | 1.054052037 | 0.848235697 |
| KCNK13 | 1.054040012 | 0.988319273 |
| AQP9 | 1.054028076 | 0.881894873 |
| RBBP7 | 1.054017881 | 0.855407047 |
| ANKRD28 | 1.053993271 | 0.896882494 |
| ITPR2 | 1.053965181 | 0.93770279 |
| PRAMEF9 | 1.053951858 | 0.901336074 |

| | | |
|---|---|---|
| GORAB | 1.053934696 | 1.057108141 |
| NUPL2 | 1.053934696 | 1.045362495 |
| SPINK6 | 1.053934696 | 1.068853787 |
| NID2 | 1.053910807 | 0.905789654 |
| TXN2 | 1.053869768 | 0.968202466 |
| ZNF665 | 1.053858235 | 0.929931146 |
| DFNB59 | 1.05385195 | 1.028091812 |
| CSTF1 | 1.053840894 | 1.116478246 |
| MTBP | 1.053792685 | 0.889688249 |
| STARD8 | 1.053792685 | 0.889688249 |
| MMP24 | 1.053783905 | 0.917585983 |
| FAM136A | 1.053778106 | 0.908464966 |
| TRIM38 | 1.053776907 | 1.0798183 |
| PTN | 1.053774108 | 1.160934458 |
| CECR6 | 1.053711536 | 0.986981843 |
| RASA1 | 1.053692279 | 0.948085659 |
| MAGEH1 | 1.05367565 | 1.006166495 |
| ZNF85 | 1.053651978 | 1.006480356 |
| IL18R1 | 1.053635723 | 0.841661259 |
| MOCS3 | 1.053618765 | 0.900769542 |
| RNF112 | 1.053606334 | 0.978585334 |
| CKAP2L | 1.053582037 | 1.008505468 |
| LOC284009 | 1.05358118 | 1.107913669 |
| CDH10 | 1.05354602 | 0.872216513 |
| TDRD12 | 1.053545767 | 0.9145403 |
| SNW1 | 1.053541298 | 0.872160934 |
| TMLHE | 1.053539858 | 1.081004455 |
| OR5K2 | 1.053535327 | 1.023980815 |
| DNAJB11 | 1.053532699 | 1.009249743 |
| PLEKHA8 | 1.053521877 | 1.034021871 |
| SSUH2 | 1.053497018 | 0.975293641 |
| PCDHGA3 | 1.053474859 | 0.928311057 |
| RASSF5 | 1.053454075 | 1.036046983 |
| PER1 | 1.053416389 | 0.978417266 |
| CNIH3 | 1.053392539 | 1.04981774 |
| ADAD2 | 1.053365416 | 0.938677629 |
| RIMS2 | 1.053354179 | 0.909215485 |
| PSD2 | 1.053339205 | 1.075334143 |
| CMTM4 | 1.053282254 | 0.981368975 |
| IL20 | 1.053263424 | 0.966769442 |
| NTRK1 | 1.053221473 | 0.994159637 |
| UBE2V2 | 1.05320402 | 0.801427644 |
| SPINK9 | 1.053192197 | 0.834189791 |
| RPL15 | 1.053185627 | 0.960945529 |

| | | |
|---|---|---|
| UBXN6 | 1.053167231 | 0.994861254 |
| PVRL3 | 1.053109414 | 1.034426893 |
| TFPT | 1.053109414 | 1.034426893 |
| CTAG1A | 1.053049101 | 0.963953017 |
| HADH | 1.052991589 | 0.905224787 |
| CABP5 | 1.0529695 | 1.021582734 |
| ZNF446 | 1.052953384 | 0.942486837 |
| S1PR3 | 1.052904281 | 0.923426347 |
| TRAPPC10 | 1.052858809 | 0.889003083 |
| CDY2 | 1.052824016 | 0.842310188 |
| RANBP17 | 1.052810328 | 0.961288112 |
| NOBOX | 1.052807237 | 0.94653706 |
| TSPYL4 | 1.052804709 | 0.995203837 |
| INTS4 | 1.052800258 | 0.752531389 |
| TRIM16 | 1.052790074 | 1.096951011 |
| NMD3 | 1.052778608 | 0.923045768 |
| ING5 | 1.052734978 | 1.071942446 |
| EXOG | 1.052703001 | 1.157040883 |
| NAMPT | 1.052606189 | 1.020116807 |
| RASA3 | 1.052573574 | 0.940866748 |
| CTDSP2 | 1.052557618 | 1.050616483 |
| FAM166A | 1.052542684 | 1.050017129 |
| DMGDH | 1.052538782 | 0.917375456 |
| CRCT1 | 1.052522787 | 0.856622114 |
| PRPF38A | 1.052518712 | 0.954637505 |
| NHLH2 | 1.052513923 | 1.101660591 |
| RPP21 | 1.052491348 | 1.005823912 |
| GANC | 1.052483053 | 0.882138518 |
| PAGE4 | 1.052483053 | 1.029161604 |
| TNS2 | 1.052466972 | 0.957177115 |
| THEM6 | 1.052463391 | 0.870392872 |
| PATL1 | 1.0524609 | 1.010277492 |
| OR51B5 | 1.052401046 | 1.019184652 |
| MAP1S | 1.052327525 | 1.060999351 |
| C4orf50 | 1.05230866 | 0.926344639 |
| C14orf142 | 1.052213419 | 1.084275437 |
| GGTLC1 | 1.052192758 | 1.069544365 |
| HSFX1 | 1.052153063 | 0.878493317 |
| TMEM56 | 1.052143079 | 0.953017416 |
| DDX20 | 1.052092444 | 1.004050223 |
| RND3 | 1.052069524 | 1.101881895 |
| U2AF1 | 1.052062493 | 1.072627612 |
| SDCCAG3 | 1.052051528 | 1.000342583 |
| OPHN1 | 1.052049196 | 0.943296881 |

| | | |
|---|---|---|
| C6orf222 | 1.052025407 | 0.931551235 |
| TRPM3 | 1.051880509 | 1.081765088 |
| ARFRP1 | 1.051877134 | 0.96495782 |
| RPGRIP1 | 1.051862093 | 0.961117861 |
| DNAJC28 | 1.05186082 | 0.897225077 |
| LEMD1 | 1.051858373 | 0.973963686 |
| KRT73 | 1.0518435 | 0.986634265 |
| RSRC1 | 1.051784918 | 1.025921426 |
| RIMBP2 | 1.051769483 | 1.051437829 |
| RD3 | 1.05176607 | 0.951397327 |
| UFSP2 | 1.05176006 | 1.01417578 |
| ZNF641 | 1.051692819 | 1.078979344 |
| TTN | 1.051684251 | 1.06164828 |
| MTFR1 | 1.051642999 | 1.055488052 |
| TCERG1L | 1.051611209 | 1.014045906 |
| THAP6 | 1.051606995 | 0.994861254 |
| HSFY1 | 1.051517431 | 0.882948562 |
| TIMM9 | 1.051412708 | 0.863651936 |
| KCNJ6 | 1.051410773 | 0.955223881 |
| SMIM19 | 1.051369858 | 1.026326448 |
| COX16 | 1.051366991 | 1.077766358 |
| ZPBP | 1.051364906 | 0.953751285 |
| PAOX | 1.051268683 | 1.021414666 |
| FANCG | 1.051259051 | 1.051914341 |
| FXYD7 | 1.051233037 | 1.143413368 |
| TRAPPC13 | 1.051196128 | 0.918465228 |
| ALOX15 | 1.051177881 | 0.935107073 |
| KISS1 | 1.051159518 | 1.075025694 |
| GTPBP10 | 1.051146485 | 1.097205346 |
| OR14J1 | 1.051112758 | 0.889003083 |
| ADRA2B | 1.051055713 | 0.940298507 |
| BRD3 | 1.051051926 | 0.970798183 |
| KIAA1841 | 1.051041345 | 0.934386391 |
| COL20A1 | 1.051018727 | 1.024706359 |
| POLR3F | 1.0509718 | 1.001370332 |
| SYNPR | 1.0509718 | 1.001370332 |
| SLC4A5 | 1.050971709 | 0.936411503 |
| EPHB3 | 1.050934863 | 1.006489293 |
| MTRF1 | 1.050934863 | 0.975989617 |
| DNAAF2 | 1.050906962 | 0.99109284 |
| IQSEC1 | 1.050883311 | 0.99554642 |
| FOLH1 | 1.050825702 | 1.042180402 |
| CDH26 | 1.050820723 | 1.00526529 |
| CBX4 | 1.050803301 | 0.954232483 |

| | | |
|---|---|---|
| ZC3H4 | 1.050800485 | 0.942446043 |
| VMA21 | 1.050749602 | 0.974991435 |
| IFNL1 | 1.050746197 | 0.981773998 |
| PRRC2A | 1.050680052 | 0.988352175 |
| C1QTNF1 | 1.050676835 | 1.153819801 |
| ADIG | 1.050641159 | 1.036857027 |
| IZUMO1 | 1.050628423 | 1.011340624 |
| EYA1 | 1.050590916 | 0.99156392 |
| SRR | 1.05056836 | 0.930564568 |
| C10orf113 | 1.050549823 | 0.987849332 |
| DDT | 1.050504971 | 1.057754705 |
| PROSER2 | 1.050499855 | 1.000342583 |
| DDX1 | 1.050484823 | 0.989874443 |
| SLC48A1 | 1.050399268 | 1.046248715 |
| SLC4A2 | 1.050349791 | 0.971447112 |
| BRD9 | 1.050344806 | 1.007879411 |
| RET | 1.050330775 | 0.940947437 |
| CD300LD | 1.05031907 | 1.064063035 |
| RBM15B | 1.050291556 | 0.995949777 |
| TVP23B | 1.050291295 | 1.044878383 |
| VASP | 1.050289722 | 0.917375456 |
| ST8SIA3 | 1.050257672 | 1.007138222 |
| DLG2 | 1.050213958 | 0.946138871 |
| TGFBR1 | 1.050213958 | 1.103828683 |
| BTG4 | 1.050086904 | 0.985954094 |
| AMPH | 1.050055459 | 1.046591298 |
| ZZEF1 | 1.050055459 | 1.046591298 |
| CPA2 | 1.050013501 | 0.987021415 |
| HEXDC | 1.049995135 | 1.137707574 |
| ATG4D | 1.049984194 | 0.956521739 |
| ZRSR2 | 1.049956064 | 1.07290401 |
| EDEM1 | 1.049893815 | 0.862697448 |
| SLC36A4 | 1.049878177 | 0.929526124 |
| GCFC2 | 1.049856505 | 0.927029805 |
| UGT1A4 | 1.049830955 | 0.970838396 |
| AIF1 | 1.049818309 | 0.992120589 |
| CRYBA2 | 1.049817221 | 0.964028777 |
| LKAAEAR1 | 1.049814774 | 0.907845152 |
| IPP | 1.049771821 | 1.117848578 |
| OR5AP2 | 1.049740829 | 0.981843097 |
| RAVER1 | 1.049729909 | 0.89756766 |
| GCA | 1.049729411 | 1.042480301 |
| NUP155 | 1.049718174 | 0.958204865 |
| PM20D2 | 1.049714006 | 0.930113052 |

| | | |
|---|---|---|
| CRYGN | 1.049681881 | 1.083932854 |
| FAM161A | 1.049663531 | 0.882836588 |
| TTYH2 | 1.049654741 | 1.032202809 |
| CCR3 | 1.049648644 | 0.972096042 |
| MN1 | 1.049643142 | 0.976019185 |
| LHX1 | 1.049625921 | 0.990684488 |
| TMEM225 | 1.049621438 | 1.138112596 |
| OSR1 | 1.049541615 | 1.020251114 |
| SMTNL2 | 1.049541615 | 1.020251114 |
| HYAL3 | 1.049533159 | 0.872823005 |
| SPACA3 | 1.049476177 | 1.012978585 |
| CHAT | 1.049451247 | 0.888618874 |
| IER3IP1 | 1.049426652 | 0.96882494 |
| TOMM7 | 1.049404813 | 1.131551901 |
| SEPT1 | 1.049402141 | 0.957472661 |
| FZD10 | 1.049382658 | 0.951979234 |
| TOX3 | 1.049359606 | 1.026326448 |
| ZNF428 | 1.0493295 | 1.081409478 |
| CD86 | 1.049316964 | 1.053861129 |
| BHLHB9 | 1.049285589 | 1.055893074 |
| APP | 1.049281154 | 0.824789098 |
| VIPR1 | 1.049272345 | 0.957170668 |
| RPL13A | 1.049257013 | 0.915724563 |
| OR51D1 | 1.049200331 | 1.096265844 |
| WDR5B | 1.049153481 | 0.942446043 |
| KIF2B | 1.049145453 | 0.965573107 |
| SCUBE1 | 1.049141383 | 0.979445015 |
| FBLN7 | 1.049099562 | 1.025351148 |
| TRIML2 | 1.049006406 | 0.912983899 |
| DDB2 | 1.048953103 | 0.985419198 |
| RBX1 | 1.048948403 | 0.972744971 |
| ZNF688 | 1.048918515 | 0.808424463 |
| NUDT10 | 1.048866491 | 1.04412719 |
| ZNF519 | 1.048849544 | 0.883521754 |
| CLN6 | 1.048848864 | 0.920520726 |
| YAF2 | 1.0488427 | 0.977936405 |
| DERL1 | 1.048776686 | 1.016786571 |
| SLC25A29 | 1.048768394 | 1.087889834 |
| ATXN10 | 1.048701713 | 0.910895099 |
| TMEM38A | 1.048670934 | 0.973963686 |
| CXCL5 | 1.048663412 | 1.015416238 |
| KIAA0408 | 1.048637427 | 0.926690968 |
| CSN2 | 1.048599833 | 1.080194411 |
| SLCO3A1 | 1.048592849 | 0.922128488 |

| | | |
|---|---|---|
| KRTAP5-11 | 1.048572295 | 1.046591298 |
| VCPKMT | 1.04850066 | 0.907502569 |
| SLC25A17 | 1.048433693 | 0.971223022 |
| NPAS1 | 1.048430919 | 0.998702141 |
| SKIDA1 | 1.04841788 | 0.934224049 |
| SNRPD1 | 1.048415327 | 1.031186715 |
| CCDC63 | 1.04839541 | 0.976103686 |
| DEFB127 | 1.048395315 | 0.984583762 |
| CDC42EP5 | 1.048382765 | 0.989037342 |
| GNMT | 1.048288582 | 0.902011681 |
| UMODL1 | 1.048255477 | 1.064803564 |
| LTN1 | 1.04823145 | 1.00908501 |
| PTGR1 | 1.048227511 | 1.121911705 |
| RARS2 | 1.048219055 | 0.883758607 |
| NSDHL | 1.048201379 | 0.856217092 |
| CPNE8 | 1.048183221 | 0.972935937 |
| BARD1 | 1.048162216 | 0.942894225 |
| C9orf169 | 1.048157576 | 0.935936965 |
| WDYHV1 | 1.048131998 | 0.858242203 |
| ARHGAP33 | 1.048098266 | 0.958204865 |
| RBFOX3 | 1.048040764 | 0.917375456 |
| FLJ33360 | 1.047977017 | 1.057553957 |
| LAMA5 | 1.047971791 | 0.990279465 |
| UBE2A | 1.047939779 | 1.024659312 |
| DZIP1L | 1.047865479 | 1.008100446 |
| ECH1 | 1.047843116 | 0.852571891 |
| STRADB | 1.047830618 | 1.208306295 |
| ZNF672 | 1.047807153 | 1.010125557 |
| C17orf80 | 1.047757129 | 1.140137708 |
| PSMD4 | 1.047755272 | 1 |
| CTNND2 | 1.047708512 | 1.0222679 |
| C2orf49 | 1.047674759 | 0.943296881 |
| CCDC19 | 1.047592338 | 1.03199676 |
| SLC27A2 | 1.047585767 | 0.974888619 |
| CDK5RAP3 | 1.047547799 | 0.983898595 |
| CDH8 | 1.047538964 | 0.988352175 |
| SAG | 1.04753031 | 0.992805755 |
| PLEKHG5 | 1.047513177 | 1.063588497 |
| RAP2B | 1.047484701 | 0.992709599 |
| CEP170 | 1.04745592 | 0.878493317 |
| SNCA | 1.047454099 | 0.969167523 |
| COASY | 1.04745259 | 0.979234263 |
| LDLRAD2 | 1.047388985 | 0.923855812 |
| FAM101A | 1.047388017 | 0.967193196 |

| | | |
|---|---|---|
| TMEM8A | 1.047366289 | 0.881808839 |
| MAP3K7CL | 1.047348487 | 0.982989064 |
| ZNF74 | 1.047331491 | 0.899623159 |
| TMEM120A | 1.047320164 | 1.044878383 |
| IFRD1 | 1.047234755 | 0.953066119 |
| FAM155A | 1.047193297 | 0.902706406 |
| CYSTM1 | 1.047178755 | 0.916160389 |
| C14orf105 | 1.047170189 | 0.916067146 |
| INSR | 1.047135975 | 1.107073329 |
| TRIM13 | 1.047130953 | 1.255029202 |
| OR10Z1 | 1.047114911 | 1.028776978 |
| DYNLRB2 | 1.047093661 | 0.887735237 |
| MCEMP1 | 1.047017937 | 0.93600648 |
| UBE2G1 | 1.046988699 | 0.892926671 |
| FAM104B | 1.046975198 | 0.981368975 |
| CWC27 | 1.046890841 | 0.808019441 |
| OTX1 | 1.046890841 | 0.808019441 |
| USP1 | 1.046853277 | 0.974691759 |
| PHF14 | 1.046849161 | 0.862426995 |
| RABL3 | 1.046838025 | 1.066118534 |
| ZNF689 | 1.046832361 | 0.971648441 |
| WDR18 | 1.046832316 | 0.876327509 |
| RHOBTB2 | 1.046826193 | 1.051265412 |
| OBP2A | 1.046816927 | 1.119481571 |
| TRAFD1 | 1.046797009 | 1.046577562 |
| YTHDF3 | 1.046784423 | 0.911956149 |
| RFX3 | 1.046767908 | 0.791044776 |
| BOC | 1.046732136 | 0.848235697 |
| TSSK4 | 1.046721477 | 0.944192083 |
| SIGLEC11 | 1.046634902 | 0.928400137 |
| TMEM155 | 1.046578068 | 1.025111381 |
| C14orf93 | 1.046520857 | 1.044193217 |
| AARS2 | 1.046505122 | 0.804374241 |
| TMEM215 | 1.046466305 | 0.971565605 |
| COG5 | 1.046387511 | 0.916752312 |
| OXER1 | 1.046374897 | 0.847501622 |
| RMDN3 | 1.046349856 | 1.0036452 |
| MLANA | 1.046334454 | 0.983556012 |
| REG3A | 1.046301711 | 1.005840363 |
| IDI2 | 1.046265601 | 1.066828676 |
| CADM3 | 1.046232897 | 1.053057918 |
| PLEKHG1 | 1.046232897 | 1.053057918 |
| TXLNA | 1.046224696 | 0.978128797 |
| C1QTNF6 | 1.04621354 | 1.068853787 |

| | | |
|---|---|---|
| DNAJC1 | 1.046192635 | 1.01781432 |
| OR9Q1 | 1.046158743 | 1.08016444 |
| SCAMP2 | 1.046122329 | 1.133679429 |
| SHC2 | 1.046121045 | 0.927372388 |
| TMC6 | 1.046110845 | 1.027541515 |
| PCSK7 | 1.046062068 | 0.863075925 |
| CERS5 | 1.046051652 | 0.893479141 |
| STARD3NL | 1.045987382 | 0.939362795 |
| MCTP2 | 1.045971625 | 0.911300122 |
| EXD3 | 1.045966902 | 0.992805755 |
| NDC1 | 1.045962054 | 1.006166495 |
| RIMS3 | 1.045929205 | 1.108598835 |
| NAT8 | 1.045877641 | 1.112596193 |
| HTT | 1.045859718 | 0.873458793 |
| ORC5 | 1.045851317 | 0.973621103 |
| KLHDC3 | 1.045849097 | 0.982528263 |
| SLC12A7 | 1.045760988 | 0.887808829 |
| ZNF202 | 1.045746924 | 0.847550531 |
| TRMT2A | 1.045726625 | 0.990065091 |
| FAM118A | 1.045684883 | 0.767517213 |
| C16orf82 | 1.045606057 | 1.006509078 |
| NXNL2 | 1.045606057 | 1.006509078 |
| CASC4 | 1.045603481 | 1.016200891 |
| FGF21 | 1.045535447 | 1.052563271 |
| XAF1 | 1.045517188 | 0.895909275 |
| SCML4 | 1.04549822 | 1.107571086 |
| EGFL6 | 1.045488795 | 1.018499486 |
| SYAP1 | 1.045488297 | 1.014045906 |
| ARL6IP4 | 1.04548508 | 0.987324426 |
| PAF1 | 1.045480002 | 1.051842851 |
| BANF2 | 1.045478782 | 0.938335046 |
| STARD7 | 1.04547049 | 0.880438506 |
| ZNF449 | 1.045460746 | 0.990684488 |
| DNAH3 | 1.04537434 | 0.899415964 |
| MFSD2A | 1.045364571 | 1.087484812 |
| STAG1 | 1.045350012 | 0.932511134 |
| PBX2 | 1.045330187 | 0.917780478 |
| BACE1 | 1.045280574 | 0.904607398 |
| CIR1 | 1.045249599 | 0.982989064 |
| LRRC1 | 1.045243897 | 0.989037342 |
| GABRA1 | 1.045195946 | 0.950681376 |
| WDHD1 | 1.045193739 | 0.868790682 |
| FBXL3 | 1.045147046 | 1.119402985 |
| EIF4EBP3 | 1.045141097 | 0.987039287 |

| | | |
|---|---|---|
| HLA-DRA | 1.045128158 | 1.050222762 |
| UQCC1 | 1.045106745 | 0.956491949 |
| SH120 | 1.045079867 | 0.690968003 |
| SPRED2 | 1.04502857 | 1.038477116 |
| SUMO2 | 1.045008831 | 0.999657417 |
| SNAPC5 | 1.044998002 | 0.981843097 |
| SLC44A1 | 1.044953339 | 1.007138222 |
| PROZ | 1.044909296 | 1.094094744 |
| TRAM2 | 1.044897868 | 1.011647825 |
| OR4K13 | 1.044857905 | 1.117163412 |
| PMCH | 1.044820343 | 1.063720452 |
| LOXL1 | 1.044813472 | 1.058403634 |
| ZNF257 | 1.044806931 | 1.045906132 |
| PPP1R1B | 1.044790915 | 1.01752109 |
| SLITRK6 | 1.044778749 | 1.010277492 |
| CTTNBP2 | 1.044752424 | 0.979102432 |
| ZNF484 | 1.044705303 | 0.955852572 |
| SMAD4 | 1.044675402 | 1.052652896 |
| CHML | 1.044601078 | 0.987021415 |
| MARS2 | 1.044584728 | 0.928311057 |
| CTAGE6 | 1.044576272 | 1.056703119 |
| CNIH1 | 1.044529954 | 0.998629668 |
| ADAMTS3 | 1.04451966 | 0.992212849 |
| SLC22A16 | 1.044505396 | 0.976361768 |
| ANKRD13B | 1.044487687 | 0.963953017 |
| EFCAB12 | 1.044436186 | 1.078574322 |
| PSEN2 | 1.044425591 | 0.95018226 |
| ALOX5AP | 1.044421414 | 1.046982584 |
| CCDC78 | 1.044397523 | 0.999594978 |
| UBR3 | 1.044362588 | 1.089552239 |
| HDAC7 | 1.044316885 | 0.954232483 |
| CLCN6 | 1.044280429 | 1.007787151 |
| ATG2B | 1.044222981 | 1.122302158 |
| EHD4 | 1.044209044 | 0.958282706 |
| CCDC92 | 1.044191725 | 1.007695423 |
| HKR1 | 1.044190675 | 1.012332991 |
| RGMA | 1.044176086 | 1.057108141 |
| PKD1L3 | 1.044125085 | 1.018170019 |
| TRIM43 | 1.044077356 | 1.015416238 |
| FARSB | 1.04407281 | 0.977287476 |
| IRF6 | 1.04407281 | 0.977287476 |
| OCLM | 1.04405307 | 1.076396026 |
| UBE2K | 1.044048525 | 1.023361454 |
| DSG4 | 1.044043147 | 0.993148338 |

| | | |
|---|---|---|
| C1orf85 | 1.044039566 | 1.013770757 |
| CTSE | 1.04402638 | 1.069435432 |
| FAM20A | 1.043999922 | 1.040767386 |
| TMEM246 | 1.043980578 | 0.950587282 |
| NXF1 | 1.043927039 | 0.952612394 |
| NPNT | 1.043907529 | 0.986229243 |
| FAR1 | 1.043875502 | 1.034943474 |
| FSHR | 1.043868072 | 0.900713822 |
| C2orf50 | 1.043856131 | 0.938841636 |
| POPDC2 | 1.043848543 | 1.087016101 |
| TGFBR2 | 1.043837036 | 0.992861778 |
| CAPZA1 | 1.043821719 | 1.003768414 |
| CPLX3 | 1.043807492 | 0.929427886 |
| NDP | 1.043771013 | 0.951979234 |
| RRP15 | 1.043735861 | 0.844066424 |
| C17orf70 | 1.043710289 | 1.043742406 |
| STRAP | 1.043708627 | 1.006851662 |
| EPHA5 | 1.043691077 | 0.957170668 |
| C9orf156 | 1.043674463 | 0.989037342 |
| PDZK1IP1 | 1.043631892 | 0.980558931 |
| ZG16 | 1.043590731 | 1.066461117 |
| ABHD16A | 1.043560147 | 0.992120589 |
| DNAJC25-GNG10 | 1.043504054 | 0.908872902 |
| TCF4 | 1.043463206 | 1.064892927 |
| FZD1 | 1.043448388 | 0.880597015 |
| TMEM132A | 1.043379942 | 0.92345079 |
| NDUFS7 | 1.043364007 | 0.885788449 |
| SLC25A26 | 1.043355846 | 0.955121617 |
| ZNF835 | 1.043353045 | 0.874038072 |
| IL17RD | 1.043343658 | 1.002740665 |
| LINC00696 | 1.043334687 | 0.946214457 |
| KLK4 | 1.043314531 | 1.029201817 |
| ZDHHC9 | 1.043281674 | 0.976019185 |
| MGC34761 | 1.043260243 | 0.967112025 |
| TEX26 | 1.043257991 | 1.046172539 |
| METTL8 | 1.043234019 | 0.752757949 |
| HCN3 | 1.043081885 | 0.998702141 |
| UBE2B | 1.043032244 | 1.049967554 |
| ZNF761 | 1.042988772 | 1.091940057 |
| RHOJ | 1.042963422 | 1.055158988 |
| L3HYPDH | 1.04295179 | 0.99108951 |
| C16orf45 | 1.042931057 | 1.060348319 |
| CNTRL | 1.0429063 | 0.890238963 |
| HLA-DMB | 1.042834504 | 0.968202466 |

| | | |
|---|---|---|
| TRIM17 | 1.042799517 | 0.922128488 |
| IQCK | 1.042735192 | 1.086269745 |
| HSPA14 | 1.042722462 | 0.927319922 |
| TRIM48 | 1.042670022 | 0.783906554 |
| APOO | 1.04264783 | 0.951802349 |
| PRDM7 | 1.042631618 | 1.124015074 |
| ZNF512 | 1.042614355 | 1.037684138 |
| AGBL4 | 1.042569636 | 0.920210612 |
| VN1R5 | 1.042564154 | 1.086307592 |
| ZNF7 | 1.042521697 | 0.890373416 |
| DDO | 1.042506012 | 1.027136493 |
| SELT | 1.042484543 | 1.109284001 |
| POFUT2 | 1.042403801 | 0.932168551 |
| IL3RA | 1.04239593 | 0.857027136 |
| PFN2 | 1.042363883 | 0.957519699 |
| DBP | 1.042349867 | 0.953277093 |
| ZNF37A | 1.04233339 | 1.021582734 |
| HLA-DRB1 | 1.042322371 | 1.055807917 |
| GTF2F2 | 1.042277834 | 0.958468527 |
| DNAI2 | 1.042255865 | 1.033573141 |
| FBXO16 | 1.042244493 | 0.758598313 |
| NOSIP | 1.042230356 | 0.990407674 |
| TRMT44 | 1.042204395 | 0.952207371 |
| SCN8A | 1.04219938 | 1.014925373 |
| TMEM42 | 1.042102566 | 0.974078574 |
| TMEM256 | 1.042049232 | 1.029566626 |
| ADAR | 1.042006497 | 1.1330305 |
| GCDH | 1.041993325 | 0.927968851 |
| OR4Q3 | 1.041944407 | 0.880438506 |
| FBXO9 | 1.041928784 | 0.984425698 |
| AAR2 | 1.041894521 | 1.029462145 |
| CT83 | 1.041864523 | 1.002025111 |
| ADIPOR1 | 1.041853153 | 0.986296677 |
| OR4F3 | 1.041843548 | 0.952038369 |
| TMEM173 | 1.041835487 | 1.146618064 |
| MOG | 1.041818945 | 0.977389517 |
| TEX35 | 1.04181292 | 0.968408262 |
| TLR6 | 1.041808025 | 0.943131209 |
| SHOC2 | 1.041804413 | 0.914945322 |
| NFKBIZ | 1.041801636 | 0.972935937 |
| USP8 | 1.041794268 | 0.994808566 |
| CLECL1 | 1.041778664 | 0.998287085 |
| FAM210A | 1.041726694 | 0.984926345 |
| TUBGCP5 | 1.041726694 | 0.984926345 |

| | | |
|---|---|---|
| PSMD9 | 1.041716061 | 1.15898767 |
| SNRNP35 | 1.041683696 | 0.852346694 |
| HCP5 | 1.041669151 | 1.03117506 |
| TAGLN2 | 1.041658866 | 0.907502569 |
| FANCD2 | 1.041619718 | 0.898595409 |
| HNRNPCL1 | 1.041618495 | 0.958204865 |
| WDR5 | 1.041566389 | 1.064063035 |
| SCGB2B2 | 1.041541152 | 0.910585817 |
| ECHDC3 | 1.041520574 | 1.071688943 |
| MR1 | 1.041500464 | 1.076053443 |
| HES2 | 1.041491402 | 0.98663926 |
| ARMC2 | 1.041484379 | 0.927029805 |
| CTTNBP2NL | 1.04147213 | 0.964763062 |
| EYA4 | 1.041469996 | 1.020765737 |
| LGSN | 1.041462961 | 0.946942082 |
| COG6 | 1.041453885 | 0.9777321 |
| CDC7 | 1.041403397 | 0.918234912 |
| TSTD2 | 1.041402459 | 0.994176088 |
| OR2L2 | 1.041392511 | 1.077764277 |
| SLC5A10 | 1.041364129 | 0.8669695 |
| CDKN2AIPNL | 1.041341517 | 1.035971223 |
| ARID2 | 1.041312927 | 1.056868791 |
| MMP7 | 1.041297776 | 1.144062297 |
| RBAKDN | 1.041266938 | 1.018156903 |
| SPRYD4 | 1.041262155 | 1.010530579 |
| ASB5 | 1.041261719 | 1.092796885 |
| YIPF7 | 1.041230063 | 0.901579587 |
| SUZ12 | 1.041203928 | 1.097988319 |
| RUVBL2 | 1.04117849 | 0.990266061 |
| CEP162 | 1.041127979 | 1.016605913 |
| LACTB | 1.041099745 | 0.927372388 |
| C15orf27 | 1.041058955 | 0.834345889 |
| LAYN | 1.041055083 | 1.038477116 |
| PPARGC1B | 1.041042657 | 1.057105775 |
| SLTM | 1.04100506 | 0.836371 |
| KLRG2 | 1.040999711 | 0.929526124 |
| TM2D1 | 1.040985346 | 1.060348319 |
| ANO5 | 1.040973649 | 1.082562521 |
| CSAD | 1.040968638 | 1.042527339 |
| ANKRD16 | 1.040944243 | 0.87084618 |
| GOLGA8B | 1.040944243 | 0.87084618 |
| PROC | 1.040929133 | 1.011031798 |
| CTNNBIP1 | 1.040896124 | 1.039397054 |
| IDI1 | 1.040896048 | 0.971243418 |

| | | |
|---|---|---|
| ME3 | 1.040868378 | 0.959766385 |
| SLC5A4 | 1.040868378 | 0.903309539 |
| GOSR2 | 1.040814292 | 1.134328358 |
| FSIP2 | 1.040807363 | 0.945529291 |
| PAQR5 | 1.040762534 | 0.901984609 |
| C1orf146 | 1.040760496 | 0.939651681 |
| RRAGA | 1.040748763 | 1.0266061 |
| SLC43A3 | 1.040738583 | 0.932168551 |
| SLC35G3 | 1.040737369 | 0.921830701 |
| TRAPPC6A | 1.040713275 | 1.054678007 |
| FGF4 | 1.040602639 | 0.924075276 |
| ZNF322B_pseudogene | 1.040588653 | 1.023295649 |
| ATP5L | 1.040579138 | 0.926344639 |
| ESAM | 1.040569079 | 1.090441932 |
| SETD4 | 1.040558773 | 1.00526529 |
| RAB8A | 1.040533558 | 0.987444309 |
| MAP3K6 | 1.040460055 | 1.052563271 |
| DVL1 | 1.040454673 | 0.950325454 |
| NDUFAF1 | 1.040454673 | 0.950325454 |
| CNOT6 | 1.040432594 | 1.106520859 |
| RGL3 | 1.040420872 | 1.059266872 |
| TTLL13 | 1.040417159 | 0.973673552 |
| MIOS | 1.040381226 | 0.936964714 |
| S100A7 | 1.040373886 | 1.004110997 |
| CSN3 | 1.040354584 | 1.045906132 |
| ZNF598 | 1.040271305 | 1.007194245 |
| CPM | 1.040261409 | 1.011680727 |
| ELTD1 | 1.040249966 | 0.8935756 |
| PLCZ1 | 1.040249966 | 0.8935756 |
| UBA7 | 1.0402391 | 1.073329007 |
| YBX2 | 1.040129245 | 1.025516403 |
| PHRF1 | 1.040091329 | 1.027255029 |
| RTP3 | 1.04008289 | 0.95018226 |
| GPR82 | 1.040078981 | 1.08890331 |
| PELI1 | 1.040071852 | 0.992463172 |
| ITPRIPL2 | 1.040046841 | 0.988009592 |
| HTR2C | 1.040046134 | 0.970798183 |
| SLC17A1 | 1.040014522 | 0.791044776 |
| C8orf37 | 1.040013153 | 1.108950992 |
| CCDC37 | 1.039973521 | 0.779667882 |
| INADL | 1.039947205 | 0.928400137 |
| PSMB9 | 1.039929729 | 0.981181051 |
| C1orf64 | 1.039927064 | 1.015795869 |
| ODF2 | 1.039917663 | 1.007536828 |

| | | |
|---|---|---|
| DPH2 | 1.039903071 | 0.958282706 |
| EIF2AK1 | 1.03987235 | 0.924724205 |
| TRH | 1.039866826 | 0.998629668 |
| NFE2L3 | 1.039853201 | 0.980153908 |
| ESYT2 | 1.039823655 | 0.922640745 |
| NRP1 | 1.039815629 | 0.99156392 |
| EFTUD1 | 1.0398098 | 0.98217902 |
| PRKRIP1 | 1.039784498 | 0.943473792 |
| CLDN1 | 1.039773734 | 0.944511948 |
| CACNB3 | 1.03976607 | 1.058403634 |
| PREP | 1.03976607 | 1.058403634 |
| MRPS34 | 1.039762614 | 0.980815348 |
| ZNF69 | 1.039736017 | 0.976361768 |
| GPR124 | 1.039634435 | 0.945489942 |
| AGGF1 | 1.039595443 | 0.893074119 |
| RASGRF2 | 1.039592476 | 0.9331963 |
| ARL13B | 1.039555506 | 1.053867963 |
| PGAP1 | 1.039510934 | 0.9963548 |
| FLRT1 | 1.039510285 | 0.998972251 |
| ITIH4 | 1.039502129 | 0.978074683 |
| NRK | 1.039495062 | 0.89422453 |
| OR52B2 | 1.039481468 | 1.052757794 |
| CHCHD6 | 1.039436204 | 0.948269955 |
| CLK3 | 1.039434719 | 1.027903958 |
| CCNL2 | 1.039347986 | 0.934909215 |
| COA6 | 1.039343901 | 1.004455245 |
| ACTR10 | 1.039327415 | 0.855772525 |
| SPECC1L | 1.039322353 | 0.925070879 |
| LCE1E | 1.039284779 | 0.963343611 |
| SNTB2 | 1.039169058 | 1.020212402 |
| IMPACT | 1.039138157 | 0.978417266 |
| KCTD12 | 1.039135557 | 0.992212849 |
| MARVELD3 | 1.039135303 | 0.941075711 |
| GSC | 1.039125017 | 0.994861254 |
| SLA | 1.039100006 | 1.027749229 |
| HPGD | 1.03904748 | 0.998784933 |
| PRKDC | 1.038979554 | 1.007787151 |
| MFSD6L | 1.038936076 | 0.984583762 |
| TCL1B | 1.038936076 | 0.984583762 |
| LIPM | 1.038928596 | 1.012978585 |
| ROM1 | 1.038925937 | 0.943296881 |
| SDCBP | 1.038921434 | 0.946138871 |
| SPESP1 | 1.03891152 | 1.046577562 |
| CYB561D2 | 1.03888279 | 0.945321993 |

| | | |
|---|---|---|
| CHIC2 | 1.038862358 | 1.06235012 |
| CASP8AP2 | 1.038855164 | 0.884490591 |
| TPD52L2 | 1.038839476 | 1.111682083 |
| HDC | 1.038828244 | 1.023361454 |
| NOL4L | 1.038819223 | 1.090784515 |
| COPS7A | 1.038789364 | 0.945871874 |
| KLK12 | 1.038761398 | 0.961713173 |
| PCDHA2 | 1.038739387 | 1.147653306 |
| ZNF271 | 1.038730224 | 0.97326853 |
| SCO1 | 1.038718047 | 0.953408702 |
| CCL5 | 1.038711658 | 0.969852689 |
| TIMM10B | 1.038679851 | 0.965399109 |
| GRSF1 | 1.038663584 | 1.014731072 |
| CCDC150 | 1.038633 | 1.010277492 |
| ANGPTL6 | 1.038615375 | 0.956491949 |
| AKAP12 | 1.038606404 | 0.989379925 |
| IRAK2 | 1.038606383 | 0.977287476 |
| ZNF33B | 1.038605778 | 1.021061158 |
| ANAPC1 | 1.038542789 | 0.980472765 |
| IFT22 | 1.038539481 | 0.996916752 |
| ROR1 | 1.038539207 | 1.054510058 |
| SUGT1 | 1.038507855 | 0.992463172 |
| GABRP | 1.038505949 | 0.987670344 |
| TTC1 | 1.038485155 | 0.922233642 |
| HMHB1 | 1.038475065 | 0.901984609 |
| GATA1 | 1.03845639 | 0.992861778 |
| ACCS | 1.038446959 | 0.950668037 |
| SIL1 | 1.038445192 | 0.967112025 |
| NABP1 | 1.038409913 | 1.136492507 |
| RPUSD4 | 1.038409851 | 0.99554642 |
| OR10V1 | 1.038408809 | 1.011990408 |
| SPATA20 | 1.038354133 | 0.949777238 |
| VAX1 | 1.038311937 | 0.98218568 |
| C7orf43 | 1.03830467 | 0.78290806 |
| SHROOM1 | 1.038258518 | 1.039287161 |
| GEMIN7 | 1.038215744 | 1.001712915 |
| SLC5A8 | 1.038126756 | 1.037684138 |
| TMEM257 | 1.038126756 | 1.037684138 |
| EIF2B2 | 1.038106156 | 0.876468206 |
| TMEM59L | 1.03807926 | 1.027541515 |
| AOX1 | 1.038078362 | 1.034393251 |
| FOXK2 | 1.038074163 | 0.813108371 |
| CXCR6 | 1.038033249 | 1.039584685 |
| OR8B12 | 1.03798609 | 0.96800324 |

| | | |
|---|---|---|
| NT5DC2 | 1.037979061 | 0.989874443 |
| KPNB1 | 1.037964673 | 0.972744971 |
| GTF2IRD2B | 1.037962736 | 1.015416238 |
| OLFM1 | 1.037832833 | 1.136690647 |
| LIPF | 1.037794758 | 0.839065542 |
| DEFB107A | 1.03774657 | 1.002025111 |
| AHRR | 1.037745204 | 1.078451524 |
| MRPS35 | 1.03767357 | 0.963343611 |
| GIP | 1.037671304 | 1.008221994 |
| CSN1S1 | 1.037612628 | 1.015758822 |
| HAUS3 | 1.037612628 | 1.015758822 |
| TRIM31 | 1.037512472 | 1.024659312 |
| LRRTM1 | 1.037509549 | 0.929085303 |
| ESPN | 1.037490066 | 0.794450154 |
| HS6ST3 | 1.037481354 | 0.870214147 |
| SPINK5 | 1.037469746 | 1.029850746 |
| SPRR1A | 1.037455359 | 0.981500514 |
| FGD2 | 1.037440135 | 0.965056526 |
| RPL37 | 1.037431163 | 0.920178143 |
| NHLH1 | 1.037391382 | 1.017471737 |
| GIGYF1 | 1.037377559 | 1.044147428 |
| GPSM1 | 1.037322103 | 0.980130182 |
| RNF139 | 1.03731679 | 0.968202466 |
| FXYD2 | 1.037272664 | 0.9733939 |
| NPAP1 | 1.037226047 | 1.011647825 |
| CXorf38 | 1.037217912 | 1.098015391 |
| CCDC9 | 1.037187941 | 0.950325454 |
| GTF2H1 | 1.037143005 | 1.153796236 |
| PPP3CB | 1.037143005 | 0.824140169 |
| TRADD | 1.037062978 | 1.034258308 |
| HTR7 | 1.03701738 | 1.004542505 |
| CD40LG | 1.037012318 | 0.929121102 |
| CYP4A22 | 1.037004066 | 0.974888619 |
| KLRC3 | 1.036998774 | 0.984926345 |
| TMPRSS5 | 1.036961565 | 0.927319922 |
| ZNF620 | 1.036921249 | 1.044878383 |
| PDCD5 | 1.036913229 | 1.016443988 |
| CD1C | 1.036893984 | 0.935196436 |
| KRTAP13-3 | 1.036860081 | 0.914696814 |
| C10orf118 | 1.036850657 | 0.99554642 |
| CSRNP2 | 1.036835504 | 1.052652896 |
| CLPSL1 | 1.036820571 | 0.869818431 |
| ANO3 | 1.036802085 | 0.922233642 |
| CD34 | 1.036800814 | 1.054678007 |

| | | |
|---|---|---|
| HRCT1 | 1.036773993 | 0.905789654 |
| OR10W1 | 1.03674495 | 0.929770469 |
| PPP2R2D | 1.036733036 | 1.128487995 |
| SLC12A6 | 1.036733036 | 1.128487995 |
| LZIC | 1.036732082 | 1.022610483 |
| EFHD1 | 1.036717401 | 0.953751285 |
| TBC1D7 | 1.036693297 | 1.018156903 |
| GFM1 | 1.036684823 | 0.969218307 |
| C15orf57 | 1.036669083 | 0.877683273 |
| NPTN | 1.036662814 | 1.051265412 |
| TIAL1 | 1.036662814 | 0.963659961 |
| CASQ1 | 1.03664178 | 0.892428914 |
| HPN | 1.036621982 | 0.968851395 |
| HILPDA | 1.036571867 | 0.911956149 |
| KIAA1919 | 1.036551478 | 0.883521754 |
| GTF2IRD1 | 1.036541634 | 0.979234263 |
| CCL11 | 1.036525696 | 0.95991778 |
| WFDC11 | 1.036514887 | 1.025111381 |
| RIMS4 | 1.03649398 | 1.100719424 |
| OR13D1 | 1.036455104 | 1.043850634 |
| INSC | 1.036418559 | 1.007290401 |
| YIPF3 | 1.036410511 | 1.031186715 |
| FCHSD2 | 1.036393664 | 1.010962658 |
| EGR3 | 1.036359206 | 0.912394549 |
| HIST1H1B | 1.036327467 | 0.913669065 |
| PPP1R3D | 1.036327017 | 0.824789098 |
| ASGR2 | 1.036261103 | 0.921830701 |
| RDH11 | 1.036236462 | 1.020765737 |
| TGDS | 1.036219026 | 0.947752126 |
| HOXD1 | 1.036179758 | 1.000685166 |
| BCL2L12 | 1.036089727 | 1.003768414 |
| RCAN1 | 1.036078978 | 0.948734588 |
| DHCR24 | 1.036057985 | 1.046722907 |
| ZNF324 | 1.03599204 | 1.030832477 |
| UHRF1 | 1.035946684 | 1.160285529 |
| SNRPA | 1.035905272 | 1.033915725 |
| ARHGAP5 | 1.03586489 | 0.846496557 |
| RGPD2 | 1.035857833 | 0.845152449 |
| DCTN2 | 1.03584134 | 0.917094896 |
| UBQLN4 | 1.035791486 | 0.874443094 |
| PTPN13 | 1.035758082 | 1.093445814 |
| ZBTB7A | 1.035758082 | 1.093445814 |
| LY6G6F | 1.035748184 | 0.88420692 |
| CELSR2 | 1.035736925 | 0.897469176 |

| | | |
|---|---|---|
| GDF5 | 1.035698397 | 0.90266061 |
| AMY2B | 1.035679381 | 0.804672291 |
| GPAM | 1.03563295 | 1.114211551 |
| PTDSS2 | 1.035581154 | 0.713244228 |
| TENM1 | 1.035541119 | 0.830421377 |
| LRIG1 | 1.035482624 | 1.073312778 |
| ACSL4 | 1.035479911 | 1.036988968 |
| ZSCAN23 | 1.035472837 | 1.01336074 |
| SAMD15 | 1.035442752 | 1.080849606 |
| CCPG1 | 1.035401589 | 1.044147428 |
| POMC | 1.035362554 | 0.932511134 |
| MKL2 | 1.035338933 | 1.098015391 |
| LEPROTL1 | 1.035268718 | 0.935594382 |
| KCNB2 | 1.035268588 | 0.96495782 |
| SQSTM1 | 1.035263779 | 1.039054471 |
| DUSP18 | 1.035231954 | 1.078520441 |
| ASH2L | 1.035228981 | 1.058581706 |
| NRM | 1.035174338 | 1.030147311 |
| RAB39A | 1.035118969 | 0.922233642 |
| SLC12A5 | 1.035071854 | 1.014580802 |
| HYAL2 | 1.035011575 | 1.006489293 |
| IL17D | 1.03499899 | 0.956834532 |
| FCHSD1 | 1.034982905 | 1.000342583 |
| RBMY1D | 1.034982316 | 0.932853717 |
| WDR34 | 1.034960205 | 1.031860226 |
| COL6A1 | 1.034952139 | 1.019869818 |
| CHRNB1 | 1.034947527 | 1.130434783 |
| TAC4 | 1.034922507 | 1.039397054 |
| GALNS | 1.034920525 | 1.022063595 |
| RAI2 | 1.034871812 | 1.078574322 |
| TMSL3_pseudogene | 1.034857805 | 1.066461117 |
| CRTAP | 1.034839137 | 0.986981843 |
| A26B1 | 1.034827946 | 0.919492977 |
| FAM43B | 1.034800495 | 0.825435399 |
| C19orf12 | 1.034763923 | 0.903048989 |
| TAS2R4 | 1.03473189 | 1.065090785 |
| SH3D19 | 1.034718206 | 0.954093868 |
| CPXCR1 | 1.034657265 | 0.850633779 |
| CREG1 | 1.034624803 | 0.99108951 |
| ZNF396 | 1.034579513 | 0.996231586 |
| EPHX2 | 1.034484953 | 0.981181051 |
| UBE2F | 1.034484953 | 0.981181051 |
| FAM83E | 1.034471269 | 1.027136493 |
| ATP8B3 | 1.034460421 | 0.862426995 |

| | | |
|---|---|---|
| OR4D10 | 1.034438057 | 1.026378897 |
| GPC2 | 1.034409104 | 1.002398082 |
| BEND2 | 1.034394152 | 0.990407674 |
| POLR2H | 1.034384212 | 0.953422438 |
| ZNF787 | 1.034379169 | 1.033211827 |
| GRB2 | 1.03437217 | 1.001946788 |
| ASB16-AS1 | 1.034361034 | 0.981368975 |
| PSTPIP1 | 1.034322155 | 1.036998972 |
| SLC25A52 | 1.034319708 | 1.068516615 |
| CCT8L1_pseudogene | 1.034317488 | 1.100034258 |
| ARHGEF28 | 1.034315857 | 0.898937992 |
| MACROD1 | 1.034306568 | 1.025008565 |
| PIH1D3 | 1.034297105 | 0.985419198 |
| TSGA10IP | 1.034258033 | 1.041312272 |
| OR1B1 | 1.034240734 | 0.882494005 |
| USP9Y | 1.034237489 | 1.027903958 |
| CRYZL1 | 1.034211393 | 1.033095393 |
| SLC35A1 | 1.034211393 | 1.033095393 |
| HIF1AN | 1.034152614 | 0.914341337 |
| MMP12 | 1.034152614 | 0.914341337 |
| JMY | 1.034107403 | 0.943701904 |
| FABP5 | 1.034084697 | 1.059052563 |
| SNTB1 | 1.034083818 | 1.161198866 |
| ARL2BP | 1.034077936 | 0.621445701 |
| VWCE | 1.033992075 | 1.059538275 |
| PLK3 | 1.033987638 | 1.0798183 |
| C10orf11 | 1.03397241 | 1.005823912 |
| CALHM2 | 1.033969374 | 1.056868791 |
| SCAPER | 1.033958636 | 1.076396026 |
| ITGB1BP1 | 1.033954422 | 0.942788626 |
| CLTC | 1.033921845 | 0.899280576 |
| TLR8 | 1.033904163 | 0.961064244 |
| LANCL3 | 1.033872111 | 0.98582422 |
| ATP6V0A1 | 1.033855861 | 1.041717294 |
| LOC401052 | 1.033783741 | 1.070572114 |
| SLC16A4 | 1.033744455 | 1.021871203 |
| VANGL1 | 1.033742419 | 1.117163412 |
| HLA-L | 1.033741059 | 1.046591298 |
| BRINP2 | 1.033692139 | 0.970028352 |
| DIS3 | 1.03366672 | 0.916160389 |
| SBF1 | 1.033654888 | 0.868267359 |
| C3orf17 | 1.03359627 | 1.087889834 |
| PLEKHB1 | 1.03358755 | 1.033230558 |
| SPRED1 | 1.033579938 | 1.072285029 |

| | | |
|---|---|---|
| NUDT11 | 1.033539553 | 1.038935756 |
| RNF11 | 1.033533955 | 1.188838417 |
| RNF19B | 1.033451385 | 1.059701493 |
| CD300E | 1.033382423 | 0.962332928 |
| C1orf74 | 1.033363677 | 1.020251114 |
| TOR2A | 1.033347908 | 0.944844125 |
| RPS6KA5 | 1.03331728 | 0.780661908 |
| UGT1A6 | 1.033313908 | 0.938436614 |
| BIRC8 | 1.033305096 | 1.096041531 |
| MTCH1 | 1.033299517 | 1.057553957 |
| TMED5 | 1.033291679 | 0.95991778 |
| BBS1 | 1.033187965 | 1.009592326 |
| HTN1 | 1.033187965 | 1.009592326 |
| BLOC1S6 | 1.033177861 | 1.095238095 |
| ZGRF1 | 1.033148093 | 0.833847208 |
| PGRMC1 | 1.033140121 | 0.977287476 |
| LRFN3 | 1.033139801 | 1.149366221 |
| WASF2 | 1.033124054 | 1.008505468 |
| JMJD1C | 1.033117871 | 1.051730045 |
| ELOVL5 | 1.0330859 | 0.866747671 |
| PTHLH | 1.033065221 | 0.961630695 |
| HCFC1 | 1.033025774 | 0.837767683 |
| COLGALT1 | 1.03301708 | 0.812879708 |
| CEP85 | 1.032994157 | 0.988659376 |
| C1QTNF8 | 1.032977448 | 0.960712839 |
| TMEM183B | 1.032965223 | 0.990684488 |
| GBAS | 1.032940711 | 0.999314834 |
| SPRYD7 | 1.032922995 | 0.960260363 |
| CIRBP-AS1 | 1.032909674 | 0.874614594 |
| HMX2 | 1.032884028 | 0.994861254 |
| POLR2J | 1.032860057 | 0.874107722 |
| ANKHD1-EIF4EBP3 | 1.032857567 | 1.04898938 |
| ADAM23 | 1.032837501 | 0.879299156 |
| CD96 | 1.032802698 | 1.091127098 |
| PTPN22 | 1.032800537 | 1.065541856 |
| SH3BGRL | 1.032779771 | 0.912298732 |
| CCDC85A | 1.032765333 | 1.004860267 |
| OTOL1 | 1.032756799 | 0.966426859 |
| NAA20 | 1.032753566 | 0.916970433 |
| KRTAP5-4 | 1.032716881 | 0.907845152 |
| COPS6 | 1.03271081 | 0.981500514 |
| PARD6G | 1.032701748 | 1.008564577 |
| FAM115C | 1.032692505 | 0.91538198 |
| CITED4 | 1.032688414 | 0.950992305 |

| | | |
|---|---|---|
| LRRTM2 | 1.03265897 | 0.953017416 |
| ULBP2 | 1.032644023 | 1.077766358 |
| KIAA0100 | 1.032621912 | 0.937992463 |
| SLC38A5 | 1.032621912 | 0.937992463 |
| HLTF | 1.032576536 | 0.755353666 |
| SLITRK2 | 1.03253963 | 0.813292223 |
| MECP2 | 1.032491167 | 0.967553537 |
| DGKE | 1.032472809 | 0.972744971 |
| NDUFAB1 | 1.032459285 | 0.997164844 |
| OR51Q1 | 1.032430751 | 0.978759849 |
| BCL7B | 1.032419831 | 0.796848236 |
| RSPRY1 | 1.032418722 | 0.988319273 |
| FSTL5 | 1.032415255 | 0.878040425 |
| OR7C1 | 1.03240932 | 1.01336074 |
| CADM2 | 1.032397392 | 1.123126772 |
| TBCA | 1.03239029 | 0.993833505 |
| TXNDC11 | 1.032383488 | 0.998702141 |
| CLNK | 1.032331886 | 1.016443988 |
| DNAJC25 | 1.032308007 | 0.942788626 |
| NKX6-1 | 1.032250042 | 1.040233615 |
| ADAMTSL4 | 1.032214263 | 1.015390846 |
| ATP6V1C1 | 1.032130591 | 0.864373783 |
| TBC1D4 | 1.032109234 | 1.052757794 |
| ATP2B3 | 1.032093875 | 0.874756652 |
| NPEPPS | 1.032093875 | 0.874756652 |
| KIF9 | 1.032055584 | 1.027541515 |
| SPCS2 | 1.032023327 | 0.905447071 |
| ATP6V1A | 1.032005871 | 0.900713822 |
| PHEX | 1.031999038 | 1.128487995 |
| NPEPL1 | 1.031955489 | 0.916288125 |
| KRT6C | 1.031933531 | 1.005670312 |
| SEZ6 | 1.031924144 | 1.012332991 |
| NEDD4L | 1.031893994 | 1.170019468 |
| CADPS | 1.031882233 | 0.965741692 |
| SLC35E1 | 1.031872265 | 1.104145255 |
| ZNF621 | 1.031818709 | 0.961288112 |
| TMEM30A | 1.031817576 | 1.030489894 |
| PIGA | 1.031813976 | 0.963011032 |
| DUOX1 | 1.031799158 | 0.968202466 |
| CLTCL1 | 1.031783504 | 0.803357314 |
| SGMS2 | 1.031769851 | 0.978585334 |
| TMEM129 | 1.031760487 | 0.859862292 |
| SUMO1 | 1.03175631 | 0.923855812 |
| SLC22A24 | 1.031752643 | 0.987849332 |

| | | |
|---|---|---|
| KCNH7 | 1.031727125 | 0.994159637 |
| LRRC66 | 1.031720704 | 0.937307297 |
| VPS33A | 1.031714724 | 1.048646797 |
| NDUFS5 | 1.031713212 | 0.999351071 |
| PRKG1 | 1.031713212 | 0.999351071 |
| PTBP1 | 1.031713212 | 0.999351071 |
| PFN4 | 1.031674625 | 0.95991778 |
| TRMT6 | 1.031674625 | 0.95991778 |
| CPA3 | 1.031645543 | 1.025308241 |
| RBM6 | 1.031639657 | 0.867962738 |
| EDA | 1.031604458 | 1.095990279 |
| C10orf35 | 1.031588627 | 1.005138746 |
| EPS15L1 | 1.031528139 | 0.83967112 |
| SNAP23 | 1.031526236 | 1.000685166 |
| BAZ1B | 1.031439076 | 1.113562622 |
| MED10 | 1.031350368 | 0.822195221 |
| BTN3A1 | 1.031271052 | 0.982870846 |
| OR5M11 | 1.03124296 | 0.906132237 |
| MAGEA2B | 1.031137615 | 0.92428914 |
| SLC25A33 | 1.031122258 | 0.904414743 |
| EML4 | 1.031122074 | 1.038711888 |
| EBF2 | 1.031121392 | 1.038477116 |
| MROH5 | 1.031112287 | 1.046248715 |
| CCL8 | 1.031060861 | 0.870161014 |
| NRARP | 1.031053642 | 1.010530579 |
| SCG5 | 1.031032603 | 0.978533819 |
| LRRTM3 | 1.031007033 | 0.907845152 |
| GABRG3 | 1.03098317 | 1.031148605 |
| EPS8L1 | 1.030968746 | 0.995203837 |
| PCMT1 | 1.030954274 | 1.046722907 |
| COL28A1 | 1.030932098 | 0.952612394 |
| LHB | 1.030929599 | 0.968139774 |
| CSMD2 | 1.030873052 | 0.826652963 |
| CHP1 | 1.0308156 | 0.871873929 |
| DCTPP1 | 1.030801418 | 0.951695786 |
| RAB12 | 1.030797703 | 0.886947585 |
| OTOP1 | 1.030785288 | 0.966769442 |
| MESDC2 | 1.030782071 | 1.039054471 |
| SSU72 | 1.030765982 | 1.034831916 |
| SPINK4 | 1.030739661 | 0.867420349 |
| TEX30 | 1.030719321 | 1.038882139 |
| EMX1 | 1.030702777 | 0.803374432 |
| CT47.8 | 1.030699018 | 0.905104488 |
| PDGFC | 1.03066046 | 1.022610483 |

| | | |
|---|---|---|
| ABCD3 | 1.030568029 | 0.876873228 |
| ST6GALNAC3 | 1.030529487 | 0.896820247 |
| SLC9A3R2 | 1.030529467 | 0.914945322 |
| SMPD4 | 1.03051393 | 1.057108141 |
| PRDX6 | 1.030512446 | 0.907203115 |
| MAGED2 | 1.03050174 | 1.051387461 |
| FAM162A | 1.030476897 | 1.025111381 |
| DEFB106B | 1.030463771 | 0.906817403 |
| NRBP2 | 1.030455807 | 0.943543154 |
| NCK2 | 1.030417856 | 0.969500324 |
| HERC2P3 | 1.030400247 | 0.997259335 |
| INF2 | 1.030390248 | 0.997164844 |
| SLC17A4 | 1.030374966 | 1.000648929 |
| BCR | 1.030368133 | 1.005840363 |
| CLASP2 | 1.030317658 | 1.039287161 |
| QTRT1 | 1.030304668 | 0.861077359 |
| TNFAIP1 | 1.030278648 | 1.078520441 |
| TNKS2 | 1.030278648 | 1.078520441 |
| CHCHD2 | 1.030263276 | 0.988352175 |
| BSN | 1.03025947 | 0.995889003 |
| HDDC2 | 1.030250824 | 0.829080599 |
| SPRY1 | 1.030250824 | 0.901174565 |
| SEC24A | 1.030221585 | 0.795058728 |
| FAM21A | 1.03020392 | 0.961288112 |
| CLVS1 | 1.030112304 | 0.809181226 |
| ATXN7L3B | 1.030067538 | 1.16032888 |
| CLIP3 | 1.03004039 | 0.881328473 |
| C2orf68 | 1.030003561 | 1.031591738 |
| MAGEA12 | 1.029986842 | 0.947927372 |
| RHOXF1 | 1.029985075 | 0.9554642 |
| BNC2 | 1.029984968 | 0.959497772 |
| ARHGAP25 | 1.029981631 | 0.970537855 |
| EIF1AD | 1.029964459 | 1.05344296 |
| SLC39A6 | 1.029918017 | 0.913326482 |
| ANGPTL2 | 1.029901629 | 0.929526124 |
| OR4D5 | 1.029900074 | 1.033915725 |
| IGF1 | 1.029891441 | 0.967598218 |
| MED13L | 1.029835236 | 0.984241178 |
| WWP1 | 1.029834738 | 0.991778006 |
| TRIM49B | 1.029806606 | 0.871512005 |
| GSTM1 | 1.029799342 | 0.975698663 |
| SDS | 1.0297759 | 0.907852044 |
| DOCK3 | 1.029763511 | 0.979787598 |
| NOTCH2 | 1.029721402 | 0.980532122 |

| | | |
|---|---|---|
| GNPAT | 1.02971778 | 0.985723556 |
| HECTD1 | 1.02971424 | 0.99091499 |
| CALCR | 1.029693659 | 1.022063595 |
| GPRASP1 | 1.029690366 | 1.027255029 |
| SWT1 | 1.029688932 | 0.945186708 |
| AGMO | 1.02966328 | 1.025921426 |
| CLEC1A | 1.029650357 | 1.094743673 |
| PDCL2 | 1.029620542 | 1.029971648 |
| AMZ1 | 1.029560126 | 0.805589307 |
| OR13C4 | 1.029537416 | 0.936279548 |
| LINC00518 | 1.029532109 | 1 |
| KPNA5 | 1.029501033 | 1.075025694 |
| ECSCR | 1.029363654 | 1.058581706 |
| SFN | 1.029315852 | 0.945529291 |
| RIMKLA | 1.029310672 | 1.060348319 |
| AP1S2 | 1.029209918 | 1.019527235 |
| ZNF717 | 1.029201581 | 1.00445358 |
| PLIN4 | 1.029160893 | 0.936622131 |
| EPB41L5 | 1.029136202 | 0.998379911 |
| SRSF10 | 1.02912374 | 0.992463172 |
| VWA2 | 1.029115317 | 1.000405022 |
| PRR19 | 1.029087764 | 0.939705379 |
| PCDHA12 | 1.029085634 | 1.040767386 |
| OR10AG1 | 1.029049553 | 0.926285946 |
| OSM | 1.029040959 | 1.073997945 |
| AURKC | 1.029040342 | 1.001946788 |
| CCR5 | 1.029040342 | 1.048669695 |
| CHRM3 | 1.029040342 | 1.022712524 |
| EIF2B4 | 1.029040342 | 1.0798183 |
| GFI1 | 1.029040342 | 1.033095393 |
| GRIA2 | 1.029040342 | 1.027903958 |
| NOX1 | 1.029040342 | 1.001946788 |
| POLR2B | 1.029040342 | 0.851395198 |
| PSENEN | 1.029030583 | 1.058924289 |
| KIAA0101 | 1.029026958 | 0.957862282 |
| LRRC7 | 1.029004929 | 0.930336168 |
| TLR10 | 1.028956434 | 1.056703119 |
| SEMA4C | 1.028930812 | 1.018631025 |
| CRTC2 | 1.028892117 | 0.971565605 |
| SMIM15 | 1.028818851 | 1.045563549 |
| KIAA2022 | 1.028805855 | 0.866342649 |
| PSMD3 | 1.028776079 | 1.000342583 |
| CALCB | 1.028768648 | 1.063720452 |
| ZNF221 | 1.028716678 | 1.010962658 |

| | | |
|---|---|---|
| UBN1 | 1.028699951 | 0.792223572 |
| GPT | 1.028647075 | 0.838396112 |
| ZNF823 | 1.028642869 | 1.133264817 |
| BTN2A2 | 1.028596291 | 1.088043851 |
| HOOK1 | 1.028580139 | 1.012960713 |
| MYPN | 1.028564755 | 0.882836588 |
| CLASP1 | 1.028554379 | 0.87529976 |
| ASCL4 | 1.028531226 | 0.916067146 |
| SPATA6L | 1.028519717 | 0.96437136 |
| HHIPL2 | 1.028498431 | 0.978938842 |
| KLHL3 | 1.02848804 | 1.065208586 |
| CAV2 | 1.02843005 | 0.952380952 |
| FHOD1 | 1.028427965 | 1.056526208 |
| CCR8 | 1.028384473 | 1.018170019 |
| C11orf55_pseudogene | 1.028381625 | 0.81855002 |
| C14orf166 | 1.028345094 | 1.0445358 |
| ZNF16 | 1.028345094 | 1.0445358 |
| TM6SF1 | 1.028315462 | 0.877012676 |
| LAPTM4B | 1.028302911 | 0.966084275 |
| UBR5 | 1.028297292 | 0.898767034 |
| MRO | 1.028291946 | 1.006851662 |
| SECISBP2L | 1.028260229 | 1.032545392 |
| WDR59 | 1.028251767 | 0.846852693 |
| TFPI | 1.0282111 | 0.80532122 |
| ZNRD1 | 1.028130898 | 0.97326853 |
| C4orf46 | 1.028121446 | 1.03117506 |
| PSMG1 | 1.028089095 | 0.927029805 |
| ACAP1 | 1.028070427 | 1.041795135 |
| TAS2R10 | 1.02801882 | 0.930113052 |
| TMEM179B | 1.028017461 | 0.84857828 |
| RASIP1 | 1.027960416 | 0.899965742 |
| CETN2 | 1.027942201 | 1.007194245 |
| CNGA3 | 1.027861638 | 1.1330305 |
| TULP1 | 1.027839657 | 1.112264763 |
| VPS29 | 1.027833977 | 1.107073329 |
| RAB30 | 1.027826479 | 1.051842851 |
| RBP4 | 1.027780796 | 1.060350422 |
| PSMD14 | 1.027779818 | 0.919400567 |
| UBE3B | 1.027768365 | 1.049967554 |
| ART4 | 1.027755605 | 1.039584685 |
| CDC20B | 1.027740391 | 0.969623329 |
| PCDHGB6 | 1.027672972 | 0.978759849 |
| SNF8 | 1.027656034 | 1.072093965 |
| FAAH2 | 1.027636401 | 0.951330305 |

| | | |
|---|---|---|
| CUX1 | 1.027597133 | 0.925373134 |
| SNAPIN | 1.027546014 | 1.018156903 |
| IER3 | 1.027529431 | 1.010620075 |
| CLEC4F | 1.027521683 | 0.944158959 |
| COIL | 1.02747959 | 0.895854745 |
| METTL10 | 1.027466677 | 0.921548476 |
| DAK | 1.027437449 | 0.955852572 |
| RIPPLY2 | 1.027324178 | 1.012332991 |
| STX7 | 1.027309906 | 0.920178143 |
| WDR6 | 1.027298714 | 1.030489894 |
| PTPRT | 1.027265124 | 1.128487995 |
| TMED9 | 1.027251976 | 1.074524099 |
| PRPF4B | 1.027142956 | 1.055807917 |
| ADAM32 | 1.027133489 | 1.050616483 |
| FIBIN | 1.027122234 | 0.988352175 |
| RIMS1 | 1.027122234 | 0.988352175 |
| SLC30A6 | 1.02709976 | 1.006509078 |
| RPL26L1 | 1.027095773 | 1.05789654 |
| GSK3B | 1.027055035 | 1.00908501 |
| TMEM62 | 1.027052953 | 1.000405022 |
| MLF1 | 1.027017478 | 0.924974306 |
| PCBP3 | 1.027017478 | 0.847893114 |
| CHST15 | 1.027013297 | 0.988319273 |
| ZNF497 | 1.026996726 | 0.994518671 |
| VN1R4 | 1.02696983 | 0.967553537 |
| ASXL2 | 1.026954135 | 0.979445015 |
| DMRT1 | 1.02692871 | 0.914135277 |
| C15orf40 | 1.026926061 | 1.066828676 |
| PCNX | 1.026918159 | 1.041452552 |
| SUGP1 | 1.026887756 | 0.956834532 |
| TSPAN14 | 1.02678291 | 0.970537855 |
| LPGAT1 | 1.026778487 | 0.827865533 |
| ARHGAP1 | 1.026775383 | 0.884490591 |
| CLEC16A | 1.026752507 | 0.937307297 |
| OVCA2 | 1.026745274 | 1.091130012 |
| PPL | 1.026735924 | 0.9554642 |
| RRP8 | 1.026712861 | 1.087016101 |
| KLHDC4 | 1.026701262 | 0.835965978 |
| C20orf166-AS1 | 1.026657266 | 0.976704351 |
| XBP1 | 1.026617109 | 0.950587282 |
| SLC30A10 | 1.026596083 | 1.092796885 |
| KDM4A | 1.026574395 | 0.840698869 |
| KIR2DL4 | 1.026568414 | 1.059952038 |
| HNRNPA0 | 1.026533707 | 1.02672148 |

| | | |
|---|---|---|
| DEFB1 | 1.026500676 | 1.081192189 |
| MPV17L | 1.026481964 | 1.021061158 |
| SLC25A1 | 1.026474162 | 1.040882544 |
| C14orf177 | 1.026463677 | 1.077359255 |
| DHRS12 | 1.026353697 | 0.874443094 |
| NUBPL | 1.026222068 | 1.001215067 |
| GNGT1 | 1.026215135 | 0.954436451 |
| ANKIB1 | 1.026157876 | 0.926670993 |
| DEFA3 | 1.02611927 | 1.071688943 |
| UCHL1 | 1.026091853 | 0.905905256 |
| C2CD5 | 1.026069374 | 1.069544365 |
| TMEM95 | 1.026056308 | 0.967193196 |
| MICU2 | 1.026051622 | 0.910895099 |
| CPLX4 | 1.026045406 | 0.872901679 |
| IGDCC3 | 1.026024065 | 1 |
| ABCE1 | 1.025986809 | 0.874756652 |
| COMMD8 | 1.025957958 | 1.093867763 |
| CERK | 1.025950092 | 0.864373783 |
| KRTAP11-1 | 1.025937569 | 0.977389517 |
| BPY2C | 1.025932449 | 1.068174032 |
| UFD1L | 1.025912553 | 1.067488644 |
| MLLT4-AS1 | 1.025912422 | 0.842754368 |
| MYD88 | 1.025814599 | 0.929085303 |
| LOC653192 | 1.025735805 | 1.010382868 |
| DMRTA1 | 1.025730821 | 0.997259335 |
| ATRN | 1.025722303 | 0.942788626 |
| ZNF567 | 1.025705765 | 0.852004111 |
| OR2G6 | 1.025668883 | 0.98218568 |
| CCDC83 | 1.025655511 | 0.963547995 |
| CASC5 | 1.025641146 | 0.965573107 |
| RTKN | 1.025626832 | 0.967598218 |
| CXCR5 | 1.02562154 | 0.781310837 |
| TTLL3 | 1.025574261 | 1.036046983 |
| C11orf30 | 1.025553121 | 1.100445525 |
| CBX6 | 1.025512519 | 0.962658445 |
| HIST1H2AE | 1.025505542 | 0.944501542 |
| PIP | 1.025465941 | 1.067146283 |
| GHRHR | 1.025454836 | 0.744970798 |
| CENPI | 1.025419098 | 1.060348319 |
| PILRA | 1.025410301 | 0.940047962 |
| LCE2C | 1.025406566 | 1.062373431 |
| RAPGEF1 | 1.025401149 | 0.921891059 |
| SERPINB7 | 1.025391543 | 0.903734156 |
| CLSTN1 | 1.02538751 | 1.015758822 |

| | | |
|---|---|---|
| DCAF16 | 1.02534335 | 0.883353584 |
| TARBP2 | 1.025324938 | 0.953751285 |
| LPAR4 | 1.025314639 | 0.896171317 |
| POLR3K | 1.025289284 | 1.068137573 |
| VIP | 1.025204006 | 0.912983899 |
| GDF6 | 1.025198932 | 1.03562864 |
| FTCD | 1.025196304 | 0.988694758 |
| FANK1 | 1.025161137 | 0.778857837 |
| ACPT | 1.025157155 | 1.031797534 |
| SLC4A11 | 1.025133777 | 0.944844125 |
| SPERT | 1.02510444 | 0.851356825 |
| GARNL3 | 1.025075557 | 1.052652896 |
| CDC16 | 1.025051462 | 1.056703119 |
| RLN2 | 1.025049863 | 0.77834875 |
| GPNMB | 1.025047054 | 0.987324426 |
| SYNGR1 | 1.025041798 | 1.016101405 |
| MFAP4 | 1.025036814 | 1.044878383 |
| FAM98B | 1.025031545 | 0.795058728 |
| ANP32A | 1.025028437 | 0.994329688 |
| IFT74 | 1.025015804 | 1.062778453 |
| TRMT11 | 1.025003171 | 0.998379911 |
| IKBKE | 1.024994237 | 0.990266061 |
| NSUN7 | 1.024962062 | 0.938031592 |
| ST6GAL1 | 1.024962062 | 0.938031592 |
| C6orf25 | 1.024953897 | 0.982870846 |
| TSPAN2 | 1.024945558 | 1.074929121 |
| MAEA | 1.024944065 | 0.878383008 |
| CRLF1 | 1.024938855 | 0.935936965 |
| PRR24 | 1.024879001 | 0.967797191 |
| GBP2 | 1.024859571 | 0.885783718 |
| KIRREL3 | 1.024841213 | 0.989037342 |
| CYP2A6 | 1.024840128 | 0.953926022 |
| ARMCX1 | 1.024808046 | 1.067831449 |
| BPIFA3 | 1.024788835 | 0.780061665 |
| UBL7 | 1.024773125 | 1.036857027 |
| CA3 | 1.024729651 | 1.084360805 |
| ATG16L1 | 1.024728029 | 0.840698869 |
| WDR81 | 1.024694983 | 0.980558931 |
| FERMT3 | 1.024657301 | 0.911956149 |
| BBC3 | 1.024622322 | 0.9331963 |
| GRM4 | 1.024580152 | 1.048020766 |
| KLK15 | 1.024580152 | 1.048020766 |
| SEPT2 | 1.024559533 | 1.075334143 |
| ARID3B | 1.024541444 | 0.986296677 |

| | | |
|---|---|---|
| GOLGA3 | 1.024506873 | 1.046933881 |
| XKR9 | 1.024445841 | 0.981843097 |
| NUDT4 | 1.024443413 | 1.016872161 |
| UBE2T | 1.024443413 | 1.016872161 |
| DYDC2 | 1.024438958 | 1.025111381 |
| C7orf63 | 1.024418882 | 0.963686194 |
| BOLA2 | 1.024375435 | 0.962737951 |
| TEFM | 1.024340541 | 0.894289186 |
| CBR3 | 1.024323056 | 0.99091499 |
| METAP1D | 1.024298194 | 0.985723556 |
| STRADA | 1.024298194 | 0.985723556 |
| ZNF780B | 1.024297904 | 1.051437829 |
| NNAT | 1.024294507 | 0.980472765 |
| CHTOP | 1.024283233 | 0.978938842 |
| PRAF2 | 1.024232977 | 0.854744776 |
| PDS5A | 1.024182168 | 0.920615634 |
| SYT12 | 1.024179647 | 1.086673518 |
| CTAG1B | 1.024174301 | 0.997259335 |
| HERPUD2 | 1.024107643 | 1.011340624 |
| ZC3H14 | 1.024065566 | 1.019441069 |
| OPN1MW2 | 1.024061117 | 1.064063035 |
| PMS2 | 1.024055072 | 0.942891859 |
| ETHE1 | 1.024009827 | 0.956491949 |
| C17orf85 | 1.023973059 | 1.037667072 |
| ACAT2 | 1.023971207 | 1.053861129 |
| POLR3G | 1.02394972 | 0.918234912 |
| ZGRF1 | 1.023944685 | 0.963142973 |
| FAM199X | 1.023895732 | 1.134467396 |
| F7 | 1.02389514 | 1.038286827 |
| HSPA12B | 1.023886857 | 1.0445358 |
| SUPT7L | 1.023877648 | 1.138517618 |
| AUNIP | 1.023849223 | 0.981368975 |
| FBXO40 | 1.023843194 | 1.027903958 |
| TSSC1 | 1.023814633 | 0.936964714 |
| BAX | 1.023801538 | 0.892277742 |
| NKAIN1 | 1.023737155 | 1.0036452 |
| CHST2 | 1.023736009 | 1.007138222 |
| SCCPDH | 1.023678217 | 0.931146213 |
| TSLP | 1.023641278 | 0.864337102 |
| SH3YL1 | 1.023616902 | 0.906817403 |
| CDK15 | 1.023545843 | 0.850746269 |
| CT45A4 | 1.023522522 | 0.994861254 |
| ARHGEF12 | 1.023518367 | 1.050222762 |
| COX11 | 1.023494578 | 0.967598218 |

| | | |
|---|---|---|
| MRS2 | 1.023460312 | 1.029804728 |
| TNFRSF6B | 1.023450027 | 1.061664954 |
| SHKBP1 | 1.023444136 | 0.926687222 |
| GFRA3 | 1.023417171 | 0.950032446 |
| SIGLEC12 | 1.023351741 | 0.997974889 |
| ERCC5 | 1.023329826 | 1.169370539 |
| DHDH | 1.023306048 | 1.008100446 |
| KRT36 | 1.02327691 | 1.106925881 |
| OR2A7 | 1.023264031 | 1.042137718 |
| CD97 | 1.023255571 | 1.038935756 |
| C11orf58 | 1.023240386 | 0.744025921 |
| SND1 | 1.023157405 | 1.042527339 |
| ERG | 1.023126323 | 0.903309539 |
| CNNM3 | 1.023119901 | 0.87084618 |
| MNDA | 1.023119901 | 1.062007537 |
| HAPLN3 | 1.02306707 | 0.948269955 |
| AIG1 | 1.023065892 | 0.927029805 |
| THOP1 | 1.023022596 | 0.887735237 |
| HIST1H3I | 1.023021262 | 1.015073655 |
| TCTEX1D2 | 1.023009708 | 0.908872902 |
| SPAG4 | 1.022972373 | 1.018156903 |
| ADAMTS14 | 1.022951386 | 0.987021415 |
| CCT8 | 1.022951386 | 1.096690461 |
| AP2B1 | 1.022936681 | 0.998379911 |
| JUP | 1.022929737 | 1.053100377 |
| SEPW1 | 1.022919633 | 1 |
| TNFRSF25 | 1.022886598 | 0.976638546 |
| AGFG2 | 1.022881708 | 0.840698869 |
| BPIFA1 | 1.022862089 | 0.971223022 |
| SPATC1 | 1.022804727 | 0.953066119 |
| KAT2A | 1.022803699 | 0.856586632 |
| ODF2L | 1.022791043 | 1.034831916 |
| HAGH | 1.022786905 | 0.961064244 |
| SHROOM3 | 1.022767814 | 1.04090725 |
| NOA1 | 1.022753392 | 0.956149366 |
| CGRRF1 | 1.022752905 | 0.95587281 |
| RAB3D | 1.022752905 | 0.95587281 |
| EIF2S2 | 1.022718576 | 0.950681376 |
| GLTPD1 | 1.022667213 | 0.884892086 |
| TSR3 | 1.022638477 | 0.968408262 |
| HIP1R | 1.02261778 | 0.997259335 |
| SMAD6 | 1.022616825 | 1.039584685 |
| TMEM229B | 1.022614586 | 0.974483597 |
| SIAH1 | 1.022512145 | 0.778857837 |

| | | |
|---|---|---|
| CDK18 | 1.022452343 | 1.013627515 |
| FNTB | 1.022452343 | 1.013627515 |
| USP47 | 1.022452343 | 1.013627515 |
| TNRC6B | 1.022376899 | 1.039287161 |
| NIPSNAP1 | 1.022361781 | 0.924631723 |
| NEK7 | 1.022349604 | 0.998053212 |
| MEIS2 | 1.022346393 | 1.09733939 |
| ZDHHC16 | 1.022337178 | 0.959575197 |
| ZBTB5 | 1.022309765 | 1.050702295 |
| NXPE3 | 1.022281998 | 1.06763872 |
| MARC2 | 1.022210885 | 0.968813285 |
| TLR1 | 1.022186689 | 0.853786958 |
| TRMT2B | 1.022185823 | 1.098015391 |
| FAXC | 1.022158024 | 0.936964714 |
| RABEP1 | 1.022119193 | 0.950668037 |
| PENK | 1.022069851 | 0.953751285 |
| RIMKLB | 1.022054652 | 1.015390846 |
| FICD | 1.022035294 | 1.021466181 |
| CGB5 | 1.022020962 | 0.911270983 |
| TMSB15A | 1.021972434 | 1.041717294 |
| SS18L1 | 1.0219723 | 0.95991778 |
| SPTBN5 | 1.02196718 | 1.085988352 |
| PKD2L1 | 1.021943586 | 0.846852693 |
| TEX261 | 1.021937049 | 0.973621103 |
| GNPTAB | 1.021917928 | 1.059943297 |
| AMER3 | 1.021900252 | 1.066018631 |
| MRPL45 | 1.0218699 | 1.081534772 |
| TNFSF11 | 1.021863239 | 1.035971223 |
| KRT78 | 1.021814469 | 1.096395302 |
| EFNB1 | 1.021791605 | 1.01781432 |
| POP1 | 1.021758302 | 1.066461117 |
| GGN | 1.021669314 | 0.862966769 |
| GMDS | 1.021661343 | 0.866342649 |
| TRPV4 | 1.021658418 | 0.904607398 |
| BCL11B | 1.021597408 | 1.019527235 |
| PHACTR4 | 1.021531211 | 0.977046934 |
| RBMS1 | 1.021507242 | 0.931483385 |
| SGSM2 | 1.021472353 | 0.850976362 |
| C18orf56 | 1.021467233 | 0.969510106 |
| ZNF441 | 1.021457311 | 0.993833505 |
| MYL12A | 1.021448928 | 0.958890031 |
| APRT | 1.021448494 | 0.967553537 |
| GOLGA6 | 1.021445254 | 0.937221547 |
| ROBO4 | 1.021439957 | 0.923946557 |

| | | |
|---|---|---|
| FAM213A | 1.02143017 | 0.948269955 |
| MON1A | 1.021395893 | 0.795868773 |
| HAUS7 | 1.021320094 | 0.983799109 |
| S100A7A | 1.021279393 | 1 |
| FAM179B | 1.021258991 | 0.99554642 |
| TMEM63B | 1.021254534 | 1.010125557 |
| KRTAP13-1 | 1.02122664 | 0.925659472 |
| GPR12 | 1.021224904 | 1.025308241 |
| AKR1C4 | 1.021185059 | 1.020116807 |
| GNG10 | 1.021161021 | 0.847501622 |
| TXNDC15 | 1.021144886 | 1.014925373 |
| RAI14 | 1.021128227 | 0.980472765 |
| ZSWIM7 | 1.021081083 | 1.087079789 |
| KCNN2 | 1.021063303 | 1.08825438 |
| SRP19 | 1.021038605 | 0.962315862 |
| LIN7A | 1.021027188 | 1.113406237 |
| TMEFF1 | 1.021020526 | 1.073312778 |
| BHLHE41 | 1.021019665 | 1.04898938 |
| VTCN1 | 1.021013587 | 0.933576347 |
| FAM214A | 1.021008697 | 0.935601458 |
| IGIP | 1.020999638 | 1.062692703 |
| FOXN2 | 1.020989849 | 0.943701904 |
| LRRN2 | 1.020980425 | 0.947752126 |
| COPRS | 1.020971205 | 0.951802349 |
| RNF207 | 1.020882604 | 1.146009085 |
| CAMLG | 1.020877756 | 1.071942446 |
| LRRC63 | 1.020857608 | 1.004455245 |
| RAB1B | 1.020853431 | 1.006480356 |
| CLCA3P | 1.020851733 | 0.978585334 |
| HTATIP2 | 1.020831893 | 0.894873459 |
| POLN | 1.020808019 | 0.9733939 |
| XCR1 | 1.020787767 | 1.051914341 |
| SIGLEC10 | 1.02077155 | 0.980815348 |
| APEX1 | 1.02074677 | 1.046722907 |
| BPIFB2 | 1.02074373 | 1.032545392 |
| BRMS1 | 1.020718583 | 1.084275437 |
| MRFAP1L1 | 1.02071141 | 0.865127582 |
| CCL22 | 1.02066992 | 0.837958205 |
| CYYR1 | 1.020661357 | 1.038711888 |
| DEFB124 | 1.020625698 | 0.803014731 |
| RBFOX1 | 1.020620578 | 1.041795135 |
| PSMA2 | 1.020586652 | 0.868916288 |
| DACH2 | 1.020584104 | 1.145255224 |
| MRPS28 | 1.020575223 | 0.944501542 |

| | | |
|---|---|---|
| SEPT6 | 1.020535858 | 1.020765737 |
| RDH13 | 1.020535858 | 0.942245295 |
| RANBP3L | 1.020515138 | 0.96435804 |
| GPR65 | 1.02049552 | 1.094094744 |
| DSCR4 | 1.020474274 | 1.154504967 |
| IL10 | 1.020469648 | 0.990684488 |
| DNAJB13 | 1.020466235 | 0.992709599 |
| LHX8 | 1.020444097 | 0.991778006 |
| ZFHX2 | 1.020407215 | 1.236724906 |
| SYCP3 | 1.02039616 | 1.08461802 |
| KCNIP4 | 1.020392945 | 0.926670993 |
| CPVL | 1.020359275 | 1 |
| CALY | 1.020288731 | 0.990407674 |
| MCM10 | 1.020288731 | 0.848920863 |
| PLD4 | 1.020287901 | 1.068137573 |
| ARSD | 1.020268143 | 0.989617132 |
| FAM196A | 1.020253978 | 1.083247688 |
| SFXN2 | 1.020143753 | 0.900713822 |
| C10orf131 | 1.020137034 | 1.040767386 |
| CRABP1 | 1.020131252 | 0.93236128 |
| RBM19 | 1.020128335 | 1.054470709 |
| PEX26 | 1.02011157 | 1.081877355 |
| TIMD4 | 1.02004496 | 0.997164844 |
| SLC35E3 | 1.020041748 | 0.943473792 |
| CPNE6 | 1.020031644 | 0.891743748 |
| TMEM5 | 1.02001786 | 1.019441069 |
| ANKS6 | 1.02001302 | 1.023491292 |
| ANAPC13 | 1.020006724 | 0.932853717 |
| RAPGEFL1 | 1.020002737 | 1.011990408 |
| ASAH2 | 1.019994172 | 1.039692183 |
| BAK1 | 1.019989588 | 1.043742406 |
| AADACL2 | 1.01998047 | 1.051842851 |
| MAPRE3 | 1.019976554 | 1.105126541 |
| ZNF14 | 1.019961958 | 1.173347037 |
| SYNE2 | 1.01994252 | 0.977046934 |
| KRT86 | 1.019938851 | 1.090319968 |
| FER | 1.019933746 | 1.0266061 |
| CLIC1 | 1.019890609 | 1.094743673 |
| RAD52 | 1.019840721 | 1.016223232 |
| TBC1D13 | 1.019834819 | 1.051730045 |
| ACCSL | 1.019792772 | 1.068516615 |
| CERS3 | 1.019779723 | 0.989379925 |
| MRPL10 | 1.019767172 | 1.030489894 |
| ZBTB24 | 1.019732238 | 1.019869818 |

| | | |
|---|---|---|
| PDGFB | 1.019728519 | 0.892264075 |
| OR5K3 | 1.019721953 | 1.060979788 |
| MYH11 | 1.019720866 | 0.913326482 |
| GIF | 1.019697561 | 1.000648929 |
| LGALS3BP | 1.019697561 | 1.000648929 |
| COL12A1 | 1.019690508 | 1.036656389 |
| PDRG1 | 1.019688277 | 0.932766302 |
| MTX2 | 1.019665724 | 0.971223022 |
| TUBA3E | 1.019662246 | 0.961117861 |
| GPSM3 | 1.019647736 | 1.067146283 |
| TPPP | 1.019614705 | 1.042822885 |
| CCR7 | 1.019599596 | 0.848799481 |
| HCFC1R1 | 1.019588471 | 0.949982871 |
| LGALS4 | 1.019576463 | 1.058403634 |
| UNC119B | 1.019509632 | 0.887632751 |
| KIAA1279 | 1.019504784 | 0.997259335 |
| CAMK2D | 1.019499574 | 0.979883193 |
| ZNF415 | 1.019465909 | 0.890715999 |
| MFSD6 | 1.019388837 | 1.006509078 |
| OR2T4 | 1.019316659 | 1.04008222 |
| MCOLN2 | 1.019293025 | 0.959117456 |
| VGLL4 | 1.019269582 | 0.988352175 |
| HES6 | 1.01926369 | 0.89671932 |
| OR5R1 | 1.019258073 | 0.933538883 |
| KIT | 1.019240008 | 0.953926022 |
| FNTA | 1.019231853 | 0.963547995 |
| ZNF790 | 1.019216214 | 1 |
| CCDC68 | 1.019214533 | 1.004050223 |
| CDRT15 | 1.019211986 | 1.010125557 |
| ZNF317 | 1.019182492 | 1.089104901 |
| CEP104 | 1.019139226 | 1.069201781 |
| CAPN12 | 1.019132083 | 0.943543154 |
| MAP3K10 | 1.019077338 | 0.93835172 |
| HABP2 | 1.019036752 | 1.001297859 |
| TBX3 | 1.019036752 | 1.001297859 |
| KIAA1522 | 1.019004838 | 1.009934909 |
| HMGCLL1 | 1.018965872 | 0.927968851 |
| TRIM51 | 1.018901989 | 1.053861129 |
| CLMP | 1.018862339 | 0.96437136 |
| DDX11 | 1.018851854 | 0.917585983 |
| OR1J1 | 1.018830985 | 1.039054471 |
| SSR3 | 1.018830351 | 0.793764988 |
| ADH6 | 1.018803548 | 0.876873228 |
| DNAH5 | 1.018803548 | 0.986229243 |

| | | |
|---|---|---|
| ERMN | 1.018803548 | 1.065208586 |
| TBC1D3C | 1.018803548 | 0.963953017 |
| YWHAQ | 1.018803548 | 0.933576347 |
| ZNF268 | 1.018783184 | 0.738951696 |
| FAS | 1.018770519 | 0.975340688 |
| SRSF7 | 1.018768141 | 1.001027749 |
| ANXA8 | 1.018694205 | 0.839065542 |
| ZNF839 | 1.018654051 | 0.908187736 |
| CIB3 | 1.018645578 | 1.040767386 |
| SHISA3 | 1.018600042 | 0.985954094 |
| RIF1 | 1.018575847 | 1.002740665 |
| ITGAM | 1.018517171 | 1.022610483 |
| OR56A3 | 1.01844744 | 1.028776978 |
| DNAJC3 | 1.018424909 | 1.089914945 |
| MS4A15 | 1.018410512 | 0.882494005 |
| RAB11FIP2 | 1.018393734 | 1.006885379 |
| PLLP | 1.018377382 | 0.968408262 |
| GPX2 | 1.018376685 | 1.001946788 |
| C10orf12 | 1.018373177 | 1.065433368 |
| RBBP9 | 1.01837298 | 0.939000649 |
| S1PR4 | 1.01837298 | 0.939000649 |
| PABPC1L2A | 1.018346869 | 0.903604698 |
| AK9 | 1.018333531 | 0.993833505 |
| TUSC2 | 1.018329363 | 1.024323398 |
| DDAH1 | 1.018323182 | 0.859052248 |
| NOXRED1 | 1.018281833 | 0.983213429 |
| GDAP1L1 | 1.01821192 | 0.928400137 |
| SLC22A14 | 1.018173815 | 0.900993491 |
| STOML1 | 1.018158636 | 1.009249743 |
| ZDHHC18 | 1.018157503 | 0.978759849 |
| LSMEM1 | 1.018152474 | 0.856800274 |
| ADRA1A | 1.018131773 | 0.918234912 |
| ST5 | 1.018093164 | 0.975989617 |
| HIAT1 | 1.018058752 | 1.033744322 |
| GADD45A | 1.01803374 | 1.072093965 |
| NFIX | 1.018003349 | 1.028552888 |
| CYP2A13 | 1.017975359 | 0.965606749 |
| ZNF480 | 1.017949892 | 0.966788173 |
| RASGEF1A | 1.017935215 | 1.014045906 |
| GNAI1 | 1.017896762 | 0.910085055 |
| UNC5A | 1.017880345 | 1.003425831 |
| KNTC1 | 1.017875496 | 0.878383008 |
| ZNF324B | 1.017873991 | 0.887808829 |
| HSD11B2 | 1.017855085 | 0.955223881 |

| | | |
|---|---|---|
| GRHL2 | 1.017829521 | 0.847306602 |
| SLC9A6 | 1.017717523 | 1.002595717 |
| FSD1L | 1.017682288 | 1.175859831 |
| LZTFL1 | 1.017641563 | 1.169921206 |
| FBXO33 | 1.01761503 | 0.876703439 |
| WFDC2 | 1.017505544 | 0.811579308 |
| SSPO | 1.017501873 | 0.950992305 |
| RNF215 | 1.017423135 | 1.092147956 |
| HIVEP3 | 1.017412983 | 0.890238963 |
| TMEM167A | 1.017406666 | 0.88618874 |
| RPH3A | 1.017379175 | 0.985611511 |
| GLTSCR1 | 1.017326027 | 1.106200754 |
| CRHBP | 1.017308673 | 1.025351148 |
| CNRIP1 | 1.017304324 | 0.843782117 |
| WWP2 | 1.017197981 | 1.071382219 |
| BOLA2B | 1.017184752 | 1.037684138 |
| OTOP2 | 1.017184752 | 1.037684138 |
| RAP1A | 1.017164146 | 0.95587281 |
| OOEP | 1.017162968 | 1.006075334 |
| ALK | 1.017143977 | 0.898118105 |
| RAB3GAP2 | 1.017142898 | 0.993924666 |
| PARP4 | 1.017139485 | 0.991899554 |
| TXNDC5 | 1.017118854 | 0.979748886 |
| CCDC124 | 1.01710128 | 0.969623329 |
| FYN | 1.01708207 | 1.060999351 |
| GGPS1 | 1.017074743 | 0.892926671 |
| PPARG | 1.017064959 | 0.949372215 |
| TTC9C | 1.016888503 | 1.077359255 |
| DAO | 1.016741005 | 0.977287476 |
| TAS2R3 | 1.016672436 | 0.986296677 |
| GTF2B | 1.016662139 | 0.809214796 |
| DIP2C | 1.016640475 | 1.144592953 |
| FAM124A | 1.016631204 | 0.949777238 |
| ISM2 | 1.016617246 | 0.943701904 |
| CTNNB1 | 1.01661256 | 0.941676792 |
| FBXO30 | 1.016608958 | 0.966904607 |
| MARVELD2 | 1.016593508 | 0.933576347 |
| TBCE | 1.016561473 | 0.908530319 |
| MAP3K1 | 1.016541865 | 0.961713173 |
| IGFBP4 | 1.016480912 | 0.988009592 |
| VSIG8 | 1.016435135 | 1.045362495 |
| APOL6 | 1.016427953 | 1.215876873 |
| TMEM194A | 1.016385944 | 0.907159986 |
| C9orf114 | 1.016375568 | 1.017129154 |

| | | |
|---|---|---|
| CARD10 | 1.016336139 | 1.156391953 |
| CYP21A2 | 1.016269704 | 0.993510707 |
| UPF3A | 1.016237555 | 0.829393628 |
| CYP2E1 | 1.016202693 | 0.988319273 |
| TMEM39B | 1.0161867 | 0.946132037 |
| KCNK9 | 1.016154369 | 1.08825438 |
| GCM2 | 1.016066366 | 0.977936405 |
| FGD4 | 1.016059714 | 1.070914697 |
| CACNB1 | 1.016051054 | 0.925373134 |
| CNR2 | 1.016044385 | 1.130434783 |
| WNT7A | 1.016041469 | 1.045767517 |
| SEPT11 | 1.01603393 | 0.872809864 |
| NEK10 | 1.01603393 | 0.872809864 |
| EXTL1 | 1.016019871 | 1.037667072 |
| LY86 | 1.016013453 | 0.944158959 |
| TMEM231 | 1.01594345 | 1.046591298 |
| COL7A1 | 1.015924782 | 0.953408702 |
| TC2N | 1.015879926 | 0.91983556 |
| TMX2 | 1.015877269 | 0.862426995 |
| OR52A1 | 1.015862935 | 0.883179171 |
| GNAQ | 1.015849886 | 0.922918808 |
| FRS2 | 1.015843724 | 0.942788626 |
| KIR3DL3 | 1.015803324 | 0.962737951 |
| GFOD2 | 1.015799351 | 1.098825435 |
| SLC9A4 | 1.015760354 | 1.068174032 |
| PPP1R14B | 1.015752879 | 0.904607398 |
| ANKRD22 | 1.015718865 | 0.936411503 |
| GCGR | 1.015712623 | 0.951979234 |
| NOP56 | 1.015688855 | 1.037684138 |
| CETN1 | 1.015686365 | 0.794248684 |
| RANBP1 | 1.01567123 | 0.961288112 |
| SPG20 | 1.015641507 | 0.984241178 |
| FADD | 1.01563952 | 0.9467878 |
| PGBD4 | 1.015587498 | 0.970537855 |
| SPON2 | 1.01555995 | 0.953751285 |
| AGBL3 | 1.015547299 | 1.013770757 |
| COMMD4 | 1.015502995 | 0.940047962 |
| CES4A | 1.015494474 | 0.997569866 |
| STPG1 | 1.015489583 | 0.871607938 |
| AKAP17A | 1.015474149 | 1.115431349 |
| MTHFD1L | 1.015464665 | 0.983776768 |
| FAM24A | 1.015451297 | 0.985954094 |
| IPO7 | 1.015439764 | 0.981368975 |
| ZNF81 | 1.015398058 | 0.992120589 |

| | | |
|---|---|---|
| FUT11 | 1.015391307 | 0.952380952 |
| P4HA2 | 1.015352953 | 1.073329007 |
| KLHL14 | 1.0153508 | 0.895854745 |
| MIA2 | 1.015257644 | 0.885234669 |
| ZAN | 1.015202683 | 0.973963686 |
| OR13J1 | 1.015164397 | 1.082562521 |
| CREG2 | 1.01507926 | 0.94655704 |
| OR7A17 | 1.015052618 | 0.949640288 |
| ABCC11 | 1.01502473 | 0.905256327 |
| DES | 1.014909426 | 0.847711624 |
| RSPH1 | 1.014863133 | 1.036998972 |
| NME1-NME2 | 1.014820588 | 1.085988352 |
| GHITM | 1.014815332 | 0.934566632 |
| DIO1 | 1.0148139 | 1.126541207 |
| AXIN2 | 1.014770838 | 0.983556012 |
| FCGR2C | 1.014755161 | 0.897910243 |
| SPTLC1 | 1.014643326 | 0.974042829 |
| FANCF | 1.014569799 | 1.054128126 |
| RBFOX2 | 1.014567363 | 0.974078574 |
| TFPI2 | 1.014566272 | 0.968851395 |
| HSD11B1L | 1.014542962 | 0.774807614 |
| CTC1 | 1.014531857 | 0.965978129 |
| TTC38 | 1.014531014 | 1.014388489 |
| TSHB | 1.014525033 | 1.060294621 |
| TECRL | 1.014510443 | 0.994518671 |
| HSPBAP1 | 1.014480856 | 1.066461117 |
| LOC652591_pseudogene | 1.014474399 | 0.779364049 |
| ZNF235 | 1.014468532 | 0.931825968 |
| NOLC1 | 1.014463412 | 1.000685166 |
| GINS2 | 1.014445514 | 0.852166869 |
| KIF5C | 1.014409446 | 0.958468527 |
| OR4K1 | 1.014393952 | 0.918122645 |
| ZFAT | 1.014339507 | 0.831915755 |
| CMTR1 | 1.014322545 | 0.97327852 |
| POM121L2 | 1.014315114 | 0.881466256 |
| CLIC5 | 1.014312305 | 1.042829332 |
| CCDC159 | 1.014308997 | 1.091127098 |
| TMEM175 | 1.014286427 | 1.004860267 |
| C16orf59 | 1.014285561 | 0.913325233 |
| TSPY1 | 1.014283397 | 1.048304214 |
| ERVW-1 | 1.014269849 | 0.933538883 |
| ZPBP2 | 1.014255354 | 0.907249899 |
| LRCH2 | 1.01421072 | 1.011647825 |
| YJEFN3 | 1.014178732 | 0.96882494 |

| | | |
|---|---|---|
| NPIP | 1.014166317 | 1.01781432 |
| CTU1 | 1.014131977 | 1.059943297 |
| OR9G4 | 1.014130704 | 0.880095923 |
| VEGFC | 1.014126748 | 1.074626866 |
| MS4A14 | 1.014123011 | 1.057918186 |
| MTR | 1.014106609 | 0.966383151 |
| HGF | 1.014092008 | 1.116807268 |
| C20orf85 | 1.014071937 | 0.863309353 |
| SASH3 | 1.014041942 | 0.987324426 |
| PRSS35 | 1.014039732 | 0.890330954 |
| FOXI2 | 1.013984556 | 1.027541515 |
| TMEM170A | 1.0139463 | 1.019441069 |
| RWDD4 | 1.013943651 | 0.933981369 |
| KCNJ4 | 1.013907357 | 1.059052563 |
| PTGDR | 1.013862738 | 0.879948086 |
| TTL | 1.013832854 | 0.922128488 |
| ZNF57 | 1.013821649 | 0.828270555 |
| TUBGCP2 | 1.013786857 | 0.987039287 |
| OC90 | 1.013780497 | 1.006489293 |
| LIPC | 1.013766175 | 0.982989064 |
| MRPS18A | 1.013755784 | 0.980963953 |
| CCDC96 | 1.013685894 | 1.048197651 |
| GAS6 | 1.013660843 | 1.042822885 |
| TRMT12 | 1.013635972 | 1.038072094 |
| OR2AG1 | 1.013629554 | 0.87727825 |
| LIN37 | 1.013538422 | 0.940461725 |
| FRK | 1.013514675 | 1.075275795 |
| LRRC18 | 1.013485495 | 1.096608428 |
| KCTD7 | 1.01347022 | 0.943543154 |
| SEC23IP | 1.013447952 | 1.001620089 |
| KIAA0513 | 1.013446118 | 0.924260834 |
| PNOC | 1.013439415 | 0.94655704 |
| PODXL2 | 1.013402533 | 1.004796163 |
| GET4 | 1.013324555 | 0.909900651 |
| ZNF394 | 1.013318665 | 0.99109284 |
| ZSWIM2 | 1.013209009 | 1.012329656 |
| SF1 | 1.013186724 | 1.093867763 |
| TMED7 | 1.013186724 | 1.093867763 |
| FAM57B | 1.013154463 | 0.934566632 |
| tcag7.955_pseudogene | 1.013152243 | 1.044878383 |
| SSSCA1 | 1.013124423 | 1.021925317 |
| ELMOD3 | 1.013119853 | 0.922777417 |
| C2orf73 | 1.013115497 | 1.051044878 |
| PQLC1 | 1.013115497 | 1.051044878 |

| | | |
|---|---|---|
| DYNC1I1 | 1.013089843 | 0.938841636 |
| PAQR7 | 1.013076078 | 1.002055498 |
| GBP6 | 1.013069212 | 1.079384366 |
| MEN1 | 1.013065035 | 0.934791414 |
| CDKL1 | 1.013044856 | 1.001946788 |
| PTTG2 | 1.013043183 | 1.063377869 |
| GPR37 | 1.013037694 | 0.959766385 |
| CCNY | 1.012967484 | 0.989874443 |
| TOMM40 | 1.012961627 | 0.996755354 |
| LOC642678_pseudogene | 1.012950679 | 0.954574951 |
| CDH16 | 1.012919397 | 0.981773998 |
| ORC6 | 1.012902406 | 0.916409729 |
| OSBPL11 | 1.012855465 | 1.039054471 |
| YWHAG | 1.012853125 | 1.155094095 |
| TMEM202 | 1.012798821 | 1.030781693 |
| ATP9A | 1.012792289 | 0.986372485 |
| TRAPPC4 | 1.012764982 | 0.967112025 |
| REPS1 | 1.012692407 | 0.945321993 |
| ELF3 | 1.012666416 | 1.060350422 |
| VRK1 | 1.01265909 | 0.896820247 |
| GPR4 | 1.012643942 | 1.018170019 |
| SCRN2 | 1.012631923 | 0.854639844 |
| RCC1 | 1.01260078 | 0.904419322 |
| FAM3D | 1.01258421 | 1.061563386 |
| SBDS | 1.01257228 | 1.030147311 |
| GPR171 | 1.012500864 | 0.928617781 |
| ZAP70 | 1.01245287 | 1.086956522 |
| KRTAP4-11 | 1.012433134 | 1.087016101 |
| HIST1H2BG | 1.012404952 | 0.780404248 |
| CXorf65 | 1.012388396 | 1.029161604 |
| N4BP1 | 1.012383883 | 1.002740665 |
| FLNB | 1.012340204 | 0.947584789 |
| PPFIBP1 | 1.012340204 | 0.947584789 |
| USP34 | 1.012339346 | 1.039584685 |
| ZNF511 | 1.012317284 | 0.890643985 |
| FAM153A | 1.012302643 | 0.985954094 |
| TRIM4 | 1.012280732 | 0.79688514 |
| ARMC6 | 1.012268932 | 0.992120589 |
| FAM229B | 1.012268932 | 0.992120589 |
| PUS10 | 1.012252757 | 0.963001028 |
| SPAG7 | 1.01223563 | 0.966084275 |
| EMB | 1.012230635 | 0.941676792 |
| DRGX | 1.012218004 | 0.936964714 |
| EXOC4 | 1.012191258 | 0.811259619 |

| | | |
|---|---|---|
| ICMT | 1.012170831 | 0.950032446 |
| TMEM63A | 1.012166399 | 1.119076549 |
| PCDH19 | 1.012120951 | 1.151764303 |
| ZNF593 | 1.012095215 | 1.058239123 |
| DDX5 | 1.012069655 | 0.865671642 |
| IFT140 | 1.012047957 | 0.967797191 |
| TAS2R8 | 1.01203808 | 1.003083248 |
| CACNA1F | 1.012037055 | 0.981829981 |
| SOAT2 | 1.011984451 | 0.939649578 |
| CTSC | 1.011967245 | 0.860480208 |
| HNRNPUL1 | 1.011966989 | 0.983213429 |
| FGGY | 1.011904159 | 0.897124342 |
| B3GALNT2 | 1.011855286 | 0.971447112 |
| MUTYH | 1.011855286 | 0.971447112 |
| PSMB7 | 1.011855286 | 0.971447112 |
| SERHL | 1.011846602 | 1.145940391 |
| ABHD14B | 1.011793873 | 1.045425049 |
| UCHL5 | 1.011793873 | 1.045425049 |
| OR2AK2 | 1.011782987 | 0.984926345 |
| SPON1 | 1.011722317 | 0.997259335 |
| SIGLEC5 | 1.011703913 | 0.929931146 |
| CST9L | 1.01168983 | 0.968139774 |
| CTAGE4 | 1.011602609 | 0.912983899 |
| ZNF385D | 1.01156286 | 0.968813285 |
| FBXO22 | 1.011553324 | 0.992861778 |
| KLHL28 | 1.011549412 | 1.023896314 |
| SIAH3 | 1.011507731 | 1.043508051 |
| TM4SF19 | 1.011502974 | 1.081877355 |
| RNF40 | 1.011496439 | 1.103828683 |
| SYT2 | 1.011415436 | 1.02672148 |
| HYPK | 1.011402885 | 0.991435423 |
| SPG21 | 1.01139994 | 1.106543337 |
| PWP1 | 1.011374883 | 0.8441247 |
| C2orf88 | 1.011361064 | 1.000685166 |
| SHD | 1.011347335 | 1.003768414 |
| IFNA21 | 1.011303793 | 0.974648852 |
| ASCC1 | 1.011300938 | 0.936279548 |
| RNF39 | 1.011298287 | 0.903309539 |
| TDRD6 | 1.01124856 | 0.986981843 |
| SLC6A6 | 1.011189825 | 0.867152693 |
| MARCH1 | 1.011179824 | 0.972096042 |
| KAT5 | 1.011172744 | 1.00908501 |
| KCNS1 | 1.011172744 | 1.00908501 |
| DOCK9 | 1.011170582 | 1.085303186 |

| | | |
|---|---|---|
| VKORC1 | 1.011129817 | 0.914135277 |
| RANBP10 | 1.011119428 | 0.976361768 |
| CMPK1 | 1.011101452 | 0.818948735 |
| LRRIQ1 | 1.011092831 | 0.982528263 |
| LRRC16B | 1.011081679 | 0.855002025 |
| TRIM49 | 1.011065806 | 1.188838417 |
| SCUBE2 | 1.011052959 | 1.033230558 |
| ATP5F1 | 1.011016938 | 1.083932854 |
| SFR1 | 1.01099161 | 1.048646797 |
| C1QTNF4 | 1.010920338 | 1.067146283 |
| SNX15 | 1.010920338 | 0.853717026 |
| RCL1 | 1.01085795 | 0.934791414 |
| SPATC1L | 1.010850665 | 0.98582422 |
| NVL | 1.010817394 | 0.91983556 |
| SLC18B1 | 1.010797684 | 1.056526208 |
| AREL1 | 1.010773394 | 0.877352369 |
| AKAP1 | 1.010765379 | 0.932168551 |
| C20orf166 | 1.010752511 | 0.935251799 |
| ACSM1 | 1.010682002 | 0.9145403 |
| DTD2 | 1.010650338 | 1.052072628 |
| HELLS | 1.010641556 | 0.85945727 |
| GPR21 | 1.010630645 | 1.051914341 |
| LIN52 | 1.010587356 | 1.004455245 |
| GRP | 1.010573493 | 0.933881466 |
| PNPLA2 | 1.010554689 | 0.844809866 |
| GPR87 | 1.010554579 | 1.083711875 |
| CCDC132 | 1.010532238 | 1.147428109 |
| SUN5 | 1.010522247 | 1.0890716 |
| TBX4 | 1.010505267 | 0.972744971 |
| BCAS3 | 1.010490621 | 1.003083248 |
| DSC2 | 1.01046697 | 0.961630695 |
| POU2F2 | 1.010427225 | 1.004542505 |
| CLDND2 | 1.010365358 | 0.929121102 |
| OR10A3 | 1.010336886 | 0.947927372 |
| PRPSAP1 | 1.010312872 | 1.004796163 |
| ACTL6B | 1.010303822 | 1.068043742 |
| PIANP | 1.010303822 | 1.068043742 |
| GOLGA8F | 1.010303447 | 0.957177115 |
| IFI35 | 1.010292483 | 0.960260363 |
| ZNF445 | 1.010281518 | 0.963343611 |
| ASIC2 | 1.010259861 | 1.031148605 |
| CLVS2 | 1.010249167 | 0.972593354 |
| ZRANB1 | 1.010231844 | 1.010935601 |
| IRX5 | 1.010206068 | 0.959902795 |

| | | |
|---|---|---|
| FBXO42 | 1.010193427 | 1.062946139 |
| DNMT3B | 1.010172187 | 0.9201817 |
| CNTNAP4 | 1.010149032 | 0.949640288 |
| OXLD1 | 1.010142598 | 1.057211374 |
| PRR27 | 1.010107076 | 0.961973279 |
| TALDO1 | 1.010100895 | 0.951979234 |
| FAM47C | 1.010020799 | 1.033616849 |
| TMPRSS11D | 1.010007541 | 1.052563271 |
| RNF8 | 1.009892124 | 0.941596366 |
| PPP1R9A | 1.009883971 | 0.976019185 |
| LACTB2 | 1.009756742 | 1.019184652 |
| STON2 | 1.009747505 | 0.911300122 |
| DNAJB4 | 1.009738346 | 0.965399109 |
| MTFR1L | 1.009731764 | 1.046172539 |
| INSIG1 | 1.009684564 | 1.10688592 |
| HSF4 | 1.00968383 | 0.95018226 |
| GEMIN8 | 1.009676545 | 1.085054678 |
| PGBD5 | 1.009625147 | 0.944106926 |
| SPSB2 | 1.009624491 | 0.89422453 |
| ZNF451 | 1.009607673 | 1.011647825 |
| SGOL2 | 1.009589504 | 0.954779034 |
| WFDC13 | 1.009582934 | 1.020897568 |
| DCBLD1 | 1.009566713 | 1.027064063 |
| POMGNT1 | 1.009555715 | 0.803159174 |
| C1QL2 | 1.009523669 | 0.913326482 |
| SMIM11 | 1.009497118 | 0.922576225 |
| C11orf63 | 1.009492753 | 0.842041312 |
| S1PR5 | 1.00948166 | 1.058403634 |
| DCTN4 | 1.009480263 | 0.994861254 |
| USP32 | 1.009471699 | 0.989617132 |
| NBL1 | 1.009462547 | 0.934909215 |
| FARP1 | 1.009454119 | 0.937992463 |
| RNLS | 1.009425166 | 1.016443988 |
| TBRG4 | 1.009425166 | 1.016443988 |
| DYNLT1 | 1.009425003 | 0.835965978 |
| DARS | 1.009403353 | 0.921830701 |
| NLRX1 | 1.009335144 | 1.089509923 |
| MITF | 1.009289756 | 0.997569866 |
| GABRE | 1.009225717 | 1.011031798 |
| PRPF3 | 1.009212735 | 0.94653706 |
| B3GNT5 | 1.009189495 | 1.042829332 |
| ROBO2 | 1.009185433 | 0.82288455 |
| LETM1 | 1.009174514 | 1.127440904 |
| ANGPTL1 | 1.009155495 | 1.074626866 |

| | | |
|---|---|---|
| GLRX5 | 1.009148805 | 0.897529364 |
| AQPEP | 1.009135275 | 1.070572114 |
| CYP4B1 | 1.009071376 | 0.805589307 |
| BEX2 | 1.009038723 | 1.01417578 |
| PGAM2 | 1.009020074 | 1.000648929 |
| TRPM5 | 1.009020074 | 1.000648929 |
| OCRL | 1.008961542 | 1.064243997 |
| SHCBP1L | 1.008931443 | 1.044957473 |
| CCDC27 | 1.008923293 | 1.085864723 |
| PFDN4 | 1.008918107 | 1.015416238 |
| WDR55 | 1.008900268 | 1 |
| CARD8 | 1.008878461 | 0.949297705 |
| OLAH | 1.008863083 | 1.059052563 |
| GPR45 | 1.008810149 | 0.990266061 |
| ZNF579 | 1.008809499 | 0.983213429 |
| SRBD1 | 1.008802514 | 0.948967193 |
| RNASE2 | 1.008795586 | 1.030781693 |
| CSNK1G2 | 1.008786193 | 1.022063595 |
| KRT5 | 1.008779686 | 0.998629668 |
| SLC47A1 | 1.008768689 | 0.781287971 |
| TIMM10 | 1.008700938 | 1.041795135 |
| MRC1 | 1.008682795 | 1.016872161 |
| FBF1 | 1.008664554 | 0.966426859 |
| PET112 | 1.008655793 | 0.884490591 |
| APOL3 | 1.008645009 | 1.080467229 |
| CNTD1 | 1.008638996 | 1.055488052 |
| PLA2G2D | 1.008614549 | 0.948085659 |
| ZMYM2 | 1.008611284 | 0.971648441 |
| MYEOV | 1.008580731 | 0.912641316 |
| MSR1 | 1.008554042 | 0.847501622 |
| UNC80 | 1.008525136 | 0.943473792 |
| C16orf93 | 1.008503886 | 0.955806783 |
| SCAMP5 | 1.00849351 | 0.961973279 |
| TMEM68 | 1.008482151 | 0.959497772 |
| TMEM106A | 1.008480521 | 1.039287161 |
| ATIC | 1.008428715 | 0.994329688 |
| EFCAB5 | 1.008401716 | 0.714054273 |
| ATAT1 | 1.008391745 | 1.02672148 |
| TNFRSF9 | 1.008356137 | 1.033095393 |
| TMEM164 | 1.008343543 | 0.986229243 |
| MED15 | 1.008322421 | 1.076053443 |
| TIMELESS | 1.008322421 | 0.717368962 |
| NBPF10 | 1.008311683 | 0.963686194 |
| BRWD3 | 1.008278696 | 0.980153908 |

| | | |
|---|---|---|
| UBA5 | 1.008258419 | 0.996106424 |
| WIPI1 | 1.008167952 | 0.931146213 |
| TMEM139 | 1.008149868 | 1.045767517 |
| PDGFRA | 1.008149588 | 0.99091499 |
| RBM7 | 1.008066734 | 1.037667072 |
| OGFOD3 | 1.008039522 | 1.049318624 |
| RAB6A | 1.008039522 | 0.953926022 |
| FAR2P1 | 1.008034615 | 1.045563549 |
| PYDC2 | 1.008030668 | 1.072498987 |
| IL24 | 1.007916855 | 0.997944502 |
| ANXA4 | 1.007845266 | 1.071942446 |
| MGAT2 | 1.007841781 | 1.102530824 |
| ZDHHC6 | 1.007827323 | 0.910243234 |
| CYCS | 1.007816923 | 0.976508708 |
| ANKMY1 | 1.007814591 | 0.922576225 |
| USP31 | 1.007791729 | 0.879948086 |
| C10orf54 | 1.007734665 | 1.00890716 |
| PREX2 | 1.007694204 | 0.847207948 |
| NF2 | 1.007681269 | 0.927501013 |
| SUN2 | 1.007661148 | 0.851761847 |
| KANSL3 | 1.007623793 | 0.927372388 |
| GJA8 | 1.007601974 | 0.996755354 |
| ZIC2 | 1.007562171 | 0.99108951 |
| ZNF792 | 1.007560388 | 0.954232483 |
| PDZK1 | 1.007558453 | 0.917375456 |
| FAM161B | 1.007530817 | 1.059952038 |
| NME9 | 1.007464111 | 0.946132037 |
| MSN | 1.007461901 | 0.889003083 |
| HSH2D | 1.00741043 | 0.975334018 |
| AVL9 | 1.007402093 | 0.990750257 |
| FAM196B | 1.007387956 | 1.018499486 |
| NDUFAF5 | 1.007387956 | 1.018499486 |
| ANKRD42 | 1.007327694 | 0.970838396 |
| PRCC | 1.007276812 | 0.869133265 |
| DCAKD | 1.007257956 | 1.036452005 |
| MEPE | 1.007230732 | 0.983213429 |
| MBD3 | 1.007226427 | 0.99554642 |
| TMEM99 | 1.007212569 | 1.032401782 |
| LOC642446 | 1.00720781 | 0.917585983 |
| MEPCE | 1.007182259 | 1.065208586 |
| C11orf52 | 1.007166977 | 1.028351559 |
| UCP2 | 1.007113847 | 0.917780478 |
| C10orf129 | 1.007063086 | 0.941760877 |
| ZNF607 | 1.007043739 | 1.052757794 |

| | | |
|---|---|---|
| ZNF614 | 1.007033363 | 1.123672491 |
| TUBB1 | 1.006994511 | 0.939000649 |
| TYSND1 | 1.006957959 | 0.907203115 |
| DFNA5 | 1.006879854 | 0.952723535 |
| MRPS17 | 1.006879854 | 0.992805755 |
| OR8B8 | 1.006879854 | 0.8663926 |
| PCDHGA10 | 1.006879854 | 0.872559096 |
| SELK | 1.006879854 | 0.98663926 |
| TPRA1 | 1.006872014 | 0.933809215 |
| ATF7IP | 1.006790843 | 0.960415315 |
| TRAPPC1 | 1.006709626 | 1.013018157 |
| GPR133 | 1.006702263 | 0.896820247 |
| NLRP7 | 1.006614939 | 0.981773998 |
| FAM63B | 1.006564406 | 0.977723775 |
| KLHDC1 | 1.006561237 | 1.082562521 |
| WBP11 | 1.006550953 | 1.098637249 |
| SARNP | 1.006533689 | 0.996231586 |
| PPP1R12A | 1.006525927 | 1.00843608 |
| SYT9 | 1.006514387 | 0.943816375 |
| GPA33 | 1.006508361 | 0.928400137 |
| ZHX2 | 1.006496044 | 0.918234912 |
| ARF6 | 1.006476698 | 0.976638546 |
| ZCRB1 | 1.006443729 | 0.934791414 |
| EBP | 1.006436342 | 0.967598218 |
| MRPL2 | 1.006422945 | 1.033211827 |
| CEACAM16 | 1.006379137 | 0.9963548 |
| LRRC48 | 1.006379137 | 0.9963548 |
| FAIM | 1.00635825 | 0.991778006 |
| TIGD3 | 1.006356574 | 0.988694758 |
| ADRA1D | 1.006356094 | 0.971447112 |
| TNN | 1.006349959 | 0.976361768 |
| SSNA1 | 1.00634656 | 0.970195272 |
| ZNF395 | 1.00634656 | 0.970195272 |
| PHACTR3 | 1.006334463 | 0.948612539 |
| CRYGD | 1.006325089 | 1.025111381 |
| MEIG1 | 1.006308381 | 0.924665857 |
| PAQR4 | 1.006296539 | 0.886947585 |
| FKBP3 | 1.006295951 | 0.725799919 |
| RLN3 | 1.006194048 | 1.005823912 |
| BOD1L1 | 1.006177057 | 1.045767517 |
| PSME1 | 1.006150109 | 1.010935601 |
| CD7 | 1.006149282 | 0.944158959 |
| TFIP11 | 1.006116841 | 0.904076739 |
| APPBP2 | 1.006103656 | 0.8886605 |

| | | |
|---|---|---|
| TMED6 | 1.006066321 | 1.059952038 |
| KIAA1217 | 1.006034332 | 1.019869818 |
| HRK | 1.006018791 | 1.001370332 |
| TPGS1 | 1.006010726 | 0.992120589 |
| PI16 | 1.00597996 | 0.958204865 |
| PCNXL4 | 1.005956218 | 0.933538883 |
| BEST4 | 1.005945585 | 1.091130012 |
| RAC3 | 1.005934429 | 0.961927906 |
| C9orf116 | 1.005924637 | 0.902706406 |
| FAM167A | 1.005918022 | 0.896539911 |
| IL11RA | 1.005907329 | 0.959902795 |
| ACBD7 | 1.005894325 | 0.874957177 |
| GPC1 | 1.005887302 | 0.868790682 |
| TBC1D2B | 1.005883896 | 0.894289186 |
| CDC25A | 1.005878702 | 1.00908501 |
| QRSL1 | 1.005878702 | 1.00908501 |
| VPS35 | 1.005851462 | 1.006166495 |
| OR8S1 | 1.005812134 | 0.969167523 |
| DPF3 | 1.005758921 | 1.147306944 |
| PIGN | 1.005758921 | 1.003893576 |
| ATP8A1 | 1.005717067 | 0.851356825 |
| ZNF720 | 1.005702396 | 1.070878898 |
| ATCAY | 1.00568572 | 1.010962658 |
| OR51T1 | 1.00567403 | 0.858170606 |
| HNF4A | 1.005657722 | 0.941676792 |
| CPXM1 | 1.005653054 | 1.142115509 |
| IL1RAP | 1.005567026 | 0.881894873 |
| KIAA1524 | 1.00553898 | 0.96435804 |
| CXCL14 | 1.005538464 | 0.899965742 |
| PRAMEF8 | 1.005537829 | 1.028091812 |
| IQUB | 1.005507601 | 0.993114621 |
| LAD1 | 1.005492973 | 1.119218911 |
| KLK7 | 1.005474495 | 0.935107073 |
| ANKZF1 | 1.005425791 | 0.987039287 |
| PIBF1 | 1.005424374 | 0.947927372 |
| ASB7 | 1.005392007 | 0.988319273 |
| SLBP | 1.005354869 | 0.904761905 |
| LRGUK | 1.005296658 | 1.069258809 |
| CALHM3 | 1.005242184 | 1.053100377 |
| CSRNP3 | 1.005230792 | 0.972863507 |
| URB2 | 1.005223879 | 0.937307297 |
| CLDN9 | 1.005214186 | 1.032401782 |
| MPZL1 | 1.005202856 | 1.028434395 |
| RPAP3 | 1.005182557 | 1.016101405 |

| | | |
|---|---|---|
| CCNE2 | 1.005174426 | 0.951330305 |
| VARS | 1.005174045 | 0.968813285 |
| TWF2 | 1.005167594 | 1.062946139 |
| TRIM68 | 1.005140921 | 0.977936405 |
| GCNT3 | 1.0051337 | 1.026326448 |
| SLC35D3 | 1.005094656 | 0.869475848 |
| C20orf141 | 1.005081879 | 0.863309353 |
| PCCA | 1.005079096 | 1.031148605 |
| EGR1 | 1.005075392 | 0.919532771 |
| FNDC9 | 1.005070733 | 1.048646797 |
| GTPBP6 | 1.005035997 | 0.899149453 |
| FUOM | 1.005004626 | 1.011647825 |
| OR8D4 | 1.004993117 | 1.005481329 |
| OR2V2 | 1.004987227 | 1.002398082 |
| TMEM160 | 1.004955715 | 1.042932361 |
| DPM1 | 1.004942318 | 0.952612394 |
| RPH3AL | 1.004933399 | 0.974648852 |
| RAB9A | 1.004905896 | 0.771972458 |
| THRAP3 | 1.004893364 | 0.860267315 |
| NKG7 | 1.004886538 | 1.008100446 |
| CDRT4 | 1.004815426 | 0.884973674 |
| GPSM2 | 1.004799555 | 0.994861254 |
| C16orf62 | 1.004744232 | 0.888317917 |
| FAM114A2 | 1.004726153 | 0.960945529 |
| NUDT1 | 1.00464196 | 0.930564568 |
| RC3H2 | 1.004622449 | 0.957170668 |
| PARVA | 1.00460853 | 1.063035286 |
| SYNM | 1.004578489 | 0.899280576 |
| HGS | 1.004512383 | 1.019869818 |
| FAM35A | 1.004505178 | 0.871531346 |
| GIGYF2 | 1.004505178 | 1.016786571 |
| CDC45 | 1.004453661 | 0.85303186 |
| KLK5 | 1.004434176 | 1.058403634 |
| KIAA0895L | 1.004433816 | 1.128126071 |
| WDR27 | 1.004426854 | 1.061968408 |
| LANCL2 | 1.00442498 | 0.913326482 |
| IDNK | 1.004392673 | 0.94491697 |
| IFNE | 1.004382888 | 0.897910243 |
| NDUFA2 | 1.004366589 | 0.9201817 |
| TMCO4 | 1.00434365 | 1.088043851 |
| BTG2 | 1.004343373 | 0.9733939 |
| C1orf52 | 1.004313512 | 0.825840421 |
| IL17C | 1.004300787 | 1.003083248 |
| NAV3 | 1.004231055 | 1.041795135 |

| | | |
|---|---|---|
| RNGTT | 1.004203753 | 0.994808566 |
| TGIF1 | 1.004127835 | 1.068448765 |
| CD207 | 1.004099431 | 0.992463172 |
| LRRC25 | 1.004073242 | 0.983213429 |
| ORC4 | 1.004068745 | 0.980153908 |
| DNASE1L2 | 1.004030387 | 0.754151478 |
| PRPF31 | 1.003966855 | 1.006509078 |
| SPINT3 | 1.003963502 | 0.946214457 |
| POLG | 1.003916586 | 1.081004455 |
| NDNF | 1.003914039 | 0.9145403 |
| ERVFRD-1 | 1.003862386 | 0.883758607 |
| FLJ46321 | 1.003856834 | 1.021466181 |
| CDCP1 | 1.003839463 | 0.992709599 |
| EPN3 | 1.003836363 | 0.963343611 |
| OR8K1 | 1.003821321 | 0.845837616 |
| LHPP | 1.003787856 | 0.846203764 |
| C6orf118 | 1.00377728 | 1.056183625 |
| ZSWIM4 | 1.003723497 | 0.983556012 |
| CLEC6A | 1.003713665 | 0.980472765 |
| PLIN5 | 1.003646502 | 1.007290401 |
| ACSS3 | 1.003641403 | 0.86761843 |
| MYO16 | 1.003622662 | 0.978533819 |
| RTTN | 1.003600165 | 0.94655704 |
| DEF8 | 1.003567768 | 0.937307297 |
| CATSPERB | 1.003551366 | 1.036313806 |
| NCKIPSD | 1.003545838 | 0.931140802 |
| S100A7L2 | 1.003519508 | 1.025921426 |
| CCDC152 | 1.00351308 | 0.87084618 |
| SLC7A2 | 1.003489745 | 0.915724563 |
| UNC13C | 1.00347946 | 0.963686194 |
| RHOT2 | 1.003466773 | 0.909558068 |
| GPKOW | 1.003462423 | 0.807468311 |
| P2RY4 | 1.003415426 | 0.990266061 |
| PSMD8 | 1.003213175 | 0.940733128 |
| CHP2 | 1.003212133 | 1.081534772 |
| ARF4 | 1.003144006 | 0.979883193 |
| CNTLN | 1.003061887 | 0.948612539 |
| NRROS | 1.003036876 | 1.121959575 |
| EHMT1 | 1.003035321 | 0.942081814 |
| HIST1H2BI | 1.003025322 | 1.029119561 |
| C3AR1 | 1.003006033 | 0.974691759 |
| ACSM5 | 1.002977453 | 1.042932361 |
| RNF144B | 1.002952029 | 0.921479559 |
| C18orf8 | 1.002938373 | 0.962658445 |

| | | |
|---|---|---|
| C12orf45 | 1.002935581 | 1.01417578 |
| GDPD2 | 1.002836282 | 0.942894225 |
| OR11H4 | 1.002810554 | 0.890030832 |
| TMC3 | 1.002794469 | 0.92874272 |
| ATXN3 | 1.002792541 | 0.979343864 |
| GPR110 | 1.002789358 | 1.01752109 |
| CEP250 | 1.002767193 | 0.922576225 |
| SLAMF7 | 1.002766559 | 0.96437136 |
| C17orf104 | 1.002759429 | 1.080194411 |
| ARL6 | 1.002740642 | 1 |
| PLEKHG3 | 1.002726064 | 0.975293641 |
| ARSE | 1.002725722 | 0.96430889 |
| DACH1 | 1.002725722 | 0.96430889 |
| DAZL | 1.002705076 | 0.897124342 |
| GADD45B | 1.002699772 | 0.948955122 |
| FAM122B | 1.002692647 | 0.97326853 |
| KIAA1191 | 1.002691571 | 1.029462145 |
| PSMC4 | 1.002687689 | 1.038935756 |
| ZSCAN1 | 1.002595288 | 0.885577252 |
| SNRPE | 1.002511647 | 0.947584789 |
| OSBPL7 | 1.002491937 | 1.021925317 |
| IRAK4 | 1.00248007 | 0.980532122 |
| ZNF804B | 1.002475285 | 1.086674767 |
| ARHGAP24 | 1.002445689 | 0.983799109 |
| MDGA2 | 1.002429235 | 1.033211827 |
| MAGOHB | 1.002424698 | 1.045220966 |
| SKA2 | 1.002424698 | 0.929085303 |
| OR7C2 | 1.002334214 | 0.910585817 |
| ARPC4 | 1.002320936 | 0.926285946 |
| HELQ | 1.002286528 | 1.023361454 |
| FASTKD2 | 1.00224473 | 0.946942082 |
| PHOX2B | 1.002239109 | 1.071382219 |
| GLI1 | 1.002201431 | 1.019036047 |
| STEAP4 | 1.002101843 | 0.914135277 |
| EHHADH | 1.002094468 | 0.916937054 |
| PSMD13 | 1.00205402 | 1.000685166 |
| C9orf84 | 1.002039158 | 0.997601918 |
| RNASE9 | 1.002039158 | 0.997601918 |
| SRCIN1 | 1.002025611 | 1.101404591 |
| MUC1 | 1.002024659 | 1.030147311 |
| LECT2 | 1.002023617 | 0.958890031 |
| TAT | 1.00197691 | 1.060999351 |
| SART1 | 1.001960274 | 0.94655704 |
| TOR1B | 1.001940474 | 0.907845152 |

| | | |
|---|---|---|
| LNP1 | 1.00192371 | 0.904761905 |
| HSPA2 | 1.00189541 | 1.082414017 |
| DMRT2 | 1.001880921 | 0.88513952 |
| ZNF234 | 1.001865532 | 0.859883522 |
| ACOT9 | 1.00185249 | 0.949372215 |
| SFTPB | 1.001836489 | 0.957519699 |
| DEFB121 | 1.00182865 | 0.921891059 |
| SUPT6H | 1.001817038 | 0.981181051 |
| HSDL2 | 1.00180858 | 0.922640745 |
| ITLN1 | 1.001720179 | 0.935936965 |
| RAD51B | 1.001710826 | 1.038072094 |
| PARPBP | 1.001696766 | 1.0854597 |
| BEST3 | 1.0016952 | 0.927968851 |
| TRIM62 | 1.001672232 | 0.975989617 |
| ST3GAL3 | 1.001669186 | 1.146658014 |
| SCIN | 1.001668662 | 0.926687222 |
| RBBP5 | 1.001662862 | 0.991778006 |
| CPEB1 | 1.00164389 | 1.034021871 |
| BRCA2 | 1.001577464 | 0.838396112 |
| ABHD17A | 1.001543335 | 0.908464966 |
| USP17L9P | 1.001543335 | 0.908464966 |
| ANKRD10 | 1.001505094 | 0.8663926 |
| FMR1NB | 1.001503327 | 1.058581706 |
| RNASEH1 | 1.001499106 | 1.066839714 |
| CNTNAP5 | 1.001489372 | 0.895854745 |
| PCDHGA2 | 1.001487605 | 1.055498458 |
| SCN1A | 1.00145778 | 0.944192083 |
| PLN | 1.00145261 | 1.093965168 |
| CDT1 | 1.001444243 | 1.078794108 |
| ATOH1 | 1.001425485 | 0.948612539 |
| IFFO2 | 1.001421893 | 1.04455245 |
| ACTA2 | 1.00139617 | 0.974991435 |
| ZNF397 | 1.00134207 | 1.027749229 |
| BPIFC | 1.001317014 | 0.992120589 |
| HELZ | 1.001305317 | 0.939000649 |
| RECQL | 1.001305317 | 0.939000649 |
| SRD5A1 | 1.001305317 | 0.939000649 |
| C10orf88 | 1.001288242 | 0.894484412 |
| ZNF813 | 1.001275746 | 0.941676792 |
| USH1C | 1.001267762 | 1.044878383 |
| RGL2 | 1.001257749 | 1.012332991 |
| WWC3 | 1.001230969 | 1.009720535 |
| NEK6 | 1.001228427 | 1.056456846 |
| NONO | 1.001223268 | 0.823227133 |

| | | |
|---|---|---|
| CRTAC1 | 1.00120587 | 1.003083248 |
| SPACA5B | 1.001190238 | 0.879068174 |
| SPRED3 | 1.001155485 | 1.114765331 |
| XIAP | 1.001137048 | 0.789746918 |
| SPANXB2 | 1.001114707 | 1.017129154 |
| TAF12 | 1.0010999 | 0.978938842 |
| MAPT | 1.001099256 | 0.984583762 |
| MSS51 | 1.001099256 | 0.984583762 |
| LOC441294 | 1.001093275 | 1.072970195 |
| ASXL1 | 1.001044431 | 0.975334018 |
| PEG10 | 1.001025945 | 0.913326482 |
| CNOT6L | 1.000968286 | 0.832576249 |
| ABI2 | 1.000964098 | 0.874614594 |
| THOC5 | 1.000960375 | 0.948157149 |
| SH3BGRL2 | 1.000955081 | 1.018842069 |
| ANKRD13A | 1.0009211 | 0.830700689 |
| TMEM41B | 1.000912399 | 0.953751285 |
| MEIOB | 1.000877394 | 1.10490077 |
| PDIA2 | 1.000877394 | 0.966788173 |
| SLC25A11 | 1.000874437 | 1.019467878 |
| TRIM27 | 1.000870157 | 0.805970149 |
| CRB3 | 1.000807508 | 0.908872902 |
| RPA4 | 1.000797519 | 0.985419198 |
| NPHP3 | 1.00079609 | 1.020554985 |
| GINS4 | 1.00078453 | 0.847306602 |
| OR4F29 | 1.000770218 | 0.988009592 |
| ABCA12 | 1.000744949 | 1.096800324 |
| OR2S2 | 1.000733608 | 0.926002055 |
| BUD31 | 1.00069569 | 0.865937627 |
| BTG3 | 1.000684764 | 1.05789654 |
| MRPL37 | 1.000659798 | 0.887290168 |
| CNFN | 1.000655702 | 0.81855002 |
| ANKRD27 | 1.000630075 | 1.048646797 |
| SUPT20HL1 | 1.000616256 | 0.881123672 |
| BMP8B | 1.000613802 | 1.057105775 |
| MMP19 | 1.000552389 | 0.961064244 |
| HHLA2 | 1.000543715 | 0.598835218 |
| GEMIN2 | 1.000483136 | 0.997601918 |
| BPIFB6 | 1.000446164 | 0.965056526 |
| USP45 | 1.000437466 | 1.003893576 |
| BARX2 | 1.000424128 | 0.987849332 |
| ATP6V1B2 | 1.000415213 | 0.942486837 |
| NDUFA10 | 1.000360045 | 1.029161604 |
| STMN3 | 1.000289211 | 1.15142172 |

| | | |
|---|---|---|
| GAL | 1.000280829 | 0.993148338 |
| NDRG2 | 1.000237667 | 1.020116807 |
| WFDC3 | 1.000233815 | 0.977723775 |
| FLG2 | 1.000185271 | 1.030489894 |
| RUNX3 | 1.000144642 | 1.13238157 |
| KLHL6 | 1.000076709 | 1.064748201 |
| ARMC12 | 1.00006289 | 1.011990408 |
| DACT2 | 1.000021069 | 1.005823912 |
| TMEM145 | 0.99999623 | 0.899554475 |
| COL24A1 | 0.999958496 | 0.946899623 |
| CLCF1 | 0.999953899 | 0.897529364 |
| BTC | 0.999951963 | 1.006885379 |
| LIX1 | 0.999928531 | 0.9181855 |
| NEU1 | 0.999913962 | 1.004860267 |
| CMTM8 | 0.99988676 | 1.068853787 |
| DAZ2 | 0.999833213 | 0.983127839 |
| UBXN4 | 0.999829455 | 0.905104488 |
| CETN3 | 0.999820665 | 0.928400137 |
| RSPO1 | 0.999802811 | 0.912110166 |
| SENP6 | 0.999791146 | 1.004542505 |
| NUP50 | 0.999782695 | 1.117848578 |
| CEP170B | 0.999782287 | 1.044878383 |
| EMILIN3 | 0.999760428 | 1.040097205 |
| TMEM223 | 0.999740104 | 1.038711888 |
| SETD7 | 0.999727345 | 0.865671642 |
| IDH3G | 0.999719931 | 0.951397327 |
| SUSD2 | 0.999697513 | 1.032545392 |
| PIWIL3 | 0.999617043 | 0.973621103 |
| MPHOSPH8 | 0.999613736 | 0.996916752 |
| PLAC4 | 0.999611021 | 1.03199676 |
| CCNB2 | 0.999580813 | 1.073309032 |
| MEX3D | 0.999565992 | 1.042180402 |
| ATL3 | 0.999563086 | 0.879303362 |
| PFKFB4 | 0.999558994 | 0.951330305 |
| C1orf86 | 0.999535426 | 1.027946537 |
| ZNF311 | 0.999498106 | 1.051387461 |
| RAB18 | 0.999485644 | 0.994159637 |
| DAZAP1 | 0.999476357 | 0.955121617 |
| MTMR2 | 0.999470168 | 0.903309539 |
| MAGEA3 | 0.999408936 | 1.015758822 |
| SLC26A11 | 0.999363065 | 0.967553537 |
| RP13-36C9.3 | 0.999348629 | 0.89311408 |
| CREB3L1 | 0.999293805 | 1.000342583 |
| ZNF20 | 0.999275046 | 0.907159986 |

| | | |
|---|---|---|
| TRAF2 | 0.999261861 | 1.064063035 |
| SPAG11A | 0.999243103 | 0.880781089 |
| PAX9 | 0.999228664 | 0.906439854 |
| ULBP3 | 0.999216914 | 0.967797191 |
| CLDN2 | 0.999203774 | 0.921205892 |
| SYT15 | 0.999199108 | 0.988009592 |
| LOC348174 | 0.999196751 | 0.897910243 |
| KCNG2 | 0.999157586 | 0.871512005 |
| COL18A1 | 0.999134723 | 1.135662898 |
| UBD | 0.999126386 | 0.892926671 |
| TRDN | 0.999104818 | 0.798561151 |
| GLUD1 | 0.999093045 | 1.048020766 |
| POLR2C | 0.999084809 | 0.941676792 |
| NARG2 | 0.999067574 | 0.949297705 |
| HRG | 0.999058331 | 1.146625557 |
| ZNRF1 | 0.99899047 | 1 |
| PRTG | 0.998989007 | 0.983556012 |
| C1orf213 | 0.998976561 | 0.811664642 |
| KCNK12 | 0.998951449 | 0.887735237 |
| ZNF227 | 0.998929198 | 1.08461802 |
| LY6G6D | 0.998926117 | 0.954093868 |
| MDH1B | 0.998922142 | 1.064243997 |
| ZNF260 | 0.998915636 | 0.912920211 |
| AP1M2 | 0.998891546 | 1.014731072 |
| TBC1D10B | 0.998888782 | 0.971223022 |
| NDUFAF4 | 0.998864315 | 1.118876328 |
| PDCD10 | 0.998776913 | 0.979102432 |
| LAMP3 | 0.998759922 | 1.019527235 |
| MRPL21 | 0.998725577 | 0.972935937 |
| RBMY1J | 0.998725577 | 0.972935937 |
| SEMA3B | 0.998725577 | 0.972935937 |
| ANKRD24 | 0.99869966 | 0.969852689 |
| KCNJ12 | 0.998693366 | 0.968202466 |
| MFSD12 | 0.998666901 | 0.986981843 |
| ASIC1 | 0.998657884 | 1.011031798 |
| C6orf89 | 0.998620957 | 0.960602946 |
| USP51 | 0.998593949 | 1.041312272 |
| MRPL44 | 0.998593635 | 0.853374443 |
| RBM46 | 0.998570754 | 1.058581706 |
| LUC7L2 | 0.998558566 | 0.994861254 |
| ZC3H18 | 0.998508624 | 1.016605913 |
| PCDHGA11 | 0.998497443 | 1.049331963 |
| IL9R | 0.998410514 | 0.971243418 |
| PIK3C3 | 0.99836432 | 0.957819598 |

| | | |
|---|---|---|
| TOR1AIP2 | 0.998361815 | 1.130417173 |
| RELL1 | 0.99834697 | 1.051044878 |
| DUSP4 | 0.998342669 | 1.000648929 |
| SFTPD | 0.998263147 | 0.940733128 |
| KLHDC2 | 0.998245703 | 1.038711888 |
| JMJD8 | 0.998208892 | 0.981773998 |
| CXCL9 | 0.998124455 | 0.945529291 |
| PPP1R18 | 0.998084537 | 0.90202124 |
| FTL | 0.998067999 | 1.037341555 |
| PURB | 0.998054361 | 0.898937992 |
| LSM8 | 0.998053228 | 1.074683111 |
| CHRDL2 | 0.998050736 | 0.976704351 |
| OR51V1 | 0.99804761 | 0.878725591 |
| EFEMP2 | 0.998022735 | 0.973621103 |
| SPIN4 | 0.998019599 | 1.052247874 |
| ACOT12 | 0.998017327 | 1.011680727 |
| CHRNA10 | 0.998002015 | 1.118754056 |
| CDNF | 0.998001802 | 0.990750257 |
| AGAP2 | 0.997965373 | 1.102774923 |
| PLEKHA8P1 | 0.997883588 | 0.986229243 |
| KIAA0930 | 0.997880236 | 0.958204865 |
| C19orf43 | 0.997863365 | 0.965573107 |
| RPF2 | 0.997863365 | 0.965573107 |
| BRWD1 | 0.997858424 | 1.044147428 |
| CDK8 | 0.997857291 | 0.899415964 |
| GPR62 | 0.997857291 | 0.920830629 |
| NOTCH2NL | 0.997843309 | 0.992463172 |
| GEMIN6 | 0.99781055 | 0.931825968 |
| ZNF140 | 0.997762975 | 0.945871874 |
| SERPINA9 | 0.997655863 | 1.065775951 |
| DCSTAMP | 0.997614133 | 1.005138746 |
| ACOX2 | 0.997551954 | 1.060350422 |
| SMYD3 | 0.997539112 | 1.01752109 |
| AKAP14 | 0.997458696 | 1.00526529 |
| KRTAP20-1 | 0.997444403 | 1.078451524 |
| CERS6 | 0.997443363 | 1.0818145 |
| ARMCX6 | 0.997415652 | 1.003240178 |
| SURF4 | 0.99738106 | 0.980472765 |
| ZCCHC2 | 0.99738106 | 1.089414183 |
| MARK4 | 0.997377594 | 1.012329656 |
| S100A2 | 0.997339511 | 0.940047962 |
| RP9 | 0.997336974 | 0.885577252 |
| HPS5 | 0.997279793 | 1.059952038 |
| MRPL20 | 0.997261261 | 0.896539911 |

| | | |
|---|---|---|
| SLURP1 | 0.99725193 | 1.053057918 |
| NDUFB2 | 0.99723079 | 1.013770757 |
| UBR2 | 0.99715458 | 0.921479559 |
| MAL2 | 0.997154376 | 0.904419322 |
| MRPL55 | 0.997150072 | 1.099006509 |
| PRAP1 | 0.997138019 | 1.0445358 |
| CPSF3 | 0.997119397 | 1.007194245 |
| AP3M1 | 0.997099234 | 0.969852689 |
| NCBP2 | 0.997094205 | 0.986981843 |
| ATHL1 | 0.997092691 | 1.026326448 |
| CTAGE1 | 0.997080204 | 1.038369305 |
| LIPT1 | 0.997049453 | 1.001946788 |
| IFNB1 | 0.997040732 | 0.815309842 |
| ERBB2IP | 0.997018175 | 1.049331963 |
| DDA1 | 0.997013123 | 1.0036452 |
| ZPLD1 | 0.997013123 | 1.0036452 |
| TMEM203 | 0.997012195 | 0.926344639 |
| MOSPD1 | 0.997008479 | 0.943473792 |
| LY6K | 0.997001592 | 0.9777321 |
| FAM32A | 0.996938929 | 0.971565605 |
| CPS1 | 0.996903329 | 1.018170019 |
| NR3C2 | 0.996882831 | 0.830629461 |
| CCER1 | 0.996864836 | 1.072093965 |
| ZBTB43 | 0.996846599 | 0.939651681 |
| SBSPON | 0.996840743 | 1.047961631 |
| GHSR | 0.996839117 | 0.953926022 |
| FAM81A | 0.996791125 | 0.993519644 |
| CEPT1 | 0.99676552 | 0.889682025 |
| PSPC1 | 0.996754156 | 1.090099349 |
| FAM180A | 0.99675035 | 1.021582734 |
| NEURL2 | 0.996738601 | 0.723556132 |
| KRT12 | 0.996716459 | 0.984241178 |
| PCDHGB3 | 0.996709391 | 0.915724563 |
| YBX3 | 0.996700197 | 1.008100446 |
| GUK1 | 0.996695628 | 1.11486048 |
| MORF4L1 | 0.99668882 | 1.015416238 |
| OR5H1 | 0.996686736 | 0.998287085 |
| KLHL15 | 0.99668225 | 0.964028777 |
| ZNF329 | 0.996655675 | 0.894289186 |
| VBP1 | 0.996646637 | 0.943816375 |
| CLPP | 0.996598981 | 1.029201817 |
| PIGQ | 0.996579307 | 1.071599863 |
| ITK | 0.996572225 | 1.007787151 |
| ADAD1 | 0.996544318 | 0.986372485 |

| | | |
|---|---|---|
| SH2D1A | 0.996514967 | 1.048304214 |
| HMG20B | 0.996497147 | 1.190136275 |
| CAPRIN1 | 0.99649664 | 1.128392062 |
| TRIP4 | 0.996491631 | 1.066839714 |
| SLC25A45 | 0.996448807 | 0.867557716 |
| LTBP4 | 0.996416147 | 1.021925317 |
| AXL | 0.996391608 | 1.104477612 |
| PSMG2 | 0.996319547 | 0.947242206 |
| FEZ1 | 0.996312207 | 0.99554642 |
| SCRT1 | 0.9962944 | 0.961288112 |
| RPL27A | 0.99628117 | 0.878725591 |
| SMARCE1 | 0.99626364 | 0.915350344 |
| CENPBD1 | 0.996199431 | 1.065775951 |
| UNC5CL | 0.996179132 | 1.031517643 |
| KITLG | 0.996163441 | 1.002430134 |
| CACNA2D4 | 0.996074377 | 1.05344296 |
| MMEL1 | 0.995994234 | 0.868916288 |
| ACTR2 | 0.995939445 | 0.961630695 |
| CHN1 | 0.99591389 | 1.038026721 |
| ITPRIP | 0.995911036 | 1.069201781 |
| MCPH1 | 0.995904257 | 0.955042527 |
| FN3K | 0.995889107 | 0.987021415 |
| GNGT2 | 0.99586559 | 1.01781432 |
| GPR15 | 0.995845476 | 1.046073978 |
| SRP54 | 0.995815296 | 1.028776978 |
| NUDT21 | 0.995791526 | 0.986229243 |
| SLC9A8 | 0.995762918 | 0.977389517 |
| KLHL10 | 0.995761197 | 0.946214457 |
| MB21D2 | 0.995720872 | 0.911705144 |
| AEBP1 | 0.995713841 | 0.981829981 |
| LDLR | 0.995713841 | 0.981829981 |
| TTPAL | 0.9957032 | 1.064748201 |
| EPPIN | 0.995614348 | 0.933881466 |
| C19orf25 | 0.995598807 | 1.024323398 |
| MAPKAPK2 | 0.995560144 | 1.057105775 |
| ORC1 | 0.995556896 | 0.990065091 |
| TMEM184C | 0.995552821 | 0.976103686 |
| FRMPD1 | 0.995538998 | 1.049331963 |
| SUGP2 | 0.995493553 | 1.060294621 |
| MRGBP | 0.995485941 | 0.983898595 |
| RNF114 | 0.995484653 | 0.955223881 |
| KCTD9 | 0.995479009 | 0.862281603 |
| DKKL1 | 0.995434487 | 0.9181855 |
| USP19 | 0.9954325 | 0.953422438 |

| | | |
|---|---|---|
| GHRL | 0.995426449 | 1.00890716 |
| ORMDL2 | 0.995382782 | 0.986634265 |
| RHBG | 0.995330874 | 0.914135277 |
| EXOC2 | 0.995320425 | 0.999657417 |
| SLC4A9 | 0.995290535 | 1.00908501 |
| UBTD1 | 0.995258034 | 0.979445015 |
| SCGB2A1 | 0.995227313 | 1.06763872 |
| AAAS | 0.995214038 | 1.049674546 |
| PARP9 | 0.995205867 | 0.804374241 |
| OR14C36 | 0.995184406 | 0.97327852 |
| TRAPPC2L | 0.995151921 | 0.924665857 |
| FRG2 | 0.995132979 | 0.924974306 |
| PAQR9 | 0.995131846 | 0.998287085 |
| IL36B | 0.99511601 | 0.885788449 |
| RASL11B | 0.995060178 | 0.955852572 |
| USP6NL | 0.995047344 | 0.932853717 |
| GLTPD2 | 0.995046211 | 1.020212402 |
| BAZ2A | 0.995034295 | 1.048304214 |
| MCOLN3 | 0.994976637 | 0.842959117 |
| COLEC11 | 0.994963068 | 0.940733128 |
| ZNF780A | 0.994940418 | 1.072093965 |
| SLX4IP | 0.994909191 | 1.001620089 |
| TBX2 | 0.994842614 | 1.073977936 |
| SLC26A3 | 0.994841798 | 0.96435804 |
| CNPY4 | 0.994827638 | 1.001712915 |
| DENND1B | 0.994754537 | 0.943701904 |
| CXorf58 | 0.994714396 | 0.993519644 |
| CHPF2 | 0.994710219 | 0.599432969 |
| GTSF1 | 0.994668244 | 1.128392062 |
| LAMP1 | 0.994600909 | 0.87084618 |
| RHOQ | 0.99459902 | 0.794938352 |
| ZMYM1 | 0.9945638 | 1.0222679 |
| TCF7 | 0.994543699 | 1.025957171 |
| PAH | 0.994473065 | 1.004542505 |
| FAM81B | 0.994464789 | 1.085054678 |
| NUAK2 | 0.994439972 | 1.042180402 |
| PLOD2 | 0.994439972 | 1.042180402 |
| REXO1 | 0.994432974 | 0.945489942 |
| ATP2B4 | 0.994372549 | 0.789746918 |
| STXBP3 | 0.994368742 | 0.978759849 |
| PPP2R1B | 0.99436835 | 1.020765737 |
| OR10H4 | 0.99435089 | 0.908530319 |
| WBP2NL | 0.99433968 | 1.045767517 |
| CHSY3 | 0.994339455 | 1.058403634 |

| | | |
|---|---|---|
| MRPL16 | 0.994333174 | 0.989722508 |
| ARHGAP19 | 0.994293936 | 0.863309353 |
| STRA8 | 0.994286906 | 1.060348319 |
| NCOR2 | 0.994275139 | 0.942081814 |
| CD200R1 | 0.994259093 | 0.991899554 |
| HNRNPAB | 0.994238975 | 0.941418294 |
| DDX54 | 0.994227153 | 0.92345079 |
| CIC | 0.994157645 | 0.854059609 |
| TMEM183A | 0.994113149 | 0.952612394 |
| ELAVL2 | 0.994098335 | 0.904076739 |
| CNDP2 | 0.994091827 | 1.031797534 |
| ZNF302 | 0.994090496 | 1.038369305 |
| OR10G8 | 0.994062993 | 0.915039397 |
| MYBPC2 | 0.993971241 | 0.961973279 |
| RSPH6A | 0.993939343 | 0.972935937 |
| RHPN2 | 0.993899516 | 0.863651936 |
| SDHD | 0.993865849 | 0.892277742 |
| NES | 0.993858466 | 1.019869818 |
| VWA9 | 0.993858143 | 1.042122317 |
| TMEM26 | 0.993844032 | 0.90927501 |
| WNT2 | 0.993822399 | 1.069886948 |
| CARD11 | 0.993807203 | 1.023491292 |
| CBLN3 | 0.993775161 | 0.840826245 |
| GUF1 | 0.993767775 | 0.807614419 |
| A26C1A | 0.993674871 | 0.940390545 |
| OR6B3 | 0.993674871 | 0.940390545 |
| RBMXL2 | 0.993616104 | 0.88420692 |
| BLOC1S2 | 0.993604958 | 1.026378897 |
| OSTF1 | 0.993570025 | 0.915755367 |
| ZNF585A | 0.99351588 | 0.94210346 |
| ITIH6 | 0.993488649 | 0.953066119 |
| CAMK2N1 | 0.993456044 | 0.919532771 |
| TFAP2B | 0.99343916 | 1.073714054 |
| NUP62 | 0.993380554 | 1.048669695 |
| IFNA2 | 0.993343851 | 0.988659376 |
| NT5DC1 | 0.993323526 | 1.036452005 |
| RIC3 | 0.993307229 | 0.965741692 |
| MTA2 | 0.993306919 | 0.776832726 |
| ALG11 | 0.99330264 | 1.003240178 |
| SLC45A4 | 0.99330264 | 1.003240178 |
| COL23A1 | 0.993291371 | 1.040767386 |
| SLC35G5 | 0.993262418 | 0.84857828 |
| NOXO1 | 0.993247103 | 1.062692703 |
| KIAA0368 | 0.993205781 | 0.895512162 |

| | | |
|---|---|---|
| RFTN1 | 0.993159651 | 0.949372215 |
| MAP3K9 | 0.993069784 | 0.9467878 |
| DMAP1 | 0.993042642 | 0.993114621 |
| DNAJC15 | 0.993042642 | 0.993114621 |
| ERMP1 | 0.993039585 | 0.897225077 |
| RNPEP | 0.993003925 | 1.000648929 |
| LCTL | 0.992958977 | 0.925373134 |
| REEP4 | 0.99295151 | 1.05346294 |
| RANBP2 | 0.992883097 | 0.939246659 |
| ADCY10 | 0.992871467 | 0.941596366 |
| OR4F4 | 0.992865551 | 0.95991778 |
| NBEA | 0.992857939 | 1.020897568 |
| RFFL | 0.992842983 | 1.033095393 |
| IBA57 | 0.992819574 | 1.06399352 |
| ZNF425 | 0.992815703 | 1.016200891 |
| STAG3L1 | 0.992804519 | 1.053785543 |
| ABCB4 | 0.992793589 | 1.086956522 |
| MYPOP | 0.992780233 | 0.831791709 |
| SPICE1 | 0.99277923 | 0.856217092 |
| RASGRF1 | 0.992772757 | 1.014731072 |
| PPAN-P2RY11 | 0.992757624 | 0.903734156 |
| PAPL | 0.992749532 | 0.918590522 |
| OR7D4 | 0.992712767 | 1.022610483 |
| DUSP13 | 0.992705092 | 0.974042829 |
| GLIS2 | 0.992593544 | 0.861129137 |
| SHFM1 | 0.992589263 | 0.952628164 |
| FKBP15 | 0.992561053 | 0.974888619 |
| MIB2 | 0.992459275 | 0.839714471 |
| DOK4 | 0.992455998 | 1.112024666 |
| AMELX | 0.992437693 | 1.015073655 |
| GNG7 | 0.992410371 | 1.036998972 |
| GPRC5A | 0.992403707 | 1.038935756 |
| PXMP4 | 0.992393697 | 1.011990408 |
| NSUN3 | 0.992341144 | 1.029161604 |
| YTHDF2 | 0.992341144 | 0.904414743 |
| S100Z | 0.992310282 | 0.875984926 |
| SLC19A3 | 0.992266638 | 1.071382219 |
| ATP6V0B | 0.992193111 | 0.927714971 |
| PHGDH | 0.992165706 | 1.068853787 |
| ZNF671 | 0.992131169 | 1.021061158 |
| TMEM50B | 0.991985276 | 0.984241178 |
| ITGB3BP | 0.991874993 | 1 |
| LDLRAD4 | 0.991843956 | 0.974991435 |
| MSLNL_pseudogene | 0.991809991 | 1.00891049 |

| | | |
|---|---|---|
| ACRC | 0.991799869 | 0.892086331 |
| SEC61B | 0.991781927 | 0.993833505 |
| SPATA9 | 0.991736436 | 1.070572114 |
| KRT7 | 0.991687637 | 0.987667009 |
| PLGRKT | 0.991624113 | 1.017471737 |
| UBE2G2 | 0.991597268 | 1.034393251 |
| SLCO1A2 | 0.991591458 | 1.000810045 |
| MAGED1 | 0.99158352 | 0.980532122 |
| DNHD1 | 0.99153629 | 0.832320778 |
| OSCAR | 0.991525248 | 0.898595409 |
| CKAP2 | 0.991516277 | 0.920520726 |
| PKD1 | 0.99143007 | 1.083062946 |
| TBL2 | 0.99141646 | 1.081877355 |
| OR6Y1 | 0.991415916 | 0.903391572 |
| NEDD1 | 0.991409398 | 0.948967193 |
| MYL9 | 0.991386782 | 1.013018157 |
| TXNL4A | 0.991296953 | 0.93770279 |
| PCGF2 | 0.991271268 | 1.007787151 |
| MIA | 0.9911934 | 1.055498458 |
| MYH1 | 0.991165565 | 1.040233615 |
| SDCBP2 | 0.991147411 | 0.877012676 |
| BLVRB | 0.991099789 | 0.862426995 |
| PFDN1 | 0.991086864 | 0.932511356 |
| ZNF491 | 0.99105063 | 1.002398082 |
| ATP6V1E2 | 0.991043788 | 0.936622131 |
| SAFB2 | 0.991040073 | 0.903734156 |
| BTD | 0.991022863 | 0.772377481 |
| ZC3H15 | 0.99099994 | 1.038882139 |
| ATPIF1 | 0.990987785 | 0.911613566 |
| TMEM207 | 0.990986339 | 1.068043742 |
| RP6-166C19.9 | 0.990943699 | 0.952380952 |
| ACTR6 | 0.990903192 | 0.993148338 |
| HMG1L1_pseudogene | 0.990843384 | 0.957177115 |
| GKAP1 | 0.990834079 | 0.693398137 |
| AKR1B1 | 0.990792806 | 0.733290071 |
| LRP1 | 0.99077313 | 0.959766385 |
| WDR60 | 0.990766583 | 1.089104901 |
| LRFN5 | 0.990734595 | 0.940047962 |
| ASTN2 | 0.990730246 | 1.046591298 |
| KCNG1 | 0.990725054 | 0.975989617 |
| OR51F1 | 0.990721365 | 1.024665982 |
| ARRDC1 | 0.990702335 | 0.980815348 |
| KLRC1 | 0.99069726 | 0.969852689 |
| CFHR4 | 0.990678355 | 0.982989064 |

| | | |
|---|---|---|
| LSP1 | 0.990677641 | 1.032545392 |
| TXLNGY | 0.990672612 | 1.021582734 |
| POLR3A | 0.990652527 | 1.078520441 |
| C19orf48 | 0.990645702 | 0.80599433 |
| XX-FW88277B6.1 | 0.990645109 | 1.06235012 |
| HECW2 | 0.990633428 | 1.00843608 |
| HYAL4 | 0.990627721 | 0.922640745 |
| DKK3 | 0.99062182 | 0.922918808 |
| ZNF366 | 0.990615476 | 0.911956149 |
| FBL | 0.990612622 | 0.816375471 |
| SLC5A1 | 0.990611448 | 0.93835172 |
| SH3BP2 | 0.990586297 | 1.037341555 |
| LRIT2 | 0.990503477 | 1.064398542 |
| SLC9B2 | 0.990503477 | 1.020656136 |
| WLS | 0.990503477 | 1.04981774 |
| PLIN2 | 0.990472117 | 0.86193902 |
| TUBGCP6 | 0.990467088 | 0.998287085 |
| IFNA4 | 0.990459566 | 0.987324426 |
| GDF9 | 0.990444692 | 0.974888619 |
| STAC2 | 0.99043152 | 0.880781089 |
| LRP4 | 0.990413803 | 1.05789654 |
| CCNF | 0.990394032 | 1.019467878 |
| SNX31 | 0.990331748 | 1.010962658 |
| SLC26A8 | 0.990318004 | 0.92709599 |
| BRF2 | 0.990316483 | 1.051914341 |
| CHPF | 0.990313026 | 0.965606749 |
| NALCN | 0.990279356 | 1.068137573 |
| UPK1B | 0.990277099 | 1.012555691 |
| ADK | 0.99027277 | 0.981829981 |
| CAPZB | 0.990209729 | 1.034600891 |
| CRCP | 0.990198492 | 0.920178143 |
| C11orf45 | 0.990187954 | 0.908464966 |
| CNGA1 | 0.990090095 | 1.062946139 |
| IQCF1 | 0.99006692 | 0.918995545 |
| PPM1L | 0.990066715 | 0.976638546 |
| SAMSN1 | 0.990051327 | 0.973621103 |
| LMAN2 | 0.990029125 | 0.9109284 |
| CHAC2 | 0.990010729 | 0.981500514 |
| GPR85 | 0.989962823 | 0.922777417 |
| CASZ1 | 0.989950726 | 0.829485622 |
| C17orf58 | 0.989946803 | 1.029566626 |
| PAGE5 | 0.989939722 | 0.914945322 |
| PSD4 | 0.989908955 | 1.156743621 |
| PDXK | 0.98989894 | 1.057754705 |

| | | |
|---|---|---|
| OLIG2 | 0.989892107 | 0.955223881 |
| NRIP2 | 0.989846088 | 0.868916288 |
| PRKAR1B | 0.989825919 | 0.987670344 |
| MSMP | 0.989811186 | 0.980130182 |
| NRAP | 0.989797865 | 0.898595409 |
| FBXO41 | 0.989782206 | 1.122647631 |
| PTPRB | 0.989775291 | 0.901362751 |
| BST2 | 0.9897415 | 0.966084275 |
| UBQLN1 | 0.989728983 | 1.036046983 |
| NCAPD3 | 0.989717929 | 1.021466181 |
| GABRB1 | 0.989709432 | 0.933809215 |
| SOCS2 | 0.989709432 | 0.933809215 |
| FAM3C | 0.989708831 | 0.944158959 |
| PEX12 | 0.98964721 | 0.840698869 |
| CDK10 | 0.989643738 | 0.847501622 |
| BRI3BP | 0.989613953 | 0.948955122 |
| THRA | 0.989566331 | 0.917375456 |
| TNFSF9 | 0.989539645 | 0.905104488 |
| ASB2 | 0.989518195 | 0.912394549 |
| NAT8B | 0.989517377 | 0.929931146 |
| HMGN4 | 0.989500771 | 0.971648441 |
| AMER2 | 0.989499905 | 1.027946537 |
| PRICKLE4 | 0.989489547 | 0.928617781 |
| PAFAH1B3 | 0.989485489 | 1.013365735 |
| C2orf44 | 0.989470665 | 0.998784933 |
| CLN8 | 0.989469869 | 1.069886948 |
| TXK | 0.98945141 | 0.871607938 |
| RRP7A | 0.989383965 | 1.023896314 |
| CD3G | 0.98935116 | 0.99473471 |
| KIR2DL5A | 0.98935116 | 0.99473471 |
| HAO2 | 0.989323754 | 0.825840421 |
| PHOSPHO2 | 0.989301493 | 1.049007695 |
| CREBBP | 0.989291101 | 0.992709599 |
| IRGQ | 0.989251266 | 0.82381531 |
| AADAT | 0.989243236 | 0.939649578 |
| CASKIN1 | 0.989235256 | 1.065433368 |
| IVNS1ABP | 0.989235256 | 1.065433368 |
| PIGW | 0.98923522 | 0.948967193 |
| GGACT | 0.989195066 | 1.043508051 |
| RUNDC1 | 0.989177803 | 0.886605002 |
| GPR156 | 0.989134816 | 1.020765737 |
| ASB1 | 0.989115058 | 1.036988968 |
| NT5C2 | 0.989107402 | 0.869565217 |
| TFAP2A | 0.989103947 | 0.778047793 |

| | | |
|---|---|---|
| KIF18B | 0.989086776 | 0.988694758 |
| SNX8 | 0.989040107 | 0.966769442 |
| SLC35F4 | 0.989034081 | 0.918122645 |
| PON3 | 0.989031663 | 0.817739976 |
| ZBTB47 | 0.989030695 | 1.011745646 |
| DCAF12L2 | 0.98902178 | 0.928311057 |
| MCM5 | 0.989009147 | 0.997164844 |
| MXRA7 | 0.989007348 | 0.974648852 |
| F2 | 0.988983753 | 0.966904607 |
| BAP1 | 0.988965559 | 0.983127839 |
| CLNS1A | 0.988965559 | 0.983127839 |
| COX6B1 | 0.988940531 | 0.953422438 |
| TNMD | 0.98892058 | 0.89311408 |
| RGS8 | 0.988912198 | 0.998287085 |
| FNBP1 | 0.988906489 | 0.930798219 |
| GPR114 | 0.988898137 | 1.048020766 |
| GLIPR2 | 0.988894074 | 1.054813292 |
| UBE2Q2 | 0.988838781 | 1.112913692 |
| AP5S1 | 0.988835489 | 0.860226105 |
| CISH | 0.988831866 | 0.896820247 |
| FAM220A | 0.988831049 | 0.984241178 |
| ZFAND1 | 0.98882724 | 1.059943297 |
| PLP2 | 0.988800555 | 0.992120589 |
| IGJ | 0.988758281 | 0.913669065 |
| MSLN | 0.988691994 | 0.948269955 |
| PCDH7 | 0.988663313 | 0.937307297 |
| ECM2 | 0.988637894 | 0.869818431 |
| MACC1 | 0.988632729 | 0.945186708 |
| DHX30 | 0.988624906 | 0.859182349 |
| TTC19 | 0.988555118 | 0.94127177 |
| VOPP1 | 0.988516328 | 0.99554642 |
| PIAS3 | 0.988477794 | 1.070084361 |
| MEF2D | 0.98845446 | 0.978938842 |
| KIF3B | 0.98845178 | 1.118754056 |
| CFL1 | 0.988378542 | 1.016101405 |
| GDF7 | 0.988371111 | 0.838643371 |
| ZMYND19 | 0.988371111 | 0.978417266 |
| CCDC15 | 0.988362054 | 1.043742406 |
| NPDC1 | 0.988344061 | 0.986296677 |
| CST1 | 0.988339575 | 1.088386434 |
| DDX52 | 0.988320392 | 0.967553537 |
| MAGEA1 | 0.988319866 | 0.892086331 |
| GALNT10 | 0.988292732 | 1.032446463 |
| WDR70 | 0.988280401 | 0.96437136 |

| | | |
|---|---|---|
| FOXA3 | 0.988280054 | 1.064892927 |
| MRPS30 | 0.988274044 | 1.081116158 |
| SLC44A3 | 0.988268273 | 1 |
| CD99L2 | 0.98826563 | 1.009934909 |
| ZNF211 | 0.98825974 | 0.99109284 |
| FAM57A | 0.988240801 | 0.934566632 |
| PGM3 | 0.988239441 | 0.810044552 |
| HS3ST3A1 | 0.988215907 | 1.052247874 |
| ZNF704 | 0.988152402 | 0.939362795 |
| VSX2 | 0.988134776 | 0.901678657 |
| ADAMTS7 | 0.988090052 | 1.011031798 |
| PRSS21 | 0.988086759 | 1.027255029 |
| KARS | 0.988035782 | 0.952207371 |
| PELO | 0.987991552 | 0.922233642 |
| KRTAP25-1 | 0.987986659 | 1.058924289 |
| HPSE2 | 0.987916565 | 0.945871874 |
| MRPL32 | 0.987878728 | 0.989617132 |
| RANBP9 | 0.987845248 | 0.969510106 |
| C6orf183 | 0.987828075 | 1.022953066 |
| GPANK1 | 0.9877921 | 1.011990408 |
| DCTN1 | 0.987707507 | 1.043508051 |
| LUZP1 | 0.98769627 | 0.997944502 |
| POLR3GL | 0.987674515 | 1.073714054 |
| DCTN6 | 0.987654268 | 1.013703323 |
| ACTG1 | 0.987650734 | 1.040424803 |
| EDDM3A | 0.987647335 | 0.914696814 |
| ENOSF1 | 0.987634883 | 1.098825435 |
| PLEKHG7 | 0.987631296 | 0.869133265 |
| PLEKHS1 | 0.987624726 | 0.922576225 |
| NOP58 | 0.987595592 | 1.010620075 |
| SPHKAP | 0.987594742 | 1.004860267 |
| SLAMF8 | 0.98754036 | 0.847207948 |
| NIPAL3 | 0.987520421 | 0.883758607 |
| DNAJB3 | 0.987516844 | 0.935251799 |
| ATP5G2 | 0.987462913 | 1.027903958 |
| TUBA8 | 0.987462349 | 0.961117861 |
| OR2K2 | 0.987434516 | 0.966769442 |
| ZNF12 | 0.987420063 | 1.028091812 |
| USB1 | 0.987415484 | 0.94653706 |
| SLC6A7 | 0.9874029 | 0.898118105 |
| EFCC1 | 0.987311719 | 0.969623329 |
| TET2 | 0.987299952 | 0.733495342 |
| NDNL2 | 0.987298729 | 0.929931146 |
| TNKS | 0.98729802 | 1.103828683 |

| | | |
|---|---|---|
| C16orf70 | 0.987294144 | 0.98217902 |
| GTPBP2 | 0.987265873 | 0.902794654 |
| PIK3C2A | 0.987261715 | 1.038935756 |
| PIGT | 0.987218344 | 1 |
| LCE4A | 0.987174802 | 0.989037342 |
| TMEM179 | 0.987133072 | 1.003240178 |
| BCS1L | 0.987128691 | 0.950992305 |
| C12orf43 | 0.987107398 | 0.911300122 |
| LTF | 0.987100691 | 1.098637249 |
| EML1 | 0.987047436 | 1.017471737 |
| TCHHL1 | 0.987030989 | 0.860226105 |
| PAFAH2 | 0.986981405 | 0.952628164 |
| MGST1 | 0.986965928 | 0.936404932 |
| TRAPPC8 | 0.986957723 | 0.960945529 |
| DYNC1LI1 | 0.986949793 | 0.9201817 |
| C12orf66 | 0.986925032 | 0.94491697 |
| ZKSCAN1 | 0.986925032 | 0.94491697 |
| FHL3 | 0.98691042 | 0.984583762 |
| ST13 | 0.986901634 | 1.093445814 |
| PEX11A | 0.986847802 | 0.981500514 |
| CARD16 | 0.986844495 | 0.95544755 |
| EMILIN2 | 0.986811079 | 0.915755367 |
| ZNF333 | 0.98678822 | 0.901336074 |
| ARAP1 | 0.986765928 | 0.874614594 |
| RASAL3 | 0.986764025 | 0.951695786 |
| TMEM199 | 0.986708485 | 0.951397327 |
| UAP1L1 | 0.986707252 | 0.983213429 |
| RALYL | 0.986676079 | 1.041452552 |
| THY1 | 0.986653334 | 0.852689277 |
| HOXB9 | 0.986523114 | 0.881123672 |
| CARD9 | 0.986515683 | 0.973963686 |
| FCN2 | 0.986496155 | 0.896882494 |
| ATOH7 | 0.986491397 | 0.989722508 |
| LOC643923 | 0.986483105 | 0.904761905 |
| ZHX1 | 0.986454948 | 1.050616483 |
| PRR35 | 0.986451615 | 0.878040425 |
| TRIM21 | 0.986432638 | 1.034393251 |
| CCM2L | 0.986425879 | 0.893799246 |
| POT1 | 0.986407836 | 0.993519644 |
| TCP11 | 0.986372459 | 1.026036314 |
| TCF12 | 0.986319913 | 0.698663426 |
| SEMA7A | 0.98631913 | 0.863994519 |
| SNCAIP | 0.986293221 | 0.939246659 |
| HPRT1 | 0.986288534 | 0.951802349 |

| | | |
|---|---|---|
| PLEKHF2 | 0.986278351 | 0.996231586 |
| UXT | 0.986258325 | 1.011990408 |
| SMPDL3A | 0.986222974 | 0.937221547 |
| PLEKHA3 | 0.986213604 | 0.717711545 |
| ZNF34 | 0.986190632 | 0.900308325 |
| CUX2 | 0.986109664 | 0.905104488 |
| ZNF587 | 0.986087626 | 0.895504253 |
| OSER1 | 0.986029647 | 0.909900651 |
| PCDHGA6 | 0.986015597 | 0.86836776 |
| MOSPD2 | 0.986011931 | 0.925659472 |
| GUSB | 0.98593911 | 0.99156392 |
| ABCA13 | 0.985906922 | 0.975340688 |
| EPCAM | 0.98586298 | 0.876873228 |
| WISP1 | 0.98584279 | 0.95101062 |
| ZSCAN22 | 0.985812244 | 1.050627785 |
| METTL9 | 0.985810715 | 1.038072094 |
| LRP5L | 0.985795077 | 0.925070879 |
| CHCHD5 | 0.98570156 | 0.855087359 |
| SPANXN1 | 0.985690497 | 1.071688943 |
| CSRP2BP | 0.985688103 | 0.968482357 |
| OR2A1 | 0.985672471 | 1.089756766 |
| NELL2 | 0.985650496 | 1.023638232 |
| NUCKS1 | 0.985638631 | 0.953926022 |
| HDGF | 0.985604255 | 0.994329688 |
| SEMA5B | 0.985566402 | 0.938677629 |
| LRR1 | 0.985559765 | 1.10577547 |
| TNRC6A | 0.985546438 | 1.017415958 |
| FAM177B | 0.985534458 | 0.993833505 |
| KCNC3 | 0.985525272 | 1.089552239 |
| TTC3 | 0.985525272 | 1.089552239 |
| MOGAT1 | 0.985522061 | 0.997404283 |
| OR8G2 | 0.985491913 | 0.943473792 |
| APOL5 | 0.98546808 | 0.990750257 |
| LAIR2 | 0.985460558 | 1.006509078 |
| YARS2 | 0.985443335 | 0.940056703 |
| DDX12P | 0.985384087 | 0.856586632 |
| RIPK1 | 0.985376184 | 1.024659312 |
| DPY19L4 | 0.985315679 | 0.961117861 |
| C11orf83 | 0.985313302 | 1.047619048 |
| SRPK2 | 0.985278055 | 1.068137573 |
| OR2T35 | 0.985268536 | 0.973621103 |
| SLC35F3 | 0.985266271 | 0.981500514 |
| VPS52 | 0.985253675 | 1.028776978 |
| COL4A1 | 0.985249688 | 1.0445358 |

| | | |
|---|---|---|
| LIPJ | 0.985219194 | 0.883841661 |
| LINC00478 | 0.985174982 | 0.871202916 |
| FREM2 | 0.98516949 | 1.136005481 |
| CEMP1 | 0.985147288 | 0.865022268 |
| RHBDD3 | 0.985089301 | 0.783313082 |
| DLGAP2 | 0.985058436 | 1.003768414 |
| SURF1 | 0.985031076 | 0.953017416 |
| FAM134C | 0.985020582 | 1.087079789 |
| HIST1H2BA | 0.984991196 | 0.984926345 |
| HES1 | 0.984981691 | 1.1064244 |
| PSCA | 0.984918429 | 0.926687222 |
| DMRTC1 | 0.984915846 | 0.895169579 |
| CUTA | 0.984874843 | 0.875658161 |
| AP3S1 | 0.984856536 | 0.501199041 |
| IGLL1 | 0.984813175 | 0.758136348 |
| GSR | 0.984801239 | 1.041531473 |
| SLC6A15 | 0.984752586 | 1.025308241 |
| DEFB123 | 0.984722329 | 1.004110997 |
| LRRC24 | 0.984671718 | 1.056183625 |
| SNX29P1 | 0.984668382 | 0.942891859 |
| LMX1A | 0.984616094 | 0.916937054 |
| YTHDC1 | 0.984610005 | 1.001620089 |
| CD1A | 0.984606699 | 1.053100377 |
| NKAPL | 0.984582232 | 0.990065091 |
| NLRC5 | 0.984510638 | 0.824789098 |
| CNTNAP1 | 0.98442461 | 0.927968851 |
| WDFY4 | 0.984424283 | 0.952380952 |
| POFUT1 | 0.984417036 | 0.987670344 |
| TSPY2 | 0.984414768 | 0.936622131 |
| PTPN12 | 0.984410368 | 1.047371836 |
| RNPS1 | 0.984409331 | 1.043850634 |
| XKRX | 0.984405938 | 0.911705144 |
| BOLA3 | 0.984379347 | 1.066828676 |
| CPE | 0.984355788 | 1.031148605 |
| JMJD6 | 0.984351228 | 0.922235723 |
| CSMD3 | 0.984337832 | 0.82631038 |
| ZNF625 | 0.984287131 | 0.862624186 |
| METRNL | 0.984217154 | 0.966383151 |
| TAS2R40 | 0.984210648 | 0.95101062 |
| TBL1Y | 0.984169779 | 0.903734156 |
| C3orf43 | 0.984144564 | 0.96435804 |
| CITED1 | 0.984098616 | 0.867962738 |
| SLC2A13 | 0.984071618 | 0.819765087 |
| DLGAP1 | 0.984065476 | 1.020554985 |

| | | |
|---|---|---|
| TRIM29 | 0.984038596 | 0.890330954 |
| NLRP4 | 0.984021595 | 0.937221547 |
| ROMO1 | 0.984018852 | 1.041109969 |
| TNFSF4 | 0.9840039 | 1.025351148 |
| TMEM222 | 0.983982727 | 1.054678007 |
| LRRC34 | 0.983956235 | 0.978074683 |
| SLC13A1 | 0.983923262 | 1.080467229 |
| SFXN1 | 0.983901281 | 1.020765737 |
| GRAMD1A | 0.983884871 | 1.004455245 |
| ZNF791 | 0.983783653 | 0.895504253 |
| C5orf51 | 0.983735169 | 1.020897568 |
| OR1F1 | 0.983733266 | 1.072970195 |
| S100A10 | 0.983728826 | 0.908872902 |
| PDP1 | 0.983705434 | 1.075275795 |
| DDB1 | 0.983686969 | 0.975293641 |
| CAPRIN2 | 0.983659155 | 1.080194411 |
| ZNF496 | 0.983644233 | 1.002055498 |
| SEMA5A | 0.98363354 | 0.994176088 |
| PMFBP1 | 0.983630141 | 0.94210346 |
| CALHM1 | 0.983626335 | 0.890030832 |
| SLC38A6 | 0.983613869 | 0.97326853 |
| POP5 | 0.983583563 | 1.006851662 |
| LRRC74 | 0.983583458 | 0.960712839 |
| RNF219 | 0.983575611 | 1.042829332 |
| ATP6V1G3 | 0.983571131 | 0.843256379 |
| TMEFF2 | 0.983503909 | 0.907502569 |
| ZNF624 | 0.983488413 | 0.988009592 |
| FTHL17 | 0.983476406 | 0.980130182 |
| CLEC4M | 0.983457279 | 0.992304577 |
| SELE | 0.983452211 | 1.008564577 |
| UBA2 | 0.983449903 | 0.820246593 |
| FGD6 | 0.983440022 | 1.000685166 |
| ZNF521 | 0.983429329 | 1.036998972 |
| PDDC1 | 0.983425493 | 0.979748886 |
| GAS2L2 | 0.983393655 | 1.072093965 |
| MCUR1 | 0.983325039 | 0.942081814 |
| ABHD16B | 0.983321719 | 0.929770469 |
| ACTR1B | 0.983281167 | 0.98663926 |
| CACTIN | 0.983273538 | 0.998784933 |
| IL23A | 0.983248577 | 0.940056703 |
| ZDHHC11 | 0.983244965 | 1.043508051 |
| ARID4A | 0.983227488 | 1.055488052 |
| ENHO | 0.983183578 | 1.13527744 |
| BCMO1 | 0.983169009 | 0.961117861 |

| | | |
|---|---|---|
| RCN3 | 0.983162184 | 0.996231586 |
| GUCD1 | 0.983157608 | 1.177800617 |
| TDRP | 0.983154797 | 1.09963548 |
| HABP4 | 0.983132687 | 0.871531346 |
| PACRGL | 0.983098966 | 0.843661401 |
| SNX2 | 0.98309377 | 0.95909275 |
| CPNE4 | 0.983085022 | 0.920520726 |
| TOR1AIP2 | 0.983079222 | 1.094210346 |
| KRT18P55 | 0.98307741 | 1.05789654 |
| CENPC | 0.983030061 | 0.997944502 |
| PRR30 | 0.983021588 | 1.049412718 |
| MAGEE1 | 0.983014384 | 0.990065091 |
| PURG | 0.982993406 | 0.945871874 |
| SLC4A1AP | 0.982963867 | 1.043478261 |
| PRPH | 0.982831424 | 0.874957177 |
| ALG8 | 0.982819304 | 0.917375456 |
| C3 | 0.982786341 | 0.887632751 |
| CCDC65 | 0.98276867 | 0.881733495 |
| RAB37 | 0.98276713 | 1.001370332 |
| PLEKHB2 | 0.982722545 | 1.070914697 |
| LGR4 | 0.982712452 | 0.951330305 |
| AMOTL2 | 0.982655849 | 0.982528263 |
| STX2 | 0.982651862 | 1.022063595 |
| CCNT2 | 0.982637949 | 0.978585334 |
| DIRAS3 | 0.982622802 | 0.935107073 |
| ZFYVE26 | 0.982599846 | 1.044193217 |
| ACVR1C | 0.982583945 | 1.049318624 |
| TP53I11 | 0.982577192 | 0.922576225 |
| PRB3 | 0.982559974 | 0.971565605 |
| ANGPTL5 | 0.982558252 | 0.943131209 |
| BICD1 | 0.982556349 | 0.914696814 |
| RBM10 | 0.982548329 | 1.077423775 |
| SP100 | 0.982506056 | 1.004796163 |
| VIT | 0.982506056 | 1.004796163 |
| HID1 | 0.982502793 | 0.976361768 |
| MYO3A | 0.982458237 | 0.946138871 |
| THAP4 | 0.982431068 | 1.001712915 |
| ELMO3 | 0.982404607 | 0.965399109 |
| RUVBL1 | 0.982363648 | 0.929915639 |
| TNP2 | 0.982317341 | 0.933881466 |
| PICK1 | 0.98229746 | 0.957170668 |
| ACTRT1 | 0.982291651 | 0.974991435 |
| OR13A1 | 0.982291651 | 0.974991435 |
| CEACAM8 | 0.982264158 | 0.959902795 |

| | | |
|---|---|---|
| ZIM3 | 0.98223601 | 0.881808839 |
| C1orf115 | 0.982204304 | 1.10490077 |
| MKNK2 | 0.982140387 | 0.968202466 |
| DENND1B | 0.982104869 | 0.989469421 |
| FCN1 | 0.982060934 | 0.806097979 |
| NPR3 | 0.982003447 | 1.031591738 |
| ZFP42 | 0.981983545 | 1.032202809 |
| CACNA1A | 0.981976729 | 1.049967554 |
| ANK1 | 0.981951783 | 0.954779034 |
| PPP1R3B | 0.981941725 | 0.995889003 |
| DYNC1I2 | 0.981941412 | 0.935756003 |
| PCDHB8 | 0.981936967 | 1.016443988 |
| RBM14 | 0.981905114 | 0.918465228 |
| MUC13 | 0.981878382 | 1.06235012 |
| KCNAB2 | 0.981780717 | 0.87605451 |
| TPR | 0.98176171 | 0.961288112 |
| CAPSL | 0.981736925 | 0.891743748 |
| DAZ | 0.981736016 | 0.974042829 |
| DCAF4L2 | 0.981693425 | 0.956257594 |
| HAL | 0.981610647 | 0.876468206 |
| SHBG | 0.981589397 | 1.091127098 |
| OR9K2 | 0.98158473 | 1.050017129 |
| FAM22G | 0.981561849 | 0.885577252 |
| MRPL30 | 0.98155069 | 0.98582422 |
| MFI2 | 0.981478706 | 0.903048989 |
| LIN28B | 0.981451204 | 1.082624544 |
| PHLDA1 | 0.981419667 | 1.077081192 |
| OR5I1 | 0.981398734 | 0.920520726 |
| C9orf78 | 0.981395925 | 0.994861254 |
| MYOD1 | 0.981395017 | 0.893479141 |
| CLCA1 | 0.98137791 | 0.854639844 |
| RTN2 | 0.981277753 | 0.956662616 |
| HTR5A | 0.981260928 | 0.936404932 |
| SBNO1 | 0.981248805 | 0.901678657 |
| KRT10 | 0.981204311 | 0.873244262 |
| SAMD13 | 0.981198184 | 0.954637505 |
| MED19 | 0.981157177 | 0.876873228 |
| EIF3G | 0.981148619 | 0.778047793 |
| SH3BGRL3 | 0.981137751 | 0.998287085 |
| CYTH1 | 0.981137071 | 0.770469339 |
| MTUS1 | 0.981131453 | 0.996759822 |
| C11orf42 | 0.981100175 | 1.01782098 |
| SLC46A2 | 0.981086568 | 0.84944841 |
| ATG9B | 0.981081069 | 0.949297705 |

| | | |
|---|---|---|
| C11orf71 | 0.981070121 | 0.984204131 |
| FAM214B | 0.981051255 | 0.787941076 |
| PDZD2 | 0.98099641 | 1.036857027 |
| AZI2 | 0.98098775 | 0.927501013 |
| SCN2A | 0.980977325 | 1.083711875 |
| CDC37L1 | 0.980976222 | 1.025351148 |
| NDUFA12 | 0.980964482 | 1.236210253 |
| SUMF1 | 0.98094514 | 1.017471737 |
| TIGD7 | 0.98094514 | 1.017471737 |
| IPO13 | 0.980929418 | 0.83076396 |
| TMEM263 | 0.980919745 | 0.914945322 |
| PIK3IP1 | 0.980890455 | 0.93600648 |
| NAA11 | 0.980797591 | 1.009720535 |
| PITX3 | 0.980797591 | 1.009720535 |
| ZCCHC3 | 0.980782054 | 0.955042527 |
| SETX | 0.980769319 | 0.965573107 |
| IL17F | 0.980759597 | 1.036656389 |
| KCNK1 | 0.980746079 | 1.078520441 |
| TXNDC9 | 0.980741519 | 0.936964714 |
| INTS5 | 0.980730961 | 0.856217092 |
| GIPC2 | 0.980701692 | 1.054128126 |
| PAX8 | 0.980678747 | 0.898339409 |
| WDPCP | 0.980592658 | 0.896314297 |
| STK19 | 0.980551549 | 0.99156392 |
| TMEM45A | 0.980538662 | 0.948967193 |
| PPFIA4 | 0.980512446 | 1.073329007 |
| STARD4 | 0.980500245 | 1.006851662 |
| PANX3 | 0.980495035 | 0.810551559 |
| MMP9 | 0.980478858 | 0.866320571 |
| OR2A12 | 0.980475098 | 0.986296677 |
| JAM2 | 0.980463678 | 1.033211827 |
| SRF | 0.980421092 | 0.989064399 |
| MCF2L | 0.980413387 | 0.899280576 |
| ARGFX | 0.980337674 | 1.000685166 |
| CDK11A | 0.980337673 | 0.850097339 |
| DDX41 | 0.980337673 | 0.850097339 |
| KLHL13 | 0.980285438 | 1.071283921 |
| TDRD3 | 0.980284979 | 1.018156903 |
| ORM2 | 0.98024978 | 1.006075334 |
| ZNF250 | 0.980212492 | 1.069258809 |
| CTPS1 | 0.980201593 | 0.779364049 |
| CLMN | 0.98017968 | 0.981843097 |
| VWC2L | 0.980166721 | 1.007194245 |
| NLRP14 | 0.980165947 | 0.997404283 |

| | | |
|---|---|---|
| EIF2AK4 | 0.980150635 | 1.133679429 |
| LAT2 | 0.980074801 | 1.044147428 |
| PCBD1 | 0.980038428 | 0.926670993 |
| RGS22 | 0.980019836 | 0.989469421 |
| TMEM41A | 0.979987489 | 0.861077359 |
| MORN3 | 0.979940268 | 0.987444309 |
| ERCC6 | 0.979906629 | 0.96495782 |
| LPHN1 | 0.979906629 | 0.96495782 |
| PRKRIR | 0.979898726 | 0.910447761 |
| ATP6V0D1 | 0.979894363 | 0.883192732 |
| MED29 | 0.979876898 | 0.922235723 |
| CYB5R4 | 0.979871142 | 0.943296881 |
| PDZD8 | 0.979848117 | 1.038072094 |
| ZNF335 | 0.97975608 | 0.941075711 |
| KIAA1715 | 0.979711903 | 0.9331963 |
| PDCD1 | 0.979709547 | 0.983898595 |
| PHLDA3 | 0.979666956 | 1.02672148 |
| ZNF564 | 0.979666956 | 0.976019185 |
| MEFV | 0.979662211 | 1.014276444 |
| ZSCAN9 | 0.979627718 | 1.069544365 |
| ETV4 | 0.97956721 | 1.0798183 |
| LYVE1 | 0.979538502 | 0.987849332 |
| TMEM236 | 0.979526633 | 0.861482382 |
| RNMTL1 | 0.979518075 | 0.787768327 |
| TARP | 0.979484359 | 0.919150394 |
| CATSPERG | 0.979467911 | 0.843096951 |
| OR4A5 | 0.979462384 | 1.025351148 |
| TOMM20 | 0.979454454 | 0.987324426 |
| MORC3 | 0.979452515 | 0.954232483 |
| ZNF618 | 0.979434065 | 1.080849606 |
| HOXB8 | 0.97941644 | 1.004796163 |
| NUCB1 | 0.979386626 | 1.047619048 |
| NCAPH2 | 0.979381861 | 1.004860267 |
| DND1 | 0.979348067 | 0.920863309 |
| DEFA4 | 0.979321039 | 1.066018631 |
| PDGFD | 0.979309781 | 1.057211374 |
| DOCK4 | 0.979294149 | 0.900308325 |
| MGEA5 | 0.979290357 | 1.020116807 |
| SOWAHC | 0.979260878 | 0.886998785 |
| OR10C1 | 0.979257222 | 1.023980815 |
| RIN2 | 0.979253416 | 0.811579308 |
| UVRAG | 0.979242132 | 1.05346294 |
| ACOT4 | 0.979238363 | 1.042932361 |
| DEFB106A | 0.979230467 | 1.021871203 |

| | | |
|---|---|---|
| ARHGAP6 | 0.979218346 | 0.841726619 |
| C11orf87 | 0.979130038 | 1.093867763 |
| KLF8 | 0.979114723 | 0.987667009 |
| DESI1 | 0.979082598 | 1.005138746 |
| OCIAD1 | 0.979067357 | 1.007290401 |
| COL5A1 | 0.979066423 | 1.047961631 |
| GUCY1A3 | 0.979049477 | 0.975698663 |
| PRKCD | 0.979035814 | 1.014925373 |
| SFT2D3 | 0.978993799 | 1.015795869 |
| SUSD3 | 0.978988445 | 0.989379925 |
| SULT4A1 | 0.978947975 | 0.933160286 |
| MED30 | 0.978938786 | 1.036998972 |
| YEATS2 | 0.978935625 | 1.045362495 |
| C19orf26 | 0.978870595 | 0.948269955 |
| ARF1 | 0.978850504 | 0.929526124 |
| ARTN | 0.978845357 | 1.021240151 |
| GAST | 0.978841641 | 0.918995545 |
| ZNF827 | 0.978834982 | 1.008564577 |
| PVRL1 | 0.978795834 | 0.915039397 |
| FMNL2 | 0.978791414 | 0.95909275 |
| OR5M1 | 0.978779251 | 0.945186708 |
| ARFIP2 | 0.978722543 | 0.885378696 |
| HOMEZ | 0.97872076 | 1.060348319 |
| GNG3 | 0.978674495 | 0.972250771 |
| TCN1 | 0.978612376 | 0.994518671 |
| ADAM21 | 0.97855908 | 1.031797534 |
| NISCH | 0.978493139 | 1.070084361 |
| CRTC1 | 0.978459538 | 0.889833941 |
| SOX3 | 0.978456588 | 0.884490591 |
| SNRNP40 | 0.97844283 | 1.012150668 |
| SEC62 | 0.97843035 | 1.001620089 |
| TTC27 | 0.97843035 | 1.001620089 |
| VPRBP | 0.978412107 | 0.902706406 |
| OPN1MW | 0.97841024 | 1.015574302 |
| RLIM | 0.978406206 | 0.857235561 |
| CTU2 | 0.978347722 | 0.731414868 |
| NLRC3 | 0.978335041 | 0.989064399 |
| ARHGAP28 | 0.978331501 | 0.942446043 |
| MAGEC1 | 0.978326563 | 1.032887975 |
| FLYWCH1 | 0.978321236 | 1.100850547 |
| APITD1 | 0.97831741 | 0.839328537 |
| MPP3 | 0.978280581 | 0.868267359 |
| OR10G3 | 0.978236623 | 0.909215485 |
| PAPOLG | 0.978003728 | 1.125243348 |

| | | |
|---|---|---|
| APOA1 | 0.977991704 | 0.829080599 |
| BHLHE40 | 0.977979631 | 0.919805589 |
| TNNI1 | 0.977952985 | 0.930113052 |
| HNRNPR | 0.977948454 | 1.025351148 |
| TIPIN | 0.977940305 | 1.040097205 |
| RAB7B | 0.977884341 | 1.082562521 |
| HOXD3 | 0.977842551 | 1.027541515 |
| INVS | 0.977831646 | 0.914354231 |
| TBC1D22A | 0.977814381 | 0.835560956 |
| ELOVL1 | 0.977776232 | 0.954093868 |
| LMO2 | 0.977733369 | 1.041452552 |
| POLE4 | 0.977696827 | 0.923426347 |
| TBC1D8 | 0.977654984 | 0.938335046 |
| SERPINA12 | 0.977649184 | 0.873244262 |
| OGDH | 0.977616937 | 0.911705144 |
| IPO8 | 0.977580042 | 1.065433368 |
| ACTA1 | 0.977548189 | 0.970195272 |
| AMY1C | 0.977524855 | 0.940047962 |
| ITGB7 | 0.977512102 | 0.979343864 |
| GNL1 | 0.977473338 | 0.87975334 |
| HCK | 0.977452574 | 0.737832576 |
| KRTAP13-2 | 0.977451272 | 0.984583762 |
| GZMK | 0.977424913 | 1.022063595 |
| ARMCX5 | 0.977340637 | 0.90562981 |
| RMI1 | 0.977317409 | 1.034426893 |
| SAR1A | 0.977315728 | 0.964763062 |
| HOXC5 | 0.977298561 | 0.855002025 |
| LMBR1 | 0.977287433 | 1.109669046 |
| ZFYVE16 | 0.977199441 | 0.918122645 |
| CERS4 | 0.977136979 | 1.119076549 |
| OR5A1 | 0.977121509 | 1.054813292 |
| MRPL35 | 0.976983133 | 0.969167523 |
| CDR2 | 0.976964148 | 0.807125728 |
| HES3 | 0.976955132 | 0.881808839 |
| DRP2 | 0.976953319 | 0.973963686 |
| MUSTN1 | 0.97694761 | 0.956491949 |
| TMEM206 | 0.976934899 | 1.064398542 |
| RAB11A | 0.976902603 | 0.876468206 |
| CCDC8 | 0.9768714 | 1.080507023 |
| C16orf91 | 0.976812814 | 0.940733128 |
| SLC6A17 | 0.976808329 | 0.997944502 |
| ARL14EP | 0.976779328 | 1.089914945 |
| OR5B21 | 0.976771492 | 0.985268928 |
| OR1K1 | 0.976751284 | 1.064748201 |

| | | |
|---|---|---|
| VEPH1 | 0.9766803 | 0.999189955 |
| PPIL1 | 0.976635218 | 1.037262049 |
| TEKT4 | 0.976617083 | 0.978128797 |
| ARSG | 0.976605138 | 0.815055159 |
| HNRNPK | 0.976569456 | 0.961630695 |
| DIMT1 | 0.976544795 | 1.055807917 |
| HDGFRP3 | 0.976529517 | 0.976103686 |
| AEN | 0.976501653 | 1.043742406 |
| IFNA5 | 0.976477151 | 1.111381126 |
| ARL3 | 0.976420297 | 1.007879411 |
| SERPINB8 | 0.97641554 | 1.109626584 |
| SRGAP1 | 0.976398186 | 1.052415211 |
| KIF1A | 0.976390196 | 0.933981369 |
| CYGB | 0.976324286 | 1.0668037 |
| KIAA1598 | 0.976308654 | 1.049331963 |
| OR7A5 | 0.976298096 | 0.931956258 |
| PSMB2 | 0.976269013 | 0.809863725 |
| CEP89 | 0.976264781 | 0.970028352 |
| RCOR1 | 0.976232761 | 1.001712915 |
| DUSP9 | 0.976232461 | 0.935756003 |
| PTGES2 | 0.976221227 | 0.910895099 |
| PDE6B | 0.976166415 | 0.919400567 |
| GPR1 | 0.976164791 | 0.820895522 |
| RNF126 | 0.976164791 | 0.985074627 |
| ANKLE1 | 0.976142525 | 1.141352774 |
| RWDD3 | 0.976139921 | 0.892086331 |
| TIGD2 | 0.976069601 | 0.83967112 |
| BRD4 | 0.976063204 | 0.83192732 |
| CD83 | 0.976042868 | 0.99554642 |
| PSMB5 | 0.976026982 | 0.919532771 |
| LSM5 | 0.976026919 | 0.933538883 |
| PPP1R14D | 0.976023884 | 0.955806783 |
| CD79A | 0.975997017 | 0.881245944 |
| OR6F1 | 0.975996683 | 0.963953017 |
| SCRN3 | 0.975989992 | 1.049674546 |
| FLII | 0.975983693 | 1.069663832 |
| C4orf22 | 0.975922157 | 1.116241393 |
| ATG12 | 0.975910696 | 1.048602673 |
| ZNF559 | 0.97589901 | 1.046591298 |
| MTMR11 | 0.975857361 | 0.970433374 |
| KIAA0586 | 0.975844168 | 1.114216282 |
| PDLIM5 | 0.975807485 | 1.043508051 |
| TMEM168 | 0.975802091 | 0.873228028 |
| CASC3 | 0.975782565 | 1.026036314 |

| | | |
|---|---|---|
| PLAUR | 0.975758773 | 1.040233615 |
| SYCP2 | 0.975718134 | 0.92428914 |
| NUDCD3 | 0.975686661 | 0.928311057 |
| NOP10 | 0.975645552 | 0.898744431 |
| EPHA8 | 0.975522094 | 0.985723556 |
| ASIC4 | 0.975506452 | 0.898118105 |
| AKR1C1 | 0.975491552 | 1.03504218 |
| SNAPC4 | 0.975481392 | 0.972250771 |
| MEX3C | 0.975438371 | 1.133679429 |
| KLHL9 | 0.975414968 | 0.838643371 |
| HCAR2 | 0.975368561 | 1.007787151 |
| WNK1 | 0.975368561 | 1.007787151 |
| PTCRA | 0.975346958 | 0.951695786 |
| SF3B5 | 0.975331608 | 1.110571081 |
| PLEKHG4 | 0.975325255 | 1.082219938 |
| SEMG2 | 0.975308219 | 1.010620075 |
| FILIP1 | 0.975288599 | 0.939020212 |
| RP6-166C19.5 | 0.975244423 | 0.948612539 |
| CBFA2T2 | 0.975238737 | 0.980532122 |
| SIPA1L2 | 0.975238125 | 0.975676602 |
| PDZRN4 | 0.975191731 | 1.128487995 |
| NMBR | 0.975026096 | 0.96430889 |
| KIAA0430 | 0.974970849 | 1.007290401 |
| PRKAB1 | 0.974952335 | 0.975340688 |
| RFC1 | 0.974952335 | 0.975340688 |
| SERPINB13 | 0.974942017 | 0.95018226 |
| UHRF2 | 0.974880302 | 0.739779364 |
| ART5 | 0.974865913 | 0.751316322 |
| PRSS45 | 0.974798417 | 0.740785743 |
| ERI3 | 0.974794453 | 0.972935937 |
| SP140L | 0.974768627 | 0.950668037 |
| CD55 | 0.974739938 | 0.92709599 |
| ALX1 | 0.974725781 | 0.909798832 |
| ZSWIM1 | 0.974718747 | 1.020251114 |
| PLEKHH1 | 0.974717517 | 1.051044878 |
| SAMD12 | 0.974674632 | 1.075334143 |
| TIMM22 | 0.974612444 | 1.020897568 |
| CCNH | 0.974607483 | 1.030499676 |
| GLMN | 0.97458979 | 0.966769442 |
| CAB39 | 0.974571173 | 0.848926691 |
| LGMN | 0.974542859 | 1.041531473 |
| ENOPH1 | 0.974526148 | 0.992212849 |
| PAXBP1 | 0.97450796 | 0.927029805 |
| MPEG1 | 0.974483765 | 0.99554642 |

| | | |
|---|---|---|
| PRAMEF4 | 0.974466638 | 1.000342583 |
| ADAMTS16 | 0.97445996 | 1.003244646 |
| HPCA | 0.974449692 | 1.005138746 |
| ATP5G1 | 0.974446217 | 0.874443094 |
| KIAA1024 | 0.974435193 | 0.946214457 |
| CD8B | 0.974432106 | 0.948562171 |
| ZNF45 | 0.974417728 | 1.063595068 |
| KLHDC10 | 0.974399942 | 1.019527235 |
| EDAR | 0.974380249 | 0.984609154 |
| ZNF652 | 0.974366801 | 0.89145403 |
| TMEM200A | 0.974364148 | 0.933538883 |
| VCPIP1 | 0.974364148 | 0.933538883 |
| KRT18 | 0.974352436 | 1.039692183 |
| INS-IGF2 | 0.974335648 | 1.038711888 |
| PER3 | 0.974335648 | 1.038711888 |
| tcag7.1231_pseudogene | 0.974335648 | 1.038711888 |
| CAP1 | 0.97431851 | 0.853381936 |
| PRNP | 0.974294779 | 0.952723535 |
| OSTN | 0.974211953 | 0.976704351 |
| PLK4 | 0.974209203 | 1.047371836 |
| PRKAR2A | 0.974209203 | 1.047371836 |
| CECR5 | 0.974165375 | 0.854059609 |
| PRMT3 | 0.974139393 | 0.948734588 |
| PRR11 | 0.974137825 | 0.970028352 |
| AQP6 | 0.974068546 | 0.857837181 |
| CDH13 | 0.974042676 | 0.887632751 |
| CTDP1 | 0.973994175 | 1.031148605 |
| SLC38A10 | 0.973897821 | 0.930798219 |
| CDV3 | 0.97388725 | 0.982989064 |
| NEURL4 | 0.973873037 | 1.013703323 |
| SALL4 | 0.973857224 | 0.830421377 |
| CSF2RB | 0.973856329 | 0.853786958 |
| LOC440350_pseudogene | 0.973799636 | 1.001027749 |
| PRKCZ | 0.973784498 | 0.833874108 |
| ASF1B | 0.973746487 | 1.098321343 |
| IQGAP3 | 0.973727503 | 0.868790682 |
| TUBE1 | 0.973715593 | 0.965606749 |
| SIGLEC15 | 0.973710919 | 0.993148338 |
| GAMT | 0.973687106 | 0.896314297 |
| ZNF628 | 0.973686189 | 1.070068854 |
| MAMDC2 | 0.973645039 | 1.017129154 |
| DKK2 | 0.973635615 | 0.939020212 |
| COL8A1 | 0.973607523 | 0.990065091 |
| CLIC4 | 0.97359927 | 0.987670344 |

| | | |
|---|---|---|
| POLDIP3 | 0.973567968 | 0.963001028 |
| ACSS1 | 0.973543044 | 0.998702141 |
| KCNH2 | 0.97345718 | 0.889033095 |
| SKOR2 | 0.97345492 | 0.918465228 |
| C18orf32 | 0.97344835 | 0.964713943 |
| MZB1 | 0.973429603 | 0.782098015 |
| OR2M4 | 0.973414232 | 1.0668037 |
| EVI5 | 0.97335166 | 1.003083248 |
| TECPR2 | 0.973341193 | 0.961630695 |
| NAA50 | 0.973306364 | 0.98247891 |
| BRK1 | 0.973293084 | 0.915755367 |
| MTX3 | 0.973267665 | 0.824625354 |
| GPR183 | 0.973189136 | 0.872809864 |
| EFCAB14 | 0.973169923 | 0.936279548 |
| KIF26B | 0.97313585 | 0.950668037 |
| FADS6 | 0.973126153 | 1.038882139 |
| RP6-166C19.3 | 0.973103363 | 0.914011648 |
| TRAF3IP3 | 0.973029372 | 0.998629668 |
| PLEKHG4B | 0.973026019 | 0.947584789 |
| POMGNT2 | 0.973014645 | 1.045362495 |
| RPL10L | 0.972991824 | 1.01782098 |
| TLR5 | 0.97296003 | 0.988319273 |
| DEF6 | 0.972942015 | 1.041795135 |
| RFX6 | 0.972937938 | 0.879068174 |
| SPEF1 | 0.972925659 | 0.939705379 |
| OR5D18 | 0.972923529 | 1.051387461 |
| BMP2 | 0.972915413 | 0.935196436 |
| NOL9 | 0.972905451 | 1.060979788 |
| FCGR1C | 0.972866122 | 0.968482357 |
| ZNF638 | 0.972842467 | 1.077359255 |
| VAV2 | 0.972808846 | 1.013770757 |
| EMP3 | 0.972794759 | 0.946214457 |
| BET1 | 0.972789458 | 1.067488866 |
| C7orf66 | 0.972789458 | 1.067488866 |
| C16orf78 | 0.972764325 | 0.833535844 |
| BCAP31 | 0.972748408 | 1.030832477 |
| ATP6V1G2 | 0.972746954 | 1.039287161 |
| LPIN3 | 0.972730465 | 0.979787598 |
| PCDHB12 | 0.972730465 | 0.979787598 |
| ALAS2 | 0.972703808 | 0.89509923 |
| PSMD1 | 0.972695713 | 0.998972251 |
| ZNF266 | 0.972656837 | 0.957519699 |
| DDX19A | 0.972628213 | 0.840016201 |
| RALB | 0.972609874 | 0.973673552 |

| | | |
|---|---|---|
| TERF2IP | 0.972561777 | 1.080507023 |
| LRRC37A2 | 0.972548186 | 1.043742406 |
| RPGR | 0.972546281 | 0.954436451 |
| FOCAD | 0.972532553 | 0.893799246 |
| CERS2 | 0.972512018 | 1.060753341 |
| FDPS | 0.972499679 | 0.944841012 |
| FAM84B | 0.972494342 | 1.024706359 |
| BMP8A | 0.972482259 | 0.922576225 |
| NPL | 0.972453578 | 0.95587281 |
| SEC22C | 0.972448496 | 0.818955043 |
| FAM129C | 0.972351149 | 1.048197651 |
| PHYHIPL | 0.972328433 | 1.023295649 |
| RMND5B | 0.972321807 | 0.869987849 |
| KDM1B | 0.972321614 | 0.824140169 |
| PTTG1IP | 0.972277388 | 1.037667072 |
| TMPRSS3 | 0.97227576 | 0.917585983 |
| SLC22A23 | 0.972232407 | 0.85945727 |
| ASPDH | 0.97222404 | 1.020212402 |
| RFX7 | 0.972188854 | 0.982584042 |
| GLYAT | 0.972187262 | 0.939649578 |
| UQCRC2 | 0.972187262 | 0.939649578 |
| CPB2 | 0.972022287 | 0.983776768 |
| GCSAM | 0.972018515 | 1.009249743 |
| SPG7 | 0.971983265 | 0.994808566 |
| C1QTNF9 | 0.971924882 | 1.021061158 |
| LAMP5 | 0.971916433 | 1.001370332 |
| UPF1 | 0.971871388 | 1.121349773 |
| ACTRT2 | 0.971870376 | 0.931956258 |
| MOB3A | 0.971854075 | 0.878898339 |
| ATP6V1C2 | 0.97183566 | 1.038935756 |
| ECD | 0.971744034 | 0.874107722 |
| CBR1 | 0.97167828 | 0.919400567 |
| CD1D | 0.971574663 | 1.016101405 |
| PTRF | 0.971574209 | 0.917375456 |
| TAS1R2 | 0.971521242 | 0.921891059 |
| COLCA1 | 0.971518116 | 0.926687222 |
| CUTC | 0.971494374 | 0.965056526 |
| LAMA2 | 0.971458881 | 0.923855812 |
| LGALS13 | 0.971423961 | 1.077871512 |
| KIAA1751 | 0.971410279 | 0.904419322 |
| GKN1 | 0.971405114 | 0.914011648 |
| CARTPT | 0.971400605 | 1.149048198 |
| PARD6A | 0.971392744 | 0.937992463 |
| C1orf100 | 0.971388074 | 0.983394087 |

| | | |
|---|---|---|
| OR4F5 | 0.971383229 | 0.957177115 |
| EEFSEC | 0.971381757 | 0.809234508 |
| SOX2 | 0.971326182 | 0.947347104 |
| TSSK1B | 0.971295619 | 0.902011681 |
| SYNE1 | 0.97128894 | 0.901336074 |
| GPR6 | 0.971288128 | 1.01752109 |
| SBF2 | 0.971277789 | 0.894289186 |
| HNRNPA1L2 | 0.971248651 | 1.023896314 |
| C3orf62 | 0.97117657 | 0.9963548 |
| TMEM192 | 0.971161666 | 0.917437479 |
| C14orf183 | 0.971159128 | 0.927029805 |
| NPPA | 0.971148526 | 0.970195272 |
| ALDH7A1 | 0.971135007 | 0.957170668 |
| NUDT3 | 0.971091952 | 1.094743673 |
| EPHX1 | 0.971069087 | 0.881733495 |
| TANK | 0.97103865 | 0.933538883 |
| KRTAP5-3 | 0.971031763 | 0.99109284 |
| KDM6B | 0.971029679 | 1.010277492 |
| RHO | 0.971023212 | 0.874756652 |
| GJA1 | 0.970965174 | 1.023361454 |
| TFDP2 | 0.970948206 | 1.043337384 |
| CXCL3 | 0.970919939 | 0.997601918 |
| SHCBP1 | 0.970919939 | 0.978417266 |
| SPANXB1 | 0.970919939 | 1.059952038 |
| WNT4 | 0.970919939 | 1.007194245 |
| FBXL2 | 0.97091331 | 1.045425049 |
| KLK14 | 0.97091331 | 1.045425049 |
| BACE2 | 0.970902444 | 0.918883842 |
| PIGV | 0.970859824 | 0.860672337 |
| DNASE1 | 0.970819215 | 0.951979234 |
| PKN1 | 0.970819215 | 0.951979234 |
| VCAN | 0.970818002 | 0.928716079 |
| CINP | 0.9708126 | 1.032887975 |
| COX5B | 0.970766857 | 0.974042829 |
| ADAMDEC1 | 0.970741584 | 0.985074627 |
| AMT | 0.970716887 | 0.996106424 |
| DDX51 | 0.970713168 | 0.926690968 |
| TSACC | 0.970694523 | 0.969218307 |
| NAALADL2 | 0.970676878 | 0.869565217 |
| FOXH1 | 0.970671142 | 0.771567436 |
| PRR13 | 0.970671142 | 1.028756582 |
| VPS11 | 0.970645842 | 1.029201817 |
| MYNN | 0.970589813 | 1.007536828 |
| DNAJC14 | 0.970579325 | 1.06229721 |

| | | |
|---|---|---|
| SYT14 | 0.970562588 | 1.077764277 |
| PSMD5 | 0.970562582 | 0.930798219 |
| BIN3 | 0.970536892 | 0.868448099 |
| DGAT2 | 0.97052351 | 0.935756003 |
| SNX20 | 0.970482565 | 1.014045906 |
| IL34 | 0.970392958 | 0.946132037 |
| OR51E2 | 0.970389228 | 1.0445358 |
| ATP5B | 0.970386845 | 1.005670312 |
| ANXA2 | 0.970326474 | 0.934224049 |
| PHYH | 0.970273741 | 1.068137573 |
| FAM13B | 0.970273416 | 0.85748544 |
| TSEN34 | 0.970217267 | 0.919532771 |
| STATH | 0.970216371 | 0.945529291 |
| SSFA2 | 0.970212747 | 0.940733128 |
| UACA | 0.970082572 | 1.059266872 |
| ZMYM3 | 0.97002589 | 0.992120589 |
| CMTM3 | 0.970008003 | 1.048602673 |
| GJD3 | 0.970003779 | 0.968139774 |
| FAM216B | 0.969993638 | 0.997569866 |
| KHNYN | 0.969910939 | 0.989037342 |
| OR52N4 | 0.969908538 | 0.877012676 |
| CALCOCO2 | 0.969883282 | 0.914341337 |
| CRNKL1 | 0.969870251 | 0.950668037 |
| KCNK7 | 0.969850929 | 0.936404932 |
| WNT3A | 0.969794358 | 1.083590271 |
| CCDC171 | 0.969778726 | 0.971565605 |
| KCNG2 | 0.969772557 | 0.950992305 |
| LATS1 | 0.969696902 | 1.057754705 |
| ZNF682 | 0.96969253 | 0.823410288 |
| OR12D2 | 0.96967909 | 0.982870846 |
| MAST4 | 0.969672617 | 1.0798183 |
| VAV3 | 0.96964932 | 1.101881895 |
| ZNF569 | 0.969628117 | 1.11579308 |
| BPIFB3 | 0.969607003 | 1.01336074 |
| KIAA1407 | 0.969607003 | 1.01336074 |
| C1orf106 | 0.969593044 | 0.84690158 |
| IFNGR1 | 0.969584696 | 0.87605451 |
| KIAA1468 | 0.969532221 | 1.038477116 |
| GEMIN4 | 0.969442057 | 0.927906035 |
| STARD6 | 0.969386028 | 1.011990408 |
| SLC25A51 | 0.969341171 | 0.983213429 |
| ACRBP | 0.96932194 | 1.11745646 |
| MAFB | 0.96925164 | 0.797533402 |
| BEAN1 | 0.969247698 | 0.994518671 |

| | | |
|---|---|---|
| PRKG2 | 0.969195389 | 0.914990266 |
| CYP4X1 | 0.969187239 | 0.926022064 |
| PARK7 | 0.969187239 | 0.926022064 |
| CHRNA1 | 0.969163859 | 0.959117456 |
| EID2B | 0.969105852 | 0.830295666 |
| SERPING1 | 0.969067909 | 0.957862282 |
| BCAS4 | 0.969060006 | 0.879303362 |
| KIAA0895 | 0.969049151 | 0.948269955 |
| ZNF658 | 0.968995414 | 0.911300122 |
| CXCL2 | 0.968942537 | 1.009249743 |
| GPRC5C | 0.968853995 | 0.909798832 |
| SERPINC1 | 0.968853995 | 0.909798832 |
| ELSPBP1 | 0.968843918 | 0.883758607 |
| NELFB | 0.96882777 | 0.94127177 |
| ZNF33A | 0.968806988 | 1.0532122 |
| GSX1 | 0.968806065 | 0.996574169 |
| SEMA3D | 0.968770905 | 1.031860226 |
| THEM5 | 0.968738472 | 1.013770757 |
| LINC00471 | 0.968713918 | 0.873228028 |
| ZMIZ2 | 0.968632414 | 1.126771972 |
| MSH5 | 0.968613821 | 0.879708384 |
| YPEL3 | 0.968607046 | 1.109761037 |
| MTHFD1 | 0.968508567 | 0.992861778 |
| PEX10 | 0.968508567 | 1.048020766 |
| RDH12 | 0.968508567 | 0.926670993 |
| ZNF277 | 0.968503179 | 0.935196436 |
| FARP2 | 0.968474308 | 0.952038369 |
| OXR1 | 0.968474308 | 0.952038369 |
| CARS2 | 0.968470017 | 0.9181855 |
| ZNF362 | 0.968462528 | 0.902363823 |
| GRHL1 | 0.968406902 | 0.835155934 |
| LSM4 | 0.968393159 | 1.053100377 |
| HFM1 | 0.968335178 | 1.005670312 |
| PYGM | 0.968216484 | 1.075924724 |
| MAGEC2 | 0.968210198 | 0.941075711 |
| B3GALT6 | 0.968183143 | 0.965606749 |
| MSRB2 | 0.968183143 | 0.965606749 |
| S1PR1 | 0.968183143 | 0.965606749 |
| MAGEL2 | 0.968153743 | 0.400822199 |
| SCHIP1 | 0.968093824 | 0.89509923 |
| SRP14 | 0.967996972 | 1.100034258 |
| SLC17A9 | 0.967992078 | 0.984583762 |
| CYB561 | 0.967854926 | 0.976704351 |
| TSSK2 | 0.967830799 | 0.927319922 |

| | | |
|---|---|---|
| POLR1D | 0.967813806 | 0.833535844 |
| SH3GLB2 | 0.967810296 | 0.927029805 |
| MVP | 0.967805451 | 1.068043742 |
| OR5AS1 | 0.967777718 | 1.093867763 |
| NUP205 | 0.967702369 | 0.999314834 |
| NEDD8 | 0.967689664 | 0.831510733 |
| ASNSD1 | 0.967680801 | 0.904009721 |
| OR56A4 | 0.967660985 | 1.121506683 |
| SIT1 | 0.967635413 | 0.814499797 |
| PPP4R2 | 0.967617413 | 0.940047962 |
| KNDC1 | 0.967611318 | 1.06399352 |
| ALPK3 | 0.967575515 | 1.010382868 |
| OTUD4 | 0.967548633 | 0.855087359 |
| DPF1 | 0.967543903 | 0.651524984 |
| TYK2 | 0.967543903 | 0.977287476 |
| ABL2 | 0.96751015 | 0.944192083 |
| OLFM3 | 0.967503376 | 0.916970433 |
| TIE1 | 0.967477244 | 0.908464966 |
| PRF1 | 0.96744433 | 1.08461802 |
| CHAF1B | 0.967433094 | 0.699554642 |
| B3GNTL1 | 0.967421159 | 0.8669695 |
| SFSWAP | 0.967416523 | 0.92345079 |
| NLGN2 | 0.967403624 | 1.137573005 |
| SPA17 | 0.967402057 | 0.882494005 |
| OR2L8 | 0.967394444 | 0.943473792 |
| GPR143 | 0.967392758 | 0.844905905 |
| ATF7IP2 | 0.967373466 | 0.969167523 |
| MCM8 | 0.967370448 | 1.104477612 |
| ZFYVE21 | 0.967351491 | 1.025351148 |
| VPS37C | 0.967339196 | 0.865937627 |
| SPIRE1 | 0.967336403 | 0.990065091 |
| PRPF40B | 0.96731342 | 0.825435399 |
| BBS9 | 0.967191113 | 0.951397327 |
| VPS51 | 0.967176415 | 0.977389517 |
| TTLL7 | 0.967156932 | 0.84690158 |
| EMC9 | 0.967130199 | 1.023980815 |
| FBXO2 | 0.967121009 | 0.905905256 |
| TIMMDC1 | 0.967109538 | 0.90202124 |
| EDA2R | 0.967064782 | 1.08825438 |
| SLC38A9 | 0.967051995 | 0.940866748 |
| RBM23 | 0.96702345 | 0.939020212 |
| CHAC1 | 0.967007117 | 0.987849332 |
| C5orf28 | 0.966986129 | 1.011340624 |
| EPHB6 | 0.966971263 | 1.022063595 |

| | | |
|---|---|---|
| CHIA | 0.966937345 | 1 |
| CCDC176 | 0.966935322 | 1.001712915 |
| SLITRK1 | 0.966871436 | 0.931140802 |
| HSF2BP | 0.966864277 | 0.956834532 |
| MAGEA4 | 0.966864277 | 0.956834532 |
| SCARB2 | 0.966857964 | 1.23126772 |
| DEFB118 | 0.966851047 | 1.008221994 |
| KRTAP5-8 | 0.966829253 | 0.921548476 |
| LOC100499484-C9ORF174 | 0.966781503 | 0.896314297 |
| ZNF2 | 0.966751909 | 0.984241178 |
| ARHGAP27 | 0.966741216 | 1.061322371 |
| TRIM40 | 0.966731455 | 1.060999351 |
| RGAG1 | 0.966719101 | 1.045767517 |
| HENMT1 | 0.966700763 | 0.958282706 |
| PTCH1 | 0.966700763 | 0.958282706 |
| TMPRSS12 | 0.966654401 | 1.016872161 |
| FAM46B | 0.966627562 | 0.751316322 |
| NFRKB | 0.966620888 | 0.830295666 |
| SLC2A3 | 0.966557264 | 1.02673147 |
| ZSCAN26 | 0.96654643 | 1.064748201 |
| SPRR2B | 0.966530255 | 0.833504625 |
| MID2 | 0.96650202 | 0.939649578 |
| C1orf186 | 0.966489884 | 0.900993491 |
| FOXD4L5 | 0.966451099 | 0.942788626 |
| FSCN3 | 0.966375477 | 0.902706406 |
| DDRGK1 | 0.966371218 | 0.974991435 |
| INSM2 | 0.966368378 | 0.952207371 |
| G3BP1 | 0.966362061 | 0.975698663 |
| CHST9 | 0.966357708 | 1.022712524 |
| MPV17 | 0.966318432 | 1.011990408 |
| PPEF2 | 0.966285757 | 0.989617132 |
| FLJ45831_pseudogene | 0.966246888 | 0.925316889 |
| RRAGD | 0.966235046 | 0.967553537 |
| KLHL42 | 0.966204615 | 1.034600891 |
| EXTL2 | 0.966181948 | 0.945489942 |
| ARPC1A | 0.966135925 | 0.927029805 |
| RP6-166C19.6 | 0.966069728 | 1.005823912 |
| CDHR3 | 0.966034794 | 0.907845152 |
| COPS7B | 0.966018211 | 0.882151422 |
| OPLAH | 0.965993051 | 0.937626569 |
| PIP5K1C | 0.965954818 | 0.984425698 |
| LONRF3 | 0.965912751 | 1.089552239 |
| ATG14 | 0.965866424 | 1.009249743 |
| ZNHIT2 | 0.96585322 | 0.84163629 |

| | | |
|---|---|---|
| VPS26A | 0.96579343 | 0.994861254 |
| STARD13 | 0.965697917 | 0.912983899 |
| ZNF347 | 0.965670369 | 0.908187736 |
| CD22 | 0.965643138 | 0.882494005 |
| CRYAB | 0.965643138 | 0.882494005 |
| FAM219B | 0.965614366 | 1.107571086 |
| OR2D2 | 0.965531903 | 0.926002055 |
| RAB32 | 0.965519328 | 0.946138871 |
| KIAA0141 | 0.965492257 | 1.041795135 |
| GCN1L1 | 0.965487633 | 1.040233615 |
| ZNF831 | 0.965487137 | 1 |
| FANCI | 0.965460777 | 0.93600648 |
| GPD1L | 0.965460777 | 0.93600648 |
| CPSF6 | 0.965426558 | 0.948612539 |
| OR6N2 | 0.965420169 | 1.048304214 |
| KLF6 | 0.965373122 | 0.902011681 |
| ZNF92 | 0.965369332 | 1.038711888 |
| LYPD5 | 0.965356706 | 0.995949777 |
| GGCX | 0.965346212 | 0.795058728 |
| TTC5 | 0.965286663 | 1.04090725 |
| OLFML2A | 0.965282319 | 0.974042829 |
| KRTAP3-3 | 0.965274743 | 0.940461725 |
| HSBP1 | 0.965267714 | 1.017415958 |
| FAM102A | 0.965233125 | 0.855002025 |
| SMURF1 | 0.965168878 | 1.03504218 |
| FUT6 | 0.965096104 | 1.012978585 |
| DNAJB9 | 0.965042786 | 1.113052415 |
| CHMP3 | 0.965035582 | 1.055155875 |
| DDTL | 0.96501492 | 0.976361768 |
| GNG11 | 0.964988052 | 0.934566632 |
| UNC93A | 0.964926612 | 0.998972251 |
| RNF152 | 0.964885686 | 1.040882544 |
| MEP1A | 0.964870209 | 0.863724854 |
| PTCD2 | 0.964866214 | 0.952380952 |
| NFKBIA | 0.964852803 | 1.169578623 |
| DEPDC5 | 0.9648422 | 1.021582734 |
| KRT81 | 0.964824381 | 0.970838396 |
| ZFR | 0.964821177 | 1.090784515 |
| TLR3 | 0.964813989 | 0.825435399 |
| HELB | 0.964770433 | 0.924724205 |
| PARVB | 0.964748863 | 0.970195272 |
| TXNDC2 | 0.96472532 | 0.830629461 |
| MPC1 | 0.964721813 | 0.912298732 |
| FAM149B1 | 0.964677636 | 0.923603974 |

| | | |
|---|---|---|
| SLC16A12 | 0.964674419 | 1.082905104 |
| SMARCAD1 | 0.964646455 | 1.057754705 |
| COMMD9 | 0.964594896 | 0.776427704 |
| ZNF518A | 0.964574013 | 0.978417266 |
| RGS1 | 0.964565268 | 0.994518671 |
| MAX | 0.964535245 | 0.97326853 |
| IMMP2L | 0.964478268 | 1.013627515 |
| RNF32 | 0.964478268 | 1.013627515 |
| SSH2 | 0.964478268 | 1.013627515 |
| SERPINB4 | 0.964396263 | 1.002740665 |
| GPHA2 | 0.964393907 | 0.917437479 |
| OVOL2 | 0.96437025 | 0.962737951 |
| CHODL | 0.964338176 | 0.977046934 |
| SOX17 | 0.964335684 | 1.094552929 |
| C3orf58 | 0.964299595 | 0.984204131 |
| IST1 | 0.964233194 | 0.994861254 |
| FASTK | 0.964196723 | 0.947436729 |
| GABRR2 | 0.964196723 | 0.947436729 |
| LHCGR | 0.964145929 | 1.00843608 |
| ATP6V0A2 | 0.964130251 | 1.028776978 |
| SOX6 | 0.964116262 | 1.018631025 |
| JTB | 0.964099485 | 1.007879411 |
| GRPEL2 | 0.964097492 | 1.04008222 |
| FAM156A | 0.964094637 | 1.088386434 |
| SASH1 | 0.964065866 | 1.035286057 |
| CDCA7L | 0.964033968 | 1.127098321 |
| ZNF90 | 0.964033968 | 0.885577252 |
| DPPA4 | 0.964000167 | 0.977389517 |
| RBMX | 0.963996451 | 0.880781089 |
| DHX37 | 0.963893444 | 0.953277093 |
| ANO8 | 0.96385285 | 0.931146213 |
| AMY1A | 0.963852282 | 1.147306944 |
| PIM2 | 0.96383944 | 0.942245295 |
| QKI | 0.963839147 | 1.051032807 |
| ARHGEF10 | 0.963814085 | 0.870214147 |
| LUZP6 | 0.963745164 | 1.143542309 |
| AK8 | 0.9637424 | 1.019527235 |
| SCT | 0.9637424 | 1.019527235 |
| SLC35C2 | 0.963733474 | 0.879410757 |
| FGF12 | 0.96373309 | 0.914135277 |
| ANKRD20A2 | 0.96372328 | 1.063035286 |
| HRSP12 | 0.963694055 | 0.874614594 |
| ADPRH | 0.96367349 | 0.935601458 |
| SLC19A1 | 0.963669415 | 0.875658161 |

| | | |
|---|---|---|
| RASGEF1C | 0.963656131 | 0.961630695 |
| BEX4 | 0.963642583 | 1.096951011 |
| VAMP5 | 0.963639638 | 0.837615622 |
| AES | 0.96363325 | 0.989037342 |
| MORC4 | 0.963629477 | 1.076954232 |
| CHI3L1 | 0.963610005 | 0.898118105 |
| OR10A7 | 0.963605339 | 0.924631723 |
| RIT1 | 0.963593952 | 0.959117456 |
| EPB42 | 0.963564682 | 0.993519644 |
| COPS3 | 0.963558897 | 0.903734156 |
| ZYG11B | 0.963503197 | 0.970028352 |
| TMTC2 | 0.963488017 | 0.910085055 |
| EXOSC3 | 0.963479922 | 0.953751285 |
| TEX11 | 0.963470407 | 0.93764988 |
| TMEM109 | 0.963433816 | 0.931551235 |
| C20orf197 | 0.963402845 | 1.012960713 |
| IL7 | 0.963401826 | 0.84163629 |
| HIRIP3 | 0.96337716 | 0.9554642 |
| MATK | 0.963356861 | 1.097988319 |
| SENP8 | 0.963356861 | 1.036988968 |
| CRIP1 | 0.963316762 | 0.874957177 |
| MFSD8 | 0.963263982 | 0.950992305 |
| RPF1 | 0.963256493 | 0.93600648 |
| GPR179 | 0.963244984 | 1.014925373 |
| OR2T8 | 0.96320086 | 0.99109284 |
| DIXDC1 | 0.963197507 | 0.947584789 |
| TM9SF2 | 0.963197507 | 1.105515588 |
| FJX1 | 0.963172859 | 0.958890031 |
| MAPK8IP2 | 0.9631401 | 0.997601918 |
| SYT7 | 0.963133983 | 0.954093868 |
| REG4 | 0.963118533 | 0.937992463 |
| CCDC90B | 0.963105914 | 0.889023896 |
| PXMP2 | 0.963087133 | 0.976704351 |
| GCLM | 0.963017788 | 1.013365735 |
| IRF8 | 0.962978717 | 1.034831916 |
| TMPRSS2 | 0.962967143 | 1.020765737 |
| TMPRSS11B | 0.962948102 | 0.92345079 |
| SUPV3L1 | 0.962938372 | 0.842041312 |
| LILRA3 | 0.962909021 | 0.96882494 |
| ZDHHC15 | 0.962906801 | 0.996231586 |
| KDM5C | 0.962882103 | 0.837767683 |
| PCSK1 | 0.962877658 | 0.948734588 |
| HCRTR1 | 0.962844811 | 0.887735237 |
| CDH11 | 0.962828914 | 1.041452552 |

| | | |
|---|---|---|
| IL17REL | 0.962828914 | 0.959232614 |
| NFIA | 0.962807609 | 1.024301337 |
| C12orf57 | 0.962750097 | 0.96435804 |
| OBFC1 | 0.962727964 | 0.960945529 |
| CCDC84 | 0.962722589 | 1.103685703 |
| SLC34A3 | 0.962671089 | 0.735115431 |
| SSR2 | 0.962668427 | 1.088386434 |
| KBTBD8 | 0.962649032 | 0.874443094 |
| NKRF | 0.962612712 | 0.895909275 |
| HIST1H4D | 0.96256861 | 0.969167523 |
| RHBDF1 | 0.962535443 | 0.925659472 |
| ZBTB8OS | 0.962532481 | 0.975293641 |
| HOXC12 | 0.962518316 | 0.936964714 |
| FMNL3 | 0.962470424 | 0.866050017 |
| DPH1 | 0.962387709 | 0.99473471 |
| COG7 | 0.962366892 | 1.050017129 |
| THAP10 | 0.962313064 | 0.940390545 |
| VPS39 | 0.96224809 | 1.03562864 |
| COX6A1 | 0.962226279 | 0.999189955 |
| MAGEB6 | 0.962219907 | 0.892086331 |
| HUNK | 0.962219566 | 1.099286178 |
| ZFPL1 | 0.962178766 | 1.014731072 |
| RBM25 | 0.962167031 | 1.026036314 |
| SLC10A4 | 0.962158682 | 1.027136493 |
| TMEM14A | 0.962146551 | 1.137375814 |
| DDX43 | 0.962144011 | 0.771972458 |
| NRG2 | 0.962137796 | 1.048602673 |
| CLDN11 | 0.962129696 | 0.971223022 |
| LCP1 | 0.962122492 | 1.07125728 |
| FOXE3 | 0.962116707 | 0.793438639 |
| CD82 | 0.962104176 | 0.924260834 |
| GCSAML | 0.962104176 | 0.924260834 |
| POU6F2 | 0.962097617 | 0.955121617 |
| HELT | 0.96209088 | 1.018631025 |
| H6PD | 0.962086703 | 0.98217902 |
| APOBEC4 | 0.962055579 | 0.930741191 |
| C19orf66 | 0.962053032 | 1.050359712 |
| GPR141 | 0.962051874 | 0.827384815 |
| WDR45B | 0.96204101 | 0.988659376 |
| C21orf91 | 0.961916741 | 0.935936965 |
| OR1A2 | 0.961863412 | 1.065090785 |
| CDC40 | 0.961862369 | 1.003768414 |
| HOMER1 | 0.96181493 | 0.93764988 |
| BTBD17 | 0.96179291 | 0.898937992 |

| | | |
|---|---|---|
| HSF2 | 0.961792269 | 0.948157149 |
| NOL12 | 0.961792269 | 0.948157149 |
| SCAP | 0.961760332 | 0.871531346 |
| TMA16 | 0.961745108 | 0.966769442 |
| ESPNL | 0.961679284 | 0.787363305 |
| CRISPLD1 | 0.961673021 | 1.007194245 |
| MT3 | 0.961666269 | 0.934566632 |
| MMRN2 | 0.96165322 | 0.957177115 |
| PIEZO2 | 0.961612442 | 0.941075711 |
| ABTB2 | 0.961573747 | 1.020212402 |
| USP24 | 0.96155176 | 1.038935756 |
| ZNF385B | 0.961540523 | 0.86472256 |
| ZKSCAN3 | 0.961538587 | 0.965573107 |
| EIF5B | 0.961530742 | 0.88618874 |
| ADAMTS8 | 0.961525828 | 0.900064893 |
| MRPL51 | 0.961521641 | 0.920178143 |
| FAM181A | 0.96150755 | 1.071942446 |
| TMEM30B | 0.961476134 | 0.914135277 |
| TEPP | 0.961443754 | 0.994518671 |
| WNK1 | 0.961443754 | 0.994518671 |
| CCBL2 | 0.961439142 | 0.977936405 |
| NIPSNAP3B | 0.961401072 | 0.978417266 |
| C5orf46 | 0.961373932 | 1.08016444 |
| MTMR8 | 0.961365298 | 1.055807917 |
| MRPS25 | 0.961357077 | 0.962315862 |
| EIF4EBP1 | 0.961301053 | 0.940056703 |
| GPER1 | 0.961297875 | 0.828033744 |
| AGTRAP | 0.961294142 | 1.036656389 |
| EGLN2 | 0.961289807 | 1.044776119 |
| LGALS14 | 0.961264328 | 0.930113052 |
| OR11H12 | 0.961252412 | 1.009249743 |
| ARRDC5 | 0.961223013 | 0.888618874 |
| OR1L3 | 0.961210681 | 0.959232614 |
| TRIM52 | 0.961134134 | 0.767034393 |
| PARD3 | 0.961112677 | 0.994861254 |
| PFKFB1 | 0.961105074 | 0.933809215 |
| FA2H | 0.961078401 | 0.972744971 |
| UBE2M | 0.961078401 | 0.972744971 |
| NT5E | 0.961053704 | 1.011680727 |
| HLA-E | 0.96105221 | 0.93600648 |
| CDHR2 | 0.961011509 | 0.942486837 |
| PCDHB6 | 0.96101141 | 0.92874272 |
| COL5A3 | 0.960984949 | 1.104830421 |
| SPCS3 | 0.960971546 | 0.833874108 |

| | | |
|---|---|---|
| SRSF3 | 0.960963608 | 0.94655704 |
| WDR90 | 0.960959448 | 0.884568651 |
| ACBD5 | 0.960924189 | 0.998287085 |
| SHPRH | 0.960870535 | 1.028552888 |
| GOLT1B | 0.960839686 | 1.033915725 |
| CAPG | 0.960838463 | 0.857828023 |
| JSRP1 | 0.960793383 | 1.024301337 |
| MRC1 | 0.960781781 | 0.995203837 |
| BCL2L13 | 0.960776471 | 0.867557716 |
| NIM1K | 0.960762774 | 1.095392602 |
| ZNF501 | 0.960757042 | 1.058239123 |
| SRRM2 | 0.960741138 | 1.013018157 |
| CEP83 | 0.960722627 | 1.037262049 |
| WDR31 | 0.960713709 | 0.967553537 |
| HCN2 | 0.960693129 | 1.045425049 |
| AGR2 | 0.960683142 | 0.985611511 |
| SNAPC1 | 0.96065138 | 0.917780062 |
| RAB40A | 0.960649744 | 0.839714471 |
| ETFB | 0.960640277 | 0.878650227 |
| MC5R | 0.960631716 | 0.917585983 |
| TIMM17A | 0.960603759 | 0.92428914 |
| CHN2 | 0.96055777 | 1.102089757 |
| ART1 | 0.960537918 | 0.977318753 |
| HBZ | 0.960535891 | 0.906554186 |
| KIF5A | 0.960534149 | 0.906439854 |
| TCTN3 | 0.960517581 | 0.948612539 |
| CD151 | 0.960487857 | 0.903391572 |
| PNRC2 | 0.960479883 | 0.892086331 |
| RAD54B | 0.960467111 | 0.990279465 |
| ZNF473 | 0.960454917 | 0.858170606 |
| FAM154B | 0.960453408 | 0.799108951 |
| ABCC12 | 0.96043768 | 1.012329656 |
| ABCG1 | 0.96043768 | 0.895522388 |
| PPARA | 0.96043768 | 0.856586632 |
| XPO6 | 0.960400501 | 0.990750257 |
| DEFB125 | 0.960366428 | 0.945529291 |
| MEDAG | 0.960362837 | 1.10854597 |
| TCF15 | 0.960295545 | 0.97326853 |
| DLEU7 | 0.960278437 | 0.958547448 |
| NR5A2 | 0.960242491 | 0.912394549 |
| NAA15 | 0.960238492 | 1.085054678 |
| PSME4 | 0.960216408 | 0.78725591 |
| TRMT10A | 0.960213734 | 0.994176088 |
| TMEM59 | 0.960181948 | 1.020656136 |

| | | |
|---|---|---|
| DGCR6 | 0.960180704 | 0.908530319 |
| PIK3R1 | 0.960175316 | 1.018170019 |
| KIF13B | 0.960153582 | 0.940298507 |
| FBXO43 | 0.960141316 | 0.901362751 |
| ZBTB39 | 0.960109432 | 0.984583762 |
| TMEM143 | 0.960107015 | 0.984204131 |
| CCDC158 | 0.960097742 | 0.97327852 |
| NCAPG2 | 0.96009428 | 0.934791414 |
| MRAP2 | 0.960066116 | 0.898937992 |
| HEMGN | 0.960031545 | 0.957177115 |
| CACNG8 | 0.960023182 | 1.073977936 |
| SDCCAG8 | 0.959998876 | 1.055841041 |
| ZXDB | 0.959995931 | 0.842411785 |
| TMEM163 | 0.959988987 | 0.848926691 |
| GLG1 | 0.959976675 | 0.952380952 |
| SNCB | 0.959975542 | 0.992805755 |
| TSPAN5 | 0.959935896 | 0.999314834 |
| CTF1 | 0.959935352 | 0.958890031 |
| SATL1 | 0.959843918 | 1.001027749 |
| METTL21A | 0.959802749 | 0.993114621 |
| CPQ | 0.959800992 | 1.118754056 |
| UMOD | 0.959799423 | 0.967112025 |
| CASP14 | 0.959797864 | 0.974042829 |
| PCDHAC2 | 0.959785785 | 0.886262419 |
| ZNF554 | 0.959767435 | 1.014045906 |
| ZNF630 | 0.959752755 | 1.072285029 |
| LHFPL4 | 0.959742487 | 1.034021871 |
| PLCB2 | 0.959742487 | 0.95018226 |
| SUPT5H | 0.959742487 | 1.034021871 |
| FAM189B | 0.959735039 | 0.921891059 |
| GRIK4 | 0.959697923 | 0.963011032 |
| PRAMEF1 | 0.95968681 | 1.074929121 |
| COQ6 | 0.95963297 | 0.88513952 |
| BTBD2 | 0.959623803 | 1.010962658 |
| KLRC4 | 0.959602055 | 0.871873929 |
| ZNF341 | 0.959595485 | 0.930113052 |
| TOP2B | 0.959560032 | 0.913043478 |
| C10orf25 | 0.959462728 | 1.054813292 |
| DNER | 0.959432053 | 0.83967112 |
| ATF6B | 0.959349367 | 1.063595068 |
| STAT5A | 0.959338481 | 0.888213852 |
| NPAS2 | 0.959274864 | 0.918883842 |
| ZNF592 | 0.959274864 | 0.918883842 |
| FOXK1 | 0.959227752 | 0.948269955 |

| | | |
|---|---|---|
| APOF | 0.959223412 | 0.879948086 |
| GORASP2 | 0.959213842 | 0.966084275 |
| UBE2H | 0.959207935 | 1.013627515 |
| SLC12A9 | 0.95919488 | 0.982989064 |
| CSRP1 | 0.959178138 | 0.972593354 |
| ZNF556 | 0.959142978 | 0.979102432 |
| EXOC3L2 | 0.959124634 | 0.96800324 |
| FNDC1 | 0.959120595 | 0.920863309 |
| CES3 | 0.959051768 | 0.963659961 |
| ADD3 | 0.959047556 | 0.933881466 |
| PLXNC1 | 0.959011354 | 1.052072628 |
| TP63 | 0.959000975 | 0.99156392 |
| GPR125 | 0.958984592 | 0.857884491 |
| EIF4EBP2 | 0.958973837 | 1.011647825 |
| SLC25A32 | 0.958921586 | 1.019846092 |
| NKAIN4 | 0.9588945 | 1.060294621 |
| C11orf84 | 0.958849761 | 1.004860267 |
| COG8 | 0.958801525 | 0.997259335 |
| TRIO | 0.958700907 | 0.768981181 |
| THBS1 | 0.958691649 | 1.088043851 |
| NDEL1 | 0.958661078 | 0.912515188 |
| AGRN | 0.958621419 | 1.000685166 |
| ANKRD39 | 0.95849807 | 1.033211827 |
| LYRM7 | 0.958489817 | 1.074119077 |
| GALE | 0.958477646 | 0.908500973 |
| CATIP | 0.95847226 | 1.086673518 |
| MTA3 | 0.95847226 | 0.961973279 |
| RAD51AP1 | 0.95843577 | 1.052652896 |
| MRPL40 | 0.958413131 | 1.010620075 |
| TMEM27 | 0.958369904 | 0.914945322 |
| KIAA1161 | 0.958346843 | 0.952380952 |
| FAM69A | 0.95834423 | 0.962332928 |
| TMEM80 | 0.95834423 | 0.962332928 |
| SYT6 | 0.958341995 | 0.916752312 |
| OR10G4 | 0.958339412 | 0.898937992 |
| FBXO21 | 0.958330287 | 0.802725503 |
| ESYT3 | 0.958322784 | 0.968813285 |
| COL4A6 | 0.958296459 | 1.018842069 |
| HMCN2 | 0.958293287 | 1.001027749 |
| OGFRL1 | 0.958232527 | 0.996231586 |
| CHMP6 | 0.958216778 | 0.960307817 |
| AFTPH | 0.958192276 | 0.925880923 |
| BTBD8 | 0.958183774 | 0.931483385 |
| ETV3L | 0.958183774 | 0.931483385 |

| | | |
|---|---|---|
| FASTKD3 | 0.958108836 | 0.951802349 |
| FGFR3 | 0.958094514 | 1.025957171 |
| KMT2B | 0.958094514 | 1.025957171 |
| ARL4D | 0.958085403 | 1.046577562 |
| ARHGEF17 | 0.95804726 | 0.931213498 |
| CXorf67 | 0.958024239 | 1.066118534 |
| MPP2 | 0.958015072 | 0.808565866 |
| FOXD4L1 | 0.957964249 | 0.881466256 |
| TCP11L1 | 0.957909558 | 0.880923451 |
| MAP7 | 0.957907159 | 1.056526208 |
| PNLIPRP3 | 0.95790645 | 0.928311057 |
| CGN | 0.957870866 | 0.943131209 |
| CYP2R1 | 0.957816585 | 0.956149366 |
| HHIP | 0.957816585 | 0.956149366 |
| TAS2R43 | 0.957792526 | 0.878383008 |
| PTPLA | 0.957747274 | 0.992861778 |
| GNRH2 | 0.957724289 | 0.957862282 |
| GINM1 | 0.957642324 | 0.935251799 |
| ACY3 | 0.9576274 | 1.03199676 |
| ZNF334 | 0.957599914 | 0.923946557 |
| ANKRD36 | 0.957498104 | 0.949982871 |
| DNAJB2 | 0.957444729 | 1.002025111 |
| RNF44 | 0.957403575 | 0.792342635 |
| HAMP | 0.957335035 | 0.971223022 |
| MAML3 | 0.957289408 | 0.984241178 |
| SLC25A15 | 0.957265063 | 0.905224787 |
| AMPD3 | 0.957246831 | 1.004542505 |
| DDR2 | 0.957246831 | 1.004542505 |
| HR | 0.957246831 | 0.837118754 |
| CNTN2 | 0.957223709 | 1.028091812 |
| OLFML2B | 0.957222078 | 0.955121617 |
| CCDC151 | 0.957177221 | 0.968139774 |
| OR2T6 | 0.957162813 | 0.797875985 |
| TEC | 0.957131497 | 1.077222583 |
| CLEC4E | 0.957056516 | 0.927714971 |
| ZDHHC14 | 0.95703436 | 1.038026721 |
| ITIH3 | 0.957014741 | 1.0445358 |
| MPPE1 | 0.956941988 | 0.815055159 |
| FBXL20 | 0.956884033 | 1.027255029 |
| MTIF2 | 0.956863123 | 0.971447112 |
| DCAF8L1 | 0.95684068 | 0.978938842 |
| TRIP10 | 0.956788148 | 0.957177115 |
| TRPV6 | 0.956783269 | 1.072031149 |
| SMG5 | 0.956768702 | 0.86472256 |

| | | |
|---|---|---|
| CD109 | 0.956754528 | 1.025351148 |
| YLPM1 | 0.956736495 | 1.031860226 |
| TAS2R5 | 0.956688603 | 0.903734156 |
| LSM3 | 0.956661462 | 0.824254882 |
| NPC1 | 0.95665159 | 1.015795869 |
| COL27A1 | 0.956641662 | 0.978417266 |
| ZNF615 | 0.956629338 | 0.923261391 |
| ZNF503 | 0.956620865 | 1.015758822 |
| EPB41 | 0.956612926 | 0.95544755 |
| GPC6 | 0.956610126 | 0.929770469 |
| UBXN2B | 0.956592999 | 0.905447071 |
| CNPY2 | 0.956572519 | 0.942788626 |
| BRINP3 | 0.956564279 | 0.987849332 |
| HS1BP3 | 0.956490008 | 0.946942082 |
| PPP2R1A | 0.95647538 | 0.966255678 |
| ABT1 | 0.956471669 | 0.959902795 |
| ZCCHC24 | 0.956436273 | 0.865022268 |
| PLD6 | 0.956421231 | 0.902363823 |
| ZDHHC20 | 0.956409217 | 0.906034832 |
| COMMD1 | 0.956403061 | 0.908872902 |
| SCUBE3 | 0.956403061 | 0.908872902 |
| MEIS3 | 0.956372976 | 0.952723535 |
| ARFGEF3 | 0.956371012 | 1.037667072 |
| EXT2 | 0.956336419 | 0.955223881 |
| LTB4R2 | 0.956336419 | 0.955223881 |
| CALR3 | 0.956297264 | 1.084275437 |
| OR2T3 | 0.956282946 | 0.954436451 |
| DAB2 | 0.956214982 | 0.830700689 |
| SFTPA1 | 0.956181724 | 0.925316889 |
| KLF14 | 0.956156487 | 0.863651936 |
| FAM177A1 | 0.956139496 | 0.870161014 |
| PPAN | 0.956096036 | 1.066018631 |
| HIST1H1D | 0.956074069 | 0.970880439 |
| C1GALT1C1 | 0.956025892 | 1.005670312 |
| FLJ22184 | 0.955997405 | 0.92874272 |
| LOC100132247 | 0.955975973 | 1.016443988 |
| MRPS14 | 0.955944846 | 0.992120589 |
| PEX1 | 0.955941178 | 0.951802349 |
| C9orf40 | 0.95590929 | 0.990684488 |
| GNLY | 0.955884085 | 0.899623159 |
| PIH1D1 | 0.955861304 | 1.055488052 |
| PGAM4 | 0.955837952 | 0.793640493 |
| HRC | 0.955833424 | 0.966904607 |
| COL9A2 | 0.955800988 | 0.938677629 |

| | | |
|---|---|---|
| SEC61A2 | 0.95578309 | 1.076738609 |
| RIPK3 | 0.955773163 | 1.011680727 |
| MSI2 | 0.955760889 | 1.045906132 |
| KCNMB1 | 0.95575744 | 1.084360805 |
| SLC10A6 | 0.955749289 | 0.920863309 |
| ERICH1 | 0.955744142 | 0.837586067 |
| SGCA | 0.95573624 | 0.927372388 |
| YEATS4 | 0.955712457 | 0.882948562 |
| SLC30A3 | 0.955687136 | 0.95587281 |
| SLC34A1 | 0.955687136 | 0.95587281 |
| B4GALT7 | 0.95568011 | 0.934791414 |
| C3orf70 | 0.955665874 | 0.872216513 |
| CHRND | 0.955632802 | 1.000648929 |
| TRIM3 | 0.955626381 | 1.073329007 |
| MRPL27 | 0.955529674 | 0.994176088 |
| FAM209B | 0.955521228 | 0.984609154 |
| IL31 | 0.955472312 | 0.875642343 |
| COLEC12 | 0.95546547 | 0.982870846 |
| PRAMEF12 | 0.955421741 | 0.878898339 |
| CCL23 | 0.955410011 | 0.965056526 |
| BIRC6 | 0.9553953 | 1.006489293 |
| ARHGEF5 | 0.955393836 | 0.921205892 |
| EVX1 | 0.955389089 | 0.989064399 |
| DIO2 | 0.955384046 | 1.008505468 |
| NACC2 | 0.955358313 | 1.054470709 |
| HBQ1 | 0.955332577 | 0.960260363 |
| ZBTB33 | 0.955313728 | 1.030147311 |
| LSM10 | 0.955294381 | 0.986296677 |
| ARSB | 0.955275441 | 0.876873228 |
| SLAMF1 | 0.955227278 | 0.922777417 |
| DLK2 | 0.955157818 | 0.963686194 |
| DYNLL2 | 0.955128326 | 0.933171324 |
| NEUROG2 | 0.955112282 | 0.932853717 |
| MARCH7 | 0.955107168 | 1.219338092 |
| REL | 0.955101406 | 0.984425698 |
| ANAPC7 | 0.955032356 | 0.997944502 |
| BTNL2 | 0.955030589 | 0.928057554 |
| TFG | 0.954993344 | 1.10688592 |
| UBE2E2 | 0.954990269 | 1.046073978 |
| DPP8 | 0.954949437 | 0.9733939 |
| PAN3 | 0.954935031 | 1.012675574 |
| CCNG1 | 0.954881603 | 0.945489942 |
| MRGPRF | 0.95486308 | 0.990266061 |
| MMP2 | 0.954846204 | 1.03504218 |

| | | |
|---|---|---|
| OR6V1 | 0.954834443 | 1.033915725 |
| NQO1 | 0.954809735 | 0.917585983 |
| PCDHB9 | 0.954740824 | 0.998379911 |
| MEOX1 | 0.954722384 | 0.953017416 |
| SLC27A1 | 0.954716831 | 0.940056703 |
| MRPS27 | 0.954699053 | 0.901174565 |
| RBM41 | 0.95463115 | 0.778047793 |
| EML5 | 0.954626336 | 0.993490922 |
| GNB5 | 0.954603953 | 0.910585817 |
| PCDHB15 | 0.954499968 | 0.931825968 |
| MCMBP | 0.954475274 | 0.970880439 |
| CLCA4 | 0.954472209 | 1.029850746 |
| FAM219A | 0.954452438 | 1.009934909 |
| HPD | 0.954419952 | 0.910085055 |
| FBN3 | 0.954393581 | 1.127098321 |
| HLA-F | 0.954390228 | 0.972593354 |
| OCEL1 | 0.954373291 | 1.024706359 |
| GOLGA2 | 0.954366486 | 0.850633779 |
| ENTPD6 | 0.954353679 | 0.832320778 |
| PREX1 | 0.954344239 | 1.070229531 |
| PI4KAP2 | 0.954326024 | 0.92874272 |
| MRPL13 | 0.954322989 | 0.935251799 |
| MOB2 | 0.954317053 | 0.948269955 |
| SMPX | 0.954297253 | 0.993833505 |
| KCNIP3 | 0.954247879 | 1.03569111 |
| G6PD | 0.954226721 | 0.946138871 |
| APOOL | 0.954216853 | 0.990279465 |
| INPP5K | 0.954201037 | 0.856586632 |
| MUC20 | 0.954192406 | 1.067146283 |
| S100A3 | 0.954190005 | 1.09733939 |
| KRT38 | 0.954154655 | 1.059538275 |
| HAND1 | 0.954124855 | 0.899554475 |
| OR10H3 | 0.954111438 | 1.088386434 |
| HNRNPH1 | 0.954070116 | 0.888317917 |
| UAP1 | 0.954000086 | 1.147955873 |
| PMM2 | 0.953996231 | 1.025111381 |
| SUN1 | 0.953974195 | 0.929085303 |
| PCDHB3 | 0.953964771 | 1.039739637 |
| OTOA | 0.953964363 | 1.046248715 |
| RPA2 | 0.953886204 | 0.943816375 |
| SLC30A4 | 0.953744719 | 1.064243997 |
| FOXD4L3 | 0.953738178 | 1.103460089 |
| NHLRC4 | 0.953716067 | 0.960260363 |
| ATG9A | 0.953707594 | 0.914696814 |

| | | |
|---|---|---|
| SLC38A7 | 0.953684121 | 0.98217902 |
| KCNE1L | 0.953677873 | 0.974691759 |
| PSG11 | 0.953627985 | 0.948955122 |
| MTMR3 | 0.953597525 | 0.88513952 |
| EGFR | 0.953590116 | 1.0532122 |
| ZNF536 | 0.953571136 | 0.910895099 |
| CNOT11 | 0.953540084 | 0.944158959 |
| ANKFY1 | 0.953517203 | 0.885577252 |
| PSIP1 | 0.953513397 | 1.095580678 |
| TMSB4X | 0.953482224 | 1.010962658 |
| FCHO1 | 0.953445296 | 0.926344639 |
| DAB1 | 0.953412457 | 0.826650466 |
| OR5AU1 | 0.953399081 | 1.006166495 |
| STRIP1 | 0.953375973 | 0.960602946 |
| NPVF | 0.953375831 | 0.882948562 |
| CLYBL | 0.953327258 | 1.014276444 |
| TM4SF20 | 0.953315167 | 1.001370332 |
| IGFL3 | 0.953314907 | 0.995544755 |
| DCTD | 0.953251843 | 0.824625354 |
| CTPS2 | 0.953230957 | 1.019441069 |
| RASSF3 | 0.953217298 | 0.977046934 |
| SCARA3 | 0.953216286 | 0.924724205 |
| ACAP2 | 0.953194236 | 0.944501542 |
| TNIK | 0.953160224 | 1.003244646 |
| SS18 | 0.953159166 | 1.011305242 |
| PHLDB2 | 0.953063925 | 0.894141829 |
| C19orf18 | 0.952991589 | 0.965573107 |
| DOCK2 | 0.952983184 | 0.995203837 |
| KIR3DL1 | 0.952933932 | 1.029462145 |
| KIAA1551 | 0.952922463 | 0.876873228 |
| MTCP1 | 0.952906293 | 0.917094896 |
| ITGAE | 0.952887405 | 1.025957171 |
| TDP2 | 0.952874885 | 0.801134062 |
| PNLDC1 | 0.952870272 | 0.884549503 |
| CEP55 | 0.952868467 | 0.970028352 |
| HLF | 0.952862751 | 1.117505995 |
| C22orf15 | 0.952823894 | 1.063183475 |
| URM1 | 0.952769949 | 0.874848117 |
| ZDHHC3 | 0.952723899 | 0.911705144 |
| PLEKHF1 | 0.952720162 | 0.980815348 |
| FAM188B | 0.95269429 | 1.028091812 |
| CCDC88C | 0.952681331 | 0.881466256 |
| MYO18A | 0.952680108 | 0.948269955 |
| LOC100129550 | 0.952655822 | 1.06235012 |

| | | |
|---|---|---|
| RHBDL2 | 0.952648668 | 0.891628812 |
| PJA1 | 0.952636648 | 0.970149254 |
| NFKB2 | 0.952589642 | 0.987021415 |
| SPP2 | 0.952584006 | 1.003768414 |
| CSNK1D | 0.952544117 | 1.003893576 |
| PCDHGA8 | 0.952531311 | 1.025008565 |
| TATDN2 | 0.952513867 | 1.011990408 |
| OR51F2 | 0.95249597 | 0.998972251 |
| FSD1 | 0.952458816 | 0.972935937 |
| MVB12A | 0.952359829 | 0.931551235 |
| FOXP4 | 0.952342415 | 0.793764988 |
| STOM | 0.952341781 | 1.163754711 |
| UBE2D1 | 0.952337815 | 1.08825438 |
| NTN5 | 0.952327599 | 0.995889003 |
| SPATA12 | 0.952311288 | 0.881808839 |
| FANCC | 0.952294167 | 0.95544755 |
| NCAPH | 0.952287036 | 0.862292426 |
| CDC26 | 0.952209477 | 0.877012676 |
| PC | 0.952202318 | 1.018842069 |
| MOAP1 | 0.952169967 | 0.952038369 |
| DLGAP5 | 0.952146496 | 0.998702141 |
| RANGAP1 | 0.952134037 | 0.932511134 |
| FMNL1 | 0.952109207 | 0.919492977 |
| THEM4 | 0.952093718 | 0.977318753 |
| DBNDD1 | 0.952090865 | 0.933981369 |
| CSRP3 | 0.952059774 | 1.028776978 |
| GPC4 | 0.951952481 | 0.929427886 |
| PLD3 | 0.951903395 | 0.8137573 |
| PSMF1 | 0.951882297 | 0.815347722 |
| EYA2 | 0.951862316 | 1.038286827 |
| C15orf48 | 0.951851293 | 1.02308627 |
| MARCH2 | 0.951844534 | 0.942894225 |
| IGF2BP3 | 0.951816807 | 1.090319968 |
| DCN | 0.951813335 | 0.945871874 |
| PPP2R4 | 0.951744676 | 0.993510707 |
| TMED10 | 0.951744676 | 0.993510707 |
| EPHB1 | 0.951741548 | 1.10577547 |
| HIST1H4F | 0.951715421 | 1.016101405 |
| KCNAB1 | 0.95165445 | 1.04412719 |
| CORO1A | 0.951566761 | 0.95101062 |
| SLC1A1 | 0.951548454 | 1.116646416 |
| SLITRK4 | 0.951546009 | 1.047276465 |
| PITPNM3 | 0.951528166 | 0.999351071 |
| ZYX | 0.95151626 | 0.999594978 |

| | | |
|---|---|---|
| AFM | 0.951481671 | 0.919400567 |
| BMP5 | 0.951445349 | 0.920830629 |
| AXIN1 | 0.951437311 | 0.967454608 |
| NDUFS6 | 0.951427777 | 1.010530579 |
| GDF10 | 0.951411832 | 0.936816525 |
| MYH13 | 0.951411832 | 0.728635075 |
| SYT16 | 0.951411832 | 0.936816525 |
| PLCXD3 | 0.951382695 | 0.826245443 |
| PSMD11 | 0.951351214 | 1.058323208 |
| CELF6 | 0.951346375 | 0.900650908 |
| HTR1A | 0.951338658 | 0.988319273 |
| C17orf77 | 0.951330328 | 0.984609154 |
| TCL6 | 0.951318555 | 1.018156903 |
| MAGI2 | 0.951318242 | 0.859182349 |
| PTOV1 | 0.951316334 | 0.921891059 |
| TMEM190 | 0.951285297 | 1.005138746 |
| ATG101 | 0.951280458 | 1.038882139 |
| TMEM47 | 0.951269484 | 0.936622131 |
| POP4 | 0.951222743 | 0.89145403 |
| ZFAND4 | 0.951198258 | 0.972593354 |
| PAQR6 | 0.951191189 | 1.000342583 |
| CLCN1 | 0.951189982 | 0.943543154 |
| ZSWIM3 | 0.951182041 | 1.012960713 |
| TLCD1 | 0.951110267 | 1.091130012 |
| NUMB | 0.951105192 | 1.057553957 |
| FAT1 | 0.951094906 | 0.878040425 |
| BOLL | 0.951048304 | 0.881894873 |
| HYI | 0.951026406 | 1.078520441 |
| LARP1 | 0.951020255 | 1.004455245 |
| SI | 0.951005414 | 0.915639195 |
| CYTIP | 0.950986027 | 0.873244262 |
| TVP23C | 0.950957395 | 1.113001215 |
| ZNF597 | 0.950942122 | 1.082219938 |
| DEGS1 | 0.950916592 | 0.826650466 |
| SOSTDC1 | 0.950885078 | 1.007194245 |
| RTP5 | 0.950883447 | 0.979445015 |
| GIMAP5 | 0.950861064 | 0.945186708 |
| FBXO31 | 0.950845789 | 1.067488644 |
| REG1A | 0.950834784 | 1.070914697 |
| OR8K5 | 0.950830911 | 1.050222762 |
| TLE2 | 0.950775238 | 0.921479559 |
| TCP10L | 0.950773661 | 1.023638232 |
| KRT222 | 0.950770648 | 0.976508708 |
| SAMD3 | 0.950727717 | 0.982870846 |

| | | |
|---|---|---|
| TSSK3 | 0.950714401 | 0.988968202 |
| VAX2 | 0.950714401 | 0.988968202 |
| NR2E3 | 0.950695961 | 0.758598313 |
| BTF3L4 | 0.950658937 | 0.980963953 |
| TDP1 | 0.950587315 | 1.022276225 |
| HNF1B | 0.950568569 | 0.961522884 |
| SAGE1 | 0.950562881 | 0.909558068 |
| MRPS33 | 0.95054009 | 1.001027749 |
| ENAH | 0.950529714 | 1.0222679 |
| DHX57 | 0.95052618 | 0.87529976 |
| PDE6D | 0.950514165 | 0.894289186 |
| CBY1 | 0.950474533 | 0.942081814 |
| GLRXL_pseudogene | 0.950468456 | 1.051730045 |
| NSMCE2 | 0.950460932 | 0.911705144 |
| ARMCX2 | 0.950436377 | 0.947242206 |
| PEA15 | 0.950315874 | 0.950032446 |
| PROK1 | 0.950301056 | 0.983776768 |
| CPEB4 | 0.95024739 | 0.965399109 |
| ZNF793 | 0.950199966 | 1.046577562 |
| DCXR | 0.950138962 | 0.804023361 |
| IFNA7 | 0.950137696 | 1.024323398 |
| MAMLD1 | 0.950093972 | 0.926344639 |
| TARBP1 | 0.950091616 | 0.972863507 |
| MRPL53 | 0.950067783 | 0.898595409 |
| TAL1 | 0.950067104 | 0.91983556 |
| FAM129A | 0.949970939 | 1.025111381 |
| ST8SIA4 | 0.949959477 | 0.923045768 |
| SPTBN2 | 0.949936929 | 0.972250771 |
| C2CD2L | 0.949934618 | 0.95101062 |
| LARP4B | 0.949934618 | 0.95101062 |
| C19orf80 | 0.94989606 | 1.044193217 |
| MRAS | 0.949889995 | 0.88618874 |
| DDHD1 | 0.949812158 | 0.873228028 |
| ZNF568 | 0.949808522 | 1.081877355 |
| GSTA4 | 0.949765955 | 1.034021871 |
| C3P1 | 0.94970431 | 0.820486468 |
| VCX3A | 0.949696167 | 0.97326853 |
| LRPPRC | 0.949661477 | 0.990684488 |
| OR1E1 | 0.949639472 | 1.000342583 |
| MPP7 | 0.949564538 | 0.834345889 |
| THAP9 | 0.949536981 | 0.99554642 |
| GAB3 | 0.949521259 | 0.917094896 |
| RHEBL1 | 0.949462492 | 0.997259335 |
| CREB3L3 | 0.949452317 | 0.999594978 |

| | | |
|---|---|---|
| PLSCR2 | 0.949452317 | 0.999594978 |
| CHMP2A | 0.949448394 | 0.862697448 |
| PRB1 | 0.949436847 | 0.954779034 |
| PTGFR | 0.949421103 | 0.889033095 |
| ERO1LB | 0.949420128 | 0.969510106 |
| CSAG2 | 0.949401682 | 1.058323208 |
| C21orf58 | 0.949393712 | 0.891058582 |
| TTC17 | 0.949377899 | 0.869818431 |
| CACNG1 | 0.949303875 | 1.063595068 |
| OR13C9 | 0.949273143 | 1.044878383 |
| FAM131C | 0.949272447 | 0.836776023 |
| BUB1B | 0.94919455 | 0.894873459 |
| HIST1H2AH | 0.949155248 | 1.018842069 |
| KCND2 | 0.949138323 | 1.103179753 |
| FAF1 | 0.94912036 | 0.925659472 |
| TM2D2 | 0.949116825 | 0.976361768 |
| BMP1 | 0.949089834 | 1.03569111 |
| PDZD4 | 0.949000987 | 1.052247874 |
| KRTAP27-1 | 0.948904812 | 1.056703119 |
| MTHFR | 0.948885658 | 0.95018226 |
| WIPF1 | 0.948885658 | 0.826245443 |
| CGB1 | 0.948824986 | 0.968482357 |
| SERF1A | 0.948810623 | 0.997944502 |
| MLIP | 0.948797665 | 0.947242206 |
| NR2C2AP | 0.94875598 | 0.9554642 |
| SREK1 | 0.948747099 | 1.065090785 |
| SAFB | 0.9487268 | 0.934224049 |
| INPP4A | 0.948724978 | 1.064243997 |
| RBPMS2 | 0.948690598 | 0.927714971 |
| GCNT7 | 0.948679769 | 0.957177115 |
| MBIP | 0.948669665 | 0.98663926 |
| GABRA6 | 0.948667105 | 1.013627515 |
| TSTA3 | 0.948667105 | 1.013627515 |
| OR4F16 | 0.948667023 | 1.01782098 |
| WDR49 | 0.94866305 | 0.817739976 |
| SPHK2 | 0.948646606 | 0.996755354 |
| C5orf45 | 0.948645271 | 1.093965168 |
| ZFPM1 | 0.948635848 | 1.052652896 |
| FLVCR1 | 0.948630906 | 0.952612394 |
| VASH1 | 0.948630906 | 0.952612394 |
| FBXL17 | 0.948625284 | 0.979883193 |
| ATP6V0E2 | 0.948614555 | 0.970028352 |
| CWC22 | 0.948562327 | 0.813292223 |
| PMVK | 0.94851777 | 1.0532122 |

| | | |
|---|---|---|
| GAGE2C | 0.948517379 | 1.019527235 |
| SIRT4 | 0.948510039 | 1.063720452 |
| MPDZ | 0.948480815 | 1.027749229 |
| GRID2 | 0.9484529 | 0.861778066 |
| PRSS57 | 0.9484529 | 0.861778066 |
| IDH3B | 0.948424333 | 0.844905905 |
| NUP43 | 0.94842087 | 0.896882494 |
| NUP93 | 0.948413666 | 0.985268928 |
| ZNF284 | 0.948379638 | 0.91983556 |
| CCDC148 | 0.948373748 | 1.081877355 |
| PGD | 0.948355594 | 1.075924724 |
| FSD2 | 0.948343502 | 1.007290401 |
| CLSPN | 0.9483404 | 0.824940048 |
| KDELC2 | 0.94832282 | 1.042122317 |
| SPACA4 | 0.948303563 | 0.980472765 |
| SPHAR | 0.948273342 | 1.077081192 |
| C17orf98 | 0.948262694 | 0.929770469 |
| C5AR2 | 0.9482573 | 0.884490591 |
| SYTL5 | 0.948190108 | 0.960945529 |
| DTNBP1 | 0.948187797 | 0.946214457 |
| GLYCTK | 0.948185536 | 1.040097205 |
| EDNRB | 0.948167238 | 0.957819598 |
| PITHD1 | 0.948165211 | 0.870392872 |
| ATG13 | 0.948145252 | 0.924974306 |
| SCAI | 0.948122098 | 0.992120589 |
| VWC2 | 0.948105357 | 0.881328473 |
| ATP5G3 | 0.94810459 | 0.924075276 |
| MGC119295 | 0.947948835 | 0.921891059 |
| CDCA4 | 0.947938459 | 1.167523124 |
| FCER1A | 0.947930229 | 0.9467878 |
| MARCH3 | 0.947816376 | 1.093445814 |
| FAM21C | 0.947802756 | 0.67146283 |
| TCP10 | 0.947802756 | 0.940047962 |
| UGT1A9 | 0.947800323 | 0.986372485 |
| TMEM74B | 0.947763201 | 1.000685166 |
| STXBP2 | 0.947749382 | 1.038369305 |
| POSTN | 0.947732617 | 1.128468654 |
| CD1E | 0.947724171 | 0.923045768 |
| EIF4E2 | 0.947724171 | 1.062373431 |
| MYO1A | 0.947724171 | 0.888213852 |
| SERP2 | 0.947724171 | 0.940461725 |
| GOLPH3 | 0.947719568 | 1.046591298 |
| PRKACA | 0.947680461 | 1.025957171 |
| POU3F2 | 0.947671907 | 0.916565411 |

| | | |
|---|---|---|
| ACVR1B | 0.947605547 | 1.082414017 |
| HSPB7 | 0.947591568 | 1.012332991 |
| CRYBG3 | 0.947584047 | 1.050017129 |
| STIP1 | 0.947566603 | 1.072970195 |
| TULP4 | 0.947532197 | 1.048669695 |
| CD101 | 0.947509558 | 0.999314834 |
| DSPP | 0.947509458 | 1.117051438 |
| MT1X | 0.947446034 | 0.887975334 |
| RBBP4 | 0.947422402 | 0.953827461 |
| KIF5B | 0.947404929 | 0.901579587 |
| FOXRED2 | 0.947378185 | 0.831915755 |
| FAM150A | 0.947340599 | 1.010962658 |
| LGI2 | 0.947327912 | 1.033915725 |
| UCN3 | 0.947278706 | 0.952038369 |
| LIM2 | 0.947238969 | 0.983213429 |
| CCDC184 | 0.947211647 | 0.991435423 |
| DNAJC16 | 0.947201634 | 1.014388489 |
| AMOTL1 | 0.947197647 | 0.916067146 |
| CALB1 | 0.947192028 | 1.037341555 |
| MYLK2 | 0.947177219 | 0.987021415 |
| C2orf43 | 0.947165154 | 0.858242203 |
| AP1S3 | 0.947159043 | 0.947242206 |
| XG | 0.947158001 | 1.068516615 |
| MYLK3 | 0.947123709 | 1.116807268 |
| C3orf30 | 0.947118034 | 0.949777238 |
| C14orf1 | 0.947112419 | 0.940733128 |
| ELOF1 | 0.947097709 | 0.995544755 |
| F2R | 0.947081395 | 0.953277093 |
| MERTK | 0.947081395 | 1.0266061 |
| FOXD4L6 | 0.947011111 | 1.143782908 |
| REST | 0.946993638 | 0.919400567 |
| IL17RC | 0.946990123 | 1.045767517 |
| TRIM43 | 0.946978573 | 0.919532771 |
| DAP | 0.946959318 | 0.967112025 |
| WRB | 0.946954878 | 0.990065091 |
| GGA2 | 0.946951253 | 0.883521754 |
| P4HTM | 0.94692424 | 0.90266061 |
| PTPN21 | 0.94692424 | 0.90266061 |
| ERGIC1 | 0.94689782 | 0.987039287 |
| F9 | 0.946884148 | 0.959117456 |
| APBB2 | 0.946867159 | 0.931140802 |
| ERMAP | 0.946791899 | 0.955806783 |
| OR7D2 | 0.946790268 | 1.024665982 |
| ZMYM6 | 0.946740924 | 1.122316727 |

| | | |
|---|---|---|
| ATP5D | 0.946739884 | 0.834532374 |
| MAPKAPK3 | 0.946717114 | 1.16807268 |
| TRIM23 | 0.946717114 | 1.038286827 |
| FAM53B | 0.946570529 | 1.039692183 |
| KLHL17 | 0.94656029 | 0.948157149 |
| GOLGA5 | 0.946554855 | 0.987670344 |
| CTIF | 0.946536081 | 1.100376841 |
| RP13-36C9.6 | 0.94646712 | 1.002055498 |
| TWISTNB | 0.946459463 | 0.90202124 |
| MMP28 | 0.946449235 | 0.897469176 |
| ALG13 | 0.946445369 | 0.712839206 |
| BSPRY | 0.946436808 | 0.910243234 |
| CAPNS2 | 0.946429527 | 0.741190765 |
| DPPA2 | 0.946429527 | 0.963547995 |
| GTF2A2 | 0.94636757 | 0.993510707 |
| ERP29 | 0.946355307 | 0.911300122 |
| PTCHD4 | 0.946329288 | 0.951353203 |
| GNS | 0.946328563 | 0.893884164 |
| RRBP1 | 0.946327974 | 0.843096951 |
| EMC10 | 0.946302057 | 1.044878383 |
| BAG5 | 0.946298457 | 0.77399757 |
| OR4N5 | 0.94629517 | 0.882494005 |
| SLC22A2 | 0.946292447 | 0.904819765 |
| ETF1 | 0.946184402 | 0.965978129 |
| KL | 0.946178885 | 0.942894225 |
| CYP2F1 | 0.946152514 | 0.97691373 |
| LY6G5C | 0.94614003 | 0.885919836 |
| FAM71E1 | 0.946116696 | 0.925316889 |
| FAM217A | 0.946108223 | 1.018842069 |
| LAMA1 | 0.946059334 | 0.981157931 |
| TMEM98 | 0.946058784 | 1.094775213 |
| KIAA0753 | 0.946031837 | 0.878898339 |
| ZFP3 | 0.946031837 | 0.992304577 |
| ZNF573 | 0.946007047 | 0.974648852 |
| PRDM4 | 0.945999795 | 0.9181855 |
| SNRPN | 0.945991913 | 1.045220966 |
| DNAJB5 | 0.945988696 | 0.951695786 |
| CFLAR | 0.945968714 | 0.892277742 |
| SNX22 | 0.945929612 | 1.078108941 |
| PPDPF | 0.945907909 | 1.023980815 |
| BUD13 | 0.945907899 | 0.950992305 |
| CYB5D1 | 0.945904079 | 0.922640745 |
| METTL21B | 0.945904079 | 0.922640745 |
| ZNF253 | 0.945896219 | 0.930455635 |

| | | |
|---|---|---|
| PDZD9 | 0.945873973 | 0.933576347 |
| LSM1 | 0.945813892 | 0.971565605 |
| ZFP30 | 0.945807598 | 1.051437829 |
| PABPC4L | 0.945798215 | 0.979787598 |
| MYOZ2 | 0.945774291 | 0.886262419 |
| HTRA1 | 0.945733352 | 0.841661259 |
| HTR3E | 0.94572482 | 0.959902795 |
| FAM24B | 0.94572001 | 0.840356286 |
| NDE1 | 0.945701161 | 0.738609113 |
| PROK2 | 0.945685031 | 1.155532717 |
| LMAN2L | 0.945678099 | 0.935594382 |
| NUF2 | 0.945658649 | 0.789793439 |
| ATAD3B | 0.945646065 | 0.991435423 |
| TBC1D29 | 0.945591603 | 0.866735183 |
| HOMER2 | 0.945561019 | 0.883179171 |
| ZNFX1 | 0.9455352 | 1.039584685 |
| ANKRD20A3 | 0.945506829 | 0.9554642 |
| ASAP2 | 0.945500938 | 1.042480301 |
| DLX6 | 0.94549445 | 0.983127839 |
| TMEM14C | 0.945493508 | 0.963686194 |
| SORD | 0.945482513 | 0.887086308 |
| OR2A4 | 0.945481002 | 1.098321343 |
| POU4F2 | 0.945475598 | 1.118754056 |
| SLC46A3 | 0.9454718 | 0.894694208 |
| NPR3 | 0.945463496 | 1.022712524 |
| DPM3 | 0.945454949 | 0.988352175 |
| RILPL2 | 0.945424818 | 1.052415211 |
| CYB5R2 | 0.945417889 | 0.966255678 |
| EXOC6 | 0.945372314 | 0.938436614 |
| IL1B | 0.945338447 | 0.8137573 |
| SLC25A19 | 0.945338447 | 0.949383517 |
| GSTT2 | 0.945302679 | 1.010530579 |
| PLA2G2A | 0.945256289 | 0.932511356 |
| PERP | 0.945249108 | 0.945871874 |
| STK17B | 0.945242211 | 0.79688514 |
| ZNF517 | 0.945222647 | 0.914696814 |
| MCEE | 0.945192653 | 0.87605451 |
| STX3 | 0.945191782 | 0.947347104 |
| TCTA | 0.945184632 | 0.995203837 |
| F10 | 0.945171002 | 0.915639195 |
| NAT9 | 0.94505511 | 0.844876468 |
| FSCB | 0.945044005 | 1.074119077 |
| TMUB1 | 0.945034988 | 1.034831916 |
| PYROXD2 | 0.94498604 | 0.936279548 |

| | | |
|---|---|---|
| HUS1B | 0.944974985 | 1.072627612 |
| DDI2 | 0.94494569 | 0.949777238 |
| BAGE3 | 0.94492605 | 0.993833505 |
| FAM13A | 0.944875749 | 0.943296881 |
| C11orf73 | 0.944874068 | 1.043742406 |
| PPP2CA | 0.944863936 | 1.023361454 |
| SGSH | 0.944840855 | 0.965168084 |
| GPR26 | 0.944783342 | 0.966904607 |
| UBLCP1 | 0.944776509 | 1.046073978 |
| AGPAT6 | 0.944723897 | 0.991494532 |
| ZSCAN31 | 0.944697735 | 0.951353203 |
| FABP2 | 0.944692786 | 0.989617132 |
| GABPA | 0.944692786 | 0.870863076 |
| NEK8 | 0.944692786 | 0.989617132 |
| MFAP5 | 0.944638198 | 1.033573141 |
| STAM | 0.944600455 | 0.994176088 |
| IDH2 | 0.944585272 | 0.81440623 |
| LRRC28 | 0.944584688 | 1.018842069 |
| ANGEL2 | 0.944547796 | 1.072498987 |
| MBLAC2 | 0.944536206 | 0.882494005 |
| ALDH1L2 | 0.944516779 | 0.995457495 |
| CLCN7 | 0.944509535 | 0.95587281 |
| TBC1D2 | 0.944486275 | 0.964713943 |
| FGF3 | 0.944473148 | 0.797534069 |
| SDR42E1 | 0.94442433 | 0.978585334 |
| RBM24 | 0.944363189 | 1.042122317 |
| B4GALNT4 | 0.944360693 | 0.970028352 |
| C11orf40 | 0.944345513 | 0.908869988 |
| ERAP1 | 0.944328506 | 0.961713173 |
| SSTR3 | 0.944312947 | 0.922128488 |
| TSPAN18 | 0.944310927 | 1.075025694 |
| SCO2 | 0.94430549 | 1.091469681 |
| ARSK | 0.944294827 | 0.797488862 |
| VDAC1 | 0.944265199 | 1.024010383 |
| BNIP1 | 0.944229306 | 0.944841012 |
| HSPA12A | 0.944201278 | 1.029119561 |
| WBP2 | 0.94417817 | 0.849948613 |
| DAB2IP | 0.944169697 | 0.882836588 |
| VNN1 | 0.94411957 | 1.069435432 |
| ZNF180 | 0.944097564 | 0.876327509 |
| MKNK1 | 0.944073716 | 0.990266061 |
| MAGIX | 0.944054339 | 0.745240988 |
| CAPNS1 | 0.944051241 | 1.092147956 |
| MARCH8 | 0.944047948 | 0.950681376 |

| | | |
|---|---|---|
| KLHDC8A | 0.944032791 | 0.987849332 |
| MCF2 | 0.944027121 | 1.052563271 |
| RERGL | 0.944018454 | 0.943816375 |
| PCDHGB1 | 0.944011476 | 1.050702295 |
| OTUD3 | 0.943978947 | 0.8327258 |
| DUT | 0.943932032 | 0.898339409 |
| GPR146 | 0.94391903 | 0.792342635 |
| MRAP | 0.943885561 | 1.004796163 |
| SLU7 | 0.943870608 | 0.815690305 |
| IRF5 | 0.94385068 | 0.891859052 |
| MID1 | 0.943833167 | 0.713173264 |
| LILRB5 | 0.943752773 | 0.9181855 |
| PDLIM7 | 0.943682891 | 0.96882494 |
| ANKRD1 | 0.943675024 | 0.860480208 |
| LRIG3 | 0.943668573 | 0.919492977 |
| CNBP | 0.943639176 | 1.010125557 |
| ITFG2 | 0.94357545 | 0.959902795 |
| FERMT1 | 0.943553189 | 1.058323208 |
| BHLHE23 | 0.943550768 | 0.972250771 |
| CHADL | 0.943550768 | 0.972250771 |
| CNP | 0.943532687 | 0.905905256 |
| ARL16 | 0.94350663 | 0.898744431 |
| AQP7 | 0.943503051 | 1.030499676 |
| EHD1 | 0.943496079 | 1.140116478 |
| ADO | 0.943442206 | 0.891743748 |
| ITGA1 | 0.943422768 | 0.858855773 |
| SLC22A15 | 0.943412085 | 1.012555691 |
| CCDC28A | 0.943336592 | 0.907654921 |
| LRRC6 | 0.943333599 | 0.944501542 |
| N4BP2L2 | 0.943320278 | 0.719081877 |
| MLF2 | 0.943289332 | 0.979102432 |
| C21orf2 | 0.943281221 | 0.870503597 |
| LOX | 0.943224798 | 0.894873459 |
| FAM170A | 0.943212169 | 0.96437136 |
| PITPNB | 0.943199392 | 0.947927372 |
| CLEC12B | 0.943192227 | 1.05872823 |
| ABCC4 | 0.943169865 | 0.916565411 |
| ALMS1 | 0.943153312 | 0.974306269 |
| NPM2 | 0.943109679 | 1.000685166 |
| FRA10AC1 | 0.94305055 | 0.934909215 |
| GAPVD1 | 0.943050257 | 1.06763872 |
| RASGRP3 | 0.942991602 | 1.079821857 |
| CCDC106 | 0.942983831 | 1.083029567 |
| YAP1 | 0.942975112 | 1.07073329 |

| | | |
|---|---|---|
| TMED8 | 0.942951005 | 0.971223022 |
| MRPL33 | 0.942935464 | 0.701267557 |
| DERA | 0.942920869 | 0.918995545 |
| DNAJC2 | 0.942895274 | 0.981157931 |
| SPATA25 | 0.942849647 | 0.999314834 |
| TTC40 | 0.942839362 | 0.931825968 |
| BLOC1S3 | 0.942796409 | 1.009249743 |
| S100B | 0.942792245 | 0.982584042 |
| MAD2L1BP | 0.942640679 | 0.988009592 |
| ELK1 | 0.942632318 | 1.105126541 |
| BMX | 0.942593215 | 1.042829332 |
| CACNA1E | 0.942589757 | 0.957819598 |
| FADS2 | 0.942561438 | 1.15055159 |
| ZNF205 | 0.942547342 | 0.922233642 |
| SLC39A12 | 0.942514366 | 1.022276225 |
| KLB | 0.942510397 | 1.003244646 |
| DPEP2 | 0.942485536 | 1.110966905 |
| ELMO2 | 0.94248409 | 0.838300788 |
| GLTP | 0.942443509 | 1.02673147 |
| ARRDC3 | 0.94243955 | 0.984926345 |
| TTC40 | 0.94243955 | 0.899280576 |
| OR4K2 | 0.942433881 | 1.015795869 |
| AIMP2 | 0.942403937 | 1.045906132 |
| C9orf89 | 0.942367236 | 0.858170606 |
| UBL5 | 0.942349456 | 1.00908501 |
| IGSF21 | 0.9423455 | 1.079384366 |
| AVPR1A | 0.942344598 | 0.708630759 |
| IL4 | 0.942323524 | 0.924075276 |
| ISX | 0.942292624 | 1.024706359 |
| ADAM18 | 0.942284338 | 1.054510058 |
| RFPL1 | 0.942253796 | 1.077222583 |
| STK32C | 0.942221361 | 0.992212849 |
| SFMBT1 | 0.942195175 | 0.937221547 |
| CENPF | 0.942160443 | 1.156560466 |
| C1orf200 | 0.942146578 | 0.932511134 |
| GATS | 0.942132487 | 0.889688249 |
| RAB11FIP5 | 0.942098287 | 1.049007695 |
| MATN3 | 0.942035207 | 1.01336074 |
| ASB8 | 0.942007073 | 0.782608696 |
| ENO3 | 0.941913389 | 0.850746269 |
| ZNF543 | 0.941902269 | 0.947584789 |
| SRSF11 | 0.941866157 | 0.931140802 |
| GPR135 | 0.941863995 | 0.74302401 |
| PIEZO1 | 0.94186004 | 0.835560123 |

| | | |
|---|---|---|
| TMEM117 | 0.94184053 | 0.970433374 |
| CCDC181 | 0.941832481 | 1.066828676 |
| SST | 0.941806043 | 0.948562171 |
| NCL | 0.941794523 | 1.020212402 |
| TMEM81 | 0.941788163 | 0.937626569 |
| SPATA18 | 0.941772015 | 0.830295666 |
| KCNIP2 | 0.941723881 | 1.032446463 |
| XAB2 | 0.941716185 | 0.990279465 |
| FLOT2 | 0.941680614 | 0.883521754 |
| KIF6 | 0.941655821 | 1.091130012 |
| SS18L2 | 0.94163757 | 0.867077766 |
| TNFSF13B | 0.941615323 | 1.021061158 |
| GIMAP6 | 0.941608618 | 0.911613566 |
| THEMIS2 | 0.941587367 | 0.903391572 |
| ZBTB11 | 0.941580208 | 0.982528263 |
| IL20RB | 0.941537663 | 1.018842069 |
| CIDEC | 0.941509317 | 1.0036452 |
| CWC25 | 0.941497193 | 0.95544755 |
| FAM90A1 | 0.941468616 | 0.859052248 |
| USP37 | 0.941462505 | 0.975989617 |
| ZNF701 | 0.941461025 | 0.896314297 |
| DOCK5 | 0.941371241 | 0.953066119 |
| NOL6 | 0.941221175 | 1.019184652 |
| TMEM130 | 0.941192673 | 0.962332928 |
| ABR | 0.941192494 | 1.055498458 |
| PDE4B | 0.941172449 | 0.988659376 |
| SOD1 | 0.941160873 | 0.942245295 |
| FUT2 | 0.941152328 | 0.839611179 |
| MPZL3 | 0.941136893 | 0.903199676 |
| KRTAP3-1 | 0.941106497 | 0.978074683 |
| PEAK1 | 0.941030308 | 0.789097988 |
| MSX2 | 0.941024957 | 0.811810513 |
| SSR1 | 0.941020861 | 0.971565605 |
| ZBTB38 | 0.940994261 | 1.07654921 |
| MRPL49 | 0.940982258 | 1.133264817 |
| IRF2BPL | 0.940981714 | 0.936964714 |
| MAPK4 | 0.940977704 | 1.084360805 |
| OR2M2 | 0.940908176 | 0.956834532 |
| ZNF273 | 0.940889146 | 0.993148338 |
| PCNP | 0.940855028 | 1.003083248 |
| VPS4A | 0.940838717 | 1.039397054 |
| GPR19 | 0.940836931 | 1.022063595 |
| HMGCS1 | 0.940836931 | 0.953926022 |
| ROBO1 | 0.940836931 | 1.044776119 |

| | | |
|---|---|---|
| MZT1 | 0.940803421 | 0.98663926 |
| ZNF285 | 0.940787036 | 0.899149453 |
| GLIPR1 | 0.940750091 | 0.970195272 |
| U2AF2 | 0.940750091 | 0.970195272 |
| PNMAL1 | 0.940732428 | 0.850951802 |
| ZBTB4 | 0.940730835 | 1.024665982 |
| ZNF177 | 0.940723463 | 0.87727825 |
| ACSL5 | 0.940685045 | 1.005191434 |
| METTL13 | 0.940685045 | 1.005191434 |
| MYL10 | 0.940683667 | 0.990065091 |
| ZFAND2B | 0.940679348 | 0.776427704 |
| CNOT7 | 0.940662742 | 0.971648441 |
| C8orf74 | 0.940656731 | 0.997974889 |
| ZNF619 | 0.940642164 | 1.036313806 |
| ARFGEF1 | 0.940601155 | 0.949777238 |
| TM4SF18 | 0.940581947 | 1.056183625 |
| STAU2 | 0.940548328 | 0.912641316 |
| MLPH | 0.940539266 | 0.967112025 |
| BEST2 | 0.940496117 | 1.022276225 |
| NTN4 | 0.940483943 | 0.914354231 |
| SPATA6 | 0.940451184 | 0.92428914 |
| PA2G4 | 0.94044746 | 0.783906554 |
| BIRC2 | 0.940423504 | 1.107722258 |
| NAA10 | 0.94039821 | 0.884163629 |
| ABCF2 | 0.94037625 | 0.994159637 |
| MIPEP | 0.94037625 | 0.994159637 |
| NPY5R | 0.940365548 | 0.971447112 |
| CDK2AP2 | 0.940345334 | 1.07290401 |
| SELM | 0.940317838 | 0.836930456 |
| FOXR1 | 0.940266502 | 0.965741692 |
| FAM104A | 0.940212533 | 1.002835156 |
| ADAM7 | 0.940165338 | 0.909149903 |
| FRG1 | 0.940161248 | 0.904761905 |
| ZNF17 | 0.940137234 | 1.051730045 |
| DNAJB12 | 0.940131661 | 0.914696814 |
| IPO5 | 0.940130021 | 0.863724854 |
| TNFAIP2 | 0.940113537 | 0.960945529 |
| TGFB3 | 0.940038215 | 1.042527339 |
| XPNPEP2 | 0.93998151 | 0.892277742 |
| GLB1L2 | 0.939963064 | 0.827338129 |
| ITGAD | 0.939957355 | 1.020554985 |
| IL21 | 0.939917527 | 1.058581706 |
| LRRFIP1 | 0.939909326 | 1.040424803 |
| ANKRD34B | 0.9398938 | 0.998784933 |

| | | |
|---|---|---|
| HDAC4 | 0.939889061 | 0.966255678 |
| ARL2 | 0.939882186 | 0.862697448 |
| ERRFI1 | 0.939870724 | 1.01417578 |
| VSIG2 | 0.939870724 | 1.01417578 |
| GRK7 | 0.939817193 | 0.898118105 |
| PSMA5 | 0.939817193 | 0.898118105 |
| CCDC3 | 0.939788259 | 1.025693731 |
| SAMD10 | 0.939781528 | 0.924260834 |
| BFSP2 | 0.939776841 | 0.965978129 |
| CPEB2 | 0.939754549 | 0.971223022 |
| IL21R | 0.939733491 | 0.996759822 |
| TUFT1 | 0.93967272 | 0.982870846 |
| CRISPLD2 | 0.939636563 | 0.974648852 |
| DENND5B | 0.939618076 | 1.179513532 |
| PI4K2A | 0.939616654 | 1.000648929 |
| VILL | 0.939585952 | 1.022610483 |
| JAK3 | 0.939558533 | 0.955223881 |
| PRR14L | 0.939554131 | 1.010125557 |
| SPARC | 0.939486228 | 1.071688943 |
| BCL3 | 0.939406099 | 1.04981774 |
| SERPINB10 | 0.939404895 | 0.953408702 |
| TRA2B | 0.939404895 | 0.953408702 |
| PPP1R1A | 0.939372977 | 0.93835172 |
| PHIP | 0.939363663 | 1.029462145 |
| CXCL12 | 0.939345267 | 1.039397054 |
| BTBD16 | 0.939293164 | 1.001620089 |
| OR2M7 | 0.939280157 | 0.872559096 |
| FLAD1 | 0.93922389 | 1.040882544 |
| WDR52 | 0.939222154 | 1.021466181 |
| ZNF833P | 0.939208432 | 0.968482357 |
| TEX28 | 0.939202898 | 0.979748886 |
| C8orf86 | 0.939086504 | 0.877697842 |
| BNC1 | 0.939009733 | 0.93600648 |
| KCNJ3 | 0.939007626 | 1.001297859 |
| USHBP1 | 0.938961042 | 0.985268928 |
| SOX12 | 0.938930956 | 1.005138746 |
| TPCN1 | 0.938920693 | 0.95587281 |
| GPN3 | 0.938897359 | 0.929526124 |
| HNRNPL | 0.938891175 | 0.958890031 |
| VIMP | 0.938852532 | 0.903199676 |
| SEPN1 | 0.938793125 | 1.028434395 |
| CHST4 | 0.938714391 | 0.791044776 |
| CA11 | 0.938706232 | 0.973673552 |
| TAS2R50 | 0.93870391 | 0.973963686 |

| | | |
|---|---|---|
| NDN | 0.938690589 | 1.069886948 |
| CEP57L1 | 0.938585607 | 1.121274409 |
| EFS | 0.938580758 | 0.891401165 |
| TBC1D9 | 0.938542051 | 0.956257594 |
| VPS13B | 0.938534156 | 1.002430134 |
| CNOT4 | 0.938514346 | 0.722907203 |
| DTWD1 | 0.938496766 | 0.86836776 |
| PROM1 | 0.938495214 | 0.912983899 |
| LCN10 | 0.938483796 | 0.922918808 |
| GSDMC | 0.938451128 | 0.846865365 |
| CLK1 | 0.938428647 | 0.950681376 |
| SEC24D | 0.938423768 | 1.042122317 |
| APCS | 0.938373286 | 1.032202809 |
| RUSC1 | 0.938372425 | 1.147995889 |
| GYS2 | 0.93834937 | 1.104477612 |
| ZNF490 | 0.938335588 | 0.956149366 |
| TSTD1 | 0.938330514 | 1.081877355 |
| C19orf54 | 0.93831766 | 1.004860267 |
| OR4F21 | 0.938291366 | 1.131551901 |
| EPHX3 | 0.938221604 | 0.933809215 |
| BAIAP2L2 | 0.938213436 | 1.029161604 |
| HSCB | 0.938173397 | 1.095180235 |
| ATP2A2 | 0.938144616 | 0.956662616 |
| ZNF432 | 0.938135003 | 1.001027749 |
| ALG6 | 0.938111097 | 0.833535844 |
| PAX3 | 0.938098006 | 0.991899554 |
| C16orf90 | 0.938038901 | 1.072285029 |
| CDON | 0.938037859 | 0.984583762 |
| GCSH | 0.937999249 | 0.968202466 |
| OR2AT4 | 0.937988109 | 0.878725591 |
| TAS2R19 | 0.937964412 | 0.918465228 |
| METTL20 | 0.937958794 | 0.928400137 |
| NPR1 | 0.93793973 | 0.842959117 |
| OR6C6 | 0.937922682 | 0.997944502 |
| SIGLECL1 | 0.937872524 | 0.989722508 |
| ZP1 | 0.937821641 | 0.981500514 |
| NDUFV2 | 0.937788419 | 0.951330305 |
| NPC2 | 0.937769898 | 0.97327852 |
| FAM132A | 0.937683047 | 0.946132037 |
| PDC | 0.937658349 | 1.00843608 |
| ST8SIA2 | 0.937647647 | 0.957170668 |
| PRL | 0.937624681 | 0.869133265 |
| TNFRSF10A | 0.937622209 | 1.139519792 |
| HRASLS | 0.937585081 | 0.985723556 |

| | | |
|---|---|---|
| VN1R10P | 0.937570098 | 0.934458144 |
| HOMER3 | 0.937560115 | 0.890715999 |
| PABPC5 | 0.93754702 | 1 |
| C9orf64 | 0.937496092 | 0.952038369 |
| HDAC10 | 0.93748871 | 1.045362495 |
| APOL1 | 0.937470487 | 1.042829332 |
| MGAT1 | 0.937448875 | 0.88254354 |
| CDC42EP2 | 0.937439908 | 1.053100377 |
| TCTE1 | 0.937392332 | 0.886998785 |
| OR8J1 | 0.937383316 | 0.945529291 |
| STXBP6 | 0.93738019 | 0.895854745 |
| LZTS2 | 0.937367167 | 0.972863507 |
| KLHL32 | 0.937318112 | 0.876063183 |
| PMS1 | 0.937317201 | 0.997404283 |
| PARVG | 0.937282276 | 1.018499486 |
| OR1C1 | 0.937263291 | 0.909215485 |
| JUN | 0.93722591 | 0.90036452 |
| GTF2A1 | 0.937218249 | 1.054510058 |
| DERL2 | 0.937178383 | 0.95544755 |
| MAMDC4 | 0.937135899 | 1.010530579 |
| NASP | 0.937099769 | 0.944158959 |
| STIM1 | 0.937092429 | 0.92428914 |
| PGS1 | 0.937073854 | 0.878493317 |
| ARF3 | 0.937059118 | 0.92926671 |
| CHMP1A | 0.937055413 | 1.00908501 |
| COL4A2 | 0.937047935 | 0.955806783 |
| EIF2B5 | 0.937026734 | 1.089914945 |
| MARCH11 | 0.937012276 | 0.957819598 |
| TREH | 0.937012276 | 0.878001298 |
| ARHGAP32 | 0.937010509 | 0.997259335 |
| UTP14C | 0.936971679 | 1.028776978 |
| ABAT | 0.936926742 | 1.014925373 |
| SH3PXD2B | 0.9368759 | 1.052652896 |
| TBC1D32 | 0.936858314 | 1.012332991 |
| TDRD1 | 0.936832714 | 0.881466256 |
| CST11 | 0.936808791 | 0.932853717 |
| OPA1 | 0.936804482 | 0.900769542 |
| C19orf73 | 0.936752607 | 0.934566632 |
| KRTAP8-1 | 0.936748121 | 1.005823912 |
| AFF3 | 0.936724251 | 0.889833941 |
| HMGCL | 0.936698295 | 1.1064244 |
| CDHR5 | 0.936689561 | 0.839206156 |
| PHF7 | 0.936673845 | 1.0266061 |
| WDFY2 | 0.936627824 | 0.856800274 |

| | | |
|---|---|---|
| CLEC12A | 0.93662777 | 0.933981369 |
| CCL24 | 0.936584372 | 1.001027749 |
| MEF2A | 0.936529048 | 0.942891859 |
| DSTYK | 0.936510763 | 0.952628164 |
| RASGRP4 | 0.936464846 | 0.9331963 |
| GIYD2 | 0.936434147 | 0.89671932 |
| WDR73 | 0.93643216 | 0.951802349 |
| ABHD17C | 0.936426711 | 1.038286827 |
| FBXL4 | 0.936426711 | 1.038286827 |
| EI24 | 0.936420216 | 0.944844125 |
| TBC1D22B | 0.936417897 | 0.861482382 |
| RAD23B | 0.936405225 | 0.929915639 |
| WHSC1L1 | 0.936405225 | 0.929915639 |
| CDS1 | 0.936352291 | 0.987021415 |
| TEX10 | 0.936304178 | 0.989722508 |
| MTG2 | 0.936271495 | 1.059943297 |
| FAM71F2 | 0.936253431 | 0.857142857 |
| HLA-DPA1 | 0.936233042 | 1.064405618 |
| ARG1 | 0.936204438 | 1.021414666 |
| HCST | 0.936148721 | 0.870503597 |
| FOXF1 | 0.936132173 | 0.848521669 |
| SEPT4 | 0.936117011 | 0.970149254 |
| GNE | 0.936117011 | 0.970149254 |
| ARHGEF19 | 0.936069882 | 0.976704351 |
| HP1BP3 | 0.936038087 | 0.837586067 |
| AHR | 0.936036663 | 1.112913692 |
| MAPK1IP1L | 0.936031278 | 0.873929428 |
| CELA3B | 0.936017595 | 0.978417266 |
| HDGFL1 | 0.936010528 | 1.011745646 |
| APOA5 | 0.935972829 | 0.948612539 |
| RUSC2 | 0.935888417 | 1.034943474 |
| PHTF2 | 0.935873194 | 1.064803564 |
| KRT80 | 0.935862241 | 0.930336168 |
| ANPEP | 0.935862221 | 0.981829981 |
| CCDC82 | 0.93584864 | 0.989874443 |
| TMEM53 | 0.93584864 | 0.989874443 |
| BSCL2 | 0.935832798 | 1.069258809 |
| DNAJB14 | 0.935819961 | 0.914945322 |
| CNN1 | 0.935758242 | 1.018499486 |
| ALG10 | 0.935752154 | 0.930564568 |
| VAMP1 | 0.935741659 | 0.977046934 |
| STX16 | 0.935703871 | 0.894141829 |
| ZCWPW1 | 0.93566717 | 0.905789654 |
| PHOSPHO1 | 0.935661552 | 1.05344296 |

| | | |
|---|---|---|
| DAPP1 | 0.935650678 | 0.927372388 |
| MRPL9 | 0.935644229 | 0.793640493 |
| MUC15 | 0.935627388 | 0.885919836 |
| HKDC1 | 0.935617062 | 0.96495782 |
| GSDMD | 0.935615925 | 0.939020212 |
| TMEM66 | 0.935553442 | 1.021061158 |
| CHGA | 0.935523539 | 1.005481329 |
| IGFALS | 0.935508949 | 0.942446043 |
| PITPNM2 | 0.93549119 | 0.913692408 |
| SNAP25 | 0.93549119 | 0.856586632 |
| ZNF35 | 0.93549119 | 0.913692408 |
| RCVRN | 0.935469238 | 1.010125557 |
| DEFB4 | 0.935447237 | 0.859540939 |
| FBRS | 0.935419979 | 0.974888619 |
| DVL3 | 0.935413074 | 0.92428914 |
| CRYBA4 | 0.935401691 | 1.078574322 |
| PRSS54 | 0.935378004 | 0.816119887 |
| SLC6A8 | 0.935350417 | 0.862426995 |
| DEFB107B | 0.935282673 | 0.992463172 |
| EXOC7 | 0.935248192 | 0.854744776 |
| ESRRB | 0.935236646 | 0.953926022 |
| CDY1B | 0.935203698 | 0.909558068 |
| GAA | 0.935133001 | 1.016872161 |
| C16orf86 | 0.935091784 | 0.934566632 |
| C2CD4A | 0.935086423 | 0.842851357 |
| TPX2 | 0.935083417 | 0.990684488 |
| CFB | 0.935029274 | 1.051265412 |
| SCRT2 | 0.934982089 | 1.110654334 |
| TRIM14 | 0.934972965 | 0.937053861 |
| SYNCRIP | 0.934959887 | 1.057553957 |
| TNFSF13 | 0.934912448 | 1.037684138 |
| DDN | 0.934911406 | 1.016101405 |
| OR4L1 | 0.934904338 | 0.886605002 |
| MSH6 | 0.934901015 | 1.028552888 |
| PAQR3 | 0.934864578 | 1.037262049 |
| TMEM91 | 0.934829305 | 0.943131209 |
| PTBP3 | 0.934814897 | 1.019527235 |
| TAS1R3 | 0.93478531 | 0.988009592 |
| PLXND1 | 0.934778489 | 0.98217902 |
| SCGB2A2 | 0.934773121 | 0.923261391 |
| AARD | 0.934749107 | 0.934909215 |
| OR4D11 | 0.934692334 | 1.013018157 |
| TENM3 | 0.934677291 | 0.871873929 |
| NR0B1 | 0.934674214 | 1.040233615 |

| | | |
|---|---|---|
| ASCL2 | 0.934644804 | 0.938335046 |
| MXI1 | 0.934639473 | 0.887403807 |
| C6orf15 | 0.934571774 | 1.028756582 |
| PCDHB14 | 0.934571584 | 0.951695786 |
| MYH7 | 0.934562666 | 1.046073978 |
| REV1 | 0.934561331 | 1.097205346 |
| PRY2 | 0.934510416 | 0.877012676 |
| PAPOLB | 0.934479931 | 0.960415315 |
| XRCC2 | 0.934479931 | 0.960415315 |
| FGF7 | 0.934451351 | 1.066423653 |
| LCOR | 0.934416967 | 1.070878898 |
| PBXIP1 | 0.934405607 | 1.02673147 |
| SLC1A7 | 0.934343154 | 0.781692993 |
| GREB1 | 0.934340868 | 0.91538198 |
| CDX4 | 0.934324103 | 0.898744431 |
| ASS1 | 0.93427791 | 1.000648929 |
| LOC644006_pseudogene | 0.934260293 | 0.789097988 |
| FNDC3B | 0.934213155 | 0.887808829 |
| TOP2A | 0.934169397 | 0.775212637 |
| MAP2K4 | 0.934164304 | 1.006489293 |
| NKD1 | 0.934115634 | 0.933881466 |
| PTCHD1 | 0.93410408 | 0.978759849 |
| TAF2 | 0.934037157 | 1.027136493 |
| GORASP1 | 0.933999233 | 0.948955122 |
| PAPD4 | 0.933978474 | 0.9145403 |
| ACTR3 | 0.933949121 | 0.950668037 |
| ZNF530 | 0.9339416 | 1.040424803 |
| PLOD1 | 0.93393711 | 1.016200891 |
| C12orf4 | 0.933901047 | 0.907502569 |
| GRK5 | 0.933892143 | 0.835171966 |
| GSK3A | 0.933783724 | 0.995457495 |
| CDC23 | 0.933722805 | 1.029850746 |
| SENP5 | 0.933713584 | 1.092796885 |
| VAMP3 | 0.93366281 | 0.892771497 |
| KLHL38 | 0.933661371 | 0.852976914 |
| FRZB | 0.933620264 | 0.882836588 |
| BLID | 0.93361025 | 0.894484412 |
| OR52B4 | 0.93350622 | 0.966084275 |
| SCLT1 | 0.933500859 | 1.025516403 |
| BCAS2 | 0.933492629 | 0.789746918 |
| ELP6 | 0.933488911 | 0.921205892 |
| FAM200A | 0.933458782 | 1.005670312 |
| GPM6B | 0.933450534 | 0.979445015 |
| SDHAF1 | 0.933448594 | 1.05872823 |

| | | |
|---|---|---|
| TFB1M | 0.933446445 | 0.950032446 |
| MORF4L2 | 0.933432138 | 0.854744776 |
| NOXA1 | 0.93342669 | 1.010125557 |
| VWA7 | 0.933423575 | 0.946214457 |
| STUB1 | 0.933404625 | 1.047371836 |
| ZDHHC7 | 0.933391949 | 1.084275437 |
| SLC51B | 0.933390046 | 1.00445358 |
| ZSCAN20 | 0.93336928 | 0.965978129 |
| YTHDC2 | 0.933343739 | 0.914696814 |
| GAS7 | 0.933315716 | 0.892926671 |
| HMBS | 0.933303618 | 0.974888619 |
| BFAR | 0.933258502 | 1.087605451 |
| LPPR3 | 0.933227837 | 0.953066119 |
| FBXO8 | 0.933218739 | 0.961713173 |
| SMAP1 | 0.933218685 | 0.976361768 |
| IAH1 | 0.933218192 | 0.853381936 |
| KIF20A | 0.933218192 | 0.853381936 |
| MAK16 | 0.933204591 | 0.935196436 |
| PDE12 | 0.933186048 | 0.910895099 |
| FAM50B | 0.933154262 | 0.780477926 |
| SH2D1B | 0.933144532 | 0.997164844 |
| OR4C16 | 0.933111528 | 1.026378897 |
| HEATR9 | 0.933084779 | 0.924260834 |
| RBBP8 | 0.933047143 | 0.901678657 |
| BANF1 | 0.933037175 | 0.948269955 |
| MEP1B | 0.932996549 | 0.87605451 |
| EZH1 | 0.932928221 | 1.042180402 |
| EOMES | 0.932902617 | 0.893479141 |
| USH2A | 0.932901518 | 0.801986982 |
| SNAI2 | 0.932880356 | 1.008505468 |
| C7orf65 | 0.932844654 | 1.01336074 |
| CENPH | 0.932757796 | 1.051730045 |
| AP1S1 | 0.932743206 | 0.900308325 |
| ZNF19 | 0.932737678 | 0.853717026 |
| CDCA5 | 0.932728163 | 0.783830079 |
| RGMB | 0.932710175 | 1.018499486 |
| RSPO2 | 0.932707414 | 0.89145403 |
| AQP10 | 0.932698704 | 0.956521739 |
| RNF111 | 0.932673101 | 0.99091499 |
| GIT2 | 0.932644962 | 0.929121102 |
| EVL | 0.932616067 | 0.998629668 |
| HMGB2 | 0.932589461 | 0.962362103 |
| CXCL16 | 0.932561114 | 0.794248684 |
| GDPD5 | 0.932543336 | 0.884973674 |

| | | |
|---|---|---|
| TIGD6 | 0.932500709 | 1.062007537 |
| MIDN | 0.932472073 | 0.980472765 |
| MS4A6E | 0.932467678 | 0.96882494 |
| EIF2AK3 | 0.932447041 | 1.036988968 |
| ALDH8A1 | 0.932435088 | 0.960307817 |
| MICU1 | 0.932417819 | 0.902794654 |
| KEL | 0.932406456 | 0.905256327 |
| TAS2R39 | 0.932396542 | 1.018842069 |
| RUFY4 | 0.932377679 | 0.969218307 |
| STAT5B | 0.932360849 | 1.00843608 |
| SZRD1 | 0.932342683 | 1.03564196 |
| ZNF546 | 0.932314713 | 1.033915725 |
| DIAPH1 | 0.932276336 | 0.964028777 |
| E2F5 | 0.932267699 | 0.925070879 |
| TMEM218 | 0.932258348 | 1.012332991 |
| ACOT1 | 0.932240246 | 1.083062946 |
| SLC39A11 | 0.93217593 | 0.955806783 |
| LRRCC1 | 0.932152727 | 1.100040502 |
| GPRC5B | 0.932146232 | 1.08890331 |
| SAMD5 | 0.932137852 | 1.071283921 |
| NPFFR2 | 0.932135201 | 1.157689812 |
| MAGEA10 | 0.932108328 | 1.054128126 |
| IL22RA1 | 0.932074529 | 0.888384166 |
| TINF2 | 0.932044215 | 0.912641316 |
| C22orf39 | 0.932026997 | 0.949297705 |
| HEY1 | 0.932014985 | 0.965168084 |
| PPP1R26 | 0.931966328 | 0.987667009 |
| GLRX | 0.931961087 | 0.963011032 |
| EVPL | 0.931930986 | 1.049331963 |
| KLHL24 | 0.931911984 | 0.949777238 |
| FBLN5 | 0.93191028 | 1.026036314 |
| PCOLCE2 | 0.931881878 | 1.020656136 |
| CRAMP1L | 0.931874893 | 0.894484412 |
| MGST3 | 0.931865628 | 0.759416768 |
| LPO | 0.931859719 | 0.925475901 |
| NXN | 0.931853244 | 0.934458144 |
| KAZALD1 | 0.931816851 | 1.066118534 |
| PRELID2 | 0.931808218 | 1.034021871 |
| PCSK9 | 0.931761289 | 1.00908501 |
| STK31 | 0.931682094 | 1.083711875 |
| SLC51A | 0.931648753 | 0.878040425 |
| KCTD13 | 0.931629407 | 0.91174562 |
| MRPS6 | 0.931628094 | 0.861482382 |
| HOXC8 | 0.931599139 | 0.953066119 |

| | | |
|---|---|---|
| ORAI3 | 0.93156773 | 1.045362495 |
| ST6GALNAC2 | 0.931537534 | 0.951979234 |
| NPAS3 | 0.93151808 | 0.894826996 |
| IRAK1BP1 | 0.931499412 | 1.018156903 |
| EIF3D | 0.931477566 | 0.921425678 |
| IL36G | 0.931474764 | 0.9331963 |
| SACS | 0.931469689 | 0.896539911 |
| LUZP4 | 0.931460093 | 0.675577157 |
| IYD | 0.931357754 | 0.688942892 |
| SIGLEC9 | 0.931322841 | 0.840013703 |
| CRLF3 | 0.931312827 | 1.01336074 |
| RASSF4 | 0.931309565 | 0.951695786 |
| EGFL7 | 0.931258139 | 0.978417266 |
| CXCL1 | 0.931239018 | 0.966769442 |
| LHX9 | 0.93123349 | 0.991778006 |
| KYNU | 0.931233268 | 0.969500324 |
| IL26 | 0.931199372 | 0.943473792 |
| TMEM79 | 0.931191435 | 0.824220332 |
| IRF4 | 0.931188225 | 0.937221547 |
| MC1R | 0.931146664 | 0.866320571 |
| TSSC4 | 0.931141104 | 1.045220966 |
| HK3 | 0.931124519 | 1.084360805 |
| MRPL57 | 0.931087276 | 0.935251799 |
| NAAA | 0.931036515 | 0.981181051 |
| P2RY2 | 0.931002845 | 0.9467878 |
| MAFK | 0.930824869 | 0.848926691 |
| GLI2 | 0.930749948 | 0.998702141 |
| DCAF7 | 0.930729458 | 0.866747671 |
| CLTA | 0.93070302 | 0.913325233 |
| BFSP1 | 0.930683613 | 1.014045906 |
| CD2 | 0.93067918 | 0.959902795 |
| INTS6 | 0.93067918 | 0.959902795 |
| SLC25A30 | 0.930637171 | 0.72387804 |
| FAM221B | 0.930629332 | 0.927372388 |
| BMF | 0.930626568 | 1.029119561 |
| OR2W1 | 0.930618307 | 1.015390846 |
| SSX2IP | 0.930595622 | 0.954093868 |
| SIN3A | 0.93056752 | 1.085009734 |
| MYOM3 | 0.930555523 | 0.917437479 |
| GTF2E2 | 0.930549894 | 0.902389631 |
| LAMC3 | 0.930539347 | 0.930798219 |
| KCNA1 | 0.930421149 | 0.981829981 |
| POLRMT | 0.930421149 | 0.981829981 |
| EPC2 | 0.930384932 | 1.039397054 |

| | | |
|---|---|---|
| ADRB2 | 0.930383116 | 1.022063595 |
| HTR3E | 0.930383116 | 1.022063595 |
| LACE1 | 0.930340529 | 1.002740665 |
| CDKL4 | 0.930328206 | 0.987670344 |
| LPHN3 | 0.930328206 | 0.987670344 |
| CSTF2T | 0.930299388 | 1.00445358 |
| EFR3A | 0.93029676 | 0.824254882 |
| TTC39B | 0.930292818 | 0.966084275 |
| TINAGL1 | 0.930107992 | 0.924724205 |
| FST | 0.930080995 | 0.974648852 |
| CTAG2 | 0.930048146 | 0.897910243 |
| FGF2 | 0.930036453 | 0.890330954 |
| C6orf211 | 0.930021595 | 0.937992463 |
| BLZF1 | 0.930011344 | 0.930564568 |
| P4HA3 | 0.930003255 | 1.031591738 |
| PLD2 | 0.929982573 | 0.938436614 |
| CCRL2 | 0.929959069 | 0.855937703 |
| KCNE2 | 0.929947544 | 1.051265412 |
| MESP1 | 0.929921959 | 1.046591298 |
| FBXO39 | 0.929896915 | 0.976638546 |
| POTEA | 0.929879005 | 0.969852689 |
| SNX6 | 0.929842984 | 0.958204865 |
| DUSP6 | 0.929837889 | 0.902011681 |
| TCEA2 | 0.929825066 | 0.927501013 |
| OSBPL1A | 0.929820685 | 0.825435399 |
| CROT | 0.929806606 | 0.98247891 |
| ZNF691 | 0.929773362 | 0.940866748 |
| CELF2 | 0.929725089 | 0.936622131 |
| ACSF3 | 0.929716797 | 0.963343611 |
| NICN1 | 0.929682135 | 0.938335046 |
| MRE11A | 0.929677493 | 0.805589307 |
| TSPAN11 | 0.929625272 | 1.006851662 |
| LIN28A | 0.929589749 | 0.981843097 |
| CYP3A5 | 0.92954743 | 0.996916752 |
| CLEC18B | 0.929468179 | 1.0854597 |
| FAM120A | 0.929441487 | 1.09437019 |
| ACTL7A | 0.929381143 | 0.856800274 |
| TBPL2 | 0.929361205 | 0.885783718 |
| NEIL1 | 0.929354487 | 0.850746269 |
| DAD1 | 0.929304435 | 1.138221934 |
| TRIM50 | 0.929286406 | 1.01752109 |
| PNMA1 | 0.929214039 | 1.114621304 |
| PYGO2 | 0.929188578 | 0.998972251 |
| PHACTR1 | 0.929188467 | 0.999189955 |

| | | |
|---|---|---|
| ZCCHC6 | 0.929152828 | 1.014045906 |
| PRDX4 | 0.929152299 | 0.948157149 |
| RALGAPA2 | 0.929111038 | 0.961522884 |
| KLHL4 | 0.929082372 | 0.922233642 |
| PTK2 | 0.929058864 | 0.868267359 |
| GRM3 | 0.929022725 | 0.994808566 |
| VN1R2 | 0.929004531 | 0.954574951 |
| IBTK | 0.92900376 | 0.939020212 |
| SLC45A3 | 0.928915769 | 1.010962658 |
| ENDOU | 0.928900886 | 1.086956522 |
| MYLK4 | 0.928893971 | 0.9201817 |
| HPS4 | 0.928797556 | 0.947584789 |
| GBF1 | 0.928792526 | 1.002740665 |
| ZNF660 | 0.928732501 | 1.030781693 |
| HLA-DOB | 0.92871607 | 0.931862427 |
| FCER1G | 0.928694173 | 1.018170019 |
| KIAA1429 | 0.928668062 | 0.906439854 |
| PCDHGA4 | 0.928639153 | 1.009592326 |
| TNNC1 | 0.9286131 | 0.83076396 |
| FAM195B | 0.928581248 | 1.081534772 |
| SON | 0.928557188 | 0.777321 |
| NCDN | 0.928490311 | 0.849263446 |
| TMCC2 | 0.928461268 | 0.906132237 |
| ZNF471 | 0.92844744 | 1.077764277 |
| GMEB2 | 0.92844686 | 0.892771497 |
| MRPL47 | 0.928443914 | 0.809181226 |
| SP110 | 0.92844029 | 1.088386434 |
| ATG10 | 0.928421713 | 0.712230216 |
| NXPH4 | 0.928410295 | 0.978074683 |
| HPS1 | 0.928392579 | 0.922918808 |
| SLC41A2 | 0.928382899 | 0.853381936 |
| RBM38 | 0.928338989 | 0.957877683 |
| PPP3R2 | 0.928304043 | 1.047371836 |
| PRKCDBP | 0.928294121 | 0.912983899 |
| STMN1 | 0.928294121 | 0.912983899 |
| LCN6 | 0.928271421 | 0.998287085 |
| SDF2 | 0.928216172 | 1.015795869 |
| HIST1H3H | 0.928212111 | 1.015073655 |
| SPINK2 | 0.928165741 | 0.946942082 |
| CLDN10 | 0.928151894 | 0.904761905 |
| MSTN | 0.928122849 | 0.8403634 |
| TMEM150C | 0.928102575 | 0.978128797 |
| AKIRIN2 | 0.928101147 | 1.020212402 |
| AIM1 | 0.928045009 | 1.093867763 |

| | | |
|---|---|---|
| RNASE8 | 0.928040855 | 1.157040883 |
| ZNF75A | 0.928009395 | 1.010277492 |
| IBSP | 0.928007267 | 1.249188838 |
| USPL1 | 0.928007267 | 0.892277742 |
| HAUS4 | 0.927991407 | 1.025351148 |
| MLLT4 | 0.9279881 | 0.940047962 |
| PCDHGA7 | 0.927986831 | 0.911613566 |
| ERICH5 | 0.927979962 | 1.044957473 |
| ARSI | 0.927935152 | 1.076573653 |
| GNG4 | 0.927922349 | 0.971648441 |
| TXNDC8 | 0.92788338 | 0.662616444 |
| CSNK1A1L | 0.927863696 | 1.082414017 |
| STXBP1 | 0.927841569 | 0.903391572 |
| NKIRAS2 | 0.927824646 | 1.029566626 |
| MTMR9 | 0.927823275 | 0.950032446 |
| PLXNA1 | 0.927735848 | 1.001946788 |
| FLI1 | 0.927694901 | 0.869582827 |
| OR8H2 | 0.927668441 | 0.520383693 |
| SERPINB2 | 0.927667849 | 0.915639195 |
| TCF21 | 0.927663897 | 0.961713173 |
| TTF2 | 0.92758569 | 0.875405581 |
| WSB1 | 0.92758569 | 0.921479559 |
| TTLL1 | 0.927580438 | 0.940866748 |
| TMEM38B | 0.927572112 | 0.943816375 |
| AKTIP | 0.927508306 | 0.9733939 |
| CACNA2D2 | 0.927508306 | 0.9733939 |
| AHSP | 0.927494814 | 0.947242206 |
| DCST2 | 0.927449777 | 0.889833941 |
| SGCZ | 0.92744955 | 0.905447071 |
| HIST1H3E | 0.927438676 | 0.920520726 |
| ZNF547 | 0.92740672 | 0.84101233 |
| ELN | 0.927379909 | 0.952380952 |
| LPAR6 | 0.927376095 | 1.077222583 |
| SV2C | 0.927359928 | 0.923946557 |
| PRADC1 | 0.927356502 | 0.985074627 |
| C16orf71 | 0.927285597 | 0.852166869 |
| ITGA7 | 0.927267182 | 0.944841012 |
| DPY30 | 0.927266182 | 0.957519699 |
| CDC73 | 0.927247595 | 1.059133252 |
| TTC29 | 0.927236134 | 0.892264075 |
| C4B | 0.92723297 | 0.870503597 |
| IMMP1L | 0.92722495 | 1.042829332 |
| NUB1 | 0.927200577 | 0.92345079 |
| TECTB | 0.927167226 | 1.021240151 |

| | | |
|---|---|---|
| ACBD3 | 0.927160973 | 0.7550531 |
| ZNF274 | 0.927144299 | 0.889003083 |
| TRPC7 | 0.927122622 | 0.812459442 |
| TMEM108 | 0.927111279 | 1.012555691 |
| PRKCI | 0.927107063 | 1.100584036 |
| STT3B | 0.927079696 | 1.048197651 |
| ERV3-1 | 0.926990926 | 0.865707434 |
| ARHGAP12 | 0.926980777 | 0.9109284 |
| TRIB2 | 0.926976828 | 1.112264763 |
| GABPB2 | 0.926936917 | 0.942788626 |
| PSMD2 | 0.92693225 | 0.972935937 |
| FLNA | 0.926930747 | 0.754556501 |
| ARPP21 | 0.926892559 | 1.004796163 |
| SYTL1 | 0.926875024 | 1.606029462 |
| PCDHGB4 | 0.926825682 | 0.917780062 |
| SNRPC | 0.926823235 | 0.978074683 |
| ZZZ3 | 0.926823235 | 0.978074683 |
| PRAMEF3_pseudogene | 0.926809204 | 0.901579587 |
| ISL1 | 0.926752497 | 1.083711875 |
| BRIX1 | 0.926751782 | 0.996574169 |
| IER2 | 0.92675065 | 0.966426859 |
| KCNJ1 | 0.926745993 | 0.985723556 |
| AGPAT3 | 0.92669166 | 0.841661259 |
| ZBTB42 | 0.926684452 | 1.030147311 |
| OR6K3 | 0.926664743 | 0.864337102 |
| WFDC6 | 0.92662709 | 0.881123672 |
| CCDC33 | 0.926552873 | 1.019846092 |
| SNRPB2 | 0.926534342 | 0.976704351 |
| FOXD2 | 0.926467247 | 1.00908501 |
| DYX1C1 | 0.92645292 | 0.949982871 |
| CASP6 | 0.926429461 | 0.91109669 |
| SCNM1 | 0.926416446 | 0.951695786 |
| METTL14 | 0.92641119 | 0.844467283 |
| C8orf46 | 0.926392972 | 0.901984609 |
| BAG1 | 0.926384108 | 0.942081814 |
| PUS7L | 0.926358638 | 1.080194411 |
| B4GALT2 | 0.926354793 | 0.916937054 |
| SYTL2 | 0.926307929 | 0.956834532 |
| RNF165 | 0.926266378 | 1.0266061 |
| SRSF6 | 0.926246399 | 0.913326482 |
| TMCC1 | 0.926191076 | 0.946899623 |
| C6orf106 | 0.926185034 | 0.868772783 |
| MYO15A | 0.926185034 | 0.677197246 |
| CD47 | 0.926139559 | 0.980472765 |

| | | |
|---|---|---|
| USH1G | 0.926121073 | 1.059266872 |
| PEX11B | 0.92609144 | 1.014045906 |
| ELL2 | 0.926075265 | 1.139773895 |
| KLHL21 | 0.926048759 | 0.859883522 |
| OR3A1 | 0.926002679 | 1.034258308 |
| VPS33B | 0.925995842 | 0.991494532 |
| MTL5 | 0.925986503 | 0.925316889 |
| CHSY1 | 0.925965278 | 0.853381936 |
| GAS2 | 0.925947958 | 0.791008505 |
| HSD17B7 | 0.925936756 | 1.003893576 |
| VPS13D | 0.925891716 | 1.024323398 |
| SESTD1 | 0.925856102 | 0.947242206 |
| TMEM39A | 0.925781885 | 0.982528263 |
| MLC1 | 0.925747989 | 0.859831278 |
| RTKN2 | 0.925747722 | 0.984241178 |
| TNFAIP3 | 0.925744037 | 1.021414666 |
| TTC13 | 0.925695525 | 1.066461117 |
| CCDC173 | 0.925679893 | 0.923946557 |
| ATL2 | 0.925647225 | 1.021240151 |
| KRT71 | 0.925646749 | 1.045362495 |
| ICOS | 0.925624933 | 0.910585817 |
| MTCL1 | 0.925602051 | 1.071599863 |
| SNRK | 0.925557822 | 1.038935756 |
| ZNF318 | 0.925512429 | 0.996231586 |
| PECAM1 | 0.925509393 | 1.076738609 |
| ZNF443 | 0.92550092 | 0.932511134 |
| CYP27A1 | 0.925400832 | 0.998702141 |
| GIN1 | 0.925395105 | 0.949777238 |
| PNPLA8 | 0.925338595 | 0.837118754 |
| ZNF136 | 0.925319085 | 1.06229721 |
| SHANK1 | 0.925303189 | 0.974648852 |
| NXPE4 | 0.925284463 | 1.041312272 |
| AMMECR1 | 0.925257834 | 0.830421377 |
| ARHGEF35 | 0.925209262 | 1.028434395 |
| UBXN1 | 0.925169752 | 0.850633779 |
| PLEKHG2 | 0.92516185 | 0.916565411 |
| NOVA2 | 0.925159648 | 0.801986982 |
| TET1 | 0.92514667 | 1.083839611 |
| GCUD2 | 0.925124918 | 1.061563386 |
| SLC22A1 | 0.92510381 | 0.970149254 |
| CBLN1 | 0.925090008 | 1.018499486 |
| CNTD2 | 0.925088088 | 1.025921426 |
| ATP6V0C | 0.925070887 | 0.98663926 |
| ATF3 | 0.92506975 | 0.840826245 |

| | | |
|---|---|---|
| SLC39A7 | 0.925032328 | 1.005138746 |
| CHRFAM7A | 0.924995122 | 0.945727015 |
| SLC15A4 | 0.924989468 | 0.94127177 |
| TSGA13 | 0.924988423 | 0.941418294 |
| ARNTL | 0.924937509 | 0.751721345 |
| APBB1 | 0.924936499 | 1.010277492 |
| CSPG5 | 0.92486201 | 0.897910243 |
| ENPP3 | 0.92485931 | 0.993510707 |
| EMC3 | 0.924843423 | 0.838801134 |
| NCOA4 | 0.924809751 | 1.057105775 |
| BARX1 | 0.924799955 | 0.999351071 |
| NDST2 | 0.924773787 | 0.930336168 |
| LYRM5 | 0.924760186 | 0.921425678 |
| MYB | 0.924741176 | 1.005191434 |
| JAGN1 | 0.924732016 | 0.903604698 |
| TMEM134 | 0.924723051 | 1.026326448 |
| CORIN | 0.92468322 | 1.011031798 |
| NTNG2 | 0.924652589 | 0.958547448 |
| PNMA6A | 0.924652589 | 0.958547448 |
| C10orf55 | 0.924647922 | 0.841041453 |
| BDNF | 0.92464494 | 0.895522388 |
| NFIL3 | 0.924635638 | 0.968408262 |
| C11orf94 | 0.924619694 | 0.960260363 |
| ZUFSP | 0.924615979 | 0.909900651 |
| PSMC5 | 0.924565187 | 1.042122317 |
| EZR | 0.92455182 | 0.946899623 |
| CD276 | 0.924549419 | 0.930113052 |
| WDR12 | 0.924538771 | 0.862966769 |
| CCSER2 | 0.924534648 | 1.049331963 |
| ZFYVE1 | 0.9245225 | 1.015390846 |
| PPP1R32 | 0.924515215 | 1.010935601 |
| CHCHD1 | 0.92450932 | 0.898252826 |
| L1CAM | 0.924452728 | 0.952038369 |
| SMUG1 | 0.924403074 | 1.040233615 |
| C9orf85 | 0.92438721 | 0.9554642 |
| CKS1B | 0.924369157 | 0.98247891 |
| SRD5A3 | 0.924348906 | 1.046073978 |
| MTHFSD | 0.924299127 | 0.89509923 |
| MAP2K3 | 0.924287739 | 0.8669695 |
| ANGPT2 | 0.924255551 | 0.994159637 |
| OTOR | 0.924225591 | 1.032887975 |
| DEFB129 | 0.924214413 | 1.046982584 |
| CCAR1 | 0.92420203 | 0.982528263 |
| OR52N1 | 0.924184715 | 0.937626569 |

| | | |
|---|---|---|
| SF3B4 | 0.924142901 | 0.933881466 |
| IRS2 | 0.924087397 | 1.006166495 |
| PPM1N | 0.923989483 | 1.046591298 |
| BCL9 | 0.923893381 | 0.999657417 |
| BTAF1 | 0.923867891 | 0.971447112 |
| PCP4 | 0.923865923 | 0.966084275 |
| FOXO4 | 0.923852127 | 0.964763062 |
| CMTM7 | 0.923830324 | 0.955852572 |
| KPNA6 | 0.92381927 | 0.951397327 |
| SOWAHB | 0.923785548 | 0.938031592 |
| ZNF132 | 0.923773447 | 1.041795135 |
| ANKRD32 | 0.923771815 | 0.935936965 |
| FGD3 | 0.92375632 | 0.954436451 |
| KIDINS220 | 0.923739796 | 0.919532771 |
| PRR34 | 0.923731806 | 0.993924666 |
| TCEA3 | 0.923725554 | 1.052563271 |
| MFN1 | 0.923675568 | 0.971648441 |
| RP1L1 | 0.923666697 | 1.119218911 |
| MRPS16 | 0.923664005 | 0.967193196 |
| C1orf94 | 0.923628754 | 0.953827461 |
| DAAM2 | 0.923612371 | 0.944501542 |
| WDR83 | 0.923600092 | 0.963001028 |
| TMEM171 | 0.923592933 | 0.927714971 |
| VARS2 | 0.923572873 | 1.00526529 |
| ZNF200 | 0.923529421 | 0.9181855 |
| HTR3A | 0.923515877 | 0.942894225 |
| INHBB | 0.923515877 | 0.942894225 |
| KIF27 | 0.923511286 | 1.052247874 |
| NSA2 | 0.923507706 | 0.968139774 |
| TMEM106C | 0.923462282 | 0.826650466 |
| FKBP4 | 0.923432685 | 0.817739976 |
| C16orf3 | 0.923315231 | 0.924631723 |
| IGFBP2 | 0.92331301 | 1.015416238 |
| C8orf31 | 0.923179462 | 0.811259619 |
| FBXW5 | 0.923156289 | 1.0532122 |
| ADIPOQ | 0.923147115 | 0.987849332 |
| BTBD3 | 0.923144459 | 1.007194245 |
| DOLPP1 | 0.923141965 | 0.914341337 |
| PPIAL4A | 0.923119505 | 0.978938842 |
| OR5H6 | 0.923087006 | 1.010620075 |
| CIRBP | 0.922973279 | 1.054470709 |
| MYBPC1 | 0.922973279 | 0.998972251 |
| TMC2 | 0.922973279 | 0.980472765 |
| IL32 | 0.922957923 | 0.929931146 |

| | | |
|---|---|---|
| OR51B6 | 0.922946909 | 1.093182597 |
| GNG12 | 0.922940973 | 0.889688249 |
| NRXN1 | 0.922935968 | 0.981368975 |
| PPP1R27 | 0.922918908 | 1.05789654 |
| ZNF561 | 0.922913742 | 0.965399109 |
| KRAS | 0.922906109 | 0.868267359 |
| LDB2 | 0.922901991 | 1.028351559 |
| ARHGEF7 | 0.922851281 | 0.874107722 |
| LMOD3 | 0.922840341 | 1.081534772 |
| NRAS | 0.922812489 | 0.889833941 |
| CCHCR1 | 0.922799154 | 0.990750257 |
| RLN1 | 0.922795847 | 0.972250771 |
| KHDRBS3 | 0.922781257 | 0.898252826 |
| PKP3 | 0.92267383 | 0.961522884 |
| TCIRG1 | 0.922670335 | 1.117456197 |
| PRAM1 | 0.922645965 | 1.059943297 |
| DPCR1 | 0.922528384 | 0.969167523 |
| MED20 | 0.922513341 | 0.874957177 |
| OPN5 | 0.922487248 | 0.914990266 |
| KIAA0319 | 0.92245095 | 0.935594382 |
| LYG1 | 0.922410171 | 0.612538541 |
| ATM | 0.922404019 | 1.001946788 |
| NDUFA1 | 0.922373639 | 0.885783718 |
| MARCO | 0.922315211 | 0.872417983 |
| OR3A4P | 0.922271026 | 1.023295649 |
| NOL8 | 0.922256074 | 1.082219938 |
| TRAF7 | 0.922246453 | 0.944192083 |
| UNC45A | 0.922154596 | 0.930741191 |
| PRSS38 | 0.922143796 | 1.036988968 |
| MIS18BP1 | 0.922115478 | 1.072285029 |
| ASPG | 0.922017211 | 0.810044552 |
| ZNF613 | 0.921945567 | 0.867077766 |
| MASP2 | 0.921925639 | 0.903958469 |
| LCE2B | 0.92186188 | 0.931140802 |
| SNRNP70 | 0.921857485 | 1.030489894 |
| ZIC4 | 0.921857485 | 1.030489894 |
| SPNS2 | 0.921853489 | 0.995949777 |
| OSGEPL1 | 0.921815326 | 0.834523037 |
| C6orf48 | 0.921775928 | 0.936279548 |
| TSNAXIP1 | 0.921730214 | 0.884163629 |
| PHPT1 | 0.921725182 | 1.014276444 |
| NTHL1 | 0.921645246 | 1.025957171 |
| TUBD1 | 0.921636108 | 1.112913692 |
| SORCS2 | 0.921613058 | 0.944841012 |

| | | |
|---|---|---|
| VGLL3 | 0.921601794 | 1.02308627 |
| LINGO2 | 0.921581774 | 0.81490482 |
| LAX1 | 0.921577189 | 0.895504253 |
| GPR123 | 0.921570579 | 0.950681376 |
| ZNF436 | 0.921551147 | 0.869133265 |
| RNF25 | 0.921528676 | 0.956521739 |
| RPL21P44 | 0.921443471 | 1.068043742 |
| HIST1H4J | 0.921430986 | 0.875984926 |
| CLPX | 0.921393831 | 0.887086308 |
| RAB21 | 0.921345666 | 1.007695423 |
| ZDHHC2 | 0.92131087 | 0.985611511 |
| PPCS | 0.921308174 | 0.884568651 |
| CBWD5 | 0.92130089 | 1.036857027 |
| RPS27L | 0.92128459 | 0.987324426 |
| QTRTD1 | 0.921279063 | 0.967112025 |
| MMD2 | 0.921273263 | 0.946899623 |
| C5orf22 | 0.921235381 | 0.947347104 |
| ARHGDIG | 0.921211642 | 0.930113052 |
| IMMT | 0.921200181 | 1.128797084 |
| ENTPD2 | 0.921195059 | 1.034600891 |
| SSH3 | 0.921173122 | 1.00908501 |
| CYP1A2 | 0.921151431 | 0.819765087 |
| CHAD | 0.921138014 | 0.9554642 |
| XYLT2 | 0.921066922 | 0.916160389 |
| PCYT1B | 0.921054659 | 0.84101233 |
| BARHL2 | 0.920970731 | 0.861253854 |
| CD84 | 0.920872454 | 0.930455635 |
| OR56B4 | 0.920845495 | 0.932168551 |
| COX8A | 0.920829176 | 0.968851395 |
| CAPZA3 | 0.920823293 | 0.976019185 |
| IL17B | 0.9207672 | 1.107571086 |
| SRPK1 | 0.920760107 | 0.881894873 |
| ERBB4 | 0.920684946 | 0.992212849 |
| YPEL1 | 0.92066394 | 1.008221994 |
| HOXA2 | 0.920661947 | 0.882948562 |
| SMAD1 | 0.920656089 | 0.983799109 |
| DGKD | 0.920642138 | 0.899415964 |
| LDB1 | 0.920630161 | 1.012960713 |
| MINOS1 | 0.920630161 | 1.012960713 |
| GNPDA2 | 0.920605607 | 0.874038072 |
| NEUROD4 | 0.920469833 | 1.021925317 |
| MFAP2 | 0.920448039 | 1.001712915 |
| ZNF644 | 0.92041448 | 1.040097205 |
| AZIN1 | 0.92036629 | 0.934791414 |

| | | |
|---|---|---|
| GRAP2 | 0.920325522 | 0.944501542 |
| SNRPG | 0.920323619 | 0.878725591 |
| ARIH1 | 0.920290206 | 1.06229721 |
| SH3GL2 | 0.920279998 | 0.957819598 |
| PAK1IP1 | 0.920238664 | 0.86193902 |
| COMMD2 | 0.920169159 | 1.066118534 |
| DUB3 | 0.920108228 | 1.049412718 |
| HCAR3 | 0.920083163 | 0.992861778 |
| VSIG4 | 0.920067258 | 0.939705379 |
| ATP2C1 | 0.92005525 | 0.948562171 |
| FAM76B | 0.920026418 | 0.944106926 |
| GPR162 | 0.919978448 | 0.900064893 |
| SLC16A13 | 0.91993228 | 0.926344639 |
| FAM210B | 0.919917147 | 0.949982871 |
| EARS2 | 0.91990788 | 0.926285946 |
| PLXNA2 | 0.919902829 | 0.973621103 |
| GEN1 | 0.919900901 | 0.776022681 |
| DDI1 | 0.919849961 | 0.888213852 |
| THOC1 | 0.919806954 | 0.980472765 |
| ENTHD1 | 0.91969118 | 0.924260834 |
| LRRC8C | 0.919595871 | 0.910895099 |
| OR51B2 | 0.919589758 | 1.052247874 |
| AGPAT5 | 0.919574986 | 0.964763062 |
| EIF2AK2 | 0.919567984 | 0.975989617 |
| UQCRB | 0.919567984 | 1.097988319 |
| ARPC1B | 0.919561467 | 0.952038369 |
| WDR13 | 0.919542799 | 0.930113052 |
| NPR2 | 0.919539006 | 0.981829981 |
| C7orf61 | 0.919537407 | 0.953751285 |
| H2AFX | 0.919482121 | 0.993510707 |
| TOMM70A | 0.919465456 | 1.122644741 |
| TM4SF5 | 0.919436729 | 1.007879411 |
| LGALS3 | 0.91943597 | 1.083839611 |
| NAT6 | 0.919426635 | 1.005191434 |
| PCGF3 | 0.919426635 | 1.005191434 |
| GJD2 | 0.919397663 | 0.967193196 |
| C3orf22 | 0.919366284 | 0.962737951 |
| BLMH | 0.919315581 | 0.900713822 |
| ITSN2 | 0.919303066 | 1.018156903 |
| DEFA1 | 0.919270313 | 0.949372215 |
| FAM173A | 0.919156614 | 0.960712839 |
| PLK1 | 0.91910434 | 0.674237508 |
| tcag7.926_pseudogene | 0.919097043 | 0.986296677 |
| SHOX | 0.919088122 | 0.947436729 |

| | | |
|---|---|---|
| SCPEP1 | 0.919047948 | 1.092796885 |
| ENSA | 0.919045707 | 0.966084275 |
| SLCO1B3 | 0.919035376 | 0.9181855 |
| PRKCE | 0.919034612 | 0.959117456 |
| LMO7 | 0.919024816 | 1.098637249 |
| SHMT2 | 0.919023813 | 0.864317537 |
| TAGAP | 0.919007421 | 0.920520726 |
| LUC7L | 0.918959347 | 1.045563549 |
| CYP2C8 | 0.918906682 | 0.988319273 |
| MPHOSPH9 | 0.918888121 | 0.978074683 |
| COQ7 | 0.918816849 | 0.885234669 |
| SWSAP1 | 0.918803548 | 0.965573107 |
| HIST1H3B | 0.918797411 | 1.108256252 |
| ADAM8 | 0.918785996 | 0.872160934 |
| CPSF7 | 0.918777973 | 0.863309353 |
| SLC35G2 | 0.918745832 | 1.109356015 |
| F13B | 0.918732074 | 0.883841661 |
| C10orf62 | 0.918719886 | 0.966426859 |
| EBLN2 | 0.918708901 | 0.878088295 |
| TMEM11 | 0.918705648 | 0.889682025 |
| LPHN2 | 0.91869437 | 1.040882544 |
| LRRC27 | 0.918550564 | 0.929427886 |
| TBX20 | 0.918533029 | 0.880438506 |
| GLOD4 | 0.918524294 | 0.979343864 |
| VANGL2 | 0.918521022 | 1.007194245 |
| CBX5 | 0.918515223 | 1.083247688 |
| STRC | 0.918483868 | 0.909215485 |
| AP2A2 | 0.918466884 | 1.069258809 |
| DEPTOR | 0.918460669 | 1.019846092 |
| VIPAS39 | 0.918437698 | 0.811236725 |
| FGF17 | 0.918343864 | 0.813284731 |
| LAT | 0.918337654 | 1.047961631 |
| RAMP1 | 0.918335208 | 1.022610483 |
| SMIM4 | 0.918320709 | 0.895854745 |
| PROSER1 | 0.918297555 | 0.89756766 |
| FDXR | 0.918282589 | 0.994808566 |
| SORT1 | 0.9182206 | 1.012329656 |
| UROC1 | 0.918163332 | 0.865127582 |
| FLJ40296 | 0.918099009 | 0.990750257 |
| GRB7 | 0.918006305 | 0.972250771 |
| SCARA5 | 0.918003227 | 0.802349129 |
| RFPL3 | 0.917992812 | 1.082414017 |
| EVC2 | 0.917968426 | 1.028091812 |
| HOXB4 | 0.917965436 | 0.843782117 |

| | | |
|---|---|---|
| SLC25A34 | 0.917945715 | 0.887808829 |
| SPTY2D1 | 0.91789829 | 0.97326853 |
| RGS14 | 0.917874001 | 0.930113052 |
| COQ4 | 0.917871237 | 1.036656389 |
| SHISA4 | 0.917860187 | 0.968813285 |
| SORBS1 | 0.917852479 | 0.931825968 |
| RABEPK | 0.917821727 | 0.96435804 |
| CYP4Z1 | 0.91780849 | 1.146658014 |
| TRIM64C | 0.917792684 | 0.960415315 |
| NFKBID | 0.917702097 | 0.997944502 |
| AAED1 | 0.917662677 | 0.974306269 |
| RNF213 | 0.917662677 | 1.055498458 |
| GGCT | 0.917620177 | 0.923603974 |
| HOXA5 | 0.917619647 | 1.007290401 |
| TNRC6C | 0.917605842 | 1.027541515 |
| DNAH6 | 0.91758257 | 0.981157931 |
| MYO7B | 0.917575411 | 0.899965742 |
| CENPN | 0.917572833 | 0.762251924 |
| STAP2 | 0.91749911 | 0.960945529 |
| LOC400867 | 0.917494443 | 0.933881466 |
| SNN | 0.917488824 | 1.016786571 |
| UBE2V1 | 0.917487841 | 1.065541856 |
| CTH | 0.917406106 | 0.854597003 |
| COA7 | 0.917360005 | 0.850141758 |
| ABHD15 | 0.91734397 | 0.862966769 |
| PCDHA5 | 0.917310848 | 0.949297705 |
| RFWD3 | 0.917266556 | 0.926022064 |
| DUS1L | 0.917205452 | 0.87727825 |
| ZCCHC16 | 0.917205452 | 0.87727825 |
| SLC16A14 | 0.917204643 | 0.98663926 |
| SNX13 | 0.917161281 | 1.018842069 |
| TBC1D3G | 0.917128704 | 0.993490922 |
| RTFDC1 | 0.917079588 | 1.112024666 |
| RP1 | 0.917057064 | 0.924665857 |
| ZNF230 | 0.916960514 | 0.951353203 |
| CC2D1B | 0.916923193 | 0.951802349 |
| YRDC | 0.916883867 | 1.048602673 |
| CXorf21 | 0.916875564 | 0.866342649 |
| FAM26D | 0.916839243 | 0.983394087 |
| NXT2 | 0.916795667 | 0.850746269 |
| ZNF562 | 0.916787251 | 1.054813292 |
| SLC24A1 | 0.916760541 | 1.014985824 |
| COL3A1 | 0.916757935 | 0.998972251 |
| STX6 | 0.916728982 | 1.03117506 |

| | | |
|---|---|---|
| TP53TG3 | 0.916652273 | 1.127783487 |
| FSTL4 | 0.916627897 | 0.892428914 |
| AKR1A1 | 0.916622697 | 0.961117861 |
| APBA3 | 0.916600232 | 0.93835172 |
| IGF2-AS | 0.916580956 | 0.926344639 |
| GDI1 | 0.916578175 | 0.956662616 |
| PTPRS | 0.916565409 | 0.95587281 |
| FABP1 | 0.916488673 | 1.04455245 |
| IPO11 | 0.916485806 | 0.934909215 |
| LPIN1 | 0.916485806 | 0.934909215 |
| TYW1 | 0.916478281 | 0.927501013 |
| KNOP1 | 0.916470083 | 1.087358684 |
| PANK3 | 0.91646835 | 1.00843608 |
| FAM8A1 | 0.916461248 | 0.997601918 |
| IP6K2 | 0.916448181 | 1.020116807 |
| SGKL | 0.916428341 | 1.031797534 |
| PIK3CD | 0.916418627 | 1.037637897 |
| FLJ44635 | 0.916414851 | 0.850291196 |
| OR1D4 | 0.916390203 | 1.065433368 |
| ADCK1 | 0.916353591 | 1.078520441 |
| TNFAIP8L2 | 0.916343239 | 1.067233698 |
| ZBED3 | 0.916336362 | 0.914135277 |
| LRRC31 | 0.91627267 | 0.862281603 |
| DIRC1 | 0.916232842 | 0.989379925 |
| STMN2 | 0.916210459 | 0.960602946 |
| ZNF696 | 0.916203391 | 1.023295649 |
| OR4C12 | 0.916170857 | 0.767922236 |
| EMD | 0.916163791 | 0.996231586 |
| FBXO11 | 0.916156757 | 0.816353018 |
| ZNF558 | 0.916113769 | 0.938677629 |
| MS4A4A | 0.916113652 | 0.968408262 |
| NUFIP1 | 0.916084718 | 0.984204131 |
| GPX8 | 0.916078744 | 1.004796163 |
| PRR21 | 0.916073816 | 1.020251114 |
| SLC18A1 | 0.916068753 | 0.868916288 |
| GNG13 | 0.91605976 | 0.943816375 |
| CCDC14 | 0.916047022 | 1.036046983 |
| GMNN | 0.916041862 | 0.787941076 |
| TNFRSF8 | 0.916033025 | 0.892277742 |
| GPR139 | 0.916007422 | 0.909798832 |
| UBE2QL1 | 0.915990793 | 0.921479559 |
| HIVEP2 | 0.915982725 | 0.927319922 |
| KIF19 | 0.915972649 | 0.955042527 |
| SH3BP1 | 0.915949476 | 0.986296677 |

| | | |
|---|---|---|
| EGFL8 | 0.915867738 | 0.930455635 |
| ST3GAL4 | 0.91585997 | 0.853786958 |
| CA13 | 0.915845904 | 1.038286827 |
| ITGA9 | 0.915820796 | 1.06164828 |
| UPF2 | 0.915800317 | 1.033915725 |
| PRR15L | 0.915733944 | 1.04090725 |
| PSMA1 | 0.915690313 | 1.201817002 |
| KCTD18 | 0.915615046 | 0.988352175 |
| FAM101B | 0.915612095 | 0.975698663 |
| C6orf62 | 0.91559153 | 0.891743748 |
| CDH20 | 0.915590398 | 1.023980815 |
| BPY2 | 0.915512465 | 1.032545392 |
| CCDC138 | 0.915469462 | 1.050222762 |
| ASNS | 0.915434288 | 0.811161583 |
| PLA2G5 | 0.915413789 | 0.834523037 |
| TIRAP | 0.915403541 | 1.078451524 |
| BEX1 | 0.915396971 | 0.910585817 |
| CDC14B | 0.915376085 | 0.881245944 |
| ATP5I | 0.915375528 | 1.093155124 |
| BLK | 0.915354517 | 0.910447761 |
| HOXC10 | 0.915347919 | 0.969218307 |
| TK2 | 0.915346202 | 0.922128488 |
| MGAT5B | 0.915345409 | 0.949640288 |
| DPEP1 | 0.915342168 | 0.927968851 |
| ESPL1 | 0.915318871 | 0.963011032 |
| PPM1M | 0.915318871 | 0.963011032 |
| PRRG3 | 0.915297788 | 0.988694758 |
| SLC3A1 | 0.915297302 | 0.998053212 |
| PHOX2A | 0.915288777 | 0.757391657 |
| PARD6B | 0.915271418 | 1.059952038 |
| KLK11 | 0.915246344 | 1.091499027 |
| PFKL | 0.915246344 | 1.091499027 |
| CCIN | 0.915217512 | 0.991899554 |
| HADHB | 0.915203454 | 1.184944841 |
| EME2 | 0.915178941 | 1.036313806 |
| H2BFWT | 0.915153885 | 0.900650908 |
| SEMA4G | 0.915149309 | 1.039739637 |
| FAM20B | 0.915095572 | 0.941418294 |
| ZKSCAN7 | 0.915073098 | 0.978759849 |
| LAS1L | 0.915023583 | 0.890643985 |
| SHF | 0.914933164 | 0.850141758 |
| C9orf117 | 0.914905317 | 0.926344639 |
| STOX2 | 0.914885535 | 1.012555691 |
| NACC1 | 0.91485901 | 0.967112025 |

| | | |
|---|---|---|
| TMEM177 | 0.914813913 | 0.841231268 |
| MRPS23 | 0.914772695 | 1.013018157 |
| CNOT8 | 0.914763024 | 0.93600648 |
| AP4B1 | 0.914740888 | 0.907845152 |
| MRPL36 | 0.914715242 | 1.046577562 |
| SLC1A4 | 0.914715242 | 0.915755367 |
| AACS | 0.914708416 | 0.931551235 |
| EFEMP1 | 0.91470822 | 0.984241178 |
| MIP | 0.914702517 | 0.975340688 |
| OIT3 | 0.914702517 | 0.963659961 |
| PPM1B | 0.914702517 | 1.016223232 |
| RDH5 | 0.914702517 | 0.987021415 |
| SCAMP1 | 0.914702517 | 0.91109669 |
| SYNJ2 | 0.914702517 | 0.887735237 |
| TIMP2 | 0.914702517 | 1.162232317 |
| DNAJC24 | 0.914695375 | 0.963142973 |
| DNAJC8 | 0.914695375 | 0.963142973 |
| TNPO1 | 0.914649835 | 0.780477926 |
| NEUROD2 | 0.914644107 | 1.028434395 |
| NR1I3 | 0.914607758 | 0.891049008 |
| CDHR1 | 0.914546691 | 1.040424803 |
| SERINC4 | 0.914475238 | 0.936964714 |
| C10orf107 | 0.914449457 | 1.015073655 |
| INO80C | 0.91441851 | 0.981157931 |
| C6orf226 | 0.914385344 | 0.947242206 |
| SATB2 | 0.914341076 | 0.952380952 |
| ZNF714 | 0.914310828 | 0.868772783 |
| CCK | 0.914305236 | 0.994861254 |
| CWH43 | 0.914298984 | 1.033915725 |
| EDRF1 | 0.91426849 | 0.960945529 |
| HADHA | 0.914257493 | 0.990684488 |
| MFSD10 | 0.91420854 | 1.033616849 |
| SLC36A1 | 0.914200084 | 0.954637505 |
| ESRP1 | 0.91419825 | 0.938841636 |
| SEMA3F | 0.914184758 | 0.932168551 |
| PDSS2 | 0.914140993 | 0.934386391 |
| LYPD1 | 0.91412184 | 0.839611179 |
| BCAM | 0.914094395 | 0.975989617 |
| RCHY1 | 0.914094395 | 0.975989617 |
| ZNF185 | 0.914090691 | 0.970149254 |
| LIPH | 0.914083199 | 0.958468527 |
| DGCR8 | 0.914062558 | 0.867762933 |
| ADAM17 | 0.914055621 | 0.917585983 |
| CCDC61 | 0.914035146 | 1.029119561 |

| | | |
|---|---|---|
| CCKAR | 0.914030018 | 0.882543803 |
| SCOC | 0.914026446 | 0.832134293 |
| DACT1 | 0.914009047 | 1.032545392 |
| SH3GL1 | 0.913915121 | 0.964713943 |
| ZNF844 | 0.913915121 | 0.964713943 |
| TTC39C | 0.913897676 | 1.007194245 |
| SLC25A40 | 0.913892456 | 0.884973674 |
| AMN | 0.913887799 | 0.968139774 |
| DOC2A | 0.913848923 | 0.932511134 |
| PCED1B | 0.913798522 | 1.135682463 |
| NBPF1 | 0.913688899 | 1.020656136 |
| TDRD5 | 0.913656992 | 0.956491949 |
| MTCH2 | 0.91364453 | 1.03199676 |
| COL18A1-AS1 | 0.913623441 | 1.000405022 |
| ATP5SL | 0.913594918 | 0.922576225 |
| TBC1D17 | 0.913590075 | 1.027541515 |
| GSTO2 | 0.913589209 | 0.953017416 |
| WDR72 | 0.913564554 | 0.921425678 |
| NFAM1 | 0.913542313 | 0.886947585 |
| GFI1B | 0.913522249 | 1.005840363 |
| FBXL22 | 0.913508336 | 0.994159637 |
| CD180 | 0.913499949 | 0.892086331 |
| TMCC3 | 0.913499949 | 1.104487838 |
| EEF1E1 | 0.913497619 | 0.975698663 |
| DOCK11 | 0.913471947 | 0.895512162 |
| SH2D3C | 0.913455953 | 0.854744776 |
| HTR1D | 0.913433834 | 0.935756003 |
| CRY1 | 0.913417863 | 0.924075276 |
| EBAG9 | 0.913394332 | 0.948269955 |
| RIBC1 | 0.913380609 | 0.966788173 |
| BCAR1 | 0.913362208 | 0.909215485 |
| PLIN3 | 0.913337361 | 0.978128797 |
| ETV1 | 0.913332725 | 0.919400567 |
| IFT20 | 0.913308987 | 0.844876468 |
| LRRC4B | 0.913192025 | 0.975676602 |
| CLIC2 | 0.913177918 | 0.980558931 |
| SEMA6A | 0.913155324 | 0.980815348 |
| TNIP2 | 0.913127051 | 0.940047962 |
| E2F4 | 0.91309211 | 1.10577547 |
| RNASE13 | 0.913072181 | 1.082905104 |
| HEXIM1 | 0.91301654 | 0.940056703 |
| TRPA1 | 0.912983114 | 1.03569111 |
| RBM43 | 0.912976348 | 0.951397327 |
| IMPDH1 | 0.912923268 | 0.876873228 |

| | | |
|---|---|---|
| KIF7 | 0.912890514 | 0.931146213 |
| THNSL1 | 0.912880457 | 0.977287476 |
| ECE1 | 0.912869425 | 0.971447112 |
| SNX11 | 0.912818538 | 0.936964714 |
| MAP1A | 0.912750619 | 0.852004111 |
| CLDN23 | 0.912723025 | 0.950668037 |
| MTF1 | 0.912678144 | 0.874848117 |
| TAS2R46 | 0.912650167 | 0.913326482 |
| FAM65A | 0.91264814 | 0.859052248 |
| TAOK2 | 0.912633075 | 0.860480208 |
| NABP2 | 0.912609069 | 0.99473471 |
| THEMIS | 0.912521579 | 0.882836588 |
| FGF16 | 0.912514283 | 0.8137573 |
| TEX13B | 0.912472826 | 0.889688249 |
| CDC42 | 0.912472091 | 0.834345889 |
| KCNQ2 | 0.912466042 | 0.79623621 |
| KLF7 | 0.91246226 | 0.915755367 |
| STARD5 | 0.912460728 | 0.891401165 |
| KCNV2 | 0.912449577 | 1.053861129 |
| DHRS2 | 0.912427875 | 0.92709599 |
| CCDC69 | 0.912422424 | 1.008505468 |
| FAM47A | 0.912361398 | 0.705548805 |
| IRX4 | 0.912361398 | 0.841231268 |
| ELAVL3 | 0.912356516 | 1.007194245 |
| NAV1 | 0.912345027 | 1.007138222 |
| DNA2 | 0.912331279 | 1.001297859 |
| USP33 | 0.912317366 | 0.995457495 |
| PRC1 | 0.912251171 | 0.922233642 |
| FBXW10 | 0.912215203 | 0.954574951 |
| MED26 | 0.912136749 | 0.925373134 |
| KDELR1 | 0.912087558 | 0.895169579 |
| OR2F1 | 0.912086878 | 0.947927372 |
| CDC42BPG | 0.912070314 | 0.902011681 |
| BICD2 | 0.911977365 | 1.019527235 |
| AHSA1 | 0.91197247 | 0.95018226 |
| CCDC157 | 0.911929246 | 1.028091812 |
| RAP1GAP | 0.911903527 | 1.060350422 |
| CGB8 | 0.911881807 | 1.036656389 |
| GYPB | 0.911868939 | 0.874957177 |
| RBM42 | 0.911849755 | 0.902794654 |
| LOC120824_pseudogene | 0.911848535 | 0.83192732 |
| OR10J1 | 0.911828659 | 0.936279548 |
| IGFL4 | 0.911825986 | 1.046933881 |
| LRFN2 | 0.911822451 | 0.914135277 |

| | | |
|---|---|---|
| OR2C3 | 0.911817 | 1.018226002 |
| ZNF551 | 0.911798392 | 1.052072628 |
| GLCE | 0.911795707 | 0.925475901 |
| HSD17B12 | 0.911791238 | 1.019467878 |
| ALDH18A1 | 0.911787122 | 0.81440623 |
| USP36 | 0.911687758 | 0.984425698 |
| GPR101 | 0.911651453 | 0.972744971 |
| AHSG | 0.911634553 | 0.912641316 |
| OR5A2 | 0.911625491 | 1.028434395 |
| MOBP | 0.911607412 | 1.031860226 |
| HOXB1 | 0.911588279 | 0.796678817 |
| ZNF114 | 0.911518644 | 1.02308627 |
| SUCLG2 | 0.911497591 | 0.926022064 |
| RP6-166C19.4 | 0.911484442 | 1.055841041 |
| MICU3 | 0.911474835 | 1.007290401 |
| VSTM4 | 0.911467315 | 1.059266872 |
| LRP8 | 0.911456594 | 0.914341337 |
| TMEM105 | 0.911434593 | 1.029971648 |
| RAD50 | 0.911424405 | 0.953017416 |
| LRRN1 | 0.911389816 | 1.01417578 |
| LRCH4 | 0.911351611 | 1.036857027 |
| CRISP2 | 0.911295184 | 0.851661528 |
| PDYN | 0.911290064 | 0.974306269 |
| WBP4 | 0.91127339 | 0.916409729 |
| EML3 | 0.911254366 | 0.778857837 |
| PDCD6 | 0.911211353 | 1.020116807 |
| KRTAP5-7 | 0.91117099 | 0.935251799 |
| ZNF254 | 0.911152821 | 0.938677629 |
| NKX2-1 | 0.911144081 | 0.885378696 |
| ITLN2 | 0.911125771 | 0.943816375 |
| UBQLNL | 0.911125131 | 1.046172539 |
| WIPF2 | 0.911125131 | 0.79708384 |
| TAPT1 | 0.911067976 | 0.880923451 |
| ADAM30 | 0.911043743 | 0.9733939 |
| BRPF3 | 0.911043743 | 0.9733939 |
| NR1D1 | 0.911015151 | 0.914945322 |
| FBXO18 | 0.910999289 | 0.961713173 |
| GABARAPL2 | 0.910936347 | 0.799513973 |
| BCL7C | 0.910928131 | 0.983213429 |
| POU5F1 | 0.910923714 | 0.749696233 |
| SLC16A7 | 0.910879396 | 0.955852572 |
| AWAT1 | 0.910850869 | 0.978533819 |
| CDH12 | 0.910812002 | 0.940733128 |
| ANO6 | 0.910799633 | 1.010620075 |

| | | |
|---|---|---|
| RPL7L1 | 0.910793833 | 0.80952381 |
| HRH4 | 0.910785166 | 0.909149903 |
| ROGDI | 0.910721247 | 0.959575197 |
| MYO5A | 0.910708194 | 0.891628812 |
| FAM134B | 0.910685568 | 0.992304577 |
| SLC22A6 | 0.910662339 | 0.942486837 |
| KLK9 | 0.910655178 | 1.0266061 |
| TTC36 | 0.910598005 | 1.056868791 |
| RPP40 | 0.910592568 | 0.917094896 |
| KLKB1 | 0.910572937 | 0.862426995 |
| CENPP | 0.91056647 | 0.960712839 |
| CHST5 | 0.91047977 | 0.883758607 |
| ABCF1 | 0.910439069 | 0.929121102 |
| TRPM8 | 0.910420721 | 1.109020117 |
| TRIOBP | 0.910400201 | 0.879303362 |
| POC1B | 0.910372133 | 0.913325233 |
| SQRDL | 0.910346103 | 0.947347104 |
| SDC1 | 0.91030091 | 0.98218568 |
| MEF2D | 0.910293706 | 0.983898595 |
| DEFB134 | 0.910260131 | 0.915724563 |
| PPP1R10 | 0.910224814 | 0.763872013 |
| RBM34 | 0.91017744 | 0.831915755 |
| CCDC149 | 0.910134714 | 0.944844125 |
| RUNX1 | 0.910105999 | 1.033095393 |
| DDX59 | 0.910102711 | 0.96800324 |
| PRSS37 | 0.910097464 | 0.816119887 |
| DRC1 | 0.910062672 | 0.884163629 |
| ZNF585B | 0.91002357 | 0.972250771 |
| MCM2 | 0.909944646 | 0.998053212 |
| MFAP3L | 0.909899467 | 1.004796163 |
| DCLRE1A | 0.909888254 | 0.986372485 |
| C9orf43 | 0.909875634 | 0.842754368 |
| KRTAP5-2 | 0.909868203 | 1.01336074 |
| APCDD1L | 0.90986212 | 1.022681247 |
| TDG | 0.90985549 | 0.857235561 |
| OR1D5 | 0.909844597 | 0.934909215 |
| OR5M8 | 0.909690771 | 0.798561151 |
| VCY1B | 0.909684609 | 0.9777321 |
| STK33 | 0.909684587 | 0.828033744 |
| ADORA2B | 0.909676684 | 1.062946139 |
| UGT3A1 | 0.909623668 | 1.168721609 |
| NEK5 | 0.909505287 | 1.027903958 |
| HDAC5 | 0.909497032 | 0.989064399 |
| EIF4H | 0.909483991 | 0.909680032 |

| | | |
|---|---|---|
| ZNF426 | 0.909451627 | 0.952380952 |
| DEGS2 | 0.909421539 | 0.984609154 |
| SMPDL3B | 0.909384519 | 1.004542505 |
| FCGBP | 0.909334366 | 0.790339157 |
| MPHOSPH6 | 0.909318792 | 0.900769542 |
| HAP1 | 0.909307905 | 1.095923261 |
| LYPLA1 | 0.909229698 | 0.884973674 |
| NCKAP1 | 0.909227707 | 1.022610483 |
| FAM174B | 0.909217603 | 1.026036314 |
| HNRNPM | 0.909217603 | 1.026036314 |
| IQCC | 0.909201273 | 0.993924666 |
| CATSPERD | 0.909143915 | 0.880518429 |
| ACTL10 | 0.909142525 | 0.841726619 |
| NCKAP5L | 0.909135185 | 0.949297705 |
| CACNB2 | 0.909125118 | 0.957819598 |
| KRTCAP2 | 0.909124537 | 0.952723535 |
| CD200R1L | 0.909093025 | 0.932766302 |
| ZNF433 | 0.909082988 | 0.966426859 |
| DDX53 | 0.909080188 | 1.041717294 |
| RPN1 | 0.909056214 | 0.946138871 |
| KLHL18 | 0.909006631 | 1.037262049 |
| PLAGL2 | 0.908999652 | 1.025921426 |
| SLFN11 | 0.908986815 | 0.894289186 |
| MAN1C1 | 0.908985663 | 0.934458144 |
| SOX5 | 0.908946334 | 1.014388489 |
| LAMB1 | 0.908915207 | 0.911956149 |
| MT1H | 0.908900526 | 1.031517643 |
| USE1 | 0.908891099 | 0.976103686 |
| KCNN3 | 0.90889025 | 0.817001947 |
| KHDC1L | 0.908887115 | 1.036656389 |
| C12orf10 | 0.908826219 | 1.060637205 |
| MYL4 | 0.908811946 | 0.947927372 |
| MYOG | 0.908797389 | 1.005191434 |
| PRR4 | 0.908781 | 0.599520384 |
| TAL2 | 0.908760882 | 0.966769442 |
| CCNDBP1 | 0.90875182 | 0.970195272 |
| LCT | 0.908684771 | 0.887735237 |
| HNRNPH3 | 0.908676606 | 0.874614594 |
| KCNE4 | 0.908633978 | 1.075924724 |
| SERPINE1 | 0.908633978 | 1.075924724 |
| NOV | 0.90858409 | 0.970149254 |
| CRELD2 | 0.908543381 | 0.935601458 |
| WDR4 | 0.90847951 | 0.949982871 |
| ANKRD29 | 0.908411409 | 0.979102432 |

| | | |
|---|---|---|
| FNDC8 | 0.90836723 | 1.094775213 |
| PTGER4 | 0.908358442 | 1.029201817 |
| HERC1 | 0.908328312 | 1.117456197 |
| ANKRD17 | 0.908287986 | 0.8886605 |
| INSIG2 | 0.908267234 | 1.015390846 |
| GTF2F1 | 0.908264017 | 0.898937992 |
| NOL4 | 0.908256179 | 1.052757794 |
| CSF1R | 0.908215282 | 0.823491239 |
| MYF5 | 0.908211083 | 1.005840363 |
| RWDD2A | 0.908206383 | 0.924631723 |
| CPEB3 | 0.908205953 | 1.074524099 |
| C1orf101 | 0.908204374 | 1.022276225 |
| C10orf76 | 0.908130943 | 0.960602946 |
| BMP6 | 0.908118693 | 0.820170109 |
| ARRB2 | 0.908100359 | 0.976019185 |
| SGMS1 | 0.908074262 | 0.805970149 |
| YY1 | 0.90806463 | 0.89422453 |
| POLR1A | 0.908024555 | 0.908869988 |
| ELFN2 | 0.90802104 | 1.042932361 |
| SRRM1 | 0.908020161 | 1.018842069 |
| CDK9 | 0.907976709 | 0.882543803 |
| SH3KBP1 | 0.907892071 | 0.912983899 |
| AANAT | 0.907853142 | 0.953277093 |
| PKM | 0.907853142 | 0.953277093 |
| DAPL1 | 0.907851655 | 0.935251799 |
| DCPS | 0.907844787 | 0.940866748 |
| IHH | 0.90781091 | 0.947436729 |
| RDH14 | 0.90780215 | 0.95909275 |
| OR4D2 | 0.907776214 | 0.979787598 |
| MESP2 | 0.907711104 | 0.81740322 |
| STX10 | 0.907707298 | 1.024323398 |
| H1FOO | 0.907660629 | 1.056868791 |
| PLTP | 0.907636714 | 0.924075276 |
| ZNF674 | 0.907599008 | 0.882494005 |
| LOC728137 | 0.907580205 | 0.894484412 |
| SAC3D1 | 0.907554887 | 0.916160389 |
| UNC5D | 0.907536481 | 0.923603974 |
| MOGAT3 | 0.907528653 | 0.904819765 |
| TMEM196 | 0.907519173 | 0.935594382 |
| BBS10 | 0.907513676 | 0.863912515 |
| CAPN5 | 0.907500222 | 0.906554186 |
| FAXDC2 | 0.907476808 | 0.966426859 |
| AASS | 0.907426314 | 0.829890644 |
| MUC7 | 0.90737604 | 1.048646797 |

| | | |
|---|---|---|
| RAB7A | 0.90737191 | 0.907249899 |
| PDK3 | 0.907298366 | 1.042180402 |
| ALG9 | 0.907271354 | 0.961927906 |
| FAM134A | 0.907210429 | 0.998379911 |
| MOB1A | 0.907199921 | 0.923946557 |
| MSRB1 | 0.907183329 | 0.957472661 |
| NOSTRIN | 0.907172645 | 0.949640288 |
| LRRC10 | 0.907145142 | 0.977046934 |
| NEK3 | 0.907095851 | 0.936404932 |
| OS9 | 0.907086135 | 0.834750911 |
| NUP214 | 0.907019506 | 0.84163629 |
| CCDC185 | 0.90701757 | 1.037262049 |
| MPP5 | 0.907000097 | 0.918995545 |
| GDAP1 | 0.906983644 | 0.9963548 |
| MUCL1 | 0.906874071 | 1.010125557 |
| MAB21L1 | 0.906872469 | 0.912298732 |
| DTNB | 0.90683169 | 0.970537855 |
| CDCA2 | 0.906809851 | 1.004796163 |
| THSD4 | 0.906727364 | 1.060348319 |
| RPS6KL1 | 0.906696418 | 0.889682025 |
| TAAR6 | 0.906688845 | 1.036988968 |
| MEST | 0.906663586 | 0.86472256 |
| AASDH | 0.906636384 | 0.71000405 |
| CENPB | 0.906597548 | 1.025308241 |
| PDE1A | 0.906569064 | 0.948734588 |
| CRYBB2 | 0.906532058 | 0.912641316 |
| ELAVL4 | 0.906521546 | 0.941760877 |
| ZNF207 | 0.906513934 | 0.964028777 |
| ZNF79 | 0.906509448 | 0.9777321 |
| TRIM9 | 0.906501836 | 1.001712915 |
| PSMB8 | 0.906402113 | 1.072680078 |
| UFSP1 | 0.906377863 | 0.946942082 |
| NDUFV1 | 0.90636375 | 0.925373134 |
| SPRYD3 | 0.906283216 | 0.876468206 |
| TRPM4 | 0.906261011 | 0.984425698 |
| BMP3 | 0.906210547 | 0.978585334 |
| STAG3 | 0.906198961 | 0.769542325 |
| HIST3H3 | 0.906191919 | 0.969510106 |
| MRPS18B | 0.906191919 | 0.904419322 |
| NODAL | 0.906191919 | 0.815347722 |
| S100A12 | 0.906191919 | 0.979787598 |
| SHISA2 | 0.906191919 | 1.108256252 |
| UBAP1 | 0.906191919 | 0.993490922 |
| ANAPC10 | 0.906136916 | 0.94491697 |

| | | |
|---|---|---|
| RDH8 | 0.906121802 | 1.037637897 |
| SOX8 | 0.906075788 | 0.970028352 |
| VPS37B | 0.906050623 | 0.819765087 |
| NAT2 | 0.906002927 | 0.955223881 |
| ACMSD | 0.905982975 | 0.844876468 |
| LUZP2 | 0.905950271 | 1.056703119 |
| CEP95 | 0.905927144 | 0.990684488 |
| DHODH | 0.905924144 | 1.014276444 |
| MAGEE2 | 0.905876292 | 0.983556012 |
| ZDHHC1 | 0.905870085 | 0.964713943 |
| RP6-166C19.2 | 0.905857625 | 0.92874272 |
| FNBP1L | 0.905849206 | 0.931551235 |
| FFAR3 | 0.905805681 | 1.067488644 |
| GINS3 | 0.905757564 | 0.890643985 |
| FAM208A | 0.905735813 | 1.036452005 |
| ENGASE | 0.905728544 | 0.926022064 |
| CANX | 0.905626817 | 1.098663926 |
| AMZ2 | 0.905603216 | 0.947752126 |
| ANP32D | 0.905597094 | 1.043850634 |
| CDKN1C | 0.905555501 | 0.9733939 |
| CLPB | 0.905555501 | 0.843608047 |
| S100A1 | 0.905554232 | 0.973621103 |
| LDB3 | 0.905542813 | 0.956491949 |
| PCDH12 | 0.905533389 | 0.942788626 |
| AGA | 0.905509537 | 0.979748886 |
| SNX17 | 0.905508514 | 0.908530319 |
| FOXD4 | 0.90550592 | 0.943296881 |
| LOC390688_pseudogene | 0.905500721 | 0.898252826 |
| TARS | 0.905444447 | 0.961713173 |
| KIF21B | 0.905387808 | 0.95587281 |
| SLC6A20 | 0.905357596 | 1.0798183 |
| ZFC3H1 | 0.905351062 | 0.90927501 |
| NEUROG1 | 0.905345762 | 1.100719424 |
| ENPP6 | 0.905290749 | 1.00908501 |
| RNF212 | 0.905280917 | 0.995949777 |
| PARP1 | 0.905236004 | 1.003244646 |
| DEFB104A | 0.905233594 | 0.86836776 |
| ORM1 | 0.905189852 | 0.929427886 |
| MS4A5 | 0.905186136 | 0.926002055 |
| GABRB2 | 0.905097866 | 1.050616483 |
| LUC7L3 | 0.905087446 | 1.066828676 |
| LDOC1L | 0.90506921 | 0.772782503 |
| RASGEF1B | 0.90500372 | 0.783830079 |
| PTMA | 0.905003599 | 0.841231268 |

| | | |
|---|---|---|
| MRGPRE | 0.90498994 | 1.038935756 |
| ARC | 0.904973188 | 0.946132037 |
| MLYCD | 0.904970793 | 0.724989874 |
| LY6D | 0.90496276 | 1.010277492 |
| IL15RA | 0.904947921 | 0.909680032 |
| MAPK15 | 0.904947708 | 0.791044776 |
| C5orf64 | 0.904939426 | 0.991435423 |
| REEP6 | 0.904925304 | 1.014580802 |
| FAM155B | 0.90492402 | 0.979445015 |
| LZTS3 | 0.904880478 | 0.946899623 |
| OR9G9 | 0.904861221 | 0.9331963 |
| C1QBP | 0.904739701 | 1.068859198 |
| C2CD2 | 0.90471179 | 0.838985954 |
| ENY2 | 0.904663726 | 0.976103686 |
| SEPHS2 | 0.904650851 | 1.062946139 |
| LYPD8 | 0.904626835 | 0.991778006 |
| TTC16 | 0.904624161 | 0.990065091 |
| PEX13 | 0.904607998 | 0.921425678 |
| TMEM8C | 0.904602649 | 1.008100446 |
| PIK3CA | 0.904595365 | 1.057105775 |
| CDKN2C | 0.904592072 | 0.998053212 |
| SLC25A5 | 0.904590118 | 0.859862292 |
| COL14A1 | 0.904584158 | 0.989874443 |
| NPHP4 | 0.904510661 | 0.923261391 |
| FAM178A | 0.904485378 | 0.909558068 |
| ZNF229 | 0.904452619 | 0.892428914 |
| SAP18 | 0.904448006 | 0.868267359 |
| LSMEM2 | 0.904304954 | 0.882948562 |
| TMEM260 | 0.90425081 | 0.959575197 |
| LOC196346_pseudogene | 0.904236847 | 0.850746269 |
| HOXC9 | 0.904205894 | 0.791693705 |
| ERN1 | 0.904188688 | 1.016223232 |
| EEF2 | 0.904164485 | 0.844905905 |
| SPATA7 | 0.904077093 | 0.880781089 |
| TSPAN16 | 0.904050559 | 0.974483597 |
| CRYGA | 0.904035816 | 1.007879411 |
| ZKSCAN2 | 0.904029969 | 0.945489942 |
| C6orf57 | 0.904009174 | 0.853374443 |
| TBL1XR1 | 0.903967761 | 0.977046934 |
| KAT6A | 0.903959923 | 0.973621103 |
| PPHLN1 | 0.903959923 | 0.973621103 |
| EPG5 | 0.903928025 | 0.95991778 |
| MAST2 | 0.903917515 | 0.880597015 |
| CD99 | 0.903899434 | 0.947927372 |

| | | |
|---|---|---|
| SLC25A12 | 0.903877671 | 0.987021415 |
| PLGLB1 | 0.903841297 | 1.04090725 |
| FAM9C | 0.903777367 | 0.979343864 |
| TRIM37 | 0.903719775 | 0.756651525 |
| NECAP1 | 0.903715931 | 1.003083248 |
| SKI | 0.903715931 | 1.003083248 |
| ZNF675 | 0.903715067 | 1.068448765 |
| SLC13A2 | 0.903702841 | 0.892669097 |
| OR4N3P | 0.903698359 | 0.842446335 |
| TBXAS1 | 0.903634406 | 0.750811162 |
| GSTT1 | 0.903615225 | 0.963659961 |
| C1orf122 | 0.903606819 | 0.780882949 |
| TYRP1 | 0.903547637 | 1.011031798 |
| TMEM69 | 0.903506875 | 0.984204131 |
| TMTC1 | 0.90348441 | 0.908869988 |
| SLC39A10 | 0.903422823 | 0.896882494 |
| OR8G5 | 0.9033931 | 0.998287085 |
| KLHL8 | 0.903273772 | 0.925070879 |
| GALT | 0.903248228 | 1.08825438 |
| FBLN1 | 0.903241222 | 0.946899623 |
| MYBPH | 0.903235875 | 0.945186708 |
| MAGEA5 | 0.903235377 | 1.050017129 |
| MYO6 | 0.903232833 | 1.03504218 |
| C12orf55 | 0.903194662 | 0.799513973 |
| CPT1B | 0.903194662 | 0.970838396 |
| C14orf39 | 0.903120064 | 1.010620075 |
| CCT3 | 0.903106077 | 0.991494532 |
| NINJ2 | 0.903093785 | 1.002055498 |
| CORO1B | 0.90305692 | 1.062373431 |
| FAM186B | 0.903049024 | 1.00526529 |
| C9orf163 | 0.90303038 | 0.891049008 |
| LRP2BP | 0.902999103 | 0.897934386 |
| DSCC1 | 0.90296645 | 1.025921426 |
| CNTN3 | 0.902955545 | 0.959232614 |
| WT1-AS | 0.902943884 | 0.758606723 |
| SLC25A25 | 0.902940092 | 0.857884491 |
| GRIK5 | 0.902935893 | 0.958468527 |
| GPATCH2L | 0.902921744 | 0.854402193 |
| KLF2 | 0.902920706 | 0.854597003 |
| OR2A2 | 0.902908604 | 1.04008222 |
| GBP3 | 0.902869307 | 0.989469421 |
| FBXL15 | 0.902863696 | 0.952628164 |
| SPTB | 0.902860304 | 0.931825968 |
| PKNOX2 | 0.902836788 | 1.01781432 |

| | | |
|---|---|---|
| RNASE3 | 0.902826823 | 0.875253139 |
| MPRIP | 0.902818011 | 0.971243418 |
| SLC22A25 | 0.902798755 | 0.850141758 |
| DNAH1 | 0.902773536 | 0.998972251 |
| DEFB104B | 0.902743994 | 0.990407674 |
| BNIPL | 0.902733165 | 0.89756766 |
| HSPA9 | 0.902666943 | 1.03569111 |
| DEDD2 | 0.902658405 | 0.966426859 |
| HEATR6 | 0.902580833 | 1.001215067 |
| PALM2-AKAP2 | 0.902568465 | 0.942446043 |
| GHR | 0.902566097 | 0.92926671 |
| PADI4 | 0.902546998 | 0.976638546 |
| MAN1A1 | 0.902501723 | 0.886998785 |
| TBC1D10A | 0.90249831 | 1.092750101 |
| LAPTM5 | 0.902483486 | 0.925880923 |
| MAGEB2 | 0.902476758 | 1.002740665 |
| ADAMTS12 | 0.902473813 | 0.970798183 |
| FBXO32 | 0.9024599 | 1.018170019 |
| LCN8 | 0.902447421 | 0.900769542 |
| NDUFAF7 | 0.902431811 | 0.908187736 |
| OR52N5 | 0.902425241 | 0.989037342 |
| TTC18 | 0.902407616 | 1.066461117 |
| BCOR | 0.902381518 | 0.896197328 |
| ACBD4 | 0.90236882 | 0.85054678 |
| KLHL36 | 0.902345814 | 1.049331963 |
| SUCO | 0.902314314 | 0.864317537 |
| SDE2 | 0.902297962 | 0.832320778 |
| RBMS2 | 0.902194842 | 1.087358684 |
| BTBD11 | 0.902182246 | 1.006509078 |
| DNALI1 | 0.90218024 | 0.937626569 |
| F2RL3 | 0.902172345 | 0.947436729 |
| QSER1 | 0.902130777 | 0.990279465 |
| ATP11B | 0.902101712 | 1.036340039 |
| CTNND1 | 0.901989091 | 0.960260363 |
| NR1H3 | 0.901909487 | 0.835171966 |
| RAX | 0.901885037 | 1.018818949 |
| SCLY | 0.901876789 | 0.910490077 |
| IFI6 | 0.901874446 | 0.786958283 |
| SNURF | 0.901865577 | 0.932853717 |
| ENPP1 | 0.901854834 | 0.917375456 |
| PTPRH | 0.901838442 | 0.876703439 |
| GPX4 | 0.901793412 | 0.965606749 |
| MMP16 | 0.901793412 | 0.965606749 |
| GBP4 | 0.901769916 | 0.94491697 |

| | | |
|---|---|---|
| EPS8L2 | 0.901738892 | 0.836587873 |
| SPINK14 | 0.901735267 | 1.044878383 |
| GGT2 | 0.901704996 | 1.096041531 |
| CAND1 | 0.901660706 | 1.001297859 |
| CCM2 | 0.901643062 | 0.9554642 |
| OR2AE1 | 0.901607449 | 1.015758822 |
| AK1 | 0.901607279 | 0.906554186 |
| TRIM44 | 0.901504365 | 0.995544755 |
| GALNT4 | 0.901418786 | 1.029971648 |
| FRS3 | 0.901388784 | 0.969510106 |
| OR2AG2 | 0.901354576 | 0.962658445 |
| SNRNP48 | 0.901338581 | 0.895512162 |
| SNRPA1 | 0.901319914 | 0.955806783 |
| SLC22A17 | 0.901268258 | 0.835560956 |
| WFDC1 | 0.901159246 | 0.986981843 |
| PCBP4 | 0.901157921 | 1.041717294 |
| ZNF41 | 0.901155736 | 1.007787151 |
| PTPRN2 | 0.901136966 | 0.918883842 |
| DLX4 | 0.901107847 | 0.897529364 |
| RGS17 | 0.901105601 | 0.960415315 |
| SEC24C | 0.901084621 | 0.850976362 |
| SCAND1 | 0.900990921 | 1.014731072 |
| CYP2D6 | 0.90097339 | 1.037637897 |
| KLHL12 | 0.90090225 | 0.939020212 |
| C7ORF28B | 0.900901661 | 0.997601918 |
| EIF1AX | 0.900810051 | 0.650465776 |
| PCDHA9 | 0.900798895 | 0.916160389 |
| SOX15 | 0.900783358 | 1.055893074 |
| SMARCD3 | 0.900766676 | 0.937053861 |
| RNF146 | 0.900642944 | 0.717715769 |
| CD33 | 0.900621013 | 1.003083248 |
| TOLLIP | 0.90056338 | 0.93764988 |
| TMIGD1 | 0.900447 | 0.969218307 |
| MOB1B | 0.900412227 | 1.020554985 |
| KLHL40 | 0.900410777 | 1.073997945 |
| MYLIP | 0.900410299 | 1.038286827 |
| IGF2BP2 | 0.900395398 | 0.836371 |
| GJD4 | 0.900383047 | 0.908530319 |
| NT5C3B | 0.900360402 | 0.857027136 |
| TXNL1 | 0.900329458 | 1.032446463 |
| AIM2 | 0.900323556 | 1.005138746 |
| TNNI3 | 0.900313981 | 0.866320571 |
| BLVRA | 0.900309124 | 0.824789098 |
| TEAD3 | 0.900291095 | 0.700194679 |

| | | |
|---|---|---|
| TMPRSS4 | 0.900247711 | 1.0266061 |
| ADSS | 0.900245481 | 0.932766302 |
| GTF2IRD2 | 0.900201628 | 0.932853717 |
| UNKL | 0.900134932 | 0.922576225 |
| PCDHA4 | 0.900110102 | 1.020897568 |
| OR5AN1 | 0.900099817 | 1.019184652 |
| CYBA | 0.900098467 | 0.841231268 |
| IRX1 | 0.900089622 | 1.017471737 |
| SUCLA2 | 0.89998752 | 0.930741191 |
| FBXO46 | 0.899979008 | 0.967553537 |
| CISD3 | 0.899974536 | 0.998629668 |
| LYSMD4 | 0.899967959 | 0.829890644 |
| TATDN3 | 0.899953105 | 0.995203837 |
| FER1L6 | 0.899904232 | 0.884973674 |
| CBX3 | 0.89988224 | 1.033230558 |
| C2orf69 | 0.89987703 | 1.04090725 |
| PLA2G16 | 0.89987114 | 0.9331963 |
| AKAP7 | 0.899865978 | 0.9201817 |
| CPA4 | 0.899865978 | 0.9201817 |
| NSFL1C | 0.899808159 | 1.021240151 |
| PKHD1L1 | 0.899786637 | 1.01781432 |
| TEK | 0.899736813 | 0.99156392 |
| TMOD3 | 0.899728822 | 0.912641316 |
| MSRA | 0.899648233 | 0.985723556 |
| REN | 0.899640083 | 0.8669695 |
| ABCG2 | 0.899581883 | 0.951802349 |
| LGALS7B | 0.899581883 | 0.951802349 |
| EXO5 | 0.899563901 | 0.979343864 |
| SPSB4 | 0.89953792 | 0.861129137 |
| MYO9B | 0.899523251 | 1.048197651 |
| OVOS2 | 0.899508427 | 1.075739166 |
| RAD9A | 0.899479553 | 0.896820247 |
| CAT | 0.899449439 | 0.961117861 |
| FADS3 | 0.899379777 | 0.890979883 |
| MAEL | 0.899351277 | 0.894694208 |
| BVES | 0.899349082 | 0.952723535 |
| MSMB | 0.899311928 | 0.947584789 |
| TMC7 | 0.899286963 | 0.944158959 |
| MBNL1 | 0.899274412 | 0.89756766 |
| NFS1 | 0.899208746 | 0.917780478 |
| DDX3X | 0.89919922 | 0.952207371 |
| S1PR2 | 0.89919916 | 1.033744322 |
| UXS1 | 0.899197387 | 0.95909275 |
| SSTR2 | 0.899176191 | 0.879299156 |

| | | |
|---|---|---|
| SUOX | 0.899093292 | 0.950681376 |
| TPTE2 | 0.899072382 | 0.873458793 |
| C1orf159 | 0.89906372 | 1.016605913 |
| LYZL4 | 0.899062904 | 1.023491292 |
| RAB33A | 0.899056329 | 0.986372485 |
| KIAA1586 | 0.899012296 | 0.994518671 |
| ALDH1L1 | 0.899005618 | 0.83192732 |
| PRSS58 | 0.898996809 | 0.944841012 |
| CAPZA2 | 0.898986877 | 0.947927372 |
| OPRL1 | 0.898967255 | 0.86761843 |
| NFASC | 0.898944265 | 0.998379911 |
| SEC11A | 0.898899091 | 0.939000649 |
| NDUFB7 | 0.898827102 | 1.035236938 |
| ANKRD35 | 0.898823893 | 1.007695423 |
| ID2 | 0.89881778 | 0.959497772 |
| KDM8 | 0.898795716 | 0.965399109 |
| CIPC | 0.898716153 | 0.955121617 |
| TMBIM1 | 0.898690124 | 0.955042527 |
| ANXA8 | 0.89868797 | 0.868790682 |
| ICK | 0.898676736 | 0.963011032 |
| HARBI1 | 0.89867041 | 0.88618874 |
| SIGMAR1 | 0.898655168 | 0.998702141 |
| NDUFB6 | 0.8986529 | 1.029462145 |
| ADCY7 | 0.898579019 | 0.957170668 |
| KLRF1 | 0.898552017 | 0.808565866 |
| SRRM3 | 0.898537678 | 0.932853717 |
| PRSS55 | 0.898523587 | 0.931140802 |
| KLHL41 | 0.898517697 | 0.970880439 |
| VPS53 | 0.898492578 | 0.80599433 |
| REPIN1 | 0.898481086 | 0.926002055 |
| TOP3B | 0.898438582 | 0.959902795 |
| GNA15 | 0.898403688 | 0.897934386 |
| ZCCHC7 | 0.898350504 | 0.910585817 |
| SNX9 | 0.898278292 | 0.939649578 |
| MAP3K2 | 0.898263721 | 1.011031798 |
| ALKBH5 | 0.89825805 | 1.061158364 |
| SHANK2 | 0.898255535 | 0.860568688 |
| HMGXB4 | 0.89823288 | 1.092154848 |
| IL37 | 0.898200801 | 0.971223022 |
| PEX6 | 0.898184197 | 0.862426995 |
| NTPCR | 0.898084344 | 0.987849332 |
| UGGT2 | 0.898056428 | 0.820170109 |
| RRAS | 0.897966205 | 0.922128488 |
| SPG11 | 0.897947722 | 1.095990279 |

| | | |
|---|---|---|
| PRDM15 | 0.897931526 | 1.014731072 |
| PRH1 | 0.897913267 | 0.974991435 |
| ACTB | 0.897908101 | 1.049331963 |
| C10orf99 | 0.897887803 | 0.897225077 |
| LBX2 | 0.897871899 | 0.895512162 |
| GJC1 | 0.897859514 | 0.916288125 |
| SLC22A5 | 0.897859514 | 0.916288125 |
| C2orf57 | 0.897780283 | 0.959575197 |
| DYRK3 | 0.897624481 | 0.868916288 |
| ZNF197 | 0.897613198 | 0.820575132 |
| STAB2 | 0.897597957 | 0.975334018 |
| MIIP | 0.897558183 | 0.963142973 |
| POU5F2 | 0.897506432 | 0.965056526 |
| OR2C1 | 0.897475395 | 0.961630695 |
| PDCD1LG2 | 0.897420542 | 1.115026326 |
| HECA | 0.897409877 | 1.131209318 |
| RGS5 | 0.897393973 | 1.058581706 |
| KRTAP12-2 | 0.897391761 | 0.938031592 |
| SNX29 | 0.897342411 | 0.877012676 |
| KIN | 0.897293114 | 1.046591298 |
| SLC25A38 | 0.897264971 | 1.011745646 |
| TNFSF15 | 0.897251293 | 0.972250771 |
| SUPT4H1 | 0.897242007 | 1.053861129 |
| LRRK1 | 0.897238056 | 0.815704088 |
| GNA14 | 0.897148369 | 0.878493317 |
| ANKS4B | 0.897059784 | 0.772782503 |
| ECE2 | 0.897039327 | 0.940947437 |
| PMEPA1 | 0.896998554 | 1.060348319 |
| ANKRD34A | 0.896992032 | 0.877355259 |
| LCN15 | 0.89698442 | 1.045220966 |
| FGFRL1 | 0.896970999 | 0.970798183 |
| ARL8A | 0.896965697 | 1.104090725 |
| DPP6 | 0.896954535 | 0.869565217 |
| CD302 | 0.89683417 | 0.805184285 |
| MPZ | 0.896819719 | 0.960602946 |
| CD9 | 0.89681904 | 0.728674203 |
| HLA-DPB1 | 0.896817302 | 0.92926671 |
| CDH4 | 0.896768293 | 1.021240151 |
| OTUD7B | 0.896744446 | 1.024659312 |
| ENAM | 0.896659323 | 1.042137718 |
| OST4 | 0.896649627 | 0.943473792 |
| FOXP3 | 0.896645082 | 0.953277093 |
| NUDT8 | 0.89657473 | 1.000685166 |
| NELFCD | 0.896565104 | 0.9181855 |

| | | |
|---|---|---|
| IER5L | 0.89656327 | 1.019441069 |
| ATP5S | 0.896544464 | 0.9331963 |
| SHISA6 | 0.896536942 | 0.900308325 |
| KCTD5 | 0.896533781 | 0.947436729 |
| ARHGAP26 | 0.896530719 | 1.005670312 |
| RNASE1 | 0.896519127 | 0.750811162 |
| FAM110A | 0.896505657 | 0.796273795 |
| RNF5 | 0.896473602 | 0.91174562 |
| SEBOX | 0.896471198 | 0.862281603 |
| PLA2G4D | 0.896458129 | 0.927501013 |
| ZNF709 | 0.896392926 | 0.856217092 |
| SIRPB1 | 0.896363228 | 0.987444309 |
| COL19A1 | 0.896292574 | 0.778047793 |
| PACS1 | 0.896286426 | 0.971565605 |
| PRSS12 | 0.896281789 | 1.031148605 |
| TVP23A | 0.896270069 | 0.938677629 |
| CLEC9A | 0.896268887 | 0.771162414 |
| OR10G9 | 0.896233776 | 1.059952038 |
| CERS1 | 0.896185294 | 1.054813292 |
| WDR83OS | 0.896169757 | 1.093560146 |
| TACC2 | 0.896166989 | 1.021925317 |
| ARPC2 | 0.89616264 | 0.959575197 |
| SOX10 | 0.896161268 | 0.864373783 |
| RCOR2 | 0.89610272 | 0.854191981 |
| DUSP16 | 0.896102078 | 1.085009734 |
| TSPEAR | 0.896053489 | 1.041109969 |
| SPAG5 | 0.896047961 | 1.009934909 |
| DEC1 | 0.896009946 | 0.914696814 |
| EIF3M | 0.896001298 | 0.907249899 |
| BNIP3L | 0.895961828 | 0.940733128 |
| ZNF639 | 0.895849369 | 1.020554985 |
| MRPL28 | 0.895831925 | 1.018842069 |
| DNMT1 | 0.895818321 | 0.815309842 |
| OR5B17 | 0.895805736 | 0.956491949 |
| CD59 | 0.895746788 | 0.980815348 |
| SRRM4 | 0.895730794 | 0.949640288 |
| EIF3A | 0.895718122 | 1.031797534 |
| SAA2 | 0.895711945 | 0.947927372 |
| PFDN5 | 0.895651492 | 0.884568651 |
| LRMP | 0.895616387 | 0.939362795 |
| HNRNPA3 | 0.895599431 | 0.870797894 |
| PCM1 | 0.895591344 | 0.900713822 |
| PBK | 0.895566483 | 1.085658663 |
| CTSW | 0.895543185 | 0.960415315 |

| | | |
|---|---|---|
| VWA5B1 | 0.895540176 | 0.786913326 |
| ULBP1 | 0.895530843 | 1.019184652 |
| SMIM12 | 0.895528777 | 0.923855812 |
| CASP10 | 0.895521369 | 0.990266061 |
| FLVCR2 | 0.89551813 | 0.850141758 |
| HIGD1A | 0.895483899 | 0.946942082 |
| DIP2B | 0.895450686 | 0.903199676 |
| RTP4 | 0.895447273 | 1.110571081 |
| MRPL23 | 0.895409271 | 1.029971648 |
| C17orf102 | 0.895339319 | 1.029804728 |
| DMRT3 | 0.895292429 | 0.978585334 |
| RAB40B | 0.895278351 | 1.00843608 |
| SERF2 | 0.895257928 | 0.921830701 |
| RALGAPB | 0.89523624 | 0.991778006 |
| ZNF654 | 0.895219519 | 0.94491697 |
| GOLGA8A | 0.895193285 | 0.841231268 |
| ZFPM2 | 0.895187219 | 0.942894225 |
| SMIM3 | 0.895155113 | 1.032446463 |
| WBSCR28 | 0.895004345 | 1.054470709 |
| APOC3 | 0.894956158 | 0.912920211 |
| CELF1 | 0.894956158 | 0.912920211 |
| CFD | 0.894950952 | 0.871512005 |
| ANKK1 | 0.894934899 | 0.99091499 |
| SSBP3 | 0.894888035 | 1.153477218 |
| CLDND1 | 0.894841084 | 1.035236938 |
| RBM3 | 0.894818531 | 0.928057554 |
| GALNT1 | 0.894817671 | 0.955223881 |
| HSD3B7 | 0.894817671 | 1.104477612 |
| TUBB6 | 0.894817671 | 1.074626866 |
| WBSCR27 | 0.89479641 | 0.848521669 |
| NDUFA4 | 0.894784745 | 0.758606723 |
| VWA3B | 0.894761848 | 0.869133265 |
| ZNF589 | 0.89472918 | 0.974991435 |
| VPREB1 | 0.894725555 | 0.893456663 |
| GPR142 | 0.894709004 | 1.068786502 |
| DDOST | 0.894695338 | 0.949383517 |
| ADRB1 | 0.894691469 | 0.919532771 |
| SLC9A3R1 | 0.894667378 | 1.023638232 |
| PDE11A | 0.89456993 | 0.943296881 |
| SLC35D2 | 0.894446675 | 1.030832477 |
| ZBTB9 | 0.894423159 | 1.055155875 |
| GTF2I | 0.89436948 | 0.795868773 |
| PCDHB7 | 0.894365616 | 0.840013703 |
| METAP1 | 0.894362835 | 1.021414666 |

| | | |
|---|---|---|
| GJB7 | 0.894354877 | 0.944158959 |
| CBWD6 | 0.894352567 | 0.970195272 |
| CCR2 | 0.894349169 | 0.99156392 |
| RBM39 | 0.894329645 | 0.788983394 |
| NMUR2 | 0.894302656 | 0.902011681 |
| LIN9 | 0.894289496 | 0.965056526 |
| OPN3 | 0.894285039 | 0.872160934 |
| KIR3DL2 | 0.894268291 | 1.09352518 |
| OR10K1 | 0.894268291 | 0.859198356 |
| INCENP | 0.894242102 | 0.779376499 |
| ZSWIM8 | 0.89422579 | 0.95991778 |
| ORC2 | 0.894207712 | 1.010277492 |
| C10orf82 | 0.894161405 | 0.954779034 |
| CNNM1 | 0.894159684 | 1.315861596 |
| ARID1B | 0.894099603 | 0.847207948 |
| ASNA1 | 0.894097836 | 0.806619079 |
| APOA4 | 0.89407199 | 0.84690158 |
| BEGAIN | 0.894052527 | 0.946214457 |
| ID3 | 0.894035106 | 0.890330954 |
| ONECUT1 | 0.894034192 | 0.939246659 |
| MTPAP | 0.893987326 | 1.017471737 |
| LRRC46 | 0.89395442 | 0.948562171 |
| HOXD12 | 0.893930682 | 1.04090725 |
| NLRP5 | 0.89382926 | 1.020251114 |
| ZBTB48 | 0.893786623 | 0.729445119 |
| LHX3 | 0.893762732 | 0.960307817 |
| CHIC1 | 0.893759872 | 0.923946557 |
| TUFM | 0.893727227 | 0.907249899 |
| BAAT | 0.893723559 | 0.999594978 |
| IL17RB | 0.893686961 | 1.129785853 |
| PSMB3 | 0.893674859 | 1.016223232 |
| BEX5 | 0.893609708 | 0.932766302 |
| GABRB3 | 0.893603485 | 0.956521739 |
| OXSR1 | 0.893603485 | 0.956521739 |
| CAPN7 | 0.893589655 | 1.040233615 |
| ETS2 | 0.8935778 | 0.842959117 |
| PSMB10 | 0.893564464 | 0.926670993 |
| PTF1A | 0.89354891 | 0.908530319 |
| GPR42_pseudogene | 0.893543966 | 1.094094744 |
| C7 | 0.893517919 | 0.979787598 |
| LIAS | 0.893514823 | 0.980532122 |
| MAP7D2 | 0.893512157 | 0.826650466 |
| CACNG6 | 0.893473908 | 0.950681376 |
| SPTBN1 | 0.893453372 | 0.773592548 |

| | | |
|---|---|---|
| ADAM10 | 0.893391256 | 1.058403634 |
| ACTL9 | 0.893353835 | 0.97691373 |
| ILDR1 | 0.893328026 | 0.868790682 |
| MAL | 0.893292413 | 0.962658445 |
| ABHD12 | 0.893285473 | 0.900769542 |
| CYTH3 | 0.893265318 | 0.936622131 |
| NHSL2 | 0.893263773 | 0.854597003 |
| FAM131B | 0.89318451 | 0.863912515 |
| CIB2 | 0.893180097 | 0.992861778 |
| PLBD1 | 0.893144205 | 0.999314834 |
| TMTC4 | 0.893138256 | 0.857027136 |
| ZNF616 | 0.893121822 | 0.997601918 |
| ZNF507 | 0.893120988 | 0.958687728 |
| KIAA0195 | 0.893104831 | 0.877697842 |
| GRASP | 0.893097627 | 0.924631723 |
| SPI1 | 0.892979804 | 1.011031798 |
| ZNF175 | 0.892940856 | 1.030832477 |
| SEC22A | 0.892929962 | 0.908059943 |
| PHB2 | 0.892876653 | 0.897469176 |
| CX3CL1 | 0.892833517 | 1.138746146 |
| TP53TG5 | 0.892832067 | 0.952723535 |
| MRPL41 | 0.892829802 | 1.0222679 |
| ARL6IP5 | 0.892799897 | 0.973621103 |
| RSPO4 | 0.892788959 | 0.926690968 |
| RBM4 | 0.892776336 | 0.971908188 |
| ALS2 | 0.892701597 | 0.991494532 |
| DAPK1 | 0.892535012 | 1.01752109 |
| CYP24A1 | 0.89253273 | 1.003240178 |
| DNAH8 | 0.892515665 | 1.019441069 |
| AP1G2 | 0.892336788 | 0.941075711 |
| SPDL1 | 0.892319393 | 0.714459295 |
| DAZAP2 | 0.892311551 | 0.939362795 |
| CXCL11 | 0.892307835 | 0.916752312 |
| USP16 | 0.892231486 | 0.898118105 |
| RALY | 0.892203261 | 0.909900651 |
| MYL6 | 0.892200753 | 0.827865533 |
| CLDN15 | 0.892178658 | 0.996916752 |
| LRP1B | 0.892160358 | 0.948157149 |
| ELK3 | 0.892148387 | 0.96435804 |
| PGA5 | 0.892146611 | 1 |
| SOCS7 | 0.892117222 | 0.946138871 |
| CDKN2D | 0.892084457 | 0.892277742 |
| SOCS5 | 0.892064453 | 0.970149254 |
| TANGO2 | 0.891956495 | 1.024706359 |

| | | |
|---|---|---|
| LIMS3 | 0.891944543 | 1.045906132 |
| KIF26A | 0.891939175 | 0.848926691 |
| HOOK2 | 0.891895112 | 1.050222762 |
| SNX4 | 0.891890324 | 0.825840421 |
| GRAMD3 | 0.891883601 | 0.976361768 |
| COL11A2 | 0.891872183 | 0.997259335 |
| RER1 | 0.891818265 | 0.950325454 |
| WDSUB1 | 0.891791963 | 1.036340039 |
| C1orf56 | 0.891763432 | 0.997164844 |
| TMA7 | 0.891679024 | 0.684892669 |
| GALM | 0.891625842 | 0.895504253 |
| ZNF345 | 0.89161703 | 0.944106926 |
| MRPL38 | 0.89160893 | 0.992709599 |
| SBNO2 | 0.891606536 | 1.00891049 |
| SCXB | 0.891590992 | 0.999314834 |
| ZNF383 | 0.891563186 | 0.937221547 |
| HIST2H2AB | 0.891544142 | 1.102089757 |
| MYCBPAP | 0.891540744 | 0.995889003 |
| EFNA5 | 0.891495352 | 1.048669695 |
| UBE2E3 | 0.891461435 | 1.072680078 |
| OR6T1 | 0.891415281 | 0.966426859 |
| TRAIP | 0.891398046 | 0.713173264 |
| CABP1 | 0.891396976 | 0.839328537 |
| RANGRF | 0.891362722 | 0.963001028 |
| ANKRD40 | 0.891343583 | 0.942081814 |
| AKR1C6P | 0.891327413 | 0.96495782 |
| NSUN5P1 | 0.891298348 | 1 |
| RYK | 0.891294813 | 0.988968202 |
| MAGEA9B | 0.891256426 | 0.956149366 |
| PPP4C | 0.891233976 | 1.036988968 |
| FAM13C | 0.891202779 | 0.952723535 |
| TBX22 | 0.891187247 | 0.970028352 |
| TLR7 | 0.891155769 | 0.983127839 |
| PXDNL | 0.891127696 | 1.007138222 |
| PCDHGC5 | 0.891121449 | 0.947584789 |
| CDC6 | 0.891095759 | 0.865937627 |
| ATP6V0A4 | 0.891055312 | 0.907654921 |
| FAF2 | 0.891020062 | 1.072093965 |
| CUZD1 | 0.891012796 | 1.022610483 |
| ATP13A5 | 0.891010943 | 0.933171324 |
| KIAA2013 | 0.890998514 | 0.794248684 |
| TRIP11 | 0.890976915 | 0.974888619 |
| MTO1 | 0.890970459 | 0.875405581 |
| CENPQ | 0.890959545 | 0.835965978 |

| | | |
|---|---|---|
| UPP2 | 0.890914259 | 0.861482382 |
| NIPA2 | 0.890825725 | 0.986634265 |
| ALKBH8 | 0.890769691 | 0.90562981 |
| IL10RB | 0.890768723 | 0.979748886 |
| LCE5A | 0.89071552 | 0.942788626 |
| HTR3B | 0.89069303 | 1.013627515 |
| PLD1 | 0.890687473 | 0.808424463 |
| REG3G | 0.890628383 | 0.875658161 |
| FUBP3 | 0.890624085 | 0.85748544 |
| GID4 | 0.890578912 | 0.974306269 |
| RBP2 | 0.890578912 | 0.974306269 |
| TRMT1 | 0.890525535 | 0.778452815 |
| MORN5 | 0.890518831 | 0.871531346 |
| AKT1S1 | 0.890477599 | 0.948269955 |
| BIK | 0.890477599 | 0.770469339 |
| OR2Y1 | 0.890476275 | 1.003240178 |
| RELB | 0.890471029 | 0.945321993 |
| FXR2 | 0.890447287 | 0.887290168 |
| CPSF4 | 0.89044262 | 1.025008565 |
| TCEAL4 | 0.890426507 | 0.954637505 |
| OR8H1 | 0.890349464 | 0.979787598 |
| PASD1 | 0.8903313 | 0.87363305 |
| SLC43A2 | 0.890301856 | 0.940866748 |
| SLC38A11 | 0.890295998 | 0.757796679 |
| ZNF600 | 0.890293778 | 0.878725591 |
| ATP13A4 | 0.890293351 | 0.924075276 |
| LOC649055 | 0.890293351 | 0.924075276 |
| RBCK1 | 0.890277874 | 0.972096042 |
| SLC5A5 | 0.890277874 | 0.972096042 |
| UNC5C | 0.890216735 | 0.959497772 |
| TBC1D31 | 0.890188524 | 0.931140802 |
| MPLKIP | 0.890181456 | 0.969510106 |
| DECR1 | 0.890146733 | 0.870214147 |
| C10orf2 | 0.890116767 | 1.06229721 |
| SERPINA11 | 0.890112563 | 0.961927906 |
| ADSL | 0.890077312 | 0.862292426 |
| CCDC71 | 0.89006585 | 0.929526124 |
| RSPH9 | 0.890036601 | 1.037684138 |
| KIR2DL5B | 0.890009914 | 0.364508393 |
| TMEM259 | 0.890009914 | 0.940047962 |
| ZNF264 | 0.890009914 | 0.940047962 |
| SERP1 | 0.88998155 | 0.976704351 |
| CNGB1 | 0.889980852 | 0.792342635 |
| SPACA5 | 0.889969828 | 0.973673552 |

| | | |
|---|---|---|
| SEMA6C | 0.889924505 | 0.97327852 |
| NADSYN1 | 0.889918735 | 0.806804374 |
| KLHL2 | 0.889883273 | 0.875642343 |
| SPATA8 | 0.889867846 | 0.976103686 |
| APBA1 | 0.889847751 | 1.025693731 |
| GRINA | 0.889836786 | 1.002595717 |
| PPP3CA | 0.889833246 | 0.978585334 |
| MAOB | 0.889829542 | 0.954574951 |
| FPR2 | 0.889791508 | 0.762491888 |
| TLX2 | 0.889764437 | 0.885783718 |
| BOLA1 | 0.889697094 | 0.616848927 |
| MTMR4 | 0.889683994 | 0.972744971 |
| HSD17B11 | 0.889678196 | 0.929931146 |
| MX1 | 0.889573352 | 1.062946139 |
| ZNF384 | 0.889568369 | 0.957877683 |
| CTBP2 | 0.889554088 | 1.014925373 |
| OR52H1 | 0.889533982 | 0.931825968 |
| CTSH | 0.889483455 | 0.870863076 |
| NPSR1 | 0.889483455 | 0.870863076 |
| TNFRSF1A | 0.889467711 | 0.90927501 |
| ARHGAP9 | 0.889417038 | 0.888317917 |
| EEF1A2 | 0.889365423 | 0.988254354 |
| TMPRSS11F | 0.889364498 | 0.937053861 |
| ISCU | 0.889361806 | 0.937221547 |
| SNED1 | 0.889304804 | 1.032401782 |
| TNFSF18 | 0.889298462 | 0.973963686 |
| SLC5A9 | 0.889269872 | 0.898937992 |
| BCAT2 | 0.889263084 | 0.777237748 |
| KIAA1804 | 0.889243565 | 0.979234263 |
| POLI | 0.889225042 | 0.955223881 |
| DHRS11 | 0.889224624 | 0.948967193 |
| CD37 | 0.889153614 | 0.942081814 |
| SDK1 | 0.889069105 | 0.942788626 |
| AIM1L | 0.889049577 | 0.923603974 |
| ABRA | 0.889037215 | 0.7472661 |
| RGS9 | 0.889030595 | 1.063595068 |
| FCRL3 | 0.889030389 | 0.979343864 |
| C19orf68 | 0.889002727 | 1.1558753 |
| MAN1A2 | 0.888961525 | 0.99156392 |
| GRN | 0.88894174 | 1.11579308 |
| DCDC2 | 0.888912153 | 0.934224049 |
| SLC7A1 | 0.88888903 | 0.965573107 |
| C7ORF28A | 0.888866526 | 0.824254882 |
| PTH2R | 0.888836888 | 1.009733939 |

| | | |
|---|---|---|
| ADM2 | 0.888824116 | 0.911613566 |
| FBXW2 | 0.888799431 | 1.099935107 |
| ZBTB20 | 0.888761476 | 0.977318753 |
| DDX3Y | 0.888734631 | 0.986634265 |
| KIFAP3 | 0.888716696 | 0.913692408 |
| GXYLT2 | 0.888696578 | 0.935601458 |
| REM2 | 0.888682366 | 1.00526529 |
| OR8K3 | 0.888638574 | 0.990407674 |
| SLC16A5 | 0.888621366 | 0.95587281 |
| FIS1 | 0.888597754 | 0.903199676 |
| OR4N2 | 0.888592584 | 0.899623159 |
| NCAPG | 0.888590062 | 0.972863507 |
| ESYT1 | 0.888564694 | 0.98217902 |
| MIXL1 | 0.888552955 | 1.112264763 |
| TPP1 | 0.888510641 | 1.178455548 |
| MKKS | 0.888495712 | 0.842069202 |
| MEGF8 | 0.888466176 | 0.949777238 |
| CCDC51 | 0.88844636 | 1.063588497 |
| NREP | 0.888399852 | 1.047387606 |
| GIPC3 | 0.88839286 | 0.994176088 |
| DDX25 | 0.888389715 | 0.942891859 |
| ABL1 | 0.888380159 | 0.992212849 |
| RXFP3 | 0.888380159 | 0.902011681 |
| NPHS1 | 0.888359874 | 0.957519699 |
| C1orf198 | 0.888330064 | 0.743215877 |
| TAF5 | 0.888323442 | 0.857027136 |
| EFHB | 0.888305103 | 0.901174565 |
| ZBED1 | 0.888290958 | 0.919150394 |
| HOXC6 | 0.888159862 | 0.872160934 |
| SLAMF6 | 0.88806659 | 0.980532122 |
| VAT1L | 0.888034813 | 0.890330954 |
| HSPB1 | 0.888000252 | 1.006851662 |
| NME1 | 0.887980891 | 0.866320571 |
| CEP128 | 0.887977008 | 0.886262419 |
| COL2A1 | 0.887968716 | 0.936964714 |
| PSMA7 | 0.887968049 | 0.932511356 |
| SPTSSA | 0.887894137 | 1.034943474 |
| WNT8A | 0.887891214 | 0.999189955 |
| CLDN16 | 0.887879039 | 0.822600243 |
| OR2A5 | 0.887833286 | 1.031517643 |
| CCDC7 | 0.887823182 | 0.811236725 |
| GPX1 | 0.887771861 | 0.966788173 |
| NXPE2 | 0.887754083 | 0.976103686 |
| C20orf27 | 0.887729572 | 1.00890716 |

| | | |
|---|---|---|
| CCDC38 | 0.887613692 | 0.792628595 |
| H2AFB1 | 0.887576109 | 0.933881466 |
| DBX2 | 0.887481655 | 0.974078574 |
| FHL2 | 0.88742394 | 1.011340624 |
| RAVER2 | 0.88742394 | 1.011340624 |
| PTPRK | 0.887416483 | 0.957170668 |
| TRIP6 | 0.887387999 | 0.890979883 |
| ISLR | 0.88737951 | 0.874614594 |
| OIP5 | 0.887368953 | 0.923603974 |
| ZSCAN32 | 0.887357128 | 1.170263789 |
| DDHD2 | 0.887312951 | 0.98663926 |
| SCGB1D1 | 0.887285312 | 0.935936965 |
| ABCG8 | 0.887284766 | 1.01752109 |
| BICC1 | 0.887238009 | 0.949982871 |
| SEC31A | 0.887233322 | 0.953422438 |
| NANOS1 | 0.887217801 | 0.883521754 |
| ACVR2A | 0.887192646 | 0.776768332 |
| DNASE2B | 0.887172907 | 0.927906035 |
| MSRB3 | 0.887167867 | 1.077871512 |
| CREB3L4 | 0.887154276 | 0.815347722 |
| GPR119 | 0.887151238 | 0.969500324 |
| IFIT5 | 0.887079432 | 1.002430134 |
| RREB1 | 0.887068734 | 1.011745646 |
| RRP12 | 0.887068734 | 1.011745646 |
| UBASH3B | 0.886984286 | 0.770927969 |
| NAA38 | 0.886958347 | 0.860672337 |
| FERMT2 | 0.88689796 | 0.997601918 |
| PSMC2 | 0.886861662 | 0.853786958 |
| TADA2A | 0.886854462 | 0.831278391 |
| INSL4 | 0.886842637 | 0.962658445 |
| AHCTF1 | 0.886827131 | 0.915639195 |
| TMED3 | 0.886824513 | 0.913669065 |
| GATM | 0.886776749 | 1.126541207 |
| GALC | 0.886713783 | 1.012150668 |
| RHOT1 | 0.886694935 | 0.921425678 |
| LOC100652824 | 0.88666892 | 0.381637547 |
| OR11H6 | 0.886596878 | 0.966426859 |
| CMIP | 0.886569647 | 0.870161014 |
| C5orf34 | 0.886562134 | 0.833130822 |
| DSP | 0.886557821 | 1.027749229 |
| F8A3 | 0.886526014 | 0.994518671 |
| KXD1 | 0.88647467 | 0.963547995 |
| C1orf123 | 0.886469321 | 0.972863507 |
| CDC42BPB | 0.886457582 | 0.819597664 |

| | | |
|---|---|---|
| RSRC2 | 0.886441151 | 0.854191981 |
| CDKL3 | 0.886394523 | 1.006489293 |
| HTRA2 | 0.88637954 | 0.922128488 |
| DNAJA2 | 0.886361676 | 0.837767683 |
| HTR1B | 0.886283304 | 0.898118105 |
| SKAP2 | 0.886279671 | 0.987039287 |
| DYNAP | 0.886247269 | 0.965056526 |
| DPY19L2 | 0.886243656 | 0.821790198 |
| SMS | 0.886210239 | 0.914945322 |
| RHOXF2 | 0.886199603 | 0.807468311 |
| HSPA8 | 0.886170521 | 1.060999351 |
| FXYD4 | 0.886154468 | 1.018818949 |
| C7orf26 | 0.886141834 | 0.867077766 |
| PODNL1 | 0.886107799 | 0.93600648 |
| ADCYAP1 | 0.886094938 | 0.849606029 |
| SENP3 | 0.886016165 | 0.910447761 |
| AP4M1 | 0.886003066 | 1.031591738 |
| FBXL6 | 0.885989904 | 1.012978585 |
| CLUH | 0.885938287 | 0.928617781 |
| GJC3 | 0.88589322 | 1.070572114 |
| PHYHIP | 0.885880669 | 0.993490922 |
| TNFAIP8L1 | 0.885826354 | 0.999189955 |
| VEZT | 0.885818153 | 0.915350344 |
| RALGAPA1 | 0.885786425 | 1.004110997 |
| CYR61 | 0.885764722 | 0.881466256 |
| ISCA1 | 0.885733643 | 0.982989064 |
| IPPK | 0.88572507 | 1.085658663 |
| TFCP2 | 0.885643323 | 0.898767034 |
| CTGLF1 | 0.885641117 | 0.951695786 |
| PIGC | 0.885620658 | 0.910895099 |
| GSX2 | 0.885559631 | 1.006480356 |
| KIZ | 0.885549443 | 0.978533819 |
| NAIF1 | 0.885523765 | 0.871188763 |
| CA2 | 0.885484522 | 0.995457495 |
| OR6C2 | 0.885471607 | 1.00891049 |
| GRB14 | 0.885451587 | 0.972935937 |
| CCDC87 | 0.885450008 | 0.962332928 |
| MBOAT2 | 0.885398253 | 1.039287161 |
| CTAGE5 | 0.885329076 | 1.069663832 |
| TEX15 | 0.885283851 | 1.173004454 |
| CPA1 | 0.885260767 | 0.887086308 |
| MAST1 | 0.885253029 | 1.007787151 |
| CLDN12 | 0.885230204 | 0.962658445 |
| ANKRD36B | 0.885213757 | 0.976704351 |

| | | |
|---|---|---|
| PRIMPOL | 0.885176052 | 0.969623329 |
| ZADH2 | 0.885174014 | 0.846091535 |
| PYROXD1 | 0.885148783 | 0.929427886 |
| DNAJC27 | 0.885081302 | 1.001946788 |
| CPSF1 | 0.88505984 | 1.248715313 |
| EFCAB13 | 0.885045543 | 1.030376671 |
| ABCC3 | 0.885021245 | 1.002430134 |
| ADAM33 | 0.884974694 | 0.77871512 |
| ZSCAN5A | 0.884960728 | 1.032806804 |
| E2F7 | 0.884885897 | 0.903391572 |
| HIST1H1A | 0.88487632 | 0.862697448 |
| WEE1 | 0.884867921 | 0.875405581 |
| HMCN1 | 0.884844257 | 1.003425831 |
| EPN2 | 0.884783135 | 0.928057554 |
| PACRG | 0.884772849 | 0.94210346 |
| FIGNL2 | 0.884728339 | 0.981773998 |
| SCYL1 | 0.884683352 | 1.00843608 |
| SLC7A4 | 0.884617442 | 0.897934386 |
| NAV2 | 0.884565286 | 0.875642343 |
| ELMOD1 | 0.884498853 | 0.949372215 |
| KCTD17 | 0.884488532 | 1.044193217 |
| ADIPOR2 | 0.884480331 | 0.872216513 |
| PRDM13 | 0.884473489 | 0.986296677 |
| GRIN2C | 0.884374146 | 0.978585334 |
| UPP1 | 0.884253118 | 0.916970433 |
| PTPN7 | 0.884250743 | 1.033095393 |
| PTMS | 0.884244811 | 0.976019185 |
| ELAC2 | 0.88424456 | 0.965978129 |
| STAT4 | 0.884212463 | 1.051265412 |
| DLK1 | 0.884202991 | 1.002398082 |
| CCNB1 | 0.884084862 | 0.930564568 |
| UHMK1 | 0.884045594 | 0.948734588 |
| ZNF414 | 0.883961944 | 0.963953017 |
| GBX2 | 0.883912634 | 0.985014176 |
| GSTA2 | 0.883893531 | 1.064398542 |
| OR1J4 | 0.883886684 | 0.932511134 |
| SLC2A4RG | 0.883877124 | 0.890373416 |
| LCE3E | 0.883806831 | 0.978128797 |
| CCNI | 0.88369572 | 0.918883842 |
| TMEM150A | 0.883678003 | 0.961927906 |
| KCNJ5 | 0.883666989 | 1.051914341 |
| XRN2 | 0.88365615 | 0.793843661 |
| TUSC3 | 0.883606891 | 0.854597003 |
| CACNA1G | 0.88358656 | 1.1064244 |

| | | |
|---|---|---|
| CYP2B6 | 0.88356408 | 0.864337102 |
| MMP17 | 0.883537166 | 0.761842959 |
| KIF1B | 0.88352704 | 0.894873459 |
| USP38 | 0.883519548 | 1.027903958 |
| IFIT1 | 0.883483882 | 0.874957177 |
| AHNAK | 0.883472612 | 0.917780478 |
| MNX1 | 0.88346436 | 0.878088295 |
| CCDC88B | 0.883449384 | 1.018226002 |
| UVSSA | 0.883446456 | 0.941760877 |
| ST6GALNAC4 | 0.883431911 | 0.938841636 |
| LRIG2 | 0.883383884 | 0.911956149 |
| SCG2 | 0.883352348 | 0.92428914 |
| COL5A2 | 0.883228048 | 0.934386391 |
| EMR1 | 0.883224914 | 0.961713173 |
| PHF20 | 0.88318581 | 0.828682674 |
| HS6ST1 | 0.883111395 | 0.927501013 |
| CCDC122 | 0.883091376 | 0.857432159 |
| B3GALT2 | 0.88308587 | 1.08890331 |
| GTF2E1 | 0.883072039 | 0.919532771 |
| OPN1LW | 0.883072039 | 0.919532771 |
| HBEGF | 0.883032607 | 0.95587281 |
| MKLN1 | 0.883012798 | 0.843782117 |
| CNTFR | 0.883004864 | 0.786502271 |
| PGAP2 | 0.882948005 | 0.961630695 |
| EIF1AY | 0.88294592 | 1.046722907 |
| PNRC1 | 0.882876507 | 0.998629668 |
| DRD1 | 0.882837912 | 0.950032446 |
| RABGAP1 | 0.882800115 | 0.96882494 |
| CENPW | 0.882784755 | 0.928400137 |
| NDC80 | 0.882728478 | 0.703523694 |
| GALNT13 | 0.882717883 | 1.009720535 |
| ASPHD2 | 0.882674431 | 0.857837181 |
| DUOXA2 | 0.882666841 | 1.021466181 |
| EHD3 | 0.882632877 | 0.935594382 |
| ZNF321P | 0.882583502 | 0.942081814 |
| SSR4 | 0.882569308 | 0.946214457 |
| SPSB3 | 0.882551263 | 0.738481506 |
| LPCAT2 | 0.882543871 | 0.984204131 |
| FCAMR | 0.882532868 | 1.087079789 |
| LMF2 | 0.882511626 | 0.974888619 |
| FUT9 | 0.882465066 | 0.883758607 |
| AGAP1 | 0.882446156 | 0.967454608 |
| SLC37A2 | 0.882441279 | 0.93835172 |
| TRAF3 | 0.882441279 | 0.93835172 |

| | | |
|---|---|---|
| CACYBP | 0.882407759 | 0.792628595 |
| AGO3 | 0.882386699 | 1.083711875 |
| NPPC | 0.882352909 | 1.002740665 |
| STIL | 0.882335628 | 1.052247874 |
| SFRP2 | 0.88226752 | 0.909680032 |
| ACER2 | 0.882266051 | 0.830421377 |
| LPXN | 0.882188217 | 1.241401687 |
| ZNF280D | 0.882133747 | 1.006509078 |
| WDR89 | 0.882099061 | 0.987039287 |
| CHCHD7 | 0.882089841 | 1.008100446 |
| SKIL | 0.882071656 | 0.956662616 |
| HACE1 | 0.882034602 | 0.963011032 |
| FYB | 0.881986989 | 0.936279548 |
| KRTAP4-2 | 0.881986445 | 1.026036314 |
| SIGLEC8 | 0.881957266 | 1.011990408 |
| UGT2A3 | 0.881846449 | 0.837586067 |
| CCNB1IP1 | 0.881786675 | 1.068516615 |
| CIDEB | 0.881773626 | 1.029804728 |
| IFNL3 | 0.881700224 | 0.950668037 |
| TSFM | 0.881699334 | 0.93600648 |
| ENTPD1 | 0.881648259 | 0.987670344 |
| HIST1H2AD | 0.881639917 | 0.834532374 |
| PDCD11 | 0.881610013 | 1.022953066 |
| ALS2CL | 0.881578555 | 1.013365735 |
| PLAGL1 | 0.88157341 | 0.95018226 |
| TMEM64 | 0.88157341 | 0.95018226 |
| PRRG1 | 0.88156715 | 0.945529291 |
| FAM19A3 | 0.881478508 | 0.889428919 |
| MYO5C | 0.881436788 | 0.901174565 |
| OR5B2 | 0.881432581 | 0.940390545 |
| SF3A2 | 0.881384734 | 1.063720452 |
| ARL15 | 0.881269093 | 1.02308627 |
| TUBB | 0.881260943 | 1.002025111 |
| TNFRSF21 | 0.88123929 | 0.982870846 |
| HEPACAM | 0.881226354 | 1.013770757 |
| LYSMD2 | 0.881226354 | 1.013770757 |
| ACOT8 | 0.881214434 | 0.89671932 |
| XKR8 | 0.881192632 | 0.854597003 |
| CNTN6 | 0.881176535 | 0.908464966 |
| RGS18 | 0.881149333 | 1.016200891 |
| LMNA | 0.881127377 | 0.749291211 |
| ENG | 0.881115323 | 0.903734156 |
| SMN1 | 0.881114054 | 0.916067146 |
| ARV1 | 0.88109065 | 0.910895099 |

| | | |
|---|---|---|
| SIDT2 | 0.881077201 | 0.889833941 |
| DMP1 | 0.881074454 | 0.791024323 |
| MTUS2 | 0.881067703 | 0.90202124 |
| ZFP64 | 0.881059184 | 1.099349092 |
| MED13 | 0.881034004 | 0.95544755 |
| MYOF | 0.881014545 | 0.988254354 |
| THAP8 | 0.881009706 | 1.042122317 |
| C1S | 0.880992719 | 0.915639195 |
| TSPAN8 | 0.880974999 | 0.960260363 |
| RCAN3 | 0.880968429 | 0.836930456 |
| CHURC1 | 0.880882975 | 0.944501542 |
| MFSD3 | 0.8808306 | 0.864317537 |
| PIP4K2B | 0.880824698 | 0.946138871 |
| USP10 | 0.880824698 | 1.103828683 |
| LOXL2 | 0.880763104 | 0.887808829 |
| COPZ1 | 0.880758238 | 0.988694758 |
| SYNGR3 | 0.880693762 | 0.925316889 |
| HIGD2A | 0.88067457 | 1.009720535 |
| FAM53C | 0.880668254 | 0.923045768 |
| AMN1 | 0.880634939 | 0.815309842 |
| HIST1H1C | 0.880625436 | 0.983556012 |
| RDM1 | 0.880614715 | 0.913730255 |
| ABHD12B | 0.880601522 | 0.827055488 |
| CACNB4 | 0.880586207 | 0.922128488 |
| BDKRB2 | 0.880545869 | 1.025308241 |
| ZSCAN10 | 0.880533975 | 0.883353584 |
| TM4SF1 | 0.880389826 | 1.094895512 |
| FEM1A | 0.880327434 | 0.996231586 |
| HBS1L | 0.880299969 | 0.879948086 |
| ZCWPW2 | 0.880262209 | 0.825030377 |
| RASL10B | 0.880229611 | 0.872901679 |
| RNASEH2A | 0.880216261 | 0.771162414 |
| VSTM2A | 0.880180145 | 1.047792629 |
| PNMA3 | 0.880079681 | 0.963001028 |
| TGFBI | 0.880064956 | 0.903048989 |
| WDR7 | 0.880020281 | 0.948955122 |
| CCL19 | 0.879979774 | 0.994861254 |
| AVIL | 0.879877782 | 0.943816375 |
| FOXP1 | 0.879833032 | 0.953277093 |
| PLEKHG6 | 0.879820354 | 1.015390846 |
| CNR1 | 0.879790224 | 1.020116807 |
| MS4A8 | 0.879790224 | 1.020116807 |
| NUBP2 | 0.879777004 | 0.949777238 |
| ACRV1 | 0.879770715 | 1.104487838 |

| | | |
|---|---|---|
| PCDH9 | 0.879766637 | 0.963343611 |
| EXTL3 | 0.879723466 | 0.985014176 |
| UBR4 | 0.879673545 | 1.020251114 |
| AK4 | 0.879663529 | 0.965606749 |
| RAET1E | 0.879649286 | 0.912298732 |
| SLN | 0.87959138 | 0.87529976 |
| CSNK1A1 | 0.879563917 | 1.081116158 |
| NHP2 | 0.879557452 | 1.001620089 |
| SYNGR2 | 0.879539183 | 0.990065091 |
| IZUMO4 | 0.879484557 | 0.767112191 |
| IDE | 0.879450311 | 0.959766385 |
| KIAA1033 | 0.879420699 | 1.078451524 |
| GPR137 | 0.879267603 | 0.863507493 |
| SPPL2B | 0.87923092 | 1.020765737 |
| NLRP12 | 0.879227792 | 1.087605451 |
| LSS | 0.87911468 | 0.936411503 |
| PCDHA1 | 0.879094151 | 0.959902795 |
| TMEM220 | 0.879037515 | 0.891743748 |
| TAS1R1 | 0.879013048 | 0.902363823 |
| UQCRQ | 0.878985649 | 0.906034832 |
| HJURP | 0.878920753 | 0.944844125 |
| EIF3K | 0.878915128 | 0.978585334 |
| OCM2 | 0.87889701 | 0.85303186 |
| NMS | 0.878855344 | 0.863912515 |
| ID1 | 0.878847652 | 0.875658161 |
| DCK | 0.878820292 | 1.00908501 |
| RNASET2 | 0.878767727 | 0.72782503 |
| CEACAM3 | 0.878760538 | 0.97327852 |
| SNAP47 | 0.878746332 | 0.913325233 |
| LAMB3 | 0.87871806 | 0.972053463 |
| TMCO6 | 0.87871806 | 0.972053463 |
| FOXJ3 | 0.878677664 | 0.892264075 |
| INO80 | 0.878667381 | 0.87975334 |
| TSKU | 0.878643942 | 0.974483597 |
| CTXN1 | 0.878639878 | 0.923946557 |
| USP11 | 0.878623704 | 0.839065542 |
| CPNE3 | 0.878588387 | 1.051914341 |
| PRPS2 | 0.878548296 | 0.985074627 |
| TTLL5 | 0.878516846 | 0.897124342 |
| MGP | 0.878467204 | 0.996574169 |
| RAB10 | 0.878430985 | 0.900064893 |
| TAF4B | 0.878429178 | 0.94653706 |
| PEX3 | 0.87840966 | 0.759849263 |
| ADAMTS10 | 0.878400032 | 0.997404283 |

| | | |
|---|---|---|
| MYL2 | 0.878368791 | 0.959575197 |
| CCL13 | 0.878364484 | 0.831510733 |
| CA10 | 0.878344052 | 0.930564568 |
| SDHB | 0.878344052 | 0.930564568 |
| SELV | 0.878283836 | 0.945529291 |
| C19orf44 | 0.878278905 | 0.892669097 |
| ZNF697 | 0.878278905 | 1.104090725 |
| MMP1 | 0.878114452 | 0.87605451 |
| MLLT11 | 0.878109274 | 0.838801134 |
| C11orf49 | 0.878104384 | 0.815309842 |
| FAM126B | 0.878061645 | 0.98217902 |
| HNRNPA2B1 | 0.878047455 | 1.058924289 |
| DCLRE1B | 0.877969893 | 0.985723556 |
| PIM1 | 0.877969893 | 0.985723556 |
| ALKBH1 | 0.877959408 | 0.890643985 |
| C14orf181_pseudogene | 0.877943696 | 0.967112025 |
| ASPSCR1 | 0.877919909 | 1.043165468 |
| SART3 | 0.877894762 | 0.910585817 |
| C6 | 0.877854165 | 1.007879411 |
| NKAP | 0.877801017 | 1.071599863 |
| TBX5 | 0.877799507 | 1.038882139 |
| MRPS7 | 0.877778271 | 0.917780062 |
| RUFY1 | 0.877759722 | 0.803374432 |
| CPPED1 | 0.877724319 | 0.897934386 |
| KIAA2026 | 0.877716933 | 0.88618874 |
| AVPI1 | 0.877685553 | 0.95909275 |
| ATPAF2 | 0.877644427 | 0.804727646 |
| CMKLR1 | 0.877611951 | 0.992212849 |
| AP3D1 | 0.877611804 | 1.075368277 |
| POLR2F | 0.877596357 | 0.938031592 |
| UNC50 | 0.877540883 | 0.867557716 |
| SCML2 | 0.877498592 | 0.925373134 |
| HMP19 | 0.877487384 | 0.854402193 |
| EIF3J | 0.877476283 | 1.004542505 |
| CDH15 | 0.87744511 | 0.92874272 |
| SLC2A10 | 0.877429025 | 0.960602946 |
| FAHD2B | 0.877367781 | 0.85853342 |
| GRIK1 | 0.877334522 | 1.096041531 |
| NPM3 | 0.877314256 | 0.989037342 |
| CXorf22 | 0.877309362 | 0.859883522 |
| PXT1 | 0.877255975 | 0.973673552 |
| LYG2 | 0.877214742 | 0.950032446 |
| KLF11 | 0.877173333 | 1.059701493 |
| NR5A1 | 0.877166171 | 0.901362751 |

| | | |
|---|---|---|
| FURIN | 0.877078168 | 0.962362103 |
| SAA4 | 0.877071759 | 0.895512162 |
| ALG5 | 0.877065984 | 0.883192732 |
| RAP1GAP2 | 0.877051188 | 1.097293594 |
| C6orf223 | 0.877013445 | 0.936279548 |
| PGBD3 | 0.876856095 | 1 |
| UBAP2 | 0.876851146 | 0.920520726 |
| ZNF117 | 0.876820895 | 0.92709599 |
| DEFB128 | 0.876819158 | 1.067488866 |
| FKBP1B | 0.876815392 | 0.877352369 |
| CXCL13 | 0.876692653 | 0.936622131 |
| ZNF439 | 0.876676568 | 0.883521754 |
| TSHZ1 | 0.876654473 | 1.007290401 |
| ANKRD13D | 0.876609391 | 0.80963953 |
| QRICH1 | 0.876567977 | 0.997974889 |
| KLK10 | 0.876565458 | 1.011680727 |
| WBP5 | 0.87651581 | 1.014731072 |
| RHEB | 0.876465436 | 0.993510707 |
| CTNNA1 | 0.87643244 | 0.949297705 |
| C1orf116 | 0.876412558 | 0.99108951 |
| ANXA11 | 0.876411009 | 1.094552929 |
| MYO7A | 0.876366305 | 0.929931146 |
| MAP2K1 | 0.87632738 | 1.036340039 |
| GNB1L | 0.876292932 | 0.976019185 |
| TRAM1L1 | 0.876285682 | 0.965399109 |
| ZNF131 | 0.876277663 | 1.027406646 |
| PAN2 | 0.87625403 | 0.957170668 |
| COQ10B | 0.87625281 | 0.807614419 |
| ACO2 | 0.876222857 | 0.795868773 |
| ARPC5 | 0.876220029 | 1.015073655 |
| STX18 | 0.876120008 | 1 |
| BAALC | 0.87610671 | 0.939020212 |
| GTPBP3 | 0.876016317 | 1.008221994 |
| SPIRE2 | 0.876006303 | 0.997601918 |
| BTN3A3 | 0.875974813 | 0.965741692 |
| IL13RA1 | 0.875924572 | 0.780661908 |
| FAM117B | 0.875921076 | 0.964028777 |
| TLL2 | 0.875896912 | 0.963659961 |
| CCDC130 | 0.875894497 | 0.805589307 |
| DPY19L3 | 0.875894497 | 0.984609154 |
| PDE2A | 0.875894497 | 0.984609154 |
| SCAF11 | 0.875892667 | 0.891401165 |
| CLDN17 | 0.875834308 | 0.889688249 |
| SLC25A16 | 0.875780391 | 0.654921021 |

| | | |
|---|---|---|
| OOSP2 | 0.875762358 | 0.989469421 |
| CSPG4 | 0.875731637 | 0.93764988 |
| LCA5 | 0.875682385 | 1.058239123 |
| CEP68 | 0.875681414 | 0.876468206 |
| BRD7 | 0.875669443 | 0.954232483 |
| ADPGK | 0.875667477 | 1.018818949 |
| BTF3 | 0.875664019 | 0.988319273 |
| NPLOC4 | 0.875659255 | 1.03199676 |
| CSTL1 | 0.875630687 | 0.904076739 |
| KLHL25 | 0.875622425 | 1.098015391 |
| NUDT18 | 0.875599859 | 0.982584042 |
| AUP1 | 0.875594938 | 0.953751285 |
| SLC29A1 | 0.875498233 | 0.895504253 |
| GSKIP | 0.875473599 | 1.010620075 |
| SDAD1 | 0.875473599 | 1.010620075 |
| THBS2 | 0.875457786 | 1 |
| DOK5 | 0.875363315 | 1.097636177 |
| MSMO1 | 0.875292083 | 0.933809215 |
| MLLT10 | 0.875239314 | 0.842310188 |
| NEURL1 | 0.875158308 | 0.915639195 |
| APLF | 0.87515548 | 0.860226105 |
| DSCR10 | 0.875133595 | 0.959575197 |
| LYZL2 | 0.875113115 | 0.948955122 |
| PANK4 | 0.875067505 | 0.958468527 |
| ZNF280C | 0.875040439 | 0.886947585 |
| REG1B | 0.875030806 | 0.832320778 |
| PLEKHM3 | 0.875026619 | 1.035971223 |
| SYNJ2BP | 0.874973562 | 1.083932854 |
| OSCP1 | 0.874934427 | 1.021061158 |
| UFM1 | 0.874899843 | 0.972250771 |
| RAC1 | 0.874759205 | 0.891628812 |
| C10orf71 | 0.874708863 | 0.956491949 |
| DCUN1D3 | 0.87468429 | 0.830629461 |
| IL17RA | 0.87468429 | 0.934458144 |
| FAM120AOS | 0.874680134 | 0.830295666 |
| BMP2K | 0.874642553 | 0.800129786 |
| CD27 | 0.874538496 | 0.916288125 |
| TMEM128 | 0.874499755 | 0.903604698 |
| ING4 | 0.874483539 | 0.930113052 |
| UPB1 | 0.874450081 | 0.855288774 |
| CLEC2L | 0.874424455 | 0.947927372 |
| CMTM1 | 0.874421238 | 1.065090785 |
| TACC3 | 0.874378772 | 0.948562171 |
| COL11A1 | 0.874370083 | 1.063377869 |

| | | |
|---|---|---|
| ST3GAL5 | 0.874346323 | 0.922640745 |
| INTU | 0.874309799 | 0.842446335 |
| DIO3 | 0.874275567 | 0.870797894 |
| VMP1 | 0.874273982 | 0.894826996 |
| MAN2C1 | 0.874264411 | 0.939246659 |
| TMSB10 | 0.874207205 | 0.995949777 |
| CHMP2B | 0.8741717 | 0.970028352 |
| MAU2 | 0.87412523 | 1.006851662 |
| OPALIN | 0.87410253 | 0.92874272 |
| BRAT1 | 0.874090342 | 0.802672148 |
| TNFSF10 | 0.874065795 | 0.878088295 |
| GPR158 | 0.874064397 | 0.861778066 |
| IL17RE | 0.874057849 | 0.960712839 |
| P4HB | 0.873984953 | 0.894694208 |
| EXOSC2 | 0.873945668 | 0.847207948 |
| MYBPC3 | 0.873897132 | 0.925475901 |
| CHRNG | 0.873884109 | 0.959766385 |
| FLG | 0.873875076 | 0.941418294 |
| HOXA4 | 0.873772686 | 0.805184285 |
| RSG1 | 0.87376968 | 1.041312272 |
| PPIE | 0.873759441 | 0.944511948 |
| C1QL1 | 0.873738785 | 0.870503597 |
| GATA6 | 0.873737326 | 1.057754705 |
| ARHGEF9 | 0.87372927 | 0.898937992 |
| CDK5RAP1 | 0.873683286 | 0.975293641 |
| SNRPD3 | 0.873683286 | 0.975293641 |
| LDLRAD1 | 0.873627251 | 0.64236533 |
| PDE7B | 0.873621903 | 0.977723775 |
| ZNRF2 | 0.873573832 | 1.082414017 |
| TM9SF1 | 0.873539377 | 0.922233642 |
| PEX19 | 0.873536387 | 0.950668037 |
| CCAR2 | 0.873518308 | 0.931140802 |
| MED31 | 0.873497647 | 0.940047962 |
| WNT10A | 0.87343802 | 0.909900651 |
| HSD17B14 | 0.873417583 | 1.107073329 |
| CD93 | 0.873381312 | 0.97326853 |
| STK3 | 0.873370165 | 0.99091499 |
| ZNF212 | 0.873336979 | 0.926002055 |
| MRPL3 | 0.873328008 | 1.020212402 |
| MUT | 0.873327468 | 0.876468206 |
| PRR3 | 0.873317995 | 0.934909215 |
| HINFP | 0.873310201 | 0.878040425 |
| WDR17 | 0.873260176 | 1.077359255 |
| CST5 | 0.873210928 | 0.988352175 |

| | | |
|---|---|---|
| OR52E2 | 0.873162854 | 0.883521754 |
| BMPR2 | 0.873125171 | 1.027903958 |
| MLKL | 0.873040378 | 0.954574951 |
| LSAMP | 0.873031411 | 1.05789654 |
| CNTROB | 0.873010722 | 0.945321993 |
| PCDHA8 | 0.872951395 | 0.999314834 |
| PRMT8 | 0.872949905 | 0.924075276 |
| SLC25A24 | 0.872937122 | 0.821514217 |
| WNT5B | 0.872937122 | 0.933538883 |
| PSMB4 | 0.872887504 | 1.022063595 |
| OVCH2 | 0.872845574 | 0.995139733 |
| FEV | 0.872828923 | 0.921205892 |
| LPCAT1 | 0.872783859 | 0.948734588 |
| ERP27 | 0.872766362 | 0.955042527 |
| SOBP | 0.872729634 | 1 |
| CPZ | 0.872647202 | 1.016223232 |
| MAP1LC3B | 0.872629289 | 0.878725591 |
| EGFLAM | 0.872597139 | 0.889023896 |
| H3F3C | 0.87258634 | 0.874848117 |
| ZNF143 | 0.872578051 | 0.887735237 |
| ARHGAP27 | 0.87253759 | 0.978938842 |
| LLPH | 0.87251375 | 0.789793439 |
| ATP11AUN | 0.872495535 | 1.031860226 |
| MYCN | 0.87249317 | 0.922235723 |
| CGB | 0.872420774 | 0.827338129 |
| CD6 | 0.872350538 | 0.972053463 |
| DSTN | 0.87230622 | 0.988659376 |
| G0S2 | 0.872279018 | 0.901174565 |
| KCTD8 | 0.872270301 | 0.832819459 |
| LRRC14 | 0.872260469 | 0.841726619 |
| CACNG7 | 0.872219945 | 0.89422453 |
| DERL3 | 0.872209722 | 0.959232614 |
| ANGEL1 | 0.872203728 | 0.948562171 |
| LMO1 | 0.872132188 | 1.059701493 |
| PLS1 | 0.872119828 | 0.993148338 |
| SLCO5A1 | 0.872087635 | 0.804374241 |
| ARNTL2 | 0.872073729 | 0.745240988 |
| MKS1 | 0.87204943 | 0.925070879 |
| SLC10A3 | 0.872048193 | 1.009249743 |
| KMO | 0.872044843 | 0.974042829 |
| BANK1 | 0.871989608 | 0.916752312 |
| AFG3L2 | 0.871980796 | 0.778066191 |
| EIF4A3 | 0.871971492 | 0.823410288 |
| ORC3 | 0.87196206 | 0.861596437 |

| | | |
|---|---|---|
| GFPT2 | 0.871959392 | 1.041531473 |
| BGLAP | 0.871948976 | 1.078979344 |
| OLIG3 | 0.871948976 | 0.989064399 |
| DMBX1 | 0.87193723 | 0.996916752 |
| SREBF1 | 0.871925544 | 0.795058728 |
| SPANXB2 | 0.871914349 | 0.932853717 |
| TIMM8B | 0.87188816 | 0.868790682 |
| CNBD2 | 0.871860748 | 1.053057918 |
| PIP4K2A | 0.871854923 | 0.95587281 |
| FAM127A | 0.871713531 | 1.105305792 |
| MAP3K14 | 0.871657677 | 0.882543803 |
| GATAD2A | 0.87164303 | 0.911300122 |
| DISC1 | 0.871560345 | 1.07571086 |
| FLT3 | 0.87147459 | 1.029850746 |
| TMEM43 | 0.871457673 | 1.001027749 |
| MICALL1 | 0.87139709 | 1.044147428 |
| DUS3L | 0.87121987 | 0.866747671 |
| RHBDL3 | 0.871212061 | 0.846203764 |
| C12orf74 | 0.871210283 | 0.931825968 |
| LOR | 0.871187084 | 0.789311408 |
| HMGB3 | 0.871178022 | 0.744775608 |
| FBXW9 | 0.871176168 | 0.913692408 |
| GUCY1B3 | 0.871176168 | 0.913692408 |
| PPAPDC1A | 0.871172546 | 1.036340039 |
| SLC25A35 | 0.871151244 | 0.956834532 |
| SGIP1 | 0.871137754 | 0.916565411 |
| ZFP69 | 0.871103573 | 1.011340624 |
| KCTD3 | 0.871088412 | 0.993510707 |
| CER1 | 0.871085999 | 0.946214457 |
| IL36RN | 0.871072856 | 0.997164844 |
| KAT2B | 0.871017285 | 0.883192732 |
| METAP2 | 0.870974559 | 0.828033744 |
| PLCG2 | 0.870939818 | 0.963011032 |
| PRAMEF10 | 0.870874189 | 0.86472256 |
| TMEM219 | 0.870758455 | 0.823569716 |
| TCF3 | 0.870726437 | 0.877352369 |
| NCEH1 | 0.870719229 | 0.997569866 |
| FAM189A2 | 0.870711787 | 0.85748544 |
| INPP4B | 0.870672762 | 0.957170668 |
| PRSS27 | 0.870672762 | 0.957170668 |
| FOXF2 | 0.870668646 | 0.889682025 |
| IFT43 | 0.870666455 | 1 |
| SLC6A13 | 0.870633411 | 1.104477612 |
| LRRC36 | 0.870623003 | 0.827055488 |

| | | |
|---|---|---|
| HIST1H4E | 0.870609206 | 1.064405618 |
| RIN1 | 0.87059149 | 0.959232614 |
| ASPH | 0.870571146 | 0.876873228 |
| GPR64 | 0.870471235 | 1.006489293 |
| ZNF677 | 0.87046407 | 0.801134062 |
| GPR68 | 0.870452959 | 0.939000649 |
| SLC4A10 | 0.870431719 | 0.871512005 |
| PDE8B | 0.870400344 | 0.948157149 |
| NFYC | 0.870377715 | 0.883841661 |
| SLC2A11 | 0.870364035 | 0.857828023 |
| ABCC8 | 0.870353348 | 0.963659961 |
| APOB | 0.870313949 | 1.000405022 |
| SLC9B1 | 0.870294541 | 0.775212637 |
| FYCO1 | 0.870286148 | 0.907845152 |
| TNS3 | 0.87022443 | 0.920830629 |
| AGR3 | 0.870187261 | 0.844066424 |
| CCDC117 | 0.870183186 | 0.957877683 |
| C1QTNF9B | 0.870114017 | 1.023980815 |
| NAA16 | 0.870082916 | 1.037637897 |
| UBL3 | 0.870058332 | 0.979445015 |
| SLC39A9 | 0.870034339 | 0.931956258 |
| PPP4R4 | 0.870016601 | 0.858170606 |
| ZSCAN5B | 0.870003062 | 1.031591738 |
| MMADHC | 0.869966028 | 0.984204131 |
| ME2 | 0.869961654 | 1.074626866 |
| SLC12A3 | 0.869940497 | 0.914990266 |
| UPK2 | 0.869907043 | 1.001027749 |
| XPNPEP3 | 0.869877931 | 0.847501622 |
| SETMAR | 0.869855641 | 1.038882139 |
| RGL4 | 0.869803737 | 0.972593354 |
| PLEKHO1 | 0.869801237 | 1.107735925 |
| ZNF574 | 0.869797348 | 0.929770469 |
| ZNF629 | 0.86977569 | 1.031517643 |
| BCAR3 | 0.869714568 | 0.919150394 |
| ALDH6A1 | 0.869657331 | 0.948967193 |
| PRPSAP2 | 0.869651904 | 0.849606029 |
| TNFSF12-TNFSF13 | 0.869595041 | 0.924631723 |
| PELP1 | 0.869543377 | 0.904009721 |
| GOLGA7 | 0.869491509 | 0.905104488 |
| NME5 | 0.86947002 | 1.062946139 |
| DOK7 | 0.869450208 | 0.773592548 |
| DNAJC7 | 0.869421121 | 1.044147428 |
| CR1 | 0.869420214 | 0.786502271 |
| HERC2 | 0.869400456 | 0.890979883 |

| | | |
|---|---|---|
| PQLC2 | 0.869385801 | 0.978417266 |
| CGB7 | 0.869378914 | 0.944501542 |
| CHMP4C | 0.869352912 | 0.844876468 |
| C2orf76 | 0.869264507 | 0.949982871 |
| IGSF3 | 0.869256442 | 0.974991435 |
| ELMO1 | 0.869240357 | 0.882151422 |
| SMC5 | 0.86922602 | 0.935196436 |
| TSPO | 0.869220086 | 1.057105775 |
| TUBB4A | 0.869183027 | 0.954232483 |
| GUCY2D | 0.869182711 | 0.96495782 |
| SYNPO | 0.869136825 | 1.030489894 |
| LY9 | 0.869121491 | 1.025516403 |
| SCD5 | 0.869111761 | 1.00891049 |
| ITGB1 | 0.869058172 | 0.462940462 |
| PTGIR | 0.869038728 | 1.24789098 |
| HEPH | 0.869033874 | 0.892669097 |
| RRAS2 | 0.869018031 | 0.94491697 |
| TIMP4 | 0.868951146 | 0.900308325 |
| RTN3 | 0.868911908 | 1.082562521 |
| AP1B1 | 0.868883907 | 0.948612539 |
| IL25 | 0.868872495 | 1.021061158 |
| CDR2L | 0.868870458 | 0.778857837 |
| SH3RF1 | 0.86884675 | 0.983776768 |
| ZSCAN4 | 0.868843612 | 1.161198866 |
| FIBP | 0.868839911 | 0.955806783 |
| CD226 | 0.868833749 | 0.905789654 |
| ZNF552 | 0.868740864 | 1.011305242 |
| WNT8B | 0.868724689 | 0.845152449 |
| CARM1 | 0.868664048 | 0.769542325 |
| PEF1 | 0.868653643 | 1.000685166 |
| PSG4 | 0.868645035 | 1.041795135 |
| PDCD7 | 0.868598386 | 0.859862292 |
| KLK8 | 0.868572861 | 0.977936405 |
| FAM109B | 0.868554883 | 0.9145403 |
| ITGB2 | 0.86850752 | 1.054470709 |
| PNPT1 | 0.868480714 | 0.90036452 |
| FAM127B | 0.868479543 | 0.776832726 |
| FOXD3 | 0.868473837 | 0.93600648 |
| RAPH1 | 0.868403625 | 0.854402193 |
| ZBTB22 | 0.868367933 | 0.674767112 |
| ZNF582 | 0.86834948 | 0.459403905 |
| DNAJA4 | 0.868325347 | 0.872012961 |
| OR2W5 | 0.868274629 | 0.974306269 |
| CHMP4A | 0.868221378 | 0.912515188 |

| | | |
|---|---|---|
| KIAA1107 | 0.868207797 | 1.029804728 |
| SLC39A8 | 0.868185505 | 0.416581021 |
| ZNF181 | 0.868152701 | 1.011990408 |
| ISG20 | 0.868053319 | 0.941596366 |
| VLDLR | 0.868048742 | 0.938841636 |
| THAP3 | 0.868010486 | 0.97691373 |
| SLMAP | 0.867993392 | 0.910243234 |
| SULT2B1 | 0.86793703 | 0.908059943 |
| APBB3 | 0.867926606 | 0.892428914 |
| WNK4 | 0.867865375 | 0.861778066 |
| TAAR9 | 0.867829071 | 0.886437378 |
| CCL27 | 0.867808121 | 0.930113052 |
| TMEM245 | 0.867762404 | 1.001712915 |
| BTG1 | 0.867728022 | 0.863102471 |
| C9orf106 | 0.867666211 | 0.926687222 |
| MKI67 | 0.867662859 | 1.023295649 |
| GNB2 | 0.867630507 | 0.933881466 |
| KRTAP4-7 | 0.867594215 | 0.908872902 |
| LOXL4 | 0.867560203 | 0.843608047 |
| ITGA10 | 0.867550037 | 0.908464966 |
| UIMC1 | 0.86752146 | 0.965573107 |
| VWA5A | 0.86746839 | 1.018156903 |
| DPEP3 | 0.867432684 | 0.966904607 |
| FUCA1 | 0.867432684 | 0.966904607 |
| CLEC2B | 0.867424673 | 0.858647226 |
| IKZF4 | 0.86742177 | 0.989469421 |
| ZNF572 | 0.867401332 | 0.864337102 |
| NDUFA7 | 0.867379591 | 0.842041312 |
| LILRA2 | 0.86737922 | 0.91983556 |
| RGS12 | 0.867317781 | 0.950325454 |
| FAR2 | 0.867222766 | 0.971908188 |
| ZNF710 | 0.867209197 | 0.925475901 |
| MYCT1 | 0.867208766 | 0.939705379 |
| HDAC11 | 0.867172265 | 0.8327258 |
| CCSER1 | 0.867162731 | 1.129496403 |
| FXYD5 | 0.867138049 | 0.9733939 |
| IL2RA | 0.86711404 | 0.930336168 |
| SCRIB | 0.867075691 | 0.952380952 |
| BAD | 0.867061818 | 1.121349773 |
| PSG5 | 0.867054079 | 1.03117506 |
| ZMAT4 | 0.867017695 | 1.077766358 |
| NOVA1 | 0.867014433 | 0.911270983 |
| EGLN1 | 0.866975126 | 0.956662616 |
| TTYH3 | 0.866883918 | 0.868916288 |

| | | |
|---|---|---|
| LRRC8B | 0.866881753 | 0.711624139 |
| STK24 | 0.866866465 | 0.918234912 |
| USP22 | 0.866864179 | 0.866342649 |
| DNAI1 | 0.866821023 | 1.078520441 |
| LRCH3 | 0.866805284 | 1.040424803 |
| OR5M10 | 0.866792235 | 0.945529291 |
| KLHL22 | 0.86674031 | 1.038711888 |
| HSPB11 | 0.866664188 | 1.090319968 |
| TMEM132D | 0.866653032 | 0.954637505 |
| AHCYL1 | 0.866560305 | 0.875405581 |
| CAMK2G | 0.866560305 | 0.961713173 |
| INSL3 | 0.866560305 | 0.912394549 |
| NTF4 | 0.866560305 | 0.924724205 |
| LRP11 | 0.866484492 | 1.008564577 |
| FGFBP1 | 0.866466591 | 1.023896314 |
| MAPRE2 | 0.866421738 | 0.983556012 |
| RPP25 | 0.866388889 | 1.037341555 |
| TOP3A | 0.866359718 | 0.931146213 |
| AREG | 0.866314534 | 0.933576347 |
| KCNJ10 | 0.866302716 | 1.091499027 |
| RNF31 | 0.866290532 | 1.042180402 |
| RYR3 | 0.866280571 | 1.005191434 |
| SP2 | 0.86624583 | 0.89422453 |
| VEZF1 | 0.866232247 | 0.857235561 |
| SYT4 | 0.866185902 | 0.845837616 |
| IL19 | 0.866168956 | 0.519698527 |
| FBLN2 | 0.866145984 | 1.023295649 |
| MPP6 | 0.865983302 | 0.974691759 |
| ACSM2B | 0.865983016 | 0.972053463 |
| CNN2 | 0.865980559 | 1.080849606 |
| KRTAP20-3 | 0.865939282 | 0.917780062 |
| OR12D3 | 0.865916717 | 0.924974306 |
| NETO2 | 0.865909694 | 0.978759849 |
| FAM181B | 0.86588636 | 0.908872902 |
| NRXN2 | 0.865832441 | 0.899965742 |
| HGFAC | 0.865751397 | 1.042829332 |
| CAMKK2 | 0.865731886 | 1.018170019 |
| ACR | 0.865721678 | 1.005840363 |
| B3GNT1 | 0.865714 | 0.843661401 |
| ECT2 | 0.865711306 | 0.993510707 |
| ZNF532 | 0.865709109 | 0.996916752 |
| ASB4 | 0.865700604 | 0.981181051 |
| SLC24A3 | 0.865650071 | 0.867557716 |
| KATNAL1 | 0.865576207 | 0.912920211 |

| | | |
|---|---|---|
| RAB43 | 0.865574992 | 0.78725591 |
| RBBP8NL | 0.865557004 | 0.939705379 |
| C9orf142 | 0.865544421 | 0.893884164 |
| PCDHGA12 | 0.865529456 | 1.068859198 |
| ABCA2 | 0.865475779 | 1.036988968 |
| ANKRD9 | 0.86546421 | 1.036656389 |
| PPP1R11 | 0.86546421 | 0.975676602 |
| ZNF146 | 0.865406395 | 0.905789654 |
| JADE1 | 0.865361761 | 0.93835172 |
| SLC1A5 | 0.865350899 | 0.94653706 |
| SEC16B | 0.865205363 | 0.848926691 |
| TRIM39 | 0.865196868 | 1.031148605 |
| MAGEA9 | 0.865178216 | 1.036998972 |
| OR4F17 | 0.865178216 | 1.036998972 |
| RB1CC1 | 0.865174442 | 0.765492102 |
| MYCBP | 0.865146476 | 0.937221547 |
| LSM7 | 0.865134674 | 0.952723535 |
| TAS2R16 | 0.865070651 | 1.019184652 |
| ANK2 | 0.865065124 | 0.899149453 |
| HLCS | 0.865021927 | 0.944511948 |
| LIMCH1 | 0.864987599 | 1.024665982 |
| IRGC | 0.864974298 | 0.818144998 |
| LTA | 0.864953707 | 0.888317917 |
| CYP3A4 | 0.864953603 | 1.049967554 |
| UBA3 | 0.86489894 | 0.846203764 |
| MMS22L | 0.864727567 | 0.875984926 |
| TRAT1 | 0.864719037 | 1.028351559 |
| CHRD | 0.864699611 | 0.935251799 |
| RAB33B | 0.864690004 | 0.902011681 |
| VPS54 | 0.864633457 | 0.901984609 |
| DDX17 | 0.864603606 | 0.837586067 |
| ST7L | 0.864582803 | 0.910243234 |
| NUP88 | 0.864576142 | 0.962315862 |
| ACTR1A | 0.864542432 | 0.983898595 |
| ATP6V1H | 0.864531531 | 0.970149254 |
| C19orf52 | 0.864519963 | 0.775617659 |
| ARHGAP4 | 0.86447927 | 1.027064063 |
| DOPEY2 | 0.864425723 | 0.914135277 |
| TBC1D15 | 0.864417106 | 0.966084275 |
| DGKG | 0.864359064 | 0.766385464 |
| SLC16A1 | 0.864335661 | 0.96435804 |
| GPR126 | 0.864313869 | 1.001297859 |
| SLC39A2 | 0.864310904 | 0.876063183 |
| ZNF518B | 0.864275297 | 0.990684488 |

| | | |
|---|---|---|
| NANS | 0.864202554 | 0.95018226 |
| COG3 | 0.864187567 | 0.901678657 |
| ADRM1 | 0.8641602 | 0.989722508 |
| ITGB4 | 0.864101489 | 0.888213852 |
| ZNF155 | 0.864082585 | 0.921548476 |
| ZNF25 | 0.86407973 | 0.995203837 |
| IGFBP7 | 0.864065373 | 0.93600648 |
| GFAP | 0.864004199 | 0.91983556 |
| NFATC1 | 0.864002971 | 0.916970433 |
| C19orf70 | 0.863985396 | 0.826245443 |
| RNF125 | 0.863961937 | 0.865671642 |
| MAP3K8 | 0.863953705 | 0.970798183 |
| CMTM6 | 0.863909597 | 1.014985824 |
| TMEM214 | 0.863836101 | 0.835560123 |
| PRRT2 | 0.863829265 | 0.881328473 |
| HCLS1 | 0.863816074 | 1.142857143 |
| GDF11 | 0.863814167 | 0.921479559 |
| GYG2 | 0.863814167 | 0.921479559 |
| GRPR | 0.863804947 | 0.816353018 |
| ADAMTS17 | 0.863754071 | 1.001946788 |
| SMAD3 | 0.863719084 | 0.989617132 |
| HOXA6 | 0.863692741 | 0.905789654 |
| FBXO38 | 0.863574442 | 0.847501622 |
| ULK1 | 0.863566539 | 1.033095393 |
| TNFSF14 | 0.863554909 | 1.004796163 |
| LMX1B | 0.863531394 | 0.902363823 |
| PTPN5 | 0.863493271 | 1.00843608 |
| SLC7A11 | 0.863470238 | 0.903199676 |
| CYP1A1 | 0.863438197 | 0.810512654 |
| GCH1 | 0.863404609 | 0.890979883 |
| PLXNB3 | 0.863396235 | 1.037341555 |
| BBX | 0.863291162 | 0.882836588 |
| PRDX3 | 0.863247936 | 0.965573107 |
| LYST | 0.863222669 | 0.822195221 |
| IRF9 | 0.863194932 | 1.002595717 |
| METTL2B | 0.863185382 | 1.083062946 |
| DCUN1D4 | 0.86311409 | 0.897469176 |
| CCDC50 | 0.86296371 | 0.969510106 |
| KLHL7 | 0.862963075 | 0.962315862 |
| TONSL | 0.862959156 | 0.886998785 |
| SLC25A39 | 0.862941785 | 1.0854597 |
| CRTAM | 0.862921001 | 0.963547995 |
| NFE2L2 | 0.862905363 | 0.717294451 |
| BCAP29 | 0.862893217 | 0.996755354 |

| | | |
|---|---|---|
| SLC25A4 | 0.86282777 | 0.903958469 |
| QRFP | 0.86280866 | 0.958890031 |
| KRTAP2-4 | 0.862795883 | 0.908530319 |
| SPATA5L1 | 0.862776236 | 0.891628812 |
| CYP46A1 | 0.862678679 | 0.930336168 |
| ATP13A1 | 0.862627889 | 0.786502271 |
| TNIP1 | 0.862603816 | 0.847550531 |
| PRSS33 | 0.862578001 | 0.706035042 |
| SELENBP1 | 0.862545049 | 0.896197328 |
| TG | 0.862512156 | 0.992709599 |
| POLE3 | 0.862492595 | 0.815714864 |
| AARS | 0.862479197 | 0.842041312 |
| MRPS31 | 0.862435989 | 0.979102432 |
| ZNF550 | 0.86240323 | 0.92874272 |
| HMOX2 | 0.862383389 | 1.009733939 |
| SLC25A22 | 0.862373089 | 0.823005265 |
| ZNF746 | 0.862268662 | 0.933171324 |
| PPP3CC | 0.862267396 | 0.916937054 |
| CALN1 | 0.862205091 | 0.973963686 |
| SULF1 | 0.862168937 | 1.152498378 |
| SFT2D2 | 0.862168666 | 0.914135277 |
| MSANTD2 | 0.862138 | 0.863912515 |
| CCL4L2 | 0.862131836 | 0.942891859 |
| ZFHX4 | 0.862097146 | 0.971648441 |
| KLHL31 | 0.862064544 | 0.816119887 |
| LAMA4 | 0.861987166 | 0.820980154 |
| CCDC136 | 0.861958996 | 0.902389631 |
| PLCH2 | 0.86195786 | 0.855288774 |
| STEAP2 | 0.861876932 | 1.014985824 |
| C16orf13 | 0.861871939 | 0.988659376 |
| CTDSPL2 | 0.861871939 | 0.988659376 |
| CHRM5 | 0.861867716 | 0.898767034 |
| ASTE1 | 0.861848914 | 0.883353584 |
| MED12 | 0.861829518 | 1.010382868 |
| PROCR | 0.861785887 | 0.706683971 |
| OPN1MW | 0.861757523 | 0.922233642 |
| DEFA6 | 0.86175118 | 1.089756766 |
| AK9 | 0.861742661 | 1.082562521 |
| LAMTOR3 | 0.861696298 | 0.948157149 |
| ZMIZ1 | 0.861674597 | 1.058323208 |
| COL21A1 | 0.86166713 | 0.969167523 |
| ONECUT3 | 0.86159941 | 0.818955043 |
| TPM3 | 0.861591826 | 0.918807811 |
| C10orf10 | 0.861549743 | 0.986634265 |

| | | |
|---|---|---|
| TMEM65 | 0.861535683 | 0.960307817 |
| PPIC | 0.861522206 | 1.171966256 |
| HSPA13 | 0.861503234 | 1.01782098 |
| TTLL2 | 0.861457745 | 0.993924666 |
| XDH | 0.861414344 | 1.025111381 |
| SUPT20H | 0.861405378 | 0.914945322 |
| COMMD7 | 0.861358934 | 0.879410757 |
| MBD3L2 | 0.861313806 | 0.899280576 |
| FBXL8 | 0.861206743 | 0.78780013 |
| H2AFB3 | 0.861201261 | 0.845686513 |
| BRSK1 | 0.861096101 | 0.874756652 |
| KRTCAP3 | 0.861087303 | 0.984241178 |
| FGF19 | 0.861080889 | 0.96800324 |
| FRMPD2 | 0.861072986 | 0.977046934 |
| DNM1L | 0.861059138 | 1.073329007 |
| NDRG4 | 0.861049561 | 0.928400137 |
| CLIP4 | 0.860989757 | 1.004050223 |
| COX6C | 0.860860522 | 0.869987849 |
| GPI | 0.860831741 | 0.901174565 |
| CEACAM4 | 0.860820203 | 0.998972251 |
| CYP11A1 | 0.860783273 | 0.924724205 |
| TRIML1 | 0.860783273 | 0.924724205 |
| HOGA1 | 0.860726141 | 0.894484412 |
| SLFN13 | 0.86067077 | 1.002025111 |
| UBAC2 | 0.860589317 | 0.860672337 |
| PFAS | 0.860578123 | 0.987021415 |
| TUBA3D | 0.860546884 | 0.982989064 |
| PSORS1C2 | 0.860530939 | 1.01417578 |
| NRIP3 | 0.860489534 | 0.869475848 |
| TMPRSS7 | 0.860457767 | 0.875405581 |
| NUP188 | 0.860408747 | 0.983213429 |
| TAF8 | 0.860387134 | 1.003083248 |
| EIF3C | 0.860371904 | 0.937626569 |
| PGR | 0.860370586 | 0.863075925 |
| PLXDC1 | 0.860295596 | 1.004860267 |
| SSB | 0.860287227 | 0.912983899 |
| UBE3C | 0.860255992 | 0.981181051 |
| YAE1D1 | 0.860230182 | 0.999657417 |
| LRRC4C | 0.860197791 | 0.952207371 |
| DCBLD2 | 0.860143867 | 1.071942446 |
| MLLT6 | 0.860093486 | 1 |
| PTK6 | 0.860093486 | 0.913043478 |
| TP53I13 | 0.860029026 | 0.748481166 |
| ADAMTSL1 | 0.860019231 | 1.074626866 |

| | | |
|---|---|---|
| GAN | 0.860003317 | 0.900650908 |
| LYPLA2 | 0.860000448 | 0.938031592 |
| CHCHD3 | 0.859974454 | 0.940390545 |
| KANSL1L | 0.859925022 | 0.972935937 |
| ATAD2 | 0.859922336 | 0.931862427 |
| SMU1 | 0.859811974 | 0.870161014 |
| FGF13 | 0.859788843 | 0.892669097 |
| KMT2C | 0.859757937 | 0.950681376 |
| LNX2 | 0.859734969 | 1.112264763 |
| GUCA1B | 0.859722671 | 0.962332928 |
| DHX15 | 0.859702754 | 0.739165654 |
| KIAA0825 | 0.859571481 | 0.878493317 |
| SMC4 | 0.859567966 | 0.813689753 |
| HRH2 | 0.859551305 | 0.882543803 |
| PDLIM2 | 0.859526977 | 0.904761905 |
| CGB2 | 0.859522582 | 0.924631723 |
| TRDMT1 | 0.859513256 | 0.883353584 |
| RNF151 | 0.859482566 | 0.913692408 |
| MED6 | 0.859452024 | 0.870214147 |
| TPCN2 | 0.859418353 | 1.062946139 |
| CST8 | 0.859380128 | 0.8886605 |
| TEKT4P2 | 0.85937306 | 1.007879411 |
| ADAP1 | 0.859348366 | 0.941075711 |
| PRLR | 0.859331502 | 0.998784933 |
| TAAR8 | 0.859323929 | 0.932511356 |
| OTX2 | 0.859319912 | 0.973621103 |
| ANO7 | 0.85929105 | 0.926687222 |
| BECN1 | 0.859286225 | 0.889033095 |
| CFH | 0.859269347 | 1.058581706 |
| ACAD10 | 0.859264159 | 0.770757392 |
| TEKT3 | 0.859218736 | 0.793079822 |
| SERPINB12 | 0.859183897 | 0.876703439 |
| KRTAP1-3 | 0.859117016 | 0.923261391 |
| ITPRIPL1 | 0.859091144 | 0.935936965 |
| PROL1 | 0.859029523 | 1.013703323 |
| CCDC125 | 0.85901175 | 0.790198461 |
| ARMCX3 | 0.859011399 | 0.954093868 |
| LOC643834 | 0.858910889 | 1.006480356 |
| CD3E | 0.858890649 | 0.872901679 |
| ADRB3 | 0.858859049 | 0.839714471 |
| ENO2 | 0.858842832 | 1.020116807 |
| FBXL12 | 0.858842832 | 0.765087605 |
| PRKCG | 0.858842832 | 1.020116807 |
| PTPMT1 | 0.858748407 | 1.007787151 |

| | | |
|---|---|---|
| TMEM132B | 0.858746199 | 1.023491292 |
| NEB | 0.858729695 | 0.989874443 |
| CBWD3 | 0.858677377 | 0.999657417 |
| GLDN | 0.858630871 | 0.785743216 |
| ADAMTS20 | 0.858625992 | 0.815055159 |
| PATZ1 | 0.858617678 | 1.0266061 |
| ZRANB3 | 0.858614366 | 0.965978129 |
| COTL1 | 0.858598626 | 0.824220332 |
| PCDHB11 | 0.85854983 | 0.938335046 |
| SLC38A2 | 0.858542337 | 0.720939652 |
| GOLGA8E | 0.858517682 | 0.939651681 |
| MSC | 0.858514472 | 0.831510733 |
| PDAP1 | 0.858497633 | 0.943816375 |
| MTF2 | 0.858402979 | 0.896171317 |
| NLK | 0.858395405 | 1.033095393 |
| AKR1E2 | 0.858346423 | 0.821544452 |
| RBM48 | 0.858301171 | 0.998287085 |
| FOXL2 | 0.858296864 | 1.020765737 |
| CAGE1 | 0.858296143 | 1.036046983 |
| DOHH | 0.858277804 | 0.963953017 |
| CLU | 0.858272989 | 0.945871874 |
| TRA2A | 0.858221517 | 0.828365879 |
| KIAA1328 | 0.858174984 | 0.911613566 |
| IL1RN | 0.858096508 | 0.831915755 |
| ALKBH3 | 0.858025243 | 0.762251924 |
| PIAS1 | 0.858016718 | 0.921479559 |
| LIMA1 | 0.857983472 | 0.995544755 |
| TMEM254 | 0.857963351 | 0.921020656 |
| CAAP1 | 0.857895605 | 0.977046934 |
| B3GAT1 | 0.857877263 | 0.971447112 |
| CEP85L | 0.857871047 | 0.886605002 |
| TTPA | 0.857863508 | 0.853786958 |
| SYCN | 0.857857409 | 0.969852689 |
| MFRP | 0.857818669 | 0.962658445 |
| ZNF529 | 0.857788473 | 0.861077359 |
| DEK | 0.857755566 | 0.942891859 |
| GLRA1 | 0.857745052 | 1.052563271 |
| ZNF773 | 0.857719602 | 0.788983394 |
| XKR3 | 0.857700958 | 0.90927501 |
| MAGEB1 | 0.857692437 | 0.953751285 |
| GCM1 | 0.857634583 | 0.995949777 |
| RTDR1 | 0.857618809 | 1.022610483 |
| FBXO10 | 0.857533646 | 0.809863725 |
| FOXM1 | 0.857496688 | 0.931146213 |

| | | |
|---|---|---|
| TMEM156 | 0.857484462 | 0.810854597 |
| FAM171A1 | 0.857455558 | 1.019184652 |
| NHLRC3 | 0.85743072 | 0.938436614 |
| TRIM75_pseudogene | 0.857408761 | 0.890979883 |
| DNAJA1 | 0.857387319 | 0.943296881 |
| SORCS3 | 0.857336234 | 1.127839066 |
| PGAP3 | 0.857294392 | 0.9777321 |
| TMEM89 | 0.857254791 | 0.907159986 |
| HSF1 | 0.857237657 | 0.835155934 |
| TRAF5 | 0.857234318 | 1.115509409 |
| RNF19A | 0.857185748 | 0.959766385 |
| HP | 0.857148208 | 0.866320571 |
| BACH2 | 0.857092722 | 1.009733939 |
| DAPK2 | 0.857023488 | 1.090850097 |
| MS4A2 | 0.857012375 | 0.853990915 |
| RAD1 | 0.857008477 | 0.816119887 |
| ZBTB10 | 0.856974856 | 1.035236938 |
| ZNF777 | 0.856957842 | 0.950992305 |
| NCOA3 | 0.856937568 | 0.953422438 |
| MELK | 0.856931863 | 1.109669046 |
| OR51I1 | 0.856919047 | 1.014388489 |
| EIF4B | 0.856888411 | 0.784933171 |
| C9orf173 | 0.856862274 | 0.969167523 |
| EEF1A1 | 0.856849544 | 0.876468206 |
| NEFH | 0.856832778 | 0.89311408 |
| CES5A | 0.856823208 | 1.011340624 |
| GABARAPL1 | 0.856805501 | 0.936622131 |
| EXOSC7 | 0.85666891 | 0.717699473 |
| OR5M3 | 0.856654258 | 0.983898595 |
| SCAMP3 | 0.856542227 | 1.010382868 |
| RNF130 | 0.856510779 | 0.979234263 |
| C12orf40 | 0.856472128 | 0.828675577 |
| FAM76A | 0.856428625 | 0.833535844 |
| ADORA1 | 0.856402771 | 0.885788449 |
| DLG5 | 0.85638252 | 0.966904607 |
| NCOA1 | 0.856340906 | 0.937221547 |
| NKTR | 0.856340906 | 0.937221547 |
| RND1 | 0.856265565 | 0.946942082 |
| DPP9 | 0.856212358 | 0.842310188 |
| MUC21 | 0.85619509 | 0.993490922 |
| PRRG2 | 0.856097718 | 0.872417983 |
| TMEM201 | 0.856001845 | 1.008221994 |
| ERCC3 | 0.855992472 | 1.011031798 |
| MANF | 0.855986485 | 0.99554642 |

| | | |
|---|---|---|
| TMPRSS9 | 0.855977737 | 0.929915639 |
| ZFP37 | 0.855972048 | 0.838801134 |
| ZNF70 | 0.855914397 | 1.037341555 |
| TRUB1 | 0.855868181 | 0.901579587 |
| KLF12 | 0.85586199 | 0.998702141 |
| AIRE | 0.855835564 | 1.048669695 |
| IPMK | 0.855811444 | 1.098637249 |
| CASK | 0.855783618 | 1.01752109 |
| AIF1L | 0.855778786 | 0.997601918 |
| LIN7B | 0.855697319 | 0.916067146 |
| COL16A1 | 0.855688057 | 0.925880923 |
| SHE | 0.855643942 | 0.983799109 |
| HSD17B10 | 0.85564301 | 0.824140169 |
| TMEM138 | 0.85560482 | 0.781287971 |
| OXNAD1 | 0.855577234 | 1.024010383 |
| SULT2A1 | 0.855539365 | 0.892926671 |
| DHX32 | 0.855501412 | 0.899554475 |
| NRG4 | 0.855495873 | 1.077766358 |
| STK39 | 0.85546725 | 0.861778066 |
| KLRG1 | 0.85533876 | 0.75982179 |
| KATNAL2 | 0.855334163 | 1.074340528 |
| TMEM255A | 0.85531059 | 0.817739976 |
| PHYKPL | 0.855234605 | 0.968202466 |
| FGF10 | 0.85522878 | 1.049412718 |
| COQ3 | 0.855221433 | 0.818299805 |
| STEAP1 | 0.855184105 | 0.945727015 |
| C15orf56 | 0.855103174 | 0.856800274 |
| FGFR2 | 0.855099265 | 1.005840363 |
| PINK1 | 0.855021552 | 0.974691759 |
| CSDC2 | 0.855018906 | 0.9145403 |
| OR5H14 | 0.854998691 | 0.873244262 |
| CCDC23 | 0.854984318 | 0.977318753 |
| ARID4B | 0.854956235 | 1.005823912 |
| EDN1 | 0.854929186 | 0.938677629 |
| ADD2 | 0.854927554 | 0.993148338 |
| DSN1 | 0.854923495 | 0.871202916 |
| STT3A | 0.854904648 | 0.815714864 |
| PCDHA7 | 0.854898013 | 1.016786571 |
| ZNF626 | 0.854869377 | 1.0222679 |
| NADK2 | 0.854858139 | 1.057108141 |
| CUL9 | 0.854811299 | 0.981181051 |
| GTF2H3 | 0.854803124 | 0.948562171 |
| COMTD1 | 0.854777628 | 1.049967554 |
| C1QTNF9B-AS1 | 0.854721441 | 0.995203837 |

| | | |
|---|---|---|
| TNFAIP8L3 | 0.854716271 | 0.902794654 |
| SERPINB3 | 0.854670476 | 0.910085055 |
| PRKX | 0.8546618 | 0.968851395 |
| TNNT2 | 0.854656467 | 0.969852689 |
| PBX3 | 0.854653403 | 0.850097339 |
| RILPL1 | 0.854588548 | 1.040767386 |
| C2orf78 | 0.854569156 | 1.050222762 |
| ZNF438 | 0.85452983 | 0.808829486 |
| PARN | 0.854415928 | 0.825840421 |
| PLA2G4F | 0.854356073 | 0.963547995 |
| COPG1 | 0.854351579 | 1.071942446 |
| C14orf28 | 0.854351035 | 0.910243234 |
| MED11 | 0.854302535 | 0.97326853 |
| MRPL52 | 0.854278087 | 0.944844125 |
| TUBB4B | 0.854250168 | 0.982989064 |
| HTR3C | 0.854246314 | 0.981829981 |
| MEAF6 | 0.85419877 | 0.661806399 |
| COL26A1 | 0.854133413 | 0.995889003 |
| COPE | 0.854108357 | 1.001370332 |
| PCDHGB7 | 0.854078634 | 0.857828023 |
| OR2G3 | 0.854073287 | 0.923261391 |
| LRTOMT | 0.854062911 | 0.946899623 |
| DDC | 0.854031176 | 0.889023896 |
| BTBD9 | 0.854011304 | 0.957862282 |
| ZNF767P | 0.853967909 | 1.038882139 |
| RAMP3 | 0.853965858 | 0.87975334 |
| PABPC1 | 0.853868321 | 0.848116646 |
| DPYSL4 | 0.853833172 | 0.925475901 |
| HLA-G | 0.853816752 | 0.838416613 |
| MPV17L2 | 0.853781539 | 0.918122645 |
| CLSTN3 | 0.853772794 | 0.989037342 |
| ZC3H3 | 0.853731653 | 0.929085303 |
| AGAP6 | 0.853709591 | 0.949777238 |
| HES5 | 0.853613467 | 1.044147428 |
| NDUFA6 | 0.853610869 | 0.597002835 |
| MYOZ3 | 0.853568221 | 0.99109284 |
| SNX12 | 0.853481997 | 0.889345666 |
| MYO19 | 0.853452455 | 0.871188763 |
| HAVCR1 | 0.853389659 | 0.832576249 |
| SHANK3 | 0.853317214 | 0.976638546 |
| IL18RAP | 0.853269366 | 0.967193196 |
| ACOX1 | 0.853230652 | 0.894694208 |
| FZD7 | 0.853090496 | 1.127190136 |
| LINC00174 | 0.853056631 | 0.858855773 |

| | | |
|---|---|---|
| WAPAL | 0.853030508 | 0.853786958 |
| RGAG4 | 0.853012831 | 0.943296881 |
| TCEANC2 | 0.853007686 | 0.858647226 |
| JOSD1 | 0.852886538 | 1.019184652 |
| BCL11A | 0.85285507 | 0.892669097 |
| SPDEF | 0.852845008 | 0.902011681 |
| FOXJ1 | 0.852772729 | 0.958468527 |
| SMPD2 | 0.852743052 | 0.919400567 |
| IFNAR2 | 0.852677316 | 0.870863076 |
| DUSP1 | 0.852574083 | 0.96495782 |
| GATA2 | 0.852533179 | 0.779262859 |
| PCID2 | 0.852489625 | 0.788983394 |
| SLC25A27 | 0.852378071 | 0.971447112 |
| SLC22A18AS | 0.852358772 | 0.899623159 |
| TTC14 | 0.852344001 | 1.043508051 |
| OR1L4 | 0.852324971 | 0.910585817 |
| C1orf158 | 0.852323662 | 0.931956258 |
| CD164L2 | 0.852306495 | 1.04090725 |
| OR11A1 | 0.852289449 | 0.886947585 |
| KIAA0556 | 0.852214853 | 0.963547995 |
| CLEC7A | 0.852174814 | 0.975698663 |
| CX3CR1 | 0.852163784 | 1.015574302 |
| ZNF44 | 0.852160588 | 0.930798219 |
| CTNS | 0.852159074 | 0.980558931 |
| THOC2 | 0.85214909 | 0.76508708 |
| HOXB3 | 0.852051659 | 0.852693056 |
| LRRC19 | 0.851989771 | 0.916409729 |
| RIBC2 | 0.851940693 | 1.05346294 |
| PCDHB2 | 0.851878626 | 0.960260363 |
| DNAJB8 | 0.851829797 | 1.094775213 |
| DGUOK | 0.851809135 | 0.972096042 |
| ADCY2 | 0.851755577 | 0.874443094 |
| MEGF9 | 0.851747987 | 0.876873228 |
| GTDC1 | 0.851729649 | 1.026036314 |
| GDI2 | 0.851688438 | 0.896314297 |
| ZBP1 | 0.851687572 | 1.024706359 |
| LIPN | 0.851619627 | 0.903309539 |
| TWF1 | 0.851619627 | 0.903309539 |
| TXNDC12 | 0.851619627 | 1.03504218 |
| C12orf61 | 0.85161325 | 1.053867963 |
| OR10AD1 | 0.85159082 | 1.000685166 |
| CRYBB1 | 0.85151393 | 0.891401165 |
| PDCD2 | 0.851451852 | 1.097988319 |
| POR | 0.851367922 | 0.872417983 |

| | | |
|---|---|---|
| WNT3 | 0.851230563 | 0.926344639 |
| SH3BP4 | 0.851220686 | 0.931825968 |
| OSBPL10 | 0.851208174 | 0.935601458 |
| FARSA | 0.851200273 | 0.878650227 |
| GLRA4 | 0.851191052 | 0.859831278 |
| CACNG4 | 0.851138448 | 0.765736535 |
| SGSM1 | 0.851136954 | 0.981157931 |
| PYCR1 | 0.851125498 | 0.972053463 |
| RNASE12 | 0.851110312 | 0.935594382 |
| C11orf70 | 0.851058664 | 0.836371 |
| NXPH3 | 0.851040671 | 0.979445015 |
| LZTS1 | 0.851032663 | 0.941596366 |
| ESRP2 | 0.850951079 | 0.88254354 |
| CCL18 | 0.850914249 | 0.899554475 |
| TTC7B | 0.850910887 | 1.081409478 |
| PAFAH1B2 | 0.850903959 | 0.904414743 |
| TACC1 | 0.8508979 | 1.083590271 |
| ICAM4 | 0.850876152 | 0.881123672 |
| RBFA | 0.850869219 | 0.959575197 |
| FAM19A5 | 0.850844614 | 0.933576347 |
| CTCF | 0.850810225 | 0.910447761 |
| CSNK1E | 0.850708556 | 1.011031798 |
| EIF3H | 0.850703051 | 1.011340624 |
| STRN4 | 0.85057729 | 0.854059609 |
| NRN1L | 0.850573714 | 0.875658161 |
| PDCL3 | 0.85057072 | 0.859540939 |
| SH3TC2 | 0.850498541 | 0.925316889 |
| C1QTNF7 | 0.850394148 | 1.040424803 |
| ADAM9 | 0.85038305 | 1.042829332 |
| CKLF | 0.850375607 | 1.002025111 |
| AKAP2 | 0.850318209 | 1.016786571 |
| GALNTL6 | 0.850311117 | 0.788578372 |
| ADAM28 | 0.850286814 | 0.967553537 |
| ST8SIA1 | 0.850221004 | 0.84690158 |
| NCKAP5 | 0.850209829 | 1.036998972 |
| MTAP | 0.850118614 | 0.924665857 |
| SLC16A6 | 0.850095589 | 0.944106926 |
| IFT172 | 0.850090037 | 0.948967193 |
| FBXO36 | 0.850045772 | 0.936404932 |
| OR8A1 | 0.850042319 | 0.79616307 |
| PDZRN3 | 0.850035234 | 0.697598962 |
| KRT17 | 0.849986781 | 1.048602673 |
| SMIM21 | 0.849984438 | 1.051032807 |
| SLC22A9 | 0.849944843 | 0.879948086 |

| | | |
|---|---|---|
| ATP1A3 | 0.84988271 | 0.781692993 |
| DLD | 0.849870012 | 0.842310188 |
| MYO1B | 0.849865521 | 0.936964714 |
| TAOK1 | 0.849854206 | 1.043478261 |
| F8A1 | 0.84983121 | 0.830295666 |
| CDH17 | 0.84982177 | 1.024659312 |
| PPA1 | 0.849789846 | 0.874038072 |
| RPL41 | 0.849742641 | 1.033915725 |
| SFTPA2B_pseudogene | 0.849729177 | 0.946942082 |
| MARCKSL1 | 0.849693722 | 0.995544755 |
| HPS3 | 0.849692092 | 0.997974889 |
| TRIM45 | 0.849666344 | 0.848799481 |
| MXD3 | 0.849554924 | 0.997601918 |
| SLC11A1 | 0.849540636 | 0.974691759 |
| FBXO15 | 0.849499722 | 0.95587281 |
| TGIF2LX | 0.849499722 | 0.95587281 |
| S100PBP | 0.849468719 | 0.73835561 |
| SATB1 | 0.849460635 | 1.028776978 |
| JMJD4 | 0.849448718 | 0.913669065 |
| HIST1H2AC | 0.849446135 | 0.891743748 |
| EPB41L1 | 0.8494243 | 0.816119887 |
| YIPF5 | 0.849349581 | 0.944844125 |
| STK11IP | 0.849348357 | 0.939362795 |
| ZFYVE19 | 0.84934738 | 0.998379911 |
| CTSA | 0.849322562 | 0.962362103 |
| PRRX1 | 0.849283953 | 1.025308241 |
| NCR3 | 0.84927679 | 0.943543154 |
| KCNJ15 | 0.849249788 | 1.08825438 |
| ATG4B | 0.849240321 | 1.006489293 |
| OR6X1 | 0.849137849 | 0.930455635 |
| IL13 | 0.849117188 | 0.886605002 |
| NPRL2 | 0.849010665 | 1.069544365 |
| IQCG | 0.849002922 | 0.991494532 |
| THOC6 | 0.849002922 | 0.95018226 |
| SNF1LK | 0.84886929 | 0.93770279 |
| KIAA0196 | 0.848845149 | 1.09352518 |
| S100A11 | 0.848791638 | 1.016786571 |
| CAMK1D | 0.848754203 | 0.900064893 |
| XRCC6 | 0.848754203 | 0.900064893 |
| TXLNG | 0.8487495 | 0.843096951 |
| PYDC1 | 0.848647615 | 0.967598218 |
| SETDB2 | 0.848633599 | 0.925373134 |
| SEPT14 | 0.848623826 | 0.906844876 |
| SEC23B | 0.848621788 | 0.901984609 |

| | | |
|---|---|---|
| PAX7 | 0.848610123 | 0.875253139 |
| ACOT6 | 0.848596522 | 0.846091535 |
| ZNF404 | 0.848592367 | 1.007879411 |
| CCDC36 | 0.848582004 | 0.816929931 |
| TAAR2 | 0.848579513 | 0.969500324 |
| MDH2 | 0.848576961 | 0.807209397 |
| FNIP1 | 0.848574363 | 0.802349129 |
| ACOT11 | 0.848539586 | 0.717715769 |
| FLNC | 0.848506904 | 0.887735237 |
| FAM169B | 0.84828331 | 0.991778006 |
| KLRC2 | 0.848277374 | 0.911270983 |
| DGKZ | 0.848269083 | 0.980815348 |
| DNPEP | 0.848238118 | 1.089552239 |
| ZC2HC1C | 0.848227217 | 1.022953066 |
| IRX2 | 0.848168314 | 0.909558068 |
| ATP6AP1 | 0.84816404 | 0.819765087 |
| THOC3 | 0.848102706 | 0.935251799 |
| P2RY11 | 0.848068532 | 0.881894873 |
| CDK5R1 | 0.848011812 | 0.970149254 |
| BATF3 | 0.848007602 | 1.036452005 |
| EDN2 | 0.847933837 | 0.778006166 |
| SLC8B1 | 0.847871083 | 0.979445015 |
| IFITM1 | 0.847845619 | 1.021582734 |
| NAF1 | 0.847845619 | 0.510791367 |
| ARHGEF26 | 0.847836331 | 1.016101405 |
| GAGE2B | 0.847836331 | 1.016101405 |
| ADC | 0.847805523 | 0.861482382 |
| MRGPRX3 | 0.84778888 | 0.913692408 |
| GPX7 | 0.847708368 | 0.894873459 |
| CCDC93 | 0.84770079 | 1.056298096 |
| CDKN2A | 0.847656258 | 1.115509409 |
| CLTB | 0.847655727 | 0.970537855 |
| PITPNC1 | 0.847655727 | 0.970537855 |
| KCNK4 | 0.847607934 | 0.96430889 |
| C19orf53 | 0.847550465 | 0.946942082 |
| SLC12A1 | 0.847546696 | 0.944511948 |
| IL1A | 0.847521001 | 1.033744322 |
| ORMDL3 | 0.847517623 | 0.714285714 |
| TMEM178A | 0.84747594 | 0.900769542 |
| GDPD1 | 0.847445016 | 1.014925373 |
| OCA2 | 0.847388883 | 0.852166869 |
| ELF5 | 0.847375371 | 1.128487995 |
| AKR1C2 | 0.847360815 | 1.046982584 |
| HIPK2 | 0.847291154 | 1.021414666 |

| | | |
|---|---|---|
| NPM1 | 0.847276583 | 0.933160286 |
| DMXL1 | 0.847265521 | 0.974648852 |
| KANK1 | 0.847252562 | 0.884892086 |
| PPIL2 | 0.84718718 | 0.914341337 |
| GAS8 | 0.847175219 | 0.978417266 |
| TMED4 | 0.847135405 | 0.920615634 |
| ANKDD1A | 0.847058332 | 0.884163629 |
| ODF3 | 0.847019312 | 1.040097205 |
| CREBZF | 0.846991091 | 1.025516403 |
| ACAD9 | 0.846957216 | 1.008505468 |
| SH3RF2 | 0.84686579 | 0.8137573 |
| ACKR2 | 0.84681442 | 0.996755354 |
| SLC44A5 | 0.846809133 | 0.940461725 |
| MASP1 | 0.846753172 | 0.908500973 |
| DECR2 | 0.846750959 | 0.916160389 |
| STAM2 | 0.846739929 | 0.861596437 |
| PTPRF | 0.846722054 | 0.977936405 |
| GATA3 | 0.846702158 | 0.89671932 |
| F11R | 0.846695842 | 0.894289186 |
| N6AMT1 | 0.846661875 | 1.003244646 |
| IP6K3 | 0.846650268 | 0.889682025 |
| UNC93B1 | 0.846644647 | 0.874848117 |
| PTGS1 | 0.846619482 | 0.721344674 |
| ZFP2 | 0.846599769 | 1.001620089 |
| DBT | 0.84658413 | 0.852976914 |
| OR51A4 | 0.846569656 | 0.942446043 |
| EEF2K | 0.84652596 | 0.940298507 |
| RRAGC | 0.846461728 | 0.909558068 |
| SLC43A1 | 0.846458208 | 1.03504218 |
| RDH16 | 0.846404362 | 0.926285946 |
| FBXW12 | 0.846366253 | 0.9467878 |
| CAMSAP3 | 0.84631855 | 0.959232614 |
| PLIN1 | 0.846269706 | 0.885919836 |
| FZD4 | 0.846259726 | 0.927968851 |
| MRPL15 | 0.84623808 | 1.082219938 |
| CYP2C19 | 0.846215931 | 1.022712524 |
| ZNF76 | 0.846172653 | 0.94655704 |
| PNPLA7 | 0.846072843 | 0.938841636 |
| IRF1 | 0.846030003 | 0.80963953 |
| IRAK3 | 0.845942123 | 0.940947437 |
| CCNC | 0.84591284 | 0.890238963 |
| OR52K2 | 0.84586241 | 0.985419198 |
| GCC1 | 0.845821005 | 0.987324426 |
| CEP72 | 0.845779094 | 1.027749229 |

| | | |
|---|---|---|
| KIAA1456 | 0.845759837 | 1.073655361 |
| IFNA8 | 0.845731111 | 0.862624186 |
| SUPT3H | 0.845705527 | 0.903309539 |
| THSD7B | 0.845690792 | 0.934386391 |
| TMCO5A | 0.845638324 | 0.919805589 |
| ANO10 | 0.845637254 | 0.817334953 |
| TGM3 | 0.845620495 | 0.914945322 |
| USP48 | 0.845616206 | 0.859182349 |
| CDC42SE1 | 0.845606945 | 1.012555691 |
| SCARF2 | 0.845579737 | 0.884490591 |
| HSPBP1 | 0.845480154 | 0.976103686 |
| ADAP2 | 0.845456988 | 0.899623159 |
| HNRNPC | 0.845392014 | 0.908872902 |
| CDSN | 0.845352906 | 0.942081814 |
| COL1A1 | 0.845347761 | 1.034831916 |
| SLC20A2 | 0.845343482 | 0.939651681 |
| CAST | 0.845328309 | 0.918122645 |
| PTPRR | 0.84526024 | 1.042829332 |
| USP35 | 0.845236304 | 0.912920211 |
| DLX5 | 0.845231427 | 0.922777417 |
| NIPAL2 | 0.845122707 | 0.88618874 |
| OR13C2 | 0.845090071 | 1.021240151 |
| SERPINF1 | 0.845077812 | 0.92926671 |
| TAF7 | 0.845055091 | 1.15055159 |
| ACADL | 0.84503565 | 1.040097205 |
| CEP290 | 0.84503565 | 1.040097205 |
| UBE3A | 0.844856527 | 1.06229721 |
| CREB3 | 0.844839276 | 0.908464966 |
| FCHO2 | 0.844771373 | 0.893884164 |
| GALNT16 | 0.844744321 | 1.018170019 |
| ADH1C | 0.844720776 | 0.720960415 |
| NEBL | 0.844639286 | 0.907845152 |
| OR5AY1_pseudogene | 0.844624971 | 1.021061158 |
| BTNL9 | 0.84460254 | 0.86193902 |
| FNDC4 | 0.844570823 | 0.856457691 |
| NSUN6 | 0.844510916 | 0.918590522 |
| ZNF83 | 0.844498945 | 0.916160389 |
| ACE2 | 0.844492164 | 0.904607398 |
| TRIM26 | 0.844480475 | 0.980532122 |
| VENTX | 0.844476022 | 0.835560956 |
| HAND2 | 0.844462777 | 0.757391657 |
| GSG1 | 0.844356191 | 0.989037342 |
| BCAN | 0.844347854 | 0.96882494 |
| CTRC | 0.844340854 | 0.860480208 |

| | | |
|---|---|---|
| DAPK3 | 0.844340854 | 1.012329656 |
| HEATR5B | 0.844338127 | 0.884568651 |
| LINGO4 | 0.844276132 | 0.945727015 |
| KIF17 | 0.844273127 | 0.872417983 |
| IGFBP1 | 0.844248944 | 0.987324426 |
| RTN4RL1 | 0.844043419 | 1.108941418 |
| PHF5A | 0.844035929 | 0.898767034 |
| TMEM107 | 0.843979253 | 0.958282706 |
| VPS25 | 0.843979253 | 0.889833941 |
| COL15A1 | 0.84394845 | 0.847893114 |
| TTC22 | 0.843937737 | 0.88254354 |
| ARL6IP6 | 0.843934222 | 0.814094775 |
| SLC25A23 | 0.84391088 | 0.95587281 |
| USP39 | 0.84391088 | 0.95587281 |
| XPO4 | 0.8439074 | 0.961973279 |
| TFEB | 0.843906969 | 1.012150668 |
| ODF3B | 0.843874958 | 0.956491949 |
| ERF | 0.843795298 | 0.753406879 |
| TEAD4 | 0.84379434 | 0.858242203 |
| POMT1 | 0.843745947 | 1.046577562 |
| TSEN2 | 0.843614928 | 0.943543154 |
| RGS9BP | 0.84361121 | 0.829080599 |
| FAM47E | 0.843606907 | 0.962315862 |
| CMAS | 0.843595266 | 0.890238963 |
| IFIT2 | 0.843582258 | 0.94210346 |
| CDRT1 | 0.843539078 | 0.886947585 |
| YTHDF1 | 0.843521271 | 0.980815348 |
| UHRF1BP1 | 0.843499387 | 1.089414183 |
| C14orf2 | 0.843440031 | 0.920178143 |
| FAHD1 | 0.843426984 | 0.899415964 |
| KDM2B | 0.843416612 | 0.741077223 |
| PTDSS1 | 0.843388704 | 1.007138222 |
| BPIFB4 | 0.843353762 | 0.938677629 |
| OR2A25 | 0.84328548 | 0.912983899 |
| TTC23 | 0.843258704 | 0.897934386 |
| FADS1 | 0.843189399 | 1.07073329 |
| AP2S1 | 0.843145836 | 0.81740322 |
| MCAT | 0.843115257 | 0.876063183 |
| GSTM2 | 0.84299915 | 0.918883842 |
| GMPR | 0.842986633 | 0.971648441 |
| PHF3 | 0.842961199 | 0.8935756 |
| ARFGAP2 | 0.842928849 | 0.89311408 |
| MYCL | 0.842920943 | 0.868267359 |
| DPPA5 | 0.842910477 | 0.959497772 |

| | | |
|---|---|---|
| TMEM45B | 0.842725666 | 0.876468206 |
| LCE3D | 0.842672994 | 0.869177805 |
| C1orf127 | 0.842636368 | 0.9181855 |
| NRCAM | 0.842630168 | 1.064748201 |
| SLC30A9 | 0.842587984 | 0.946899623 |
| ZNF330 | 0.842505293 | 0.970149254 |
| GMPS | 0.842472188 | 0.947752126 |
| YDJC | 0.842449111 | 0.832819459 |
| OR6B1 | 0.842424054 | 0.993148338 |
| ACP6 | 0.842349456 | 0.89422453 |
| C9orf37 | 0.842283776 | 1.000685166 |
| RAPGEF3 | 0.842249658 | 0.903048989 |
| NACA2 | 0.842242753 | 0.86472256 |
| SH2B2 | 0.842216621 | 0.962737951 |
| T | 0.84219273 | 0.908869988 |
| TWIST1 | 0.842165936 | 0.855002025 |
| NUP85 | 0.842145763 | 1.008221994 |
| C12orf71 | 0.842133484 | 0.993490922 |
| KIAA1958 | 0.842110965 | 0.925316889 |
| MECOM | 0.842110965 | 0.925316889 |
| GALNT11 | 0.842060995 | 1.021414666 |
| GPR88 | 0.842060995 | 1.021414666 |
| FAM206A | 0.842051065 | 0.791413528 |
| PLSCR1 | 0.84202962 | 0.788983394 |
| GFOD1 | 0.841942665 | 0.779262859 |
| GNAO1 | 0.841928775 | 0.381637547 |
| TRAPPC2 | 0.841906211 | 1.052757794 |
| SLC10A2 | 0.841824727 | 1.034393251 |
| C1orf112 | 0.841787654 | 0.857837181 |
| ZNF570 | 0.841760858 | 1.045563549 |
| HDX | 0.841692956 | 0.987849332 |
| HIST2H4A | 0.841620366 | 0.931551235 |
| INTS7 | 0.84153173 | 1.012555691 |
| GLP1R | 0.841501691 | 0.96495782 |
| ATOH8 | 0.841463899 | 0.983213429 |
| BRCA1 | 0.841454849 | 0.996755354 |
| CCDC178 | 0.841418544 | 0.9777321 |
| OR4B1 | 0.841416188 | 0.929770469 |
| SMCR8 | 0.841360266 | 1.034831916 |
| CYP27B1 | 0.841330706 | 1.092147956 |
| C9orf172 | 0.841326339 | 0.966769442 |
| SLCO2B1 | 0.841314674 | 0.939246659 |
| PODXL | 0.841243014 | 1.003768414 |
| DHH | 0.841216935 | 0.920830629 |

| | | |
|---|---|---|
| NFATC2 | 0.841212407 | 0.83192732 |
| MLH1 | 0.841128602 | 0.895522388 |
| C3orf27 | 0.841105208 | 0.696638315 |
| OR10Q1 | 0.841086243 | 0.939362795 |
| SKIV2L2 | 0.841085654 | 0.996574169 |
| RTP1 | 0.84107632 | 0.91538198 |
| FAM19A2 | 0.841054879 | 0.951802349 |
| XRCC3 | 0.841038579 | 0.863507493 |
| SLC6A11 | 0.840981526 | 0.814499797 |
| GPR116 | 0.840936213 | 0.965606749 |
| MGRN1 | 0.840880315 | 1.029201817 |
| TECR | 0.840814023 | 0.921205892 |
| MYT1 | 0.840793135 | 1.003893576 |
| NLRP1 | 0.840781942 | 0.920210612 |
| LYPD6 | 0.840718216 | 0.954637505 |
| OLFM2 | 0.84061679 | 0.890030832 |
| CD300LB | 0.840596978 | 0.981773998 |
| DNAH6 | 0.84056695 | 0.766358342 |
| HAPLN1 | 0.840549053 | 0.969510106 |
| MAT2A | 0.840523115 | 1.013770757 |
| RDX | 0.840491509 | 1.04898938 |
| ALKBH2 | 0.840459067 | 1.023361454 |
| ARAP3 | 0.840419966 | 1.019527235 |
| MED23 | 0.840382918 | 0.934458144 |
| DGCR6L | 0.840379051 | 0.91983556 |
| BEST1 | 0.840361736 | 0.955042527 |
| DYNC1H1 | 0.840360609 | 0.998053212 |
| PRDM10 | 0.840287027 | 0.94210346 |
| PBRM1 | 0.840226796 | 0.901174565 |
| CCBL1 | 0.84013602 | 0.930741191 |
| CEP70 | 0.840102145 | 1.004455245 |
| KANK2 | 0.839976883 | 1.073655361 |
| GSTP1 | 0.839958443 | 0.874443094 |
| FAM222B | 0.839926453 | 0.985014176 |
| PRKD3 | 0.839902068 | 0.833225178 |
| TIAM1 | 0.839883714 | 0.754151478 |
| C10orf53 | 0.839876975 | 0.922233642 |
| PDHB | 0.839780509 | 0.633859862 |
| OTUB2 | 0.839753552 | 0.920520726 |
| SLCO1B7 | 0.839752848 | 0.855002025 |
| OGG1 | 0.839746532 | 0.928716079 |
| CNIH2 | 0.83973783 | 0.770811922 |
| TNNI2 | 0.839685558 | 0.941596366 |
| SFMBT2 | 0.839682416 | 0.933538883 |

| | | |
|---|---|---|
| APTX | 0.839660367 | 0.878001298 |
| BPI | 0.839643836 | 0.771567436 |
| AGRP | 0.839637242 | 1.007536828 |
| FAM111A | 0.839587649 | 1.059133252 |
| CST7 | 0.83957294 | 0.948085659 |
| SMN2 | 0.839547439 | 1.076053443 |
| POPDC3 | 0.839519075 | 0.889345666 |
| ZMYND11 | 0.839421136 | 0.890979883 |
| ERI1 | 0.839401973 | 0.752126367 |
| NBN | 0.839389085 | 1.037262049 |
| CD24 | 0.839362666 | 0.970880439 |
| ALDH3B2 | 0.839217386 | 0.935756003 |
| CFP | 0.839148494 | 0.907249899 |
| CHL1 | 0.839127147 | 0.948955122 |
| UQCR10 | 0.83909246 | 0.936816525 |
| STK38L | 0.839074967 | 0.910447761 |
| TAF5L | 0.839073103 | 0.831105711 |
| KATNB1 | 0.839066568 | 0.536485098 |
| C11orf31 | 0.839010339 | 1.021240151 |
| RGL1 | 0.838960219 | 0.890238963 |
| AHI1 | 0.838953385 | 1.015758822 |
| EDARADD | 0.838946905 | 0.960260363 |
| AMBN | 0.838940743 | 0.914011648 |
| CHMP5 | 0.838935717 | 0.922235723 |
| DUSP3 | 0.838928349 | 0.987021415 |
| TRIM35 | 0.838852282 | 0.923426347 |
| IFI27L2 | 0.8388093 | 0.893799246 |
| TERF2 | 0.83870613 | 0.956834532 |
| BIRC3 | 0.838667878 | 1.038286827 |
| KDM6A | 0.838667481 | 0.864337102 |
| SIN3B | 0.838612702 | 0.886605002 |
| LHFPL5 | 0.838594876 | 0.925070879 |
| GSG2 | 0.838578887 | 0.974691759 |
| HNRNPD | 0.838578677 | 0.890643985 |
| IMP3 | 0.838572921 | 0.989064399 |
| RCSD1 | 0.838556977 | 0.954637505 |
| VPS18 | 0.83852299 | 1.083062946 |
| KLC1 | 0.8385122 | 0.917780478 |
| DOK6 | 0.838503541 | 0.949777238 |
| CRYGS | 0.838469433 | 1.058924289 |
| CNNM4 | 0.838457652 | 0.944158959 |
| OR4K15 | 0.838405909 | 1.044193217 |
| IFNA13 | 0.838368845 | 0.823912299 |
| NXPH2 | 0.838350586 | 0.883179171 |

| | | |
|---|---|---|
| VEGFA | 0.838283173 | 0.962332928 |
| PSD3 | 0.838228769 | 0.957472661 |
| PMAIP1 | 0.838205731 | 1.068516615 |
| ANXA7 | 0.83809211 | 0.898767034 |
| CYLD | 0.838084371 | 1.007138222 |
| CMTM2 | 0.838022614 | 0.78290806 |
| HSFY2 | 0.838014479 | 0.983556012 |
| ATP6V1G1 | 0.837994801 | 0.999594978 |
| PEBP1 | 0.837953998 | 0.778047793 |
| MTNR1A | 0.837880539 | 0.91174562 |
| AIMP1 | 0.83775558 | 0.920863309 |
| TPD52 | 0.83774747 | 1.002398082 |
| B4GALNT3 | 0.837742953 | 0.916160389 |
| DHFR | 0.837742953 | 0.916160389 |
| SERHL2 | 0.837704391 | 0.852166869 |
| PNLIPRP1 | 0.837692513 | 1.071382219 |
| KLF5 | 0.837524929 | 0.928716079 |
| LIX1L | 0.837494823 | 0.926285946 |
| OR6K6 | 0.837410774 | 0.902706406 |
| GPR84 | 0.837403723 | 0.829331603 |
| DHX9 | 0.837385283 | 0.905905256 |
| SLC4A3 | 0.837372771 | 0.857432159 |
| FBXO3 | 0.837350378 | 1.09733939 |
| GP9 | 0.837342869 | 0.943701904 |
| SCAMP4 | 0.83733334 | 0.92874272 |
| WFDC10B | 0.837318569 | 0.887975334 |
| FAM188A | 0.837296231 | 0.941760877 |
| GPR63 | 0.837293739 | 0.835820896 |
| FABP3 | 0.83728065 | 0.950681376 |
| ESD | 0.83725571 | 0.965978129 |
| OR4A15 | 0.837225639 | 0.999314834 |
| SNAP91 | 0.837204797 | 0.895512162 |
| SEPT12 | 0.837201612 | 0.990684488 |
| UBE2I | 0.837185402 | 1.072031149 |
| INTS3 | 0.837145323 | 0.869987849 |
| HIST1H2BO | 0.837093063 | 0.903048989 |
| HOXD10 | 0.837056025 | 0.892264075 |
| KIF12 | 0.837005773 | 1.00843608 |
| IKBKAP | 0.836998693 | 0.970149254 |
| SERPINI1 | 0.836944936 | 1.129785853 |
| IRF2BP2 | 0.83689864 | 0.981157931 |
| PLA2G4E | 0.836830753 | 1.021414666 |
| PIKFYVE | 0.836816923 | 0.983127839 |
| WNT7B | 0.836806661 | 0.890373416 |

| | | |
|---|---|---|
| ABHD5 | 0.836749006 | 0.829980532 |
| RAD54L2 | 0.836671121 | 0.724989874 |
| DENND1A | 0.836659914 | 1.030781693 |
| CDC42BPA | 0.83664001 | 0.996106424 |
| APBB1IP | 0.836632808 | 0.968482357 |
| OR5T2 | 0.836606529 | 0.981500514 |
| KIRREL2 | 0.836560993 | 0.940733128 |
| ST8SIA5 | 0.836553159 | 1.002595717 |
| GABPB1 | 0.836507959 | 1.045767517 |
| CSHL1 | 0.836458412 | 0.903734156 |
| YBEY | 0.836422119 | 0.760877013 |
| RTCB | 0.836369045 | 0.816524909 |
| DUX5 | 0.83629778 | 0.920210612 |
| ARAP2 | 0.836227279 | 0.888213852 |
| CHTF18 | 0.836171058 | 0.913326482 |
| ALDH1A3 | 0.836058514 | 0.903199676 |
| NLRP11 | 0.836051729 | 0.958468527 |
| ALPK1 | 0.836047612 | 0.875405581 |
| ERN2 | 0.835995584 | 0.837118754 |
| MBNL2 | 0.835930807 | 0.999594978 |
| USP13 | 0.835848802 | 1.016223232 |
| RPRD1A | 0.83579304 | 1.001027749 |
| PSME3 | 0.835776094 | 0.943473792 |
| LEAP2 | 0.835719507 | 0.906034832 |
| REV3L | 0.835569561 | 0.952628164 |
| ZIC3 | 0.835548093 | 1.022276225 |
| TOMM22 | 0.835544245 | 0.914011648 |
| CYBRD1 | 0.835486003 | 0.959117456 |
| OR5D16 | 0.835484844 | 0.979102432 |
| ZMYND10 | 0.835484255 | 0.930798219 |
| TUBA4A | 0.835471276 | 0.787768327 |
| ALKBH6 | 0.835335062 | 1.0818145 |
| YY1AP1 | 0.835304243 | 1.079384366 |
| RETN | 0.835248827 | 1.023980815 |
| POU2F1 | 0.835242244 | 0.978585334 |
| SGOL1 | 0.835241305 | 0.691332648 |
| SLC25A48 | 0.835199938 | 0.817745803 |
| ABCA10 | 0.835198032 | 0.89671932 |
| SNX14 | 0.835131067 | 0.934909215 |
| FKBPL | 0.835104516 | 0.899623159 |
| NDUFB1 | 0.835081385 | 0.9467878 |
| NUDT6 | 0.835042003 | 0.78817335 |
| RAD23A | 0.834983914 | 0.863724854 |
| IGSF1 | 0.834973616 | 1.011305242 |

| | | |
|---|---|---|
| TTLL11 | 0.834962995 | 0.8327258 |
| TMEM104 | 0.83489122 | 1.047792629 |
| HMGB4 | 0.834860503 | 0.753746456 |
| HIST1H3D | 0.834845254 | 1.048646797 |
| OTOGL | 0.834831467 | 0.80072904 |
| GNRHR | 0.834827088 | 0.921479559 |
| EXPH5 | 0.834813485 | 0.872417983 |
| ABCD4 | 0.834812517 | 0.993519644 |
| PLAC9 | 0.834740861 | 0.903734156 |
| CCDC91 | 0.834723728 | 0.842851357 |
| GPRASP2 | 0.834638709 | 0.909680032 |
| PBLD | 0.834578141 | 0.667476711 |
| EMP1 | 0.834526365 | 0.987667009 |
| KANSL1 | 0.834448652 | 0.85054678 |
| TENM2 | 0.83441436 | 0.865022268 |
| PRDM5 | 0.834385384 | 0.870392872 |
| TXLNGY | 0.834353781 | 0.963686194 |
| FTSJ3 | 0.834341983 | 0.77399757 |
| FOLH1B | 0.83431298 | 0.819597664 |
| FRG2B | 0.83431264 | 0.241863652 |
| DZANK1 | 0.834257679 | 1.04008222 |
| ITGA6 | 0.834230118 | 0.791413528 |
| SLC25A31 | 0.834181378 | 0.87084618 |
| MRPS11 | 0.834177548 | 1.043742406 |
| TTLL6 | 0.834132466 | 0.994329688 |
| CCDC54 | 0.834096044 | 0.991899554 |
| SLK | 0.834090213 | 1.082414017 |
| FOLR2 | 0.834082646 | 0.94491697 |
| KDSR | 0.834082646 | 0.94491697 |
| KIFC1 | 0.834043947 | 1.037637897 |
| RAB31 | 0.833993483 | 0.794289422 |
| HNRNPLL | 0.833966605 | 0.937626569 |
| SYT10 | 0.833938428 | 0.975334018 |
| NIF3L1 | 0.833918899 | 0.949297705 |
| PPCDC | 0.833891073 | 0.852044127 |
| ZCCHC5 | 0.833885461 | 0.854402193 |
| R3HCC1L | 0.833873726 | 0.841384036 |
| CDKAL1 | 0.833857278 | 0.927029805 |
| SUGCT | 0.83382819 | 0.943816375 |
| C4A | 0.833822481 | 0.98663926 |
| ASB17 | 0.83382044 | 0.807268008 |
| DMWD | 0.833817179 | 1.029462145 |
| MDC1 | 0.833751254 | 0.908530319 |
| TMEM185A | 0.833729071 | 0.923045768 |

| | | |
|---|---|---|
| RNF133 | 0.833652912 | 0.974042829 |
| CTSL | 0.833590511 | 1.083062946 |
| MFGE8 | 0.833573822 | 0.910585817 |
| DRD3 | 0.833565732 | 0.775470474 |
| DSC1 | 0.833540881 | 0.957177115 |
| RYBP | 0.833515926 | 0.987021415 |
| WISP2 | 0.833477426 | 0.84163629 |
| ELMOD2 | 0.833474981 | 0.819360065 |
| RASSF8 | 0.833391847 | 0.859052248 |
| DFFB | 0.833385326 | 1.034831916 |
| EIF6 | 0.833311565 | 1.029971648 |
| PFKFB3 | 0.833207296 | 0.910447761 |
| PRSS23 | 0.833164077 | 0.968202466 |
| CREBRF | 0.833161685 | 0.905447071 |
| KATNBL1 | 0.833157063 | 0.824625354 |
| PCDHA13 | 0.833068891 | 0.929770469 |
| KCNQ1 | 0.8330671 | 0.923426347 |
| COPG2 | 0.833031466 | 0.933538883 |
| ARSA | 0.832943624 | 0.963547995 |
| C1orf54 | 0.832942432 | 1.00445358 |
| BUB1 | 0.832904759 | 0.994159637 |
| SIRT7 | 0.832904759 | 0.994159637 |
| ZNF213 | 0.832884737 | 1.045362495 |
| SFXN5 | 0.832842028 | 1.000648929 |
| GPATCH11 | 0.832765231 | 0.889428919 |
| ISCA2 | 0.832754076 | 1.015795869 |
| CHCHD4 | 0.832646878 | 0.958204865 |
| MEGF11 | 0.832644205 | 1.038369305 |
| ZNF418 | 0.832606757 | 0.859862292 |
| ASAH1 | 0.832565648 | 0.899554475 |
| SRP72 | 0.832551921 | 0.859831278 |
| TAF6L | 0.832543607 | 0.975340688 |
| NAA30 | 0.832432286 | 1.058323208 |
| PELI3 | 0.832411197 | 0.974078574 |
| CXCL17 | 0.832317613 | 0.945529291 |
| HTR1F | 0.832312031 | 1.059052563 |
| IQCF2 | 0.83223632 | 0.902389631 |
| KRT19 | 0.832185717 | 1.010962658 |
| SH3BP5L | 0.832129906 | 0.835560956 |
| SRPR | 0.832099698 | 0.89509923 |
| JRK | 0.831940547 | 0.572798904 |
| EFNA4 | 0.831883868 | 0.802725503 |
| GSTCD | 0.831851365 | 0.962332928 |
| ELL | 0.831824599 | 0.968482357 |

| | | |
|---|---|---|
| BBS7 | 0.831810508 | 0.934909215 |
| SNX16 | 0.831726821 | 1.033915725 |
| KCNH5 | 0.831719222 | 0.944192083 |
| MICB | 0.831719222 | 0.944192083 |
| CHRNA9 | 0.831679048 | 0.822193381 |
| CALM3 | 0.831654104 | 1.014925373 |
| UBXN8 | 0.831633799 | 0.989874443 |
| COL6A5 | 0.831545469 | 0.965168084 |
| DOK2 | 0.831536883 | 0.931483385 |
| HOXA13 | 0.831533702 | 0.885783718 |
| PSG3 | 0.831502584 | 1.064063035 |
| PCDHAC1 | 0.831484098 | 0.864337102 |
| ENKD1 | 0.83146152 | 0.980153908 |
| CDK7 | 0.831444098 | 1.040882544 |
| KLHL26 | 0.83137864 | 0.56581612 |
| UBXN10 | 0.831354875 | 1.028776978 |
| TARSL2 | 0.831274586 | 0.907502569 |
| RP2 | 0.831162399 | 0.982989064 |
| ARHGAP29 | 0.831025981 | 0.84163629 |
| WFDC5 | 0.830948538 | 0.948955122 |
| UBALD2 | 0.830909481 | 0.988254354 |
| ACTN2 | 0.830848149 | 0.965978129 |
| NSRP1 | 0.830784015 | 0.943701904 |
| TMED1 | 0.830784015 | 0.754961523 |
| AQP8 | 0.830753349 | 0.866747671 |
| FBXO6 | 0.830735706 | 0.996755354 |
| SMCO4 | 0.83072304 | 1.015795869 |
| GPHN | 0.830689674 | 0.938841636 |
| C1GALT1 | 0.830577563 | 0.945489942 |
| TMOD2 | 0.830563361 | 0.919492977 |
| ATRNL1 | 0.830492225 | 0.844809866 |
| NMRK1 | 0.830481351 | 0.930798219 |
| OR56B1 | 0.830468211 | 0.996231586 |
| MVD | 0.830443064 | 1 |
| CLDN4 | 0.830406447 | 0.961522884 |
| DZIP1 | 0.830381443 | 0.966426859 |
| RSU1 | 0.830372775 | 0.904414743 |
| TGFB2 | 0.830302427 | 0.919400567 |
| C16orf85_pseudogene | 0.830271522 | 0.895512162 |
| KCNA7 | 0.830240861 | 0.99091499 |
| OR4X2 | 0.830201655 | 0.927372388 |
| KCNK3 | 0.830198959 | 0.9201817 |
| PIK3C2G | 0.830150223 | 1.003893576 |
| C17orf62 | 0.829935912 | 0.991899554 |

| | | |
|---|---|---|
| NUDT16L1 | 0.829924247 | 1.009315512 |
| APEH | 0.829916178 | 0.959117456 |
| TUBA1C | 0.829831943 | 0.753341434 |
| GATAD2B | 0.829785435 | 1.002025111 |
| C11orf1 | 0.8297699 | 0.970880439 |
| LRRC15 | 0.829697206 | 0.765897124 |
| CHD2 | 0.829644636 | 0.870392872 |
| GALNT6 | 0.829615957 | 0.940056703 |
| GDAP2 | 0.829540361 | 0.911270983 |
| PADI3 | 0.829453509 | 0.985014176 |
| AKT1 | 0.829375809 | 0.869565217 |
| ZFX | 0.829358403 | 0.962737951 |
| RBP7 | 0.829313902 | 0.966255678 |
| SLC31A1 | 0.829313627 | 0.873228028 |
| HEXIM2 | 0.829143283 | 0.831105711 |
| MPZL2 | 0.82911495 | 1.010962658 |
| ZNF771 | 0.829085007 | 0.828675577 |
| TSPAN32 | 0.829056328 | 0.948157149 |
| MANEAL | 0.829055055 | 0.706763872 |
| NMRK2 | 0.829024783 | 0.89311408 |
| CHMP7 | 0.829005031 | 0.945727015 |
| BDP1 | 0.829000243 | 0.928311057 |
| NOS1 | 0.829000243 | 0.928311057 |
| RAB5B | 0.82897355 | 0.841231268 |
| C12orf36 | 0.828922457 | 0.856217092 |
| CD300A | 0.828865302 | 0.853786958 |
| GOSR1 | 0.828780487 | 1.003240178 |
| ARHGEF16 | 0.828733986 | 0.945871874 |
| CLEC10A | 0.828656824 | 0.960945529 |
| RCCD1 | 0.828653595 | 1.030781693 |
| FOXO1 | 0.828638721 | 0.928716079 |
| LOC652759_pseudogene | 0.828612837 | 0.992861778 |
| TMEM258 | 0.828595611 | 1.032202809 |
| POLD4 | 0.82859425 | 1.010935601 |
| KIAA1751 | 0.828586096 | 0.916067146 |
| RAB3C | 0.828558751 | 0.90266061 |
| LPAL2 | 0.82848326 | 0.915639195 |
| PRPH2 | 0.828475769 | 1.018818949 |
| STYX | 0.828446215 | 0.922128488 |
| NBPF14 | 0.828430681 | 0.869177805 |
| THADA | 0.828347269 | 0.777663583 |
| DCAF15 | 0.82834369 | 0.965056526 |
| HMBOX1 | 0.828328087 | 1.031186715 |
| PROP1 | 0.828312958 | 0.963953017 |

| | | |
|---|---|---|
| PDGFRL | 0.828252014 | 0.851395198 |
| HMGN1 | 0.828242304 | 0.944106926 |
| GUCA2B | 0.828208123 | 0.95909275 |
| PRDM11 | 0.828176689 | 0.864373783 |
| VRK2 | 0.828176689 | 0.864373783 |
| NFATC2IP | 0.828105886 | 0.707573917 |
| ANAPC4 | 0.828071644 | 0.993510707 |
| CD3EAP | 0.82805841 | 0.919400567 |
| OVGP1 | 0.827997718 | 0.896820247 |
| SLIRP | 0.827952758 | 0.988009592 |
| PKIB | 0.827948406 | 1.019467878 |
| OR1L6 | 0.827936991 | 0.991778006 |
| KIF24 | 0.827932097 | 0.650565262 |
| TTC39A | 0.82787904 | 0.971223022 |
| GOLGA4 | 0.8278601 | 1.010620075 |
| TTC26 | 0.827844054 | 0.780072904 |
| C17orf64 | 0.827809058 | 0.82746051 |
| ARMC9 | 0.827804779 | 0.957067639 |
| TSC22D1 | 0.827777882 | 0.77764277 |
| ZDHHC21 | 0.827771429 | 0.890715999 |
| ST7 | 0.8277165 | 0.840016201 |
| FAM91A1 | 0.827696633 | 0.952207371 |
| BBS2 | 0.827659152 | 1.063377869 |
| OR5T1 | 0.827655256 | 1.027749229 |
| SH3BGR | 0.827654658 | 0.837586067 |
| CCDC59 | 0.827632143 | 1.061968408 |
| CYP39A1 | 0.827587995 | 0.85853342 |
| ST18 | 0.827505689 | 0.954779034 |
| IKBKB | 0.827460147 | 0.884490591 |
| AQPEP | 0.827421785 | 1.020116807 |
| GLYATL2 | 0.827421199 | 0.940056703 |
| UCK2 | 0.827395277 | 1.0266061 |
| FBXL14 | 0.827369098 | 1.033095393 |
| F8 | 0.827343248 | 1.039584685 |
| SIVA1 | 0.827322579 | 0.793033617 |
| POLD2 | 0.82731781 | 1.046073978 |
| RHBDL1 | 0.82731781 | 1.046073978 |
| ITM2A | 0.82730542 | 0.92874272 |
| SLC15A2 | 0.827295479 | 0.855407047 |
| MARK1 | 0.827281835 | 0.923426347 |
| GPD2 | 0.827252536 | 0.932766302 |
| CD79B | 0.827207658 | 0.678412313 |
| MLNR | 0.827198277 | 0.942894225 |
| PSG6 | 0.827195499 | 0.958890031 |

| | | |
|---|---|---|
| GALNT8 | 0.827191408 | 0.835560956 |
| PTGDR2 | 0.827171193 | 0.949383517 |
| HGD | 0.82710155 | 0.80072904 |
| CWF19L2 | 0.827054224 | 0.957177115 |
| OSBPL5 | 0.827053 | 0.914011648 |
| TRIM63 | 0.826991317 | 0.710577547 |
| VCX3B | 0.826969631 | 0.893456663 |
| GP6 | 0.826884697 | 0.808424463 |
| TMEM35 | 0.826876021 | 0.966769442 |
| SUN3 | 0.826827543 | 0.86836776 |
| ERAL1 | 0.826817915 | 0.80599433 |
| TGFA | 0.826769906 | 0.953751285 |
| C3orf36 | 0.826765294 | 0.865937627 |
| NENF | 0.826727751 | 1.080599433 |
| PID1 | 0.826708801 | 0.940866748 |
| USP9X | 0.826676759 | 0.930564568 |
| HAS2 | 0.826670749 | 0.908464966 |
| SRRD | 0.826651086 | 0.938436614 |
| VAPA | 0.826635926 | 0.998972251 |
| ADAL | 0.826615428 | 0.998379911 |
| FAM46A | 0.826553383 | 1.010935601 |
| SEP15 | 0.826391526 | 0.984241178 |
| MCM6 | 0.826350595 | 1.027903958 |
| BANP | 0.826346579 | 0.890373416 |
| RTEL1 | 0.826315607 | 0.866320571 |
| SCGN | 0.826300091 | 0.963686194 |
| CGNL1 | 0.826227416 | 0.981368975 |
| HOXA3 | 0.82619529 | 0.946899623 |
| EPC1 | 0.826170822 | 0.978938842 |
| MAP3K12 | 0.826141247 | 0.918234912 |
| ERC1 | 0.826132668 | 0.73673552 |
| CENPO | 0.826116825 | 0.931551235 |
| FMO3 | 0.82605697 | 0.734305387 |
| SLC12A4 | 0.82605697 | 0.779262859 |
| ACSM2A | 0.826041373 | 0.948955122 |
| PFKP | 0.826022866 | 0.957170668 |
| PGK2 | 0.826004096 | 0.963659961 |
| LCE2A | 0.826002057 | 1.016605913 |
| HOXC4 | 0.825985196 | 0.776832726 |
| TBL3 | 0.825908118 | 1.009249743 |
| CEACAM19 | 0.82583263 | 0.994329688 |
| SLCO1B1 | 0.825829115 | 0.979343864 |
| SLC28A2 | 0.825773097 | 0.84101233 |
| RTCA | 0.825762485 | 0.946942082 |

| | | |
|---|---|---|
| FRMD3 | 0.825655409 | 0.972053463 |
| FFAR2 | 0.825642944 | 0.886437378 |
| MORC2 | 0.825522506 | 0.834750911 |
| ASB14 | 0.825493857 | 0.944841012 |
| WDR92 | 0.825478492 | 1.015416238 |
| FUT1 | 0.825463315 | 0.957819598 |
| VWA8 | 0.825438811 | 0.992304577 |
| IL12B | 0.825404372 | 0.983776768 |
| ICOSLG | 0.825385019 | 0.829890644 |
| PYCARD | 0.825308547 | 1.029201817 |
| NOX5 | 0.825261623 | 0.789746918 |
| PLEKHM1P | 0.825245076 | 0.79623621 |
| CIT | 0.825228776 | 0.802725503 |
| STX5 | 0.825213045 | 0.925070879 |
| SLC9A2 | 0.825197082 | 0.815704088 |
| PUM1 | 0.82513722 | 0.957519699 |
| CHRM4 | 0.825052193 | 0.880597015 |
| MARC1 | 0.825019778 | 0.860267315 |
| IGSF5 | 0.824964636 | 0.856115108 |
| ST3GAL2 | 0.824939081 | 0.939000649 |
| NUDT13 | 0.8248964 | 0.912920211 |
| GK | 0.824849678 | 0.99091499 |
| SLC39A13 | 0.824838041 | 0.961630695 |
| HK2 | 0.824828685 | 1.003893576 |
| SULT1C3 | 0.82479839 | 1.023361454 |
| SCNN1D | 0.824778715 | 1.036340039 |
| MUC17 | 0.82476961 | 0.823005265 |
| PAK4 | 0.824759616 | 1.049318624 |
| HPCAL4 | 0.824706037 | 0.990684488 |
| KRT34 | 0.82470023 | 0.863102471 |
| SPATS1 | 0.824552998 | 0.950668037 |
| BIRC5 | 0.82454113 | 0.816353018 |
| GP1BA | 0.82452468 | 0.988352175 |
| PRAMEF6 | 0.82452468 | 0.988352175 |
| PEX7 | 0.824512515 | 0.758201701 |
| DHX35 | 0.824451591 | 0.86836776 |
| PDE10A | 0.824435839 | 0.887735237 |
| JRKL | 0.824393442 | 0.9201817 |
| WBSCR22 | 0.824369322 | 0.939649578 |
| FCRL1 | 0.824333613 | 0.906034832 |
| USP25 | 0.824282306 | 1.01752109 |
| KRT3 | 0.824232569 | 1.003768414 |
| KRT4 | 0.824121696 | 0.907845152 |
| PSAPL1 | 0.824115398 | 0.926687222 |

| | | |
|---|---|---|
| FAM135A | 0.82399481 | 0.811259619 |
| KRTAP4-4 | 0.823982279 | 0.823227133 |
| LMAN1L | 0.82396787 | 0.898595409 |
| TNFRSF17 | 0.823962161 | 0.932511134 |
| AIPL1 | 0.823808371 | 0.927319922 |
| GRM5 | 0.823804337 | 0.933809215 |
| GLRA2 | 0.823753626 | 1.024659312 |
| C10orf95 | 0.823749788 | 0.966788173 |
| HAGHL | 0.823659624 | 0.97691373 |
| GOLT1A | 0.823583723 | 0.946942082 |
| KANK4 | 0.823416537 | 0.886998785 |
| MYO1G | 0.823387953 | 1.001027749 |
| PKHD1 | 0.823349757 | 0.918122645 |
| ATXN2L | 0.823318223 | 0.977318753 |
| CAV3 | 0.82329586 | 1.029971648 |
| EGF | 0.823232273 | 0.953926022 |
| GPR37L1 | 0.823232273 | 0.804672291 |
| NEK11 | 0.823232273 | 0.863075925 |
| P2RY13 | 0.823232273 | 0.934458144 |
| WHSC1 | 0.823232273 | 0.726800779 |
| SLC35F6 | 0.823230683 | 0.972935937 |
| GULP1 | 0.823222608 | 0.786958283 |
| EQTN | 0.823193267 | 1.012555691 |
| EHD2 | 0.823166796 | 1.095580678 |
| LYSMD1 | 0.823149254 | 0.824625354 |
| NR2F6 | 0.823126229 | 0.837181045 |
| FAM46C | 0.82305094 | 0.74200081 |
| PDGFA | 0.82305094 | 0.927501013 |
| TOM1L2 | 0.823039522 | 0.914696814 |
| ZNF483 | 0.823006112 | 1.00526529 |
| ANKRD53 | 0.822955885 | 1.042932361 |
| FAM212B | 0.822955885 | 1.042932361 |
| DYNC2LI1 | 0.822879832 | 0.842446335 |
| KLHL5 | 0.822847681 | 0.879068174 |
| ABCC1 | 0.822792338 | 1.214146658 |
| M6PR | 0.822758645 | 1.048197651 |
| NLE1 | 0.822701453 | 1.006489293 |
| ROR2 | 0.82267264 | 0.954574951 |
| CMA1 | 0.822640452 | 0.90266061 |
| DDX27 | 0.822627445 | 0.883192732 |
| DSEL | 0.822594598 | 0.837767683 |
| FOSL1 | 0.822510793 | 0.740428293 |
| WDR33 | 0.822510442 | 0.868105516 |
| HSPA5 | 0.822458047 | 0.790198461 |

| | | |
|---|---|---|
| IDH1 | 0.822458047 | 0.790198461 |
| RBM4B | 0.82241434 | 0.750101256 |
| FAM75A1 | 0.822240889 | 0.745240988 |
| KRI1 | 0.822201838 | 0.964713943 |
| PDCD2L | 0.822192292 | 1.038882139 |
| PLOD3 | 0.822171374 | 1.007138222 |
| PDLIM1 | 0.822162594 | 0.973673552 |
| ATP10B | 0.82215857 | 0.858242203 |
| CCDC97 | 0.82211145 | 0.805589307 |
| ZDHHC17 | 0.822044616 | 0.931146213 |
| BCL2L1 | 0.822023731 | 0.815714864 |
| PSMC3IP | 0.822017516 | 0.993924666 |
| ZNF354C | 0.821988203 | 0.929085303 |
| TRIM64 | 0.82195791 | 0.838416613 |
| MFAP1 | 0.821938151 | 0.825840421 |
| SPATA22 | 0.821888383 | 0.976508708 |
| LRIF1 | 0.821871878 | 1.039287161 |
| SNRPB | 0.821791695 | 0.838643371 |
| THBS4 | 0.821752004 | 0.959575197 |
| KLHL20 | 0.821700974 | 0.780882949 |
| SELP | 0.821668952 | 1.054128126 |
| ALDH2 | 0.821652161 | 1.014276444 |
| FAM73A | 0.821615751 | 1.017010936 |
| THSD1 | 0.821615751 | 0.966788173 |
| E2F3 | 0.821532546 | 0.942894225 |
| HSPB8 | 0.821496735 | 0.923426347 |
| IGFBP3 | 0.821486173 | 0.971223022 |
| CLIC6 | 0.82145936 | 0.903958469 |
| BPY2B | 0.821362704 | 0.922233642 |
| CCDC121 | 0.821328856 | 0.695828271 |
| LCAT | 0.82131388 | 0.881733495 |
| SLC39A3 | 0.821253388 | 1.0798183 |
| CRYZ | 0.821153447 | 1.027903958 |
| UBQLN2 | 0.821116582 | 0.890373416 |
| SNX19 | 0.821112776 | 0.941418294 |
| CFL2 | 0.821101947 | 0.93764988 |
| HSPA1L | 0.821090507 | 0.886998785 |
| PIGG | 0.821075276 | 0.985014176 |
| LIG4 | 0.821071536 | 0.988968202 |
| PFKFB2 | 0.821028234 | 0.969500324 |
| PTGER1 | 0.821028234 | 0.969500324 |
| PPIP5K2 | 0.82100396 | 0.761441879 |
| ZNF718 | 0.820983023 | 0.736330498 |
| SCRG1 | 0.820969054 | 0.943473792 |

| | | |
|---|---|---|
| SORCS1 | 0.820935949 | 0.930564568 |
| CYP4A11 | 0.820826433 | 0.927501013 |
| FKBP5 | 0.820726437 | 0.972863507 |
| BORA | 0.820724807 | 1.045767517 |
| UGT2B17 | 0.820714772 | 0.960307817 |
| CADPS2 | 0.820708207 | 0.955121617 |
| MKL1 | 0.820677942 | 0.922640745 |
| SMEK1 | 0.820665055 | 0.910085055 |
| KRTAP5-5 | 0.820510295 | 0.855087359 |
| LMAN1 | 0.820498429 | 0.953017416 |
| CYLC1 | 0.820483426 | 1.01781432 |
| LRRC8A | 0.820481517 | 0.880113406 |
| ENTPD4 | 0.820479055 | 0.970149254 |
| PLA2G3 | 0.820479055 | 0.970149254 |
| SPANXN2 | 0.820392321 | 1.033616849 |
| GTPBP4 | 0.820384935 | 0.865127582 |
| PPIL6 | 0.820336965 | 1.107073329 |
| SDR16C5 | 0.820335268 | 0.779667882 |
| GFRA2 | 0.820287779 | 0.961288112 |
| NEU4 | 0.820281644 | 0.905256327 |
| TMEM187 | 0.820245715 | 0.777237748 |
| A2ML1 | 0.820205574 | 0.99108951 |
| DNAJC12 | 0.82013939 | 0.999314834 |
| GFRA1 | 0.820098146 | 1.022712524 |
| FAM174A | 0.820036504 | 0.842851357 |
| LARP1B | 0.819936457 | 0.792628595 |
| JUND | 0.819935286 | 0.910895099 |
| RNF166 | 0.819844508 | 0.788449059 |
| CPT1C | 0.819792644 | 0.931862427 |
| IFNA1 | 0.819628298 | 0.9109284 |
| RAD51C | 0.819576667 | 0.94653706 |
| NELFE | 0.819575648 | 0.899623159 |
| POLR2E | 0.819391754 | 0.778047793 |
| EHF | 0.81935699 | 0.96495782 |
| LBR | 0.819330646 | 1.095392602 |
| CACNA1S | 0.81919399 | 0.926022064 |
| TCTEX1D4 | 0.819163236 | 0.841231268 |
| OPTN | 0.819130074 | 0.994861254 |
| USP21 | 0.819107303 | 0.906554186 |
| CEP131 | 0.819038178 | 1.032806804 |
| GCNT4 | 0.818995134 | 0.939651681 |
| CAMKMT | 0.818987645 | 0.962332928 |
| UGT1A8 | 0.818893757 | 0.985074627 |
| LEFTY2 | 0.818889641 | 0.861129137 |

| | | |
|---|---|---|
| FOXL1 | 0.818856711 | 0.854639844 |
| LAMB2 | 0.818856016 | 0.844066424 |
| TSPYL2 | 0.818848112 | 1.052072628 |
| POGLUT1 | 0.818634273 | 0.781287971 |
| MANBA | 0.818611059 | 0.809214796 |
| MED16 | 0.818609007 | 1.018226002 |
| TLX1NB | 0.81856171 | 0.914011648 |
| DRD4 | 0.818554832 | 0.913692408 |
| MCM4 | 0.818479753 | 1.011680727 |
| EID3 | 0.818469024 | 0.867152693 |
| PCYT2 | 0.818452998 | 0.89422453 |
| DDX50 | 0.818449129 | 1.005191434 |
| SRI | 0.818441459 | 0.91983556 |
| MAPK13 | 0.818418093 | 0.998702141 |
| PARP2 | 0.818418093 | 0.887735237 |
| TRIM6-TRIM34 | 0.818418093 | 0.776768332 |
| ETFA | 0.818354622 | 0.985723556 |
| NOP16 | 0.818332263 | 0.978759849 |
| HEYL | 0.818327461 | 0.728635075 |
| POLR2I | 0.818305506 | 0.784123127 |
| RNF144A | 0.818256411 | 0.966255678 |
| EPHA3 | 0.818242992 | 1.07073329 |
| GYLTL1B | 0.818204218 | 1.168721609 |
| TRIM6 | 0.818119013 | 0.940298507 |
| RFX4 | 0.818112194 | 0.965741692 |
| PREB | 0.818056765 | 0.766707169 |
| GLP2R | 0.818010923 | 0.920830629 |
| CD72 | 0.817979132 | 0.786958283 |
| STK17A | 0.817921108 | 1.005840363 |
| MAP2K5 | 0.817816311 | 0.986372485 |
| MRPS26 | 0.817807759 | 0.305584104 |
| ANKRD54 | 0.817674459 | 0.658566221 |
| PMS2L5 | 0.817660412 | 0.803014731 |
| MUSK | 0.817645819 | 1.051914341 |
| PP8961_pseudogene | 0.817643647 | 0.869475848 |
| FIZ1 | 0.817624792 | 0.767112191 |
| TMEM217 | 0.817603639 | 0.886605002 |
| NAT16 | 0.817580803 | 0.973963686 |
| TMEM127 | 0.817558417 | 0.852976914 |
| OPN1SW | 0.81755477 | 0.940947437 |
| PIP5K1B | 0.817504552 | 1.025957171 |
| CT45-6 | 0.817474977 | 0.916160389 |
| NANOG | 0.817434907 | 0.921479559 |
| OR4A47 | 0.817385745 | 1.010277492 |

| | | |
|---|---|---|
| PRSS48 | 0.817239142 | 0.980532122 |
| AQP2 | 0.817117798 | 0.961064244 |
| PRR18 | 0.817011986 | 1.100034258 |
| OR10J3 | 0.816993591 | 0.786570743 |
| ACE | 0.816948047 | 0.935107073 |
| MAPKAP1 | 0.816948047 | 0.935107073 |
| NMNAT3 | 0.81687338 | 1.092147956 |
| PNLIP | 0.816850659 | 0.837118754 |
| NMNAT1 | 0.816806591 | 0.982584042 |
| SYT3 | 0.816767224 | 0.961630695 |
| OSGIN1 | 0.816744393 | 0.970028352 |
| RAB9B | 0.816709652 | 0.990279465 |
| CCL3 | 0.816709001 | 1.092840014 |
| TMEM161A | 0.816671345 | 0.861482382 |
| SPDYC | 0.816615973 | 0.891859052 |
| CAMKK1 | 0.816611098 | 0.968202466 |
| LTA4H | 0.816598296 | 0.795868773 |
| PLEKHJ1 | 0.816587118 | 0.918122645 |
| LY6E | 0.816568541 | 0.935251799 |
| UBE2L6 | 0.816566397 | 0.961713173 |
| MAP3K15 | 0.816521119 | 0.955223881 |
| CEBPB | 0.816475265 | 0.948734588 |
| TLR9 | 0.816407271 | 0.907249899 |
| PAX4 | 0.816385121 | 1.014276444 |
| ERBB3 | 0.816372032 | 0.77871512 |
| NNMT | 0.816363055 | 0.937626569 |
| RAP2A | 0.816333752 | 0.92926671 |
| HOXB6 | 0.816315069 | 0.778452815 |
| CHRNB4 | 0.816271844 | 0.844256976 |
| IL20RA | 0.816265046 | 0.945321993 |
| RPS6KA2 | 0.816251181 | 0.994808566 |
| CENPA | 0.81621487 | 0.98582422 |
| JDP2 | 0.816188756 | 0.907159986 |
| RABIF | 0.816107335 | 0.930336168 |
| NPTX2 | 0.816059986 | 1.019527235 |
| TRAPPC9 | 0.816023603 | 1.032887975 |
| GNA13 | 0.81602201 | 1.011340624 |
| GPHB5 | 0.816010191 | 0.922576225 |
| ERCC6L2 | 0.816002262 | 0.939705379 |
| MICAL1 | 0.815936991 | 0.73835561 |
| GABRG2 | 0.815910034 | 1.021414666 |
| TGFB1I1 | 0.81584861 | 0.819360065 |
| LGR5 | 0.815815774 | 0.864373783 |
| RAB25 | 0.815815774 | 1.00843608 |

| | | |
|---|---|---|
| ZMAT3 | 0.815796702 | 0.766302147 |
| HIST1H3A | 0.815756004 | 1.016101405 |
| WBSCR16 | 0.815721005 | 1.095180235 |
| TRIM15 | 0.815719044 | 0.995457495 |
| NINJ1 | 0.815680065 | 0.867762933 |
| FAM108A2_pseudogene | 0.81567903 | 0.908059943 |
| ATP1B2 | 0.815589579 | 0.90562981 |
| ZBTB6 | 0.815566248 | 0.946132037 |
| ACIN1 | 0.81554954 | 0.903958469 |
| NUP153 | 0.815517928 | 0.969500324 |
| IL18BP | 0.815366251 | 0.902363823 |
| ARHGEF6 | 0.815306555 | 0.938841636 |
| ZFYVE28 | 0.815305446 | 0.929085303 |
| QRICH2 | 0.815274056 | 1.070473876 |
| TDRD10 | 0.815261728 | 0.754151478 |
| FAIM2 | 0.815229461 | 0.904419322 |
| TRAK2 | 0.815195139 | 0.930564568 |
| NDUFAF2 | 0.815136313 | 0.883353584 |
| ERCC6L2 | 0.815118997 | 0.957862282 |
| FAM46D | 0.815042838 | 0.931551235 |
| P2RX2 | 0.814964552 | 0.904607398 |
| ALYREF | 0.814962709 | 1.030376671 |
| FOSL2 | 0.814944371 | 0.83799109 |
| DLL3 | 0.814920767 | 1 |
| SMARCB1 | 0.814870355 | 0.957067639 |
| KIF25 | 0.814866636 | 0.936816525 |
| SEL1L | 0.814864751 | 0.926690968 |
| LEF1 | 0.814860413 | 0.957170668 |
| GSG1L | 0.814801382 | 1.025516403 |
| CAPN9 | 0.814786322 | 1.075275795 |
| SCARB1 | 0.81465691 | 0.996755354 |
| UROD | 0.814650191 | 0.840826245 |
| FAM69B | 0.814625078 | 0.790198461 |
| AGAP3 | 0.814561301 | 1.028351559 |
| BPIFA2 | 0.814517512 | 0.941418294 |
| RPAIN | 0.814485247 | 1.036046983 |
| ARHGEF3 | 0.814424832 | 0.934791414 |
| RAP2C | 0.814424832 | 0.934791414 |
| PDS5B | 0.814415203 | 0.750942103 |
| ITPKC | 0.814413727 | 1.029850746 |
| UBA1 | 0.814401461 | 1.08890331 |
| PRRC2C | 0.81439291 | 1.078794108 |
| RGS4 | 0.814384793 | 1.003240178 |
| CALM2 | 0.814380304 | 0.905256327 |

| | | |
|---|---|---|
| SLC5A7 | 0.814368367 | 0.96430889 |
| GTSF1L | 0.814277462 | 0.970433374 |
| TEF | 0.814240683 | 1.069435432 |
| PPAPDC1B | 0.814169418 | 0.661401377 |
| RBM44 | 0.814153066 | 0.927501013 |
| EXOC3L1 | 0.814103501 | 1.054272985 |
| IL6ST | 0.814067311 | 0.99091499 |
| FDFT1 | 0.814050625 | 0.914945322 |
| SERAC1 | 0.814014303 | 0.936279548 |
| SPPL3 | 0.813995443 | 0.925373134 |
| DDX28 | 0.813913087 | 0.803564196 |
| DCAF4L1 | 0.813837186 | 0.958282706 |
| REEP1 | 0.813837186 | 0.821385176 |
| CD14 | 0.813804041 | 1.076573653 |
| OR52A5 | 0.813791154 | 1.028091812 |
| RAPGEF4 | 0.813747792 | 1.037684138 |
| ZNF429 | 0.813747735 | 0.955852572 |
| PLA2G2F | 0.813733879 | 0.945727015 |
| OR5P3 | 0.813730303 | 0.909900651 |
| TTC30A | 0.813725831 | 0.87727825 |
| PYY | 0.813701712 | 0.879410757 |
| DDX49 | 0.813689036 | 1.063595068 |
| TPO | 0.813626964 | 0.945489942 |
| SOCS3 | 0.813622518 | 0.833874108 |
| RNF123 | 0.813534515 | 0.99156392 |
| LINC00336 | 0.813491702 | 0.984583762 |
| RASSF7 | 0.813455644 | 0.831915755 |
| MAZ | 0.81343194 | 1.090201168 |
| DMPK | 0.813340314 | 0.972096042 |
| SPEN | 0.813330586 | 0.722964763 |
| PRB4 | 0.813329041 | 0.996231586 |
| CLDN6 | 0.813292159 | 0.925659472 |
| BMP4 | 0.812998711 | 0.888213852 |
| RASSF6 | 0.812933253 | 1.134467396 |
| EED | 0.812919066 | 0.880597015 |
| GM2A | 0.812866873 | 0.979234263 |
| PNKP | 0.812855431 | 0.926670993 |
| CTLA4 | 0.812780387 | 0.933881466 |
| ABHD1 | 0.812742176 | 0.825030377 |
| NSG1 | 0.812720488 | 0.903391572 |
| VRTN | 0.812655374 | 0.898744431 |
| CEACAM21 | 0.812638309 | 0.926690968 |
| SDC4 | 0.812636303 | 0.909215485 |
| CEP112 | 0.812625115 | 0.614418793 |

| | | |
|---|---|---|
| VCX2 | 0.812589815 | 0.905447071 |
| FAM83C | 0.812555888 | 0.962332928 |
| CRADD | 0.812459731 | 0.907502569 |
| KIF13A | 0.812418859 | 0.817739976 |
| PAK6 | 0.812273076 | 0.828682674 |
| TRNP1 | 0.812255521 | 1.001370332 |
| EFCAB1 | 0.812240262 | 0.795058728 |
| CD177 | 0.812230329 | 0.850951802 |
| AMTN | 0.812122845 | 0.904414743 |
| RANBP6 | 0.812081577 | 0.818088386 |
| CCL16 | 0.812062819 | 0.972935937 |
| MNAT1 | 0.812031869 | 0.953926022 |
| SIPA1 | 0.812016686 | 0.818144998 |
| LARS2 | 0.812014088 | 0.790198461 |
| TAB1 | 0.811997046 | 1.046073978 |
| ZNF721 | 0.811899982 | 0.945321993 |
| ZDHHC5 | 0.811830879 | 0.954093868 |
| PLG | 0.811790909 | 0.980532122 |
| VAC14 | 0.811748144 | 0.795477903 |
| CCDC114 | 0.811708243 | 0.940461725 |
| DEFB105A | 0.811708243 | 0.940461725 |
| SCN2B | 0.811637458 | 0.967553537 |
| IQCF3 | 0.811581351 | 0.882138518 |
| TBC1D25 | 0.811507539 | 0.770352369 |
| IFI27 | 0.811468085 | 0.983213429 |
| LGI4 | 0.811395227 | 0.77286742 |
| MYRIP | 0.811395227 | 0.966084275 |
| DDX10 | 0.811317797 | 0.941596366 |
| RHAG | 0.811245503 | 0.956257594 |
| CHRNB2 | 0.811174308 | 0.974691759 |
| PEX5 | 0.811161465 | 1.1064244 |
| SPRY3 | 0.81115844 | 1.104145255 |
| WDR26 | 0.811151417 | 0.891743748 |
| C1orf95 | 0.811149328 | 0.953827461 |
| C4orf27 | 0.811149328 | 0.699473471 |
| DAXX | 0.811093466 | 0.968202466 |
| XPNPEP1 | 0.811040121 | 1.007787151 |
| IGFLR1 | 0.810955449 | 0.948967193 |
| YIF1B | 0.8109311 | 0.903199676 |
| DEFB105B | 0.810929037 | 0.909215485 |
| NUP37 | 0.810888349 | 0.960945529 |
| ZNF30 | 0.810868458 | 1.003083248 |
| ABCG5 | 0.810799984 | 0.988319273 |
| SIRT6 | 0.810799984 | 0.988319273 |

| | | |
|---|---|---|
| FAM150B | 0.81079784 | 0.972053463 |
| ROPN1B | 0.810785496 | 0.920863309 |
| OR4C45 | 0.810749294 | 0.907502569 |
| OPN4 | 0.810672548 | 0.935756003 |
| NSF | 0.810573958 | 0.867962738 |
| SGCG | 0.810552333 | 1.10688592 |
| RWDD2B | 0.810541821 | 0.999657417 |
| PKIA | 0.810468962 | 0.837118754 |
| HDAC3 | 0.810464682 | 0.962362103 |
| GOLGA1 | 0.810411918 | 0.962332928 |
| ZNF331 | 0.810387633 | 0.904076739 |
| CD69 | 0.810365256 | 1.005670312 |
| PGAM1 | 0.810243644 | 0.863724854 |
| CDK5R2 | 0.810145268 | 0.857235561 |
| SRGN | 0.810138725 | 0.858242203 |
| FAM129B | 0.810117331 | 0.955042527 |
| TPPP3 | 0.810057883 | 1.059538275 |
| SULT1C4 | 0.810010662 | 0.934791414 |
| KDM2A | 0.809992064 | 0.685918235 |
| MANEA | 0.809974636 | 0.887290168 |
| ALOX15B | 0.809926288 | 0.923426347 |
| LTB4R | 0.809918549 | 0.963011032 |
| PAAF1 | 0.809911387 | 1.002595717 |
| CEP152 | 0.809878421 | 0.878898339 |
| REXO4 | 0.809852594 | 0.922235723 |
| FBXL7 | 0.809832192 | 0.916937054 |
| TLX1 | 0.809828158 | 0.956521739 |
| UGGT1 | 0.809790142 | 0.754151478 |
| NDUFS1 | 0.809643837 | 0.943543154 |
| P2RX7 | 0.809643837 | 0.943543154 |
| LRRC56 | 0.809581542 | 1.07290401 |
| S100A14 | 0.809448908 | 0.953066119 |
| LEPROT | 0.809408498 | 0.981368975 |
| CCDC74B | 0.809352871 | 0.841231268 |
| OR8U1 | 0.809346962 | 0.89756766 |
| KRTAP9-3 | 0.809344334 | 0.907159986 |
| ZBTB8A | 0.809274784 | 0.951353203 |
| LRIT1 | 0.809269618 | 0.980130182 |
| GPR148 | 0.809226376 | 1.029850746 |
| HTR1E | 0.809183897 | 0.950681376 |
| EFCAB7 | 0.809099934 | 0.834532374 |
| CCT5 | 0.809076419 | 0.912920211 |
| PDIA5 | 0.809073501 | 1.056456846 |
| SCNN1B | 0.809047652 | 1.016872161 |

| | | |
|---|---|---|
| RASAL1 | 0.809038596 | 1.129136924 |
| AP5M1 | 0.808992097 | 1.028434395 |
| KMT2D | 0.808989449 | 0.93770279 |
| RNF180 | 0.808956602 | 0.785853342 |
| PTER | 0.808941122 | 0.892669097 |
| DEFB112 | 0.80893913 | 1.034258308 |
| ALDH5A1 | 0.808930883 | 0.9181855 |
| LANCL1 | 0.808928036 | 1.083062946 |
| NCAM1 | 0.808927984 | 0.967112025 |
| GRID1 | 0.808873621 | 1.116158339 |
| ACY1 | 0.808770799 | 0.997404283 |
| PPM1H | 0.808754581 | 1.070084361 |
| TBCB | 0.808717698 | 0.913325233 |
| KLF9 | 0.808716877 | 0.772874757 |
| ZNF77 | 0.808401396 | 0.952380952 |
| GRK4 | 0.808354573 | 1.077222583 |
| KIAA0020 | 0.808350267 | 1.023491292 |
| TLR2 | 0.80834173 | 1.004542505 |
| XKR4 | 0.808267947 | 0.962332928 |
| BRAF | 0.808244918 | 0.998053212 |
| B3GNT2 | 0.808154774 | 0.885788449 |
| PIGY | 0.808153638 | 0.934386391 |
| CAPN2 | 0.808146871 | 0.99156392 |
| KIAA1324L | 0.808064136 | 0.898744431 |
| ZNF646 | 0.808058312 | 0.904761905 |
| FIG4 | 0.808049522 | 1.002740665 |
| UBE4A | 0.808043473 | 0.879299156 |
| PNPLA6 | 0.808032298 | 0.906439854 |
| CENPM | 0.808001021 | 0.914135277 |
| COL9A1 | 0.807886711 | 1.095585257 |
| TRIM60 | 0.807871335 | 0.939000649 |
| FIGN | 0.807831039 | 0.830078794 |
| DCAF12 | 0.807788584 | 1.056868791 |
| GOLGB1 | 0.807773541 | 1.003083248 |
| CHST8 | 0.807747033 | 0.95018226 |
| SYNC | 0.807615455 | 0.922235723 |
| FABP4 | 0.807588202 | 0.929931146 |
| RAD51D | 0.807540063 | 1.044957473 |
| KSR1 | 0.807515534 | 0.985723556 |
| C10orf91 | 0.807515321 | 0.874614594 |
| GPR3 | 0.807501704 | 1.018818949 |
| CLCN3 | 0.80746153 | 0.846852693 |
| NAGK | 0.807434693 | 0.913043478 |
| LMBRD2 | 0.807415922 | 0.876468206 |

| | | |
|---|---|---|
| PSG1 | 0.807373684 | 0.867077766 |
| COQ10A | 0.807300288 | 0.874038072 |
| CEACAM7 | 0.807250215 | 0.869133265 |
| ALKBH7 | 0.807248685 | 0.592952612 |
| RAB23 | 0.807198919 | 0.966255678 |
| ARHGEF4 | 0.807147543 | 0.965399109 |
| ZNF286A | 0.807076864 | 0.984204131 |
| ACOT2 | 0.807050222 | 0.795058728 |
| SGCD | 0.807026023 | 1.042480301 |
| TIMP3 | 0.806938284 | 0.688513952 |
| CTSK | 0.806852585 | 0.880597015 |
| EOGT | 0.806816509 | 1.043337384 |
| DEFB110 | 0.806810032 | 0.871531346 |
| YPEL5 | 0.806768319 | 0.797893884 |
| CEBPD | 0.806689735 | 0.989037342 |
| HMGCR | 0.806684893 | 1.033095393 |
| ENTPD5 | 0.806657068 | 0.966904607 |
| APOA1BP | 0.806648096 | 0.994861254 |
| CCDC18 | 0.806607145 | 0.948967193 |
| PRRX2 | 0.80658841 | 0.834523037 |
| METTL4 | 0.806575057 | 0.966084275 |
| KRTAP12-1 | 0.806526168 | 1.012332991 |
| GLS2 | 0.806509442 | 0.793033617 |
| TMEM237 | 0.806439581 | 0.958547448 |
| SPATA16 | 0.806434559 | 0.872417983 |
| BMPR1A | 0.806411415 | 1.0798183 |
| ELF4 | 0.806399105 | 0.78290806 |
| RNPC3 | 0.806339454 | 0.80599433 |
| FUT5 | 0.80632547 | 0.821544452 |
| TBP | 0.806288507 | 0.914341337 |
| OR10S1 | 0.806261016 | 0.956834532 |
| HOXB2 | 0.806231104 | 0.908464966 |
| SPRR2E | 0.806179604 | 0.875253139 |
| JAG2 | 0.805874997 | 0.861778066 |
| PAX6 | 0.805845946 | 0.924260834 |
| PPFIA2 | 0.805826426 | 0.828682674 |
| CPSF3L | 0.805820934 | 0.939651681 |
| GGT5 | 0.805793497 | 0.888384166 |
| TMEM205 | 0.805724958 | 0.936279548 |
| OR5AC2 | 0.805696413 | 0.895854745 |
| TMEM252 | 0.805656948 | 0.901678657 |
| LCN2 | 0.805512848 | 0.90927501 |
| MAP3K7 | 0.805402791 | 0.868916288 |
| MGARP | 0.80537331 | 1.056183625 |

| | | |
|---|---|---|
| C7orf34 | 0.805363025 | 0.979102432 |
| SFT2D1 | 0.805345255 | 0.740380721 |
| COQ2 | 0.805335945 | 0.955223881 |
| MOV10L1 | 0.805168543 | 0.894289186 |
| PNLIPRP2 | 0.805147148 | 0.917375456 |
| TM7SF2 | 0.805139456 | 1.058323208 |
| CDYL2 | 0.805124174 | 0.673957068 |
| KRTAP10-11 | 0.805077157 | 1.002025111 |
| MOB3B | 0.805060357 | 0.9331963 |
| REXO1L1P | 0.805018932 | 0.930336168 |
| MB | 0.804931859 | 0.904419322 |
| LCMT2 | 0.804866208 | 0.988968202 |
| EXO1 | 0.804835996 | 0.92926671 |
| AGK | 0.804821342 | 1.015574302 |
| PXDC1 | 0.804818993 | 0.90647482 |
| MALL | 0.804779004 | 0.707573917 |
| STAG3L2 | 0.804669652 | 0.95101062 |
| SH2D3A | 0.804633359 | 1.030147311 |
| DBNDD2 | 0.804457877 | 0.779667882 |
| FREM1 | 0.804415828 | 0.887632751 |
| C14orf159 | 0.804333138 | 0.883864337 |
| C12orf60 | 0.804318606 | 0.784933171 |
| UGCG | 0.804199712 | 0.913325233 |
| KRT39 | 0.804198693 | 0.90562981 |
| S100G | 0.804195535 | 0.88254354 |
| C8orf44 | 0.80418876 | 0.836371 |
| IL1RL2 | 0.804186366 | 0.820980154 |
| ZNHIT1 | 0.804158832 | 0.718054128 |
| PSAP | 0.804097017 | 0.980153908 |
| LIPI | 0.804087348 | 0.976638546 |
| NMRAL1 | 0.804084129 | 0.926285946 |
| MAD2L2 | 0.803963781 | 0.804023361 |
| AKR1CL1 | 0.803905189 | 0.992463172 |
| MMAA | 0.803883526 | 0.998379911 |
| CD320 | 0.803830905 | 0.890643985 |
| C15orf43 | 0.803760506 | 0.972863507 |
| TAS2R60 | 0.803755486 | 1.006166495 |
| PITX1 | 0.803615988 | 0.772782503 |
| C14orf37 | 0.803613696 | 0.924260834 |
| KCNA3 | 0.8035534 | 0.977287476 |
| FBXO47 | 0.803437406 | 0.917585983 |
| DEPDC4 | 0.803316572 | 0.9777321 |
| LAP3 | 0.803310135 | 0.831278391 |
| KCNJ16 | 0.803305278 | 0.857884491 |

| | | |
|---|---|---|
| PPP1R12C | 0.803292435 | 0.93770279 |
| SIK3 | 0.803288566 | 0.96430889 |
| MBTPS2 | 0.803284944 | 0.99091499 |
| SOX18 | 0.803214835 | 0.983799109 |
| ERLIN1 | 0.803155184 | 0.840016201 |
| GPR56 | 0.803153474 | 0.931213498 |
| NUTF2 | 0.803153474 | 0.744970798 |
| TRIM64C | 0.803153474 | 1.011031798 |
| CABP7 | 0.803136013 | 0.976019185 |
| RSPH10B | 0.803102212 | 0.933538883 |
| HIST1H4K | 0.803094102 | 0.987667009 |
| SPAG9 | 0.803068547 | 0.863994519 |
| WDR93 | 0.803027225 | 1.086330935 |
| CTSB | 0.803008338 | 0.898118105 |
| MAP6D1 | 0.80299788 | 0.822195221 |
| PIH1D2 | 0.802986572 | 0.89671932 |
| GSTM3 | 0.802983147 | 0.765087605 |
| ZNF214 | 0.802939197 | 0.865937627 |
| MAGEB3 | 0.802834161 | 0.81397739 |
| HESX1 | 0.802795954 | 0.85581207 |
| HSPA4L | 0.802724734 | 0.91174562 |
| PPP1R21 | 0.802693717 | 0.801944107 |
| LEPR | 0.802675999 | 1.091499027 |
| ABHD2 | 0.802668348 | 0.987039287 |
| FAM228A | 0.802581139 | 1 |
| OR5M9 | 0.802580155 | 0.943816375 |
| ABCA8 | 0.802543851 | 0.917780478 |
| PADI1 | 0.802498055 | 0.894694208 |
| FAM175B | 0.802473088 | 0.978759849 |
| NIPSNAP3A | 0.802429455 | 0.89756766 |
| TERT | 0.802349176 | 0.972096042 |
| DCHS1 | 0.802298166 | 1.074929121 |
| GPR180 | 0.802137603 | 0.697043337 |
| TBX21 | 0.8019993 | 0.936411503 |
| SDR39U1 | 0.80188207 | 0.952380952 |
| SF3B3 | 0.80188207 | 0.952380952 |
| CCDC108 | 0.801797577 | 0.990684488 |
| ZNF184 | 0.801741882 | 0.971908188 |
| ARHGAP18 | 0.801652939 | 1.101747174 |
| ACTRT3 | 0.801644829 | 0.884892086 |
| C9orf16 | 0.801607222 | 0.495717712 |
| KAT6B | 0.801601681 | 0.839714471 |
| MYC | 0.801574663 | 0.949777238 |
| CBL | 0.801496719 | 0.786502271 |

| | | |
|---|---|---|
| AUTS2 | 0.801493767 | 0.954779034 |
| CFDP1 | 0.801493767 | 0.954779034 |
| INTS10 | 0.801375996 | 0.9963548 |
| FRRS1 | 0.801366216 | 0.772377481 |
| RAB39B | 0.801348731 | 0.929770469 |
| NDUFB3 | 0.801332748 | 0.872823005 |
| ICT1 | 0.801329273 | 1 |
| LRRC61 | 0.801328927 | 1.017010936 |
| HAUS1 | 0.801323086 | 0.891058582 |
| MAPK8 | 0.801321123 | 0.853341986 |
| SP140 | 0.801306157 | 0.829080599 |
| SLC26A10 | 0.801279468 | 0.9733939 |
| IFNA17 | 0.801226395 | 0.943473792 |
| SLC6A4 | 0.801214185 | 0.873458793 |
| COX7A1 | 0.801175286 | 0.916409729 |
| BRSK2 | 0.801088313 | 0.820246593 |
| CEACAM18 | 0.801080045 | 0.974648852 |
| HBG1 | 0.801019211 | 0.888618874 |
| ADH4 | 0.801014387 | 0.913692408 |
| PSMA3 | 0.800786022 | 0.880597015 |
| CALCOCO1 | 0.800783715 | 0.798703929 |
| ASCL3 | 0.800781228 | 0.912983899 |
| TREML1 | 0.800781228 | 0.912983899 |
| GSN | 0.80076018 | 0.976508708 |
| OR5F1 | 0.800651507 | 0.85303186 |
| RIOK1 | 0.800584166 | 1.014276444 |
| NTF3 | 0.800539793 | 0.847501622 |
| PPP2R5B | 0.800522423 | 0.940947437 |
| LMCD1 | 0.80049247 | 0.901678657 |
| DDX47 | 0.800488516 | 0.935601458 |
| CD2AP | 0.800469498 | 0.924974306 |
| ZNF775 | 0.800455711 | 0.899554475 |
| ZP3 | 0.800431982 | 0.965741692 |
| IKZF2 | 0.800318886 | 0.868772783 |
| CEBPE | 0.80029581 | 0.917780478 |
| SERPINA5 | 0.800278499 | 0.861129137 |
| LDHAL6A | 0.80026061 | 0.776022681 |
| XRN1 | 0.800249709 | 0.987444309 |
| KRT13 | 0.80021291 | 0.867077766 |
| ADCY9 | 0.800201978 | 1.028756582 |
| DDX55 | 0.800140937 | 0.994808566 |
| TCP1 | 0.800129439 | 0.985014176 |
| SALL2 | 0.799938288 | 1.022610483 |
| DSCR9 | 0.79988079 | 0.914011648 |

| | | |
|---|---|---|
| LAMTOR4 | 0.799869735 | 0.948955122 |
| BAGE4 | 0.799770779 | 0.962658445 |
| PSMC1 | 0.79976612 | 1.115509409 |
| RNF4 | 0.79968001 | 0.975340688 |
| SUMF2 | 0.799671913 | 1.005481329 |
| PIK3R5 | 0.799645599 | 0.928617781 |
| NCOA7 | 0.799581067 | 1.16478246 |
| SMAGP | 0.799581067 | 0.570743405 |
| SLC15A1 | 0.799550147 | 0.910895099 |
| SOX21 | 0.799489044 | 0.992120589 |
| MTTP | 0.799447117 | 0.808565866 |
| DNAJC21 | 0.799394165 | 0.976704351 |
| FOXI1 | 0.799362229 | 0.942081814 |
| RCN2 | 0.799281662 | 0.914696814 |
| RAG2 | 0.799233809 | 0.939651681 |
| DCAF12L1 | 0.799223575 | 1.021582734 |
| ALG14 | 0.799191834 | 0.931956258 |
| FAM83G | 0.799153221 | 0.882948562 |
| KDELR2 | 0.799151133 | 0.903604698 |
| ALDOC | 0.799098478 | 0.996106424 |
| DGKA | 0.799098478 | 0.996106424 |
| MMP14 | 0.7990872 | 0.92926671 |
| TTF1 | 0.799061631 | 0.84690158 |
| TBC1D21 | 0.799061185 | 0.878040425 |
| SNX3 | 0.799057742 | 0.872216513 |
| GNAS | 0.79901953 | 0.882543803 |
| PRSS42 | 0.798987095 | 0.969500324 |
| OSTCP1 | 0.798978751 | 0.976103686 |
| FER1L5 | 0.798975033 | 0.934791414 |
| EVA1B | 0.798964284 | 0.831510733 |
| SOHLH1 | 0.798917929 | 0.844471446 |
| IGFL2 | 0.798859557 | 1.012332991 |
| PUF60 | 0.798846871 | 0.870503597 |
| KDM3B | 0.798842521 | 0.989037342 |
| ATMIN | 0.798827816 | 0.69218307 |
| SIMC1 | 0.798780237 | 0.862697448 |
| KIF22 | 0.798769926 | 1.070084361 |
| DNAJC5B | 0.798699294 | 1.004860267 |
| TMEM136 | 0.798694964 | 0.772377481 |
| PTPRE | 0.798578196 | 0.996755354 |
| KCNJ2 | 0.798540574 | 1.016872161 |
| MTERFD3 | 0.79853491 | 0.844876468 |
| F8A2 | 0.798467629 | 0.90202124 |
| MRPS9 | 0.79845304 | 0.890373416 |

| | | |
|---|---|---|
| COX17 | 0.798414691 | 0.938031592 |
| NRL | 0.798346431 | 0.509113001 |
| LRRC41 | 0.798316172 | 0.86836776 |
| NME3 | 0.798285866 | 0.963659961 |
| SLAMF9 | 0.79826215 | 1.038369305 |
| NPBWR2 | 0.798221324 | 1.003893576 |
| MAGEA8 | 0.798184762 | 0.523124358 |
| ZNF296 | 0.798161699 | 0.836245289 |
| HECW1 | 0.798133732 | 1.064243997 |
| TOR4A | 0.798034881 | 0.985014176 |
| KRT40 | 0.797962138 | 0.956662616 |
| WDR88 | 0.79780295 | 0.899959498 |
| MORF4_pseudogene | 0.79777151 | 0.944192083 |
| WNK2 | 0.797705652 | 1.004542505 |
| FKRP | 0.797704208 | 0.92428914 |
| FAM118B | 0.797485797 | 1.001215067 |
| SMYD2 | 0.797450038 | 0.890979883 |
| SPATA32 | 0.797422733 | 0.913325233 |
| GDNF | 0.797287079 | 0.957472661 |
| KIAA0907 | 0.797246853 | 0.860568688 |
| GPATCH1 | 0.797123357 | 0.980963953 |
| ZNF467 | 0.797114443 | 0.76873228 |
| DMD | 0.797065705 | 0.907502569 |
| N4BP2L1 | 0.79687681 | 0.911613566 |
| OTOS | 0.79687681 | 0.911613566 |
| GPR17 | 0.796843316 | 1.032446463 |
| PF4 | 0.79679122 | 0.981843097 |
| SYPL1 | 0.796757691 | 0.970195272 |
| SCAF4 | 0.796731094 | 0.865022268 |
| OR52J3 | 0.796711837 | 0.859198356 |
| IGF2 | 0.796605092 | 0.872823005 |
| RPL34 | 0.796527473 | 0.956834532 |
| TSEN54 | 0.796519156 | 0.779667882 |
| ATP6V1B1 | 0.796483039 | 0.831510733 |
| PLAC8L1 | 0.796409838 | 0.82381531 |
| PALLD | 0.796323138 | 0.842041312 |
| POLR1C | 0.796322127 | 0.972744971 |
| DBR1 | 0.79629074 | 0.919805589 |
| DUS2 | 0.796249377 | 0.834345889 |
| PPP1R9B | 0.796187333 | 0.807125728 |
| NDUFA5 | 0.796139855 | 0.969218307 |
| PRORSD1P | 0.796043521 | 0.980815348 |
| SERTAD3 | 0.796022452 | 0.974991435 |
| SMARCC2 | 0.795972912 | 0.979883193 |

| | | |
|---|---|---|
| DDAH2 | 0.795940272 | 0.933171324 |
| LOC440348 | 0.795932829 | 0.900993491 |
| SLC23A1 | 0.795910017 | 0.899415964 |
| NUS1 | 0.7958551 | 0.756986634 |
| CLEC4C | 0.795843796 | 0.928400137 |
| ZNF649 | 0.795805484 | 0.956662616 |
| LCN1 | 0.795727749 | 0.910085055 |
| FABP6 | 0.795689144 | 1.047371836 |
| FUT7 | 0.795628801 | 0.987021415 |
| TICAM2 | 0.795622507 | 0.811664642 |
| B3GNT6 | 0.795509927 | 0.886437378 |
| RETSAT | 0.795497652 | 0.874038072 |
| EFHD2 | 0.795487108 | 0.995949777 |
| LRRC75A-AS1 | 0.795468718 | 0.957067639 |
| LRRC3B | 0.795462249 | 0.809234508 |
| PCDH15 | 0.795365449 | 0.9201817 |
| LINC00477 | 0.795353546 | 0.971908188 |
| KPNA2 | 0.795343634 | 1.034393251 |
| EDC4 | 0.795340553 | 0.967598218 |
| C14orf119 | 0.795299854 | 1.173004454 |
| CA12 | 0.79528963 | 0.994159637 |
| MKX | 0.79516374 | 0.830295666 |
| MITD1 | 0.795150037 | 0.734305387 |
| REEP3 | 0.795117027 | 0.861482382 |
| CYTH2 | 0.795112499 | 0.919492977 |
| SEC14L4 | 0.795063701 | 1.030832477 |
| PBX4 | 0.795039449 | 0.947436729 |
| RHD | 0.794995128 | 0.955042527 |
| RAB15 | 0.794989031 | 0.974306269 |
| PCSK1N | 0.794811284 | 0.913730255 |
| C15orf37 | 0.794778326 | 0.887808829 |
| EYA3 | 0.79477725 | 1.08890331 |
| KAT7 | 0.794735333 | 0.963142973 |
| SH2B1 | 0.794502027 | 0.851761847 |
| PARP3 | 0.794500957 | 0.895909275 |
| POLE2 | 0.79449521 | 0.948085659 |
| CLSTN2 | 0.794479868 | 0.971243418 |
| CDK2AP1 | 0.794459084 | 0.914135277 |
| DNAJC5 | 0.794398828 | 1.021925317 |
| NPY | 0.794398563 | 0.907852044 |
| SETD9 | 0.794241264 | 0.872823005 |
| EFCAB4B | 0.794214469 | 0.784528149 |
| NR2C2 | 0.794189305 | 0.84690158 |
| KCNE1 | 0.794176949 | 1.103179753 |

| | | |
|---|---|---|
| PROKR1 | 0.794039716 | 0.914990266 |
| EIF4E | 0.79402594 | 0.875658161 |
| CES2 | 0.794015701 | 0.938031592 |
| SPIB | 0.794008351 | 0.968851395 |
| PLA2G2E | 0.793979177 | 0.75617659 |
| SSPN | 0.793944599 | 0.907159986 |
| TGM6 | 0.793872865 | 0.748481166 |
| OR1N2 | 0.793871243 | 0.895512162 |
| SF3B2 | 0.793756972 | 0.924974306 |
| CCDC6 | 0.793710077 | 0.979787598 |
| SF3A3 | 0.793609988 | 1.009249743 |
| TASP1 | 0.793549893 | 0.881894873 |
| PIK3CB | 0.793476954 | 1.023361454 |
| PCDHGB2 | 0.793416923 | 0.9331963 |
| EXOSC6 | 0.793413486 | 0.816524909 |
| ALDH16A1 | 0.793370289 | 0.852976914 |
| SHISA5 | 0.793368149 | 0.902389631 |
| SPINT4 | 0.79336438 | 1.001215067 |
| OR1J2 | 0.793358655 | 0.616306954 |
| TMEM92 | 0.793348079 | 0.970028352 |
| DTL | 0.79331293 | 0.957067639 |
| TNFRSF4 | 0.793287035 | 1.016872161 |
| TOMM34 | 0.793198531 | 0.829736211 |
| IFI44L | 0.793137136 | 0.874038072 |
| SH3GL3 | 0.793002613 | 0.916067146 |
| FAM109A | 0.793000157 | 0.921020656 |
| MAGI1 | 0.792995775 | 0.936404932 |
| OTULIN | 0.792969542 | 0.889833941 |
| APPL1 | 0.792951255 | 0.668691778 |
| LPAR2 | 0.792946134 | 0.970149254 |
| CLEC1B | 0.79290984 | 0.723369785 |
| CD70 | 0.792885491 | 0.949777238 |
| SLC9A5 | 0.792885491 | 0.949777238 |
| PKMYT1 | 0.79284002 | 0.808565866 |
| M1AP | 0.79281443 | 1.11381126 |
| SRPRB | 0.792738956 | 0.867420349 |
| STK35 | 0.792698753 | 0.909798832 |
| RHOB | 0.792694061 | 0.822193381 |
| FGB | 0.792658332 | 0.943543154 |
| PKD1L1 | 0.792626498 | 0.932168551 |
| ATG4A | 0.792625979 | 0.680726801 |
| MAVS | 0.792589631 | 0.981368975 |
| CRYBB3 | 0.792538552 | 0.920520726 |
| SNAI3 | 0.79251191 | 0.955806783 |

| | | |
|---|---|---|
| GPR50 | 0.792479362 | 0.903309539 |
| NDOR1 | 0.792450513 | 0.960712839 |
| AGFG1 | 0.792402731 | 1.111786148 |
| ATP2C2 | 0.792402731 | 0.874848117 |
| LEPREL1 | 0.792349549 | 0.861887404 |
| C19orf35 | 0.792294738 | 0.848926691 |
| GPR132 | 0.792208683 | 0.96430889 |
| LIG3 | 0.792166945 | 1.031797534 |
| DPRX | 0.792152605 | 0.811236725 |
| HNMT | 0.792133686 | 0.910447761 |
| SHOX2 | 0.792036064 | 0.875253139 |
| GLRA3 | 0.792031112 | 0.890330954 |
| DNAJC30 | 0.79200929 | 1.025008565 |
| ZNF749 | 0.791906268 | 0.831105711 |
| NOTCH1 | 0.791832054 | 0.782608696 |
| MAGED4B | 0.791728779 | 1.011647825 |
| C19orf45 | 0.791684678 | 0.893884164 |
| NTS | 0.791653095 | 0.917375456 |
| ESR1 | 0.791637915 | 0.899149453 |
| USP18 | 0.791569525 | 0.944841012 |
| PTGIS | 0.791557583 | 0.867962738 |
| HLA-DOA | 0.791539499 | 0.849736736 |
| MUC16 | 0.791501506 | 0.904076739 |
| ZMYM5 | 0.791500733 | 1.016605913 |
| C15orf59 | 0.791496023 | 0.982528263 |
| PAPD7 | 0.791427271 | 0.722258274 |
| RNMT | 0.791358119 | 0.972096042 |
| CACNG5 | 0.791350463 | 0.93835172 |
| SLC45A1 | 0.791347607 | 0.912515188 |
| TMEM151B | 0.791249131 | 0.953408702 |
| OCIAD2 | 0.7911251 | 0.897124342 |
| BOP1 | 0.791119954 | 0.971223022 |
| GAS2L1 | 0.791096755 | 1.065433368 |
| P2RY8 | 0.791062414 | 1.012978585 |
| AK7 | 0.791026274 | 0.945489942 |
| BCL2A1 | 0.790961074 | 0.844256976 |
| SQLE | 0.790852061 | 0.798298906 |
| MAP2K7 | 0.790851008 | 0.857884491 |
| TBCK | 0.790803672 | 0.939000649 |
| ADSSL1 | 0.790775106 | 0.905256327 |
| CHRM2 | 0.790775106 | 0.905256327 |
| SIX1 | 0.790758891 | 0.808829486 |
| C16orf54 | 0.79068131 | 0.772377481 |
| PPP2R5A | 0.790644871 | 1 |

| | | |
|---|---|---|
| CHRAC1 | 0.790623594 | 0.824625354 |
| PIGU | 0.790487278 | 0.981368975 |
| EIF4A1 | 0.790431983 | 0.993510707 |
| RELA | 0.790392056 | 0.959766385 |
| AIFM2 | 0.790353611 | 0.844905905 |
| B3GNT4 | 0.79020864 | 0.905905256 |
| OR4E2 | 0.790207237 | 0.944844125 |
| C1QTNF5 | 0.79014906 | 0.939020212 |
| ATP5C1 | 0.790110474 | 0.940056703 |
| C1orf189 | 0.790082864 | 0.869582827 |
| TCF23 | 0.790082454 | 0.807468311 |
| PNPLA4 | 0.790082027 | 0.966904607 |
| GOLGA7B | 0.790065137 | 0.921830701 |
| PTRH1 | 0.790033198 | 0.851356825 |
| C21ORF33 | 0.790013493 | 1.002055498 |
| RALBP1 | 0.78999321 | 0.93236128 |
| PAWR | 0.789955769 | 0.861887404 |
| CHRNA3 | 0.789945206 | 1.075275795 |
| LRRC29 | 0.789931293 | 0.689811811 |
| GTSE1 | 0.789881734 | 0.913043478 |
| RAP1B | 0.789836539 | 1.007787151 |
| MORN1 | 0.789820217 | 0.958687728 |
| UBAP2L | 0.78975048 | 0.812474686 |
| NBPF15 | 0.78969903 | 0.898744431 |
| C9orf171 | 0.789667781 | 0.967112025 |
| DET1 | 0.789587764 | 0.80404248 |
| ITPK1 | 0.789568495 | 0.872809864 |
| DALRD3 | 0.78954896 | 0.867557716 |
| TOPBP1 | 0.789487982 | 0.839065542 |
| FGF14 | 0.789479174 | 0.933809215 |
| SLC16A10 | 0.789460324 | 0.825840421 |
| GMCL1 | 0.789416822 | 0.859862292 |
| AZU1 | 0.789400802 | 0.994808566 |
| C2orf54 | 0.789357018 | 0.812879708 |
| DNPH1 | 0.789324653 | 0.961064244 |
| LYZL1 | 0.789324653 | 0.961064244 |
| PI4KB | 0.78925756 | 1.022063595 |
| PSMB1 | 0.789121809 | 1.049318624 |
| IL3 | 0.789022939 | 0.968202466 |
| NLRC4 | 0.78901903 | 0.894289186 |
| TMEM54 | 0.78901256 | 0.865532604 |
| TNS1 | 0.788987905 | 0.899554475 |
| KRTAP1-1 | 0.788808077 | 1.076396026 |
| EIF5A | 0.788771359 | 0.697598962 |

| | | |
|---|---|---|
| ATP1B4 | 0.788765806 | 0.907654921 |
| DXO | 0.788722623 | 0.832476876 |
| PKP2 | 0.78870903 | 0.848235697 |
| SLC44A2 | 0.788691094 | 0.927968851 |
| TPBG | 0.788574053 | 1.045220966 |
| GABRD | 0.788536655 | 1.016223232 |
| TAP2 | 0.788477053 | 1.029850746 |
| SEC31B | 0.788435765 | 0.970838396 |
| LYRM2 | 0.788335107 | 0.991899554 |
| ABHD17B | 0.788329118 | 0.887735237 |
| AURKAIP1 | 0.788274044 | 0.779364049 |
| AKIP1 | 0.788206682 | 0.947242206 |
| ATAD1 | 0.788199046 | 0.916160389 |
| PRG3 | 0.788070301 | 0.872559096 |
| GCNT1 | 0.788062473 | 0.820246593 |
| SNUPN | 0.78804322 | 0.620494127 |
| TESK1 | 0.787966649 | 0.969500324 |
| C3orf56 | 0.787914647 | 0.82746051 |
| NEU2 | 0.787892233 | 0.945321993 |
| PLA2G4C | 0.787881026 | 0.872012961 |
| KCNB1 | 0.787871072 | 0.861129137 |
| HEATR2 | 0.787861618 | 0.83799109 |
| ABCC2 | 0.787845963 | 0.800129786 |
| CGREF1 | 0.787843345 | 0.902363823 |
| YES1 | 0.787836743 | 0.935756003 |
| ARL1 | 0.787710224 | 0.869582827 |
| JAK2 | 0.787602616 | 0.854639844 |
| VHLL | 0.787599398 | 1.028776978 |
| AKAP11 | 0.787594029 | 1.058323208 |
| KCNJ11 | 0.78753083 | 1.01752109 |
| NHEJ1 | 0.78749222 | 0.881894873 |
| CEACAM1 | 0.787470809 | 0.904761905 |
| CPNE1 | 0.787447372 | 0.888213852 |
| CYP4F3 | 0.787375954 | 0.948562171 |
| PHC3 | 0.787332117 | 1.021240151 |
| ZNF711 | 0.787135313 | 0.922640745 |
| YIPF1 | 0.787133072 | 0.784857828 |
| POMK | 0.787019767 | 1.1064244 |
| CCDC129 | 0.787017335 | 0.859862292 |
| PQLC3 | 0.786900733 | 0.84163629 |
| TMEM17 | 0.786871318 | 0.962658445 |
| PAQR8 | 0.786859879 | 0.802349129 |
| C9orf152 | 0.786822489 | 0.862697448 |
| PTK7 | 0.78670661 | 0.950681376 |

| | | |
|---|---|---|
| RNF167 | 0.78670661 | 0.760545101 |
| REXO2 | 0.786641244 | 0.700283516 |
| SLC32A1 | 0.786624924 | 0.937307297 |
| TBXA2R | 0.786565343 | 0.903309539 |
| SARDH | 0.786527639 | 0.916937054 |
| SLC25A10 | 0.786504674 | 0.847306602 |
| CCDC66 | 0.786482209 | 0.96800324 |
| FLJ44838 | 0.786432268 | 0.951353203 |
| COPB2 | 0.786381023 | 0.795477903 |
| CYP1B1 | 0.786380023 | 1.004860267 |
| ST6GAL2 | 0.78611381 | 0.984204131 |
| COX14 | 0.786084366 | 0.863507493 |
| RFXAP | 0.786081859 | 0.992212849 |
| NKX2-8 | 0.785946114 | 0.987039287 |
| TYRO3 | 0.78594298 | 0.931213498 |
| UST | 0.78588412 | 0.942486837 |
| ENTHD2 | 0.785793854 | 0.75617659 |
| PDHX | 0.785793854 | 0.75617659 |
| LRRTM4 | 0.785755598 | 0.924260834 |
| NPHS2 | 0.785743882 | 0.929526124 |
| LEPREL4 | 0.785734319 | 0.956149366 |
| NKX2-2 | 0.785720959 | 0.940056703 |
| NFE2 | 0.78572046 | 1.053861129 |
| ERCC4 | 0.785644549 | 0.895504253 |
| TTC30B | 0.785616888 | 0.743215877 |
| UBXN2A | 0.785616197 | 0.993833505 |
| KCNS3 | 0.785576477 | 0.82284231 |
| SPANXF1 | 0.785567353 | 1.029462145 |
| CD63 | 0.785526756 | 0.903048989 |
| CTDSPL | 0.785501156 | 0.920863309 |
| RGCC | 0.785337634 | 0.869582827 |
| OBSCN | 0.785252814 | 0.911270983 |
| MATN4 | 0.785192556 | 0.953827461 |
| APOE | 0.785163708 | 0.898118105 |
| ATE1 | 0.785163708 | 0.898118105 |
| IL33 | 0.785144571 | 0.985419198 |
| NOG | 0.785100166 | 0.9331963 |
| MLH3 | 0.785083854 | 0.966255678 |
| MRPL18 | 0.78492726 | 0.901579587 |
| RTF1 | 0.784819929 | 0.846496557 |
| C8orf58 | 0.784818793 | 1.020554985 |
| ASB15 | 0.784735792 | 0.721609345 |
| PTK2B | 0.78459683 | 0.898767034 |
| SLIT1 | 0.784574212 | 0.888317917 |

| | | |
|---|---|---|
| ZCCHC10 | 0.784473987 | 0.997259335 |
| PF4V1 | 0.784411143 | 0.866735183 |
| NTAN1 | 0.784382009 | 1.009249743 |
| CSNK1G1 | 0.784339405 | 0.824140169 |
| GPR78 | 0.784307382 | 0.919532771 |
| DNAJB6 | 0.784284825 | 1.001297859 |
| IL1RL1 | 0.784248482 | 0.860267315 |
| ZNF138 | 0.784172633 | 0.936279548 |
| FOXN4 | 0.784166924 | 0.793764988 |
| HS6ST2 | 0.784030693 | 0.804023361 |
| ATP6AP1L | 0.783896834 | 0.873244262 |
| CCL26 | 0.78386412 | 0.96437136 |
| HSPE1 | 0.783793026 | 1.070084361 |
| SERPINB5 | 0.783762237 | 0.947436729 |
| GPR31 | 0.783732189 | 0.852044127 |
| SLC25A43 | 0.783695017 | 0.847711624 |
| DMTN | 0.783562585 | 0.883521754 |
| ARL9 | 0.783559458 | 1.156903049 |
| CSGALNACT1 | 0.783540108 | 1.024301337 |
| APOC1 | 0.783522381 | 0.918995545 |
| HCG9 | 0.783475047 | 0.941075711 |
| CDK19 | 0.783447763 | 0.872809864 |
| MLST8 | 0.783381528 | 0.834189791 |
| CA4 | 0.783381175 | 1.011340624 |
| SPRR2G | 0.783229786 | 0.949297705 |
| ZNF750 | 0.783124946 | 0.835155934 |
| ZNF419 | 0.783070796 | 1.017010936 |
| TRIL | 0.783009144 | 0.996106424 |
| POLR2D | 0.782944768 | 0.702790396 |
| KRTAP1-5 | 0.7828872 | 0.951695786 |
| CHD3 | 0.782788271 | 0.818948735 |
| CPNE5 | 0.782787745 | 1.011305242 |
| IGF1R | 0.782762916 | 1.003244646 |
| RPS6KC1 | 0.782745463 | 0.989617132 |
| PCYT1A | 0.782744321 | 1.001620089 |
| SH3BP5 | 0.782675876 | 0.25385406 |
| CCDC73 | 0.782674583 | 0.77764277 |
| TLL1 | 0.78265038 | 0.921479559 |
| BEND3 | 0.782593843 | 0.86472256 |
| OR4N4 | 0.782589742 | 0.92428914 |
| PCSK5 | 0.782586332 | 0.880597015 |
| IK | 0.782532426 | 0.803357314 |
| KCNMA1 | 0.782478242 | 0.983127839 |
| SLC22A12 | 0.782387028 | 0.928617781 |

| | | |
|---|---|---|
| DAW1 | 0.782372355 | 0.959902795 |
| ZG16B | 0.782370489 | 0.960260363 |
| PLEKHN1 | 0.782367516 | 0.717294451 |
| POU3F3 | 0.782327324 | 0.928311057 |
| ARL13A | 0.782280969 | 1.010125557 |
| OSBPL3 | 0.782250863 | 0.989064399 |
| C15orf60 | 0.782205925 | 0.851318945 |
| NSMCE1 | 0.78219428 | 0.861253854 |
| G3BP2 | 0.782181631 | 0.963011032 |
| CLN3 | 0.782160978 | 0.891058582 |
| RNF7 | 0.78207066 | 0.77871512 |
| TMEM176B | 0.782049395 | 0.867962738 |
| VPS16 | 0.782033975 | 0.962658445 |
| TEX37 | 0.781838808 | 0.855002025 |
| GPX5 | 0.781808131 | 0.915639195 |
| IL22 | 0.78180122 | 1.068174032 |
| OR2B2 | 0.781756363 | 0.865707434 |
| ZC3H10 | 0.781755215 | 0.818144998 |
| DBN1 | 0.781726232 | 0.91538198 |
| PHLDA2 | 0.781723479 | 0.965978129 |
| FBXO25 | 0.781620763 | 0.950032446 |
| SLC37A4 | 0.781549554 | 0.717715769 |
| FUNDC2 | 0.781509021 | 0.871202916 |
| TMEM141 | 0.781455891 | 0.921425678 |
| DDX6 | 0.781351813 | 0.854639844 |
| C15orf32 | 0.781291048 | 0.990279465 |
| IL11 | 0.781254425 | 0.916288125 |
| HLA-A | 0.781133949 | 0.805589307 |
| PHKB | 0.781128882 | 0.96430889 |
| KDM5B | 0.781113405 | 0.697598962 |
| ZCCHC14 | 0.781112605 | 0.921830701 |
| CCDC71L | 0.781025429 | 0.920178143 |
| LARS | 0.781023526 | 0.910243234 |
| CKMT2 | 0.780951393 | 1.073977936 |
| EIF2S3 | 0.780888333 | 0.971447112 |
| MRGPRD | 0.780888333 | 0.971447112 |
| DENND5A | 0.7808354 | 1.027749229 |
| ZACN | 0.780786829 | 1.019467878 |
| INHBE | 0.780783022 | 0.898595409 |
| TATDN1 | 0.780739706 | 0.930455635 |
| OXTR | 0.780651325 | 0.828033744 |
| COA3 | 0.780641714 | 0.935601458 |
| SMAD7 | 0.780516452 | 0.898744431 |
| MTURN | 0.780282433 | 0.790603483 |

| | | |
|---|---|---|
| GPC5 | 0.78026613 | 0.917094896 |
| EXOSC4 | 0.78026136 | 0.944841012 |
| CSDE1 | 0.78021025 | 0.819765087 |
| EIF3F | 0.780205717 | 0.909680032 |
| PRELP | 0.780072477 | 0.996916752 |
| PPRC1 | 0.780032786 | 1.00890716 |
| OR7A10 | 0.779974699 | 0.917437479 |
| SEC61G | 0.779961333 | 0.907502569 |
| MGLL | 0.779904242 | 0.924724205 |
| SIX6 | 0.779904242 | 0.986372485 |
| HSPB9 | 0.779880093 | 0.97327852 |
| GCLC | 0.779865895 | 0.99473471 |
| ETNK1 | 0.779842582 | 0.91109669 |
| C16orf74 | 0.779836197 | 0.904819765 |
| KRT6A | 0.779761427 | 0.967454608 |
| RMDN2 | 0.779671151 | 0.828270555 |
| CPT1A | 0.779424702 | 0.929121102 |
| KBTBD4 | 0.779400944 | 0.981843097 |
| POLG2 | 0.779378448 | 0.992709599 |
| VPS13A | 0.779368668 | 0.770352369 |
| ADCK5 | 0.779360991 | 1.03504218 |
| TMEM230 | 0.779350839 | 0.913325233 |
| SDHA | 0.779345388 | 0.839206156 |
| NTRK3 | 0.779298672 | 1.021414666 |
| QPRT | 0.779287476 | 0.91174562 |
| AARSD1 | 0.779221858 | 0.886998785 |
| DAG1 | 0.779101343 | 0.980532122 |
| CHRNE | 0.779031862 | 0.966904607 |
| SIRPD | 0.778945355 | 0.868772783 |
| ARID3A | 0.778889066 | 0.990684488 |
| INHBA | 0.778820209 | 0.829980532 |
| IFNGR2 | 0.778815435 | 0.878001298 |
| SERPINB9 | 0.778813573 | 0.71526934 |
| PTPN3 | 0.778800699 | 1.070084361 |
| ATP11C | 0.778718569 | 0.861077359 |
| NOMO2 | 0.778699053 | 0.406646112 |
| ZNF676 | 0.778688005 | 0.85581207 |
| COMMD3 | 0.778538417 | 0.741077223 |
| CD2BP2 | 0.778502685 | 0.916970433 |
| TMCO2 | 0.778480016 | 0.88254354 |
| ALG12 | 0.778330201 | 0.739570676 |
| SLC35B2 | 0.778315378 | 0.9145403 |
| RNF220 | 0.778288895 | 0.974691759 |
| DEPDC1B | 0.778266346 | 0.985268928 |

| | | |
|---|---|---|
| PARP8 | 0.778130923 | 0.94127177 |
| TSPYL5 | 0.777957199 | 0.907845152 |
| FGF1 | 0.777857275 | 0.906554186 |
| ACD | 0.777757939 | 0.695018226 |
| C10orf120 | 0.777745408 | 0.960307817 |
| PAXIP1 | 0.77773303 | 0.771972458 |
| PSMC6 | 0.777730585 | 0.878088295 |
| ASPM | 0.777702015 | 0.964028777 |
| ZNF653 | 0.777664108 | 0.973673552 |
| RAB6B | 0.777466569 | 0.968851395 |
| MAS1 | 0.777438002 | 1.003244646 |
| CPAMD8 | 0.777403244 | 0.930455635 |
| MOS | 0.777300852 | 0.756002596 |
| RAB36 | 0.777295301 | 0.867962738 |
| KDM1A | 0.777122951 | 1.031148605 |
| MAP4K5 | 0.777122951 | 1.031148605 |
| PIF1 | 0.777111015 | 0.845554835 |
| CXorf66 | 0.776921909 | 0.796678817 |
| MYADM | 0.776897951 | 0.821171634 |
| POLR2K | 0.776741219 | 0.942245295 |
| MED27 | 0.776733892 | 1.011031798 |
| RRM2B | 0.776701338 | 0.794248684 |
| FAM19A4 | 0.776690386 | 0.770352369 |
| NGB | 0.776687352 | 0.913326482 |
| PIDD1 | 0.77663221 | 0.855429942 |
| COL25A1 | 0.776598137 | 0.965399109 |
| SOX7 | 0.776520522 | 1.029462145 |
| CRYGB | 0.776504392 | 0.957519699 |
| ACTR3B | 0.776441441 | 1.028351559 |
| ZEB2 | 0.776378734 | 0.932766302 |
| CD68 | 0.776327639 | 0.9777321 |
| SPIC | 0.776318077 | 0.956521739 |
| MRFAP1 | 0.77627502 | 0.898339409 |
| SPARCL1 | 0.776213595 | 0.933881466 |
| CGA | 0.776150161 | 1.059952038 |
| LRRC20 | 0.775977577 | 0.994176088 |
| OR52L1 | 0.775958819 | 0.970195272 |
| CHD5 | 0.775932311 | 1.005191434 |
| NCALD | 0.775837888 | 0.938335046 |
| MRPL34 | 0.775764531 | 1.00445358 |
| KLF10 | 0.77571119 | 0.764438676 |
| KRT28 | 0.77570775 | 1.013365735 |
| SIDT1 | 0.775705101 | 0.933576347 |
| TMEM147 | 0.775705101 | 0.933576347 |

| | | |
|---|---|---|
| CCDC58 | 0.775659764 | 0.928311057 |
| TXNRD2 | 0.775632308 | 1.082624544 |
| GSAP | 0.775615735 | 0.910585817 |
| PANX2 | 0.775615735 | 0.910585817 |
| PRR15 | 0.775613918 | 0.923045768 |
| FUT4 | 0.775514891 | 0.795463751 |
| SETD8 | 0.775500831 | 0.968408262 |
| STAT1 | 0.775393857 | 1.032401782 |
| RORC | 0.775343453 | 0.937053861 |
| SSRP1 | 0.775325844 | 0.974991435 |
| MGAT3 | 0.775268238 | 0.979343864 |
| AZGP1 | 0.775218868 | 0.877012676 |
| CDH7 | 0.77520011 | 1.097293594 |
| EPS8 | 0.775127185 | 0.907654921 |
| ARAF | 0.77506734 | 0.96495782 |
| HLA-C | 0.775053345 | 1.020116807 |
| TFF1 | 0.775032555 | 0.963343611 |
| PSMA6 | 0.774978925 | 1.096041531 |
| GABRQ | 0.774938998 | 0.951330305 |
| DPAGT1 | 0.774879808 | 0.861778066 |
| CLCA2 | 0.774871493 | 0.972096042 |
| MICA | 0.774869984 | 0.879410757 |
| KIAA1324 | 0.774818233 | 1.001215067 |
| PRIM1 | 0.77480687 | 0.71706684 |
| FNBP4 | 0.774685228 | 1.004050223 |
| MOB4 | 0.774632426 | 0.969852689 |
| H3F3A | 0.774572798 | 0.7995889 |
| RNF168 | 0.774546481 | 0.965606749 |
| NOM1 | 0.774529045 | 0.969623329 |
| FAM43A | 0.774443938 | 1.006166495 |
| BSND | 0.774296713 | 0.862426995 |
| BRIP1 | 0.774290696 | 0.759416768 |
| PSMG3 | 0.774290696 | 0.810044552 |
| ARFGAP1 | 0.774271084 | 0.842041312 |
| GCKR | 0.774181049 | 0.903958469 |
| AMDHD1 | 0.77416681 | 0.799513973 |
| IGHMBP2 | 0.774152071 | 1.055807917 |
| SLC17A2 | 0.774038961 | 0.904419322 |
| C1orf105 | 0.774010933 | 0.884973674 |
| ADAMTS4 | 0.773930045 | 0.931862427 |
| COLEC10 | 0.773893064 | 0.842411785 |
| IL12RB2 | 0.77386501 | 0.800778715 |
| PCDHGA9 | 0.773781874 | 0.930798219 |
| CPD | 0.773714605 | 1.03569111 |

| | | |
|---|---|---|
| APOLD1 | 0.773639222 | 0.962737951 |
| CYP2W1 | 0.773614253 | 1.001297859 |
| EXOSC5 | 0.773506821 | 1.063595068 |
| ACADS | 0.773506625 | 0.778857837 |
| ERICH6 | 0.773448502 | 0.960307817 |
| TMEM123 | 0.773437245 | 0.773592548 |
| SLC30A8 | 0.773409788 | 0.786958283 |
| ZNRF4 | 0.773281173 | 1.112264763 |
| GZMA | 0.773277222 | 0.780661908 |
| ANKH | 0.773267343 | 0.898118105 |
| TSPAN4 | 0.773157634 | 0.888618874 |
| ZNF10 | 0.773139928 | 0.949640288 |
| RFX2 | 0.773089221 | 0.83799109 |
| RABGGTB | 0.773032525 | 0.77399757 |
| ADD1 | 0.772914751 | 0.971648441 |
| DNHD1 | 0.772885461 | 0.880923451 |
| ERCC2 | 0.772878036 | 0.912394549 |
| FITM2 | 0.772833227 | 0.863651936 |
| PRDM8 | 0.772810375 | 0.947752126 |
| BHMT | 0.772786493 | 0.995457495 |
| UBE4B | 0.772786493 | 0.995457495 |
| RASSF1 | 0.77272398 | 0.969218307 |
| AFMID | 0.772641304 | 0.990684488 |
| ANKRD11 | 0.772636266 | 0.843782117 |
| PPP1R35 | 0.772563009 | 0.926690968 |
| METTL6 | 0.772485108 | 0.947436729 |
| TLE3 | 0.772444561 | 0.868105516 |
| DPP10 | 0.772432668 | 0.767683323 |
| CYP51A1 | 0.772369946 | 0.910895099 |
| RPS23 | 0.772328776 | 0.913043478 |
| MICAL2 | 0.772272714 | 1.016872161 |
| LRRK2 | 0.772211218 | 1.162232317 |
| QPCTL | 0.772175405 | 0.881733495 |
| CMPK2 | 0.772168037 | 0.912641316 |
| CEBPG | 0.772140257 | 0.78290806 |
| PABPN1L | 0.772135958 | 0.944844125 |
| GSDMB | 0.772091781 | 0.993148338 |
| SF3B6 | 0.772089826 | 0.929931146 |
| IFI16 | 0.772042961 | 0.844471446 |
| EPHA1 | 0.77199372 | 0.781959766 |
| ANXA10 | 0.771964597 | 0.985268928 |
| CCR4 | 0.771960297 | 0.927319922 |
| AAMP | 0.771820862 | 0.855407047 |
| NLRP10 | 0.771820862 | 0.62818955 |

| | | |
|---|---|---|
| HYAL1 | 0.771780256 | 0.851395198 |
| AKAP13 | 0.771665494 | 0.804779263 |
| MTDH | 0.771657344 | 0.764682058 |
| UBE2O | 0.771620549 | 1.045425049 |
| C2ORF27 | 0.77160579 | 0.959575197 |
| PHKA1 | 0.77157593 | 0.817001947 |
| CHD4 | 0.771506285 | 0.609344581 |
| SPDYA | 0.771461224 | 0.869177805 |
| SAYSD1 | 0.771397049 | 0.971908188 |
| CEMIP | 0.771341907 | 0.863309353 |
| VPS13C | 0.771337784 | 0.895512162 |
| FOSB | 0.77127733 | 1.035236938 |
| GGTLC2 | 0.771146038 | 0.789746918 |
| SLMO1 | 0.771133577 | 0.909680032 |
| LONP1 | 0.771071536 | 0.942245295 |
| WRN | 0.770958495 | 0.797488862 |
| ADRBK1 | 0.77079707 | 1.018818949 |
| MCTP1 | 0.770719943 | 0.795058728 |
| DPM2 | 0.770676898 | 0.900308325 |
| HMGB1 | 0.770487535 | 1.033095393 |
| ENTPD8 | 0.770376096 | 0.822195221 |
| AMACR | 0.770314866 | 0.913325233 |
| DGCR2 | 0.770311023 | 0.324426173 |
| URB1 | 0.770193717 | 0.894826996 |
| MFNG | 0.770129801 | 0.924260834 |
| PRB2 | 0.77010713 | 0.79616307 |
| SLC25A6 | 0.770073817 | 1.002025111 |
| ARHGAP30 | 0.770025934 | 0.801134062 |
| GZMB | 0.770019527 | 0.853341986 |
| TRPM2 | 0.769929465 | 0.902011681 |
| IFITM3 | 0.769879857 | 0.931483385 |
| TMEM211 | 0.769756117 | 1.010277492 |
| LILRB3 | 0.769743012 | 0.932766302 |
| BRMS1L | 0.769637723 | 0.893456663 |
| TSKS | 0.769481709 | 0.944192083 |
| TNXB | 0.769474465 | 1.013627515 |
| STOML3 | 0.769339903 | 0.85748544 |
| TXNRD1 | 0.769157098 | 0.938841636 |
| SLC5A11 | 0.769140135 | 0.839611179 |
| KCNC1 | 0.769109773 | 0.93770279 |
| PKNOX1 | 0.769041413 | 0.87727825 |
| BARHL1 | 0.768952823 | 0.900308325 |
| TH | 0.768700544 | 0.813108371 |
| SPEG | 0.768632792 | 1.007787151 |

| | | |
|---|---|---|
| RBM45 | 0.768584999 | 0.924974306 |
| TNFRSF19 | 0.768421798 | 0.994159637 |
| HBP1 | 0.768362031 | 0.979787598 |
| C1QL4 | 0.768360597 | 1.066018631 |
| ADAMTS2 | 0.768350094 | 0.68137573 |
| CLGN | 0.768350094 | 0.9733939 |
| MEIS1 | 0.768350094 | 0.9733939 |
| HOXB5 | 0.768278074 | 0.741190765 |
| CEP57 | 0.768252757 | 1.068853787 |
| TRNAU1AP | 0.768218118 | 0.676792224 |
| VPS72 | 0.768201307 | 0.880923451 |
| PTCD1 | 0.768112466 | 0.858855773 |
| PLCB3 | 0.767920532 | 0.994808566 |
| EPS15 | 0.767904087 | 0.954093868 |
| SSX5 | 0.767876675 | 0.956149366 |
| CD5 | 0.767696777 | 0.981181051 |
| PSMA4 | 0.767628449 | 1.085658663 |
| NPY1R | 0.767608527 | 0.960415315 |
| ANKRD7 | 0.767470718 | 0.890979883 |
| RGS20 | 0.767428952 | 0.86836776 |
| NAT8L | 0.767376433 | 0.787768327 |
| CYP7B1 | 0.767352033 | 0.825435399 |
| MKRN2 | 0.767284502 | 1.002595717 |
| GPIHBP1 | 0.767260102 | 0.940733128 |
| TEX264 | 0.767259118 | 0.849736736 |
| COMMD6 | 0.767210397 | 0.38643371 |
| PROX2 | 0.767187496 | 0.769137303 |
| KCNJ14 | 0.767150892 | 1.009733939 |
| BASP1 | 0.767088315 | 0.898339409 |
| NF1 | 0.767088315 | 0.898339409 |
| RPUSD1 | 0.767077058 | 0.658971243 |
| ZNF830 | 0.767069213 | 0.973673552 |
| PRKY | 0.767053668 | 0.988968202 |
| ATP2B1 | 0.766935087 | 0.946942082 |
| CYP8B1 | 0.766841358 | 0.904009721 |
| WIPI2 | 0.766733619 | 0.823410288 |
| TRIM16L | 0.766694701 | 0.75617659 |
| RUNX1T1 | 0.766619921 | 0.901579587 |
| MYO10 | 0.766506833 | 0.729445119 |
| SP3 | 0.766476677 | 0.912515188 |
| GAR1 | 0.766438631 | 0.968851395 |
| ARX | 0.766428233 | 1.020251114 |
| SLC25A42 | 0.766379024 | 0.815714864 |
| SKAP1 | 0.766372361 | 0.864373783 |

| | | |
|---|---|---|
| RGS2 | 0.766303072 | 0.905447071 |
| C2orf61 | 0.76624195 | 0.980472765 |
| ABLIM2 | 0.766153279 | 0.917780062 |
| C19orf47 | 0.766147145 | 0.958687728 |
| ZER1 | 0.766035891 | 0.853786958 |
| MAATS1 | 0.765926319 | 0.848521669 |
| RBKS | 0.765925675 | 0.969500324 |
| BTBD10 | 0.765912339 | 1.081116158 |
| LSM6 | 0.765770367 | 0.853374443 |
| EFNA2 | 0.765677273 | 1.048197651 |
| CD209 | 0.765672637 | 0.854191981 |
| RELT | 0.765671214 | 0.983776768 |
| CRIM1 | 0.765663558 | 0.927968851 |
| CASP1 | 0.765645282 | 0.816353018 |
| OR10H5 | 0.765599913 | 0.892086331 |
| SPATA2 | 0.765599913 | 0.892086331 |
| CIAO1 | 0.765574934 | 0.910895099 |
| TAS2R38 | 0.765549209 | 0.92709599 |
| TRABD | 0.765520275 | 0.873228028 |
| APAF1 | 0.76550558 | 0.851395198 |
| LSM2 | 0.765487907 | 0.851661528 |
| STX8 | 0.765364756 | 0.76053443 |
| PTCH2 | 0.765362972 | 0.90036452 |
| SLC35E4 | 0.765333039 | 0.991778006 |
| TMEM50A | 0.765206353 | 0.927029805 |
| DHX8 | 0.76516649 | 0.984425698 |
| RNF222 | 0.765113063 | 0.726151849 |
| TGIF2 | 0.765071084 | 0.852166869 |
| CCDC142 | 0.765028091 | 1.003240178 |
| TIGD5 | 0.764989728 | 0.894826996 |
| PSMD7 | 0.764971955 | 0.830700689 |
| AK4 | 0.764967255 | 0.870503597 |
| SSMEM1 | 0.764950254 | 0.973673552 |
| SLC7A10 | 0.764922567 | 0.852044127 |
| C7orf69 | 0.764916508 | 0.795477903 |
| MT4 | 0.764896119 | 0.850291196 |
| RARS | 0.76489473 | 0.911705144 |
| HEY2 | 0.764764221 | 0.779667882 |
| INPPL1 | 0.764723262 | 1.040882544 |
| FAAH | 0.76471964 | 0.922128488 |
| IL12RB1 | 0.764685467 | 0.884973674 |
| ZNF48 | 0.764632184 | 0.974078574 |
| OR52W1 | 0.764631737 | 0.901336074 |
| SMARCA2 | 0.764619296 | 0.998379911 |

| | | |
|---|---|---|
| C3orf38 | 0.764567439 | 0.887808829 |
| CYSLTR1 | 0.764429945 | 0.726800779 |
| SAPCD1 | 0.764415701 | 0.950325454 |
| PPIG | 0.764394052 | 1.06229721 |
| HTRA4 | 0.764351656 | 0.828675577 |
| HSPG2 | 0.764116612 | 0.92428914 |
| PURA | 0.76404566 | 0.992861778 |
| NDUFA8 | 0.764006435 | 1.072093965 |
| DENND2D | 0.763870475 | 0.888618874 |
| CDYL | 0.763866196 | 0.872417983 |
| DUX1 | 0.763789633 | 0.988659376 |
| ALOX12B | 0.763787035 | 0.980558931 |
| BDH1 | 0.763676037 | 0.967193196 |
| PPT2 | 0.763656271 | 0.986372485 |
| CCDC17 | 0.76345786 | 0.799918996 |
| NOS2 | 0.763346861 | 0.818955043 |
| PARP12 | 0.763323785 | 0.92709599 |
| ALDH3A1 | 0.763308949 | 0.748864374 |
| PHLPP1 | 0.763299153 | 0.944841012 |
| ANKRD50 | 0.763245083 | 0.962332928 |
| FAM222A | 0.763235353 | 0.713649251 |
| ADAMTS13 | 0.763223086 | 0.916937054 |
| POC5 | 0.763186807 | 1.013770757 |
| SLC35A2 | 0.763183191 | 0.897529364 |
| HNRNPDL | 0.763165202 | 0.872559096 |
| ETNK2 | 0.763154182 | 0.987021415 |
| EPHA2 | 0.763142327 | 0.889033095 |
| GATA4 | 0.763062106 | 0.892264075 |
| YWHAE | 0.763024852 | 0.896171317 |
| NME7 | 0.762995709 | 1.064243997 |
| TEDDM1 | 0.762990433 | 0.927501013 |
| PRAMEF2 | 0.762918047 | 0.870797894 |
| COA1 | 0.762858854 | 0.746456055 |
| MROH8 | 0.762834351 | 1.011305242 |
| MYH9 | 0.762787487 | 0.94127177 |
| BCL2L10 | 0.762610012 | 0.760226812 |
| CCDC57 | 0.762548883 | 0.863102471 |
| LRRC47 | 0.762540071 | 0.990279465 |
| C15orf39 | 0.762509354 | 0.906439854 |
| ZNF174 | 0.762473441 | 0.949777238 |
| NECAP2 | 0.762405042 | 0.928400137 |
| SLC4A8 | 0.762361573 | 0.903958469 |
| OR1Q1 | 0.762303322 | 0.507022953 |
| FXN | 0.762252084 | 0.946138871 |

| | | |
|---|---|---|
| EDEM3 | 0.762177937 | 0.803969218 |
| SLC35B4 | 0.762172106 | 0.959232614 |
| CIB4 | 0.762151397 | 0.898744431 |
| RERG | 0.762097095 | 0.874443094 |
| UBE2NL | 0.762046111 | 0.960415315 |
| SEMA4A | 0.761974284 | 0.83521754 |
| FBXO45 | 0.761961153 | 0.680077872 |
| ZNF664 | 0.761916818 | 0.99108951 |
| ZNF217 | 0.761789965 | 0.780404248 |
| NXPE1 | 0.761722277 | 1.040097205 |
| ZNF684 | 0.761665326 | 0.677197246 |
| CR2 | 0.761514714 | 1.073329007 |
| TMEM216 | 0.761431001 | 0.926690968 |
| CAMK1 | 0.761428028 | 1.080467229 |
| ATP6V1E1 | 0.761426722 | 0.848116646 |
| VCX | 0.761388023 | 0.996916752 |
| CDH18 | 0.761349737 | 1.003083248 |
| NIPA1 | 0.761314399 | 0.850951802 |
| PRICKLE1 | 0.761305202 | 0.940298507 |
| BCL2L2 | 0.761175078 | 0.986634265 |
| FGF5 | 0.761145533 | 0.802349129 |
| ANKRA2 | 0.761124341 | 0.935196436 |
| TSR1 | 0.761101113 | 0.962332928 |
| GNPNAT1 | 0.761067747 | 0.883758607 |
| DTX2 | 0.760780227 | 0.940947437 |
| HIF3A | 0.760763889 | 0.865127582 |
| GOT1 | 0.760752733 | 0.954637505 |
| CHST12 | 0.760746416 | 0.829890644 |
| MYBL2 | 0.76065722 | 0.778452815 |
| CDC42EP3 | 0.760596827 | 0.904076739 |
| TJP2 | 0.760594999 | 0.955223881 |
| LIMS1 | 0.760541096 | 0.865364851 |
| HDDC3 | 0.76049141 | 0.971243418 |
| PIAS4 | 0.760487233 | 0.82746051 |
| LMO4 | 0.760463776 | 0.88513952 |
| TF | 0.760452425 | 0.92874272 |
| LDLRAD3 | 0.760375317 | 0.830295666 |
| MTMR12 | 0.760368746 | 0.911705144 |
| EPOR | 0.760334259 | 0.930741191 |
| R3HDML | 0.760320439 | 0.902363823 |
| CCDC183 | 0.760276579 | 0.859540939 |
| HPR | 0.760222816 | 0.780012979 |
| PLEKHH3 | 0.760011961 | 0.958282706 |
| H3F3B | 0.759969833 | 0.968139774 |

| | | |
|---|---|---|
| CALML4 | 0.759910029 | 0.787363305 |
| UQCRFS1 | 0.759896072 | 0.760226812 |
| RPP38 | 0.759817115 | 0.890643985 |
| RP11-94I2.2 | 0.759743506 | 0.828270555 |
| IRS1 | 0.759701056 | 0.968482357 |
| GPR107 | 0.759514672 | 1.048020766 |
| CLEC4D | 0.75944571 | 0.863912515 |
| TAP1 | 0.759423092 | 0.771567436 |
| CXXC1 | 0.759419835 | 1.012978585 |
| SERPINB6 | 0.75932544 | 0.820575132 |
| EGLN3 | 0.75930486 | 0.989064399 |
| UNC13B | 0.759251359 | 0.885234669 |
| FAM103A1 | 0.759233187 | 1.038072094 |
| TFAM | 0.759174351 | 0.983799109 |
| C8B | 0.75915281 | 1.036656389 |
| RNF121 | 0.759057366 | 0.774172615 |
| LMBRD1 | 0.759050567 | 0.837181045 |
| TBC1D8B | 0.758950877 | 0.820980154 |
| DDX4 | 0.758909055 | 0.97326853 |
| HOXD4 | 0.758909055 | 0.97326853 |
| DCT | 0.758858522 | 1.003240178 |
| ADCY4 | 0.758779871 | 0.910895099 |
| ASPA | 0.758734228 | 0.86472256 |
| SPRR4 | 0.758638766 | 0.981773998 |
| NTSR2 | 0.75853379 | 0.957819598 |
| EXOC3L4 | 0.758513606 | 1.033616849 |
| KCND3 | 0.75837787 | 1.021414666 |
| CALU | 0.758363472 | 0.888317917 |
| XAGE5 | 0.758347338 | 0.914135277 |
| INO80E | 0.758289979 | 0.780882949 |
| KRCC1 | 0.758158502 | 0.919805589 |
| SFTA2 | 0.758112147 | 0.998784933 |
| NET1 | 0.758104527 | 0.966426859 |
| LENEP | 0.75807784 | 0.886605002 |
| RTN4R | 0.758051019 | 0.971648441 |
| OR2H2 | 0.757848582 | 0.841231268 |
| ZNF417 | 0.757807779 | 0.852166869 |
| ABCB9 | 0.757787036 | 0.930564568 |
| PCDHA11 | 0.75768555 | 0.921891059 |
| EVI2A | 0.757562307 | 0.969167523 |
| ZC3H7A | 0.757510391 | 0.860672337 |
| PRKAG2 | 0.757507384 | 0.959117456 |
| CARD6 | 0.757288909 | 0.850633779 |
| EPHA7 | 0.757218348 | 0.825438027 |

| | | |
|---|---|---|
| IFNL2 | 0.757127788 | 1.051730045 |
| C3orf20 | 0.757107607 | 1.002835156 |
| PAM16 | 0.757033998 | 0.904819765 |
| WFS1 | 0.757012501 | 0.945727015 |
| TPRKB | 0.756982548 | 0.825840421 |
| ERBB2 | 0.756935238 | 1.023361454 |
| SNTG2 | 0.756870786 | 1.041312272 |
| DPH5 | 0.756766664 | 0.801539085 |
| CST4 | 0.756766626 | 0.966084275 |
| SLC34A2 | 0.756720003 | 0.782503038 |
| JUNB | 0.756700187 | 0.752531389 |
| SLC7A6OS | 0.756684192 | 0.834345889 |
| MFHAS1 | 0.756642828 | 0.774402592 |
| DHRS7 | 0.75660578 | 0.946138871 |
| MOV10 | 0.756512097 | 1.016872161 |
| NCR1 | 0.756483695 | 0.960415315 |
| BCORL1 | 0.756322773 | 1.112367249 |
| SLC4A4 | 0.756317015 | 1.020251114 |
| NUDT9 | 0.756307276 | 0.981829981 |
| STAMBP | 0.756250144 | 0.988968202 |
| KDF1 | 0.756162921 | 0.81855002 |
| RAB4A | 0.756124054 | 0.840421223 |
| DUS4L | 0.756122831 | 0.788578372 |
| PYCR2 | 0.756101715 | 0.756002596 |
| FKBP6 | 0.756087226 | 0.9467878 |
| CAMK2B | 0.755958884 | 0.77027904 |
| UBE2D2 | 0.755923979 | 1.031797534 |
| PPP2CB | 0.755916982 | 0.968202466 |
| ALDH1A2 | 0.755873377 | 1.015390846 |
| TXNDC16 | 0.755869047 | 0.764277035 |
| FAM86C1 | 0.755819737 | 0.79708384 |
| TSNAX | 0.755800988 | 0.807125728 |
| AMIGO1 | 0.75579498 | 1.018226002 |
| GNG2 | 0.755755497 | 0.868790682 |
| GTF2H4 | 0.755749643 | 0.778047793 |
| CCDC120 | 0.755692132 | 0.821790198 |
| CNPY3 | 0.755684723 | 0.985954094 |
| FAM122C | 0.755604973 | 0.898339409 |
| BRAP | 0.755536237 | 0.883841661 |
| FAM187B | 0.755442219 | 0.988659376 |
| OR5C1 | 0.755420659 | 0.992463172 |
| CCDC94 | 0.755386644 | 0.958687728 |
| CYP26A1 | 0.755328674 | 0.811259619 |
| GJC2 | 0.755185328 | 0.740380721 |

| | | |
|---|---|---|
| FAM131A | 0.755113197 | 0.68853787 |
| CFHR2 | 0.754990686 | 0.950992305 |
| PLA2G1B | 0.754990686 | 0.950992305 |
| ACHE | 0.754924209 | 0.899149453 |
| DSG1 | 0.754870736 | 0.894484412 |
| CSGALNACT2 | 0.754864812 | 0.781692993 |
| OTUB1 | 0.754847391 | 0.858242203 |
| RBM26 | 0.754847391 | 0.858242203 |
| HSD17B4 | 0.754842959 | 0.880113406 |
| RALA | 0.754838731 | 0.901984609 |
| RABGGTA | 0.754764292 | 0.99091499 |
| C1orf226 | 0.754750808 | 0.937626569 |
| FIGF | 0.754629611 | 0.77871512 |
| GSTM5 | 0.754582275 | 0.941596366 |
| PRKAR1A | 0.754527284 | 0.870863076 |
| SPIN2A | 0.754484238 | 0.825840421 |
| ZBTB37 | 0.754430446 | 0.959902795 |
| PSEN1 | 0.754421989 | 0.92709599 |
| KRTDAP | 0.754369159 | 0.981500514 |
| EIF1 | 0.754306508 | 0.842446335 |
| TNNI3K | 0.75422845 | 0.998702141 |
| EMR3 | 0.75415971 | 0.757949384 |
| EXOSC8 | 0.754105871 | 1.020116807 |
| DIS3L | 0.754085682 | 0.916565411 |
| ARMC5 | 0.754064033 | 0.883758607 |
| MYL1 | 0.754017511 | 0.905789654 |
| AOC1 | 0.753936614 | 0.963659961 |
| CA1 | 0.753920314 | 0.878650227 |
| SIRPG | 0.753877999 | 0.675982179 |
| ZNF233 | 0.753853898 | 0.990407674 |
| OR1S2 | 0.753757887 | 0.922576225 |
| NSUN5P2 | 0.753752636 | 0.916970433 |
| GAREM | 0.753730222 | 0.89509923 |
| DNAJC22 | 0.753689017 | 0.879410757 |
| MOGS | 0.753618435 | 0.836469825 |
| TLE1 | 0.753605215 | 0.933576347 |
| PAG1 | 0.753581884 | 0.843256379 |
| CYFIP2 | 0.753489201 | 0.929085303 |
| TOMM6 | 0.753478055 | 0.922918808 |
| SECISBP2 | 0.753466796 | 0.957170668 |
| DR1 | 0.753323617 | 0.851761847 |
| SUV39H1 | 0.753256733 | 0.865532604 |
| CKAP4 | 0.753227799 | 0.901174565 |
| LDHAL6B | 0.753218018 | 0.947752126 |

| | | |
|---|---|---|
| EMX2 | 0.753164582 | 0.66018631 |
| ACYP2 | 0.752956309 | 0.851395198 |
| INTS2 | 0.752947883 | 0.887808829 |
| MMP25 | 0.752824263 | 0.986372485 |
| CHCHD10 | 0.752798955 | 0.767043508 |
| HIBCH | 0.752774482 | 0.84690158 |
| TFAP2E | 0.752684216 | 0.885378696 |
| ETFDH | 0.752625205 | 0.688513952 |
| REP15 | 0.752513958 | 0.9777321 |
| CCNO | 0.752446564 | 0.958468527 |
| BCL6B | 0.752378983 | 0.80072904 |
| FAM63A | 0.752347247 | 0.789793439 |
| OR8J3 | 0.752310246 | 0.907845152 |
| RARRES1 | 0.752189178 | 0.87084618 |
| FXYD3 | 0.752063843 | 1.058403634 |
| NXF5 | 0.751909518 | 0.913730255 |
| RBP3 | 0.751898374 | 0.831278391 |
| CYP20A1 | 0.751884557 | 0.987849332 |
| DNMT3L | 0.751884557 | 0.878088295 |
| RPE65 | 0.751868155 | 0.927501013 |
| MNS1 | 0.75181107 | 0.927029805 |
| MRPS12 | 0.75176789 | 1.220280918 |
| FLT1 | 0.751714957 | 1.023361454 |
| MON1B | 0.751683547 | 0.946942082 |
| BID | 0.751616005 | 0.753406879 |
| LRRC45 | 0.75150979 | 1.015795869 |
| ELANE | 0.751498365 | 0.93835172 |
| C12orf50 | 0.751386923 | 1.070878898 |
| CFHR5 | 0.751323553 | 0.936411503 |
| EMC6 | 0.751168644 | 0.829485622 |
| TMEM133 | 0.751156113 | 0.804779263 |
| FBXO27 | 0.751097204 | 0.718364698 |
| OR2A42 | 0.751075378 | 1.076738609 |
| UPK3B | 0.751055759 | 0.882494005 |
| RBM33 | 0.751041039 | 0.758201701 |
| GPATCH4 | 0.750960859 | 0.824220332 |
| CDX1 | 0.750852306 | 0.840826245 |
| ALB | 0.750807122 | 0.791413528 |
| OFCC1 | 0.750730798 | 0.934566632 |
| UGT1A1 | 0.750726256 | 0.832576249 |
| CYP2J2 | 0.750714236 | 1.046073978 |
| CUL2 | 0.750698595 | 1.0532122 |
| LYPD6B | 0.750557107 | 0.929121102 |
| RFPL4B | 0.750492211 | 0.925373134 |

| | | |
|---|---|---|
| OR4C6 | 0.750478608 | 0.893479141 |
| IFT52 | 0.75047005 | 0.764277035 |
| KIF23 | 0.750460351 | 0.939649578 |
| CNTN5 | 0.750454093 | 1.0445358 |
| TROAP | 0.750429604 | 0.97326853 |
| PRKAB2 | 0.750384696 | 0.975340688 |
| MACROD2 | 0.750265831 | 1.006480356 |
| FGFBP2 | 0.75024464 | 0.863507493 |
| SVEP1 | 0.750209899 | 0.891049008 |
| TAF1D | 0.750205416 | 0.838801134 |
| MAP6 | 0.750190074 | 0.989037342 |
| SGK494 | 0.750156661 | 0.826086957 |
| GJA3 | 0.750126695 | 0.8403634 |
| ATXN7L2 | 0.750082447 | 0.882948562 |
| INS | 0.750022791 | 0.90647482 |
| SLC24A5 | 0.750014544 | 0.854744776 |
| RBM12 | 0.749987585 | 0.900308325 |
| SERPINA6 | 0.74990329 | 0.929931146 |
| MZF1 | 0.749880775 | 0.877683273 |
| PLCB4 | 0.749823005 | 1.018818949 |
| LTK | 0.749793616 | 1.040233615 |
| TMEM198 | 0.749518404 | 0.321000343 |
| PTGER2 | 0.749457572 | 0.912394549 |
| OR2H1 | 0.749430368 | 0.903048989 |
| PXN | 0.749382631 | 0.693803159 |
| DPP3 | 0.749380271 | 0.983776768 |
| MC3R | 0.749380271 | 0.983776768 |
| CCDC88A | 0.749370106 | 0.938335046 |
| NOTCH3 | 0.749052691 | 0.897934386 |
| AFF1 | 0.749037409 | 0.732685298 |
| PCK1 | 0.748942064 | 0.884490591 |
| GJB2 | 0.748924859 | 0.913043478 |
| PIK3R2 | 0.748908806 | 0.941596366 |
| PAK7 | 0.74889728 | 0.963011032 |
| ARHGEF18 | 0.748872419 | 1.012978585 |
| CALML3 | 0.748827388 | 0.980472765 |
| DOK1 | 0.7488007 | 0.976361768 |
| UGT2B11 | 0.748770686 | 0.867962738 |
| LXN | 0.748666992 | 0.916409729 |
| OTUD7A | 0.74853575 | 0.934386391 |
| CXorf40B | 0.748450095 | 0.848235697 |
| SETD6 | 0.748410183 | 0.769137303 |
| CDK1 | 0.748392968 | 0.992212849 |
| SENP7 | 0.748392968 | 0.906554186 |

| | | |
|---|---|---|
| C8orf33 | 0.74830005 | 0.887808829 |
| LYPD4 | 0.748273765 | 1.0036452 |
| ZNF596 | 0.748261081 | 0.9181855 |
| TMEM176A | 0.74823994 | 0.689347914 |
| KLHDC7B | 0.748154565 | 0.879708384 |
| MAGEC3 | 0.748118836 | 0.881808839 |
| NKX3-1 | 0.748109534 | 0.780477926 |
| NCMAP | 0.748072551 | 0.868772783 |
| MTFP1 | 0.74800261 | 1.031591738 |
| OTC | 0.747910801 | 1.007138222 |
| HK1 | 0.747881823 | 0.950032446 |
| DDIT3 | 0.74787392 | 0.935756003 |
| LCN9 | 0.747870051 | 0.928617781 |
| PPP2R5C | 0.747861819 | 0.914341337 |
| TAAR1 | 0.747835722 | 0.871512005 |
| CLUL1 | 0.747772172 | 0.805412813 |
| INTS1 | 0.747416747 | 0.748076144 |
| LONRF2 | 0.747401056 | 0.979234263 |
| RAB34 | 0.747390802 | 0.998972251 |
| F12 | 0.747386402 | 0.96495782 |
| CDPF1 | 0.747187724 | 0.941075711 |
| ST6GALNAC6 | 0.747187516 | 0.847711624 |
| GPR182 | 0.747146677 | 0.779364049 |
| SRP68 | 0.747135799 | 0.870503597 |
| COL4A3BP | 0.747074233 | 0.736534718 |
| SYNE4 | 0.746984265 | 0.994329688 |
| RPS6KA3 | 0.74692745 | 0.994159637 |
| TMEM52B | 0.746898787 | 0.737545565 |
| CDK13 | 0.746895179 | 0.972744971 |
| RPL36AL | 0.746880298 | 0.906132237 |
| YPEL2 | 0.746875966 | 0.892264075 |
| FOLR1 | 0.746789229 | 0.908500973 |
| CRK | 0.746754983 | 0.897934386 |
| TPD52L3 | 0.746695875 | 0.85853342 |
| IGFL1 | 0.746623201 | 0.826245443 |
| CACUL1 | 0.74662236 | 0.82284231 |
| SYCE1L | 0.746620176 | 0.848235697 |
| AAK1 | 0.746520856 | 1.037637897 |
| PHF6 | 0.746454092 | 1.001946788 |
| MRPS10 | 0.746372604 | 0.94210346 |
| PHF11 | 0.746336946 | 0.944841012 |
| KRTAP10-8 | 0.746257349 | 0.953827461 |
| MAGEB4 | 0.746182485 | 0.979787598 |
| XCL2 | 0.746075328 | 0.911270983 |

| | | |
|---|---|---|
| TREM1 | 0.74607417 | 0.837767683 |
| PSPH | 0.746038112 | 1.031148605 |
| GAB2 | 0.745985838 | 1.064803564 |
| ORAI2 | 0.745876316 | 1.103685703 |
| RAF1 | 0.745805138 | 0.93835172 |
| TBC1D3B | 0.745532164 | 0.761036857 |
| CNDP1 | 0.745529272 | 1.01752109 |
| ZNF300 | 0.74549609 | 0.936964714 |
| PM20D1 | 0.745471893 | 0.831278391 |
| TAS2R41 | 0.745414798 | 0.982584042 |
| CORT | 0.745373845 | 0.951695786 |
| ADH5 | 0.745252255 | 1.082414017 |
| SGPL1 | 0.7452418 | 0.924724205 |
| SSTR4 | 0.7452418 | 0.924724205 |
| CKS2 | 0.74521027 | 1.068137573 |
| SYT17 | 0.745155818 | 0.811259619 |
| FAM159A | 0.745091158 | 0.924974306 |
| CEP135 | 0.745087406 | 0.797488862 |
| NUDCD2 | 0.745043444 | 0.595382746 |
| MSH4 | 0.744978283 | 0.853341986 |
| NAA60 | 0.744954197 | 0.78655326 |
| CHRDL1 | 0.744947754 | 0.962658445 |
| DGKB | 0.744932758 | 0.98247891 |
| RAB3A | 0.744907485 | 0.975340688 |
| EIF2B1 | 0.744829196 | 0.953926022 |
| PKN3 | 0.744775109 | 0.939649578 |
| WRAP53 | 0.744662361 | 0.939246659 |
| GMPPB | 0.744632196 | 1.033095393 |
| NDRG1 | 0.744632196 | 0.903958469 |
| HMGA2 | 0.744371959 | 0.863309353 |
| STK36 | 0.744135376 | 1.0266061 |
| HCAR1 | 0.744098001 | 0.904607398 |
| PCDH10 | 0.744082975 | 1.074340528 |
| TFDP1 | 0.744070028 | 0.829080599 |
| LIPE | 0.744042674 | 0.848521669 |
| RLF | 0.74398832 | 0.851356825 |
| MCMDC2 | 0.7439818 | 1.051032807 |
| ASZ1 | 0.7439764 | 0.743215877 |
| NAGLU | 0.743853329 | 0.748886189 |
| RNFT1 | 0.743769696 | 0.8403634 |
| PDK1 | 0.743761875 | 1.048669695 |
| ZNF343 | 0.743687672 | 0.848926691 |
| CISD1 | 0.743652217 | 0.832320778 |
| SHARPIN | 0.743585521 | 0.916409729 |

| | | |
|---|---|---|
| SKA1 | 0.743585521 | 0.916409729 |
| HOXB13 | 0.743564629 | 0.905256327 |
| ACTL7B | 0.743542114 | 0.935251799 |
| ANGPT1 | 0.74350556 | 1.02673147 |
| CD180 | 0.74350556 | 0.821385176 |
| NLRP6 | 0.743432054 | 1.162818955 |
| POLR2G | 0.743423527 | 0.977287476 |
| PRR16 | 0.743343723 | 0.829890644 |
| KLHL23 | 0.743200123 | 0.799513973 |
| PAK3 | 0.743195821 | 1.121349773 |
| GBP1 | 0.743072314 | 0.882948562 |
| OR4F15 | 0.742991466 | 1.084275437 |
| OMG | 0.742989354 | 0.898767034 |
| PIGO | 0.742989354 | 0.898767034 |
| TFB2M | 0.742929172 | 0.833130822 |
| DAOA | 0.742926991 | 0.867077766 |
| PTPRM | 0.742890072 | 0.970798183 |
| TTLL4 | 0.74287757 | 0.79708384 |
| SETD3 | 0.742775435 | 0.84163629 |
| CCDC104 | 0.742728315 | 0.863912515 |
| SYNPO2 | 0.742685853 | 0.808838643 |
| OXCT1 | 0.742467858 | 0.839206156 |
| DEAF1 | 0.742436696 | 1.036998972 |
| MROH2B | 0.742379062 | 0.951353203 |
| FANCB | 0.742351332 | 0.949297705 |
| SPN | 0.742344328 | 0.80599433 |
| KCTD20 | 0.742308459 | 0.878001298 |
| CDC25C | 0.742128253 | 1.007787151 |
| RPL5 | 0.742126733 | 1.012332991 |
| SERTAD1 | 0.742068569 | 0.921479559 |
| GPX6 | 0.742018769 | 0.700688538 |
| STC2 | 0.741911512 | 1.054470709 |
| UGT1A3 | 0.741847254 | 0.959497772 |
| RARRES2 | 0.74160583 | 1.073329007 |
| ANO2 | 0.741496072 | 1.02673147 |
| SMG9 | 0.741427914 | 0.859862292 |
| CA14 | 0.741420356 | 0.835820896 |
| TCF7L1 | 0.741359298 | 0.820980154 |
| WDR65 | 0.741338107 | 0.865532604 |
| GPR176 | 0.741191909 | 0.944192083 |
| C1orf174 | 0.741123037 | 0.857432159 |
| TTC21A | 0.741077634 | 0.881808839 |
| PIGR | 0.741023296 | 0.996759822 |
| FAM216A | 0.740999253 | 0.732280275 |

| | | |
|---|---|---|
| ISYNA1 | 0.740967413 | 0.915639195 |
| MYH4 | 0.740948058 | 0.801944107 |
| SERGEF | 0.740948058 | 0.957877683 |
| SLC22A18 | 0.740905013 | 0.871607938 |
| RSPO3 | 0.740727283 | 1.019441069 |
| CSF2RA | 0.740714105 | 0.822193381 |
| SLC14A1 | 0.74062744 | 0.93600648 |
| PCMTD2 | 0.740409518 | 0.766302147 |
| ABI1 | 0.740269447 | 0.972935937 |
| PLD5 | 0.740257818 | 0.869582827 |
| SERINC1 | 0.740176569 | 0.875253139 |
| OR4S1 | 0.740135829 | 0.981500514 |
| NAPSA | 0.739984062 | 0.924075276 |
| CCRN4L | 0.739922484 | 0.974691759 |
| PDIK1L | 0.73991417 | 0.859182349 |
| YIF1A | 0.73981362 | 0.925880923 |
| NFXL1 | 0.739747856 | 0.906439854 |
| ALX4 | 0.739732153 | 0.953277093 |
| MECOM | 0.739642503 | 1.054510058 |
| TACO1 | 0.739622187 | 0.792223572 |
| CCDC113 | 0.739603288 | 0.892669097 |
| RFPL2 | 0.739527498 | 0.657365347 |
| DENND3 | 0.739463752 | 0.890030832 |
| ACSL3 | 0.739338399 | 0.862292426 |
| ACVR1 | 0.739253114 | 0.903309539 |
| PGM1 | 0.73920726 | 1.004542505 |
| CHKA | 0.739141566 | 0.997404283 |
| SERPINE2 | 0.739104492 | 0.934566632 |
| SHC1 | 0.73908273 | 0.985419198 |
| FANCM | 0.739061383 | 1.040882544 |
| EMC4 | 0.738991293 | 0.862697448 |
| KIAA0922 | 0.738969651 | 0.894826996 |
| PIK3R4 | 0.738882001 | 0.817001947 |
| GFRA4 | 0.738765933 | 0.773592548 |
| SLC25A14 | 0.738765933 | 0.773592548 |
| MAN2A1 | 0.738722376 | 0.903958469 |
| PCBP1 | 0.738715808 | 0.991494532 |
| MPPED1 | 0.738694566 | 0.832320778 |
| METRN | 0.738688822 | 0.926687222 |
| NR2F1 | 0.738653637 | 0.947436729 |
| FOXJ2 | 0.738579628 | 0.940298507 |
| C8orf4 | 0.738518911 | 0.901678657 |
| ZNF705A | 0.738403341 | 0.855002025 |
| GPR160 | 0.738374396 | 0.969500324 |

| | | |
|---|---|---|
| PRDM16 | 0.738374396 | 0.969500324 |
| ACOX3 | 0.738139064 | 0.961522884 |
| STK11 | 0.738116148 | 0.803374432 |
| PEPD | 0.738107422 | 0.99156392 |
| CCL14 | 0.738103762 | 0.936411503 |
| MAPK9 | 0.738070294 | 0.846852693 |
| SLC26A2 | 0.738066576 | 0.911300122 |
| SCTR | 0.738028886 | 0.890330954 |
| H2AFY2 | 0.73798462 | 0.933881466 |
| ASAP3 | 0.737964095 | 0.887632751 |
| DSC3 | 0.737888881 | 0.868790682 |
| C19orf10 | 0.737815186 | 0.933981369 |
| USP29 | 0.737802497 | 0.963011032 |
| DRAM1 | 0.737690128 | 0.886593763 |
| MBOAT1 | 0.737617742 | 0.833535844 |
| MPDU1 | 0.737518656 | 0.909215485 |
| C1orf87 | 0.7374502 | 1.029566626 |
| OR1I1 | 0.737431861 | 0.970838396 |
| MYH10 | 0.737407512 | 0.947242206 |
| ISG20L2 | 0.737403394 | 0.883841661 |
| NBPF8 | 0.737338279 | 0.972593354 |
| ZEB1 | 0.73729203 | 0.738760632 |
| CLK2 | 0.737222941 | 1.043478261 |
| ARHGAP44 | 0.737212816 | 0.936964714 |
| MYH6 | 0.737201662 | 0.901174565 |
| HLA-B | 0.737187348 | 0.778047793 |
| EXOSC1 | 0.737167688 | 1.006509078 |
| ZNF606 | 0.737072311 | 1.029804728 |
| SPTLC3 | 0.737041507 | 0.758606723 |
| SKIV2L | 0.736987102 | 0.890238963 |
| FAM204A | 0.736983187 | 0.939362795 |
| FMO1 | 0.736884407 | 0.93236128 |
| SEPT9 | 0.736855626 | 0.870797894 |
| KLHL35 | 0.736800137 | 0.859540939 |
| COL4A4 | 0.736786227 | 0.844809866 |
| LCA10 | 0.736700971 | 0.688942892 |
| OR6C3 | 0.736688403 | 0.952380952 |
| MRPL12 | 0.736685911 | 0.910243234 |
| B4GALT5 | 0.736679526 | 1.019441069 |
| TNFRSF1B | 0.736514174 | 0.837586067 |
| CCDC135 | 0.736484476 | 0.974888619 |
| DEFB136 | 0.736416953 | 0.855772525 |
| TFF3 | 0.736318088 | 1.043508051 |
| FMO5 | 0.736267333 | 1.001297859 |

| | | |
|---|---|---|
| SHH | 0.736176531 | 0.914341337 |
| FTSJ2 | 0.73613257 | 0.950681376 |
| PLA2R1 | 0.736039874 | 0.943543154 |
| FN3KRP | 0.735945696 | 0.936404932 |
| WISP3 | 0.735913436 | 0.827055488 |
| OGFOD1 | 0.735660009 | 0.964763062 |
| CBLN4 | 0.735634641 | 0.818955043 |
| GABBR1 | 0.735630481 | 0.951330305 |
| DCP1B | 0.735534957 | 0.962315862 |
| RAB40AL | 0.735533998 | 0.755353666 |
| BAG4 | 0.735524253 | 0.906034832 |
| LGALS1 | 0.735511569 | 0.945321993 |
| ATP13A3 | 0.73547028 | 0.864373783 |
| TNIP3 | 0.735460105 | 0.945529291 |
| FASLG | 0.735451613 | 0.9145403 |
| SEC63 | 0.735336587 | 0.929915639 |
| NRIP1 | 0.7353236 | 0.813689753 |
| TCP11L2 | 0.73527969 | 1.052247874 |
| ITGAX | 0.735268585 | 0.858647226 |
| AJUBA | 0.735134737 | 0.981843097 |
| KCNH3 | 0.735028863 | 0.908500973 |
| TRMT5 | 0.734858312 | 0.825435399 |
| CYP4F12 | 0.734851456 | 1.075924724 |
| CAD | 0.734838614 | 1.003244646 |
| TRIM2 | 0.734718916 | 0.923426347 |
| VDR | 0.734642931 | 0.988968202 |
| GREM2 | 0.734629489 | 0.938031592 |
| STK16 | 0.734628278 | 0.952628164 |
| CDKL5 | 0.734576496 | 1.054510058 |
| SEMA4F | 0.73450331 | 1.129202106 |
| FOXS1 | 0.734418847 | 0.93835172 |
| TNFRSF11A | 0.734311745 | 0.931213498 |
| KBTBD2 | 0.734307429 | 1.042137718 |
| HIPK1 | 0.734220613 | 1.06229721 |
| NEUROD1 | 0.73417904 | 0.785853342 |
| ABI3BP | 0.734049029 | 1.017129154 |
| GNB1 | 0.733945515 | 0.851761847 |
| ZNF460 | 0.733776088 | 0.972863507 |
| PXDN | 0.733764321 | 0.584852167 |
| TEX13A | 0.733715368 | 0.933576347 |
| AGTR1 | 0.7336376 | 0.82284231 |
| ATP5H | 0.733628791 | 0.953926022 |
| RRP1 | 0.733607986 | 0.950587282 |
| SEC14L3 | 0.73352103 | 1.012329656 |

| | | |
|---|---|---|
| NFAT5 | 0.733520522 | 0.914135277 |
| DNAJC4 | 0.733495481 | 0.668722165 |
| ITM2C | 0.733275367 | 0.910243234 |
| CLDN14 | 0.733218232 | 0.848920863 |
| SMAP2 | 0.733147538 | 0.75982179 |
| OR13C8 | 0.7331184 | 0.98217902 |
| TMEM87A | 0.732877962 | 0.799513973 |
| PLA2G7 | 0.732848773 | 0.861482382 |
| TRIM66 | 0.732801552 | 0.985419198 |
| SEMA3A | 0.732713574 | 0.968139774 |
| MAT2B | 0.73258979 | 0.860480208 |
| HTATSF1 | 0.732489951 | 0.802754151 |
| PRR7 | 0.732457604 | 0.870392872 |
| HBM | 0.732455108 | 0.746456055 |
| OR8B2 | 0.732374975 | 0.953751285 |
| IL1R1 | 0.732374775 | 0.822600243 |
| POLA2 | 0.732287005 | 0.904607398 |
| ACTN4 | 0.732213084 | 0.992120589 |
| NHS | 0.732161476 | 0.956149366 |
| ADCY1 | 0.732012293 | 0.948734588 |
| METTL1 | 0.732012293 | 0.948734588 |
| RSC1A1 | 0.731923054 | 0.737183647 |
| GDE1 | 0.731913206 | 0.842851357 |
| PDE6C | 0.7319112 | 0.795587281 |
| DCUN1D5 | 0.731892183 | 0.912394549 |
| S100A4 | 0.731833435 | 0.920520726 |
| TNFRSF13C | 0.731830174 | 0.859862292 |
| CXorf40A | 0.731713572 | 0.958687728 |
| SLC6A19 | 0.731691209 | 0.741595788 |
| FBXW4 | 0.731615577 | 0.810512654 |
| FAM73B | 0.731544909 | 0.769947347 |
| WDR82 | 0.731505328 | 0.933576347 |
| FAM126A | 0.731404619 | 0.95909275 |
| ABCA5 | 0.731361473 | 0.72782503 |
| OR2B3 | 0.731355011 | 0.90202124 |
| DFFA | 0.731333303 | 0.733495342 |
| SLC14A2 | 0.731329839 | 0.978938842 |
| SAMD11 | 0.7312681 | 0.995949777 |
| FTSJ1 | 0.731223965 | 1.103179753 |
| CPO | 0.731145017 | 1.154445165 |
| ZNF816 | 0.731136216 | 0.948157149 |
| GRK6 | 0.731122378 | 0.927968851 |
| SNRNP200 | 0.731101551 | 0.898767034 |
| C6orf165 | 0.730961452 | 0.930113052 |

| | | |
|---|---|---|
| CXCL10 | 0.730961452 | 0.930113052 |
| STK4 | 0.730880677 | 0.942894225 |
| RINL | 0.730747542 | 0.875253139 |
| GPR32 | 0.730679562 | 0.877352369 |
| SF3B1 | 0.730408856 | 1.024323398 |
| HN1L | 0.730392234 | 0.991494532 |
| TTC4 | 0.730312935 | 0.822541966 |
| TMEM71 | 0.730279198 | 0.805184285 |
| DHX33 | 0.730191714 | 0.907203115 |
| NOC3L | 0.730190613 | 0.714864318 |
| LRRC73 | 0.730167079 | 0.842041312 |
| HLA-DRB5 | 0.730131787 | 0.899280576 |
| TTC25 | 0.730118533 | 0.859052248 |
| VMAC | 0.73011461 | 0.737545565 |
| NEDD9 | 0.730015175 | 1.025921426 |
| GDPGP1 | 0.729912072 | 1.074119077 |
| BEND7 | 0.729911447 | 0.338129496 |
| STK32B | 0.729876005 | 0.944192083 |
| C11orf57 | 0.729863475 | 0.961117861 |
| RSBN1L | 0.729850893 | 0.966788173 |
| CA6 | 0.729838464 | 0.972458485 |
| ADPRHL2 | 0.729770466 | 0.834523037 |
| MGMT | 0.729721505 | 0.882138518 |
| APMAP | 0.729669267 | 0.974306269 |
| ADCK2 | 0.729637432 | 1.06164828 |
| PPP1CA | 0.729620556 | 0.776119403 |
| EXD1 | 0.72961881 | 0.774807614 |
| EPHX4 | 0.729600471 | 0.938841636 |
| C20orf195 | 0.729550601 | 0.96435804 |
| NUP98 | 0.729521356 | 0.900713822 |
| TTLL9 | 0.729383975 | 0.707168894 |
| LIME1 | 0.729337057 | 0.821514217 |
| MS4A1 | 0.729324681 | 0.916565411 |
| NYAP1 | 0.729323866 | 1.100445525 |
| SIX2 | 0.72931974 | 0.919400567 |
| PSMC3 | 0.729229932 | 0.93770279 |
| VAV1 | 0.729178839 | 0.806399352 |
| RAB41 | 0.729173694 | 0.809234508 |
| BRD8 | 0.72906942 | 0.871607938 |
| MTOR | 0.729042153 | 1.003893576 |
| APOBEC3B | 0.728996134 | 0.901362751 |
| CBLC | 0.728960653 | 0.974691759 |
| KIR3DP1_pseudogene | 0.728924138 | 0.976508708 |
| C1QA | 0.728840691 | 0.894484412 |

| | | |
|---|---|---|
| MRPL42 | 0.728755735 | 0.979787598 |
| CHID1 | 0.728695824 | 0.903199676 |
| NME8 | 0.728663172 | 0.934386391 |
| EIF3B | 0.728619363 | 0.979748886 |
| RSRP1 | 0.728432005 | 0.824220332 |
| OR1L1 | 0.728390336 | 0.875253139 |
| ZFP69B | 0.728388146 | 0.878088295 |
| LILRB4 | 0.728361669 | 0.872809864 |
| LINGO1 | 0.72835911 | 0.917780478 |
| UCKL1 | 0.728312851 | 0.894873459 |
| MTHFD2L | 0.72822235 | 0.854744776 |
| EDF1 | 0.728179817 | 0.961064244 |
| FCGR2A | 0.728118418 | 0.733900365 |
| IFT88 | 0.72808439 | 0.801944107 |
| D2HGDH | 0.728060912 | 0.859198356 |
| FUBP1 | 0.727827803 | 1.086307592 |
| ZNF576 | 0.727725848 | 0.895854745 |
| GNAL | 0.727716805 | 0.731470231 |
| RASL10A | 0.727716805 | 0.844876468 |
| ZBTB7B | 0.727716805 | 0.694613204 |
| ZNF707 | 0.727716805 | 0.745646011 |
| C1orf162 | 0.72769395 | 0.838300788 |
| AGT | 0.727631133 | 1.005840363 |
| ZNF557 | 0.727575465 | 0.947242206 |
| SLC6A2 | 0.727400216 | 0.925373134 |
| AKIRIN1 | 0.727365776 | 0.839611179 |
| CA7 | 0.727332135 | 0.99156392 |
| NOX5 | 0.727275335 | 0.909215485 |
| CDHR4 | 0.727232472 | 0.926344639 |
| RETNLB | 0.727212173 | 0.907159986 |
| ZNF485 | 0.727141037 | 0.766358342 |
| SMAD5 | 0.727063548 | 0.902389631 |
| GLRX2 | 0.726800697 | 0.965168084 |
| RAD9B | 0.726605659 | 1.014731072 |
| SFRP1 | 0.726506467 | 0.974078574 |
| YARS | 0.726455068 | 0.934386391 |
| CRBN | 0.726381384 | 0.992861778 |
| NAT1 | 0.726373564 | 0.877683273 |
| FAM212A | 0.726275759 | 0.818144998 |
| RPS19BP1 | 0.726267761 | 1.017010936 |
| MAPK7 | 0.726045176 | 0.934458144 |
| PABPN1 | 0.725783018 | 0.898252826 |
| GATAD1 | 0.725775134 | 0.906817403 |
| PTCD3 | 0.725742007 | 0.895504253 |

| | | |
|---|---|---|
| ACAD8 | 0.725714401 | 0.706683971 |
| HPDL | 0.725703802 | 0.878493317 |
| ZNF232 | 0.725701325 | 0.913326482 |
| PAPOLA | 0.725664019 | 0.87605451 |
| GPS2 | 0.7255574 | 0.818955043 |
| PRPF6 | 0.72550376 | 0.799108951 |
| SPAG1 | 0.725487599 | 1.046248715 |
| CCDC25 | 0.725282985 | 0.847711624 |
| RGPD5 | 0.725258967 | 0.813292223 |
| TMEM167B | 0.725233166 | 0.830700689 |
| TOPORS | 0.725228447 | 0.817650876 |
| CDK12 | 0.725094507 | 0.854639844 |
| GGNBP2 | 0.7250833 | 1.007290401 |
| MED4 | 0.725026553 | 0.876468206 |
| XKR5 | 0.725007093 | 0.978938842 |
| ATP10D | 0.724928646 | 0.739975699 |
| THOC7 | 0.724926251 | 0.950587282 |
| LHX5 | 0.724902768 | 1.047387606 |
| PRMT6 | 0.724813725 | 0.913730255 |
| TRAF1 | 0.724758115 | 0.953066119 |
| RAD17 | 0.72471108 | 0.88254354 |
| EDDM3B | 0.724582903 | 0.837615622 |
| LTBR | 0.724500697 | 1.008100446 |
| PMPCA | 0.724444401 | 0.811161583 |
| TEX14 | 0.724403074 | 1.032446463 |
| USMG5 | 0.724396272 | 0.445700582 |
| FGD1 | 0.724390514 | 0.797488862 |
| NMT1 | 0.724366724 | 0.79181855 |
| ACTL6A | 0.72422939 | 0.760631835 |
| SIRT3 | 0.723998867 | 1.055158988 |
| C9orf170 | 0.723928496 | 0.969167523 |
| GLUD2 | 0.723924818 | 0.855002025 |
| EIF3I | 0.723899501 | 0.849331713 |
| C4orf19 | 0.7238317 | 0.986229243 |
| ZFY | 0.723829132 | 0.883521754 |
| DDX26B | 0.723802613 | 0.903604698 |
| IMPDH2 | 0.723777829 | 1.025957171 |
| SALL3 | 0.723768417 | 0.733470367 |
| C16orf89 | 0.723592163 | 1.04898938 |
| JARID2 | 0.723556572 | 0.921020656 |
| FGFR1 | 0.723544031 | 0.996755354 |
| CORO7 | 0.723532835 | 0.93770279 |
| TMTC3 | 0.723440326 | 0.827055488 |
| TCAP | 0.723430544 | 0.896882494 |

| | | |
|---|---|---|
| ROCK1 | 0.723402682 | 0.738481506 |
| GUCA1C | 0.723320359 | 1.0266061 |
| MDFIC | 0.723192146 | 0.781692993 |
| CCDC126 | 0.723138149 | 1.029971648 |
| GGT6 | 0.723109447 | 0.864373783 |
| CLDN19 | 0.723101279 | 0.871531346 |
| TIAM2 | 0.723032475 | 0.93835172 |
| LAG3 | 0.72294096 | 0.987444309 |
| KRT76 | 0.722791905 | 0.919150394 |
| OR5J2 | 0.722786102 | 1.016200891 |
| EPRS | 0.722721375 | 1.116158339 |
| UTP23 | 0.722680757 | 0.702308627 |
| EVI5L | 0.722639751 | 0.870797894 |
| KAAG1 | 0.72262997 | 0.985014176 |
| CSTA | 0.722517708 | 0.975989617 |
| CCDC77 | 0.722510057 | 0.962332928 |
| SERINC3 | 0.722363808 | 0.99108951 |
| JAKMIP2 | 0.72233574 | 0.931146213 |
| OR3A3 | 0.72230161 | 0.889688249 |
| KISS1R | 0.722236574 | 0.909798832 |
| FAM160A2 | 0.722232382 | 0.859862292 |
| PTPRZ1 | 0.722133588 | 0.887735237 |
| GAS1 | 0.721985983 | 0.857828023 |
| EPT1 | 0.721955013 | 1.02308627 |
| RAB4B | 0.7219207 | 0.880597015 |
| SLC22A8 | 0.72179137 | 0.821544452 |
| LGALS9 | 0.721681468 | 1.036340039 |
| SLCO1C1 | 0.721673263 | 0.829080599 |
| C4orf26 | 0.721589415 | 0.817739976 |
| STK10 | 0.721558148 | 0.977287476 |
| CXCR1 | 0.721389139 | 1.007138222 |
| BSG | 0.721370271 | 0.987667009 |
| TMEM186 | 0.721255757 | 0.866747671 |
| PHC2 | 0.721126471 | 0.849736736 |
| SMR3B | 0.721071048 | 0.975698663 |
| FBXO28 | 0.721033585 | 0.852044127 |
| LELP1 | 0.72103218 | 0.970028352 |
| ACSBG1 | 0.721028207 | 0.881328473 |
| EDN3 | 0.721004487 | 1.037684138 |
| CBR4 | 0.720945684 | 0.827055488 |
| SMC1A | 0.720944429 | 0.8669695 |
| DCDC1 | 0.720913641 | 0.891401165 |
| INSRR | 0.72085058 | 1.022712524 |
| PGM2 | 0.720829897 | 0.898744431 |

| | | |
|---|---|---|
| NEFL | 0.720762307 | 0.9777321 |
| DDX18 | 0.72073632 | 0.844471446 |
| OTOG | 0.720697045 | 0.755771567 |
| EPB41L3 | 0.720496838 | 1.044878383 |
| CSNK2A2 | 0.720328239 | 1.090201168 |
| PEX16 | 0.720320648 | 0.916565411 |
| A1BG | 0.720290045 | 0.971908188 |
| HERPUD1 | 0.720263448 | 0.926687222 |
| DPYSL3 | 0.719881391 | 0.902794654 |
| TGFB1 | 0.719765972 | 0.831278391 |
| CADM4 | 0.719690518 | 0.771162414 |
| ERAS | 0.71950859 | 0.798218568 |
| MAP3K4 | 0.719435855 | 0.972744971 |
| THUMPD2 | 0.719389919 | 0.853990915 |
| SMARCC1 | 0.719345653 | 1.021061158 |
| ARSF | 0.719312746 | 0.771567436 |
| PRAMEF16 | 0.719294153 | 0.874848117 |
| CSNK2A1 | 0.719260013 | 0.750162232 |
| HIPK3 | 0.719133894 | 1.06229721 |
| PRKD2 | 0.719129695 | 0.780012979 |
| SNX33 | 0.718947567 | 0.878725591 |
| TMEM234 | 0.718875373 | 0.866747671 |
| TAB3 | 0.7188172 | 0.861077359 |
| GLIS3 | 0.71875276 | 0.723369785 |
| CHFR | 0.718694864 | 1.144711226 |
| ADCY5 | 0.718676859 | 0.912920211 |
| SERPINA10 | 0.718658063 | 0.976103686 |
| PDCD6IP | 0.718599589 | 0.410414526 |
| ZXDC | 0.718563161 | 0.901579587 |
| APOBR | 0.718464159 | 1.071599863 |
| RGS7BP | 0.718387112 | 0.758201701 |
| FMO4 | 0.718328989 | 0.910490077 |
| SERBP1 | 0.718179632 | 1.019846092 |
| ALDH1B1 | 0.718176722 | 0.900064893 |
| RNASEL | 0.71816709 | 0.803374432 |
| NOC4L | 0.717962729 | 0.997164844 |
| SPHK1 | 0.717900856 | 0.907852044 |
| MDP1 | 0.71783273 | 0.715674362 |
| SLPI | 0.7178284 | 0.804779263 |
| CARF | 0.717793404 | 0.712839206 |
| CYB5R3 | 0.71774543 | 0.98247891 |
| GAGE2E | 0.717708349 | 1.139773895 |
| PSME2 | 0.717694122 | 0.752655019 |
| LCORL | 0.717673542 | 0.704333738 |

| | | |
|---|---|---|
| SCGB1C1 | 0.717638597 | 0.848116646 |
| KRT84 | 0.717632993 | 0.876873228 |
| HPCAL1 | 0.717630779 | 0.847893114 |
| NETO1 | 0.717539571 | 0.845837616 |
| TARDBP | 0.717523115 | 1.04090725 |
| NEK9 | 0.717471706 | 1.005191434 |
| COL8A2 | 0.717466895 | 0.995203837 |
| YOD1 | 0.71745158 | 1.042822885 |
| SSTR1 | 0.717424946 | 1.03504218 |
| ZC3H11A | 0.717357865 | 0.796678817 |
| ZNF514 | 0.717337853 | 0.807125728 |
| LCMT1 | 0.717333771 | 0.652895909 |
| IFT57 | 0.717306925 | 0.906034832 |
| MORC1 | 0.717250503 | 0.998053212 |
| SNAPC3 | 0.717097395 | 1.019527235 |
| ANKAR | 0.717095676 | 0.860267315 |
| IL2RB | 0.716967476 | 0.834523037 |
| NTM | 0.716935826 | 1.038882139 |
| UGT1A7 | 0.716843579 | 0.939000649 |
| BPTF | 0.716828103 | 0.953926022 |
| XYLT1 | 0.716724119 | 0.938436614 |
| PIK3R6 | 0.716712218 | 0.881466256 |
| RGP1 | 0.716689019 | 0.961630695 |
| MMACHC | 0.716685557 | 0.855002025 |
| CNOT2 | 0.716521173 | 0.921425678 |
| CYP4F8 | 0.716490711 | 0.892669097 |
| ZNF784 | 0.716482764 | 0.708383961 |
| C4orf6 | 0.716390565 | 0.920863309 |
| SLCO4A1 | 0.716356555 | 0.924724205 |
| CLEC11A | 0.716340831 | 0.954574951 |
| ZNF611 | 0.716291428 | 0.931825968 |
| CNOT3 | 0.716104662 | 0.947242206 |
| DIAPH3 | 0.716095508 | 0.962362103 |
| ADM | 0.715938013 | 0.936622131 |
| RABL2A | 0.715868782 | 0.945321993 |
| PIK3CG | 0.715854136 | 0.746268657 |
| SSX4 | 0.715854136 | 0.910447761 |
| VGLL2 | 0.715738016 | 1.010620075 |
| C19orf55 | 0.715688454 | 0.784123127 |
| ADI1 | 0.715614744 | 0.925475901 |
| FATE1 | 0.715612916 | 0.824254882 |
| ADCY6 | 0.715573826 | 0.828682674 |
| GNG8 | 0.715520402 | 1.014985824 |
| TBX15 | 0.715414687 | 0.911270983 |

| | | |
|---|---|---|
| PCF11 | 0.715319356 | 0.857142857 |
| LOC729974 | 0.715219507 | 0.731992213 |
| PPTC7 | 0.715163198 | 1.00843608 |
| CSK | 0.714965951 | 1.046073978 |
| APOC2 | 0.714953191 | 1.031148605 |
| PTCHD3 | 0.714949872 | 0.969623329 |
| CD4 | 0.714857288 | 0.879410757 |
| ATP7B | 0.714798784 | 0.958282706 |
| METTL3 | 0.714721864 | 0.793843661 |
| TRO | 0.714689552 | 0.901336074 |
| EFNA3 | 0.71465905 | 0.777417262 |
| SKP2 | 0.714548819 | 0.889682025 |
| MICALCL | 0.714545458 | 0.984609154 |
| ZW10 | 0.714404914 | 0.929931146 |
| HMGN5 | 0.714319742 | 1.034021871 |
| CEP350 | 0.714232278 | 0.655731065 |
| IGFBP6 | 0.714231427 | 0.89756766 |
| BMP10 | 0.714099214 | 0.869475848 |
| CNKSR2 | 0.714052998 | 0.899965742 |
| GLI4 | 0.714007507 | 0.986981843 |
| SLC52A2 | 0.713980412 | 0.854402193 |
| DHPS | 0.713972804 | 0.987670344 |
| NOS1AP | 0.713913953 | 0.76873228 |
| RRM2 | 0.713888483 | 0.682057513 |
| RCC1 | 0.71376699 | 0.887403807 |
| EZH2 | 0.713689409 | 0.861482382 |
| TUBA1A | 0.71364178 | 0.774807614 |
| TBC1D20 | 0.713536967 | 0.913669065 |
| RNF186 | 0.713426589 | 0.861129137 |
| CELF4 | 0.713271801 | 0.754961523 |
| ZNF189 | 0.713206614 | 0.961973279 |
| TSHZ2 | 0.713162483 | 0.729040097 |
| GAL3ST4 | 0.713144879 | 0.771576898 |
| CHRNA5 | 0.713125698 | 0.936404932 |
| ZNF645 | 0.713091369 | 0.756651525 |
| ACOT7 | 0.713077703 | 1.003893576 |
| INTS8 | 0.712974871 | 0.879708384 |
| PDLIM4 | 0.712856553 | 0.862281603 |
| ASB3 | 0.712854772 | 0.92926671 |
| MPC2 | 0.712669433 | 0.920615634 |
| GPR144 | 0.712568287 | 0.989617132 |
| PRSS36 | 0.712534946 | 0.839714471 |
| FZD3 | 0.712517988 | 0.974691759 |
| PVALB | 0.712516866 | 0.958547448 |

| | | |
|---|---|---|
| NCF1 | 0.71248177 | 0.90927501 |
| TRIM56 | 0.712470158 | 0.892277742 |
| NOP2 | 0.712412532 | 0.944841012 |
| C1orf210 | 0.712386155 | 0.903604698 |
| TAPBP | 0.712381774 | 0.845686513 |
| PLAG1 | 0.712337915 | 0.900769542 |
| AUH | 0.712307899 | 0.98247891 |
| IRF2 | 0.712300243 | 0.914990266 |
| TANC1 | 0.712191309 | 0.892264075 |
| PLRG1 | 0.712173327 | 0.834345889 |
| VPS37D | 0.712141795 | 0.889428919 |
| SPATS2L | 0.712115917 | 0.906844876 |
| HIC2 | 0.712081305 | 0.906817403 |
| CA9 | 0.712044754 | 0.996759822 |
| FASTKD5 | 0.71204292 | 0.921425678 |
| FAM27L | 0.711980315 | 0.767922236 |
| TNFRSF12A | 0.711835229 | 1.027064063 |
| RNF38 | 0.711728261 | 0.900713822 |
| MTERFD1 | 0.711719093 | 0.829080599 |
| TFAP4 | 0.711454969 | 0.869987849 |
| TAF13 | 0.711384009 | 0.812069664 |
| MYBBP1A | 0.711355501 | 0.826086957 |
| FLT4 | 0.711350068 | 0.773523686 |
| SPRTN | 0.7112508 | 0.733900365 |
| FOS | 0.711006542 | 0.899554475 |
| RSPH4A | 0.710962068 | 0.954093868 |
| CIAPIN1 | 0.710953666 | 0.89671932 |
| BCL9L | 0.710924597 | 0.818773553 |
| GAD1 | 0.71066402 | 0.743215877 |
| TCEB2 | 0.710544371 | 0.962362103 |
| FAM203A | 0.71042832 | 0.920615634 |
| MAP3K5 | 0.710369763 | 0.804672291 |
| NAPB | 0.710302498 | 0.880113406 |
| CTNNBL1 | 0.710100053 | 0.953066119 |
| ZNF599 | 0.709774231 | 0.883521754 |
| ZNF695 | 0.709764926 | 0.886593763 |
| TMIE | 0.709690545 | 0.842069202 |
| MIEN1 | 0.709556887 | 0.892669097 |
| CHST13 | 0.709535288 | 0.648440664 |
| CDC5L | 0.709491938 | 0.921830701 |
| GMFG | 0.709474101 | 0.8663926 |
| TMPO | 0.709362503 | 1.039397054 |
| FH | 0.709325904 | 1.031797534 |
| CRMP1 | 0.709029788 | 0.987021415 |

| | | |
|---|---|---|
| THBD | 0.709003856 | 0.949383517 |
| VWF | 0.708986255 | 0.849736736 |
| KHDRBS2 | 0.708974608 | 0.862624186 |
| FAM71B | 0.708936996 | 0.878898339 |
| DUSP27 | 0.708707765 | 0.713244228 |
| NDUFA11 | 0.708693603 | 0.914135277 |
| NGF | 0.708616633 | 0.617253949 |
| PLCB1 | 0.70860563 | 0.987039287 |
| PCSK6 | 0.7085724 | 0.829331603 |
| ACKR1 | 0.708519631 | 0.950032446 |
| MFSD11 | 0.708515007 | 0.90562981 |
| HS3ST5 | 0.70836264 | 1.116158339 |
| CLDN8 | 0.708302213 | 0.999314834 |
| OSBP | 0.708267145 | 1.040882544 |
| ZFYVE27 | 0.708206972 | 0.396711202 |
| RHOU | 0.708158171 | 0.874038072 |
| AHCYL2 | 0.70809863 | 0.988319273 |
| GABRA3 | 0.708085623 | 0.807268008 |
| SVOP | 0.707888036 | 0.743215877 |
| VPS9D1 | 0.7078536 | 0.816119887 |
| LOC643847 | 0.707830167 | 0.874443094 |
| LINC00207 | 0.707690897 | 0.912515188 |
| UNG | 0.707641842 | 0.822195221 |
| ZMYM4 | 0.707443582 | 0.974078574 |
| PADI2 | 0.707435636 | 0.857432159 |
| SNIP1 | 0.707372234 | 0.904076739 |
| OR4K5 | 0.70724949 | 1.020554985 |
| PDE3B | 0.707170024 | 0.989617132 |
| FOXRED1 | 0.707033259 | 0.855002025 |
| UNC13D | 0.707028878 | 0.935251799 |
| EIF4G3 | 0.706838158 | 0.776427704 |
| CABIN1 | 0.706763952 | 0.283316204 |
| DIS3L2 | 0.706649018 | 1.007290401 |
| AGO4 | 0.706623548 | 0.782503038 |
| NOS3 | 0.70658957 | 0.753341434 |
| IFIH1 | 0.706545273 | 0.998053212 |
| A2M | 0.706438533 | 0.969623329 |
| WNT9A | 0.70641235 | 0.907502569 |
| ANAPC15 | 0.706303847 | 0.908464966 |
| ITGB5 | 0.706287088 | 0.990279465 |
| ACAD11 | 0.706199267 | 0.835560956 |
| KCNH8 | 0.706145017 | 0.88513952 |
| CLEC17A | 0.706123547 | 0.8441247 |
| ZKSCAN4 | 0.706122857 | 0.95544755 |

| | | |
|---|---|---|
| PDIA4 | 0.706108054 | 0.953277093 |
| HIST1H3C | 0.706084535 | 1.033230558 |
| SCGB1D2 | 0.706034559 | 0.905789654 |
| PDPN | 0.705964433 | 0.745240988 |
| VCY | 0.705943803 | 1.009249743 |
| OR2A14 | 0.70583642 | 0.912641316 |
| DSE | 0.705755527 | 0.885783718 |
| MMAB | 0.705675755 | 0.815714864 |
| GAPDH | 0.705627616 | 0.885788449 |
| PRMT2 | 0.705608004 | 0.78655326 |
| DRG2 | 0.705504035 | 0.889023896 |
| HOXB7 | 0.705475203 | 0.848116646 |
| STK38 | 0.705472601 | 0.984425698 |
| EIF4G2 | 0.705426963 | 0.938841636 |
| ZNF575 | 0.70540036 | 1.003425831 |
| ZNF648 | 0.705325541 | 0.868772783 |
| ZFP36 | 0.705324104 | 0.882494005 |
| ARL8B | 0.705258351 | 0.94491697 |
| RAE1 | 0.705235886 | 0.707978939 |
| SAMD4A | 0.705216494 | 1.113562622 |
| GDA | 0.705154382 | 0.927501013 |
| HOXA10 | 0.705035742 | 0.766707169 |
| YME1L1 | 0.704993315 | 0.962362103 |
| OR2T29 | 0.704963032 | 0.937992463 |
| SERPIND1 | 0.70495627 | 0.682024659 |
| NPY2R | 0.704822165 | 0.947436729 |
| NFKBIB | 0.704816648 | 0.810854597 |
| GIMAP8 | 0.704740697 | 0.705548805 |
| WDR78 | 0.704647986 | 0.843256379 |
| MEX3A | 0.704543748 | 0.985723556 |
| CLEC3A | 0.704541113 | 0.890238963 |
| IL27RA | 0.704440863 | 0.861077359 |
| ZIM2 | 0.704400015 | 0.912298732 |
| GYPC | 0.704308139 | 0.925373134 |
| NOD1 | 0.704295327 | 0.893479141 |
| CABS1 | 0.704070171 | 0.884973674 |
| MVB12B | 0.703931308 | 0.978938842 |
| KLF4 | 0.703892746 | 0.952612394 |
| KRTAP4-5 | 0.703838357 | 0.882151422 |
| CRKL | 0.703832229 | 0.850097339 |
| OR2D3 | 0.703809087 | 1.027749229 |
| ALDH3A2 | 0.703786786 | 1.077871512 |
| RGR | 0.703782917 | 0.827384815 |
| CEBPA | 0.703745835 | 0.82381531 |

| | | |
|---|---|---|
| DTX1 | 0.70372101 | 0.880597015 |
| LRRD1 | 0.703616345 | 1.002835156 |
| NSL1 | 0.703568359 | 0.903199676 |
| ZKSCAN8 | 0.703513125 | 1.29736211 |
| SLC39A14 | 0.703495952 | 0.836245289 |
| TNRC18 | 0.703347926 | 0.798903734 |
| KRT83 | 0.703311723 | 0.929931146 |
| UTP20 | 0.703263177 | 1.000405022 |
| MBP | 0.703181413 | 1.055155875 |
| CDCP2 | 0.703146422 | 0.815714864 |
| DUOX2 | 0.703134655 | 0.695423248 |
| TMX4 | 0.702982802 | 0.989064399 |
| PTPLAD2 | 0.702959777 | 0.84528149 |
| OSTC | 0.70288971 | 0.984926345 |
| SPATA5 | 0.702805327 | 0.780882949 |
| ZC3H8 | 0.702709355 | 0.825030377 |
| EEF1G | 0.702708133 | 0.836776023 |
| ASTN1 | 0.702623113 | 0.786958283 |
| C4orf45 | 0.702570731 | 0.998629668 |
| PRKACG | 0.702563051 | 0.996106424 |
| C1orf192 | 0.702357002 | 0.801944107 |
| MILR1 | 0.702316606 | 0.928311057 |
| FCRLA | 0.702286806 | 0.846091535 |
| SLC2A8 | 0.702205659 | 0.681652491 |
| KLRAP1 | 0.702176368 | 0.796273795 |
| TSPO2 | 0.702142136 | 0.887403807 |
| PIGP | 0.701911275 | 0.799513973 |
| TMEM119 | 0.701852389 | 0.923045768 |
| MRGPRX4 | 0.701847251 | 0.928617781 |
| THPO | 0.701714697 | 0.861482382 |
| SYT11 | 0.701644648 | 0.913669065 |
| CH25H | 0.701628863 | 0.858647226 |
| JPH2 | 0.701608038 | 1 |
| POC1A | 0.701571301 | 0.811664642 |
| ATL1 | 0.701343597 | 1.104477612 |
| APOBEC3H | 0.701260769 | 0.835560956 |
| LOC651503_pseudogene | 0.7012389 | 0.973963686 |
| TMEM101 | 0.701136526 | 0.90927501 |
| ALG1 | 0.701112126 | 0.753341434 |
| ITPKA | 0.701033733 | 0.830629461 |
| ESF1 | 0.701011621 | 0.838801134 |
| ZFAND6 | 0.700940543 | 0.71641791 |
| FAM26F | 0.700890332 | 0.780072904 |
| DGKI | 0.70088481 | 0.868916288 |

| | | |
|---|---|---|
| SLC9A3 | 0.70074058 | 0.968202466 |
| LARGE | 0.700601909 | 0.739165654 |
| CPLX1 | 0.700559936 | 0.923946557 |
| COPB1 | 0.700461518 | 0.756986634 |
| NPTX1 | 0.700321562 | 1.028434395 |
| FUK | 0.700292165 | 1.029850746 |
| RNF181 | 0.700045936 | 0.984425698 |
| EPM2AIP1 | 0.699930631 | 0.960260363 |
| C2orf83 | 0.699873719 | 0.931551235 |
| ACSF2 | 0.699747432 | 0.77871512 |
| NOP14 | 0.699712748 | 0.757796679 |
| LAIR1 | 0.699685139 | 0.799108951 |
| MPP1 | 0.699673012 | 0.717715769 |
| WFDC10A | 0.699510702 | 0.929770469 |
| MISP | 0.699508376 | 0.98217902 |
| DHRS3 | 0.699437671 | 0.781692993 |
| CKAP5 | 0.699424523 | 0.710174717 |
| MAD2L1 | 0.699284392 | 0.767112191 |
| NAB2 | 0.699260298 | 0.994329688 |
| PRKAA1 | 0.699247977 | 0.909149903 |
| COPA | 0.699233724 | 0.828023296 |
| OR51M1 | 0.699190272 | 0.901336074 |
| PUS3 | 0.699178946 | 0.722964763 |
| TCEB1 | 0.698970795 | 0.825438027 |
| FEZF1 | 0.698880997 | 0.950587282 |
| NT5DC3 | 0.698736938 | 1.006885379 |
| TCFL5 | 0.698684228 | 0.703439325 |
| WNT9B | 0.69865136 | 0.90647482 |
| IP6K1 | 0.6984678 | 1.001946788 |
| TRIM8 | 0.698463931 | 0.894873459 |
| XXYLT1 | 0.698444491 | 0.936411503 |
| GNA11 | 0.698393551 | 0.824220332 |
| BAG3 | 0.698251208 | 0.910447761 |
| PRKCQ | 0.698251208 | 0.910447761 |
| SURF2 | 0.698228259 | 1.031517643 |
| ETV3 | 0.698214801 | 0.839206156 |
| DTX4 | 0.698168967 | 0.879948086 |
| C11orf53 | 0.698109457 | 0.756581612 |
| PLXNB1 | 0.697953326 | 0.990279465 |
| TECTA | 0.697810642 | 0.975698663 |
| TCEAL8 | 0.697738766 | 0.78655326 |
| SNAPC2 | 0.697679726 | 0.863507493 |
| COQ5 | 0.697652677 | 0.715674362 |
| CCND1 | 0.697624507 | 0.910895099 |

| | | |
|---|---|---|
| ATP2A3 | 0.697355746 | 0.931956258 |
| UQCC2 | 0.69733498 | 0.903734156 |
| MS4A7 | 0.69720532 | 0.840421223 |
| FAM71C | 0.696948734 | 0.852571891 |
| UEVLD | 0.696886206 | 0.942894225 |
| TPTE | 0.696881761 | 0.820246593 |
| HS3ST6 | 0.69688134 | 0.6690968 |
| SPANXN3 | 0.69684609 | 0.849736736 |
| PHKA2 | 0.69681829 | 0.866320571 |
| LHFP | 0.696781025 | 0.824254882 |
| PMEL | 0.69675017 | 0.932766302 |
| NLN | 0.696709705 | 0.958468527 |
| LRRC57 | 0.696596126 | 0.814499797 |
| NR1H2 | 0.696421245 | 1.027903958 |
| ALS2CR12 | 0.696385699 | 0.871531346 |
| LST1 | 0.696343561 | 0.985268928 |
| LRP12 | 0.695840043 | 0.998053212 |
| TMEM106B | 0.695839919 | 0.702713649 |
| COMP | 0.695820643 | 1.050702295 |
| RLTPR | 0.695649047 | 0.928311057 |
| CALM1 | 0.69561209 | 0.975340688 |
| RPS6KA1 | 0.695520299 | 0.890979883 |
| GFPT1 | 0.695500871 | 0.798831927 |
| C11orf68 | 0.695484612 | 0.886998785 |
| AKAP8 | 0.695461438 | 0.99091499 |
| DRAP1 | 0.695311262 | 0.836776023 |
| KDM5A | 0.695290428 | 0.863507493 |
| PHKG1 | 0.695255959 | 0.868267359 |
| VPS26B | 0.695150495 | 0.73309032 |
| GALK1 | 0.695149598 | 0.975989617 |
| SLC2A6 | 0.695146827 | 0.923045768 |
| TNFRSF11B | 0.694989779 | 0.801539085 |
| ADRA1B | 0.694936381 | 0.799480857 |
| GLIPR1L1 | 0.6949286 | 0.970838396 |
| ICAM3 | 0.694910917 | 0.884549503 |
| RXRA | 0.6948636 | 0.834345889 |
| LYRM1 | 0.694854329 | 0.869987849 |
| LYPLAL1 | 0.694847961 | 0.89671932 |
| LACRT | 0.694769279 | 0.933881466 |
| INPP5D | 0.694757328 | 0.968851395 |
| GJB3 | 0.694710815 | 0.922777417 |
| ROS1 | 0.694602231 | 1.038286827 |
| GLT8D1 | 0.694566667 | 0.93835172 |
| CWC15 | 0.694526646 | 0.516272696 |

| | | |
|---|---|---|
| SEC11C | 0.694496363 | 0.946138871 |
| MOXD1 | 0.694389296 | 0.751721345 |
| DYNLL1 | 0.694338203 | 0.93600648 |
| COL6A2 | 0.694325336 | 0.956834532 |
| FAM96B | 0.694258584 | 0.986634265 |
| SLC22A13 | 0.694247988 | 0.959902795 |
| MBD3L1 | 0.6941972 | 0.997259335 |
| KLRD1 | 0.694092926 | 0.859052248 |
| ZNF350 | 0.693985493 | 1.004860267 |
| DHX58 | 0.693907708 | 0.897934386 |
| PRG4 | 0.69385842 | 0.966426859 |
| UTP11L | 0.69378647 | 0.990279465 |
| COPS8 | 0.693632461 | 0.93764988 |
| GRIP2 | 0.693495989 | 0.868448099 |
| USP54 | 0.693437086 | 0.821514217 |
| DNM1 | 0.693334019 | 0.934386391 |
| WDR41 | 0.69331782 | 0.993924666 |
| FAM49B | 0.693285979 | 1.041109969 |
| CD38 | 0.693253687 | 0.812474686 |
| RAN | 0.69318777 | 0.904819765 |
| OR5T3 | 0.692970307 | 0.990065091 |
| SND1-IT1 | 0.692963379 | 0.839611179 |
| SGK2 | 0.69276206 | 0.925373134 |
| VGLL1 | 0.692659724 | 1.042527339 |
| PDF | 0.692657992 | 0.899554475 |
| LAMP2 | 0.692650676 | 0.940947437 |
| XPR1 | 0.692640633 | 0.740428293 |
| ANKS1B | 0.692616985 | 0.974078574 |
| LILRA4 | 0.692587899 | 0.9145403 |
| MAPK1 | 0.692486606 | 1.033744322 |
| ASGR1 | 0.692486528 | 1.045767517 |
| PRPF38B | 0.69248286 | 0.869987849 |
| SCD | 0.692370588 | 0.992304577 |
| SSBP2 | 0.692133054 | 1.010530579 |
| H2AFB2 | 0.692033304 | 0.877012676 |
| NUBP1 | 0.691909121 | 0.790198461 |
| ANGPTL4 | 0.691845587 | 0.616992121 |
| CENPU | 0.691556971 | 0.912920211 |
| MTERF | 0.691546885 | 0.545970028 |
| COX7B | 0.691377539 | 0.859883522 |
| TUBGCP3 | 0.691136084 | 0.965741692 |
| AKNA | 0.69107111 | 0.855772525 |
| FMOD | 0.691068844 | 0.835560123 |
| FGA | 0.69091971 | 0.680842446 |

| | | |
|---|---|---|
| CHIT1 | 0.690758127 | 0.797488862 |
| IQSEC3 | 0.690757613 | 0.907845152 |
| SRCAP | 0.69066939 | 0.779667882 |
| KHDC3L | 0.690609433 | 1.000810045 |
| EPHB2 | 0.690570121 | 0.881894873 |
| CREBL2 | 0.690452811 | 1.005840363 |
| GNL3 | 0.690421718 | 0.956257594 |
| UBALD1 | 0.690354834 | 0.875658161 |
| OR2T5 | 0.69035037 | 0.906132237 |
| AKR1D1 | 0.690287917 | 0.835820896 |
| PHYHD1 | 0.690210039 | 0.851395198 |
| C16orf46 | 0.690157288 | 0.816119887 |
| PDE1C | 0.690127437 | 0.893884164 |
| ABCA9 | 0.690102278 | 0.983127839 |
| PRSS8 | 0.690098327 | 0.874756652 |
| ISLR2 | 0.690060451 | 0.962737951 |
| POMP | 0.689924789 | 0.913692408 |
| ATP13A2 | 0.689909158 | 0.912110166 |
| TCF20 | 0.689787392 | 0.828682674 |
| MYOZ1 | 0.689769003 | 0.981157931 |
| FAM209A | 0.689694287 | 0.841726619 |
| GNB4 | 0.68938315 | 0.945727015 |
| PARP14 | 0.689341035 | 0.805970149 |
| AJAP1 | 0.689328084 | 0.882948562 |
| FGF22 | 0.68931815 | 0.951802349 |
| FLJ45079 | 0.689286319 | 0.81294964 |
| CCT4 | 0.689217715 | 0.976638546 |
| RASGRP2 | 0.689173378 | 0.703523694 |
| MMP26 | 0.68914313 | 1 |
| PEAR1 | 0.68910466 | 0.847306602 |
| HINT1 | 0.689059323 | 0.739570676 |
| LRRC71 | 0.688955915 | 0.775617659 |
| OR56A1 | 0.688933043 | 0.706763872 |
| SIRT5 | 0.688691026 | 0.898744431 |
| TLK2 | 0.688635361 | 1.024010383 |
| NAIP | 0.688417287 | 0.931213498 |
| TAF9B | 0.688368222 | 0.760545101 |
| SULT1E1 | 0.688324766 | 0.995139733 |
| CENPK | 0.688296953 | 0.6654516 |
| KNG1 | 0.688160637 | 1.013365735 |
| CSF3 | 0.688150806 | 0.838416613 |
| RAB11FIP3 | 0.688092479 | 0.773592548 |
| AK5 | 0.687953971 | 0.924075276 |
| DDX56 | 0.687900297 | 1.188189487 |

| | | |
|---|---|---|
| BCCIP | 0.687734964 | 0.882494005 |
| RFC2 | 0.68768 | 0.807916937 |
| SYT14L | 0.687642728 | 0.831105711 |
| C12orf5 | 0.687564637 | 0.888213852 |
| ENTPD3 | 0.687471146 | 0.924724205 |
| NEDD4 | 0.687435582 | 0.948085659 |
| FMO2 | 0.687391848 | 0.789388416 |
| CDC14A | 0.687369394 | 0.994808566 |
| FAM110D | 0.687358737 | 0.927501013 |
| DNM2 | 0.687240501 | 1.093182597 |
| RALGPS2 | 0.687147233 | 0.813689753 |
| EIF4G1 | 0.68708719 | 0.839714471 |
| HSD17B1 | 0.687050294 | 0.963953017 |
| FBXL19 | 0.686973173 | 0.940947437 |
| INO80B | 0.686882419 | 1.007879411 |
| SH3TC1 | 0.686819665 | 0.30284344 |
| ZNF502 | 0.686725829 | 1.084960603 |
| HOXD13 | 0.686640989 | 0.961522884 |
| CD300C | 0.686594277 | 0.967598218 |
| QSOX1 | 0.686427958 | 0.913730255 |
| RP6-166C19.11 | 0.686364435 | 0.691373026 |
| MSI1 | 0.686322936 | 1.010962658 |
| ITGAV | 0.686167551 | 0.730660186 |
| UBE2E1 | 0.686026867 | 0.809863725 |
| RPRD1B | 0.685973011 | 1.157958688 |
| APEX2 | 0.68552132 | 0.880597015 |
| ABCB6 | 0.685516332 | 0.914945322 |
| FKBP2 | 0.685516332 | 0.914945322 |
| MIR22HG | 0.685495719 | 0.928400137 |
| DDX39A | 0.685325561 | 0.876063183 |
| POU4F1 | 0.685260816 | 0.840016201 |
| MPHOSPH10 | 0.685160311 | 1.038882139 |
| GAK | 0.685155146 | 1.021414666 |
| GPRC6A | 0.685155146 | 1.021414666 |
| OR52N2 | 0.685147576 | 0.948562171 |
| CD300LF | 0.685100151 | 1.002835156 |
| RAB40C | 0.685083278 | 0.943543154 |
| LDHB | 0.685057565 | 0.822195221 |
| BATF | 0.684969837 | 0.842310188 |
| FAM58A | 0.684968878 | 0.88254354 |
| KCNK6 | 0.684710107 | 1.014276444 |
| IRAK1 | 0.684699981 | 1.006489293 |
| MFF | 0.684622332 | 0.96435804 |
| SF3A1 | 0.684598272 | 0.861596437 |

| | | |
|---|---|---|
| HAS1 | 0.684547144 | 1.0036452 |
| MUM1L1 | 0.684467474 | 0.783718104 |
| PCNA | 0.684207194 | 0.733939001 |
| ZBTB45 | 0.684166825 | 0.886593763 |
| AMFR | 0.684157636 | 0.952628164 |
| GAL3ST1 | 0.684137127 | 0.919805589 |
| CHD1 | 0.684019404 | 0.856622114 |
| HOXC11 | 0.683985478 | 0.862697448 |
| A4GNT | 0.6838485 | 0.947347104 |
| NLRP2 | 0.683792285 | 0.79688514 |
| OR3A2 | 0.683681387 | 0.865022268 |
| WNK3 | 0.683438131 | 1.031797534 |
| HSP90AB1 | 0.683292233 | 0.975293641 |
| C8orf76 | 0.683187653 | 0.966383151 |
| NAP1L2 | 0.683007225 | 0.899965742 |
| ACTN1 | 0.682948489 | 0.954637505 |
| FTMT | 0.682928877 | 0.848926691 |
| AIFM1 | 0.682924802 | 0.960712839 |
| GPLD1 | 0.682922705 | 0.860480208 |
| ODF3L2 | 0.682695087 | 0.406988695 |
| PRKCH | 0.682594235 | 0.907852044 |
| BRD2 | 0.682564599 | 0.900064893 |
| STK40 | 0.682564599 | 0.900064893 |
| NUP133 | 0.682559385 | 0.905104488 |
| KRTAP26-1 | 0.682467766 | 0.631834751 |
| KIAA1432 | 0.682453043 | 0.939362795 |
| P4HA1 | 0.682452128 | 0.761846902 |
| DNAJB7 | 0.682309231 | 1.019184652 |
| CNKSR1 | 0.682255448 | 0.95101062 |
| ART3 | 0.682196306 | 0.964763062 |
| PIGX | 0.68218133 | 0.692588092 |
| TIMP1 | 0.681856537 | 0.853990915 |
| NFIC | 0.681775133 | 0.962332928 |
| RAB27B | 0.681702696 | 0.935196436 |
| EDC3 | 0.681625929 | 0.908059943 |
| HARS | 0.681414222 | 0.808424463 |
| LARP4 | 0.681395374 | 0.878088295 |
| NUPR1 | 0.681223756 | 1.020656136 |
| PLCE1 | 0.681163647 | 0.911705144 |
| NRF1 | 0.680906552 | 0.887808829 |
| SCARF1 | 0.680891629 | 0.87084618 |
| NELL1 | 0.680869824 | 0.742405832 |
| ADRA2A | 0.680774069 | 0.847501622 |
| RAMP2 | 0.680767383 | 0.966788173 |

| | | |
|---|---|---|
| AP4S1 | 0.680764174 | 0.900650908 |
| TSPAN13 | 0.680732082 | 1.078979344 |
| GPBAR1 | 0.680554595 | 0.894873459 |
| ADAM20 | 0.680506518 | 0.887086308 |
| CERCAM | 0.680449468 | 0.957819598 |
| CDK11B | 0.680400157 | 1.028552888 |
| FAM102B | 0.680391547 | 0.80963953 |
| EDIL3 | 0.680285704 | 0.967454608 |
| TMOD4 | 0.680246693 | 0.901336074 |
| SPANXC | 0.680188696 | 0.997259335 |
| AMD1 | 0.680089551 | 0.824789098 |
| TRPC4AP | 0.680009037 | 0.852571891 |
| ARID5A | 0.679899531 | 0.9109284 |
| CSTB | 0.67983048 | 1.005840363 |
| SLC40A1 | 0.679620567 | 0.986634265 |
| RIPK4 | 0.679591331 | 0.691758598 |
| DPCD | 0.679202348 | 0.899149453 |
| PPP6R2 | 0.679202348 | 0.965978129 |
| SPRR1B | 0.679068019 | 1.023896314 |
| KLF1 | 0.679028897 | 0.793033617 |
| DHRS4 | 0.67902116 | 0.826086957 |
| FECH | 0.678954396 | 0.944192083 |
| AURKB | 0.678891337 | 0.873458793 |
| DLG3 | 0.678843425 | 0.99156392 |
| EEPD1 | 0.678829318 | 0.85748544 |
| WNT1 | 0.678740058 | 0.944501542 |
| DHTKD1 | 0.678576905 | 0.659781288 |
| RABAC1 | 0.678506505 | 0.790603483 |
| RBMS3 | 0.678502228 | 0.951695786 |
| ZNF708 | 0.678459592 | 0.917437479 |
| MC2R | 0.678425469 | 0.937053861 |
| TRPC6 | 0.678425469 | 0.937053861 |
| AVP | 0.677980803 | 1.013365735 |
| CCT7 | 0.677879337 | 0.819597664 |
| STAT3 | 0.677834224 | 1.032446463 |
| FAH | 0.677811987 | 0.791413528 |
| TNC | 0.677678715 | 0.906554186 |
| PNP | 0.67718094 | 0.612393682 |
| NLRP13 | 0.677015283 | 0.691778048 |
| ZNF829 | 0.676999399 | 0.880438506 |
| GAB4 | 0.676995874 | 0.685702714 |
| SLC5A12 | 0.676971525 | 0.801539085 |
| MAPK11 | 0.676928422 | 0.978585334 |
| SFXN4 | 0.676923132 | 0.905224787 |

| | | |
|---|---|---|
| TAF1L | 0.676894175 | 0.828682674 |
| ACAN | 0.676754163 | 0.78655326 |
| RAC2 | 0.676685343 | 0.710409072 |
| STAB1 | 0.676603024 | 0.899554475 |
| B3GAT2 | 0.676561589 | 1.081765088 |
| PRSS1 | 0.676537469 | 0.797534069 |
| DEFB126 | 0.676310014 | 0.9777321 |
| PARP11 | 0.676309845 | 0.916288125 |
| SNAP29 | 0.676212689 | 0.927372388 |
| NKX2-3 | 0.676134332 | 1.03504218 |
| ALOXE3 | 0.676057913 | 0.656136087 |
| SNRPF | 0.675946812 | 0.802754151 |
| ATP1A4 | 0.675919763 | 0.921830701 |
| SIAH2 | 0.675857726 | 1.138221934 |
| FOXB2 | 0.675737041 | 0.873228028 |
| C16orf80 | 0.675714831 | 0.94653706 |
| FRMD6 | 0.675666489 | 0.894694208 |
| CNNM2 | 0.675549673 | 0.909900651 |
| DYRK2 | 0.675524317 | 1.059701493 |
| KRT33A | 0.675463339 | 0.846091535 |
| MVK | 0.675353167 | 0.917585983 |
| MPI | 0.675272212 | 0.769947347 |
| GAGE8 | 0.675137537 | 0.947242206 |
| PRKCSH | 0.675021075 | 0.728098637 |
| NOB1 | 0.674951849 | 0.938436614 |
| C16orf72 | 0.67473846 | 0.801134062 |
| TP53BP1 | 0.674613148 | 0.899149453 |
| NFYA | 0.674399287 | 0.918883842 |
| ACTC1 | 0.674295994 | 1.037262049 |
| MFSD5 | 0.674283055 | 0.810854597 |
| CCL7 | 0.674130502 | 0.673588579 |
| CLLU1OS | 0.674051532 | 0.774402592 |
| CST2 | 0.673999224 | 0.921205892 |
| TJAP1 | 0.673916744 | 0.728635075 |
| PLS3 | 0.673903304 | 0.932853717 |
| HOXD8 | 0.673877877 | 0.826650466 |
| MIER1 | 0.673811859 | 0.842041312 |
| DNAH7 | 0.67367616 | 0.792991564 |
| EAF2 | 0.673627506 | 0.888213852 |
| ADARB2 | 0.673586499 | 0.960945529 |
| GLB1 | 0.673567957 | 0.805589307 |
| PNPLA1 | 0.673554101 | 0.974078574 |
| ETS1 | 0.673083021 | 0.928617781 |
| GPD1 | 0.673083021 | 0.928617781 |

| | | |
|---|---|---|
| EBNA1BP2 | 0.672974185 | 1.03562864 |
| BTLA | 0.672897204 | 0.874272011 |
| RNF41 | 0.672884622 | 1.016223232 |
| TESK2 | 0.6727832 | 1.00843608 |
| FBXO44 | 0.672748954 | 0.603504218 |
| APC2 | 0.672517927 | 0.843661401 |
| ECI1 | 0.6722049 | 0.923855812 |
| SPCS1 | 0.67202632 | 0.890330954 |
| KPNA1 | 0.671938177 | 0.908869988 |
| NRBF2 | 0.671505593 | 0.875658161 |
| EFCAB4A | 0.671417772 | 0.989469421 |
| CUL1 | 0.671412765 | 1.066190785 |
| MATR3 | 0.671383456 | 0.883179171 |
| TDGF1 | 0.671361231 | 0.819597664 |
| AMELY | 0.671312637 | 0.96882494 |
| VMO1 | 0.671304634 | 0.940461725 |
| PNPLA3 | 0.671258482 | 1.038882139 |
| MLXIPL | 0.671048073 | 0.891628812 |
| KMT2E | 0.671015701 | 0.940866748 |
| POLR1E | 0.670979585 | 0.836371 |
| PLEK | 0.670974134 | 0.919400567 |
| PNPO | 0.670940259 | 0.965573107 |
| LEPRE1 | 0.670837003 | 0.855002025 |
| SUSD4 | 0.670798645 | 0.916565411 |
| POLH | 0.670781922 | 1.051265412 |
| PNO1 | 0.670692995 | 0.889023896 |
| NEIL2 | 0.670629789 | 0.780661908 |
| MYRFL | 0.67061037 | 0.944511948 |
| CD3D | 0.670559634 | 0.923045768 |
| TRMT61A | 0.670485159 | 0.741595788 |
| ULK2 | 0.670403071 | 0.940298507 |
| ELF2 | 0.670285349 | 0.876703439 |
| GOT2 | 0.669870821 | 0.825435399 |
| H1F0 | 0.669867805 | 0.909149903 |
| C2orf16 | 0.669823446 | 0.982584042 |
| TAS2R9 | 0.669777548 | 0.818088386 |
| TPH2 | 0.669699254 | 0.767112191 |
| ZFHX3 | 0.669547453 | 0.982989064 |
| SLC25A46 | 0.669459071 | 0.875253139 |
| SESN3 | 0.669439714 | 0.986229243 |
| EIF2B3 | 0.669406195 | 0.884568651 |
| CAPN3 | 0.669382263 | 0.989617132 |
| FGG | 0.66920074 | 0.926022064 |
| TP53BP2 | 0.669187306 | 1.057513163 |

| | | |
|---|---|---|
| TOR1AIP1 | 0.66909164 | 0.925070879 |
| NAE1 | 0.66907916 | 0.869582827 |
| MYL7 | 0.669029951 | 0.703118672 |
| ABCC6 | 0.668876222 | 1.038286827 |
| CRB2 | 0.668789255 | 0.886605002 |
| LSM12 | 0.668575972 | 0.931956258 |
| PIN4 | 0.668530958 | 0.966904607 |
| SERPINB1 | 0.668530958 | 0.966904607 |
| USF2 | 0.668462136 | 0.887086308 |
| ACBD6 | 0.668320527 | 0.980130182 |
| LBP | 0.668242926 | 0.966383151 |
| PTGES | 0.668213941 | 0.926285946 |
| ME1 | 0.668145579 | 0.870797894 |
| BMI1 | 0.668130538 | 0.895522388 |
| IDH3A | 0.668067895 | 0.790603483 |
| OR4C11 | 0.668067895 | 0.86472256 |
| MAD1L1 | 0.667875225 | 1.055155875 |
| E4F1 | 0.667813759 | 0.879948086 |
| STX4 | 0.667762916 | 0.997974889 |
| POLR2A | 0.667725832 | 0.791008505 |
| NGDN | 0.667714778 | 0.957877683 |
| HCCS | 0.667630115 | 0.92709599 |
| SEMA6D | 0.667587781 | 0.862624186 |
| HRH1 | 0.6674857 | 0.888384166 |
| YWHAH | 0.6674857 | 1.032446463 |
| SHROOM4 | 0.667398032 | 1.060348319 |
| PRM1 | 0.667309752 | 0.936816525 |
| ERH | 0.667281352 | 1.042137718 |
| WRAP73 | 0.667068034 | 0.851661528 |
| EN2 | 0.667050671 | 1.06399352 |
| NNT | 0.66704629 | 0.893884164 |
| APLP1 | 0.667009993 | 0.989037342 |
| PLXDC2 | 0.666887475 | 0.961288112 |
| TMCO1 | 0.666784559 | 0.764277035 |
| MARCH5 | 0.666758951 | 0.993510707 |
| LIMK2 | 0.666511899 | 0.889682025 |
| WTIP | 0.666343112 | 0.705953827 |
| FAM207A | 0.666317571 | 0.86193902 |
| C6orf163 | 0.666271038 | 0.980815348 |
| NAT14 | 0.666176996 | 0.876468206 |
| TNNT3 | 0.665955684 | 1.009249743 |
| XPA | 0.665921887 | 0.724989874 |
| LPPR2 | 0.665920571 | 1.107073329 |
| RBM28 | 0.665908516 | 0.892771497 |

| | | |
|---|---|---|
| TGM4 | 0.665884701 | 0.681652491 |
| SOX13 | 0.665708656 | 0.974648852 |
| ADH1B | 0.665695509 | 0.926690968 |
| CHTF8 | 0.665610286 | 0.688132847 |
| NMUR1 | 0.665564523 | 0.826735886 |
| OR13C3 | 0.665494499 | 0.933171324 |
| ZCCHC11 | 0.665416588 | 0.907203115 |
| C2orf42 | 0.665255692 | 0.986229243 |
| TAF1A | 0.665237761 | 0.896314297 |
| ING2 | 0.665068855 | 0.95587281 |
| FOXN3 | 0.665056671 | 0.891628812 |
| HOXA7 | 0.664683829 | 0.87605451 |
| PIGM | 0.6646683 | 0.776022681 |
| MAP3K11 | 0.664649254 | 0.916288125 |
| ZBTB17 | 0.664588581 | 0.996755354 |
| OAZ2 | 0.664545687 | 0.825840421 |
| PCED1A | 0.664437083 | 0.940866748 |
| SLC27A5 | 0.664437083 | 0.801134062 |
| NIN | 0.664421365 | 0.866735183 |
| TAF4 | 0.664185948 | 0.714459295 |
| KLF3 | 0.664178941 | 0.916937054 |
| RNASE11 | 0.664006231 | 0.8327258 |
| RFC3 | 0.663991738 | 0.909149903 |
| TMEM51 | 0.663792894 | 0.696233293 |
| AKT2 | 0.663680803 | 0.79688514 |
| SPAG6 | 0.663651328 | 0.907502569 |
| IFNW1 | 0.663449519 | 0.891401165 |
| TGFBRAP1 | 0.663355521 | 0.746456055 |
| OR5V1 | 0.663332485 | 0.856457691 |
| WAC | 0.663316173 | 0.76053443 |
| PBOV1 | 0.663225101 | 0.959575197 |
| PCCB | 0.663081243 | 0.950681376 |
| BTN1A1 | 0.66261373 | 0.737950587 |
| B3GNT7 | 0.662439922 | 0.853381936 |
| FAM86B1 | 0.662327752 | 0.978128797 |
| VWA1 | 0.662246299 | 0.859862292 |
| SCAF8 | 0.662034863 | 0.916752312 |
| CYP26B1 | 0.661949929 | 0.757391657 |
| SLC25A44 | 0.661850494 | 0.832320778 |
| AATK | 0.66152591 | 1.01752109 |
| HIVEP1 | 0.661449187 | 0.960712839 |
| SIGLEC6 | 0.661437165 | 0.867152693 |
| FUNDC1 | 0.661430614 | 0.971565605 |
| PDLIM3 | 0.661074436 | 0.835902706 |

| | | |
|---|---|---|
| ZNF770 | 0.660916149 | 0.789793439 |
| IL2RG | 0.660884612 | 0.74367294 |
| SLC22A7 | 0.660845546 | 0.824220332 |
| GNAI2 | 0.660825916 | 0.953926022 |
| DHX34 | 0.660762462 | 0.805589307 |
| KBTBD12 | 0.660751205 | 0.82746051 |
| COX7A2 | 0.660712228 | 0.703439325 |
| PRPS1 | 0.660580017 | 0.857235561 |
| AURKA | 0.660457931 | 1.027255029 |
| F5 | 0.660382347 | 0.81855002 |
| APLN | 0.660350569 | 0.390202124 |
| FRMPD4 | 0.660293609 | 0.90927501 |
| PPWD1 | 0.660287904 | 0.799918996 |
| CRTC3 | 0.66020696 | 0.890643985 |
| MAP3K19 | 0.660165684 | 0.9467878 |
| WFIKKN1 | 0.660076451 | 0.884568651 |
| CDK16 | 0.660025817 | 0.890330954 |
| PROS1 | 0.659999633 | 0.909680032 |
| SEC13 | 0.659612641 | 1.004050223 |
| OR11L1 | 0.659545373 | 0.795477903 |
| CCDC34 | 0.659481266 | 0.760226812 |
| ADAMTSL2 | 0.65935091 | 0.970028352 |
| COL22A1 | 0.659318156 | 0.788578372 |
| GNG5 | 0.65930863 | 0.977046934 |
| TMEM100 | 0.65889795 | 1.036452005 |
| PIK3R3 | 0.658807268 | 0.96495782 |
| AKAP8L | 0.658585819 | 0.892277742 |
| PHF19 | 0.658585819 | 1.054510058 |
| NKAIN3 | 0.658395935 | 0.965168084 |
| MAP2K6 | 0.658161195 | 1.006489293 |
| PPIL3 | 0.658147036 | 0.974042829 |
| OAS1 | 0.658131888 | 0.941596366 |
| RPL32 | 0.658108922 | 0.900993491 |
| FIP1L1 | 0.658091433 | 0.943473792 |
| ACVR2B | 0.657964114 | 1.031148605 |
| USP12 | 0.657922376 | 0.966255678 |
| SLC15A3 | 0.657866157 | 0.928311057 |
| FHIT | 0.657723236 | 0.99091499 |
| PPP1R17 | 0.657561628 | 0.948955122 |
| SP8 | 0.657514506 | 0.974991435 |
| UCK1 | 0.657065309 | 0.983776768 |
| ITGA11 | 0.65672836 | 0.80532122 |
| DDX21 | 0.656623726 | 0.895504253 |
| ESM1 | 0.6564029 | 0.748076144 |

| | | |
|---|---|---|
| SAMD7 | 0.656378143 | 0.946132037 |
| EFTUD2 | 0.656107396 | 0.786148238 |
| OR10T2 | 0.65609849 | 0.880095923 |
| CELA1 | 0.656046229 | 0.757300454 |
| DNM3 | 0.655833593 | 0.729850142 |
| VEGFB | 0.655824451 | 0.928617781 |
| FKBP8 | 0.655536062 | 0.940047962 |
| CCDC12 | 0.655480221 | 0.881328473 |
| CLEC3B | 0.655291844 | 0.76508708 |
| KCNQ4 | 0.655239873 | 0.766385464 |
| TAF1 | 0.655188586 | 0.880597015 |
| CYP2C9 | 0.655124786 | 0.864373783 |
| PRPS1L1 | 0.65502254 | 1.019467878 |
| ZBTB16 | 0.65498172 | 0.806399352 |
| MLLT3 | 0.654970372 | 0.865022268 |
| LPP | 0.654912376 | 0.900650908 |
| DZIP3 | 0.654902019 | 1.04412719 |
| DTD1 | 0.654725033 | 0.803564196 |
| PTPN23 | 0.654475091 | 0.987670344 |
| JADE2 | 0.654452452 | 0.775470474 |
| SVIL | 0.654354026 | 0.877697842 |
| HEATR1 | 0.653899842 | 0.904419322 |
| KIF1C | 0.653764657 | 0.848926691 |
| DISP2 | 0.653690784 | 1.037684138 |
| THEG | 0.653572104 | 0.858647226 |
| TBK1 | 0.653540969 | 0.931862427 |
| TAF15 | 0.653533313 | 0.972744971 |
| DTX3 | 0.653478156 | 0.711226476 |
| IL13RA2 | 0.653363147 | 0.745240988 |
| SERPINA4 | 0.653305585 | 0.934791414 |
| C1orf131 | 0.653219954 | 0.751721345 |
| KDELR3 | 0.653206557 | 0.830700689 |
| ARHGAP11A | 0.653111274 | 0.912641316 |
| GTF2H2 | 0.653079206 | 1.042527339 |
| STON1 | 0.653079206 | 0.931956258 |
| UBAC1 | 0.653079206 | 0.884568651 |
| PIP5K1A | 0.652759557 | 0.990266061 |
| ZNF442 | 0.652751833 | 0.972593354 |
| SVOPL | 0.652715392 | 0.872417983 |
| KIF15 | 0.652663789 | 0.89145403 |
| NAA40 | 0.652648507 | 0.859862292 |
| ABCA6 | 0.652631956 | 0.933160286 |
| SLC17A5 | 0.652603637 | 0.785853342 |
| MTFR2 | 0.652556305 | 1.011745646 |

| | | |
|---|---|---|
| UROS | 0.652539444 | 0.784123127 |
| TMEM40 | 0.652447905 | 0.83799109 |
| PTPRN | 0.65237272 | 0.859831278 |
| CXCL6 | 0.652147307 | 1.021871203 |
| UBIAD1 | 0.652143741 | 0.904819765 |
| TRPT1 | 0.652051132 | 0.977723775 |
| ALAD | 0.651841105 | 0.769137303 |
| CCDC105 | 0.651784154 | 0.857837181 |
| ADAM22 | 0.651725577 | 0.934458144 |
| AGBL2 | 0.651518534 | 0.967553537 |
| UTP6 | 0.651421155 | 0.861482382 |
| MAML1 | 0.6512096 | 0.820575132 |
| OAZ3 | 0.6510255 | 1.01752109 |
| SIM1 | 0.650716092 | 0.738760632 |
| PVRIG | 0.650601069 | 0.875253139 |
| UMPS | 0.650438947 | 0.813108371 |
| ANGPTL3 | 0.650300165 | 0.932766302 |
| CCDC42 | 0.650300165 | 0.81855002 |
| NIT1 | 0.649734831 | 0.854191981 |
| SMCP | 0.64971476 | 0.974888619 |
| HERC6 | 0.649632921 | 0.978585334 |
| ZNF222 | 0.649543189 | 0.783830079 |
| TMEM57 | 0.64936195 | 0.958547448 |
| B3GALTL | 0.649111119 | 0.959902795 |
| FUS | 0.648915721 | 0.839714471 |
| ANAPC2 | 0.64885299 | 0.922128488 |
| ZCCHC17 | 0.648752653 | 0.709194006 |
| PRKRA | 0.648556119 | 0.868790682 |
| NOTCH4 | 0.648492145 | 0.922640745 |
| TBX18 | 0.648284207 | 0.82746051 |
| PKN2 | 0.648269236 | 0.766385464 |
| SPRR3 | 0.648252228 | 0.912298732 |
| CSF1 | 0.648210497 | 0.945321993 |
| TBX6 | 0.648064146 | 0.706763872 |
| CENPL | 0.647833591 | 0.605103281 |
| FGF8 | 0.647829383 | 0.874107722 |
| PLA2G6 | 0.647808144 | 0.931862427 |
| PPARGC1A | 0.647806949 | 0.933171324 |
| PDGFRB | 0.647486096 | 0.808565866 |
| DMC1 | 0.647327953 | 0.759247242 |
| GPR157 | 0.647299551 | 0.965606749 |
| CAB39L | 0.647279929 | 0.832134293 |
| C4orf36 | 0.647270345 | 0.956257594 |
| TMF1 | 0.647239883 | 0.860672337 |

| | | |
|---|---|---|
| DGKQ | 0.647106668 | 0.949383517 |
| VPS4B | 0.647026647 | 0.978938842 |
| LMTK2 | 0.646825369 | 0.726800779 |
| MCHR1 | 0.646825369 | 0.908500973 |
| FAM83B | 0.646691104 | 0.778452815 |
| TBC1D24 | 0.646425808 | 0.906439854 |
| FLRT3 | 0.646308038 | 0.912641316 |
| SLC2A2 | 0.646255973 | 0.922640745 |
| TRIM25 | 0.646179214 | 0.826735886 |
| PI4K2B | 0.64610924 | 0.959117456 |
| CCDC22 | 0.64597244 | 0.923045768 |
| ABHD10 | 0.645653708 | 0.869177805 |
| ZNF584 | 0.64528676 | 0.890030832 |
| ARHGEF39 | 0.645202847 | 0.854059609 |
| DNAAF3 | 0.645021693 | 0.748481166 |
| L3MBTL4 | 0.644535119 | 0.8886605 |
| ADORA3 | 0.64452353 | 0.91174562 |
| CCL15 | 0.644477142 | 0.934791414 |
| SLC25A2 | 0.64442391 | 0.874038072 |
| MAPK3 | 0.644408853 | 0.994808566 |
| RNF10 | 0.644408853 | 0.994808566 |
| B4GALNT2 | 0.64383443 | 0.801134062 |
| POU3F1 | 0.64364045 | 0.93600648 |
| OR7G3 | 0.643559099 | 0.833535844 |
| C4orf32 | 0.643430424 | 0.888213852 |
| ENPP4 | 0.643310816 | 0.904414743 |
| MYH3 | 0.643289727 | 0.788983394 |
| CCDC11 | 0.642822045 | 0.892264075 |
| MCM3 | 0.642497226 | 1.022712524 |
| NR4A2 | 0.642357852 | 0.948085659 |
| MIOX | 0.642270006 | 0.934791414 |
| MAPK8IP1 | 0.64225872 | 0.971565605 |
| HERC3 | 0.641996207 | 0.940298507 |
| RAB28 | 0.641674821 | 0.890643985 |
| DUPD1 | 0.641536983 | 0.926687222 |
| KRIT1 | 0.641518741 | 0.919805589 |
| B4GALT3 | 0.641504121 | 0.86836776 |
| LDHA | 0.641341978 | 0.842851357 |
| MCC | 0.641288913 | 0.888317917 |
| CPT2 | 0.641236379 | 0.920210612 |
| DNASE1L1 | 0.641192567 | 0.724853991 |
| RGS6 | 0.641123547 | 0.884973674 |
| PRTFDC1 | 0.641111314 | 0.941596366 |
| C3orf65 | 0.640939032 | 0.874272011 |

| | | |
|---|---|---|
| SMAD2 | 0.640860339 | 0.753341434 |
| PLEKHA1 | 0.64085086 | 0.862281603 |
| TACR2 | 0.640475861 | 0.967553537 |
| LCLAT1 | 0.640228809 | 0.942894225 |
| RELN | 0.640149532 | 0.834523037 |
| OSBPL9 | 0.640112631 | 0.897225077 |
| CHD6 | 0.64011162 | 0.844471446 |
| PDE8A | 0.64011162 | 0.844471446 |
| RBM8A | 0.640103387 | 0.957862282 |
| C1orf141 | 0.640101585 | 0.692993115 |
| OR10H1 | 0.639940308 | 0.889833941 |
| PGGT1B | 0.63987215 | 0.90927501 |
| IGF2BP1 | 0.639775211 | 0.938436614 |
| ICA1L | 0.639747296 | 0.861077359 |
| AGMAT | 0.639736089 | 0.864317537 |
| SPATA45 | 0.639422203 | 0.903048989 |
| TSHZ3 | 0.639377419 | 0.884163629 |
| KIF4A | 0.638946625 | 0.794289422 |
| GPR55 | 0.638810462 | 0.961713173 |
| HS3ST2 | 0.638537149 | 0.778066191 |
| WDR36 | 0.638453393 | 0.452209661 |
| OGT | 0.638335387 | 0.814499797 |
| HEATR4 | 0.638023175 | 0.811664642 |
| NCAPD2 | 0.637859606 | 0.698663426 |
| CCDC112 | 0.637836988 | 0.950992305 |
| CLC | 0.637829334 | 0.988319273 |
| DEPDC7 | 0.637818803 | 0.967193196 |
| RHBDD2 | 0.637801228 | 0.983394087 |
| TUBA1B | 0.637792059 | 0.744025921 |
| ZNF391 | 0.637780903 | 1.002835156 |
| BIRC7 | 0.637634969 | 0.93835172 |
| SLC13A5 | 0.637433237 | 0.757391657 |
| LRRC40 | 0.637269515 | 0.997164844 |
| C6orf136 | 0.637166193 | 0.920863309 |
| PRDX5 | 0.63704497 | 0.880923451 |
| SLC36A3 | 0.636752217 | 0.855407047 |
| FOXN1 | 0.636551348 | 0.872809864 |
| LRP3 | 0.636547397 | 0.998702141 |
| FKBP10 | 0.63641372 | 0.761846902 |
| CCDC7 | 0.636119949 | 0.834532374 |
| PARP16 | 0.636075528 | 0.762251924 |
| PAX5 | 0.635225862 | 1.042829332 |
| SPAG16 | 0.635174804 | 0.980963953 |
| WDR48 | 0.635038997 | 0.903199676 |

| | | |
|---|---|---|
| DUSP12 | 0.634969425 | 0.959117456 |
| GBA3 | 0.634871659 | 0.685702714 |
| HOXA9 | 0.634702894 | 0.880923451 |
| IL36A | 0.634504482 | 0.803159174 |
| PAEP | 0.634486194 | 0.796678817 |
| ELOVL7 | 0.634111873 | 0.976704351 |
| LRSAM1 | 0.634096552 | 0.977287476 |
| FCGRT | 0.633853467 | 0.80072904 |
| PRTN3 | 0.633728896 | 0.868267359 |
| KSR2 | 0.633698684 | 0.927319922 |
| TIA1 | 0.633688772 | 0.913669065 |
| DNMBP | 0.633529282 | 0.886947585 |
| GART | 0.633418437 | 0.928311057 |
| LRRC17 | 0.633217779 | 1.056526208 |
| APOBEC1 | 0.633208156 | 0.873228028 |
| ZNF24 | 0.633022276 | 0.96800324 |
| C11orf80 | 0.63301978 | 0.690968003 |
| ZBTB41 | 0.632871442 | 0.863912515 |
| PLGL | 0.632824823 | 0.953926022 |
| AMIGO2 | 0.632756844 | 0.885577252 |
| HTR6 | 0.632748591 | 0.810512654 |
| TNNT1 | 0.632512172 | 0.885919836 |
| POU1F1 | 0.632451405 | 1.022063595 |
| COX7C | 0.632445684 | 0.838396112 |
| DHRS1 | 0.632337799 | 0.895522388 |
| OR51G2 | 0.632330355 | 0.933171324 |
| ENPP2 | 0.63227301 | 0.836469825 |
| MME | 0.632231766 | 0.802725503 |
| PDE4C | 0.631587342 | 0.699068449 |
| FGF11 | 0.631439817 | 0.905256327 |
| FAM127C | 0.631176458 | 0.925070879 |
| TAF6 | 0.631173657 | 0.970838396 |
| SLC35C1 | 0.631093681 | 0.774807614 |
| TMPPE | 0.631085872 | 0.917437479 |
| RPL11 | 0.630933314 | 0.976704351 |
| SLC17A3 | 0.630712139 | 0.811259619 |
| TTC37 | 0.630676726 | 0.908187736 |
| STX11 | 0.630651217 | 0.893456663 |
| NXPH1 | 0.630533366 | 0.873929428 |
| CRB1 | 0.630286293 | 0.874272011 |
| CCDC53 | 0.63028475 | 0.87727825 |
| UBTD2 | 0.63016606 | 0.978938842 |
| ADCK3 | 0.630116393 | 0.893884164 |
| MRPL43 | 0.630094573 | 0.780012979 |

| | | |
|---|---|---|
| CXCL8 | 0.629990739 | 0.936279548 |
| PAPD5 | 0.629970246 | 0.684487647 |
| HELZ2 | 0.629951602 | 0.720534629 |
| OR51A2 | 0.62993238 | 1.022610483 |
| OR10A2 | 0.629905399 | 0.979343864 |
| TIMM8A | 0.629834592 | 0.78290806 |
| RALGDS | 0.62966528 | 0.944158959 |
| KRT31 | 0.629664758 | 0.691373026 |
| LTBP2 | 0.629448519 | 0.870503597 |
| SOST | 0.629399461 | 1.052247874 |
| LPAR1 | 0.62934362 | 0.8403634 |
| POU3F4 | 0.628753693 | 0.741595788 |
| ASMT | 0.628670864 | 0.925475901 |
| TRAF4 | 0.628669805 | 0.985723556 |
| PCTP | 0.628524054 | 0.971648441 |
| UBTF | 0.6284216 | 0.935107073 |
| TUBB4Q_pseudogene | 0.628127383 | 0.867152693 |
| DNAJC18 | 0.628124578 | 0.961064244 |
| ZNF137P | 0.62808714 | 0.98217902 |
| SPRY4 | 0.628087056 | 0.92428914 |
| KIF16B | 0.628029578 | 0.798298906 |
| AGXT2 | 0.627994425 | 0.859182349 |
| MRC2 | 0.627971251 | 0.969218307 |
| ANKRD12 | 0.627885746 | 0.823227133 |
| SLC31A2 | 0.627872631 | 0.90036452 |
| C17orf82 | 0.627803302 | 0.749291211 |
| TFAP2C | 0.627620172 | 0.621304172 |
| COG4 | 0.62754371 | 0.75617659 |
| TAPBPL | 0.62713927 | 0.957862282 |
| DHX29 | 0.627035786 | 0.796273795 |
| RFTN2 | 0.626792547 | 0.776832726 |
| FASN | 0.626758918 | 1.201817002 |
| EP400 | 0.626517577 | 0.790339157 |
| GFM2 | 0.626260782 | 0.958687728 |
| PARP6 | 0.626233478 | 1.054272985 |
| KRT24 | 0.62612071 | 0.922233642 |
| ZNF223 | 0.626058652 | 1.061158364 |
| GPAA1 | 0.626011094 | 0.96430889 |
| PCNT | 0.625977079 | 0.947242206 |
| IL18 | 0.625903134 | 0.964713943 |
| ALDH3B1 | 0.625541934 | 1.118754056 |
| NRP2 | 0.625495092 | 0.794289422 |
| RHCE | 0.62535257 | 0.79181855 |
| PYGB | 0.625187676 | 0.735925476 |

| | | |
|---|---|---|
| PSG8 | 0.625132661 | 0.887808829 |
| GEM | 0.62504973 | 0.749291211 |
| SAMD8 | 0.625033506 | 0.727646454 |
| CHST7 | 0.624999918 | 0.846203764 |
| PGRMC2 | 0.624947646 | 0.742810855 |
| CPN2 | 0.624900679 | 0.957472661 |
| ASPHD1 | 0.62485035 | 0.693398137 |
| THAP2 | 0.624794418 | 0.690157959 |
| ROPN1L | 0.624607236 | 0.855288774 |
| SIM2 | 0.624591677 | 1.043478261 |
| CMSS1 | 0.624011977 | 0.991899554 |
| TSPAN1 | 0.623921558 | 1.025111381 |
| KCTD11 | 0.623800723 | 0.831106543 |
| PAPPA2 | 0.623742266 | 0.993510707 |
| JAG1 | 0.623650353 | 0.787363305 |
| SSX1 | 0.62364154 | 0.72782503 |
| PIFO | 0.623502779 | 0.992709599 |
| QPCT | 0.623375072 | 0.771162414 |
| TMEM88 | 0.62331863 | 0.959902795 |
| FBXL16 | 0.623267343 | 1.028552888 |
| SOAT1 | 0.62325022 | 0.985723556 |
| NUP35 | 0.623234579 | 0.724989874 |
| ZNF32 | 0.623127707 | 1.003240178 |
| CLK4 | 0.623058242 | 0.805970149 |
| SCP2D1 | 0.622964398 | 0.911956149 |
| OR9Q2 | 0.622932911 | 0.970433374 |
| PTEN | 0.622874332 | 1.029201817 |
| ZNF71 | 0.6228248 | 0.852346694 |
| XPO1 | 0.622673463 | 0.737832576 |
| TIFA | 0.622420198 | 0.62980964 |
| BPGM | 0.622374877 | 0.944192083 |
| SAP30 | 0.622374877 | 0.944192083 |
| S100A9 | 0.622365768 | 0.907845152 |
| RLBP1 | 0.622214502 | 0.709599028 |
| LDHD | 0.622151947 | 0.865532604 |
| GAS2L3 | 0.622092296 | 0.809234508 |
| TEKT5 | 0.622011251 | 0.892264075 |
| SMYD1 | 0.621439109 | 0.765087605 |
| NELFA | 0.621399088 | 0.856800274 |
| MAGEB10 | 0.621364616 | 1.055893074 |
| TLE6 | 0.621221678 | 0.9963548 |
| CNTF | 0.621150871 | 0.710814095 |
| VTN | 0.621060988 | 0.947242206 |
| TBR1 | 0.620930402 | 0.863912515 |

| | | |
|---|---|---|
| FLCN | 0.620605251 | 0.734710409 |
| BST1 | 0.620539334 | 0.811259619 |
| CFHR1 | 0.620511368 | 0.921020656 |
| PACSIN2 | 0.6202272 | 0.893456663 |
| BHLHA15 | 0.620161205 | 0.901579587 |
| ELAC1 | 0.620080261 | 0.881733495 |
| LGALS9C | 0.620066406 | 0.958282706 |
| FAM3A | 0.619809899 | 0.831449126 |
| WFIKKN2 | 0.619738351 | 0.712434184 |
| IMP4 | 0.61966189 | 0.935601458 |
| SMARCA4 | 0.619637218 | 0.784555483 |
| EBF3 | 0.619630867 | 0.632644795 |
| ITGA3 | 0.61943742 | 0.862426995 |
| PDCL | 0.619365367 | 0.786148238 |
| SLC20A1 | 0.619341017 | 0.829485622 |
| DHRSX | 0.619272758 | 0.722964763 |
| CELA2A | 0.619245524 | 1.0266061 |
| METTL18 | 0.619239799 | 0.736330498 |
| OR9A2 | 0.61923116 | 0.924974306 |
| GCG | 0.619081593 | 0.692017814 |
| CNN3 | 0.619042173 | 1.013018157 |
| RRN3 | 0.619015101 | 0.923426347 |
| SPO11 | 0.618971635 | 0.69046074 |
| MPP4 | 0.618926008 | 0.723369785 |
| CETP | 0.618920863 | 0.89671932 |
| ACACB | 0.618899315 | 0.823410288 |
| GUCA1A | 0.618877411 | 0.880113406 |
| CCDC127 | 0.618683176 | 1.070473876 |
| USP6 | 0.618569733 | 0.69954575 |
| GPR35 | 0.618548823 | 0.950032446 |
| ATRAID | 0.618464295 | 0.931140802 |
| ZNF786 | 0.618436167 | 0.957472661 |
| ZNF219 | 0.618168069 | 1.054678007 |
| POGZ | 0.618155181 | 0.984609154 |
| CREB5 | 0.618150444 | 0.901174565 |
| KCNJ13 | 0.618107982 | 0.976638546 |
| TLN1 | 0.617919837 | 0.94491697 |
| IFNA14 | 0.617769563 | 0.796505653 |
| CD80 | 0.617749188 | 0.855002025 |
| TAF10 | 0.617619443 | 0.768327258 |
| ZNF713 | 0.617557857 | 0.618063994 |
| SLC41A3 | 0.617095982 | 0.8137573 |
| ITGA2 | 0.61705197 | 0.772377481 |
| PIP5KL1 | 0.616983529 | 0.909149903 |

| | | |
|---|---|---|
| PDE1B | 0.616885293 | 0.765897124 |
| ETV5 | 0.616749487 | 0.618874038 |
| KCMF1 | 0.616587798 | 0.8886605 |
| MAP3K3 | 0.616356308 | 0.875405581 |
| DEFB131 | 0.616324748 | 0.883758607 |
| E2F6 | 0.616211813 | 0.954232483 |
| LRPAP1 | 0.616140539 | 0.936404932 |
| NID1 | 0.616136524 | 0.663021466 |
| PPP1R13L | 0.615928993 | 0.94127177 |
| TMED2 | 0.615760432 | 0.77399757 |
| UBE2J1 | 0.615678706 | 0.841661259 |
| SPNS3 | 0.61559126 | 0.804374241 |
| LLGL1 | 0.615535684 | 1.00891049 |
| ZNF91 | 0.615456167 | 0.968813285 |
| SLC29A4 | 0.615382744 | 0.855087359 |
| ZFP36L1 | 0.615197034 | 0.885378696 |
| ST8SIA6 | 0.61514589 | 0.811664642 |
| HMMR | 0.615025773 | 0.771567436 |
| U2AF1L4 | 0.614962251 | 0.852166869 |
| MGST2 | 0.614837804 | 0.835560956 |
| GZF1 | 0.614546936 | 0.85945727 |
| A4GALT | 0.614252785 | 0.800129786 |
| ZNF256 | 0.614160608 | 0.763275094 |
| OR2B11 | 0.614118379 | 1.056526208 |
| ZMAT5 | 0.614116491 | 0.786148238 |
| TP53I3 | 0.613900199 | 0.914135277 |
| ANKRD23 | 0.613447526 | 0.943816375 |
| CDH23 | 0.61338872 | 0.661907852 |
| ADH7 | 0.61323329 | 0.888213852 |
| TPRN | 0.613164023 | 0.24049332 |
| NCF2 | 0.613070282 | 0.969218307 |
| USP20 | 0.612908035 | 0.828033744 |
| UTS2B | 0.612764997 | 0.946132037 |
| RARA | 0.612761911 | 1.002595717 |
| LAMB4 | 0.612601777 | 1.624186365 |
| FAM178B | 0.612146279 | 0.864679685 |
| KCNH4 | 0.612115015 | 0.855288774 |
| GFER | 0.612023966 | 0.890330954 |
| NACA | 0.611504831 | 0.7995889 |
| DMTF1 | 0.611448691 | 0.961064244 |
| MED22 | 0.611238547 | 0.756766016 |
| LINC00303 | 0.61119227 | 0.9181855 |
| LRRN4CL | 0.61119227 | 0.9181855 |
| POLK | 0.610802701 | 0.988319273 |

| | | |
|---|---|---|
| MYF6 | 0.610548437 | 0.787363305 |
| IL23R | 0.610473243 | 0.883841661 |
| FBXO17 | 0.61026299 | 0.857884491 |
| ADAT2 | 0.610130932 | 0.716889429 |
| STYK1 | 0.609041313 | 0.780661908 |
| SNX1 | 0.608982174 | 0.468996232 |
| RFX5 | 0.608748078 | 0.939000649 |
| FPGS | 0.608506459 | 0.763789747 |
| LCK | 0.608475999 | 0.895522388 |
| SYN1 | 0.608353934 | 0.817334953 |
| GC | 0.608299745 | 0.922128488 |
| CASC10 | 0.608282414 | 0.742810855 |
| SLC39A4 | 0.608270603 | 0.904607398 |
| R3HDM1 | 0.608105448 | 0.90927501 |
| CNKSR3 | 0.608096839 | 0.855002025 |
| NANP | 0.608008157 | 0.992861778 |
| FAM83D | 0.607599714 | 1.008505468 |
| SNTA1 | 0.607580114 | 0.899280576 |
| ESRRG | 0.607251487 | 0.778047793 |
| CXXC4 | 0.607203954 | 0.841041453 |
| NAP1L3 | 0.607089366 | 0.943473792 |
| ZNF208 | 0.607073281 | 0.948612539 |
| SRFBP1 | 0.607035195 | 0.893884164 |
| COL10A1 | 0.606859782 | 0.795477903 |
| CYP2A7 | 0.606739691 | 0.635884974 |
| ANKRD33 | 0.606643109 | 0.925070879 |
| GBP7 | 0.606430688 | 0.69744836 |
| PGM5 | 0.606430688 | 0.782503038 |
| RCOR3 | 0.606330794 | 0.983394087 |
| RFXANK | 0.606256318 | 1.004542505 |
| GTF3C6 | 0.606192825 | 1.052415211 |
| DENND4A | 0.606177464 | 0.776022681 |
| NADK | 0.605981441 | 0.828682674 |
| FCN3 | 0.60597187 | 0.963547995 |
| SBK1 | 0.605917641 | 0.916937054 |
| WNT5A | 0.605915304 | 0.868448099 |
| E2F2 | 0.605774645 | 1.031797534 |
| EXOC3 | 0.605741518 | 0.997974889 |
| MT1A | 0.605724958 | 0.907159986 |
| VNN3 | 0.605647703 | 0.714471123 |
| DDX39B | 0.605596168 | 0.846852693 |
| ESCO2 | 0.605533071 | 0.875658161 |
| EFNB2 | 0.605480966 | 0.917780062 |
| PDE5A | 0.605445454 | 0.947347104 |

| | | |
|---|---|---|
| PDE6A | 0.60501793 | 0.785853342 |
| CTSL3P | 0.604872275 | 0.914135277 |
| RAD54L | 0.604631175 | 0.682462535 |
| CCL3L3 | 0.604559961 | 0.945321993 |
| POLR3H | 0.604553186 | 0.784933171 |
| GABARAP | 0.604048821 | 0.928617781 |
| C19orf57 | 0.603915975 | 0.761846902 |
| VIM | 0.603882443 | 0.843096951 |
| MLN | 0.60371955 | 0.960307817 |
| GRHL3 | 0.603495107 | 0.953827461 |
| NFYB | 0.603313811 | 0.724989874 |
| HNF4G | 0.603310755 | 0.755771567 |
| MGC50273 | 0.603173076 | 0.975334018 |
| PRKAG1 | 0.602751983 | 1.010382868 |
| SLC28A3 | 0.602564176 | 0.965573107 |
| GOT1L1 | 0.602298676 | 0.606318348 |
| KPNA4 | 0.60222248 | 0.913692408 |
| PANK2 | 0.60205594 | 0.842959117 |
| BCKDHA | 0.601765841 | 0.884568651 |
| CDK5 | 0.601697999 | 0.976638546 |
| TRIM67 | 0.601659554 | 0.923426347 |
| EIF5 | 0.601514961 | 1.039287161 |
| ANGPTL7 | 0.601207843 | 0.761846902 |
| SREBF2 | 0.601127612 | 0.861482382 |
| LHX4 | 0.600613555 | 0.818299805 |
| HOXC13 | 0.600514292 | 0.89671932 |
| GDF2 | 0.599731698 | 0.835965978 |
| DPPA3 | 0.599672811 | 0.966788173 |
| LGI3 | 0.59942728 | 0.925880923 |
| PTGFRN | 0.599296211 | 0.957067639 |
| RFESD | 0.599014308 | 0.861077359 |
| NTMT1 | 0.598567715 | 0.999594978 |
| FAHD2A | 0.598409351 | 0.955223881 |
| SLC35F1 | 0.598337591 | 0.965056526 |
| PIN1 | 0.598281024 | 0.717294451 |
| INSM1 | 0.598238998 | 0.803564196 |
| JPH3 | 0.598146475 | 0.830421377 |
| NDUFS2 | 0.598014454 | 0.852166869 |
| NFKBIL1 | 0.597878201 | 0.92926671 |
| KRBA1 | 0.597567454 | 0.842446335 |
| PKP4 | 0.59747177 | 0.979445015 |
| RBP1 | 0.597396702 | 0.766707169 |
| HSPA4 | 0.596787583 | 0.957819598 |
| ASL | 0.59670526 | 0.966904607 |

| | | |
|---|---|---|
| FAM69C | 0.596394709 | 0.920210612 |
| C1orf35 | 0.596289416 | 0.369647139 |
| FAN1 | 0.595957489 | 0.872417983 |
| CCL4 | 0.595799574 | 0.646111682 |
| SYPL2 | 0.595749371 | 0.95101062 |
| ALAS1 | 0.595437441 | 0.817739976 |
| TOX2 | 0.595437441 | 0.817739976 |
| ATP2B2 | 0.595342336 | 0.85945727 |
| BOD1 | 0.595236635 | 0.956662616 |
| LPL | 0.595082694 | 0.748886189 |
| KRT37 | 0.595049022 | 0.828675577 |
| NKX3-2 | 0.595049022 | 0.828675577 |
| DEFB103A | 0.594688447 | 0.98663926 |
| SECTM1 | 0.594300506 | 1.000342583 |
| PGF | 0.594257463 | 0.770757392 |
| ERCC1 | 0.594085158 | 0.944192083 |
| RFT1 | 0.594074435 | 0.906439854 |
| SOS1 | 0.59392136 | 0.993519644 |
| CCNA1 | 0.593834201 | 0.95544755 |
| SOX4 | 0.593728398 | 0.719319563 |
| UTP14A | 0.593619412 | 0.925659472 |
| BRF1 | 0.593313236 | 0.730255164 |
| MDN1 | 0.593170043 | 0.911300122 |
| XRCC5 | 0.593139071 | 0.709599028 |
| ILF3 | 0.592862741 | 0.946138871 |
| CTNNA2 | 0.59281716 | 0.777663583 |
| C4orf17 | 0.592743979 | 1.037262049 |
| ZRANB2 | 0.592436097 | 0.884568651 |
| AMBP | 0.592415415 | 1.093560146 |
| AIP | 0.592395854 | 0.703523694 |
| CCNJ | 0.592282326 | 0.157245632 |
| TEAD2 | 0.592165019 | 0.865022268 |
| MBOAT7 | 0.59198059 | 0.829485622 |
| YPEL4 | 0.591781974 | 0.791413528 |
| PAGE3 | 0.591697719 | 0.990279465 |
| SLC12A2 | 0.591682641 | 0.669501823 |
| EN1 | 0.591619475 | 0.948562171 |
| C6orf201 | 0.591369519 | 0.900308325 |
| DPH7 | 0.591269872 | 0.861887404 |
| THRSP | 0.591231667 | 0.963142973 |
| ARVCF | 0.591225351 | 0.827055488 |
| UTP18 | 0.591142522 | 1.012150668 |
| UNK | 0.591122016 | 0.985074627 |
| FAM168B | 0.591088678 | 0.813284731 |

| | | |
|---|---|---|
| GMPPA | 0.59100208 | 0.687727825 |
| GSS | 0.590891511 | 0.931213498 |
| DGKH | 0.590863768 | 0.940298507 |
| KRT8 | 0.590826591 | 0.831105711 |
| IGFN1 | 0.590826302 | 0.801301816 |
| SEPSECS | 0.590012516 | 0.849736736 |
| ERLIN2 | 0.590004824 | 0.888213852 |
| CORO2A | 0.589829083 | 1.100034258 |
| ARL4A | 0.589612291 | 0.960415315 |
| TRPC3 | 0.588824128 | 0.952628164 |
| PDK2 | 0.588431117 | 0.935107073 |
| EXT1 | 0.588023041 | 0.808565866 |
| UGT1A10 | 0.588023041 | 0.926670993 |
| ARL6IP1 | 0.587860294 | 0.89145403 |
| ACOT13 | 0.587627066 | 0.963659961 |
| SOD2 | 0.587569476 | 0.78655326 |
| MARCH6 | 0.587299749 | 1.055158988 |
| PBX1 | 0.587238994 | 0.9733939 |
| UGT3A2 | 0.587184249 | 0.909798832 |
| HIRA | 0.587167915 | 0.920615634 |
| AP1G1 | 0.587158037 | 0.914696814 |
| SCIMP | 0.586893792 | 0.8886605 |
| FAM154A | 0.586879084 | 0.889428919 |
| DEFB116 | 0.586527851 | 0.893479141 |
| TRIM42 | 0.586499649 | 1.001946788 |
| BBS5 | 0.586328013 | 0.879708384 |
| WDR74 | 0.586302899 | 0.756581612 |
| SALL1 | 0.585885598 | 0.84163629 |
| RARG | 0.585431873 | 0.814094775 |
| STAR | 0.58527997 | 0.891859052 |
| PPP1R3F | 0.585167224 | 0.949640288 |
| OR2T34 | 0.585036687 | 0.883521754 |
| TMEM115 | 0.584958945 | 0.846496557 |
| CD247 | 0.584765983 | 0.81855002 |
| HN1 | 0.584039921 | 0.417266187 |
| UNKL | 0.583933881 | 0.933160286 |
| GLIS1 | 0.583428141 | 0.657756177 |
| BTN2A1 | 0.582796807 | 0.27166838 |
| PAIP2 | 0.582489874 | 0.931551235 |
| AP3B2 | 0.582017754 | 0.954779034 |
| DOLK | 0.581714361 | 0.641960308 |
| TPPP2 | 0.581131351 | 0.848521669 |
| ZNF141 | 0.581072324 | 0.88420692 |
| IVL | 0.580846592 | 0.871188763 |

| | | |
|---|---|---|
| AGPAT9 | 0.5808279 | 0.788578372 |
| SH3GLB1 | 0.580472969 | 0.949297705 |
| ATP5A1 | 0.580331182 | 0.991899554 |
| CALB2 | 0.579993956 | 0.797533402 |
| TEAD1 | 0.579910722 | 0.807614419 |
| PIGK | 0.579830809 | 0.838416613 |
| LYZL6 | 0.579134292 | 0.872823005 |
| WASH5P | 0.577937444 | 0.889003083 |
| PLEKHA6 | 0.577883201 | 0.906844876 |
| KRTAP5-6 | 0.577727343 | 0.905447071 |
| SETD2 | 0.577608787 | 0.671526934 |
| RPP30 | 0.5772844 | 1.097205346 |
| PLXNA3 | 0.57678699 | 0.815055159 |
| KCNS2 | 0.576752332 | 0.981829981 |
| C19orf24 | 0.576387292 | 0.92709599 |
| MRPL17 | 0.576284957 | 0.958890031 |
| VASH2 | 0.575950735 | 0.852571891 |
| OAF | 0.575916044 | 1.017415958 |
| FOXA1 | 0.575611003 | 0.983776768 |
| HS3ST3B1 | 0.575433102 | 1.030499676 |
| BLM | 0.57522336 | 0.720939652 |
| TMEM125 | 0.575012162 | 0.839611179 |
| NCOA2 | 0.574779291 | 0.864373783 |
| VPS41 | 0.574695239 | 0.715768981 |
| BRINP1 | 0.574648296 | 1.034021871 |
| INSL5 | 0.574236193 | 0.789311408 |
| TTC23L | 0.574002985 | 0.381294964 |
| RORA | 0.573716499 | 0.564601053 |
| KHSRP | 0.573625927 | 0.751721345 |
| ZNF267 | 0.573497919 | 0.952723535 |
| DNAH11 | 0.5733225 | 0.726800779 |
| DNAH2 | 0.573107763 | 0.838801134 |
| ZNF416 | 0.572821276 | 1.008505468 |
| DENND2C | 0.5727657 | 0.745646011 |
| SYCP2L | 0.572612217 | 0.832320778 |
| MARCKS | 0.572607684 | 0.659376266 |
| PCOLCE | 0.572331741 | 0.260363138 |
| CYP17A1 | 0.572163282 | 0.81855002 |
| TPM1 | 0.572104395 | 0.901579587 |
| CENPE | 0.571801708 | 0.869582827 |
| OR5H15 | 0.571457084 | 0.91538198 |
| CSRP2 | 0.571302352 | 0.904761905 |
| SLC4A1 | 0.571260163 | 0.61036857 |
| ZFAND5 | 0.571244167 | 1.000648929 |

| | | |
|---|---|---|
| TAF1B | 0.570860385 | 0.849331713 |
| SLC23A2 | 0.570571146 | 0.802754151 |
| CTBP1 | 0.570247829 | 0.701903605 |
| DCAF6 | 0.570169891 | 0.825030377 |
| FCGR2B | 0.569975237 | 0.909149903 |
| C1orf50 | 0.569331415 | 0.89509923 |
| UTRN | 0.569256391 | 0.853990915 |
| EGR4 | 0.569132494 | 0.732280275 |
| HOPX | 0.568893286 | 0.845837616 |
| PDPR | 0.568856711 | 0.995457495 |
| TAX1BP1 | 0.568856223 | 0.780404248 |
| HAVCR2 | 0.568851406 | 0.884973674 |
| HLA-DMA | 0.568779937 | 0.66018631 |
| AMY2A | 0.568328526 | 0.949383517 |
| XKR7 | 0.568273235 | 1.009315512 |
| SPX | 0.568178894 | 0.842446335 |
| IDUA | 0.567974114 | 0.813689753 |
| RAB5A | 0.567754307 | 0.784933171 |
| ROPN1 | 0.567517741 | 0.75617659 |
| NCSTN | 0.567298116 | 1.045220966 |
| ZKSCAN5 | 0.567176741 | 0.970149254 |
| NGFR | 0.566880887 | 0.782503038 |
| DCX | 0.566872478 | 0.772182254 |
| BYSL | 0.566798034 | 0.90647482 |
| JMJD7-PLA2G4B | 0.566537703 | 0.85581207 |
| WDR11 | 0.566501536 | 0.917780478 |
| PPOX | 0.566444229 | 0.990266061 |
| SH2D2A | 0.566369949 | 0.978417266 |
| HPSE | 0.566227477 | 1.016605913 |
| SDPR | 0.566135608 | 0.828023296 |
| LPAR5 | 0.565508708 | 1.152498378 |
| CDK14 | 0.565368841 | 0.774821544 |
| FLT3LG | 0.565353111 | 0.381637547 |
| TCEAL1 | 0.565332783 | 0.992212849 |
| TNFRSF14 | 0.565189928 | 0.84690158 |
| NTN3 | 0.565144914 | 0.950325454 |
| CLPSL2 | 0.56509197 | 0.755771567 |
| ZNF706 | 0.564823413 | 0.778047793 |
| OXT | 0.564813972 | 0.997601918 |
| RAB17 | 0.564607885 | 0.822195221 |
| USP26 | 0.564568001 | 0.927319922 |
| CYB5R1 | 0.564232405 | 0.906844876 |
| SLC44A4 | 0.564225274 | 0.903199676 |
| RAB3B | 0.564112506 | 0.861778066 |

| | | |
|---|---|---|
| TMEM19 | 0.56406573 | 0.842069202 |
| CRIPAK | 0.563893399 | 0.761036857 |
| GGH | 0.563857716 | 0.947436729 |
| RNF122 | 0.563451451 | 0.900713822 |
| IGLL3P | 0.563237289 | 0.912110166 |
| RPTOR | 0.563192411 | 0.897529364 |
| TTBK2 | 0.562941458 | 0.958468527 |
| PKIG | 0.562822178 | 0.865127582 |
| RPRML | 0.562711281 | 0.835965978 |
| FAM9A | 0.562438751 | 0.836371 |
| TAZ | 0.562365295 | 0.693398137 |
| RND2 | 0.562004291 | 1.0266061 |
| KLHL30 | 0.561840153 | 0.716079384 |
| SDC3 | 0.561790146 | 0.889688249 |
| ERP44 | 0.561477764 | 0.962737951 |
| MGAT4C | 0.561391523 | 0.84528149 |
| LOXL3 | 0.561228107 | 0.768327258 |
| CTNNAL1 | 0.561072804 | 0.915724563 |
| SMARCD2 | 0.561017564 | 0.87605451 |
| NAT10 | 0.560869794 | 0.857235561 |
| EME1 | 0.560846483 | 0.913326482 |
| C9orf9 | 0.560569369 | 0.836245289 |
| GNL2 | 0.560472914 | 0.886437378 |
| NSMCE4A | 0.560300893 | 1.00526529 |
| PLCL2 | 0.560175071 | 0.865532604 |
| SMC2 | 0.560166921 | 0.825030377 |
| AAMDC | 0.560089899 | 0.73673552 |
| ELP5 | 0.55947118 | 0.947752126 |
| HSD3B2 | 0.559263853 | 0.612393682 |
| LGALSL | 0.559180494 | 0.971565605 |
| SSX4B | 0.55913967 | 0.80507023 |
| NQO2 | 0.558937989 | 0.778857837 |
| OR2J2 | 0.55861294 | 0.775617659 |
| FKBP1A | 0.55831387 | 0.86472256 |
| STAU1 | 0.558280249 | 0.890643985 |
| FAM115A | 0.558279231 | 0.920210612 |
| C6orf195 | 0.558044311 | 0.862281603 |
| UQCRC1 | 0.558017294 | 0.852044127 |
| ACER1 | 0.557965439 | 0.798703929 |
| GBA2 | 0.557739163 | 0.776832726 |
| SRSF5 | 0.5576924 | 0.965573107 |
| LITAF | 0.55734835 | 0.847711624 |
| HRAS | 0.557281901 | 0.968202466 |
| HBB | 0.557168624 | 0.853341986 |

| | | |
|---|---|---|
| CNOT1 | 0.55688836 | 0.955852572 |
| PRPF18 | 0.556836615 | 0.844256976 |
| SIGLEC1 | 0.556585572 | 0.900769542 |
| CIDEA | 0.556469836 | 0.889833941 |
| PATE2 | 0.556311055 | 0.875253139 |
| MALT1 | 0.556238043 | 0.624269955 |
| MMP20 | 0.556150615 | 0.826086957 |
| CDR1 | 0.556072301 | 0.150736554 |
| SFRP5 | 0.555917287 | 0.90927501 |
| NTSR1 | 0.555681785 | 1.038286827 |
| RAX2 | 0.554961955 | 0.851356825 |
| TAF7L | 0.554786931 | 0.895522388 |
| MYO1E | 0.554702466 | 0.892669097 |
| PLCD4 | 0.554343703 | 0.838416613 |
| CIB1 | 0.553899139 | 0.890238963 |
| SKP1 | 0.553668405 | 0.955223881 |
| SP4 | 0.553524669 | 0.984425698 |
| AADACL4 | 0.553371195 | 0.961927906 |
| HHAT | 0.55323274 | 0.771972458 |
| SNRPD2 | 0.553064739 | 0.921425678 |
| PPIH | 0.553059543 | 0.8403634 |
| NAGA | 0.552963554 | 0.859831278 |
| MED14 | 0.55290005 | 0.787363305 |
| GCK | 0.552842209 | 1.062946139 |
| DLAT | 0.552803722 | 0.903199676 |
| LRRC8D | 0.55251265 | 0.8327258 |
| RNF214 | 0.552504931 | 0.773523686 |
| C17orf50 | 0.552165187 | 0.878088295 |
| FAM83H | 0.552081951 | 0.971648441 |
| ZNF549 | 0.551851905 | 0.874848117 |
| BLOC1S5 | 0.551834534 | 0.987039287 |
| C19orf71 | 0.551239489 | 0.948955122 |
| SH3D21 | 0.551156062 | 0.86472256 |
| GHRH | 0.551008539 | 0.917375456 |
| PCBD2 | 0.550950833 | 0.669046074 |
| TMEM87B | 0.55064126 | 0.880518429 |
| AKR7A3 | 0.550470889 | 0.863724854 |
| TPI1 | 0.550448146 | 0.869582827 |
| OR1A1 | 0.550274383 | 0.980532122 |
| WDR46 | 0.550100225 | 0.922640745 |
| ONECUT2 | 0.549983062 | 0.772782503 |
| RNF34 | 0.549592829 | 0.923426347 |
| IARS2 | 0.549440804 | 0.705953827 |
| TNFRSF10C | 0.549195126 | 0.953277093 |

| | | |
|---|---|---|
| DBF4 | 0.548821543 | 0.866320571 |
| ZDHHC12 | 0.548425129 | 0.962315862 |
| FEN1 | 0.548358529 | 0.974483597 |
| SDHC | 0.547874801 | 0.847306602 |
| EEF1B2 | 0.547743834 | 0.753341434 |
| C2orf48 | 0.547612001 | 1.009720535 |
| OR13F1 | 0.547400634 | 0.859198356 |
| TERF1 | 0.547304034 | 0.865022268 |
| OR5P2 | 0.547202812 | 1.032545392 |
| TMEM135 | 0.547160365 | 0.912515188 |
| DTYMK | 0.546902589 | 0.927968851 |
| TIGD1 | 0.546411707 | 0.874957177 |
| PRM2 | 0.546285927 | 0.869475848 |
| SGTA | 0.546226021 | 1.008505468 |
| TBC1D28 | 0.545868522 | 0.910895099 |
| DAND5 | 0.54476363 | 0.64763062 |
| SNTG1 | 0.544291897 | 0.861253854 |
| MX2 | 0.543600105 | 0.808424463 |
| DIP2A | 0.543518754 | 0.876873228 |
| HUWE1 | 0.543029198 | 0.737832576 |
| DNASE2 | 0.542661685 | 0.707168894 |
| SGK223 | 0.542340729 | 1.04412719 |
| WDR75 | 0.54213965 | 0.787768327 |
| CIRH1A | 0.541521668 | 0.905789654 |
| DLGAP1-AS1 | 0.540001895 | 0.802349129 |
| MSL3 | 0.539575938 | 0.840013703 |
| TMEM169 | 0.539396267 | 0.81855002 |
| AGPAT2 | 0.538764456 | 0.765897124 |
| RASGRP1 | 0.538449697 | 0.87363305 |
| MAFG | 0.538317758 | 0.8137573 |
| HIST1H2BK | 0.53788802 | 0.831106543 |
| TAF1C | 0.537283523 | 1.013770757 |
| LTV1 | 0.537210322 | 0.979343864 |
| PRPF8 | 0.537196369 | 0.771497088 |
| DGKK | 0.537018862 | 0.905256327 |
| EP300 | 0.536890602 | 0.776119403 |
| TCTN2 | 0.536592316 | 0.857432159 |
| LILRA5 | 0.536323097 | 0.980963953 |
| TGM5 | 0.536204932 | 0.846852693 |
| DNAJC10 | 0.535763515 | 0.927319922 |
| SPPL2A | 0.53571124 | 0.78780013 |
| PDSS1 | 0.535580339 | 0.859052248 |
| IFT46 | 0.535489767 | 0.901579587 |
| C14orf178 | 0.535296143 | 0.948157149 |

| | | |
|---|---|---|
| ERCC8 | 0.534793211 | 0.9181855 |
| SYCE1 | 0.534719093 | 0.945321993 |
| AWAT2 | 0.5346111 | 0.949372215 |
| SCAND3 | 0.533934667 | 0.9331963 |
| WASL | 0.53352602 | 0.77389517 |
| PAGR1 | 0.532691521 | 0.8327258 |
| ABCD1 | 0.532599421 | 0.871607938 |
| NOL11 | 0.532567278 | 0.863912515 |
| NIP7 | 0.532138565 | 0.852976914 |
| INHA | 0.532104083 | 0.873458793 |
| NPAS4 | 0.531709014 | 0.874107722 |
| USP7 | 0.531329174 | 0.81440623 |
| CLIP2 | 0.531117604 | 0.88513952 |
| HYKK | 0.530313977 | 0.8663926 |
| CHD7 | 0.530262348 | 0.805970149 |
| RIPK2 | 0.529896586 | 0.846852693 |
| ADRA2C | 0.529676618 | 0.837118754 |
| OR51A7 | 0.529370903 | 0.272696129 |
| GCNT2 | 0.528887105 | 0.889428919 |
| WASF1 | 0.528774986 | 0.905224787 |
| CTRL | 0.52862675 | 0.899415964 |
| ZNF474 | 0.528181583 | 0.878898339 |
| CMC2 | 0.528007776 | 0.820575132 |
| CCDC116 | 0.527647527 | 0.836776023 |
| MEF2BNB-MEF2B | 0.527357161 | 0.962362103 |
| TREM2 | 0.5270582 | 0.78290806 |
| TRERF1 | 0.526967373 | 0.845686513 |
| RPAP2 | 0.526819036 | 0.959497772 |
| RAB20 | 0.526770608 | 0.872160934 |
| CDAN1 | 0.526654244 | 0.908869988 |
| TOB1 | 0.526576654 | 1.024659312 |
| USP27X | 0.526412482 | 0.897529364 |
| RRNAD1 | 0.526286813 | 0.940866748 |
| ARF5 | 0.525797583 | 0.729850142 |
| RHOV | 0.525782963 | 0.780882949 |
| HSD3B1 | 0.524838176 | 0.735115431 |
| ABCC5 | 0.523791661 | 0.716889429 |
| SRPK3 | 0.523409598 | 0.939000649 |
| TIMM23B | 0.523374206 | 0.871202916 |
| NTRK2 | 0.523240835 | 0.918883842 |
| WT1 | 0.522687129 | 0.899415964 |
| RPS6KB1 | 0.522517872 | 1.001946788 |
| BNIP3 | 0.522315933 | 0.685269306 |
| CD5L | 0.522123574 | 0.778452815 |

| | | |
|---|---|---|
| DCAF13 | 0.521967646 | 0.853786958 |
| FCF1 | 0.521967646 | 0.853786958 |
| HRASLS5 | 0.521917113 | 0.774807614 |
| CCDC147 | 0.521799579 | 0.892428914 |
| SIGLEC7 | 0.521725634 | 0.886605002 |
| ZNF420 | 0.521085311 | 0.882948562 |
| NOL10 | 0.520674122 | 0.76873228 |
| GTF2A1L | 0.520640247 | 0.899554475 |
| PANK1 | 0.520531742 | 0.944192083 |
| tcag7.873_pseudogene | 0.520380845 | 0.936622131 |
| RS1 | 0.520301553 | 0.214799589 |
| ASF1A | 0.520228854 | 0.781287971 |
| MAP2 | 0.519488948 | 0.818144998 |
| LRRN4 | 0.519473068 | 0.806783145 |
| IMPAD1 | 0.519362158 | 0.5799919 |
| CAMK2A | 0.519340772 | 1.038935756 |
| ALKBH4 | 0.518919029 | 0.910490077 |
| TFEC | 0.518893763 | 0.652085865 |
| SMO | 0.518843869 | 0.926670993 |
| CEL | 0.518835131 | 0.787363305 |
| GDF15 | 0.518703261 | 0.957819598 |
| H2AFJ | 0.518654751 | 0.847306602 |
| GNA12 | 0.517939246 | 0.724989874 |
| SLC10A1 | 0.517441713 | 1.000810045 |
| EWSR1 | 0.517291335 | 1.084360805 |
| EIF2A | 0.517159199 | 0.718104496 |
| CD46 | 0.516104431 | 0.92345079 |
| RPP14 | 0.516104431 | 0.92345079 |
| MAPK14 | 0.515776094 | 0.797534069 |
| OR10A5 | 0.51531265 | 0.900650908 |
| ANXA2R | 0.515200549 | 0.925070879 |
| EEF1D | 0.51509495 | 0.761036857 |
| CENPJ | 0.514893991 | 0.601053058 |
| CALD1 | 0.514881756 | 0.105515588 |
| PHB | 0.51447404 | 0.854191981 |
| UBC | 0.514454486 | 0.908187736 |
| MED17 | 0.513717018 | 1.004050223 |
| ZNF800 | 0.513281225 | 0.691373026 |
| TES | 0.513212761 | 0.812069664 |
| PLCG1 | 0.513102017 | 0.836371 |
| PSG9 | 0.512939314 | 0.933171324 |
| ZNF580 | 0.512806259 | 0.883864337 |
| MYBPHL | 0.512751396 | 0.80072904 |
| EIF5A2 | 0.512680538 | 0.69218307 |

| | | |
|---|---|---|
| TAS2R14 | 0.512259874 | 0.938436614 |
| AATF | 0.511394197 | 0.931956258 |
| ZDHHC24 | 0.511304339 | 0.867557716 |
| KNSTRN | 0.511295475 | 0.762656946 |
| ANKRD49 | 0.511147188 | 0.82746051 |
| PCGF1 | 0.510871111 | 0.731992213 |
| AICDA | 0.510600345 | 0.860672337 |
| SERPINA1 | 0.510404009 | 0.9733939 |
| TJP1 | 0.510307643 | 0.68327258 |
| IFITM5 | 0.509629273 | 0.906844876 |
| CTHRC1 | 0.509560823 | 0.112709832 |
| NEUROG3 | 0.509454346 | 0.974991435 |
| CNIH4 | 0.508749536 | 0.948967193 |
| PIWIL1 | 0.508284656 | 0.738760632 |
| MAP1LC3A | 0.507944579 | 0.99108951 |
| SRMS | 0.507819801 | 0.9467878 |
| APC | 0.507799303 | 0.89422453 |
| PDCD4 | 0.507526332 | 0.211031175 |
| MAN2A2 | 0.507468543 | 0.853786958 |
| DCD | 0.506876366 | 0.903391572 |
| SDF4 | 0.506867449 | 0.895504253 |
| PACSIN1 | 0.506514896 | 0.917585983 |
| TMPRSS6 | 0.506171855 | 0.759896171 |
| CLEC2D | 0.505896895 | 0.92926671 |
| GPR128 | 0.505548256 | 0.855937703 |
| SLC11A2 | 0.505432757 | 0.771576898 |
| IQCJ | 0.505343315 | 0.906132237 |
| SP1 | 0.504915767 | 0.9733939 |
| EMG1 | 0.504718843 | 0.793033617 |
| DMXL2 | 0.504711966 | 0.923855812 |
| DDX24 | 0.503478501 | 0.835965978 |
| OGFR | 0.503369572 | 0.838416613 |
| CAPN1 | 0.50325893 | 0.859831278 |
| LIPK | 0.50298917 | 1.029566626 |
| CCL3L1 | 0.502685183 | 0.914011648 |
| NDUFAF3 | 0.502649755 | 0.825030377 |
| PDE9A | 0.502269734 | 0.872160934 |
| TRPC1 | 0.501910887 | 1.085658663 |
| TMEM31 | 0.501595849 | 0.841726619 |
| PLA2G10 | 0.501534573 | 0.839206156 |
| MTHFD2 | 0.501517661 | 0.942081814 |
| SRSF2 | 0.50151002 | 0.810449575 |
| ZNF148 | 0.501323579 | 0.86836776 |
| ERAP2 | 0.501097945 | 0.746268657 |

| | | |
|---|---|---|
| UBE2U | 0.501097945 | 0.895522388 |
| SEZ6L2 | 0.500974129 | 0.881328473 |
| NAPA | 0.500710726 | 0.821514217 |
| TMOD1 | 0.500597325 | 0.820170109 |
| ABHD6 | 0.500238655 | 0.865022713 |
| ATP2A1 | 0.499993327 | 0.767517213 |
| NEO1 | 0.499489267 | 0.866320571 |
| ACACA | 0.499413526 | 0.929526124 |
| MND1 | 0.499141658 | 0.744025921 |
| TMEM52 | 0.499067591 | 0.81855002 |
| ATAD3A | 0.498857292 | 0.790339157 |
| GLI3 | 0.498838105 | 0.703118672 |
| LOC157708_pseudogene | 0.49854902 | 0.608343459 |
| MARCH4 | 0.498080716 | 0.954574951 |
| C17orf47 | 0.496544779 | 0.930741191 |
| CCND2 | 0.495923024 | 0.861778066 |
| RPS6KA4 | 0.495437235 | 0.927319922 |
| RDH10 | 0.495386606 | 0.959766385 |
| TBC1D26 | 0.495030353 | 0.758821514 |
| SULT1C2 | 0.494542464 | 0.885788449 |
| C20orf196 | 0.493587803 | 0.861077359 |
| UGT2A1 | 0.493214014 | 0.736534718 |
| TEKT1 | 0.493187294 | 0.975676602 |
| ATP5J | 0.492969491 | 0.778452815 |
| CHST6 | 0.492756709 | 0.813108371 |
| SCYL3 | 0.492440587 | 0.998053212 |
| B3GALT1 | 0.492311141 | 0.808829486 |
| ZC3HAV1L | 0.491528037 | 0.92345079 |
| NCCRP1 | 0.491293907 | 0.848150552 |
| CTGF | 0.491283781 | 0.804672291 |
| CILP2 | 0.491206857 | 0.916409729 |
| AKR1C3 | 0.491101311 | 0.890643985 |
| WFDC8 | 0.490762203 | 0.874443094 |
| FASTKD1 | 0.490452486 | 0.740380721 |
| C19orf40 | 0.490441381 | 0.816119887 |
| PPT1 | 0.48958523 | 0.707978939 |
| BCL2 | 0.489333873 | 0.927968851 |
| IFI30 | 0.488511464 | 0.885788449 |
| HBA1 | 0.487387122 | 0.891743748 |
| RECQL4 | 0.486375336 | 0.878088295 |
| LEP | 0.486347598 | 0.900713822 |
| NPHP1 | 0.485500235 | 1.10490077 |
| CMTM5 | 0.485375686 | 0.850141758 |
| PSD | 0.485349045 | 0.960712839 |

| | | |
|---|---|---|
| CECR1 | 0.485324897 | 0.880597015 |
| NUDT22 | 0.484898937 | 0.799918996 |
| RAB38 | 0.484836739 | 0.957472661 |
| PCYOX1 | 0.484416075 | 0.809234508 |
| CLIP1 | 0.484119909 | 0.767112191 |
| PDHA1 | 0.483864495 | 0.767517213 |
| C17orf53 | 0.483738622 | 0.908464966 |
| UBE2D3 | 0.483495839 | 0.828682674 |
| SYTL4 | 0.482834208 | 0.88513952 |
| PRPF19 | 0.482411642 | 0.852693056 |
| CLEC4G | 0.480219216 | 0.558525719 |
| WDR62 | 0.478832268 | 0.762656946 |
| FKBP11 | 0.478677359 | 0.926690968 |
| SLC2A9 | 0.478632531 | 0.905224787 |
| NR3C1 | 0.478484346 | 0.840826245 |
| CD52 | 0.478366318 | 0.884163629 |
| KIF21A | 0.477467275 | 0.6654516 |
| NDUFA9 | 0.476338288 | 0.736330498 |
| RNASE6 | 0.47629478 | 0.987021415 |
| SUCNR1 | 0.47629478 | 0.987021415 |
| DPH6 | 0.476046197 | 0.637100041 |
| ATF7 | 0.475672296 | 0.715674362 |
| RIOK2 | 0.474385457 | 0.923426347 |
| GPX3 | 0.474139149 | 0.865937627 |
| SLC16A8 | 0.473088967 | 0.806804374 |
| ACADVL | 0.471816379 | 0.77399757 |
| SDR9C7 | 0.471612399 | 0.894873459 |
| ACAP3 | 0.470512463 | 0.85945727 |
| HEBP1 | 0.469616065 | 0.836245289 |
| MS4A3 | 0.469369716 | 0.906132237 |
| LGALS2 | 0.468410931 | 0.912394549 |
| P2RY14 | 0.467450548 | 1.028552888 |
| MECR | 0.466497613 | 0.605913325 |
| ETNPPL | 0.465598605 | 0.791693705 |
| HECTD2 | 0.465451494 | 0.952628164 |
| LGALS8 | 0.465248978 | 0.826735886 |
| BAMBI | 0.465022844 | 0.971648441 |
| NYX | 0.464462551 | 0.792052073 |
| TXLNB | 0.464278099 | 0.791008505 |
| PSMA8 | 0.464061383 | 0.8403634 |
| PI4KA | 0.46349607 | 1.014276444 |
| EPGN | 0.463188808 | 0.861253854 |
| TNK2 | 0.462861522 | 0.969500324 |
| OR4K17 | 0.462816589 | 1.069886948 |

| | | |
|---|---|---|
| ANO1 | 0.460977971 | 0.649880096 |
| RASAL2 | 0.460827971 | 0.828270555 |
| GTF3A | 0.460765417 | 0.644795464 |
| ITGB3 | 0.460739065 | 0.788449059 |
| MED8 | 0.460671738 | 0.774807614 |
| BMS1 | 0.459704027 | 0.830295666 |
| SMOC2 | 0.45935992 | 0.148681055 |
| TNR | 0.459108537 | 0.988659376 |
| C9orf139 | 0.458150963 | 0.79708384 |
| ICAM1 | 0.458078378 | 0.816353018 |
| PPP1R12B | 0.45773081 | 0.781959766 |
| CSRNP1 | 0.456279276 | 0.877697842 |
| UTP15 | 0.45621707 | 0.845686513 |
| TOP1MT | 0.456055794 | 0.986229243 |
| TUBB8 | 0.456035876 | 0.85054678 |
| FGF18 | 0.455347572 | 0.702308627 |
| NOC2L | 0.454147167 | 0.817739976 |
| TXNDC17 | 0.453234115 | 0.74200081 |
| OR4S2 | 0.453095959 | 0.137033231 |
| CCNE1 | 0.452950328 | 0.924665857 |
| POLR2J3 | 0.451906718 | 0.913669065 |
| LRRC37A | 0.451653085 | 0.9181855 |
| ITGB6 | 0.451181256 | 0.76963011 |
| RPL28 | 0.450867316 | 0.905447071 |
| PVR | 0.450017617 | 0.890330954 |
| IL1RAPL2 | 0.449588137 | 0.879299156 |
| DCDC5 | 0.449472132 | 1.00526529 |
| PRKAG3 | 0.447722857 | 1.109669046 |
| RPS4Y2 | 0.446822076 | 1.014388489 |
| GRAMD4 | 0.446157761 | 0.897124342 |
| SSTR5 | 0.446120847 | 0.766385464 |
| UCN | 0.445982272 | 1.008505468 |
| TP73 | 0.445667082 | 1.066839714 |
| C11orf35 | 0.445108342 | 0.709194006 |
| TK1 | 0.444264168 | 1.159986297 |
| GSTK1 | 0.444260239 | 0.826650466 |
| DENND6A | 0.443479173 | 0.68853787 |
| SAMD1 | 0.442557207 | 0.89509923 |
| TMEM55A | 0.442025137 | 0.961522884 |
| GLDC | 0.441233628 | 0.832320778 |
| DLL4 | 0.438676381 | 0.921830701 |
| HSD17B2 | 0.43848084 | 0.865022713 |
| PGLYRP4 | 0.437648717 | 0.70798183 |
| PRDM9 | 0.43753802 | 0.844066424 |

| | | |
|---|---|---|
| RPS4Y1 | 0.437314582 | 0.816375471 |
| DHRS4L1 | 0.437002792 | 0.948967193 |
| RRP36 | 0.436688135 | 1.015795869 |
| IGF2R | 0.436412304 | 0.807916937 |
| CTSZ | 0.436342247 | 0.918234912 |
| KRT35 | 0.43630374 | 0.903199676 |
| YBX1 | 0.435714719 | 0.965978129 |
| SCML1 | 0.435386155 | 0.663426488 |
| NPTXR | 0.434299193 | 0.859862292 |
| TOM1 | 0.432426625 | 0.988319273 |
| NIFK | 0.430535858 | 0.901336074 |
| BTNL8 | 0.429090713 | 0.899149453 |
| TEP1 | 0.427823117 | 0.776427704 |
| HOXA1 | 0.425809814 | 0.828033744 |
| C12orf29 | 0.425791776 | 0.799513973 |
| H1FX | 0.423811144 | 0.853374443 |
| KIF3C | 0.423107965 | 1.022712524 |
| TMEM60 | 0.422862461 | 0.839206156 |
| CHEK1 | 0.422170386 | 0.759247242 |
| RAB11FIP1 | 0.421484587 | 0.766015759 |
| ZIC1 | 0.421480097 | 0.910490077 |
| PHF12 | 0.420845229 | 0.723556132 |
| ATXN1 | 0.420788686 | 0.833535844 |
| CT55 | 0.420606983 | 0.750506278 |
| TAF11 | 0.42023624 | 0.765897124 |
| CXorf57 | 0.417769462 | 0.893884164 |
| OR10K2 | 0.417379855 | 0.747516273 |
| INHBC | 0.416337992 | 0.926690968 |
| KLHL1 | 0.416315477 | 0.882138518 |
| LALBA | 0.41602685 | 0.892669097 |
| CMBL | 0.415744947 | 0.903199676 |
| OR51I2 | 0.415639151 | 0.944844125 |
| BAI1 | 0.41536596 | 0.926022064 |
| ARL17P1 | 0.414192053 | 0.925659472 |
| HEXA | 0.413727308 | 0.877683273 |
| ANXA9 | 0.413467504 | 0.865671642 |
| GLTSCR2 | 0.412988449 | 0.78177458 |
| APOM | 0.412709727 | 0.699878493 |
| MSX1 | 0.410962021 | 0.863507493 |
| KLF15 | 0.410807876 | 0.929121102 |
| ARRDC2 | 0.41077947 | 0.751969853 |
| ATOX1 | 0.409569265 | 0.886593763 |
| CYP11B2 | 0.408564572 | 0.949372215 |
| PHF23 | 0.406946269 | 0.800778715 |

| | | |
|---|---|---|
| NFE2L1 | 0.406920914 | 0.853341986 |
| RHOA | 0.406529919 | 0.832320778 |
| LINC00176 | 0.405602452 | 0.946132037 |
| FAM89B | 0.404654149 | 0.892264075 |
| ELOVL6 | 0.40412036 | 0.819597664 |
| GNPDA1 | 0.402171193 | 0.876703439 |
| RUNX2 | 0.401771292 | 0.559740786 |
| CASKIN2 | 0.401525619 | 0.815690305 |
| FAM133A | 0.401522755 | 0.852976914 |
| ACSL1 | 0.401520666 | 0.935196436 |
| AKR1B10 | 0.401284915 | 0.807209397 |
| ENO1 | 0.401260617 | 0.724989874 |
| NDUFS3 | 0.4000948 | 0.788983394 |
| SLC1A6 | 0.398757518 | 0.941676792 |
| RPS17 | 0.395456354 | 0.832134293 |
| PLP1 | 0.394607014 | 0.920210612 |
| PCBP2 | 0.393773236 | 0.868267359 |
| VNN2 | 0.39346896 | 0.880923451 |
| ZBTB18 | 0.392901843 | 0.903199676 |
| C7orf49 | 0.392361979 | 0.925475901 |
| MDM2 | 0.38850669 | 0.797534069 |
| EPX | 0.385956863 | 0.673552045 |
| POLB | 0.385949171 | 0.807209397 |
| FCRL5 | 0.383956442 | 0.703928716 |
| TMEM241 | 0.380700448 | 0.840016201 |
| NCOR1 | 0.379948061 | 0.760226812 |
| RNF141 | 0.3794322 | 0.788449059 |
| TBL1X | 0.378799278 | 0.954779034 |
| GNL3L | 0.378510428 | 0.844876468 |
| LZTR1 | 0.378327603 | 0.692588092 |
| ISL2 | 0.375416831 | 0.956257594 |
| TRIB3 | 0.374737224 | 0.898118105 |
| UBB | 0.37402652 | 0.739165654 |
| SLC36A2 | 0.373850726 | 0.855407047 |
| SNPH | 0.373027016 | 0.823912299 |
| GJB1 | 0.372122835 | 0.659781288 |
| ZIK1 | 0.369501046 | 0.80963953 |
| UTS2 | 0.36937171 | 0.252141144 |
| PES1 | 0.36673084 | 0.846522782 |
| ECM1 | 0.365524911 | 0.795868773 |
| PDHA2 | 0.363541758 | 0.930741191 |
| CYP26C1 | 0.363051067 | 0.756766016 |
| NUSAP1 | 0.362958454 | 0.902363823 |
| KRT32 | 0.359040124 | 0.948157149 |

| | | |
|---|---|---|
| PWP2 | 0.3552407 | 0.92345079 |
| TIAF1 | 0.354293084 | 0.9331963 |
| GJA4 | 0.353015786 | 0.584447145 |
| SMPD1 | 0.349050484 | 0.811161583 |
| GLB1L3 | 0.344313624 | 0.904819765 |
| HOXD11 | 0.344037329 | 0.869565217 |
| POU2F3 | 0.34352462 | 0.870863076 |
| PGLYRP3 | 0.342285798 | 0.819765087 |
| TMEM165 | 0.33808848 | 0.90927501 |
| EXOSC9 | 0.337594258 | 0.959497772 |
| NDST1 | 0.333831257 | 0.834345889 |
| ZNF470 | 0.333699016 | 0.908869988 |
| FANCD2OS | 0.333275092 | 0.89145403 |
| ZNF768 | 0.330374507 | 0.924260834 |
| C1QB | 0.328987086 | 0.787941076 |
| TMC8 | 0.327589557 | 0.62980964 |
| PITX2 | 0.324766197 | 0.686107736 |
| MYLPF | 0.324067349 | 0.770352369 |
| NPFFR1 | 0.323541068 | 0.74302401 |
| ZCCHC9 | 0.321580246 | 0.885378696 |
| CBFA2T3 | 0.316839702 | 0.755366545 |
| CDK6 | 0.315898838 | 0.845554835 |
| MDM4 | 0.312973564 | 0.95587281 |
| MPST | 0.309323479 | 0.840421223 |
| MTIF3 | 0.307750403 | 0.972096042 |
| PLCD1 | 0.30771112 | 0.737545565 |
| CD19 | 0.302838692 | 0.912920211 |
| UGT1A5 | 0.302373456 | 0.846091535 |
| ADNP2 | 0.301040912 | 0.609963548 |
| BMS1P20 | 0.299245006 | 0.954436451 |
| NSMF | 0.29908461 | 0.903048989 |
| GMEB1 | 0.299070982 | 0.678780013 |
| ALDOB | 0.29843145 | 0.698258404 |
| FBLIM1 | 0.298000038 | 0.810044552 |
| RMI2 | 0.292513628 | 0.57837181 |
| RSL24D1 | 0.291084476 | 0.810551559 |
| ARCN1 | 0.287679958 | 0.669064748 |
| APOA2 | 0.282680493 | 0.71526934 |
| SMG8 | 0.275960057 | 1.009315512 |
| PLCD3 | 0.273000942 | 0.687727825 |
| SNX18 | 0.27292371 | 0.727989037 |
| SEMG1 | 0.266895252 | 0.89671932 |
| PKDCC | 0.261620417 | 0.918883842 |
| GRHPR | 0.256193281 | 0.829485622 |

| | | |
|---|---|---|
| DOCK8 | 0.253083998 | 0.8218568 |
| CCL17 | 0.25253992 | 0.94655704 |
| ARMC8 | 0.25049007 | 0.95544755 |
| MRPS2 | 0.247727769 | 0.820829051 |
| C9orf41 | 0.245538908 | 0.809181226 |
| METTL5 | 0.242572285 | 0.714459295 |
| RPL36 | 0.236816176 | 0.838985954 |
| RPS25 | 0.234882544 | 0.819458719 |
| PIGF | 0.232932419 | 0.871512005 |
| ASCC3 | 0.232088688 | 0.782608696 |
| MTSS1L | 0.230119908 | 0.788983394 |
| RPS2 | 0.227726792 | 0.93835172 |
| FAU | 0.224969257 | 0.786570743 |
| SCGB3A1 | 0.221022429 | 0.056183625 |
| RPSA | 0.22073362 | 0.962737951 |
| RPL4 | 0.21733332 | 0.811161583 |
| NKIRAS1 | 0.201965476 | 0.82746051 |
| UBA52 | 0.201046566 | 0.820980154 |
| RPL7 | 0.18696674 | 0.780404248 |
| RPS24 | 0.185622631 | 0.878040425 |
| APOBEC3A | 0.179151 | 0.875253139 |
| SPANXA1 | 0.17805611 | 0.819458719 |
| DNTTIP2 | 0.177021192 | 0.885783718 |
| RPL6 | 0.173255467 | 0.707776636 |
| ADTRP | 0.170714324 | 0.77286742 |
| SYNGR4 | 0.169337936 | 0.743215877 |
| VCP | 0.168571439 | 0.783313082 |
| FILIP1L | 0.159610756 | 0.762656946 |
| RNF17 | 0.157405798 | 1.018170019 |
| SPANXA2 | 0.150645707 | 0.803699897 |
| RPL27 | 0.147463252 | 0.76841384 |
| GTF3C5 | 0.146702719 | 0.660996355 |
| RPS10 | 0.144284376 | 0.720794793 |
| LOC653314_pseudogene | 0.142315175 | 0.872559096 |
| RPS9 | 0.140767671 | 0.705721137 |
| C17orf78 | 0.137980589 | 0.732685298 |
| ADAM2 | 0.136611444 | 0.899415964 |
| RPS14 | 0.135799474 | 0.720109627 |
| PUSL1 | 0.129949849 | 0.822193381 |
| CARD14 | 0.129598586 | 0.780072904 |
| RPL10A | 0.128581293 | 0.507022953 |
| RPS28 | 0.105801758 | 0.58684481 |
| NUMA1 | 0.105646922 | 0.898339409 |
| C17orf59 | 0.105482182 | 0.801944107 |

| | | |
|---|---|---|
| RPL35A | 0.10416001 | 0.774922919 |
| PLAA | 0.103347228 | 0.618874038 |
| KDM3A | 0.102202092 | 0.045563549 |
| RPL18 | 0.097027509 | 0.687906817 |
| MAML2 | 0.096634484 | 0.7472661 |
| RPS16 | 0.096578265 | 0.755395683 |
| RPS12 | 0.095238233 | 0.651935594 |
| RPL8 | 0.087250877 | 0.76498801 |
| HINT2 | 0.085688255 | 0.698258404 |
| GALK2 | 0.084966626 | 1.131732641 |
| RPL17 | 0.084755043 | 0.677629325 |
| RPS6 | 0.079890967 | 0.680027407 |
| RPLP0 | 0.076579606 | 0.729701953 |
| RPS3A | 0.076098735 | 0.754710517 |
| RPL12 | 0.073079983 | 0.594724221 |
| RPL38 | 0.069642118 | 0.735525865 |
| RPS7 | 0.068581647 | 0.678999657 |
| RPL9 | 0.065429005 | 0.664268585 |
| RPS19 | 0.064550678 | 0.625214114 |
| FGR | 0.061144836 | 0.961064244 |
| RPS27A | 0.060920474 | 0.611510791 |
| RPL24 | 0.060579927 | 0.743062693 |
| RPL37A | 0.058413765 | 0.797190819 |
| RPS3 | 0.057965382 | 0.642685851 |
| RPS11 | 0.053995899 | 0.661185337 |
| ARHGAP36 | 0.051762301 | 0.677602268 |
| RPS29 | 0.051566334 | 0.560145808 |
| RPL19 | 0.0492214 | 0.599177801 |
| RPL3 | 0.048863273 | 0.698869476 |
| RPS5 | 0.046324304 | 0.703665639 |
| MRPL22 | 0.046234274 | 0.033573141 |
| RPS18 | 0.046151176 | 0.571771155 |
| RPS4X | 0.043847999 | 0.743405276 |
| RPL13 | 0.043590187 | 0.640972936 |
| DRAM2 | 0.043169661 | 0.621304172 |
| RPS13 | 0.041473323 | 0.748544022 |
| CCBE1 | 0.041102865 | 0.752936412 |
| COL13A1 | 0.03974526 | 0.039054471 |
| RPS21 | 0.03786319 | 0.614936622 |
| RPL23 | 0.037343082 | 0.623501199 |
| XPO5 | 0.037139007 | 0.041795135 |
| RPL26 | 0.03687753 | 0.839206156 |
| RPL14 | 0.036021265 | 0.603288798 |
| HES4 | 0.034835649 | 0.668379582 |

| | | |
|---|---|---|
| PARK2 | 0.034349037 | 0.933160286 |
| RPL30 | 0.033687413 | 0.829393628 |
| KPRP | 0.032430096 | 0.827055488 |
| GP5 | 0.025032917 | 0.062007537 |
| HTR2B | 0.0227832 | 0.703439325 |
| PPIL4 | 0.020987401 | 1.018170019 |
| RPL21 | 0.018810415 | 0.660157588 |
| RPL7A | 0.014868725 | 0.693803159 |
| TSSK6 | 0.010719143 | 0.996755354 |
| PLSCR3 | 0.00562704 | 0.949383517 |
| ABLIM3 | 0 | 0.036656389 |
| COL4A3 | 0 | 0.031517643 |
| KCNT1 | 0 | 1.020116807 |
| LINC00452 | 0 | 0.040424803 |
| MYEF2 | 0 | 1.001712915 |
| NAB1 | 0 | 0.922918808 |
| SLC35B1 | 0 | 0.032887975 |
| SOCS1 | 0 | 0.791044776 |
| TUBGCP4 | 0 | 0.039054471 |
