## Supplemental Table 2 for "Functional analysis across model systems implicates ribosomal proteins in growth and proliferation defects associated with hypoplastic left heart syndrome"

| Proband | Gene | MOI | Gene Alias |
|---|---|---|---|
| 323H | *ACAP2* | De Novo | |
| 87H | *ACTR6* | Cmpd Het | |
| 314H | *AIFM1* | Xlinked | |
| 145H | *AJUBA* | Cmpd Het | |
| 68H | *AKAP9* | Cmpd Het | |
| 87H | *AMMECR1* | Xlinked | |
| 205H | *AMOTL2* | Cmpd Het | |
| 49H | *ANKRD50* | Cmpd Het | |
| 87H | *APOO* | Xlinked | |
| 219H | *ARHGAP26* | Cmpd Het | |
| 48H | *ARHGAP31* | Cmpd Het | |
| 151H | *ARHGEF6* | Xlinked | |
| 49H | *ARHGEF9* | Xlinked | |
| 49H | *ARID1B* | De Novo | |
| 314H | *ARSD* | Xlinked | |
| 72H | *ATM* | Cmpd Het | |
| 249H | *ATP11C* | Xlinked | |
| 87H | *ATP13A1* | Cmpd Het | |
| 314H | *B4GALT2* | Cmpd Het | |
| 197H | *BAG6* | Cmpd Het | |
| 68H | *BCAP31* | Xlinked | |
| 219H | *BECN1* | De Novo | |
| 56H | *BPHL* | Hom Rec | |
| 151H | *C1GALT1C1* | Xlinked | |
| 314H | *C7orf55-LUC7L2* | Hom Rec | |
| 108H | *CAMSAP1* | Cmpd Het | |
| 74H | *CAND2* | Cmpd Het | |
| 201H | *CAPN6* | Xlinked | |
| 197H | *CCDC8* | Hom Rec | |
| 197H | *CDKN1A* | Cmpd Het | |
| 87H | *CELSR1* | Cmpd Het | |
| 201H | *CENPI* | Xlinked | |
| 325H | *CIPC* | De Novo | KIAA1737 |
| 323H | *CLDND1* | Cmpd Het | |
| 96H | *CLIC1* | Cmpd Het | |
| 201H | *CLTA* | De Novo | |
| 250H | *CMTM3* | De Novo | |
| 146H | *COL4A5* | Xlinked | |
| 68H | *COL4A5* | Xlinked | |
| 201H | *COL4A6* | Xlinked | |
| 48H | *COL6A3* | Cmpd Het | |
| 314H | *COL6A3* | Cmpd Het | |
| 219H | *COL6A6* | Cmpd Het | |
| 96H | *COX18* | Cmpd Het | |
| 15H | *CTBP2* | Cmpd Het | |
| 96H | *CTC1* | Hom Rec | |
| 219H | *CTDSP1* | Cmpd Het | |
| 151H | *CTPS2* | Xlinked | |
| 207H | *CTSL* | Cmpd Het | |
| 108H | *CTSZ* | Cmpd Het | |
| 48H | *CUL7* | Cmpd Het | |
| 68H | *CUL7* | Cmpd Het | |
| 250H | *CYHR1* | Cmpd Het | |
| 323H | *CYTH1* | Cmpd Het | |

| | | | |
|---|---|---|---|
| 48H | *DAP3* | Cmpd Het | |
| 15H | *DAP3* | Cmpd Het | |
| 96H | *DCTN1* | De Novo | |
| 15H | *DDAH1* | Cmpd Het | |
| 76H | *DDX3X* | Xlinked | |
| 205H | *DIDO1* | Cmpd Het | |
| 267H | *DKC1* | Xlinked | |
| 68H | *DMD* | Xlinked | |
| 207H | *DMD* | Xlinked | |
| 68H | *DNAJB1* | Cmpd Het | |
| 207H | *DNAJB4* | Cmpd Het | |
| 68H | *DNHD1* | De Novo | |
| 96H | *DOCK6* | Cmpd Het | |
| 219H | *DOCK9* | Cmpd Het | |
| 15H | *DPP9* | Cmpd Het | |
| 249H | *DPYSL3* | Cmpd Het | |
| 267H | *DST* | Cmpd Het | |
| 197H | *DTNA* | Cmpd Het | |
| 151H | *ELF1* | Cmpd Het | |
| 56H | *ELP1* | Cmpd Het | IKBKAP |
| 323H | *EPHA4* | Cmpd Het | |
| 108H | *EZR* | Cmpd Het | |
| 219H | *FAM129A* | Cmpd Het | |
| 207H | *FAM50A* | Xlinked | |
| 68H | *FAM98B* | De Novo | |
| 323H | *FAT4* | Cmpd Het | |
| 151H | *FAT4* | Cmpd Het | |
| 205H | *FBXO11* | Cmpd Het | |
| 49H | *FGF13* | Xlinked | |
| 197H | *FGGY* | Cmpd Het | |
| 250H | *FHL1* | Xlinked | |
| 49H | *FLNA* | Xlinked | |
| 48H | *FLNA* | Xlinked | |
| 314H | *FLNA* | Xlinked | |
| 250H | *GBF1* | Cmpd Het | |
| 146H | *GLYR1* | Cmpd Het | |
| 96H | *GNL3L* | Xlinked | |
| 197H | *GOLGA3* | Cmpd Het | |
| 323H | *GOT2* | Cmpd Het | |
| 249H | *GPC3* | Xlinked | |
| 76H | *GPC4* | Xlinked | |
| 250H | *GPRASP1* | Xlinked | |
| 76H | *GPSM1* | Cmpd Het | |
| 250H | *GRIPAP1* | Xlinked | |
| 250H | *GUCY1A1* | Cmpd Het | GUCY1A3 |
| 87H | *GYS1* | Cmpd Het | |
| 146H | *HDLBP* | Cmpd Het | |
| 219H | *HEY2* | Cmpd Het | |
| 74H | *HIST1H4H* | Cmpd Het | |
| 249H | *HMCN1* | Cmpd Het | |
| 146H | *HMGA1* | Cmpd Het | |
| 151H | *HPS4* | Cmpd Het | |
| 146H | *HTATSF1* | Xlinked | |
| 249H | *HTATSF1* | Xlinked | |
| 250H | *IDS* | Xlinked | |
| 49H | *IFIT1* | Cmpd Het | |

| | | | |
|---|---|---|---|
| 72H | *JMJD1C* | Hom Rec | |
| 49H | *JMJD1C* | Cmpd Het | |
| 49H | *JUP* | De Novo | |
| 323H | *KALRN* | De Novo | |
| 76H | *KDM6A* | Xlinked | |
| 56H | *KIAA1671* | Cmpd Het | |
| 249H | *KIFC1* | Cmpd Het | |
| 314H | *KLHL21* | Cmpd Het | |
| 314H | *KMT2C* | Cmpd Het | |
| 325H | *KMT2D* | De Novo | |
| 250H | *LATS2* | Cmpd Het | |
| 207H | *LDLRAD4* | Cmpd Het | |
| 76H | *LGALS3BP* | Hom Rec | |
| 74H | *LIMCH1* | Cmpd Het | |
| 207H | *LIMS1* | Cmpd Het | |
| 72H | *LINS1* | Cmpd Het | LINS |
| 197H | *LRBA* | De Novo | |
| 87H | *LRBA* | Cmpd Het | |
| 56H | *LRRFIP2* | De Novo | |
| 314H | *LUC7L2* | Hom Rec | |
| 205H | *LUC7L2* | Cmpd Het | |
| 201H | *LYST* | Cmpd Het | |
| 207H | *MAGT1* | Xlinked | |
| 151H | *MAP3K15* | Xlinked | |
| 15H | *MAP7D3* | Xlinked | |
| 145H | *MARCH7* | Cmpd Het | |
| 151H | *MDC1* | Cmpd Het | |
| 145H | *MDM2* | Cmpd Het | |
| 48H | *MED13* | Cmpd Het | |
| 151H | *MED14* | Xlinked | |
| 197H | *MEF2C* | Cmpd Het | |
| 48H | *MEIS2* | Cmpd Het | |
| 49H | *MESDC2* | De Novo | MESD |
| 96H | *MID1* | Xlinked | |
| 250H | *MIER1* | Cmpd Het | |
| 145H | *MLEC* | Cmpd Het | |
| 323H | *MLLT10* | Cmpd Het | |
| 201H | *MMRN1* | Cmpd Het | |
| 267H | *MPDZ* | Cmpd Het | |
| 314H | *MRPL47* | Cmpd Het | |
| 49H | *MRPL50* | De Novo | |
| 249H | *MSL3* | Xlinked | |
| 249H | *MSN* | Xlinked | |
| 207H | *MTCP1* | Xlinked | |
| 87H | *MXRA5* | Xlinked | |
| 145H | *MXRA5* | Xlinked | |
| 151H | *MXRA5* | Xlinked | |
| 151H | *MYADM* | Cmpd Het | |
| 219H | *MYBPC3* | Cmpd Het | |
| 323H | *MYH6* | Cmpd Het | |
| 219H | *MYOF* | Cmpd Het | |
| 48H | *MZT2B* | Cmpd Het | |
| 250H | *NAA10* | Xlinked | |
| 314H | *NBAS* | Cmpd Het | |
| 250H | *NEBL* | Cmpd Het | |
| 145H | *NEDD9* | Cmpd Het | |

| | | |
|---|---|---|
| 146H | *NFAT5* | De Novo |
| 68H | *NHSL2* | Xlinked |
| 250H | *NNT* | Cmpd Het |
| 146H | *NOL6* | Hom Rec |
| 74H | *NOP9* | Cmpd Het |
| 108H | *NPEPL1* | Cmpd Het |
| 323H | *NPNT* | Cmpd Het |
| 146H | *NR2F2* | Cmpd Het |
| 205H | *NSUN5* | Cmpd Het |
| 72H | *NUBP2* | Cmpd Het |
| 249H | *NUMB* | Cmpd Het |
| 250H | *NUP153* | Cmpd Het |
| 145H | *NUP214* | Cmpd Het |
| 325H | *OBSCN* | Cmpd Het |
| 74H | *OSBP* | Cmpd Het |
| 48H | *OTUD5* | Xlinked |
| 151H | *OTUD5* | Xlinked |
| 49H | *PCDH17* | Cmpd Het |
| 314H | *PDE4D* | Cmpd Het |
| 96H | *PDE4DIP* | Cmpd Het |
| 151H | *PDHA1* | Xlinked |
| 314H | *PGD* | Cmpd Het |
| 87H | *PGK1* | Xlinked |
| 72H | *PGRMC1* | Xlinked |
| 323H | *PHF11* | Cmpd Het |
| 145H | *PHF6* | Xlinked |
| 250H | *PIEZO1* | Cmpd Het |
| 145H | *PKD1* | Cmpd Het |
| 146H | *PKMYT1* | Cmpd Het |
| 201H | *PLEC* | Cmpd Het |
| 49H | *PLXNA3* | Xlinked |
| 15H | *PLXNA3* | Xlinked |
| 145H | *PLXNA3* | Xlinked |
| 76H | *PPAN* | Cmpd Het |
| 249H | *PPID* | Hom Rec |
| 49H | *PPP1CB* | Cmpd Het |
| 323H | *PPP3CC* | Cmpd Het |
| 87H | *PRIMPOL* | Cmpd Het |
| 49H | *PRPF19* | Cmpd Het |
| 201H | *PRRC2A* | De Novo |
| 68H | *PRRC2C* | Cmpd Het |
| 267H | *PSPH* | Cmpd Het |
| 201H | *PTGER3* | Cmpd Het |
| 68H | *PTK7* | Cmpd Het |
| 201H | *PTPRD* | Cmpd Het |
| 49H | *RAB11FIP5* | Cmpd Het |
| 207H | *RAB5B* | Cmpd Het |
| 197H | *RABL6* | Cmpd Het |
| 48H | *RAD23A* | Cmpd Het |
| 201H | *RAD50* | Cmpd Het |
| 314H | *RAD51AP1* | Cmpd Het |
| 68H | *RAD51C* | Cmpd Het |
| 249H | *RALBP1* | Cmpd Het |
| 267H | *RAN* | Cmpd Het |
| 197H | *RASL11B* | Cmpd Het |
| 76H | *RBFOX1* | De Novo |

| | | | |
|---|---|---|---|
| 314H | *RBM24* | Cmpd Het | |
| 267H | *RDX* | Cmpd Het | |
| 201H | *RENBP* | Xlinked | |
| 56H | *RERE* | Cmpd Het | |
| 56H | *RGS3* | Cmpd Het | |
| 201H | *RGS3* | Cmpd Het | |
| 219H | *RGS5* | Cmpd Het | |
| 96H | *RNF103* | De Novo | |
| 96H | *RNH1* | Cmpd Het | |
| 76H | *RPL10* | Xlinked | |
| 201H | *RPL13A* | Cmpd Het | |
| 145H | *RPL26L1* | Cmpd Het | |
| 145H | *RPL36A* | Xlinked | |
| 145H | *RPL36A-HNRNPH2* | Xlinked | |
| 151H | *RPL39* | Xlinked | |
| 96H | *RPL3L* | Cmpd Het | |
| 145H | *RPS15* | Cmpd Het | |
| 325H | *RPS17* | Hom Rec | |
| 76H | *RPS28* | Cmpd Het | |
| 146H | *RTCA* | Cmpd Het | |
| 250H | *RTL8C* | Xlinked | FAM127A |
| 87H | *RYK* | Cmpd Het | |
| 151H | *SBSPON* | Cmpd Het | |
| 151H | *SCAF11* | Cmpd Het | |
| 249H | *SCD* | Cmpd Het | |
| 219H | *SDF4* | Cmpd Het | |
| 108H | *SEC16A* | Cmpd Het | |
| 68H | *SEMA6C* | Cmpd Het | |
| 49H | *SENP7* | De Novo | |
| 146H | *SEPT2* | Cmpd Het | |
| 323H | *SF1* | Cmpd Het | |
| 314H | *SGCA* | Cmpd Het | |
| 197H | *SLC2A14* | Cmpd Het | |
| 49H | *SLC35A2* | Xlinked | |
| 250H | *SLC6A8* | Xlinked | |
| 207H | *SMS* | Xlinked | |
| 72H | *SPSB3* | Cmpd Het | |
| 207H | *SRPK3* | Xlinked | |
| 314H | *SRPK3* | Xlinked | |
| 68H | *SRRM2* | Cmpd Het | |
| 201H | *STARD13* | Cmpd Het | |
| 15H | *STT3B* | Cmpd Het | |
| 201H | *SULF2* | Cmpd Het | |
| 74H | *SUN1* | Cmpd Het | |
| 219H | *SYNCRIP* | Cmpd Het | |
| 72H | *SYNE1* | Cmpd Het | |
| 15H | *SYNE2* | Cmpd Het | |
| 197H | *SYNGR2* | Cmpd Het | |
| 314H | *SZT2* | Cmpd Het | |
| 74H | *TAF1* | Xlinked | |
| 267H | *TAZ* | Xlinked | |
| 145H | *TBC1D1* | Hom Rec | |
| 207H | *TBCD* | Cmpd Het | |
| 325H | *TFRC* | Hom Rec | |
| 48H | *TIA1* | Cmpd Het | |

| | | | |
|---|---|---|---|
| 201H | *TIMM17B* | Xlinked | |
| 72H | *TIMM8A* | Xlinked | |
| 87H | *TIMP1* | Xlinked | |
| 267H | *TIPARP* | De Novo | |
| 151H | *TKT* | De Novo | |
| 15H | *TMEM167A* | De Novo | |
| 207H | *TMEM185A* | Xlinked | |
| 151H | *TMEM185A* | Xlinked | |
| 201H | *TMSB15A* | Xlinked | |
| 145H | *TP53BP1* | Cmpd Het | |
| 96H | *TRAPPC8* | Cmpd Het | |
| 87H | *TRO* | Xlinked | |
| 56H | *TSEN54* | Cmpd Het | |
| 145H | *TSPAN6* | Xlinked | |
| 72H | *TTI1* | Cmpd Het | |
| 249H | *TTN* | De Novo | |
| 74H | *TTN* | Cmpd Het | |
| 72H | *TTN* | Cmpd Het | |
| 207H | *TTN* | Cmpd Het | |
| 314H | *TTN* | Cmpd Het | |
| 108H | *TTN* | Cmpd Het | |
| 267H | *TUBB6* | Cmpd Het | |
| 146H | *UBN1* | Cmpd Het | |
| 201H | *UBN2* | Cmpd Het | |
| 250H | *UBQLN2* | Xlinked | |
| 151H | *UNC13B* | Cmpd Het | |
| 207H | *USP11* | Xlinked | |
| 76H | *USP2* | Cmpd Het | |
| 72H | *VMA21* | Xlinked | |
| 76H | *VPS13B* | Cmpd Het | |
| 323H | *VPS26A* | Cmpd Het | |
| 76H | *WAPL* | Cmpd Het | WAPAL |
| 49H | *WDR43* | Cmpd Het | |
| 68H | *WDR45* | Xlinked | |
| 48H | *YY1AP1* | Cmpd Het | |
| 146H | *ZCCHC24* | Cmpd Het | |
| 151H | *ZDHHC9* | Xlinked | |
| 207H | *ZNF185* | Xlinked | |
| 49H | *ZNF189* | De Novo | |
| 151H | *ZNF445* | Cmpd Het | |
