## Supplemental Table 3 for "Functional analysis across model systems implicates ribosomal proteins in growth and proliferation defects associated with hypoplastic left heart syndrome"

| Gene Symbol | Consequence | Variant | MAF% | CADD | TFs |
|---|---|---|---|---|---|
| **RPS15A** | Regulatory | c.-95G>A | 0.662 | | MAX; SRF; GTF2B; BRCA1; CHD2; NR2C2; ETS1; YY1; RXRA; HEY1; POLR2A; SIN3A; MYC; E2F1; E2F4; ZNF143; EGR1; ZNF263; TBP; SETDB1; STAT1; PAX5; ZBTB33; GTF2F1; POU2F2; NFKB1; FOSL2; REST; ELF1; Hltf; SP1; IRF1; BCL3; TAF7; TAF1 |
| **WDR33** | Missense | p.Y280C | 0.518 | 21 | N/A |
| **NADK2** | Regulatory | c.300+371C>G | 0.708 | | POLR2A |
| **RSL1D1** | Missense | p.A7V | 0.191 | <10 | N/A |
| **GDE1** | Missense | p.V201L | 0.045 | 16 | N/A |
| **ACSF2** | Missense | p.A260D | 0.11 | 20 | N/A |
