## Supplemental Table 4 for "Functional analysis across model systems implicates ribosomal proteins in growth and proliferation defects associated with hypoplastic left heart syndrome"

| Gene | relative hPSC-CM proliferation rate | Fly heart defect(s) |
|---|---|---|
| AIFM1 | 0.99 | dilated |
| ARID1B | 1.04 | lethal_hits |
| CAMSAP1 | 1.02 | lethal_hits |
| CAND2 | 0.77 | |
| CMTM3 | 0.77 | |
| COL6A3 | 0.98 | no.heart, constricted |
| CTBP2 | 1.02 | constricted |
| CTDSP1 | 0.86 | |
| CTSL | 0.75 | |
| CYTH1 | 0.42 | |
| DKC1 | 1.02 | constricted |
| DNAJB1 | 0.76 | lethal_hits |
| DNAJB4 | 1.07 | lethal_hits |
| DOCK6 | 1.06 | constricted |
| DOCK9 | 0.96 | constricted |
| FAM98B | 0.85 | |
| GBF1 | 0.59 | |
| GNL3L | 0.52 | |
| GPSM1 | 0.85 | |
| HEY2 | 0.86 | |
| HMCN1 | 1.15 | dilated |
| HPS4 | 0.78 | |
| IFIT1 | 1 | dilated |
| JUP | 1.01 | constricted |
| KALRN | | constricted |
| KDM6A | 1.11 | dilated |
| KIFC1 | 0.83 | dilated |
| MAP7D3 | | dilated |
| MED14 | 1.02 | no.heart, constricted |
| MEF2C | 1.2 | dilated |
| MIER1 | 0.57 | |
| MRPL47 | 0.85 | |
| MRPL50 | 0.94 | constricted |
| MSL3 | 0.73 | |
| MYBPC3 | 1.06 | dilated |
| MYH6 | 1.07 | dilated |
| NEBL | 0.89 | |
| NOL6 | 1.02 | ostia.bulging.out, constricted |
| OBSCN | 0.77 | dilated |
| PCDH17 | 0.97 | constricted |
| PGD | 0.32 | constricted |
| PPP1CB | 0.98 | constricted |
| PRPF19 | 0.97 | constricted, dilated |
| PRRC2C | 0.84 | |
| PTK7 | 0.85 | |
| RABL6 | 0.87 | |
| RAN | 0.98 | constricted |
| RASL11B | 0.87 | |
| RBM24 | 0.73 | |
| RERE | 1.23 | constricted |
| RPL10 | 0.79 | constricted |
| RPL26L1 | 0.53 | lethal_hits, no.heart |
| RPL36A | 1.06 | lethal_hits, no.heart |
| 36A-HNRNPH2 | 0.99 | lethal_hits, no.heart |
| RPL39 | 0.26 | lethal_hits |
| RPL3L | 1.3 | no.heart |
| RPS15 | 0.22 | lethal_hits |
| RPS28 | 0.07 | ostia.bulging.out |
| SCAF11 | 0.76 | |
| SCD | 0.77 | |
| SEC16A | 1 | constricted, dilated |
| SF1 | 0.5 | |
| SRPK3 | 0.84 | |
| STARD13 | 0.71 | dilated |
| SYNCRIP | 1.04 | dilated |
| TAF1 | 0.85 | |
| TIMM8A | 0.85 | |
| TMEM167A | 1.03 | constricted, dilated |
| TTN | 1.02 | dilated, dilated |
| TTN | 1.08 | dilated, dilated |
| USP11 | 1.11 | lethal_hits |
| USP2 | 1.14 | lethal_hits |
| WDR45 | 0.86 | |
| YY1AP1 | 0.98 | no.heart, constricted |
