## Supplemental Table 5 for "Functional analysis across model systems implicates ribosomal proteins in growth and proliferation defects associated with hypoplastic left heart syndrome"

| ENSG | baseMean | log2FoldChange | lfcSE | pvalue | padj | gene |
|---|---|---|---|---|---|---|
| ENSG00000000003 | 436.233393 | 0.09268074 | 0.18943058 | 0.45745352 | 0.726537795 | TSPAN6 |
| ENSG00000000005 | 11.96205387 | 0.018232672 | 0.23956997 | 0.63057256 | NA | TNMD |
| ENSG00000000419 | 351.7405756 | -0.166689773 | 0.18823467 | 0.20706823 | 0.505936081 | DPM1 |
| ENSG00000000457 | 281.5344808 | 0.203559714 | 0.2248445 | 0.14706995 | 0.426552403 | SCYL3 |
| ENSG00000000460 | 83.38523874 | -1.08739924 | 0.48943672 | 0.00110204 | 0.018633614 | C1orf112 |
| ENSG00000000938 | 1.537745985 | 0.01398106 | 0.24175946 | 0.46543284 | NA | FGR |
| ENSG00000000971 | 109.5009018 | 0.720660007 | 0.56233275 | 0.00836659 | 0.076033569 | CFH |
| ENSG00000001036 | 1031.022948 | -0.219983079 | 0.14179717 | 0.06097325 | 0.263577734 | FUCA2 |
| ENSG00000001084 | 416.2099784 | 0.000252191 | 0.16150139 | 0.99972718 | 0.999727184 | GCLC |
| ENSG00000001167 | 519.6341893 | -0.023923049 | 0.17025259 | 0.84157895 | 0.937972108 | NFYA |
| ENSG00000001460 | 141.4108736 | 0.058413706 | 0.19740495 | 0.62431969 | 0.826637958 | STPG1 |
| ENSG00000001461 | 653.6454341 | 0.794621974 | 0.18880917 | 2.20E-06 | 0.000101429 | NIPAL3 |
| ENSG00000001497 | 671.1297061 | -0.430373954 | 0.19427167 | 0.00484194 | 0.052877381 | LAS1L |
| ENSG00000001561 | 109.8440289 | 0.131165593 | 0.25436305 | 0.22645956 | 0.524413205 | ENPP4 |
| ENSG00000001617 | 311.1756207 | -0.105768702 | 0.22483629 | 0.36223788 | 0.654133479 | SEMA3F |
| ENSG00000001626 | 17.54156138 | 0.033492632 | 0.23885832 | 0.52853003 | NA | CFTR |
| ENSG00000001629 | 620.9469868 | 0.098150865 | 0.14874828 | 0.40829687 | 0.691421151 | ANKIB1 |
| ENSG00000001630 | 1109.825518 | -0.194285397 | 0.18214267 | 0.1435645 | 0.422111699 | CYP51A1 |
| ENSG00000001631 | 290.9032828 | 0.069280903 | 0.20017562 | 0.56323479 | 0.791662607 | KRIT1 |
| ENSG00000002016 | 220.4658588 | 0.124710389 | 0.19548956 | 0.3325832 | 0.627313557 | RAD52 |
| ENSG00000002079 | 0.281046574 | 0.005473671 | 0.24149293 | 0.78105731 | NA | MYH16 |
| ENSG00000002330 | 789.9075204 | -0.386831347 | 0.16420446 | 0.00426408 | 0.04819944 | BAD |
| ENSG00000002549 | 1355.715115 | 0.125728919 | 0.15206709 | 0.29602337 | 0.594088738 | LAP3 |
| ENSG00000002586 | 5427.859497 | -0.128314487 | 0.13094328 | 0.244437 | 0.5434308 | CD99 |
| ENSG00000002587 | 14.0636511 | 0.024616498 | 0.24041588 | 0.51789756 | NA | HS3ST1 |
| ENSG00000002726 | 4.427006818 | -0.044111576 | 0.24730444 | 0.01103398 | NA | AOC1 |
| ENSG00000002745 | 0.426141397 | -0.005955552 | 0.24153919 | 0.73324866 | NA | WNT16 |
| ENSG00000002746 | 6.778424773 | -0.031455165 | 0.24178519 | 0.38881271 | NA | HECW1 |
| ENSG00000002822 | 579.2106434 | -0.336513153 | 0.1756107 | 0.01440023 | 0.108372214 | MAD1L1 |
| ENSG00000002834 | 3509.829377 | -0.413264198 | 0.14407637 | 0.00085264 | 0.015214534 | LASP1 |
| ENSG00000002919 | 400.5560554 | -0.110691655 | 0.17204351 | 0.37640371 | 0.665876422 | SNX11 |
| ENSG00000002933 | 324.1652802 | -0.445337787 | 0.25768942 | 0.01194159 | 0.096801968 | TMEM176A |
| ENSG00000003056 | 2017.297461 | 0.029720973 | 0.13858161 | 0.79873657 | 0.919105508 | M6PR |
| ENSG00000003096 | 1376.217893 | 0.022008104 | 0.12694099 | 0.84106491 | 0.937759244 | KLHL13 |
| ENSG00000003137 | 64.15857327 | -0.105995494 | 0.25468946 | 0.22606904 | 0.524022131 | CYP26B1 |
| ENSG00000003147 | 727.89575 | 0.195158081 | 0.2086429 | 0.15859141 | 0.443361007 | ICA1 |
| ENSG00000003249 | 350.6961354 | 0.247517791 | 0.23671054 | 0.09470232 | 0.336466256 | DBNDD1 |
| ENSG00000003393 | 1578.209098 | 0.232674511 | 0.12998116 | 0.03647192 | 0.198309958 | ALS2 |
| ENSG00000003400 | 24.21657902 | 0.045188954 | 0.23660532 | 0.51201703 | 0.761290603 | CASP10 |
| ENSG00000003402 | 1530.027151 | 0.665660381 | 0.14785379 | 7.57E-07 | 4.02E-05 | CFLAR |
| ENSG00000003436 | 93.27344418 | 0.177819133 | 0.28661867 | 0.13180816 | 0.404056658 | TFPI |
| ENSG00000003509 | 329.8625828 | 0.073076588 | 0.16758797 | 0.55262867 | 0.785766299 | NDUFAF7 |
| ENSG00000003756 | 3207.397135 | 0.28121186 | 0.11786374 | 0.00704881 | 0.067576211 | RBM5 |
| ENSG00000003987 | 16.06949436 | -0.015167823 | 0.23452232 | 0.79249174 | NA | MTMR7 |
| ENSG00000003989 | 541.3129517 | 0.154742975 | 0.19135101 | 0.24826282 | 0.547401439 | SLC7A2 |
| ENSG00000004059 | 1739.898844 | -0.045425239 | 0.12800567 | 0.67551239 | 0.859908366 | ARF5 |
| ENSG00000004139 | 199.5666842 | -0.05822593 | 0.20574631 | 0.61407227 | 0.820924662 | SARM1 |
| ENSG00000004142 | 2932.948681 | -0.008012457 | 0.10885343 | 0.92889055 | 0.973660183 | POLDIP2 |
| ENSG00000004399 | 1126.29213 | -0.216964567 | 0.20434755 | 0.12042328 | 0.384325002 | PLXND1 |
| ENSG00000004455 | 2132.599504 | 0.004408114 | 0.12059698 | 0.96785545 | 0.987047963 | AK2 |
| ENSG00000004468 | 1.917479523 | 0.003236534 | 0.2408063 | 0.86990336 | NA | CD38 |
| ENSG00000004478 | 3612.052308 | -0.320643139 | 0.13177524 | 0.00503948 | 0.054323714 | FKBP4 |
| ENSG00000004487 | 2321.849971 | -0.232597851 | 0.12285343 | 0.0293418 | 0.172143391 | KDM1A |
| ENSG00000004534 | 1419.609979 | -0.003231735 | 0.13429337 | 0.97551247 | 0.990312003 | RBM6 |
| ENSG00000004660 | 175.5099327 | -0.617308918 | 0.3946209 | 0.00814613 | 0.074796793 | CAMKK1 |
| ENSG00000004700 | 178.3814168 | -0.09613021 | 0.21432328 | 0.41686141 | 0.697193579 | RECQL |
| ENSG00000004766 | 325.6194011 | 0.230706852 | 0.21638821 | 0.10782369 | 0.361500578 | VPS50 |
| ENSG00000004776 | 487.6706965 | -0.107949499 | 0.18457232 | 0.39207049 | 0.677786006 | HSPB6 |
| ENSG00000004777 | 442.7739649 | -1.383031933 | 0.20601278 | 1.15E-12 | 2.09E-10 | ARHGAP33 |
| ENSG00000004779 | 1707.548609 | -0.069623563 | 0.12827336 | 0.52100827 | 0.766000406 | NDUFAB1 |
| ENSG00000004799 | 88.45048004 | 0.849455322 | 0.49624603 | 0.00387509 | 0.045053521 | PDK4 |
