## Supplemental Table 6 for "Functional analysis across model systems implicates ribosomal proteins in growth and proliferation defects associated with hypoplastic left heart syndrome"

| | baseMean | log2FoldChange | lfcSE | pvalue | padj | ENSG | gene |
|---|---|---|---|---|---|---|---|
| ENSG00000000003 | 2923.838943 | -0.002537397 | 0.08838725 | 0.97381323 | 0.98907078 | ENSG00000000003 | TSPAN6 |
| ENSG00000000005 | 21.97529732 | -0.144260105 | 0.28196543 | 0.10680297 | NA | ENSG00000000005 | TNMD |
| ENSG00000000419 | 1551.595726 | -0.170069826 | 0.11509838 | 0.09397821 | 0.24059987 | ENSG00000000419 | DPM1 |
| ENSG00000000457 | 273.3008654 | 0.12741839 | 0.15511522 | 0.29263928 | 0.51551178 | ENSG00000000457 | SCYL3 |
| ENSG00000000460 | 1172.058855 | 0.002608323 | 0.10021421 | 0.97963428 | 0.99191824 | ENSG00000000460 | C1orf112 |
| ENSG00000000938 | 8.944371215 | 0.01314923 | 0.23324599 | 0.8080063 | NA | ENSG00000000938 | FGR |
| ENSG00000000971 | 0 | NA | NA | NA | NA | ENSG00000000971 | CFH |
| ENSG00000001036 | 2274.615867 | -0.154706996 | 0.08065658 | 0.04088466 | 0.13013453 | ENSG00000001036 | FUCA2 |
| ENSG00000001084 | 1808.83403 | -0.261072984 | 0.09232867 | 0.00223105 | 0.01341339 | ENSG00000001084 | GCLC |
| ENSG00000001167 | 2017.948124 | -0.038123427 | 0.08073934 | 0.61375663 | 0.79007086 | ENSG00000001167 | NFYA |
| ENSG00000001460 | 267.5774669 | -0.008718871 | 0.15295153 | 0.9396064 | 0.97485747 | ENSG00000001460 | STPG1 |
| ENSG00000001461 | 473.9583056 | 0.258137934 | 0.14000226 | 0.02805993 | 0.09872391 | ENSG00000001461 | NIPAL3 |
| ENSG00000001497 | 3902.752227 | -0.166979311 | 0.06834179 | 0.01050721 | 0.0460909 | ENSG00000001497 | LAS1L |
| ENSG00000001561 | 75.42690795 | 0.060530809 | 0.20701531 | 0.58488746 | 0.76967843 | ENSG00000001561 | ENPP4 |
| ENSG00000001617 | 1771.237269 | -0.05651774 | 0.09100832 | 0.50049242 | 0.70720854 | ENSG00000001617 | SEMA3F |
| ENSG00000001626 | 61.93122916 | 0.208493345 | 0.28195121 | 0.12067689 | 0.28503666 | ENSG00000001626 | CFTR |
| ENSG00000001629 | 2086.76753 | 0.090742068 | 0.07672777 | 0.20911752 | 0.41418089 | ENSG00000001629 | ANKIB1 |
| ENSG00000001630 | 2786.544179 | -0.594451961 | 0.09830264 | 2.01E-10 | 6.55E-09 | ENSG00000001630 | CYP51A1 |
| ENSG00000001631 | 1033.403651 | 0.443477874 | 0.12750521 | 0.00010359 | 0.001009 | ENSG00000001631 | KRIT1 |
| ENSG00000002016 | 584.8033736 | 0.105381112 | 0.12946282 | 0.33509196 | 0.56040801 | ENSG00000002016 | RAD52 |
| ENSG00000002079 | 0.490351291 | 0.007644257 | 0.23929939 | 0.53338695 | NA | ENSG00000002079 | MYH16 |
| ENSG00000002330 | 572.7702846 | 0.027345091 | 0.11566045 | 0.78923406 | 0.89691366 | ENSG00000002330 | BAD |
| ENSG00000002549 | 2661.345656 | -0.068118972 | 0.08545224 | 0.39254407 | 0.61674026 | ENSG00000002549 | LAP3 |
| ENSG00000002586 | 939.7273394 | 0.04397094 | 0.10129451 | 0.62155939 | 0.79597947 | ENSG00000002586 | CD99 |
| ENSG00000002587 | 78.49177266 | 1.887847382 | 0.36773607 | 1.54E-08 | 3.58E-07 | ENSG00000002587 | HS3ST1 |
| ENSG00000002726 | 6.936825543 | -0.014742473 | 0.23417701 | 0.76901059 | NA | ENSG00000002726 | AOC1 |
| ENSG00000002745 | 3.702536636 | -0.004666659 | 0.23637822 | 0.89758369 | NA | ENSG00000002745 | WNT16 |
| ENSG00000002746 | 385.6919765 | 0.445938632 | 0.17804159 | 0.0022004 | 0.01326026 | ENSG00000002746 | HECW1 |
| ENSG00000002822 | 1020.22236 | -0.114810472 | 0.11088037 | 0.24234327 | 0.4572328 | ENSG00000002822 | MAD1L1 |
| ENSG00000002834 | 15647.24734 | -0.094343635 | 0.05160756 | 0.06530493 | 0.18419924 | ENSG00000002834 | LASP1 |
| ENSG00000002919 | 716.4469346 | -0.098870219 | 0.11162463 | 0.31872621 | 0.54435772 | ENSG00000002919 | SNX11 |
| ENSG00000002933 | 5.77917767 | -0.022803287 | 0.23763766 | 0.55516778 | NA | ENSG00000002933 | TMEM176A |
| ENSG00000003056 | 4447.872613 | -0.006479666 | 0.06721047 | 0.91899289 | 0.96433041 | ENSG00000003056 | M6PR |
| ENSG00000003096 | 254.0304233 | -0.359281765 | 0.20841797 | 0.01854559 | 0.0720665 | ENSG00000003096 | KLHL13 |
| ENSG00000003137 | 4.040896169 | -0.039256943 | 0.24085977 | 0.30517045 | NA | ENSG00000003137 | CYP26B1 |
| ENSG00000003147 | 657.5689868 | -0.184695759 | 0.11566463 | 0.06859053 | 0.19066579 | ENSG00000003147 | ICA1 |
| ENSG00000003249 | 2943.411796 | -0.029552002 | 0.07088961 | 0.6620469 | 0.8227249 | ENSG00000003249 | DBNDD1 |
| ENSG00000003393 | 2212.685474 | -0.08212158 | 0.08026579 | 0.28290128 | 0.50503429 | ENSG00000003393 | ALS2 |
| ENSG00000003400 | 289.9504135 | 1.603071254 | 0.19830413 | 3.78E-17 | 2.44E-15 | ENSG00000003400 | CASP10 |
| ENSG00000003402 | 404.4600004 | 0.53389132 | 0.1755232 | 0.00033093 | 0.0026911 | ENSG00000003402 | CFLAR |
| ENSG00000003436 | 76.85577442 | 0.006957266 | 0.19969119 | 0.95120444 | 0.98056851 | ENSG00000003436 | TFPI |
| ENSG00000003509 | 1532.920777 | 0.285262515 | 0.09417842 | 0.00103778 | 0.00707875 | ENSG00000003509 | NDUFAF7 |
| ENSG00000003756 | 3694.953878 | 0.048020395 | 0.06697429 | 0.45544098 | 0.66864464 | ENSG00000003756 | RBM5 |
| ENSG00000003987 | 182.9471554 | -0.013936424 | 0.16600694 | 0.90590901 | 0.95699527 | ENSG00000003987 | MTMR7 |
| ENSG00000003989 | 1147.046134 | -0.344765203 | 0.11334797 | 0.00071982 | 0.00522552 | ENSG00000003989 | SLC7A2 |
| ENSG00000004059 | 3837.880708 | -0.196481589 | 0.06513132 | 0.00166059 | 0.01045426 | ENSG00000004059 | ARF5 |
| ENSG00000004139 | 386.485291 | -0.004387056 | 0.14072068 | 0.96798148 | 0.98738413 | ENSG00000004139 | SARM1 |
| ENSG00000004142 | 4822.483128 | -0.079517169 | 0.08105056 | 0.29499137 | 0.51828461 | ENSG00000004142 | POLDIP2 |
| ENSG00000004399 | 1757.908474 | 0.131004436 | 0.08465691 | 0.09721961 | 0.24609067 | ENSG00000004399 | PLXND1 |
| ENSG00000004455 | 4367.782977 | -0.241781301 | 0.06839005 | 0.00019886 | 0.00176295 | ENSG00000004455 | AK2 |
| ENSG00000004468 | 10.05626945 | 0.046129931 | 0.23869012 | 0.41253503 | NA | ENSG00000004468 | CD38 |
| ENSG00000004478 | 16194.2816 | -0.455919285 | 0.06045853 | 9.01E-15 | 4.55E-13 | ENSG00000004478 | FKBP4 |
| ENSG00000004487 | 13185.99056 | -0.498892137 | 0.05462562 | 1.08E-20 | 9.04E-19 | ENSG00000004487 | KDM1A |
| ENSG00000004534 | 3640.259212 | -0.246991009 | 0.07591608 | 0.00058008 | 0.00436152 | ENSG00000004534 | RBM6 |
| ENSG00000004660 | 315.556503 | -0.507115772 | 0.17855468 | 0.00068583 | 0.00501671 | ENSG00000004660 | CAMKK1 |
| ENSG00000004700 | 1574.919169 | 0.128056214 | 0.11080081 | 0.1923405 | 0.39236951 | ENSG00000004700 | RECQL |
| ENSG00000004766 | 369.3893506 | 0.506598236 | 0.21434154 | 0.00243161 | 0.01440495 | ENSG00000004766 | VPS50 |
| ENSG00000004776 | 22.25104063 | -0.15182639 | 0.29076529 | 0.08671688 | NA | ENSG00000004776 | HSPB6 |
| ENSG00000004777 | 658.491433 | -0.101210922 | 0.11939178 | 0.32692372 | 0.55246152 | ENSG00000004777 | ARHGAP33 |
| ENSG00000004779 | 3114.024751 | -0.118278671 | 0.07306629 | 0.08826306 | 0.22933167 | ENSG00000004779 | NDUFAB1 |
| ENSG00000004799 | 4.13358193 | 0.044882438 | 0.24303502 | 0.19835666 | NA | ENSG00000004799 | PDK4 |
| ENSG00000004809 | 5.270047132 | -0.053741908 | 0.2444349 | 0.2045809 | NA | ENSG00000004809 | SLC22A16 |
| ENSG00000004838 | 6.401083155 | 0.027255248 | 0.23711702 | 0.54463129 | NA | ENSG00000004838 | ZMYND10 |
| ENSG00000004846 | 0 | NA | NA | NA | NA | ENSG00000004846 | ABCB5 |
| ENSG00000004848 | 1.449516205 | 0.017869244 | 0.23925429 | 0.41410314 | NA | ENSG00000004848 | ARX |
| ENSG00000004864 | 4491.862377 | 0.001639805 | 0.06494097 | 0.98918176 | 0.99578988 | ENSG00000004864 | SLC25A13 |
| ENSG00000004866 | 731.8491828 | -0.138701687 | 0.11122652 | 0.15846423 | 0.34502201 | ENSG00000004866 | ST7 |
| ENSG00000004897 | 1658.559281 | 0.06548376 | 0.10848832 | 0.49779812 | 0.7051528 | ENSG00000004897 | CDC27 |
