## Supplemental Table 7 for "Functional analysis across model systems implicates ribosomal proteins in growth and proliferation defects associated with hypoplastic left heart syndrome"

| | baseMean | log2FoldChange | lfcSE | pvalue | padj | ENSG | gene |
|---|---|---|---|---|---|---|---|
| ENSG00000000003 | 436.233393 | -0.133017342 | 0.19431072 | 0.23983516 | 0.59910224 | ENSG00000000003 | TSPAN6 |
| ENSG00000000005 | 11.96205387 | 0.006321223 | 0.20774455 | 0.82372234 | NA | ENSG00000000005 | TNMD |
| ENSG00000000419 | 351.7405756 | -0.150794155 | 0.18145599 | 0.19780171 | 0.54933731 | ENSG00000000419 | DPM1 |
| ENSG00000000457 | 281.5344808 | 0.415012062 | 0.29674211 | 0.01616249 | 0.14754252 | ENSG00000000457 | SCYL3 |
| ENSG00000000460 | 83.38523874 | -0.576897996 | 0.60337868 | 0.01128617 | 0.1189959 | ENSG00000000460 | C1orf112 |
| ENSG00000000938 | 1.537745985 | 0.015004932 | 0.21024807 | 0.17640287 | NA | ENSG00000000938 | FGR |
| ENSG00000000971 | 109.5009018 | 0.32435281 | 0.49312637 | 0.02643244 | 0.19902896 | ENSG00000000971 | CFH |
| ENSG00000001036 | 1031.022948 | -0.050348697 | 0.12309751 | 0.6143764 | 0.86422068 | ENSG00000001036 | FUCA2 |
| ENSG00000001084 | 416.2099784 | -0.06479482 | 0.15647532 | 0.54468923 | 0.83058814 | ENSG00000001084 | GCLC |
| ENSG00000001167 | 519.6341893 | -0.084061171 | 0.1678764 | 0.43684566 | 0.76601886 | ENSG00000001167 | NFYA |
| ENSG00000001460 | 141.4108736 | 0.026739695 | 0.17636389 | 0.78506234 | 0.93385564 | ENSG00000001460 | STPG1 |
| ENSG00000001461 | 653.6454341 | 0.704146634 | 0.19079888 | 1.79E-05 | 0.00074148 | ENSG00000001461 | NIPAL3 |
| ENSG00000001497 | 671.1297061 | -0.181688127 | 0.16718134 | 0.12287513 | 0.44606719 | ENSG00000001497 | LAS1L |
| ENSG00000001561 | 109.8440289 | 0.111928875 | 0.22773461 | 0.19196351 | 0.5432496 | ENSG00000001561 | ENPP4 |
| ENSG00000001617 | 311.1756207 | -0.109315151 | 0.21275375 | 0.26087001 | 0.62365197 | ENSG00000001617 | SEMA3F |
| ENSG00000001626 | 17.54156138 | -0.003433876 | 0.20567133 | 0.92912965 | NA | ENSG00000001626 | CFTR |
| ENSG00000001629 | 620.9469868 | 0.050125486 | 0.13784307 | 0.63278566 | 0.87304973 | ENSG00000001629 | ANKIB1 |
| ENSG00000001630 | 1109.825518 | -0.083057029 | 0.15495967 | 0.44168307 | 0.76948073 | ENSG00000001630 | CYP51A1 |
| ENSG00000001631 | 290.9032828 | -0.051708255 | 0.18110551 | 0.59558822 | 0.85558022 | ENSG00000001631 | KRIT1 |
| ENSG00000002016 | 220.4658588 | 0.088663872 | 0.17659306 | 0.411515 | 0.74785444 | ENSG00000002016 | RAD52 |
| ENSG00000002079 | 0.281046574 | 0.0035829 | 0.20955353 | 0.82142336 | NA | ENSG00000002079 | MYH16 |
| ENSG00000002330 | 789.9075204 | -0.174042988 | 0.1464748 | 0.12017644 | 0.44166397 | ENSG00000002330 | BAD |
| ENSG00000002549 | 1355.715115 | 0.097046084 | 0.14387073 | 0.37066776 | 0.7182969 | ENSG00000002549 | LAP3 |
| ENSG00000002586 | 5427.859497 | -0.079849807 | 0.12366476 | 0.42528155 | 0.75902789 | ENSG00000002586 | CD99 |
| ENSG00000002587 | 14.0636511 | 0.003244259 | 0.20758609 | 0.91441067 | NA | ENSG00000002587 | HS3ST1 |
| ENSG00000002726 | 4.427006818 | -0.028232703 | 0.21198606 | 0.08555968 | NA | ENSG00000002726 | AOC1 |
| ENSG00000002745 | 0.426141397 | 0.000327077 | 0.20941147 | 0.99643794 | NA | ENSG00000002745 | WNT16 |
| ENSG00000002746 | 6.778424773 | -0.03432081 | 0.21225294 | 0.19601233 | NA | ENSG00000002746 | HECW1 |
| ENSG00000002822 | 579.2106434 | -0.195746681 | 0.1606326 | 0.09570945 | 0.39897929 | ENSG00000002822 | MAD1L1 |
| ENSG00000002834 | 3509.829377 | -0.336378482 | 0.14391504 | 0.00433844 | 0.06273718 | ENSG00000002834 | LASP1 |
| ENSG00000002919 | 400.5560554 | -0.192913874 | 0.19033823 | 0.11868947 | 0.43915722 | ENSG00000002919 | SNX11 |
| ENSG00000002933 | 324.1652802 | -0.310728279 | 0.24769471 | 0.0351981 | 0.23300058 | ENSG00000002933 | TMEM176A |
| ENSG00000003056 | 2017.297461 | 0.125887212 | 0.14323321 | 0.24730739 | 0.60918607 | ENSG00000003056 | M6PR |
| ENSG00000003096 | 1376.217893 | -0.129161271 | 0.13139952 | 0.21545208 | 0.57237943 | ENSG00000003096 | KLHL13 |
| ENSG00000003137 | 64.15857327 | -0.084001841 | 0.22106414 | 0.21405424 | 0.57078448 | ENSG00000003137 | CYP26B1 |
| ENSG00000003147 | 727.89575 | 0.247152487 | 0.23051139 | 0.07140124 | 0.34107189 | ENSG00000003147 | ICA1 |
| ENSG00000003249 | 350.6961354 | 0.263728862 | 0.25141864 | 0.05958909 | 0.3132288 | ENSG00000003249 | DBNDD1 |
| ENSG00000003393 | 1578.209098 | 0.235267176 | 0.13115603 | 0.02886147 | 0.20797688 | ENSG00000003393 | ALS2 |
| ENSG00000003400 | 24.21657902 | 0.017359991 | 0.20358825 | 0.73984238 | 0.91751062 | ENSG00000003400 | CASP10 |
| ENSG00000003402 | 1530.027151 | 0.662661454 | 0.14857355 | 7.55E-07 | 4.63E-05 | ENSG00000003402 | CFLAR |
| ENSG00000003436 | 93.27344418 | 0.049404068 | 0.19839689 | 0.53569544 | 0.82558886 | ENSG00000003436 | TFPI |
| ENSG00000003509 | 329.8625828 | 0.046787222 | 0.15467303 | 0.66108517 | 0.88686411 | ENSG00000003509 | NDUFAF7 |
| ENSG00000003756 | 3207.397135 | 0.398952226 | 0.12139782 | 0.00019425 | 0.00550908 | ENSG00000003756 | RBM5 |
| ENSG00000003987 | 16.06949436 | -0.025158615 | 0.20679958 | 0.56813118 | NA | ENSG00000003987 | MTMR7 |
| ENSG00000003989 | 541.3129517 | 0.014700253 | 0.15724464 | 0.88766073 | 0.96856819 | ENSG00000003989 | SLC7A2 |
| ENSG00000004059 | 1739.898844 | 0.016444607 | 0.12177752 | 0.86767201 | 0.96139929 | ENSG00000004059 | ARF5 |
| ENSG00000004139 | 199.5666842 | -0.046045059 | 0.18535691 | 0.62031255 | 0.86714017 | ENSG00000004139 | SARM1 |
| ENSG00000004142 | 2932.948681 | 0.016574618 | 0.10546121 | 0.85753493 | 0.95814418 | ENSG00000004142 | POLDIP2 |
| ENSG00000004399 | 1126.29213 | -0.243468642 | 0.21628853 | 0.06858458 | 0.33342731 | ENSG00000004399 | PLXND1 |
| ENSG00000004455 | 2132.599504 | 0.074739696 | 0.11900821 | 0.45407236 | 0.77851952 | ENSG00000004455 | AK2 |
| ENSG00000004468 | 1.917479523 | -0.002174479 | 0.20914331 | 0.87704591 | NA | ENSG00000004468 | CD38 |
| ENSG00000004478 | 3612.052308 | -0.321775563 | 0.13341838 | 0.00388045 | 0.05770234 | ENSG00000004478 | FKBP4 |
| ENSG00000004487 | 2321.849971 | -0.225409576 | 0.12322038 | 0.0285945 | 0.20780628 | ENSG00000004487 | KDM1A |
| ENSG00000004534 | 1419.609979 | 0.139686673 | 0.14093843 | 0.19434557 | 0.54608562 | ENSG00000004534 | RBM6 |
| ENSG00000004660 | 175.5099327 | -0.512906823 | 0.41822554 | 0.01201227 | 0.12438399 | ENSG00000004660 | CAMKK1 |
| ENSG00000004700 | 178.3814168 | -0.242029631 | 0.30479698 | 0.06324465 | 0.32180364 | ENSG00000004700 | RECQL |
| ENSG00000004766 | 325.6194011 | 0.008138672 | 0.15914688 | 0.93854996 | 0.98469445 | ENSG00000004766 | VPS50 |
| ENSG00000004776 | 487.6706965 | -0.25254246 | 0.23215635 | 0.06383287 | 0.32317385 | ENSG00000004776 | HSPB6 |
| ENSG00000004777 | 442.7739649 | -1.35902878 | 0.20571982 | 2.42E-12 | 5.92E-10 | ENSG00000004777 | ARHGAP33 |
| ENSG00000004779 | 1707.548609 | -0.105399991 | 0.12774501 | 0.30616285 | 0.66593472 | ENSG00000004779 | NDUFAB1 |
| ENSG00000004799 | 88.45048004 | 0.047387909 | 0.193674 | 0.57756819 | 0.84696829 | ENSG00000004799 | PDK4 |
| ENSG00000004809 | 0.207067359 | -0.00454838 | 0.20960728 | 0.71692093 | NA | ENSG00000004809 | SLC22A16 |
| ENSG00000004838 | 2.971271355 | 0.006368466 | 0.20883001 | 0.74734834 | NA | ENSG00000004838 | ZMYND10 |
| ENSG00000004846 | 0 | NA | NA | NA | NA | ENSG00000004846 | ABCB5 |
| ENSG00000004848 | 0.219074038 | 0.004784327 | 0.20959965 | 0.74157219 | NA | ENSG00000004848 | ARX |
| ENSG00000004864 | 820.9199753 | -0.316419746 | 0.17135107 | 0.01341863 | 0.13220132 | ENSG00000004864 | SLC25A13 |
| ENSG00000004866 | 792.2572323 | -0.049548461 | 0.13836872 | 0.63373126 | 0.87360696 | ENSG00000004866 | ST7 |
| ENSG00000004897 | 1080.348288 | -0.324642871 | 0.18926117 | 0.01763597 | 0.15568382 | ENSG00000004897 | CDC27 |
| ENSG00000004939 | 0.109343154 | 0.004041433 | 0.20956579 | 0.82142048 | NA | ENSG00000004939 | SLC4A1 |
