## Supplemental Table 8 for "Functional analysis across model systems implicates ribosomal proteins in growth and proliferation defects associated with hypoplastic left heart syndrome"

| | baseMean | log2FoldChange | lfcSE | pvalue | padj | ENSG | gene |
|---|---|---|---|---|---|---|---|
| ENSG00000000003 | 2923.838943 | -0.062063724 | 0.08422641 | 0.38938233 | 0.68623541 | ENSG00000000003 | TSPAN6 |
| ENSG00000000005 | 21.97529732 | -0.022442385 | 0.15973972 | 0.57762432 | NA | ENSG00000000005 | TNMD |
| ENSG00000000419 | 1551.595726 | -0.100742712 | 0.10603345 | 0.22365965 | 0.51896091 | ENSG00000000419 | DPM1 |
| ENSG00000000457 | 273.3008654 | 0.11018324 | 0.14210209 | 0.22300125 | 0.51817978 | ENSG00000000457 | SCYL3 |
| ENSG00000000460 | 1172.058855 | -0.065621451 | 0.09458045 | 0.4025841 | 0.69698461 | ENSG00000000460 | C1orf112 |
| ENSG00000000938 | 8.944371215 | 0.014552243 | 0.16176923 | 0.57962992 | NA | ENSG00000000938 | FGR |
| ENSG00000000971 | 0 | NA | NA | NA | NA | ENSG00000000971 | CFH |
| ENSG00000001036 | 2274.615867 | -0.079843907 | 0.07634979 | 0.23597114 | 0.53278456 | ENSG00000001036 | FUCA2 |
| ENSG00000001084 | 1808.83403 | -0.135176559 | 0.08792734 | 0.07202479 | 0.2639128 | ENSG00000001084 | GCLC |
| ENSG00000001167 | 2017.948124 | 0.020896695 | 0.07573459 | 0.75694542 | 0.90659629 | ENSG00000001167 | NFYA |
| ENSG00000001460 | 267.5774669 | -0.006434137 | 0.12608896 | 0.93751562 | 0.97899657 | ENSG00000001460 | STPG1 |
| ENSG00000001461 | 473.9583056 | 0.131974787 | 0.12658038 | 0.14467464 | 0.40249885 | ENSG00000001461 | NIPAL3 |
| ENSG00000001497 | 3902.752227 | -0.170374321 | 0.06851221 | 0.00622264 | 0.04969514 | ENSG00000001497 | LAS1L |
| ENSG00000001561 | 75.42690795 | 0.075142918 | 0.16876752 | 0.25850773 | 0.55928858 | ENSG00000001561 | ENPP4 |
| ENSG00000001617 | 1771.237269 | -0.089991272 | 0.08813893 | 0.22944878 | 0.52533561 | ENSG00000001617 | SEMA3F |
| ENSG00000001626 | 61.93122916 | 0.170271558 | 0.26602398 | 0.04410458 | 0.19352728 | ENSG00000001626 | CFTR |
| ENSG00000001629 | 2086.76753 | 0.105968554 | 0.07520532 | 0.11299858 | 0.34767711 | ENSG00000001629 | ANKIB1 |
| ENSG00000001630 | 2786.544179 | -0.36023107 | 0.09895614 | 3.48E-05 | 0.00071899 | ENSG00000001630 | CYP51A1 |
| ENSG00000001631 | 1033.403651 | 0.198565082 | 0.12299319 | 0.0346277 | 0.16510236 | ENSG00000001631 | KRIT1 |
| ENSG00000002016 | 584.8033736 | 0.114664682 | 0.1240727 | 0.19846182 | 0.48434432 | ENSG00000002016 | RAD52 |
| ENSG00000002079 | 0.490351291 | -0.000390398 | 0.16297784 | 0.9975057 | NA | ENSG00000002079 | MYH16 |
| ENSG00000002330 | 572.7702846 | -0.064088371 | 0.10706888 | 0.4344795 | 0.72273431 | ENSG00000002330 | BAD |
| ENSG00000002549 | 2661.345656 | -0.05427923 | 0.08070977 | 0.43465622 | 0.72294711 | ENSG00000002549 | LAP3 |
| ENSG00000002586 | 939.7273394 | -0.002987182 | 0.09184924 | 0.96130595 | 0.98660698 | ENSG00000002586 | CD99 |
| ENSG00000002587 | 78.49177266 | 0.101817032 | 0.18768385 | 0.1418495 | 0.39795414 | ENSG00000002587 | HS3ST1 |
| ENSG00000002726 | 6.936825543 | 0.001090134 | 0.1610844 | 0.96776173 | NA | ENSG00000002726 | AOC1 |
| ENSG00000002745 | 3.702536636 | 0.006718233 | 0.16220242 | 0.71473047 | NA | ENSG00000002745 | WNT16 |
| ENSG00000002746 | 385.6919765 | -0.039134646 | 0.12244813 | 0.63082667 | 0.84634109 | ENSG00000002746 | HECW1 |
| ENSG00000002822 | 1020.22236 | -0.08791551 | 0.10339291 | 0.27199322 | 0.57298869 | ENSG00000002822 | MAD1L1 |
| ENSG00000002834 | 15647.24734 | -0.153301558 | 0.05193158 | 0.00151707 | 0.01727898 | ENSG00000002834 | LASP1 |
| ENSG00000002919 | 716.4469346 | -0.083520098 | 0.1039849 | 0.30541187 | 0.60697332 | ENSG00000002919 | SNX11 |
| ENSG00000002933 | 5.77917767 | 0.0131499 | 0.16265896 | 0.49390008 | NA | ENSG00000002933 | TMEM176A |
| ENSG00000003056 | 4447.872613 | 0.020131828 | 0.06438178 | 0.73410489 | 0.89792567 | ENSG00000003056 | M6PR |
| ENSG00000003096 | 254.0304233 | -0.014169321 | 0.1272118 | 0.85844773 | 0.94815731 | ENSG00000003096 | KLHL13 |
| ENSG00000003137 | 4.040896169 | -0.007138034 | 0.16206549 | 0.70724836 | NA | ENSG00000003137 | CYP26B1 |
| ENSG00000003147 | 657.5689868 | -0.177392653 | 0.11635513 | 0.04872545 | 0.20540933 | ENSG00000003147 | ICA1 |
| ENSG00000003249 | 2943.411796 | 0.096220619 | 0.06963085 | 0.12587474 | 0.37244648 | ENSG00000003249 | DBNDD1 |
| ENSG00000003393 | 2212.685474 | 0.012411649 | 0.07472801 | 0.83658372 | 0.93976935 | ENSG00000003393 | ALS2 |
| ENSG00000003400 | 289.9504135 | 1.163971179 | 0.2029941 | 5.12E-10 | 2.91E-08 | ENSG00000003400 | CASP10 |
| ENSG00000003402 | 404.4600004 | 0.243979768 | 0.17355492 | 0.03034149 | 0.1523036 | ENSG00000003402 | CFLAR |
| ENSG00000003436 | 76.85577442 | 0.058949204 | 0.1606049 | 0.36273618 | 0.66438621 | ENSG00000003436 | TFPI |
| ENSG00000003509 | 1532.920777 | 0.059053601 | 0.0841867 | 0.41492798 | 0.70671296 | ENSG00000003509 | NDUFAF7 |
| ENSG00000003756 | 3694.953878 | 0.102459367 | 0.06614906 | 0.08937801 | 0.3012989 | ENSG00000003756 | RBM5 |
| ENSG00000003987 | 182.9471554 | 0.021947155 | 0.13389607 | 0.77250399 | 0.91262637 | ENSG00000003987 | MTMR7 |
| ENSG00000003989 | 1147.046134 | -0.04862368 | 0.09416571 | 0.52585298 | 0.78552379 | ENSG00000003989 | SLC7A2 |
| ENSG00000004059 | 3837.880708 | -0.093143053 | 0.06271056 | 0.10632955 | 0.33580378 | ENSG00000004059 | ARF5 |
| ENSG00000004139 | 386.485291 | -0.005760435 | 0.11895239 | 0.94134314 | 0.97986415 | ENSG00000004139 | SARM1 |
| ENSG00000004142 | 4822.483128 | -0.046868996 | 0.07651906 | 0.50284609 | 0.77214232 | ENSG00000004142 | POLDIP2 |
| ENSG00000004399 | 1757.908474 | 0.088413262 | 0.08087131 | 0.20906771 | 0.49846585 | ENSG00000004399 | PLXND1 |
| ENSG00000004455 | 4367.782977 | -0.23983956 | 0.06902318 | 0.0001536 | 0.00259293 | ENSG00000004455 | AK2 |
| ENSG00000004468 | 10.05626945 | 0.021606338 | 0.16289014 | 0.41848452 | NA | ENSG00000004468 | CD38 |
| ENSG00000004478 | 16194.2816 | -0.299232198 | 0.06056394 | 1.55E-07 | 5.77E-06 | ENSG00000004478 | FKBP4 |
| ENSG00000004487 | 13185.99056 | -0.262045614 | 0.05446895 | 4.37E-07 | 1.49E-05 | ENSG00000004487 | KDM1A |
| ENSG00000004534 | 3640.259212 | -0.152254888 | 0.07430033 | 0.02144312 | 0.11915018 | ENSG00000004534 | RBM6 |
| ENSG00000004660 | 315.556503 | -0.091101761 | 0.13059584 | 0.29657016 | 0.59872303 | ENSG00000004660 | CAMKK1 |
| ENSG00000004700 | 1574.919169 | 0.132443321 | 0.10872073 | 0.11795339 | 0.357211 | ENSG00000004700 | RECQL |
| ENSG00000004766 | 369.3893506 | 0.237928493 | 0.2077203 | 0.04103339 | 0.18493522 | ENSG00000004766 | VPS50 |
| ENSG00000004776 | 22.25104063 | -0.062248343 | 0.17540989 | 0.11916497 | NA | ENSG00000004776 | HSPB6 |
| ENSG00000004777 | 658.491433 | -0.016075509 | 0.10292232 | 0.84036524 | 0.94057572 | ENSG00000004777 | ARHGAP33 |
| ENSG00000004779 | 3114.024751 | -0.182521546 | 0.07447472 | 0.00625276 | 0.04972781 | ENSG00000004779 | NDUFAB1 |
| ENSG00000004799 | 4.13358193 | 0.018140787 | 0.16370107 | 0.27256354 | NA | ENSG00000004799 | PDK4 |
| ENSG00000004809 | 5.270047132 | 0.004642191 | 0.16158872 | 0.83669125 | NA | ENSG00000004809 | SLC22A16 |
