## Supplemental Table 9 for "Functional analysis across model systems implicates ribosomal proteins in growth and proliferation defects associated with hypoplastic left heart syndrome"

| RPS15a KD | log2FoldCh | pvalue |
|---|---|---|
| KIFC1 | -2.16879 | 2.6E-15 |
| PKMYT1 | -2.00214 | 2.5E-14 |
| HMGA1 | -1.08576 | 3.77E-08 |
| MDC1 | -0.85598 | 1.81E-10 |
| RABL6 | -0.46948 | 3.14E-05 |
| EWSR1 | -0.32331 | 0.001473 |
| FAT4 | 0.254803 | 0.046054 |
| TTN | 0.594686 | 2.66E-05 |
| COL4A5 | 0.614801 | 2.05E-05 |
| DMD | 0.642083 | 9.03E-09 |
| SYNE1 | 0.852181 | 1.04E-12 |
| EPHA4 | 0.980925 | 1.46E-09 |

| RPL39KD-C | log2FoldCh | pvalue |
|---|---|---|
| KIFC1 | -2.59969 | 9.44E-21 |
| PKMYT1 | -2.35329 | 1.53E-18 |
| HMGA1 | -0.93341 | 1.39E-06 |
| MDC1 | -1.07829 | 3.47E-15 |
| RABL6 | -0.47047 | 3.5E-05 |
| EWSR1 | -0.45131 | 2.48E-05 |
| FAT4 | 0.367179 | 0.011637 |
| TTN | 0.638153 | 4.91E-06 |
| COL4A5 | 0.453809 | 0.001024 |
| DMD | 0.482461 | 1.02E-05 |
| SYNE1 | 0.834024 | 2.69E-12 |
| EPHA4 | 1.012869 | 4.8E-10 |

| RPS15a KD | Normalized |
|---|---|
| EWSR1 | 0.517291 |
| PKMYT1 | 0.79284 |
| DMD | 0.797066 |
| MDC1 | 0.833751 |
| KIFC1 | 0.834044 |
| SYNE1 | 0.971289 |
| TTN | 1.058523 |
| HMGA1 | 1.11364 |
| RABL6 | 1.118545 |
| COL4A5 | 1.122295 |
| FAT4 | 1.247476 |
| EPHA4 | 1.308838 |

d % EdU+CM
