## Supplementary material for "Functional analysis across model systems implicates ribosomal proteins in growth and proliferation defects associated with hypoplastic left heart syndrome": Table 1

| **Gene** | **Proband** (sex) | **Mode of Inheritance** | | **Variant** | **Type** | **MAF** | **CADD score** | **TFBS affected** | **hPSC-CM proliferation** | ***Fly gene* and defects** | ***Zebrafish gene* and defects** | **Cardiac Phenotype** |
|---|---|---|---|---|---|---|---|---|---|---|---|---|
| *RPL26L1* | 145H (m) | Compound heterozygous | maternal | -1248A>G | Regulatory | 0.029 | - | Pdx1; NFE2L1::MAFG; FOXC1 | reduced | *RpL26:* lethal, no heart | *rpl26:* n.t. | MS/AS; Restrictive ASD, plastic bronchitis at 1-5y |
| | | | paternal | V97M | Missense | 0.032 | 21.3 | - | | | | |
| *RPL36A* | | X-linked recessive | maternal | -1321C>T | Regulatory | 0.055 | - | PAX2 | no effect | *RpL36A*: lethal, no heart | *rpl36a:* n.t. | |
| *RPS15* | | Compound heterozygous | paternal | -1558C>T | Regulatory | 0 | - | FOXC1 | reduced | *RpS15:* lethal | *rps15:* n.t. | |
| | | | maternal | T101S | Missense | 0.102 | 23.5 | - | | | | |
| *RPL39* | 151H (m) | X-linked recessive | maternal | -1359T>C | Regulatory | 0.653 | - | HOXA5 | reduced | *RpL39:* lethal | *rpl39*: morphants mild edema, reduced ventricular size | MS/AS; Reduced RV function at 15-20y |
| *RPL3L* | 96H (m) | Compound heterozygous | maternal | R200Q | Missense | 0.966 | 21.2 | - | elevated | *RpL3:* no heart | *rpl3:* n.t. | MA/AA; Reduced RV function <5y |
| | | | paternal | R242W | Missense | 0.432 | 14.94 | | | | | |
| RPL13A | 201H (m) | Compound heterozygous | maternal | -92-645C>T | Regulatory | 0.72 | - | FOXD1; GATA2; ETS1; ELF5 | no effect | *RpL13A:* no phenotype | *rpl13a:* n.t. | Failing Fontan circulation, transplant at 10-15y |
| | | | paternal | -29-191C>T | Regulatory | 0.046 | - | FOXC1 | | | | |
| RPS17 | 325 (f) | Homozygous Recessive | maternal + paternal | S136N | Missense | 0 | < 10 | - | reduced | *RpS17:* lethal | *rps17:* CRISPR mutants show systolic dysfunction in atrium, shortened heart period | MA/AA; Reduced RVEF & increased RVEDP at <5y |
| RPL10 | 76 (m) | X-linked Recessive | maternal | 24-218G>A | Regulatory | 0.583 | - | ELK1; ETS1; SPIB; POLR2A; HEY1; Hltf | reduced | *RpL10:* constricted | *rpl10, n.t.* | MS/AA; Reduced RVEF at 5-10y |
| RPS28 | | Compound Heterozygous | paternal | -589G>A | Regulatory | 0.061 | - | CTCF | reduced | *RpS28b,* ostia defect | *rps28,* CRISPR mutants show no heart phenotype | |
| | | | maternal | -505A>T | Regulatory | 0.08 | | CTCF | | | | |
